# Compartment-specific total RNA profile of Hippocampal and Cortical cells from Mesial Temporal Lobe Epilepsy tissue

**DOI:** 10.1101/2021.12.03.21266858

**Authors:** Vamshidhar R. Vangoor, Giuliano Giuliani, Marina de Wit, Morten T. Venø, Noora Puhakka, Andreia Gomes-Duarte, Peter C. van Rijen, Peter H. Gosselaar, Pieter van Eijsden, Jørgen Kjems, Pierre N.E. de Graan, R. Jeroen Pasterkamp

**Author notes:** **Correspondence:** R. Jeroen Pasterkamp, Department of Translational Neuroscience, UMC Utrecht, Universiteitsweg 100, 3584 CG, Utrecht, The Netherlands. Equal contribution (Vamshidhar R. Vangoor, Giuliano Giuliani). Noora Puhakka: A.I. Virtanen Institute for Molecular Sciences, University of Eastern Finland, Kuopio, Finland.

## Abstract

Mesial temporal lobe epilepsy (mTLE) is a chronic neurological disease characterized by recurrent seizures. The pathogenic mechanisms underlying mTLE involve defects in post-transcriptional regulation of gene expression. So far, transcriptome profiles from epileptic tissue have been generated using whole cells, thereby lacking information on RNA localization and function at a subcellular level. In line with this, we have previously observed by *in situ* hybridization that a few microRNAs (miRNAs) display subcellular mis-localization with aberrant enrichment in the nucleus in human hippocampal mTLE tissue samples (Kan et al., 2012). To further investigate the possible mechanisms leading to the mis-localization of miRNAs, we set out to understand the compartment-specific total RNA (coding and non-coding) profile of human mTLE tissue samples. For this, we have successfully established a protocol to isolate cytoplasmic and nuclear compartments from human hippocampal tissue. After confirming the purity of the isolated cell compartments, we performed total RNA-sequencing (RNA-seq) on five resected hippocampal (HC) mTLE (no hippocampal sclerosis (non-HS)) samples and five HC postmortem control samples. Similarly, six neo-cortical (Cx) tissue samples from mTLE non-HS and HS International League Against Epilepsy (ILAE) Type 1, or mTLE+HS, samples were compared with six Cx postmortem controls. Our dataset provides a comprehensive overview of compartment-specific transcriptomic profiles of pharmacoresistant mTLE patient HC and Cx tissue, which in further studies can be used to investigate disease mechanisms.

## Introduction

Epilepsy is one of the most common and severe neurological diseases affecting 70 million individuals worldwide, and is characterized by recurrent unprovoked seizures (Chang and Lowenstein, 2003; Ngugi et al., 2010). Mesial temporal lobe epilepsy (mTLE) is the most common form of focal epilepsy where seizures originate from temporal lobe structures in the brain (Engel, 2001). Approximately one-third of mTLE patients develop drug-resistance for antiepileptic drugs (AEDs) (Kwan et al., 2011; Kwan and Brodie, 2000). Even in respondents, available AEDs often have side-effects, acting as seizure suppressors rather than disease modifiers (Löscher et al., 2013). Currently, in approximately 60% of drug-resistant patients, surgical resection of epileptic *foci* is performed as a last resort to attain seizure freedom (Schuele and Lüders, 2008). However, not all resection surgeries yield in complete seizure freedom, and a significant number of patients do for several reasons not qualify for surgery (e.g., *foci* not localized, other brain functions are impaired (memory, language), suspected genetic or metabolic cause (Lamberink et al., 2020; Moshé et al., 2015)). Hence, there is an urgent and unmet need for developing disease modifying therapies to treat mTLE.

Resected tissue from 60-80% of drug-resistance mTLE patients is associated with hippocampal sclerosis (HS), referred as HS International League Against Epilepsy Type1 (HS ILAE Type1) (Blümcke et al., 2013). HS ILAE Type 1, or mTLE+HS, tissue is characterized by predominant neuronal cell loss in Cornus ammonis 1 (CA1) together with variable cell loss in other CA regions, and displays gliosis, granule cell dispersion and mossy fiber sprouting (Blümcke et al., 2013; Proper et al., 2000). Approximately 20% of mTLE patient tissue displays focal lesions in the temporal lobe, but shows no apparent hippocampal damage with reactive gliosis only, and it is classified as mTLE non-HS (Blümcke et al., 2013). Structural and functional changes such as neurodegeneration and reorganization of neuronal networks are key pathological processes occurring during epileptogenesis (Pitkänen and Lukasiuk, 2011). Accumulating evidence from transcriptomics studies performed on resected human TLE patient tissue indicates large-scale deregulation of the gene expression landscape within the brain, affecting critical pathways and key functional networks underlying inflammation, gliosis, synaptic structure and neuronal function (Dixit et al., 2016; Goldberg and Coulter, 2013; Guelfi et al., 2019; Johnson et al., 2015; Okamoto et al., 2010; Salman et al., 2017a; Srivastava et al., 2018; van Gassen et al., 2009; Wang et al., 2010).

Previous studies have provided important information about the pathways and specific genes contributing towards epileptogenesis and seizure generation. However, these studies did not take into account the subcellular localization of gene expression. It is becoming increasingly evident that it is important to understand the subcellular repertoire of different RNA molecules and investigate the mechanisms that govern their distribution within the cells (Martin and Ephrussi, 2009; Taliaferro et al., 2014; Zaghlool et al., 2021; Zhang et al., 2016). Moreover, it has been proven that mRNA subcellular localization provides a means to spatially control protein production and transport (Barthelson et al., 2007; Russo et al., 2006; Steward and Schuman, 2003). Further, long non-coding RNA (lncRNAs) molecules often exert specific functions in distinct cell compartments, such as the nucleus or cytoplasm (Chen, 2016; Vangoor et al., 2020; Zaghlool et al., 2021). Similarly, in our previous study, we observed miRNAs displaying disease-specific nuclear mis-localization in mTLE hippocampus (Kan et al., 2012). Therefore, understanding disease-specific alterations in RNA localization in cytoplasmic and nuclear compartments will help dissecting mechanisms contributing to refractory mTLE.

In this study, we established a protocol to isolate pure nuclear and cytoplasmic fractions from human tissue. Subcellular RNA was isolated using resected hippocampal and cortical tissue from mTLE non-HS and mTLE+HS patients and postmortem control tissue. This tissue was used for total RNA sequencing (RNA-seq). The generated dataset constitutes a resource for further dissection of the pathogenic mechanisms underlying mTLE.

## Materials and Methods

### Human tissue

Human pharmacoresistant mTLE patient hippocampus (HC) and neocortical (Cx) tissue was obtained after surgery, as described previously (Kan et al., 2012; Vangoor et al., 2019) at the University Medical Center Utrecht. Informed consent was obtained from all patients for use of tissue and clinical information for research purposes and all procedures were approved by the Review board of University Medical Center Utrecht (Utrecht, The Netherlands). Postmortem human HC and Cx tissue was obtained from the Netherlands Brain Bank (Amsterdam, The Netherlands). A written informed consent was obtained from all donors for brain autopsy and for use of the material and clinical information for research purposes. Use of postmortem tissue and clinical information for research purposes was approved by the medical ethics board of the Amsterdam University Medical Center (Amsterdam, The Netherlands). For HC, a total of five HC mTLE (no hippocampal sclerosis (mTLE non-HS) (male and female)) and five control HC (male and female) samples were used. For Cx, six mTLE (non-HS (male and female) and six HS International League Against Epilepsy (ILAE) Type 1 (male and female) or mTLE+HS) and six control samples were used (Supplementary Table S1).

### Nuclear-cytoplasmic fractionation of human hippocampus and cortex

Before fractionation, frozen brain samples were powdered in dry ice using a mortar pestle. Approximately 500 mg of powdered tissue was resuspended in Hypotonic Lysis Buffer (HLB) (10 mM Tris (pH 7.4), 3 mM CaCl_2_, 2 mM MgCl_2_, 1% Nonidet P-40 (Sigma), Protease inhibitor (Roche), 60 U Superase-In/mL (ThermoFisher Scientific)) for 10 minutes (min) on ice to lyse: homogenization and lysis were enhanced with the help of 30-80 strokes in a tightly fitting glass Douncer. The cytoplasmic fraction was collected following a gentle centrifugation (500 g) for 5 min at 4°C, and frozen in Qiazol (Qiagen) until RNA purification. The semi-pure nuclear fraction (pellet) was resuspended in 2 mL Sucrose Buffer I (0.32 M Sucrose, 5 mM CaCl_2_, 3 mM Mg(CH_3_COO)_2_, 0.1 mM EDTA, 10 mM Tris (pH 8.0), 0.5% Nonidet P-40, 1 mM DTT, 25 U Superase-In/mL) and placed on top of two thicker sucrose cushions: Sucrose Buffer II (1.6 M Sucrose, 3 mM Mg(CH_3_COO)_2_, 0.1 mM EDTA, 10 mM Tris (pH 8.0), 1 mM DTT, Protease Inhibitor, 35 U Superase-In/mL), and a Sucrose Buffer III (2.0 M Sucrose, 3 mM Mg(CH_3_COO)_2_, 0.1 mM EDTA, 10 mM Tris (pH 8.0), 1 mM DTT, 35 U Superase-In/mL, Protease Inhibitor), respectively, in ultracentrifugation tubes (Beckman Coulter). Samples were centrifuged using a SW41.4 rotor at 30000 g at 4°C for 45 min. Pure nuclei were collected and stored in Qiazol (Qiagen) at −80°C until RNA extraction was performed.

### RNA purification

Total RNA was extracted from fractionated human samples using miRNeasy Mini Kit (Qiagen) according to the manufacturer’s instructions and DNaseI treated using RNase free-DNase (Catalogue#79254, Qiagen) to eliminate genomic DNA contamination. When required, RNA was further purified using RNA Clean-Up and Concentration Kit (Catalogue#23600, Norgen Biotek Corporation). Finally, RNA integrity (RIN) value was estimated in an Agilent 2100 Bioanalyzer (Agilent Technologies) (Supplementary Table S1).

### Quality control fractionation procedure

The efficiency of the fractionation protocol was estimated at the protein level by western blotting (WB) and at the RNA level by quantitative PCR (RT-qPCR). For WB analysis, a fraction of cytoplasmic and nuclear samples were collected in RIPA buffer (50 mM Tris, pH.7.5, 150 mM NaCl, 0.5% NP-40, 0.5% NaDoc, 1% Triton, Protease inhibitors in MilliQ). Nuclear sample was lysed by passing through 27 g needle (BD Plastipak) for five times and by sonication. 12 µl of the cytosol and 6 µl of nuclear protein samples were resuspended in 4x NuPAGE LDS Sample buffer (ThermoFisher Scientific) and heated at 95°C for 5 min. The samples were separated in SDS-PAGE gels (8%) for approximately 1 h at 160 V, until the dye ran out of the gel. Separated proteins were transferred onto nitrocellulose blotting membrane (GE Healthcare) for 1 h at 100 V under cold conditions. Then blots were blocked for 1 h at RT in 5% milk powder in 1xTBS-Tween. Blots were incubated overnight at 4°C with antibodies against the nuclear-enriched protein fibrillarin (1:1000, Catalogue# 2639S, Cell signalling) or the cytoplasmic-enriched protein β-tubulin (1:2000, Catalogue# T5168, Sigma) in blocking buffer. Blots were stained with peroxidase-conjugated secondary antibodies (1:30000 for HRP-anti-mouse; 1:50000 for HRP-anti-rabbit) for 1 h at RT in 1xTBS-Tween, and signal was detected by incubating blots with Pierce ECL substrate (Thermo Fisher Scientific). Images were acquired using a FluorChem M imaging system (Protein Simple).

RNA levels of nuclear-enriched markers (*NEAT1, GOMAFU, pre-GAPDH*) and cytoplasmic-enriched markers designed to amplify an intron spanning amplicon (*GAPDH*) were measured by RT-qPCR, both in cytoplasmic and nuclear preparations. Briefly, 100 ng of total RNA reaction was reverse transcribed using the SuperScript III First-Strand Synthesis System (ThermoFisher Scientific) with random hexamers, according to the manufacturer’s instructions. RT-qPCR quantification of mRNA was conducted using FastStart Universal SYBR Green Master (Rox) (Roche) for all samples, in triplicates, in a QuantStudio(tm) 6 Flex Real-Time PCR System (ThermoFisher Scientific). Real-time reactions were carried out as follows: pre-denaturation at 95°C for 10 min, followed by 40 cycles at 95°C for 15 s and 60°C for 1 min. Melting curves were generated for the final PCR products by decreasing the temperature to 60°C for 1 min followed by an increase in temperature to 95°C. Primer sequences used for RT–qPCR reactions are listed in Supplementary Table S3.

### Total RNA library preparation and sequencing

A total of 2 µg of RNA per sample in a 5 µl volume was rRNA-depleted using the Ribo-Zero Magnetic Kit (human/mouse/rat; Illumina) before library preparation. Stranded, paired-end sequencing libraries were prepared using ScriptSeq v2 (Epicentre), after which samples were subjected to quality control in the 2100 Bioanalyzer (Agilent). Sequencing was performed as paired end 100 bp reads on an Illumina HiSeq 4000 sequencer at the Beijing Genomics Institute (BGI), Hongkong, China.

### Read mapping and differential expression analysis

Read mapping and differential expression analysis was performed as described previously (Vangoor et al., 2019). Briefly, following trimming of low-quality bases and adapter sequences with FASTQ-MCF (version 0.0.13), processed reads were mapped to the hg19/GCh37 reference human genome (iGenomes) with TopHat2 (version 2.0.13) (Kim et al., 2013). For the alignments of the total RNA-seq data, the ‘fr-secondstrand’ option was chosen. Mapped counts for each gene were summarised using the python script htseq-count (Anders et al., 2015). Alignment statistics were obtained from Bam files generated using Bowtie (Langmead et al., 2009). All raw and processed RNA-seq data are deposited at NCBI Gene Expression Omnibus (GEO) with reference number GSEXXXX.

Mapped counts were visualized using a principal component analysis (PCA; R version 4.0.3, package ggfortify). For differential expression analysis, genes and transcripts count data was analysed for differential expression in R using the Bioconductor package EdgeR version 3.12.1 (Robinson et al., 2010), with the trimmed mean of M-values (TMM) normalisation method (Robinson and Oshlack, 2010). Gene expression levels were corrected for batch and gender effects by including the series of sequencing rounds, sexes of controls and patients to a generalised linear model. Using the Benjamini-Hochberg false discovery rate (FDR) adjusted P-values for multiple testing were calculated and only genes with an FDR < 0.05 were considered significantly differentially expressed (DE). Visualisation of data was performed in R using the ggplot2 library (version 2.1.0). Using the Bioconductor heatmap package in R, gene expression heatmaps with hierarchical clustering of expression profiles were created. To estimate proportion of protein coding RNAs, total RNA-seq reads were pseudoaligned to the human protein coding transcriptome (Homo_sapiens.GRCh38.cds.all.fa from the Ensembl FTP site; (Yates et al., 2019)) using the Kallisto algorithm (Bray et al., 2016). To set the number of bootstraps to 100, the extra argument -b 100 was passed to the function.

### Validating the expression of differentially expressed (DE) genes by RT-qPCR

RNA from an independent set of mTLE-HS and control samples were used to validate DE genes. Reverse transcription (RT) was performed with the High-Capacity RNA-to-cDNA Kit (Catalogue# 4387406, Thermo Fisher) according to manufacturer’s instructions. Briefly, 2x RT Buffer Mix and 20x RT Enzyme Mix were mixed with RNA (20 ng) and nuclease-free water in a final reaction volume of 20 μl. Reactions were then incubated in a thermal cycler with the following program: 60 min at 37°C and 5 min at 95°C, with the infinite hold set at 4°C. For RT-qPCR, reaction mixes were prepared according to manufacturer’s instructions. Briefly, 1x reaction was prepared by mixing 2x Universal Master Mix (Catalogue # 4440040, Thermo Fisher), 20x Taqman Gene Expression Assay, and nuclease-free water in a final volume of 9 μl. Next, 1 μl of cDNA was added to the reaction mix. Finally, 384-well RT-qPCR plates were spun down for 5 min in a centrifuge at 5000 g. The RT-qPCR reactions were performed in a QuantStudioTM 6 Flex Real-Time PCR System, using the following program: 10 min at 95°C and 40x (15 s at 95°C + 1 min at 60°C). Pre-designed gene expression assays were obtained from ThermoFisher Scientific: *BTG2* (Hs00198887_m1), *IL1ß* (Hs01555410_m1), *FAM126A* (Hs00261521_m1), *WISP1* (Hs05047584_s1) and *HLA-C* (Hs05049205_g1). *GAPDH* (Hs03929097_g1) was used as endogenous control. Every sample was run in triplicate and fold changes were estimated using the 2^-ΔCt^ method. Statistical significance between conditions was estimated by performing Mann-Whitney tests using Graphpad Prism software (Version 9.2.0).

### Pathway analysis using Reactome

Pathway enrichment analysis was performed using the Reactome pathway analysis tool (https://reactome.org), by applying the hypergeometric distribution for P value comparison (Fabregat et al., 2017). Analysis was performed by using the DE gene list containing 174 genes for the cytoplasm and 337 genes for the nuclear compartment of the HC, 99 genes for the cytoplasm and 138 genes for the nuclear compartment of Cx mTLE non-HS and 223 genes for the cytoplasm and 409 genes for the nuclear compartment of Cx mTLE+HS. Enrichment of pathways was considered significant with a corrected P value of < 0.05 (Benjamin-Hochberg).

## Results

### Total RNA profile of individual cell compartments

To understand the subcellular RNA profile of human mTLE patient hippocampal and neo-cortical tissue, total RNA-seq was performed on 1) five mTLE non-HS HC samples and compared to five control HC, and 2) six mTLE non-HS, six mTLE+HS Cx samples were compared separately to six control Cx samples (Figure 1A). Nuclear and cytoplasmic fractions of human brain tissue from patients and controls were successfully isolated by lysing tissue in hypotonic lysis buffer followed with ultracentrifugation. The purity of nucleus samples had evidently improved after ultracentrifugation (Figure 1B). Fractionation success was examined at both the protein and RNA level. The cytoplasmic protein β-Tubulin was only observed in the cytoplasmic samples, whereas the nucleus-enriched protein Fibrillarin was observed in nuclear samples (Figure 1C). At the RNA level, the cytoplasmic marker gene *GAPDH* was enriched in cytoplasmic samples, whereas in nuclear samples several nucleus-specific lncRNAs were found (*GOMAFU, MALAT1, NEAT1)* in addition to *GAPDH* intronic transcripts (Figure 1D).

**Figure 1:**
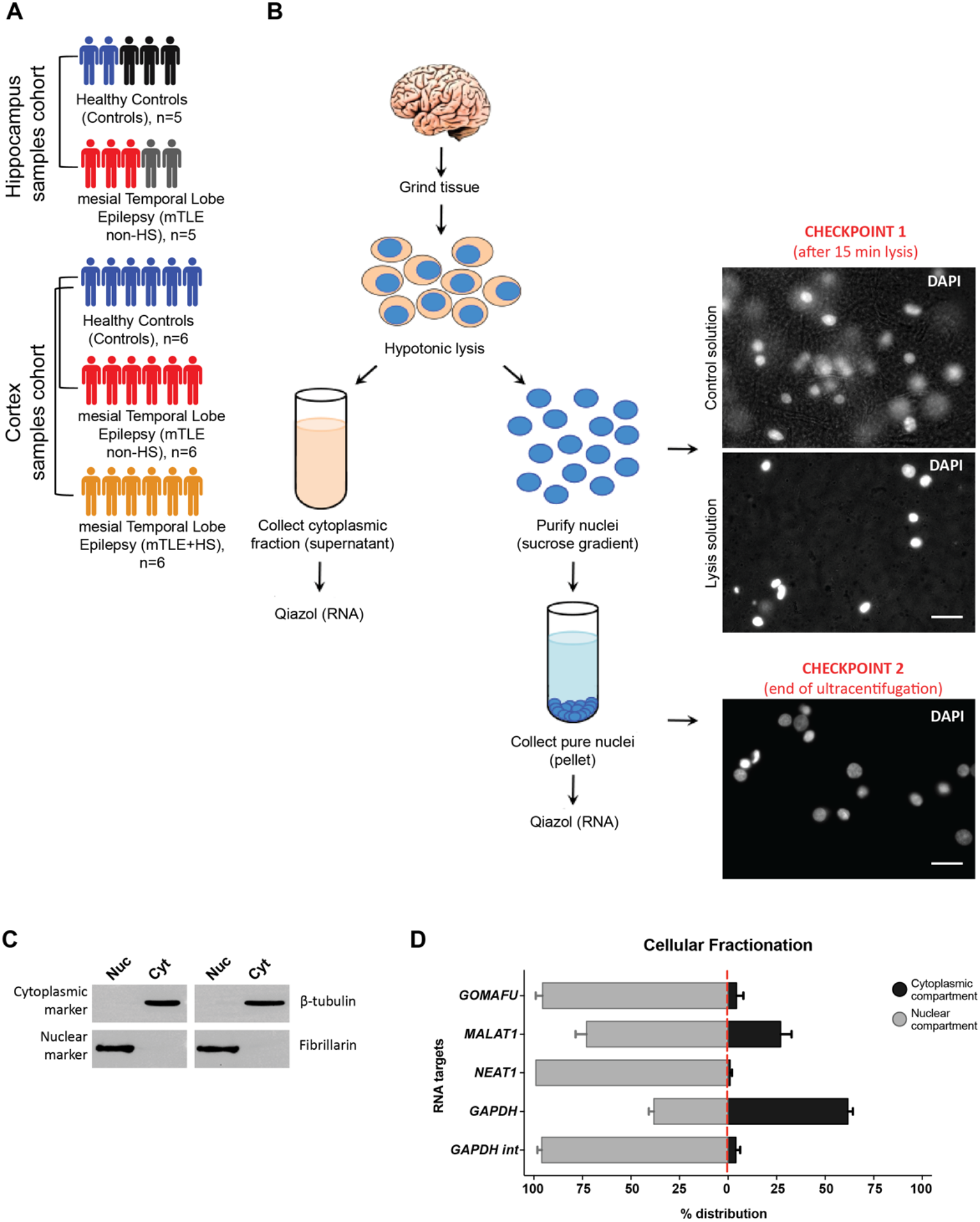
Nuclear fractionation and RNA-seq of mTLE brain tissue. (A) Summary legend of the cohorts (cortical and hippocampal tissue) used for the RNA-seq experiment. For hippocampus (HC), a total of five mTLE non-HS HC samples and five control HC samples were used. For Cx, six mTLE non-HS, six mTLE+HS and six control samples were used. Similar color codes indicate that both HC and Cx tissue were obtained from the same individual control or mTLE non-HS patient. (B) Overview of the cell fractionation procedure for the isolation of RNA from nuclear and cytoplasmic fractions. DAPI (4',6-diamidino-2-phenylindole). Scale bars = 20 µm. (C) Western blot analysis of the lysates of the nuclear and cytoplasmic fractions. Fibrillarin and β-Tubulin were used as nuclear and cytoplasmic markers, respectively. Consistent with its reported subcellular localisation, fibrillarin was specifically detected in the nuclear fraction, whereas anti-β-Tubulin antibody showed signal only in cytoplasmic fractions. Nuc: nucleus, Cyt: cytoplasm. (D) Bar plot showing the enrichment of transcripts in the nuclear and cytoplasmic fractions. As expected, RT-qPCR detected nuclear enrichment for *GOMAFU, MALAT1, NEAT1* and *GAPDH int* (an intron retained variant of *GAPDH*). Higher levels of the spliced form of *GAPDH* were reported in the cytoplasmic fraction. Enrichment is represented as a percentage of the distribution (% distribution) of a marker across the nuclear and cytoplasmic compartments.

Total RNA-seq was performed and an average of 61 million high-quality reads per cytoplasmic and 65 million reads per nuclear samples were obtained for Cx samples. For HC samples, 51 and 58 million reads in cytoplasm and nuclear samples, respectively, were obtained. Of these, 59% reads were uniquely mapped to the reference human genome in cytoplasmic Cx samples, and 62% in the nuclear Cx samples. For HC samples, 63% of reads mapped to the genome in cytoplasmic fractions and 66% in nuclear samples. We observed that approximately 36% of reads from cytoplasmic samples were aligned to exons, whereas in the nuclear samples approximately 15% of reads were aligned to exons. We hypothesize that the lower level of alignment in nucleus could be due to the high level of intronic or intergenic sequences present in this compartment (Supplementary Table S2). Due to the presence of nascent transcripts, an increased alignment of nuclear samples with intronic and intergenic regions has also been shown in recent studies performing total RNA-seq from human brain nuclear and cytosolic fractions (Zaghlool et al., 2021). Principal component analysis (PCA) showed a clear separation between cytoplasmic and nuclear samples, indicating differential RNA profiles between different cell compartments (Figure 2A, D, E). It is, however, important to note that the separation of control and mTLE samples was less clear (for both compartments (nuclear and cytoplasmic) and tissue types) (HC – Figure 2B, 2C, Cx – Figure 2D, 2E).

**Figure 2:**
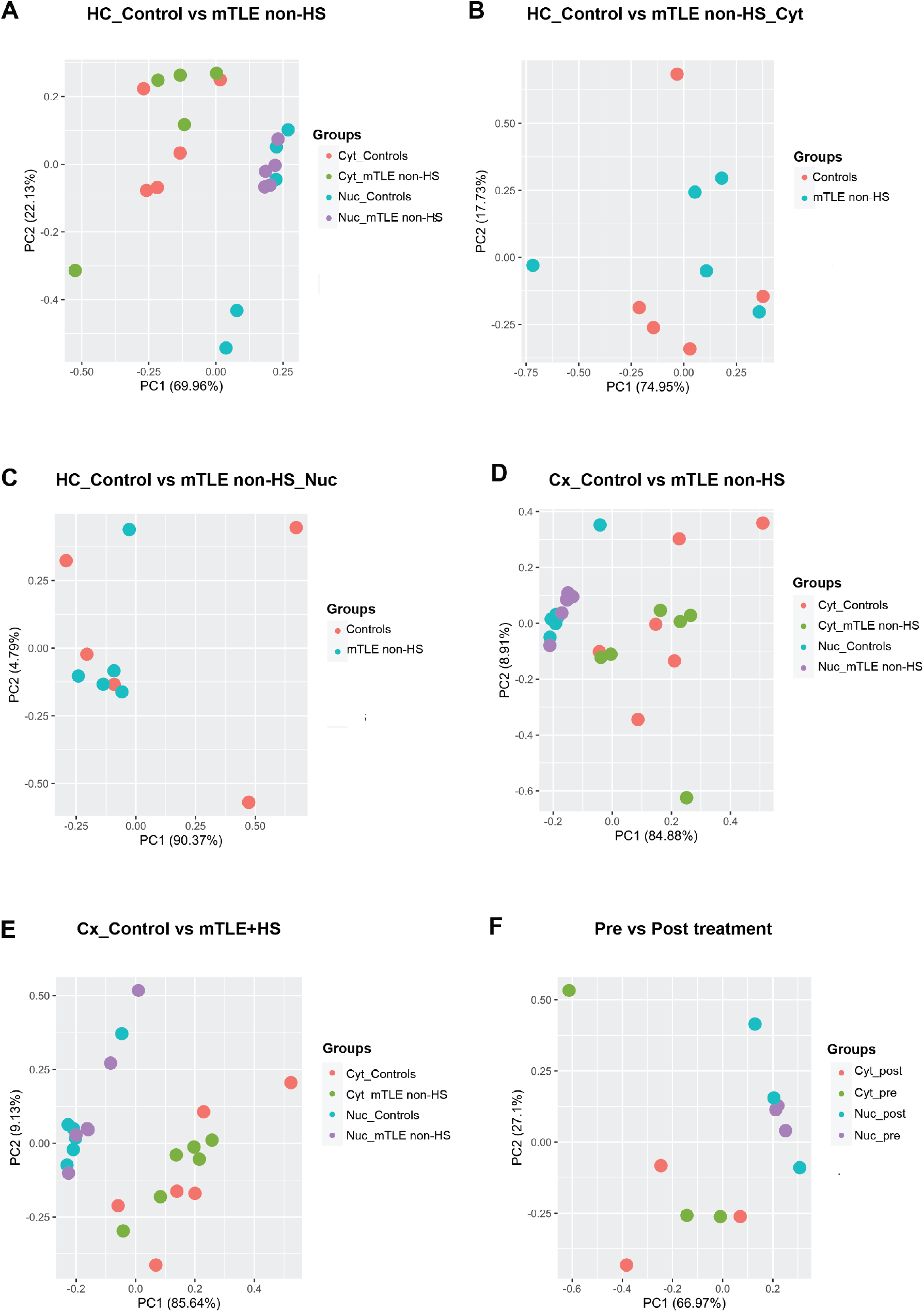
Principal component analysis (PCA) plots of mRNA raw count data. PCA plots showing variance captured by the first two PCs along the respective axes. (A) Cytoplasmic and nuclear samples are clustered separately for HC control *versus* mTLE non-HS group. However, individual comparison of HC control *versus* mTLE non-HS group for each compartment did not result in separate clusters either in Cyt (Cytoplasmic) (B) or Nuc (Nuclear) samples (C). For Cx, separate clustering of cytoplasmic and nuclear samples was observed for control *versus* mTLE non-HS (D) and control *versus* mTLE+HS groups (E). (F) Similarly, pre- and postmortem treatment groups were separated on basis of compartments.

### Postmortem effect on gene expression

Postmortem delay is a common feature of control tissue samples obtained from brain banks (Ferrer et al., 2008). It is often not clear to which extent postmortem delay in control tissues will affect transcript levels and affect differential gene expression analysis when compared with samples collected without any postmortem delay. To estimate this potential confounding effect, resected Cx tissue obtained directly from the surgery room was subjected to postmortem delay treatment. Briefly, tissue was cut into two halves, where one half (pre-) was stored immediately on dry ice and then at −80°C until processing. The second half (post-) was incubated at 4°C in saline for 24 h to mimic postmortem conditions observed for tissue samples collected from brain banks. Cytoplasmic and nuclear fractions were collected from the pre- and postmortem samples as described above. RNA-seq of the pre- and postmortem samples did not show strong differences in the overall coverage and mapping of reads (Supplementary Table S2). PCA plots of the pre- and postmortem-treated samples showed an overall similar RNA profile in cytoplasmic and nuclear fractions (Figure 2F). Finally, no major changes in RNA expression were found in the same sample before and after postmortem delay, either in cytoplasmic or nuclear fractions (Supplemental Figure S1A). In comparison, significant differences in gene expression were observed in Cx samples between control and mTLE+HS or mTLE non-HS (Supplemental Figure S1B-S1C). Together, these data show that postmortem delay is unlikely to cause major differences in the transcriptomic profiles of control tissue.

### Differential gene expression and pathway analysis

Several transcripts were found to be differentially expressed in cytoplasmic and nuclear compartments (Figure 3A-B). Several of the differentially expressed genes (DEGs) were unique to a given compartment, whereas other transcripts were altered in both the nuclear and cytoplasmic compartments, in mTLE non-HS HC nucleus and cytoplasm (Figure 3C), mTLE non-HS Cx nucleus and cytoplasm (Figure 3D) and mTLE+HS Cx nucleus and cytoplasm (Figure 3E). A few transcripts commonly upregulated in hippocampal cytoplasmic and nuclear samples (*IL1β, FAM126A, BTG2, WISP1*) were confirmed using RT-qPCR in an independent set of unfractionated patient and control samples. A significant upregulation of the selected transcripts in mTLE non-HS samples compared to control samples was found, confirming the RNA-seq analysis: *IL1β* (FC = 12.09, n = 7, p = 0.0011), *FAM126A* (FC = 2.00, n = 7, p = 0.0011), *BTG2* (FC = 3.05, n = 7, p = 0.0005), *WISP1* (FC = 6.08, n = 7, p = 0.004) (Figure 3F). In contrast, *HLA-C* was specifically downregulated in the nuclear compartment by RNA-seq analysis, but was unchanged using RT-qPCR analysis of unfractionated tissue; *HLA-C* (FC = 1.14, n = 7, p = 0.7103) (Figure 3F).

**Figure 3:**
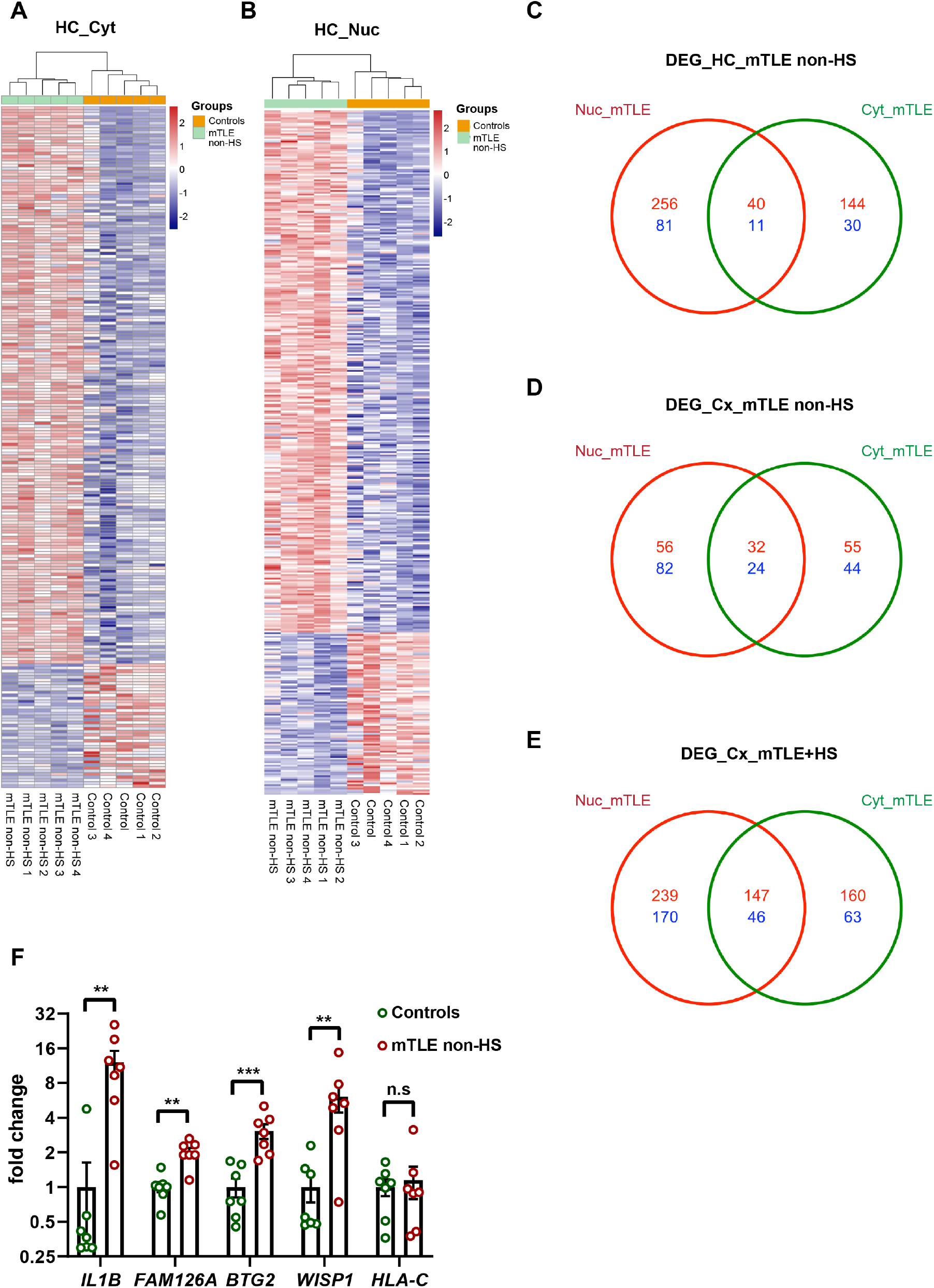
Differentially expressed genes and experimental validation. Heatmaps of differentially expressed mRNAs across the 10 hippocampal samples (five controls and five mTLE non-HS patients) for cytoplasmic (A) and nuclear (B) fractions. Hierarchical clustering is shown at the top. HC: hippocampus, Controls, mTLE non-HS, Cyt: cytoplasm, Nuc: nucleus. Venn diagrams showing overlap of the number of differentially expressed genes (DEGs) between nuclear (Nuc) and cytoplasmic (Cyt) compartments in HC mTLE non-HS (C), Cx mTLE non-HS (D) and Cx mTLE+HS (E). Color code: red indicates upregulated genes and blue indicates downregulated genes. (F) RT-qPCR validation of a selection of DEGs in an independent set of HC samples, comparing expression levels between controls and mTLE non-HS. *GAPDH* was used for normalization. n = 7 mTLE non-HS and 7 control samples. Each colored dot is a sample. Error bars represent standard error of mean. Mann-Whitney tests. ***p < 0.001, **p < 0.01, n.s. non-significant.

In order to identify pathways that are enriched in the cytoplasm and nucleus based on DEGs identified by RNA-seq, reactome pathway enrichment analysis was performed. Pathways enriched in HC mTLE non-HS cytoplasmic samples were related to Neuronal system (Upregulated transcripts, entities = 17 out of 487, p = 5.22e-05, FDR = 0.021), Adrenoceptors (Upregulated transcripts, entities = 3 out of 11, p = 3.01e-04, FDR = 0.061), Transmission across Chemical Synapses (Upregulated transcripts, entities = 12 out of 341, p = 6.35e-04, FDR = 0.065), GABA receptor activation (Upregulated transcripts, entities = 5 out of 67, p = 0.001, FDR = 0.092), Potassium channels (Upregulated transcripts, entities = 6 out of 107, p = 0.002, FDR = 0.092), Activation of AMPA receptor (Upregulated transcripts, entities = 2 out of 7, p = 0.003, FDR = 0.127), Caspase mediated cleavage of cytoskeletal proteins (Downregulated transcripts, entities = 3 out of 12, p = 3.61e-06, FDR = 5.12e-04) and Apoptotic cleavage of cellular proteins (Downregulated transcripts, entities = 3 out of 38, p = 1.10e-04, FDR = 0.005). Pathways derived from nuclear samples included NGF-stimulated transcription (Upregulated transcripts, entities = 8 out of 56, p = 1.93e-05, FDR = 0.016), Signaling by Tyrosine kinase receptors (NTRK1) (Upregulated transcripts, entities = 9 out of 143, p = 0.002, FDR = 0.542), Endosomal/vacuolar pathway (Downregulated transcripts, entities = 14 out of 82, p = 4.11e-15, FDR = 1.27e-12), Antigen presentation (Downregulated transcripts, entities = 14 out of 102, p = 7.76e-14, FDR = 1.20e-11), and Interferon alpha/beta signaling (Downregulated transcripts, entities = 15 out of 186, p = 1.72e-11, FDR = 1.77e-09) (Supplementary file 2). No common pathways were found when comparing HC and Cx mTLE non-HS conditions. Importantly, pathways identified for Cx mTLE+HS in nuclear and cytoplasmic samples were commonly enriched for Interleukin-4 and Interleukin-13 signaling (Nuc: entities = 18 out of 211, p = 1.96e-04, FDR = 0.087. Cyt: entities = 16 out of 211, p = 1.88e-06, FDR = 0.001) hinting at activation of immune response genes which is a pathological signature of mTLE+HS tissue (Supplementary file 3).

## Discussion

Thus far, most profiling studies of resected tissue from mTLE patients have focused on profiling gene expression changes at the level of the entire cell without taking into account subcellular RNA localization and function. Here, we have for the first time performed RNA-seq on purified cell compartments in resected mTLE patient HC and Cx tissue in comparison to postmortem control tissue.

The observed changes in gene expression were specific to tissue type and cell compartment, as underscored by pathway analysis. Here, we focus on pathways altered in the hippocampus, as this is considered the seizure generating region in refractory mTLE (Tatum, 2012). Enrichment analysis using DEGs in HC cytoplasmic samples revealed key pathways such as Neuronal system, Adrenoreceptors, Transmission across Chemical synapses, Gamma-aminobutyric acid (GABA) and Ionotropic AMPA receptor activation, Potassium channel pathways, all linked to genes upregulated in mTLE tissue. Several of these pathways were detected in previous transcriptomics studies performed on mTLE tissue (Dixit et al., 2016; Mirza et al., 2015). We also observed upregulation of genes coding for the AMPA receptor subunits Glutamate ionotropic receptor AMPA type subunit 1 (*GRIA1*) and *GRIA2*, which are strongly associated with pathways altered in mTLE tissue (Pfisterer et al., 2020). AMPA receptor subunit proteins are known to be one of the major driving forces of seizures (Barker-Haliski and White, 2015; Leo et al., 2018). Overall, upregulated genes were mainly associated with synaptic transmission, receptor activation (GABA and AMPA), potassium channels, all of which are biological processes known to be deregulated in refractory epilepsy (Aronica and Gorter, 2007; Dixit et al., 2016; Pfisterer et al., 2020). However, in the nuclear compartment, upregulated genes were associated with pathways related to transcription factors: Nerve growth factor (NGF)-stimulated transcription, Kinase and transcription factor activation, and signaling by Tyrosine kinase receptors (NTRK1 or TRK). The upregulation of genes associated with factors promoting neuronal growth and regulating synaptic strength could be due to the compensatory mechanisms in response to abnormal neural activity (Huang and Reichardt, 2003; Sofroniew et al., 2001). Passive transport by Aquaporins (AQP) was also a key pathway associated with upregulated genes *AQP1, AQP4* and *AQP6* in the nucleus. AQPs are cell membrane proteins that regulate water transport in and out of cells, where AQP4 is a key astrocytic protein that is found to be upregulated in the hippocampus in human mTLE (Lee et al., 2004; TAKATA, 2004). An increase in AQP4 protein levels was shown previously, but AQP1 protein expression was unchanged in the sclerotic mTLE hippocampus in this study (Salman et al., 2017b). The main pathways associated with downregulated genes in the cytoplasm were caspase-mediated cleavage of cytoskeletal proteins, apoptotic cleavage of cellular proteins and apoptosis-related pathways. In contrast to published data, we observed that Vimentin (VIM) and Gelsolin (GSN), genes associated with these pathways, were downregulated in our dataset. VIM and GSN are mainly expressed in astrocytes and function as anti-inflammatory and neuroprotective agents. These proteins were reported to be upregulated at the chronic phase of experimental epilepsy and in human mTLE tissue (Bitsika et al., 2016; Bruxel et al., 2021; do Canto et al., 2021). Even though reactive gliosis is observed in mTLE non-HS HC tissue, VIM and GSN displayed lower expression in our dataset (Blümcke et al., 2013). Activation of caspase and apoptotic pathways has been reported in studies using mTLE+HS tissue that displays severe cell loss (Aronica and Gorter, 2007; Henshall and Simon, 2005; Teocchi and D’Souza-Li, 2016). The key pathways associated with genes downregulated in the nucleus were: Endosomal and vacuolar pathway, Interferon signaling and GABA-B receptor activation. Endosomal-vacuolar (or endolysosomal) pathway activity is known to be required for protecting cells from neurotoxicity, and reduced function of this pathway could be detrimental for neurons (Winckler et al., 2018).

Frequently, genes encoding members of the same gene family, but for different subunits, were observed to display opposite expression trends in individual compartments. For example, genes associated with GABA-A receptor subunits (*GABRA3, GABRA5, GABRQ*) were upregulated in the cytoplasm. GABA-A receptors are fast-acting ligand gated chloride ion channels that mediate membrane depolarization and provide fast inhibitory action by inhibiting neurotransmitter release (Michels and Moss, 2007). Interestingly, genes associated with GABA-B receptor subunits (*ADCY1, GABBR2, KCNJ4*) were downregulated in the nucleus (Pinard et al., 2010). GABA-B receptors are slow acting metabotropic G-protein coupled receptors that mediate cell membrane hyperpolarization and lead to inhibitory action via second messengers and other channels. Previous studies show that GABA receptor subunit transcript or protein expression is mainly downregulated in mTLE tissue (Bruxel et al., 2021). GABA-A receptor binding is known to mediate the early phase of inhibition due to its fast-acting property, whereas GABA-B binding mediates the late phase (Treiman, 2001). Reduced nuclear expression of GABA-B subunits could delay the late inhibitory phase. GABA receptors are the most common drug targets in epilepsy and changes in their expression are considered to be one of the causes of drug resistance observed in refractory mTLE (Rogawski and Löscher, 2004). Our study sheds light on their compartment-specific expression regulation in refractory mTLE, which has not been addressed previously. Our dataset constitutes a valuable resource for studies on the subcellular localization of RNA in hippocampal and cortical tissue from healthy control and drug refractory mTLE patients. Future experimental studies are required to understand how changes in compartment-specific mRNA localization and function contribute towards the development of pharmacoresistant mTLE.

## Supporting information

Other supplemental files_Reactome

## Data Availability

The datasets generated for this study are deposited in the NCBI Gene Expression Omnibus (GEO) repository with reference number GSEXXXX (provided upon request to the authors).

## ETHICS AND PATIENT CONSENT STATEMENT

An informed consent was obtained from all patients at the University Medical Center Utrecht for use of tissue and clinical information for research purposes. Pharmacoresistant mesial temporal lobe epilepsy patient hippocampus and neocortical tissue was obtained after surgery at the University Medical Center Utrecht, Utrecht, The Netherlands. All procedures and use of tissue and clinical information for research purposes were approved by the Institutional Review board of University Medical Center Utrecht (Utrecht, The Netherlands) for this study. Postmortem human hippocampus and neocortical tissue was obtained from the Netherlands Brain Bank (Amsterdam, The Netherlands). A written informed consent was obtained from all donors for brain autopsy and for use of the material and clinical information for research purposes. Use of postmortem tissue and clinical information for research purposes was approved by the medical ethics board of the Amsterdam University Medical Center (Amsterdam, The Netherlands).

## DATA AVAILABILITY STATEMENT

The datasets generated for this study are deposited in the NCBI Gene Expression Omnibus (GEO) repository with reference number GSEXXXX (provided upon request).

## AUTHOR CONTRIBUTIONS

V.R.V: investigation, formal analysis, visualization, writing-original draft. G.G.: methodology, investigation, formal analysis, visualization. M.d.W.: formal analysis, visualization. M.T.V.: investigation, formal analysis, visualization. N.P.: formal analysis, visualization, writing-review & editing. A.G-D.: investigation, formal analysis, visualization, writing-review & editing. P.C.v.R., P.H.G., P.v.E.: epilepsy surgery material. J.K.: resources. P.N.E.d.G.: conceptualization, resources. R.J.P.: conceptualization, resources, writing-review & editing, supervision, funding acquisition.

## ACKNOWLEDGMENTS

We thank all the Pasterkamp lab members for their valuable input throughout the project.

---

**Supplementary Figure S1:**
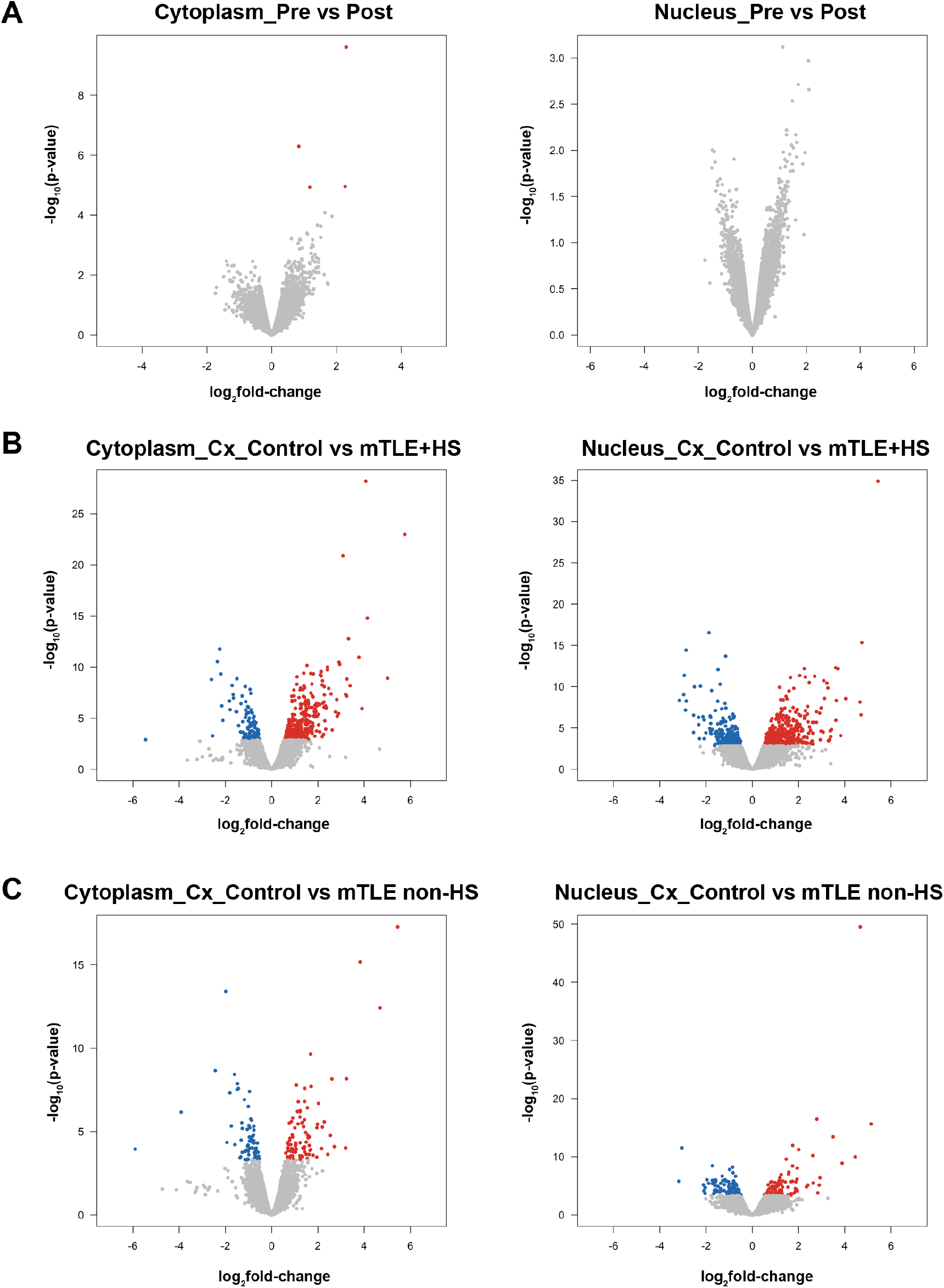
(A) Volcano plots showing overall gene expression changes observed between Pre- *versus* Postmortem-treated nuclear and cytoplasmic samples. (B) Volcano plots for Cx Control *versus* mTLE+HS Cytoplasm and Nucleus. (C) Volcano plots for Cx Controls *versus* mTLE non-HS Cytoplasm and nucleus. Blue colored dots indicate significantly downregulated genes and the red colored dots indicate significantly upregulated genes.

## Supplementary file 1

- Supplementary Table S1: Patient and postmortem control tissue details used for RNA-seq. AED, Anti-epileptic drugs used.
- Supplementary Table S2: Different parameters of the RNA-seq analysis quality control.
- Supplementary Table S3: Primers sequences used in the study.

## Supplementary file 2

Reactome pathway results of HC mTLE non-HS. up = upregulated genes compared to controls, down = downregulated genes compared to controls. CYT = cytoplasm, NUC = nucleus.

- Table 1: mTLE non-HS_CYT_up_result
- Table 2: mTLE non-HS_NUC_up_result
- Table 3: mTLE non-HS_CYT_down_result
- Table 4: mTLE non-HS_NUC_down_result

## Supplementary file 3

Reactome pathway results of Cx mTLE+HS and mTLE non-HS. up = upregulated genes compared to controls, down = downregulated genes compared to controls. CYT = cytoplasm, NUC = nucleus.

- Table 1: mTLE non-HS_CYT_updown_result
- Table 2: mTLE non-HS_NUC_updown_result
- Table 3: mTLE+HS_CYT_updown_result
- Table 4: mTLE+HS_NUC_updown_result

## Other supplemental files

HC and Cx Reactome pathway reports and figures.

- HC: mTLE non-HS_Cytoplasm (Upregulated & downregulated DEGs were analyzed separately) and Nucleus (Upregulated & downregulated DEGs were analyzed separately).
- Cx: mTLE non-HS_Cytoplasm (Combined analysis for Upregulated & downregulated DEGs) and Nucleus (Combined analysis for Upregulated & downregulated DEGs).
- Cx: mTLE+HS_Cytoplasm (Combined analysis for Upregulated & downregulated DEGs) and Nucleus (Combined analysis for Upregulated & downregulated DEGs).

