## Supplementary material for "Compartment-specific total RNA profile of Hippocampal and Cortical cells from Mesial Temporal Lobe Epilepsy tissue": Other supplemental files_Reactome: Cx_mTLE non-HS_Cyt_updownspecific_report.pdf

### Pathway Analysis Report

This report contains the pathway analysis results for the submitted sample ''. Analysis was performed against Reactome version 77 on 13/09/2021. The web link to these results is:

https://reactome.org/PathwayBrowser/#/ANALYSIS=MjAyMTA5MTMxMDIxNThfNTE4Mzc%3D

Please keep in mind that analysis results are temporarily stored on our server. The storage period depends on usage of the service but is at least 7 days. As a result, please note that this URL is only valid for a limited time period and it might have expired.

### Table of Contents

1. Introduction
2. Properties
3. Genome-wide overview
4. Most significant pathways
5. Pathways details
6. Identifiers found
7. Identifiers not found

### 1. Introduction

Reactome is a curated database of pathways and reactions in human biology. Reactions can be considered as pathway 'steps'. Reactome defines a 'reaction' as any event in biology that changes the state of a biological molecule. Binding, activation, translocation, degradation and classical biochemical events involving a catalyst are all reactions. Information in the database is authored by expert biologists, entered and maintained by Reactome’s team of curators and editorial staff. Reactome content frequently cross-references other resources e.g. NCBI, Ensembl, UniProt, KEGG (Gene and Compound), ChEBI, PubMed and GO. Orthologous reactions inferred from annotation for Homo sapiens are available for 17 non-human species including mouse, rat, chicken, puffer fish, worm, fly, yeast, rice, and Arabidopsis. Pathways are represented by simple diagrams following an SBGN-like format.

Reactome's annotated data describe reactions possible if all annotated proteins and small molecules were present and active simultaneously in a cell. By overlaying an experimental dataset on these annotations, a user can perform a pathway over-representation analysis. By overlaying quantitative expression data or time series, a user can visualize the extent of change in affected pathways and its progression. A binomial test is used to calculate the probability shown for each result, and the p-values are corrected for the multiple testing (Benjamini–Hochberg procedure) that arises from evaluating the submitted list of identifiers against every pathway.

To learn more about our Pathway Analysis, please have a look at our relevant publications:


Fabregat A, Sidiropoulos K, Garapati P, Gillespie M, Hausmann K, Haw R, … D’Eustachio P (2016). The reactome pathway knowledgebase. Nucleic Acids Research, 44(D1), D481–D487. https://doi.org/10.1093/nar/gkv1351.

Fabregat A, Sidiropoulos K, Viteri G, Forner O, Marin-Garcia P, Arnau V, … Hermjakob H (2017). Reactome pathway analysis: a high-performance in-memory approach. BMC Bioinformatics, 18.

### 2. Properties

- This is an **overrepresentation** analysis: A statistical (hypergeometric distribution) test that determines whether certain Reactome pathways are over-represented (enriched) in the submitted data. It answers the question ‘Does my list contain more proteins for pathway X than would be expected by chance?’ This test produces a probability score, which is corrected for false discovery rate using the Benjamani-Hochberg method.
- 63 out of 99 identifiers in the sample were found in Reactome, where 332 pathways were hit by at least one of them.
- All non-human identifiers have been converted to their human equivalent.
- This report is filtered to show only results for species 'Homo sapiens' and resource 'all resources'.
- The unique ID for this analysis (token) is MjAyMTA5MTMxMDIxNThfNTE4Mzc%3D. This ID is valid for at least 7 days in Reactome’s server. Use it to access Reactome services with your data.

### 3. Genome-wide overview

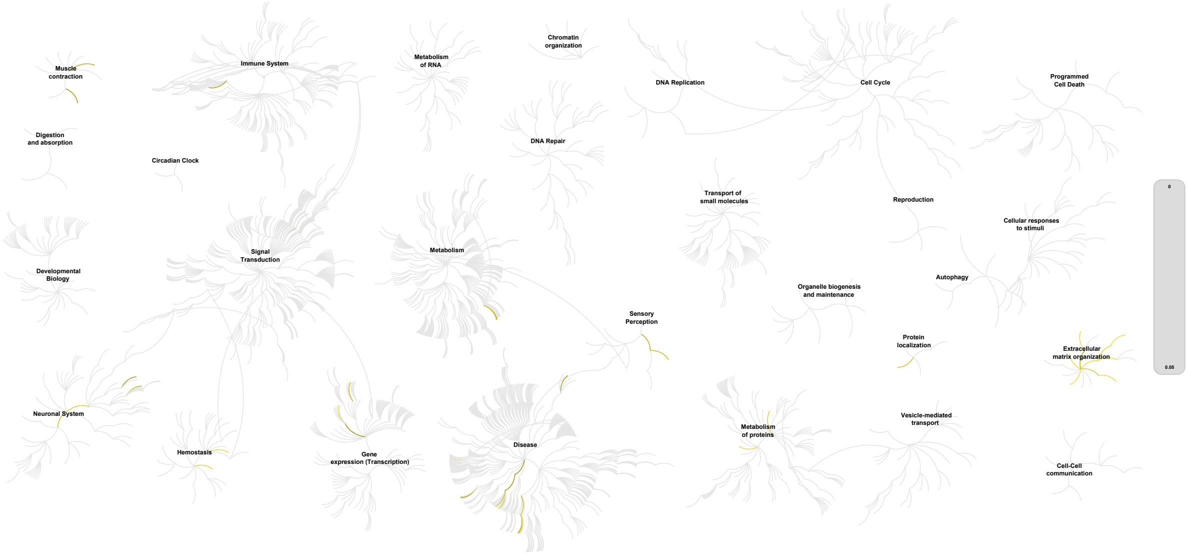


This figure shows a genome-wide overview of the results of your pathway analysis. Reactome pathways are arranged in a hierarchy. The center of each of the circular "bursts" is the root of one top-level pathway, for example "DNA Repair". Each step away from the center represents the next level lower in the pathway hierarchy. The color code denotes over-representation of that pathway in your input dataset. Light grey signifies pathways which are not significantly over-represented.

### 4. Most significant pathways

The following table shows the 25 most relevant pathways sorted by p-value.

| Pathway name | Entities | | | | Reactions | |
|---|---|---|---|---|---|---|
| | found | ratio | p-value | FDR* | found | ratio |
| Integrin cell surface interactions | 6 / 86 | 0.006 | 6.30e-05 | 0.022 | 8 / 55 | 0.004 |
| Assembly of collagen fibrils and other multimeric structures | 5 / 67 | 0.005 | 1.93e-04 | 0.033 | 18 / 26 | 0.002 |
| Collagen chain trimerization | 4 / 44 | 0.003 | 4.15e-04 | 0.047 | 4 / 28 | 0.002 |
| Collagen formation | 5 / 104 | 0.007 | 0.001 | 0.065 | 42 / 77 | 0.006 |
| Toxicity of botulinum toxin type G (botG) | 2 / 7 | 4.81e-04 | 0.001 | 0.065 | 2 / 5 | 3.70e-04 |
| Syndecan interactions | 3 / 29 | 0.002 | 0.002 | 0.065 | 2 / 15 | 0.001 |
| Collagen degradation | 4 / 69 | 0.005 | 0.002 | 0.065 | 12 / 34 | 0.003 |
| Defective B4GALT1 causes B4GALT1-CDG (CDG-2d) | 2 / 9 | 6.19e-04 | 0.002 | 0.065 | 3 / 3 | 2.22e-04 |
| Defective CHST6 causes MCDC1 | 2 / 9 | 6.19e-04 | 0.002 | 0.065 | 1 / 1 | 7.40e-05 |
| Defective ST3GAL3 causes MCT12 and EIEE15 | 2 / 9 | 6.19e-04 | 0.002 | 0.065 | 1 / 1 | 7.40e-05 |
| Toxicity of botulinum toxin type F (botF) | 2 / 9 | 6.19e-04 | 0.002 | 0.065 | 2 / 5 | 3.70e-04 |
| Toxicity of botulinum toxin type D (botD) | 2 / 9 | 6.19e-04 | 0.002 | 0.065 | 2 / 5 | 3.70e-04 |
| Collagen biosynthesis and modifying enzymes | 4 / 76 | 0.005 | 0.003 | 0.077 | 24 / 51 | 0.004 |
| Regulation of Insulin-like Growth Factor (IGF) transport and uptake by Insulin-like Growth Factor Binding Proteins (IGFBPs) | 5 / 127 | 0.009 | 0.003 | 0.077 | 14 / 14 | 0.001 |
| ECM proteoglycans | 4 / 79 | 0.005 | 0.004 | 0.077 | 9 / 23 | 0.002 |
| GP1b-IX-V activation signalling | 2 / 12 | 8.25e-04 | 0.004 | 0.085 | 5 / 7 | 5.18e-04 |
| MECP2 regulates transcription of neuronal ligands | 2 / 13 | 8.94e-04 | 0.005 | 0.095 | 2 / 8 | 5.92e-04 |
| Platelet Adhesion to exposed collagen | 2 / 16 | 0.001 | 0.007 | 0.134 | 4 / 6 | 4.44e-04 |
| Neurotransmitter receptors and postsynaptic signal transmission | 6 / 231 | 0.016 | 0.01 | 0.173 | 28 / 109 | 0.008 |
| Post-translational protein phosphorylation | 4 / 109 | 0.007 | 0.011 | 0.181 | 1 / 1 | 7.40e-05 |
| Non-integrin membrane-ECM interactions | 3 / 61 | 0.004 | 0.012 | 0.194 | 4 / 22 | 0.002 |
| Keratan sulfate degradation | 2 / 22 | 0.002 | 0.013 | 0.194 | 2 / 7 | 5.18e-04 |
| Extracellular matrix organization | 7 / 329 | 0.023 | 0.015 | 0.205 | 77 / 319 | 0.024 |

| Pathway name | Entities | | | | Reactions | |
|---|---|---|---|---|---|---|
| | found | ratio | p-value | FDR* | found | ratio |
| Defective F8 binding to von Willebrand factor | 1 / 2 | 1.38e-04 | 0.015 | 0.213 | 1 / 1 | 7.40e-05 |
| Insertion of tail-anchored proteins into the endoplasmic reticulum membrane | 2 / 25 | 0.002 | 0.016 | 0.213 | 7 / 7 | 5.18e-04 |

* False Discovery Rate

### 5. Pathways details

For every pathway of the most significant pathways, we present its diagram, as well as a short summary, its bibliography and the list of inputs found in it.

#### 1. Integrin cell surface interactions (R-HSA-216083)

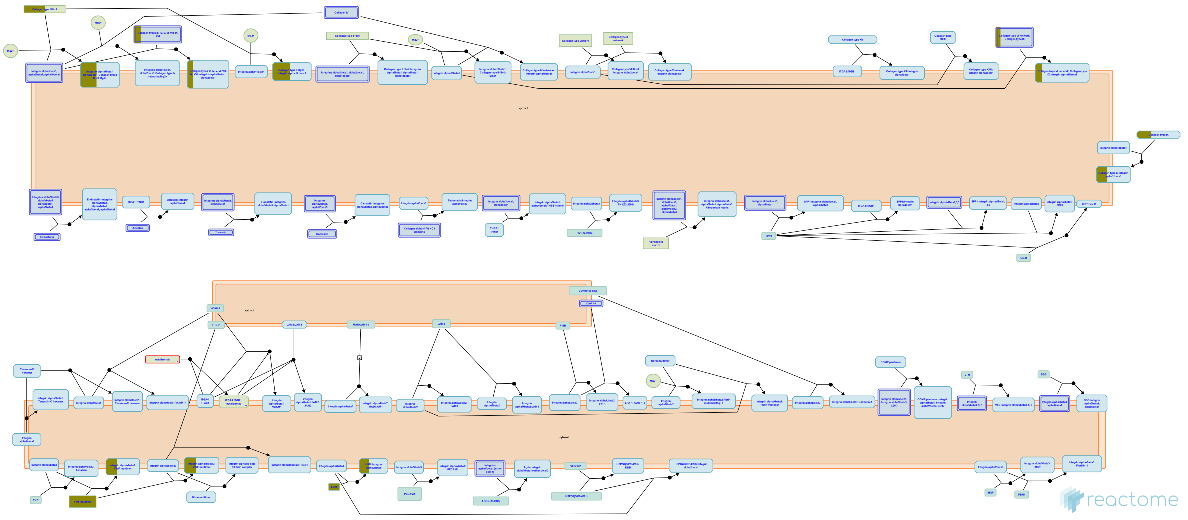

The extracellular matrix (ECM) is a network of macro-molecules that underlies all epithelia and endothelia and that surrounds all connective tissue cells. This matrix provides the mechanical strength and also influences the behavior and differentiation state of cells in contact with it. The ECM are diverse in composition, but they generally comprise a mixture of fibrillar proteins, polysaccharides synthesized, secreted and organized by neighboring cells. Collagens, fibronectin, and laminins are the principal components involved in cell matrix interactions; other components, such as vitronectin, thrombospondin, and osteopontin, although less abundant, are also important adhesive molecules.

Integrins are the receptors that mediate cell adhesion to ECM. Integrins consists of one alpha and one beta subunit forming a noncovalently bound heterodimer. 18 alpha and 8 beta subunits have been identified in humans that combine to form 24 different receptors.

The integrin dimers can be broadly divided into three families consisting of the beta1, beta2/beta7, and beta3/alphaV integrins. beta1 associates with 12 alpha-subunits and can be further divided into RGD-, collagen-, or laminin binding and the related alpha4/alpha9 integrins that recognise both matrix and vascular ligands. beta2/beta7 integrins are restricted to leukocytes and mediate cell-cell rather than cell-matrix interactions, although some recognize fibrinogen. The beta3/alphaV family members are all RGD receptors and comprise aIIbb3, an important receptor on platelets, and the remaining b-subunits, which all associate with alphaV. It is the collagen receptors and leukocyte-specific integrins that contain alpha A-domains.

#### Edit history

| Date | Action | Author |
|---|---|---|
| 2008-03-11 | Edited | Garapati P V |
| 2008-03-11 | Created | Garapati P V |
| 2008-05-07 | Reviewed | Hynes R, Humphries MJ, Yamada KM |
| 2008-05-07 | Authored | Geiger B, Horwitz AR |
| 2012-08-08 | Authored | Jupe S |
| 2013-08-13 | Edited | Jupe S |
| 2013-08-13 | Reviewed | Ricard-Blum S |
| 2021-05-22 | Modified | Shorser S |

#### Entities found in this pathway (5)

| Input | UniProt Id | Input | UniProt Id | Input | UniProt Id |
|---|---|---|---|---|---|
| COL1A2 | P08123 | COL3A1 | P02461 | COL8A2 | P25067, Q14050 |
| LUM | P51884 | VWF | P04275 | | |

#### 2. Assembly of collagen fibrils and other multimeric structures (R-HSA-2022090)

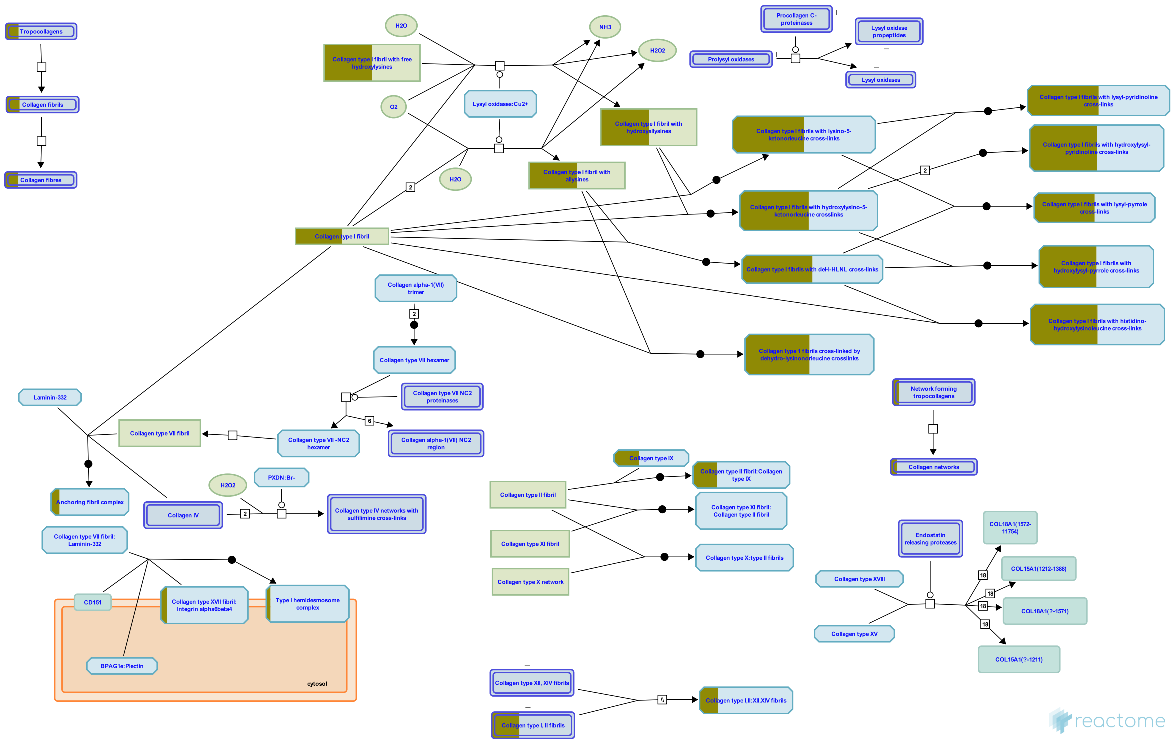


Collagen trimers in triple-helical form, referred to as procollagen or collagen molecules, are exported from the ER and trafficked through the Golgi network before secretion into the extracellular space. For fibrillar collagens namely types I, II, III, V, XI, XXIV and XXVII (Gordon & Hahn 2010, Ricard-Blum 2011) secretion is concomitant with processing of the N and C terminal collagen propeptides. These processed molecules are known as tropocollagens, considered to be the units of higher order collagen structures. They form within the extracellular space via a process that can proceed spontaneously, but in the cellular environment is regulated by many collagen binding proteins such as the FACIT (Fibril Associated Collagens with Interrupted Triple helices) family collagens and Small Leucine-Rich Proteoglycans (SLRPs). The architecture formed ultimately depends on the collagen subtype and the cellular conditions. Structures include the well-known fibrils and fibres formed by the major structural collagens type I and II plus several different types of supramolecular assembly (Bruckner 2010). The mechanical and physical properties of tissues depend on the spatial arrangement and composition of these collagen-containing structures (Kadler et al. 1996, Shoulders & Raines 2009, Birk & Bruckner 2011).

Fibrillar collagen structures are frequently heterotypic, composed of a major collagen type in association with smaller amounts of other types, e.g. type I collagen fibrils are associated with types III and V, while type II fibrils frequently contain types IX and XI (Wess 2005). Fibres composed exclusively of a single collagen type probably do not exist, as type I and II fibrils require collagens V and XI respectively as nucleators (Kadler et al. 2008, Wenstrup et al. 2011). Much of the structural understanding of collagen fibrils has been obtained with fibril-forming collagens, particularly type I, but some central features are believed to apply to at least the other fibrillar collagen subtypes (Wess 2005). Fibril diameter and length varies considerably, depending on the tissue and collagen types (Fang et al. 2012). The reasons for this are poorly understood (Wess 2005).

Some tissues such as skin have fibres that are approximately the same diameter while others such as tendon or cartilage have a bimodal distribution of thick and thin fibrils. Mature type I collagen fibrils in tendon are up to 1 cm in length, with a diameter of approx. 500 nm. An individual fibrillar collagen triple helix is less than 1.5 nm in diameter and around 300 nm long; collagen molecules must assemble to give rise to the higher-order fibril structure, a process known as fibrillogenesis, prevented by the presence of C-terminal propeptides (Kadler et al. 1987). In electron micrographs, fibrils have a banded appearance, due to regular gaps where fewer collagen molecules overlap, which occur because the fibrils are aligned in a quarter-stagger arrangement (Hodge & Petruska 1963). Collagen microfibrils are believed to have a quasi-hexagonal unit cell, with tropocollagen arranged to form supertwisted, right-handed microfibrils that interdigitate with neighbouring microfibrils, leading to a spiral-like structure for the mature collagen fibril (Orgel et al. 2006, Holmes & Kadler 2006).

Neighbouring tropocollagen monomers interact with each other and are cross-linked covalently by lysyl oxidase (Orgel et al. 2000, Maki 2006). Mature collagen fibrils are stabilized by lysyl oxidase-mediated cross-links. Hydroxylysyl pyridinoline and lysyl pyridinoline cross-links form between (hydroxy) lysine and hydroxylysine residues in bone and cartilage (Eyre et al. 1984). Arginoline cross-links can form in cartilage (Eyre et al. 2010); mature bovine articular cartilage contains roughly equimolar amounts of arginoline and hydroxylysyl pyridinoline based on peptide yields. Mature collagen fibrils in skin are stabilized by the lysyl oxidase-mediated cross-link histidino-hydroxylysinonorleucine (Yamauch et al. 1987). Due to the quarter-staggered arrangement of collagen molecules in a fibril, telopeptides most often interact with the triple helix of a neighbouring collagen molecule in the fibril, except for collagen molecules in register staggered by 4D from another collagen molecule. Fibril aggregation in vitro can be unipolar or bipolar, influenced by temperature and levels of C-proteinase, suggesting a role for the N- and C- propeptides in regulation of the aggregation process (Kadler et al. 1996). In vivo, collagen molecules at the fibril surface may retain their N-propeptides, suggesting that this may limit further accretion, or alternatively represents a transient stage in a model whereby fibrils grow in diameter through a cycle of deposition, cleavage and further deposition (Chapman 1989).

In vivo, fibrils are often composed from more than one type of collagen. Type III collagen is found associated with type I collagen in dermal fibrils, with the collagen III on the periphery, suggesting a regulatory role (Fleischmajer et al. 1990). Type V collagen associates with type I collagen fibrils, where it may limit fibril diameter (Birk et al. 1990, White et al. 1997). Type IX associates with the surface of narrow diameter collagen II fibrils in cartilage and the cornea (Wu et al. 1992, Eyre et al. 2004). Highly specific patterns of crosslinking sites suggest that collagen IX functions in interfibrillar networking (Wess 2005). Type XII and XIV collagens are localized near the surface of banded collagen I fibrils (Nishiyama et al. 1994). Certain fibril-associated collagens with interrupted triple helices (FACITs) associate with the surface of collagen fibrils, where they may serve to limit fibril fusion and thereby regulate fibril diameter (Gordon & Hahn 2010). Collagen XV, a member of the multiplexin family, is almost exclusively associated with the fibrillar collagen network, in very close proximity to the basement membrane. In human tissues collagen XV is seen linking banded collagen fibers subjacent to the basement membrane (Amenta et al. 2005). Type XIV collagen, SLRPs and discoidin domain receptors also regulate fibrillogenesis (Ansorge et al. 2009, Kalamajski et al. 2010, Flynn et al. 2010).

Collagen IX is cross-linked to the surface of collagen type II fibrils (Eyre et al. 1987). Type XII and XIV collagens are found in association with type I (Walchli et al. 1994) and type II (Watt et al. 1992, Eyre 2002) fibrils in cartilage. They are thought to associate non-covalently via their COL1/NC1 domains (Watt et al. 1992, Eyre 2002).

Some non-fibrillar collagens form supramolecular assemblies that are distinct from typical fibrils. Collagen VII forms anchoring fibrils, composed of antiparallel dimers that connect the dermis to the epidermis (Bruckner-Tuderman 2009). During fibrillogenesis, the nascent type VII procollagen molecules dimerize in an antiparallel manner. The C-propeptides are then removed by Bone morphogenetic protein 1 (Rattenholl et al. 2002) and the processed antiparallel dimers aggregate laterally. Collagens VIII and X form hexagonal networks and collagen VI forms beaded filament (Gordon & Hahn 2010, Ricard-Blum et al. 2011).

#### Edit history

| Date | Action | Author |
|---|---|---|
| 2011-08-05 | Authored | Jupe S |
| 2011-11-25 | Created | Jupe S |
| 2012-10-08 | Reviewed | Kalamajski S, Raleigh S |
| 2012-11-12 | Edited | Jupe S |
| 2012-11-19 | Reviewed | Ricard-Blum S |
| 2021-05-22 | Modified | Shorser S |

#### Entities found in this pathway (4)

| Input | UniProt Id | Input | UniProt Id |
|---|---|---|---|
| COL1A2 | P08123 | COL3A1 | P02461 |
| COL8A2 | P25067, Q14050 | ITGB4 | P16144 |

#### 3. Collagen chain trimerization (R-HSA-8948216)

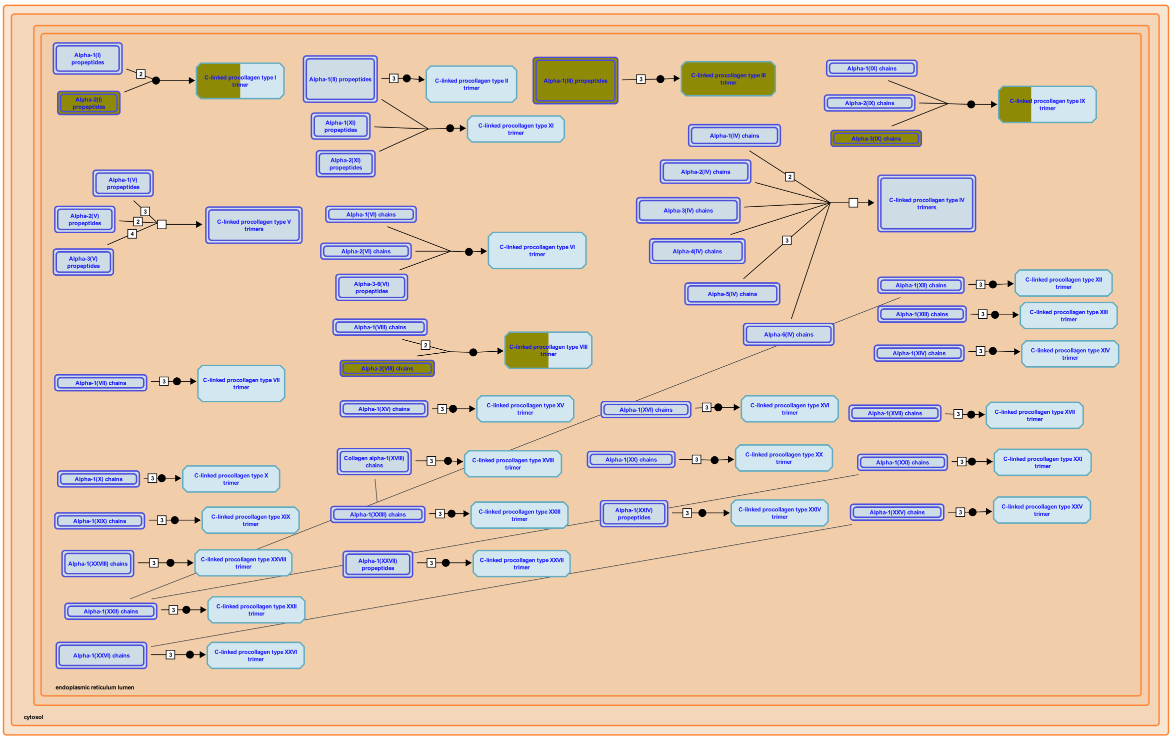

The C-propeptides of collagen propeptide chains are essential for the association of three peptide chains into a trimeric but  non-helical procollagen. This initial binding event determines the composition of the trimer, brings the individual chains into the correct register and initiates formation of the triple helix at the C-terminus, which then proceeds towards the N-terminus in a zipper-like fashion (Engel & Prockop 1991). Most early refolding studies were performed with collagen type III, which contains a disulfide linkage at the C-terminus of its triple helix (Bächinger et al. 1978, Bruckner et al. 1978) that acts as a permanent linker even after removal of the non-collagenous domains.

Mutations within the C-propeptides further suggest that they are crucial for the correct interaction of the three polypeptide chains and for subsequent correct folding (refs. in Boudko et al. 2011).


##### Edit history

| Date | Action | Author |
|---|---|---|
| 2012-04-11 | Authored | Jupe S |
| 2012-05-24 | Reviewed | Canty-Laird EG |
| 2016-11-03 | Edited | Jupe S |

| Date | Action | Author |
|---|---|---|
| 2016-11-11 | Created | Jupe S |
| 2021-05-22 | Modified | Shorser S |

#### Entities found in this pathway (3)

| Input | UniProt Id | Input | UniProt Id | Input | UniProt Id |
|---|---|---|---|---|---|
| COL1A2 | P08123 | COL3A1 | P02461 | COL8A2 | P25067, Q14050 |

### 4. Collagen formation (R-HSA-1474290)

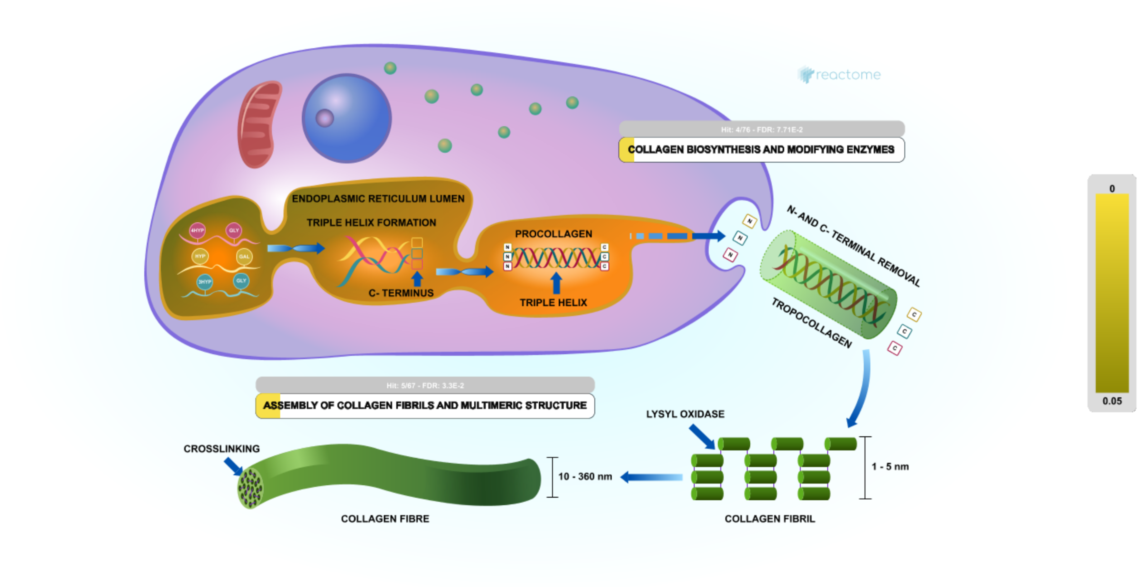

Collagen is a family of at least 29 structural proteins derived from over 40 human genes (Myllyharju & Kivirikko 2004). It is the main component of connective tissue, and the most abundant protein in mammals making up about 25% to 35% of whole-body protein content. A defining feature of collagens is the formation of trimeric left-handed polyproline II-type helical collagenous regions. The packing within these regions is made possible by the presence of the smallest amino acid, glycine, at every third residue, resulting in a repeating motif Gly-X-Y where X is often proline (Pro) and Y often 4-hydroxyproline (4Hyp). Gly-Pro-Hyp is the most common triplet in collagen (Ramshaw et al. 1998). Collagen peptide chains also have non-collagenous domains, with collagen subclasses having common chain structures. Collagen fibrils are mostly found in fibrous tissues such as tendon, ligament and skin. Other forms of collagen are abundant in cornea, cartilage, bone, blood vessels, the gut, and intervertebral disc. In muscle tissue, collagen is a major component of the endomysium, constituting up to 6% of muscle mass. Gelatin, used in food and industry, is collagen that has been irreversibly hydrolyzed.

On the basis of their fibre architecture in tissues, the genetically distinct collagens have been divided into subgroups. Group 1 collagens have uninterrupted triple-helical domains of about 300 nm, forming large extracellular fibrils. They are referred to as the fibril-forming collagens, consisting of collagens types I, II, III, V, XI, XXIV and XXVII. Group 2 collagens are types IV and VII, which have extended triple helices (>350 nm) with imperfections in the Gly-X-Y repeat sequences. Group 3 are the short-chain collagens. These have two subgroups. Group 3A have continuous triple-helical domains (type VI, VIII and X). Group 3B have interrupted triple-helical domains, referred to as the fibril-associated collagens with interrupted triple helices (FACIT collagens, Shaw & Olsen 1991). FACITs include collagen IX, XII, XIV, XVI, XIX, XX, XXI, XXII and XXVI plus the transmembrane collagens (XIII, XVII, XXIII and XXV) and the multiple triple helix domains and interruptions (Multiplexin) collagens XV and XVIII (Myllyharju & Kivirikko 2004). The non-collagenous domains of collagens have regulatory functions; several are biologically active when cleaved from the main peptide chain. Fibrillar collagen peptides all have a large triple helical domain (COL1) bordered by N and C terminal extensions, called the N- and C-propeptides, which are cleaved prior to formation of the collagen fibril. The intact form is referred to as a collagen propeptide, not procollagen, which is used to refer to the trimeric triple-helical precursor of collagen before the propeptides are removed. The C-propeptide, also called the NC1 domain, directs chain association during assembly of the procollagen molecule from its three constituent alpha chains (Hulmes 2002).

Fibril forming collagens are the most familiar and best studied subgroup. Collagen fibres are aggregates or bundles of collagen fibrils, which are themselves polymers of tropocollagen complexes, each consisting of three polypeptide chains known as alpha chains. Tropocollagens are considered the subunit of larger collagen structures. They are approximately 300 nm long and 1.5 nm in diameter, with a left-handed triple-helical structure, which becomes twisted into a right-handed coiled-coil 'super helix' in the collagen fibril. Tropocollagens in the extracellular space polymerize spontaneously with regularly staggered ends (Hulmes 2002). In fibrillar collagens the molecules are staggered by about 67 nm, a unit known as D that changes depending upon the hydration state. Each D-period contains slightly more than four collagen molecules so that every D-period repeat of the microfibril has a region containing five molecules in cross-section, called the 'overlap', and a region containing only four molecules, called the 'gap'. The triple-helices are arranged in a hexagonal or quasi-hexagonal array in cross-section, in both the gap and overlap regions (Orgel et al. 2006). Collagen molecules cross-link covalently to each other via lysine and hydroxylysine side chains. These cross-links are unusual, occuring only in collagen and elastin, a related protein.

The macromolecular structures of collagen are diverse. Several group 3 collagens associate with larger collagen fibers, serving as molecular bridges which stabilize the organization of the extracellular matrix. Type IV collagen is arranged in an interlacing network within the dermal-epidermal junction and vascular basement membranes. Type VI collagen forms distinct microfibrils called beaded filaments. Type VII collagen forms anchoring fibrils. Type VIII and X collagens form hexagonal networks. Type XVII collagen is a component of hemidesmosomes where it is complexed wtih alpha6Beta4 integrin, plectin, and laminin-332 (de Pereda et al. 2009). Type XXIX collagen has been recently reported to be a putative epidermal collagen with highest expression in suprabasal layers (Soderhall et al. 2007). Collagen fibrils/aggregates arranged in varying combinations and concentrations in different tissues provide specific tissue properties. In bone, collagen triple helices lie in a parallel, staggered array with 40 nm gaps between the ends of the tropocollagen subunits, which probably serve as nucleation sites for the deposition of crystals of the mineral component, hydroxyapatite (Ca10(PO4)6(OH)2) with some phosphate. Collagen structure affects cell-cell and cell-matrix communication, tissue construction in growth and repair, and is changed in development and disease (Sweeney et al. 2006, Twardowski et al. 2007). A single collagen fibril can be heterogeneous along its axis, with significantly different mechanical properties in the gap and overlap regions, correlating with the different molecular organizations in these regions (Minary-Jolandan & Yu 2009).


#### Edit history

| Date | Action | Author |
|---|---|---|
| 2011-08-05 | Authored | Jupe S |
| 2011-08-05 | Created | Jupe S |
| 2012-04-11 | Edited | Jupe S |
| 2012-05-24 | Reviewed | Canty-Laird EG |
| 2021-05-22 | Modified | Shorser S |

#### Entities found in this pathway (4)

| Input | UniProt Id | Input | UniProt Id |
|---|---|---|---|
| COL1A2 | P08123 | COL3A1 | P02461 |
| COL8A2 | P25067, Q14050 | ITGB4 | P16144 |

#### 5. Toxicity of botulinum toxin type G (botG) (R-HSA-5250989)

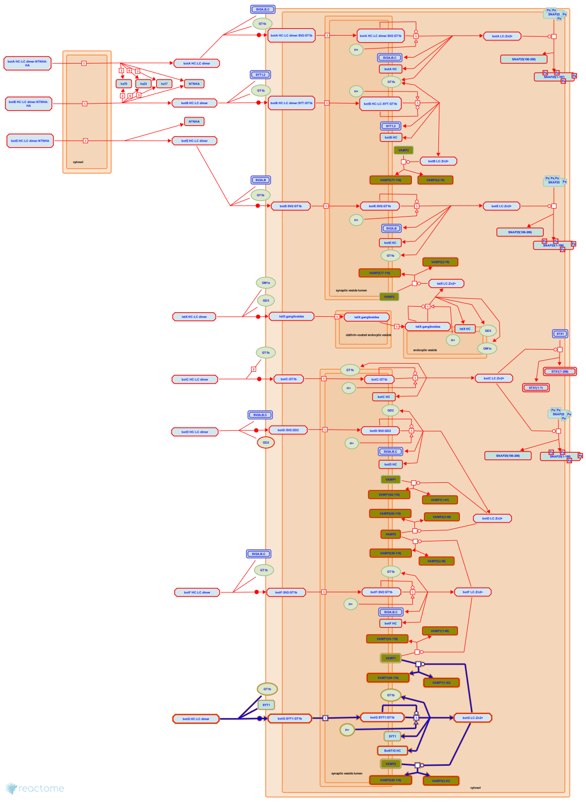

**Diseases:** botulism.

Botulinum toxin type G (botG) is rarely if ever associated with human disease (Hatheway 1995) and a pathway by which it might enter the circulation from the human gut has not been described. Nevertheless, the toxin itself, a disulfide-bonded heavy chain (HC) - light chain (LC) heterodimer ("dichain"), is capable of binding to neurons by interactions with cell-surface ganglioside and syntagmin 1 (SYT1) (Peng et al. 2012; Willjes et al. 2013), the bound toxin can enter synaptic vesicles and release its LC moiety into the cytosol of targeted cells (Montal 2010), and the botG LC can cleave vesicle-associated membrane proteins 1 and 2 (VAMP1 and 2) on the cytosolic face of the synaptic vesicle membrane (Schiavo et al. 1994; Yamasaki et al. 1994). These four events are annotated here.

#### Edit history

| Date | Action | Author |
|---|---|---|
| 2006-06-15 | Authored | Krupa S, Gopinathrao G |
| 2007-08-03 | Reviewed | Ichtchenko K |
| 2014-02-01 | Created | D'Eustachio P |
| 2014-02-11 | Revised | D'Eustachio P |
| 2014-02-11 | Edited | D'Eustachio P |
| 2014-11-18 | Reviewed | Sharma S, Thirunavukkarasu N |
| 2020-10-08 | Modified | D'Eustachio P |

#### Entities found in this pathway (1)

| Input | UniProt Id |
|---|---|
| VAMP1 | P23763, P63027 |

#### 6. Syndecan interactions (R-HSA-3000170)

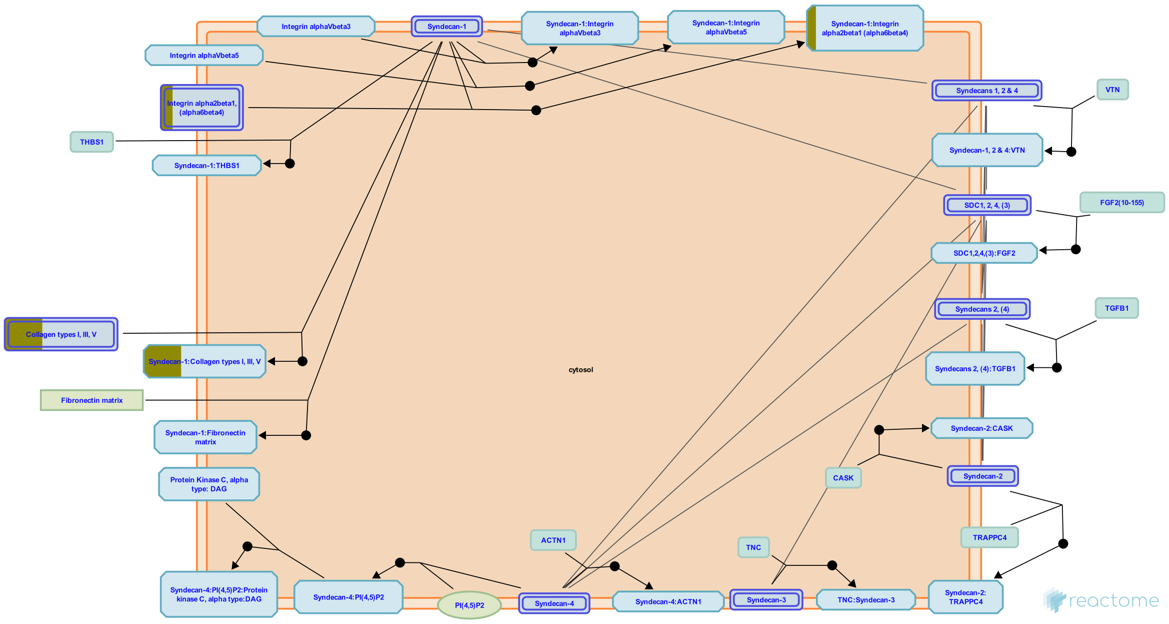


Syndecans are type I transmembrane proteins, with an N-terminal ectodomain that contains several consensus sequences for glycosaminoglycan (GAG) attachment and a short C-terminal cytoplasmic domain. Syndecan-1 and -3 GAG attachment sites occur in two distinct clusters, one near the N-terminus and the other near the membrane-attachment site, separated by a proline and threonine-rich 'spacer'. Syndecan ectodomain sequences are poorly conserved in the family and between species, but the transmembrane and cytoplasmic domains are highly conserved. Syndecan-1 and -3 form a subfamily. Syndecan core proteins form dimers (Choi et al. 2007) and at least syndecan-3 and -4 form oligomers (Asundi & Carey 1995, Shin et al. 2012). Syndecan-1 is the major syndecan of epithelial cells including vascular endothelium. Syndecan-2 is present mostly in mesenchymal, neuronal and smooth muscle cells. Syndecan-3 is the major syndecan of the nervous system, while syndecan-4 is ubiquitously expressed but at lower levels than the other syndecans (refs in Alexopoulou et al. 2007). The core syndecan protein has three to five heparan sulfate or chondroitin sulfate chains, which interact with a variety of ligands including fibroblast growth factors, vascular endothelial growth factor, transforming growth factor-beta, fibronectin, collagen, vitronectin and several integrins. Syndecans may act as integrin coreceptors. Interactions between fibronectin and syndecans are modulated by tenascin-C.

Syndecans bind a wide variety of soluble and insoluble ligands, inckluding extracellular matrix components, cell adhesion molecules, growth factors, cytokines, and proteinases. As the cleaved ectodomains of syndecans retain the ability to bind ligands, ectodomain shedding is a mechanism for releasing soluble effectors that may compete for ligands with their cell-bound counterparts (Kainulainen et al. 1998). Shed ectodomains are found in inflammatory fluids (Subramanian et al. 1997) and may induce the proliferation of cancer cells (Maeda et al. 2004).

#### Edit history

| Date | Action | Author |
|---|---|---|
| 2012-07-31 | Authored | Jupe S |
| 2013-01-24 | Created | Jupe S |
| 2013-04-26 | Edited | Jupe S |
| 2013-05-22 | Reviewed | Ricard-Blum S, Fuentes J |
| 2021-05-22 | Modified | Shorser S |

#### Entities found in this pathway (3)

| Input | UniProt Id | Input | UniProt Id | Input | UniProt Id |
|---|---|---|---|---|---|
| COL1A2 | P08123 | COL3A1 | P02461 | ITGB4 | P16144 |

### 7. Collagen degradation (R-HSA-1442490)

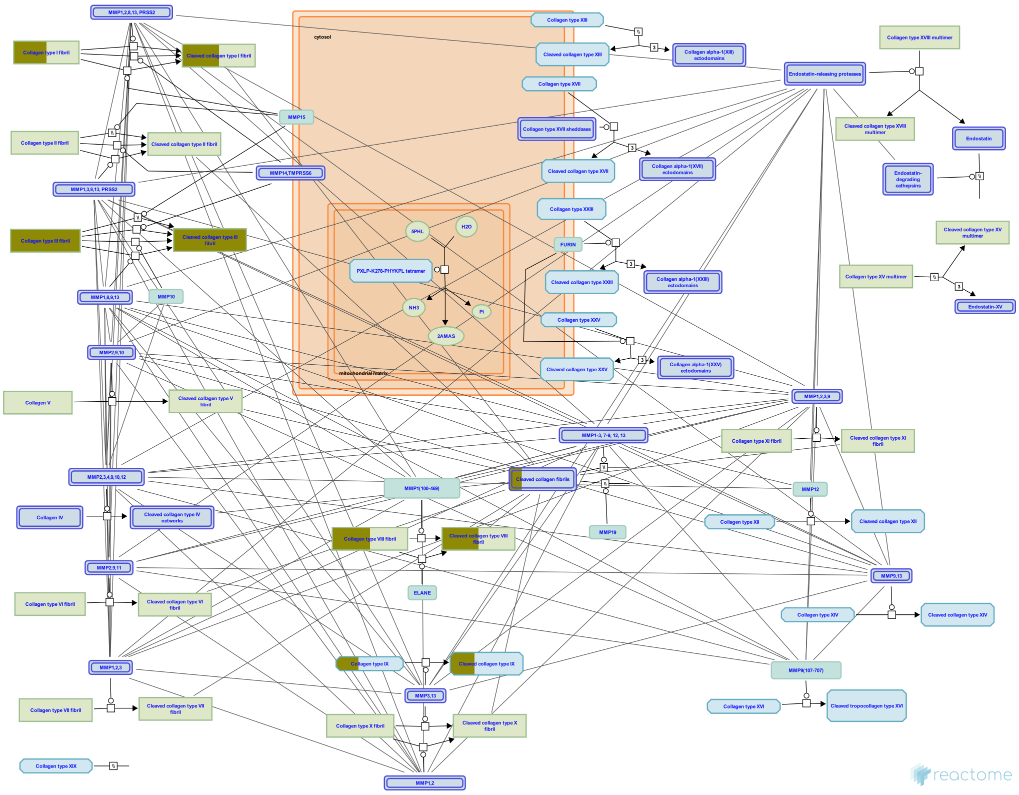

Collagen fibril diameter and spatial organisation are dependent on the species, tissue type and stage of development (Parry 1988). The lengths of collagen fibrils in mature tissues are largely unknown but in tendon can be measured in millimetres (Craig et al. 1989). Collagen fibrils isolated from adult bovine corneal stroma had ~350 collagen molecules in transverse section, tapering down to three molecules at the growing tip (Holmes & Kadler 2005).

The classical view of collagenases is that they actively unwind the triple helical chain, a process termed molecular tectonics (Overall 2002, Bode & Maskos 2003), before preferentially cleaving the alpha2 chain followed by the remaining chains (Chung et al. 2004). More recently it has been suggested that collagen fibrils exist in an equilibrium between protected and vulnerable states (Stultz 2002, Nerenberg & Stultz 2008). The prototypical triple-helical structure of collagen does not fit into the active site of collagenase MMPs. In addition the scissile bonds are not solvent-exposed and are therefore inaccessible to the collagenase active site (Chung et al. 2004, Stultz 2002). It was realized that collagen must locally unfold into non-triple helical regions to allow collagenolysis. Observations using circular dichroism and differential scanning calorimetry confirm that there is considerable heterogeneity along collagen fibres (Makareeva et al. 2008) allowing access for MMPs at physiological temperatures (Salsas-Escat et al. 2010).

Collagen fibrils with cut chains are unstable and accessible to proteinases that cannot cleave intact collagen strands (Woessner & Nagase 2000, Somerville et al. 2003). Continued degradation leads to the formation of gelatin (Lovejoy et al. 1999). Degradation of collagen types other than I-III is less well characterized but believed to occur in a similar manner.

Metalloproteinases (MMPs) play a major part in the degradation of several extracellular macromolecules including collagens. MMP1 (Welgus et al. 1981), MMP8 (Hasty et al. 1987), and MMP13 (Knauper et al. 1996), sometimes referred to as collagenases I, II and III respectively, are able to initiate the intrahelical cleavage of the major fibril forming collagens I, II and III at neutral pH, and thus thought to define the rate-limiting step in normal tissue remodeling events. All can cleave additional substrates including other collagen subtypes. Collagenases cut collagen alpha chains at a single conserved Gly-Ile/Leu site approximately 3/4 of the molecule's length from the N-terminus (Fields 1991, Chung et al. 2004). The cleavage site is characterised by the motif G(I/L)(A/L); the G-I/L bond is cleaved. In collagen type I this corresponds to G953-I954 in the Uniprot canonical alpha chain sequences (often given as G775-I776 in literature). It is not clear why only this bond is cleaved, as the motif occurs at several other places in the chain. MMP14, a membrane-associated MMP also known as Membrane-type matrix metalloproteinase 1 (MT-MMP1), is able to cleave collagen types I, II and III (Ohuchi et al. 1997).


#### Edit history

| Date | Action | Author |
|---|---|---|
| 2011-07-12 | Authored | Jupe S |
| 2011-07-12 | Created | Jupe S |
| 2012-10-08 | Reviewed | Sorsa T |
| 2012-11-12 | Edited | Jupe S |
| 2021-05-22 | Modified | Shorser S |

#### Entities found in this pathway (3)

| Input | UniProt Id | Input | UniProt Id | Input | UniProt Id |
|---|---|---|---|---|---|
| COL1A2 | P08123 | COL3A1 | P02461 | COL8A2 | P25067, Q14050 |

#### 8. Defective B4GALT1 causes B4GALT1-CDG (CDG-2d) (R-HSA-3656244)

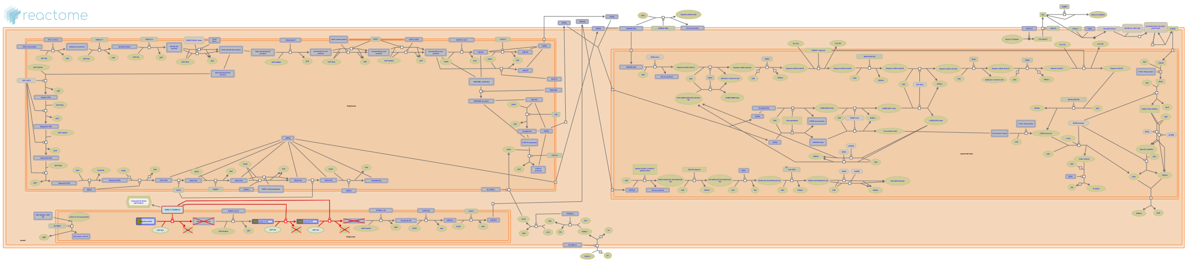

**Diseases:** congenital disorder of glycosylation type II.

Congenital disorders of glycosylation (CDG, previously called carbohydrate-deficient glycoprotein syndromes, CDGSs), are a group of hereditary multisystem disorders. They are characterized biochemically by hypoglycosylation of glycoproteins, diagnosed by isoelectric focusing (IEF) of serum transferrin. There are two types of CDG, types I and II. Type I CDG has defects in the assembly of lipid-linked oligosaccharides or their transfer onto nascent glycoproteins, whereas type II CDG comprises defects of trimming, elongation, and processing of protein-bound glycans. Clinical symptoms are dominated by severe psychomotor and mental retardation, as well as blood coagulation abnormalities (Jaeken 2013). B4GALT1-CDG (CDG type IId) is a multisystem disease, characterized by dysmorphic features, hydrocephalus, hypotonia and blood clotting abnormalities (Hansske et al. 2002).


##### Edit history

| Date | Action | Author |
|---|---|---|
| 2013-05-31 | Edited | Jassal B |
| 2013-05-31 | Authored | Jassal B |
| 2013-05-31 | Created | Jassal B |
| 2014-07-09 | Reviewed | Spillmann D |
| 2018-01-25 | Modified | Jassal B |

##### Entities found in this pathway (2)

| Input | UniProt Id | Input | UniProt Id |
|---|---|---|---|
| LUM | P51884 | OGN | P20774 |

#### 9. Defective CHST6 causes MCDC1 (R-HSA-3656225)

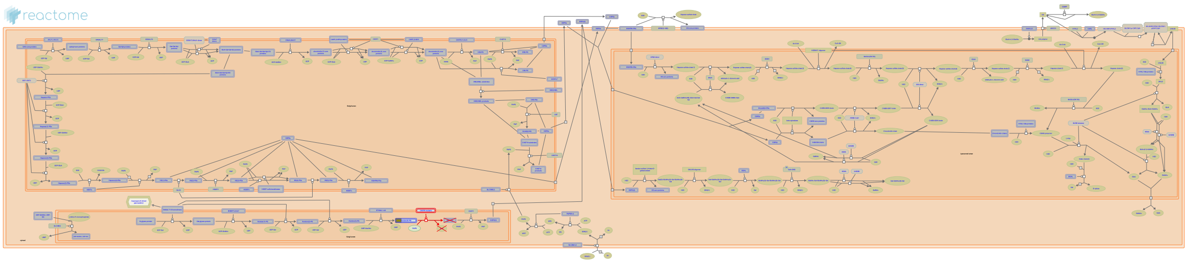

**Diseases:** macular corneal dystrophy.

Carbohydrate sulfotransferase 6 (CHST6) catalyzes the transfer of sulfate to position 6 of non-reducing ends of N-acetylglucosamine (GlcNAc) residues on keratan sulfate (KS). KS plays a central role in maintaining corneal transparency. Defective CHST6 (Nakazawa et al. 1984) results in unsulfated keratan deposited within the intracellular space and the extracellular corneal stroma leading to macular dystrophy, corneal type I (MCDC1; MIM:217800). MCDC1 is an early-onset, ocular disease characterized by bilateral, progressive corneal opacification, and reduced corneal sensitivity (Jones & Zimmerman 1961). MCD can be subdivided into 2 types on the basis of immunohistochemical studies and serum analysis for keratan sulfate; MCD type I, in which there is a virtual absence of sulfated KS-specific antibody response in the serum and cornea and MCD type II, in which the normal KS-specific antibody response is present in cornea and serum (Yang et al. 1988).


##### Edit history

| Date | Action | Author |
|---|---|---|
| 2013-05-31 | Edited | Jassal B |
| 2013-05-31 | Authored | Jassal B |
| 2013-05-31 | Created | Jassal B |
| 2014-07-09 | Reviewed | Spillmann D |
| 2015-02-09 | Modified | Wu G |

##### Entities found in this pathway (2)

| Input | UniProt Id | Input | UniProt Id |
|---|---|---|---|
| LUM | P51884 | OGN | P20774 |

#### 10. Defective ST3GAL3 causes MCT12 and EIEE15 (R-HSA-3656243)

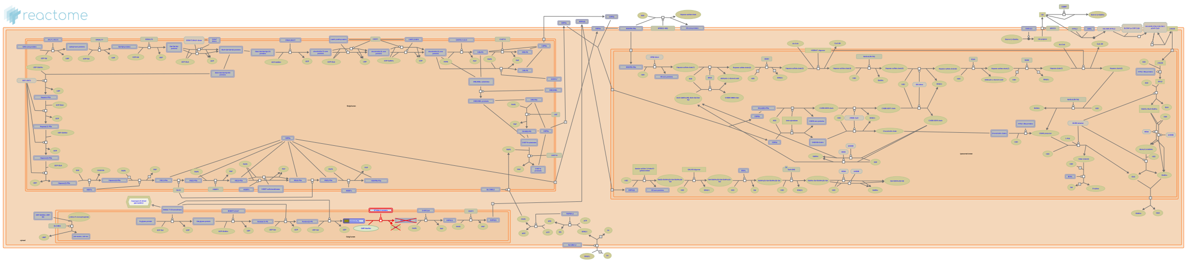

**Diseases:** developmental disorder of mental health.

CMP-N-acetylneuraminate-beta-1,4-galactoside alpha-2,3-sialyltransferase (ST3GAL3) mediates the transfer of sialic acid from CMP-sialic acid to galactose-containing glycoproteins and forms the sialyl Lewis a epitope on proteins which are required for attaining and/or maintaining higher cognitive functions. Some defects in ST3GAL3 result in mental retardation, autosomal recessive 12 (MRT12; MIM:611090), a disorder characterised by below average general intellectual function and impaired adaptive behaviour (Najmabadi et al. 2007, Hu et al. 2011). Another defect of ST3GAL3 can cause early infantile epileptic encephalopathy-15 (EIEE15: MIM:615006), resulting in severe mental retardation (Edvardson et al. 2012).


##### Edit history

| Date | Action | Author |
|---|---|---|
| 2013-05-31 | Edited | Jassal B |
| 2013-05-31 | Authored | Jassal B |
| 2013-05-31 | Created | Jassal B |
| 2014-07-09 | Reviewed | Spillmann D |
| 2015-09-01 | Modified | Jassal B |

##### Entities found in this pathway (2)

| Input | UniProt Id | Input | UniProt Id |
|---|---|---|---|
| LUM | P51884 | OGN | P20774 |

#### 11. Toxicity of botulinum toxin type F (botF) (R-HSA-5250981)

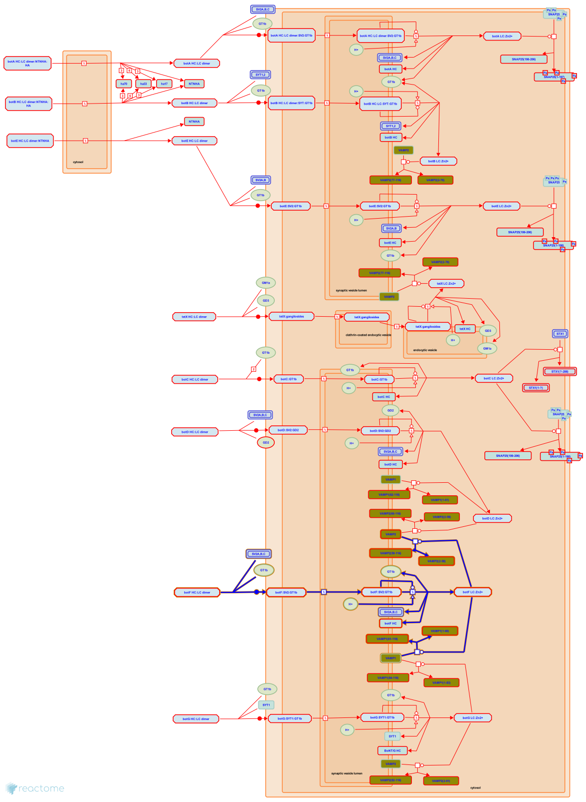

**Diseases:** botulism.

Botulinum toxin type F (botF) is only very rarely associated with human disease (Hatheway 1995) and a pathway by which it might enter the circulation from the human gut has not been described. Nevertheless, the toxin itself, a disulfide-bonded heavy chain (HC) - light chain (LC) heterodimer ("dichain"), is capable of binding to neurons by interactions with cell-surface ganglioside and synaptic vesicle protein 2 (SV2) (Fu et al. 2009; Rummel et al. 2009), the bound toxin can enter synaptic vesicles and release its LC moiety into the cytosol of targeted cells (Montal 2010), and the botF LC can cleave vesicle-associated membrane proteins 1 and 2 (VAMP1 and 2) on the cytosolic face of the synaptic vesicle membrane (Yamasaki et al. 1994). These four events are annotated here.

#### Edit history

| Date | Action | Author |
|---|---|---|
| 2006-06-15 | Authored | Krupa S, Gopinathrao G |
| 2007-08-03 | Reviewed | Ichtchenko K |
| 2014-02-01 | Created | D'Eustachio P |
| 2014-02-11 | Revised | D'Eustachio P |
| 2014-02-11 | Edited | D'Eustachio P |
| 2014-11-18 | Reviewed | Sharma S, Thirunavukkarasu N |
| 2020-10-08 | Modified | D'Eustachio P |

#### Entities found in this pathway (1)

| Input | UniProt Id |
|---|---|
| VAMP1 | P23763, P63027 |

#### 12. Toxicity of botulinum toxin type D (botD) (R-HSA-5250955)

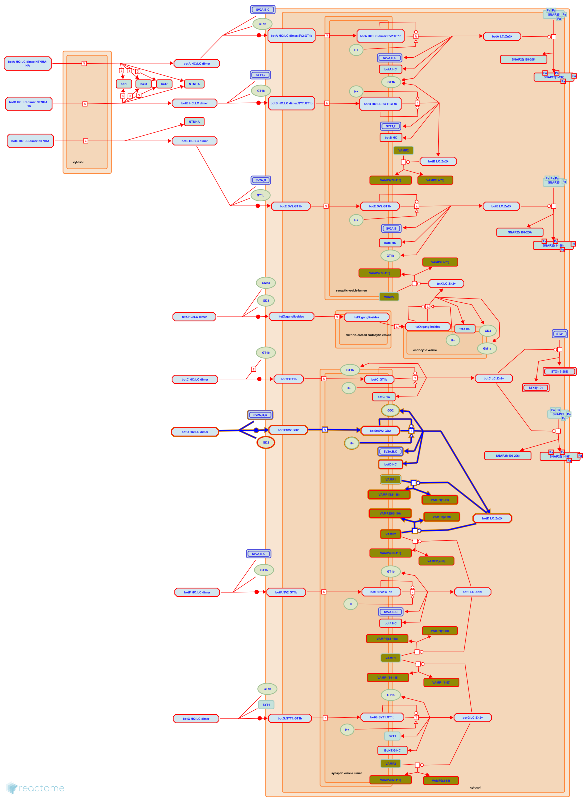

**Diseases:** botulism.

Botulinum toxin type D (botD) is only very rarely associated with human disease (Hatheway 1995) and a pathway by which it might enter the circulation from the human gut has not been described. Nevertheless, the toxin itself, a disulfide-bonded heavy chain (HC) - light chain (LC) heterodimer ("dichain"), is capable of binding to neurons by interactions with cell surface ganglioside (Kroken et al. 2011) and synaptic vesicle protein 2 (SV2) (Peng et al. 2011), the bound toxin can enter synaptic vesicles and release its LC moiety into the cytosol of targeted cells (Montal 2010), and the botD LC can cleave vesicle associated membrane proteins 1 and 2 (VAMP1 and 2) on the cytosolic face of the synaptic vesicle membrane (Schiavo et al. 1993; Yamasaki et al. 1994). These four events are annotated here.

#### Edit history

| Date | Action | Author |
|---|---|---|
| 2006-06-15 | Authored | Krupa S, Gopinathrao G |
| 2007-08-03 | Reviewed | Ichtchenko K |
| 2014-02-01 | Created | D'Eustachio P |
| 2014-02-11 | Revised | D'Eustachio P |
| 2014-02-11 | Edited | D'Eustachio P |
| 2014-11-18 | Reviewed | Sharma S, Thirunavukkarasu N |
| 2020-10-08 | Modified | D'Eustachio P |

#### Entities found in this pathway (1)

| Input | UniProt Id |
|---|---|
| VAMP1 | P23763, P63027 |

#### 13. Collagen biosynthesis and modifying enzymes (R-HSA-1650814)

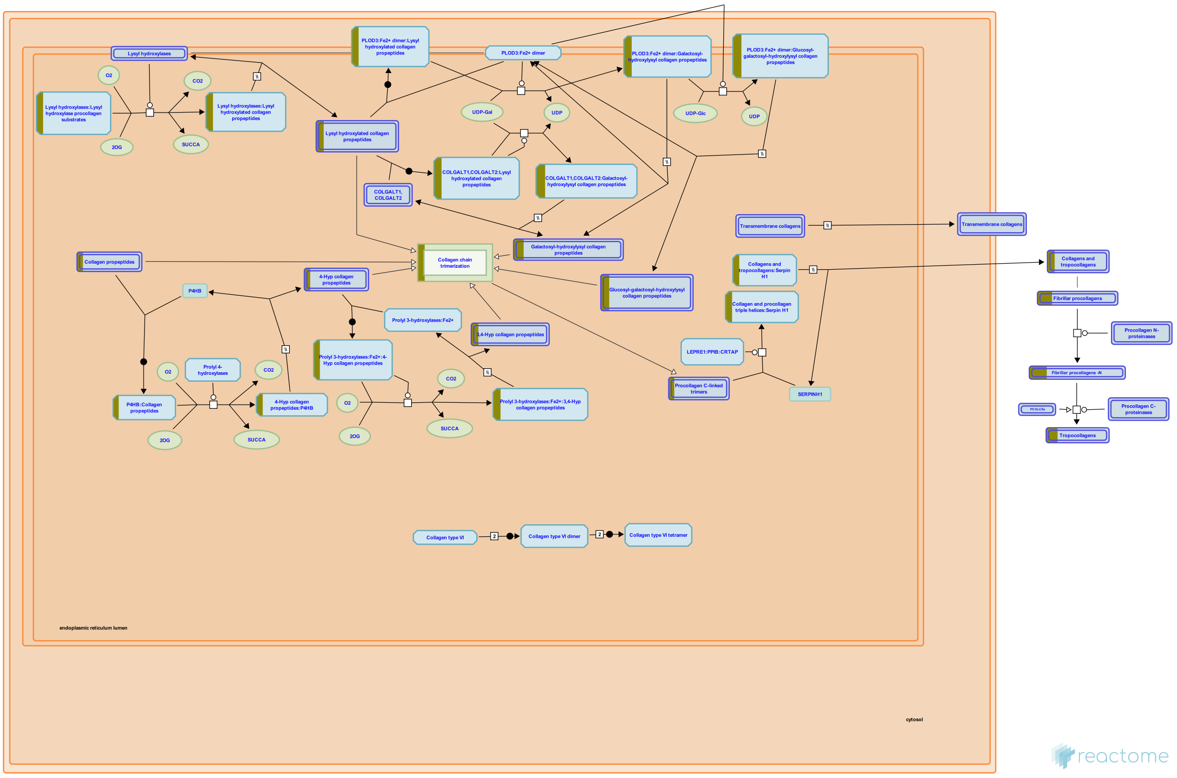

The biosynthesis of collagen is a multistep process. Collagen propeptides are cotranslationally translocated into the ER lumen. Propeptides undergo a number of post-translational modifications. Proline and lysine residues may be hydroxylated by prolyl 3-, prolyl 4- and lysyl hydroxylases. 4-hydroxyproline is essential for intramolecular hydrogen bonding and stability of the triple helical collagenous domain. In fibril forming collagens approximately 50% of prolines are 4-hydroxylated; the extent of this and of 3-hydroxyproline and lysine hydroxylation varies between tissues and collagen types (Kivirikko et al. 1972, 1992). Hydroxylysine molecules can form cross-links between collagen molecules in fibrils, and are sites for glycosyl- and galactosylation. Collagen peptides all have non-collagenous domains; collagens within the subclasses have common chain structures. These non-collagenous domains have regulatory functions; some are biologically active when cleaved from the main peptide chain. Fibrillar collagens all have a large triple helical domain (COL1) bordered by N and C terminal extensions, called the N and C propeptides, which are cleaved prior to formation of the collagen fibril. The C propeptide, also called the NC1 domain, is highly conserved. It directs chain association during intracellular assembly of the procollagen molecule from three collagen propeptide alpha chains (Hulmes 2002). The N-propeptide has a short linker (NC2) connecting the main triple helix to a short minor one (COL2) and a globular N-terminal region NC3. NC3 domains are variable both in size and the domains they contain.

Collagen propeptides typically undergo a number of post-translational modifications. Proline and lysine residues are hydroxylated by prolyl 3-, prolyl 4- and lysyl hydroxylases. 4-hydroxyproline is essential for intramolecular hydrogen bonding and stability of the triple helical collagenous domain. Prolyl 4-hydroxylase may also have a role in alpha chain association as no association of the C-propeptides of type XII collagen was seen in the presence of prolyl 4-hydroxylase inhibitors (Mazzorana et al. 1993, 1996). In fibril forming collagens approximately 50% of prolines are 4-hydroxylated; the extent of this is species dependent, lower hydroxylation correlating with lower ambient temperature and thermal stability (Cohen-Solal et al. 1986, Notbohm et al. 1992). Similarly the extent of 3-hydroxyproline and lysine hydroxylation varies between tissues and collagen types (Kivirikko et al. 1992). Hydroxylysine molecules can form cross-links between collagen molecules in fibrils, and are sites for glycosyl- and galactosylation.

Collagen molecules fold and assemble through a series of distinct intermediates (Bulleid 1996). Individual collagen polypeptide chains are translocated co-translationally across the membrane of the endoplasmic reticulum (ER). Intra-chain disulfide bonds are formed within the N-propeptide, and hydroxylation of proline and lysine residues occurs within the triple helical domain (Kivirikko et al. 1992). When the peptide chain is fully translocated into the ER lumen the C-propeptide folds, the conformation being stabilized by intra-chain disulfide bonds (Doege and Fessler 1986). Pro alpha-chains associate via the C-propeptides (Byers et al. 1975, Bachinger et al. 1978), or NC2 domains for FACIT family collagens (Boudko et al. 2008) to form an initial trimer which can be stabilized by the formation of inter-chain disulfide bonds (Schofield et al. 1974, Olsen et al. 1976), though these are not a prerequisite for further folding (Bulleid et al. 1996). The triple helix then nucleates and folds in a C- to N- direction. The association of the individual chains and subsequent triple helix formation are distinct steps (Bachinger et al. 1980). The N-propeptides associate and in some cases form inter-chain disulfide bonds (Bruckner et al., 1978). Procollagen is released via carriers into the exracellular space (Canty & Kadler 2005). Fibrillar procollagens undergo removal of the C- and N-propeptides by procollagen C and N proteinases respectively, both Zn2+ dependent metalloproteinases. Propeptide processing is a required step for normal collagen I and III fibril formation, but collagens can retain some or all of their non-collagenous propeptides. Retained collagen type V and XI N-propeptides contribute to the control of fibril growth by sterically limiting lateral molecule addition (Fichard et al. 1995). Processed fibrillar procollagen is termed tropocollagen, which is considered to be the unit of higher order fibrils and fibres. Tropocollagens of the fibril forming collagens I, II, III, V and XI sponteneously aggregate in vitro in a manner that has been compared with crystallization, commencing with a nucleation event followed by subsequent organized aggregation (Silver et al. 1992, Prockop & Fertala 1998). Fibril formation is stabilized by lysyl oxidase catalyzed crosslinks between adjacent molecules (Siegel & Fu 1976).

#### Edit history

| Date | Action | Author |
|---|---|---|
| 2010-07-20 | Authored | Jupe S |

| Date | Action | Author |
| --- | --- | --- |
| 2011-10-12 | Created | Jupe S |
| 2012-05-14 | Edited | Jupe S |
| 2012-05-24 | Reviewed | Canty-Laird EG |
| 2021-05-22 | Modified | Shorser S |

#### Entities found in this pathway (3)

| Input | UniProt Id | Input | UniProt Id | Input | UniProt Id |
| --- | --- | --- | --- | --- | --- |
| COL1A2 | P08123 | COL3A1 | P02461 | COL8A2 | P25067, Q14050 |

#### 14. Regulation of Insulin-like Growth Factor (IGF) transport and uptake by Insulin-like Growth Factor Binding Proteins (IGFBPs) (R-HSA-381426)

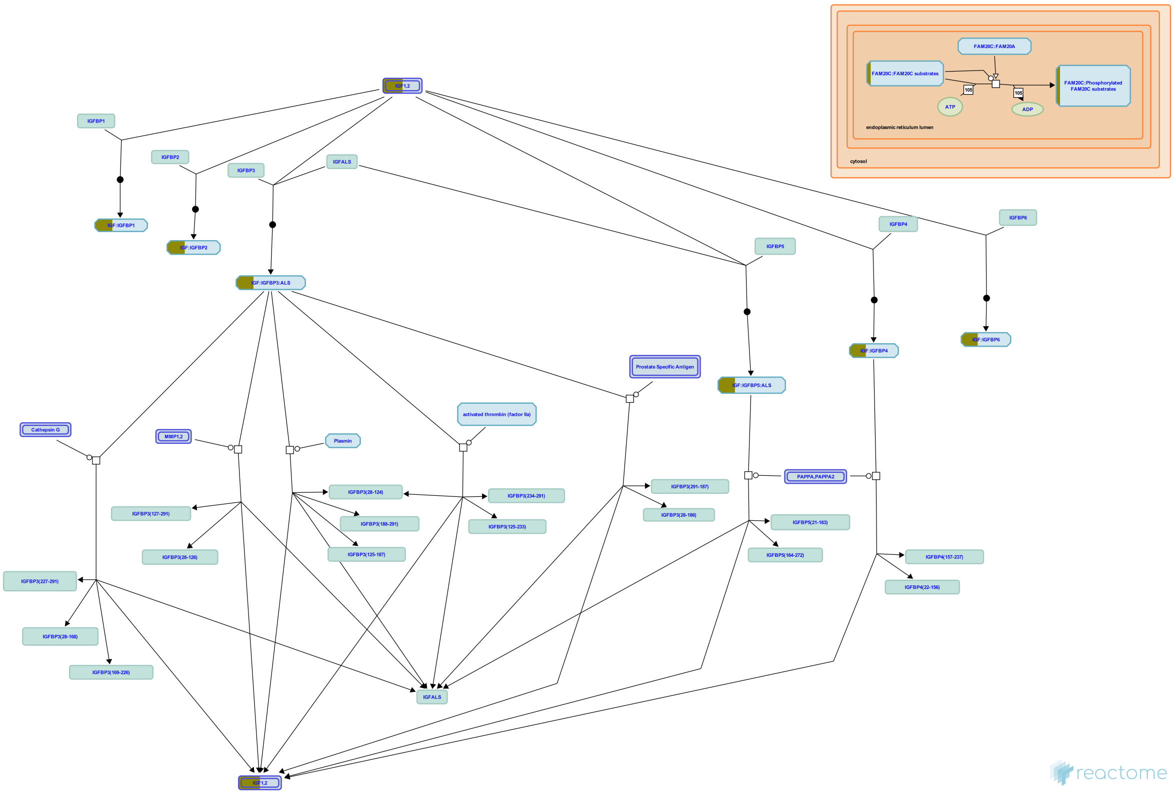


**Cellular compartments:** extracellular region.

The family of Insulin like Growth Factor Binding Proteins (IGFBPs) share 50% amino acid identity with conserved N terminal and C terminal regions responsible for binding Insulin like Growth Factors I and II (IGF I and IGF II). Most circulating IGFs are in complexes with IGFBPs, which are believed to increase the residence of IGFs in the body, modulate availability of IGFs to target receptors for IGFs, reduce insulin like effects of IGFs, and act as signaling molecules independently of IGFs.
About 75% of circulating IGFs are in 1500 220 KDa complexes with IGFBP3 and ALS. Such complexes are too large to pass the endothelial barrier. The remaining 20 25% of IGFs are bound to other IGFBPs in 40 50 KDa complexes. IGFs are released from IGF:IGFBP complexes by proteolysis of the IGFBP. IGFs become active after release, however IGFs may also have activity when still bound to some IGFBPs.
IGFBP1 is enriched in amniotic fluid and is produced in the liver under control of insulin (insulin suppresses production). IGFBP1 binding stimulates IGF function. It is unknown which if any protease degrades IGFBP1.
IGFBP2 is enriched in cerebrospinal fluid; its binding inhibits IGF function. IGFBP2 is not significantly degraded in circulation.
IGFB3, which binds most IGF in the body is enriched in follicular fluid and found in many other tissues. IGFBP 3 may be cleaved by plasmin, thrombin, Prostate specific Antigen (PSA, KLK3), Matrix Metalloprotease-1 (MMP1), and Matrix Metalloprotease-2 (MMP2). IGFBP3 also binds extracellular matrix and binding lowers its affinity for IGFs. IGFBP3 binding stimulates the effects of IGFs.
IGFBP4 acts to inhibit IGF function and is cleaved by Pregnancy associated Plasma Protein A (PAPPA) to release IGF.
IGFBP5 is enriched in bone matrix; its binding stimulates IGF function. IGFBP5 is cleaved by Pregnancy Associated Plasma Protein A2 (PAPPA2), ADAM9, complement C1s from smooth muscle, and thrombin. Only the cleavage site for PAPPA2 is known.
IGFBP6 is enriched in cerebrospinal fluid. It is unknown which if any protease degrades IGFBP6.


##### Edit history

| Date | Action | Author |
|---|---|---|
| 2008-11-20 | Edited | May B, Gopinathrao G |
| 2008-11-20 | Created | May B |
| 2008-12-02 | Reviewed | Matthews L, D'Eustachio P, Gillespie ME |

| Date | Action | Author |
|---|---|---|
| 2021-05-22 | Modified | Shorser S |

#### Entities found in this pathway (4)

| Input | UniProt Id |
|---|---|
| CABP1 | Q15084 |
| ITIH2 | P19823 |

| Input | UniProt Id |
|---|---|
| IGF1 | P05019 |
| SCG2 | O00255, P13521 |

#### 15. ECM proteoglycans (R-HSA-3000178)

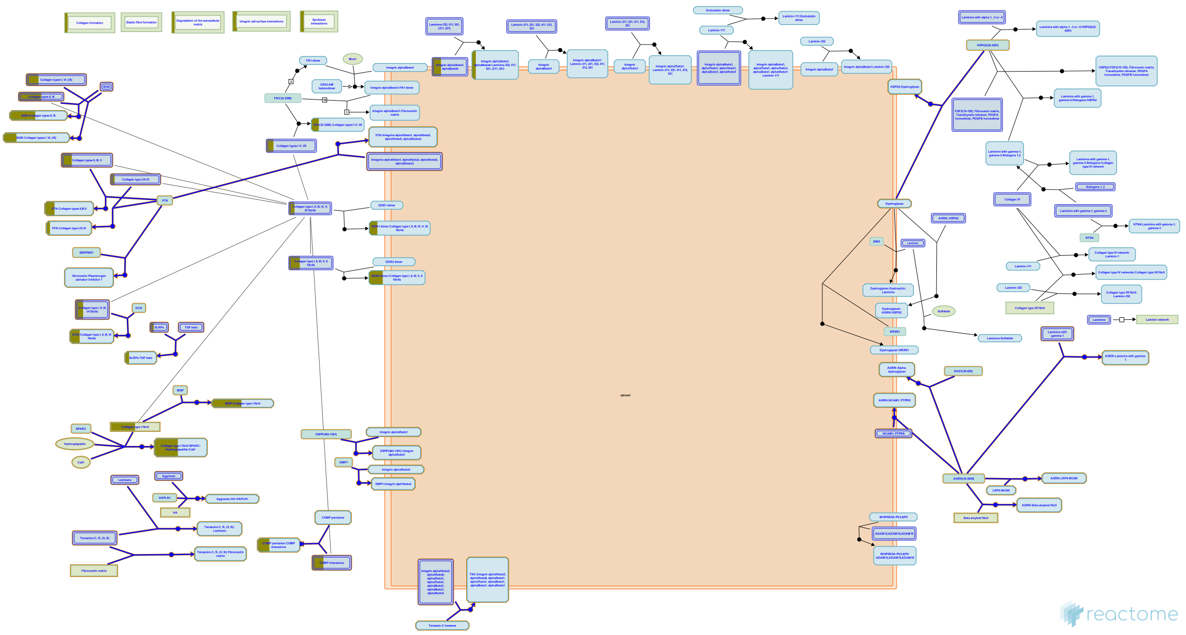

Proteoglycans are major components of the extracellular matrix. In cartilage the matrix constitutes more than 90% of tissue dry weight. Proteoglycans are proteins substituted with glycosaminoglycans (GAGs), linear polysaccharides consisting of a repeating disaccharide, generally of an acetylated amino sugar alternating

with a uronic acid. Most proteoglycans are located in the extracellular

space. Proteoglycans are highly diverse, both in terms of the core proteins and the subtypes of GAG chains, namely chondroitin sulfate (CS), keratan sulfate (KS), dermatan sulfate (DS) and heparan sulfate (HS). Hyaluronan is a non-sulfated GAG whose molecular weight runs into millions of Dalton; in articular cartilage, a single hyaluronan molecule can hold upto 100 aggrecan molecules and these aggregates are stabilized by a link protein.


##### Edit history

| Date | Action | Author |
|---|---|---|
| 2013-01-10 | Authored | Jupe S |
| 2013-01-24 | Created | Jupe S |
| 2013-04-26 | Edited | Jupe S |
| 2013-05-21 | Reviewed | Venkatesan N |
| 2013-05-22 | Reviewed | Ricard-Blum S |

| Date | Action | Author |
|---|---|---|
| 2021-05-22 | Modified | Shorser S |

#### Entities found in this pathway (4)

| Input | UniProt Id | Input | UniProt Id |
|---|---|---|---|
| COL1A2 | P08123 | COL3A1 | P02461 |
| COL8A2 | Q14050 | LUM | P51884 |

#### 16. GP1b-IX-V activation signalling (R-HSA-430116)

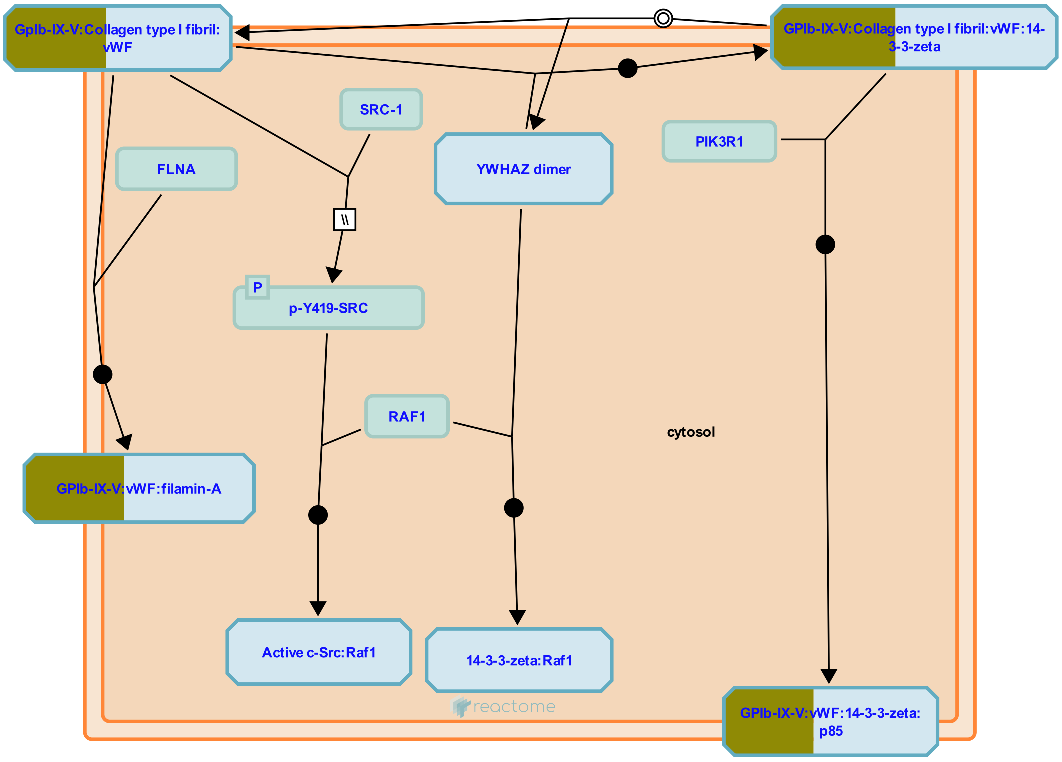


**Cellular compartments:** plasma membrane.

The platelet GPIb complex (GP1b-IX-V) together with GPVI are primarily responsible for regulating the initial adhesion of platelets to the damaged blood vessel and platelet activation. The importance of GPIb is demonstrated by the bleeding problems in patients with Bernard-Soulier syndrome where this receptor is either absent or defective. GP1b-IX-V binds von Willebrand Factor (vWF) to resting platelets, particularly under conditions of high shear stress. This transient interaction is the first stage of the vascular repair process. Activation of GP1b-IX-V on exposure of the fibrous matrix following atherosclerotic plaque rupture, or in occluded arteries, is a major contributory factor leading to thrombus formation leading to heart attack or stroke.
GpIb also binds thrombin (Yamamoto et al. 1986), at a site distinct from the site of vWF binding, acting as a docking site for thrombin which then activates Proteinase Activated Receptors leading to enhanced platelet activation (Dormann et al. 2000).

##### Edit history

| Date | Action | Author |
|---|---|---|
| 2009-06-03 | Authored | Akkerman JW |
| 2009-07-30 | Created | Jupe S |
| 2010-06-07 | Edited | Jupe S |

| Date | Action | Author |
|---|---|---|
| 2010-06-07 | Reviewed | Kunapuli SP |
| 2021-05-22 | Modified | Shorser S |

#### Entities found in this pathway (2)

| Input | UniProt Id | Input | UniProt Id |
|---|---|---|---|
| COL1A2 | P08123 | VWF | P04275 |

#### 17. MECP2 regulates transcription of neuronal ligands (R-HSA-9022702)

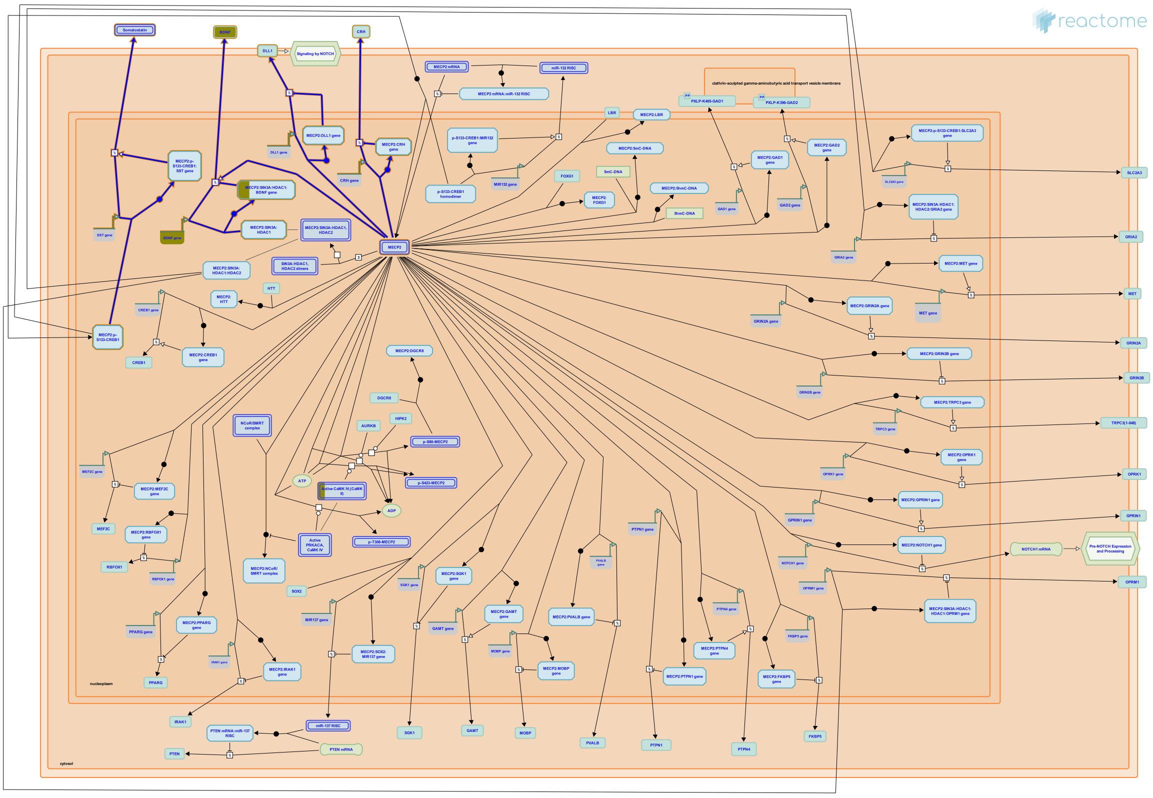

Ligands regulated by MECP2 include BDNF (reviewed by Li and Pozzo Miller 2014, and KhorshidAhmad et al. 2016), CRH (McGill et al. 2006, Samaco et al. 2012), SST (Somatostatin) (Chahrour et al. 2008), and DLL1 (Li et al. 2014).


##### Edit history

| Date | Action | Author |
|---|---|---|
| 2017-09-25 | Created | Orlic-Milacic M |

| Date | Action | Author |
| --- | --- | --- |
| 2017-10-02 | Authored | Orlic-Milacic M |
| 2018-08-07 | Reviewed | Christodoulou J, Krishnaraj R |
| 2018-08-08 | Modified | Orlic-Milacic M |
| 2018-08-08 | Edited | Orlic-Milacic M |

#### Entities found in this pathway (1)

| Input | UniProt Id |
| --- | --- |
| BDNF | P23560 |

| Input | Ensembl Id |
| --- | --- |
| BDNF | ENSG00000176697 |

#### 18. Platelet Adhesion to exposed collagen (R-HSA-75892)

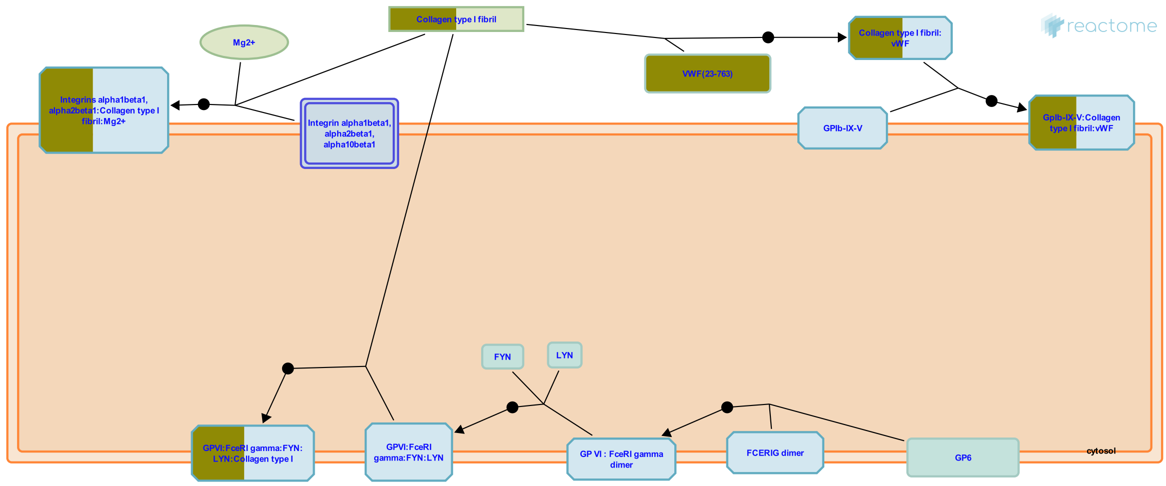


Initiation of platelet adhesion is the first step in the formation of the platelet plug. Circulating platelets are arrested and subsequently activated by exposed collagen and vWF. It is not entirely clear which type of collagen is responsible for adhesion and activation; collagen types I and III are abundant in vascular epithelia but several other types incluing IV are present (Farndale 2006). Several collagen binding proteins are expressed on platelets, including integrin alpha2 beta1, GPVI, and GPIV. Integrin alpha2 beta1, known on leukocytes as VLA-2, is the major platelet collagen receptor (Kunicki et al. 1988). It requires Mg2+ to interact with collagen and may require initiation mediated by the activation of integrin alphaIIb beta3 (van de Walle 2007). Binding occurs via the alpha2 subunit I domain to a collagen motif with the sequence Gly-Phe-Hyp-Gly-Glu-Arg (Emsley 2000). Binding of collagen to alpha2 beta1 generates intracellular signals that contribute to platelet activation. These facilitate the engagement of the lower-affinity collagen receptor, GPVI (Keely 1996), the key receptor involved in collagen-induced platelet activation. The GPVI receptor is a complex of the GPVI protein with a dimer of Fc epsilon R1 gamma (FceRI gamma). The Src family kinases Fyn and Lyn constitutively associate with the GPVI:FceRIgamma complex in platelets and initiate platelet activation through phosphorylation of the immunoreceptor tyrosine-based activation motif (ITAM) in FceRI gamma, leading to binding and activation of the tyrosine kinase Syk. Downstream of Syk, a series of adapter molecules and effectors lead to platelet activation. vWF protein is a polymeric structure of variable size. It is secreted in two directions, by the endothelium basolaterally and into the bloodstream. Shear-induced aggregation is achieved when vWF binds via its A1 domain to GPIb (part of GPIb-IX-V), and via its A3 domain mediating collagen binding to the subendothelium. The interaction between vWF and GPIb is regulated by shear force; an increase in the shear stress results in a corresponding increase in the affinity of vWF for GPIb.


##### Edit history

| Date | Action | Author |
|---|---|---|
| 2004-08-13 | Authored | de Bono B |
| 2004-09-25 | Created | Farndale R, Pace NP, de Bono B |
| 2021-05-21 | Modified | Shorser S |

#### Entities found in this pathway (2)

| Input | UniProt Id | Input | UniProt Id |
|---|---|---|---|
| COL1A2 | P08123 | VWF | P04275 |

#### 19. Neurotransmitter receptors and postsynaptic signal transmission (R-HSA-112314)

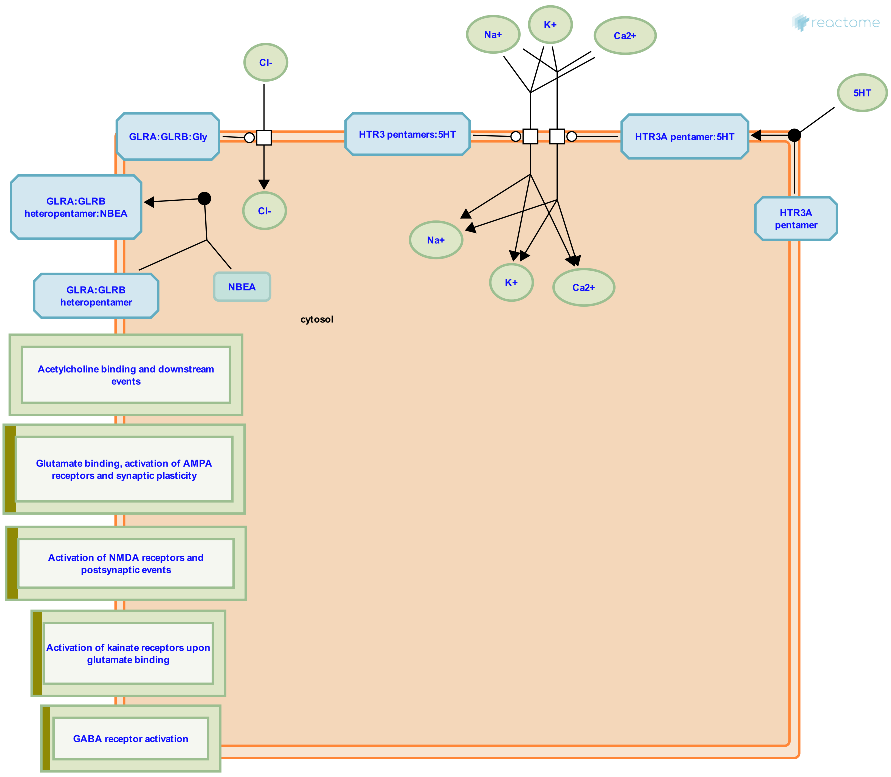


The neurotransmitter in the synaptic cleft released by the pre-synaptic neuron binds specific receptors located on the post-synaptic terminal. These receptors are either ion channels or G protein coupled receptors that function to transmit the signals from the post-synaptic membrane to the cell body.

##### References

##### Edit history

| Date | Action | Author |
|---|---|---|
| 2004-04-22 | Created | Joshi-Tope G |
| 2008-01-14 | Authored | Mahajan SS |
| 2008-12-02 | Reviewed | Restituito S, Kavalali E |
| 2021-05-22 | Modified | Shorser S |

##### Entities found in this pathway (6)

| Input | UniProt Id | Input | UniProt Id | Input | UniProt Id |
|---|---|---|---|---|---|
| CAMK2A | Q9UQM7 | GABRA5 | P31644 | GABRQ | Q9UN88 |
| GRIK1 | P39086 | GRIN3A | Q8TCU5 | NRGN | Q92686 |

#### 20. Post-translational protein phosphorylation (R-HSA-8957275)

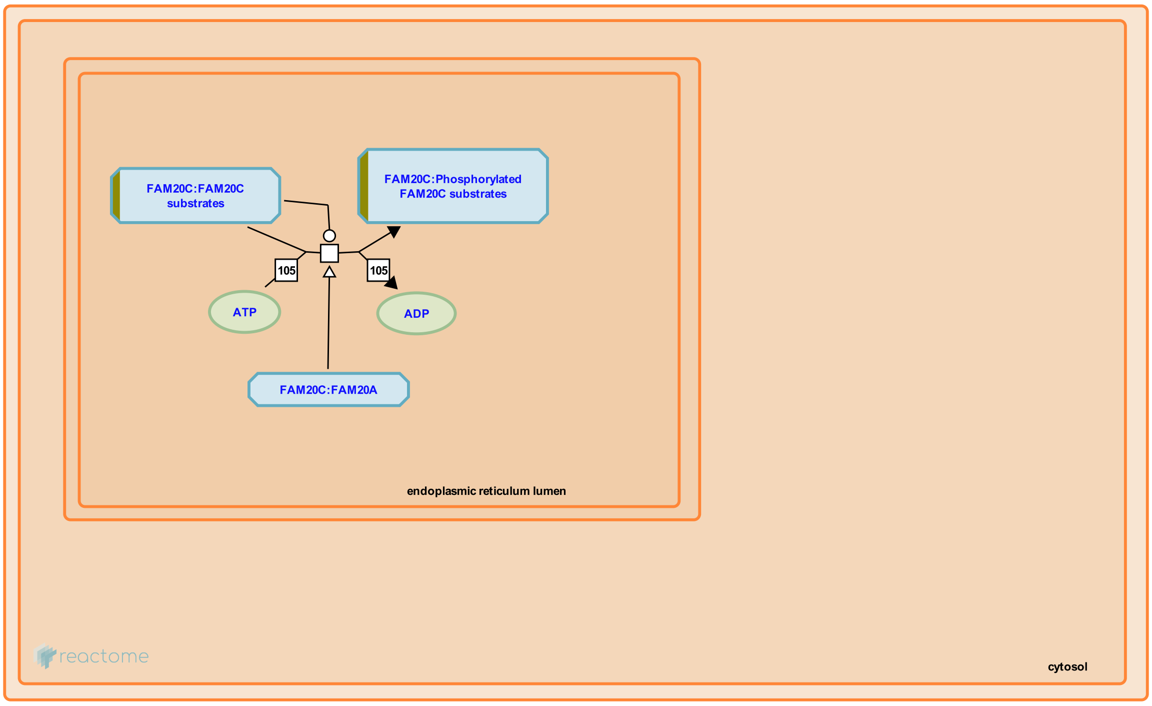


Secretory pathway kinases phosphorylate a diverse array of substrates involved in many physiological processes.


##### Edit history

| Date | Action | Author |
|---|---|---|
| 2016-12-08 | Authored | Jupe S |
| 2017-01-23 | Reviewed | Wiley SE |
| 2017-01-24 | Edited | Jupe S |
| 2017-01-24 | Created | Jupe S |
| 2021-05-31 | Modified | Shorser S |

##### Entities found in this pathway (3)

| Input | UniProt Id | Input | UniProt Id | Input | UniProt Id |
|---|---|---|---|---|---|
| CABP1 | Q15084 | ITIH2 | P19823 | SCG2 | O00255, P13521 |

#### 21. Non-integrin membrane-ECM interactions (R-HSA-3000171)

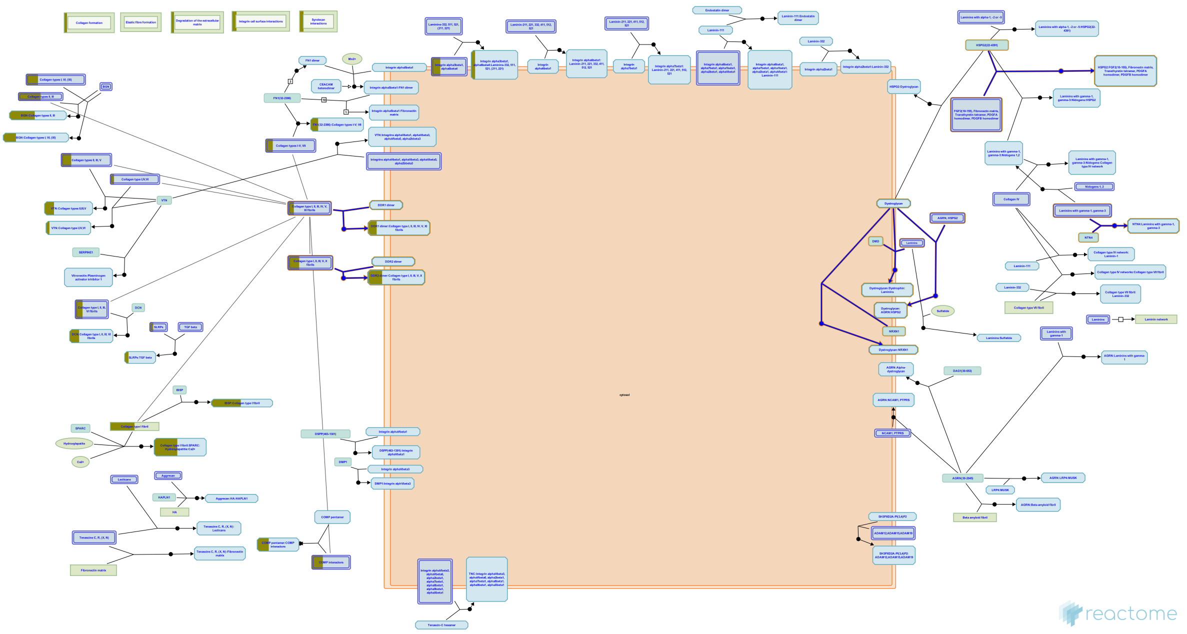

Several non-integrin membrane proteins interact with extracellular matrix proteins. Transmembrane proteoglycans may associate with integrins and growth factor receptors to influence their function, or they can signal independently, often influencing the actin cytoskeleton.


##### Edit history

| Date | Action | Author |
|---|---|---|
| 2012-07-31 | Authored | Jupe S |
| 2013-01-24 | Created | Jupe S |
| 2013-04-26 | Edited | Jupe S |
| 2013-05-22 | Reviewed | Ricard-Blum S |
| 2021-05-22 | Modified | Shorser S |

##### Entities found in this pathway (3)

| Input | UniProt Id | Input | UniProt Id | Input | UniProt Id |
|---|---|---|---|---|---|
| COL1A2 | P08123 | COL3A1 | P02461 | ITGB4 | P16144 |

#### 22. Keratan sulfate degradation (R-HSA-2022857)

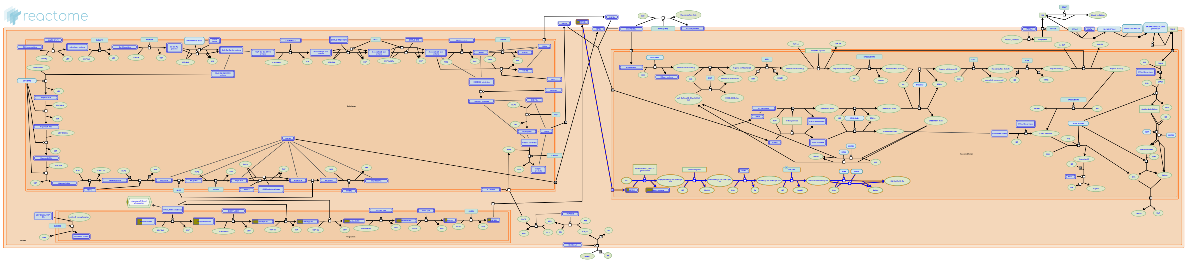

Keratan sulfate proteoglycans (KSPGs) are degraded in lysosomes as part of normal homeostasis of glycoproteins. Glycoproteins must be completely degraded to avoid undigested fragments building up and causing a variety of lysosomal storage diseases. KSPGs are Asn-linked glycoproteins and are acted upon by exo-glycosidases to release sugar monomers. The main steps of degradation are shown representing the types of cleavage reactions that occur so the full degradation of KS is not shown to avoid repetition. The proteolysis of the core protein of the glycoprotein is not shown here (Winchester 2005, Aronson & Kuranda 1989).


##### Edit history

| Date | Action | Author |
|---|---|---|
| 2011-12-01 | Edited | Jassal B |
| 2011-12-01 | Authored | Jassal B |
| 2011-12-01 | Created | Jassal B |
| 2012-03-28 | Reviewed | D'Eustachio P |
| 2021-05-22 | Modified | Shorser S |

##### Entities found in this pathway (2)

| Input | UniProt Id | Input | UniProt Id |
|---|---|---|---|
| LUM | P51884 | OGN | P20774 |

#### 23. Extracellular matrix organization (R-HSA-1474244)

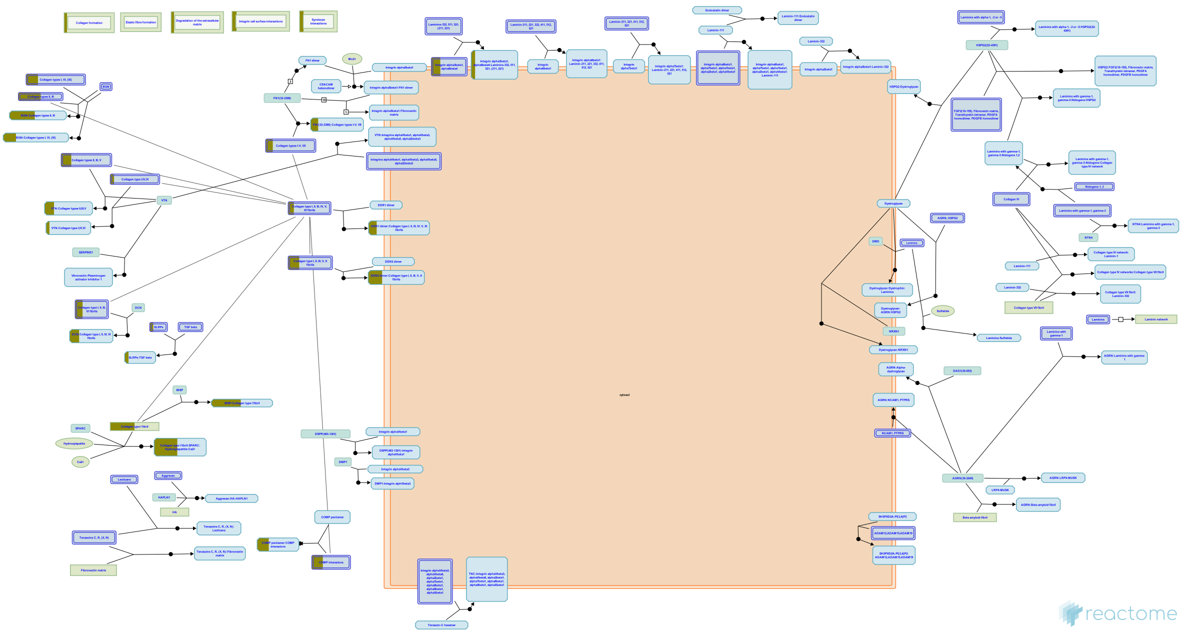


The extracellular matrix is a component of all mammalian tissues, a network consisting largely of the fibrous proteins collagen, elastin and associated-microfibrils, fibronectin and laminins embedded in a viscoelastic gel of anionic proteoglycan polymers. It performs many functions in addition to its structural role; as a major component of the cellular microenvironment it influences cell behaviours such as proliferation, adhesion and migration, and regulates cell differentiation and death (Hynes 2009).

ECM composition is highly heterogeneous and dynamic, being constantly remodeled (Frantz et al. 2010) and modulated, largely by matrix metalloproteinases (MMPs) and growth factors that bind to the ECM influencing the synthesis, crosslinking and degradation of ECM components (Hynes 2009). ECM remodeling is involved in the regulation of cell differentiation processes such as the establishment and maintenance of stem cell niches, branching morphogenesis, angiogenesis, bone remodeling, and wound repair. Redundant mechanisms modulate the expression and function of ECM modifying enzymes. Abnormal ECM dynamics can lead to deregulated cell proliferation and invasion, failure of cell death, and loss of cell differentiation, resulting in congenital defects and pathological processes including tissue fibrosis and cancer.

Collagen is the most abundant fibrous protein within the ECM constituting up to 30% of total protein in multicellular animals. Collagen provides tensile strength. It associates with elastic fibres, composed of elastin and fibrillin microfibrils, which give tissues the ability to recover after stretching. Other ECM proteins such as fibronectin, laminins, and matricellular proteins participate as connectors or linking proteins (Daley et al. 2008).

Chondroitin sulfate, dermatan sulfate and keratan sulfate proteoglycans are structural components associated with collagen fibrils (Scott & Haigh 1985; Scott & Orford 1981), serving to tether the fibril to the surrounding matrix. Decorin belongs to the small leucine-rich repeat proteoglycan family (SLRPs) which also includes biglycan, fibromodulin, lumican and asporin. All appear to be involved in collagen fibril formation and matrix assembly (Ameye & Young 2002).

ECM proteins such as osteonectin (SPARC), osteopontin and thrombospondins -1 and -2, collectively referred to as matricellular proteins (reviewed in Mosher & Adams 2012) appear to modulate cell-matrix interactions. In general they induce de-adhesion, characterized by disruption of focal adhesions and a reorganization of actin stress fibers (Bornstein 2009). Thrombospondin (TS)-1 and -2 bind MMP2. The resulting complex is endocytosed by the low-density lipoprotein receptor-related protein (LRP), clearing MMP2 from the ECM (Yang et al. 2001).

Osteopontin (SPP1, bone sialoprotein-1) interacts with collagen and fibronectin (Mukherjee et al. 1995). It also contains several cell adhesive domains that interact with integrins and CD44.

Aggrecan is the predominant ECM proteoglycan in cartilage (Hardingham & Fosang 1992). Its relatives include versican, neurocan and brevican (Iozzo 1998). In articular cartilage the major non-fibrous macromolecules are aggrecan, hyaluronan and hyaluronan and proteoglycan link protein 1 (HAPLN1). The high negative charge density of these molecules leads to the binding of large amounts of water (Bruckner 2006). Hyaluronan is bound by several large proteoglycans proteoglycans belonging to the hyalectan family that form high-molecular weight aggregates (Roughley 2006), accounting for the turgid nature of cartilage.

The most significant enzymes in ECM remodeling are the Matrix Metalloproteinase (MMP) and A disintegrin and metalloproteinase with thrombospondin motifs (ADAMTS) families (Cawston & Young 2010). Other notable ECM degrading enzymes include plasmin and cathepsin G. Many ECM proteinases are initially present as precursors, activated by proteolytic processing. MMP precursors include an amino prodomain which masks the catalytic Zn-binding motif (Page-McCawet al. 2007). This can be removed by other proteinases, often other MMPs. ECM proteinases can be inactivated by degradation, or blocked by inhibitors. Some of these inhibitors, including alpha2-macroglobulin, alpha1-proteinase inhibitor, and alpha1-chymotrypsin can inhibit a large variety of proteinases (Woessner & Nagase 2000). The tissue inhibitors of metalloproteinases (TIMPs) are potent MMP inhibitors (Brew & Nagase 2010).


#### Edit history

| Date | Action | Author |
|---|---|---|
| 2011-08-05 | Created | Jupe S |
| 2011-09-09 | Authored | Jupe S |
| 2012-02-21 | Edited | Jupe S |
| 2012-02-28 | Reviewed | D'Eustachio P |
| 2013-05-21 | Reviewed | Venkatesan N |
| 2013-05-22 | Reviewed | Ricard-Blum S |

| Date | Action | Author |
|---|---|---|
| 2021-05-22 | Modified | Shorser S |

#### Entities found in this pathway (6)

| Input | UniProt Id | Input | UniProt Id | Input | UniProt Id |
|---|---|---|---|---|---|
| COL1A2 | P08123 | COL3A1 | P02461 | COL8A2 | P25067, Q14050 |
| ITGB4 | P16144 | LUM | P51884 | VWF | P04275 |

#### 24. Defective F8 binding to von Willebrand factor (R-HSA-9672393)

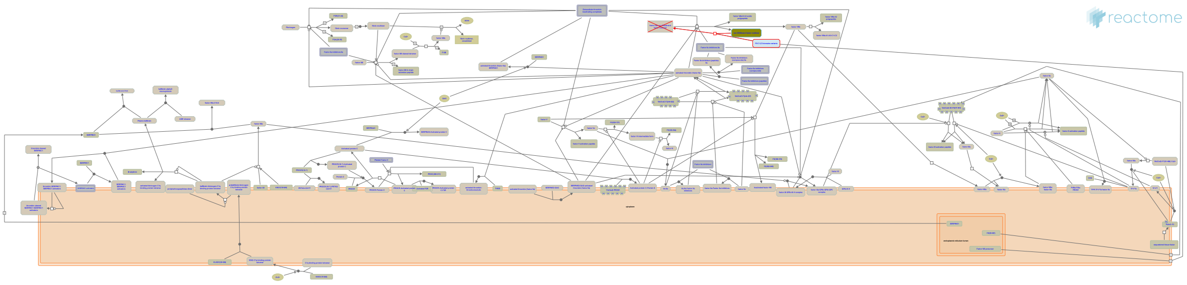

**Cellular compartments:** extracellular region.

**Diseases:** factor VIII deficiency.

Upon secretion from the cell, FVIII circulates in a tight complex with the multimeric glycoprotein von Willebrand Factor (vWF), which is essential for maintaining stable levels of FVIII in the circulation (reviewed by Pipe SW et al. 2016). Genetic mutations in the F8 gene can compromise FVIII binding to vWF thus decreasing FVIII values in the plasma causing hemophilia A (HA), an X-linked recessive bleeding disorder.


##### Edit history

| Date | Action | Author |
|---|---|---|
| 2019-09-09 | Authored | Shamovsky V |
| 2019-12-25 | Created | Shamovsky V |
| 2020-01-09 | Reviewed | D'Eustachio P |
| 2020-04-02 | Reviewed | Zhang B |
| 2020-05-26 | Edited | Shamovsky V |
| 2021-05-31 | Modified | Shorser S |

##### Entities found in this pathway (1)

| Input | UniProt Id |
|---|---|
| VWF | P04275 |

#### 25. Insertion of tail-anchored proteins into the endoplasmic reticulum membrane (R-HSA-9609523)

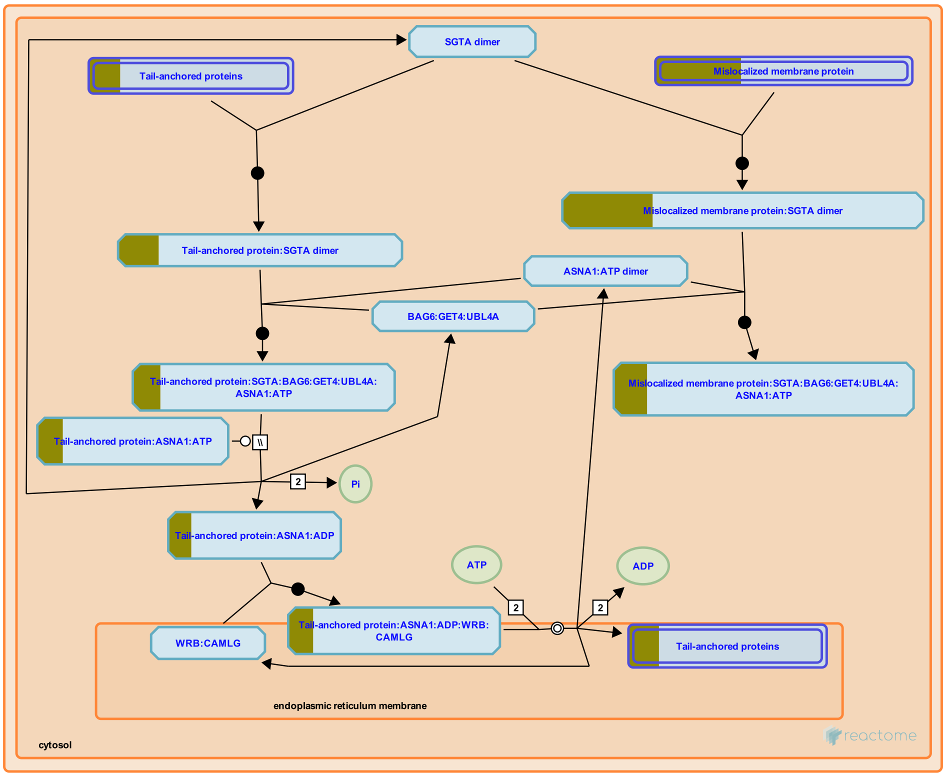


Tail-anchored (TA) proteins have a hydrophobic transmembrane domain (TMD) located near the C-terminus ("tail") of the protein. Depending on the nature of the TMD, TA proteins can be inserted into the endoplasmic reticulum (ER) membrane by at least 4 mechanisms: cotranslational insertion by the signal recognition particle (SRP), post-translational insertion by ASNA1 (TRC40), post-translational insertion by the SRP, and post-translational insertion by a SRP-independent mechanism (SND) (Casson et al. 2017, reviewed in Borgese and Fasana 2011, Casson et al. 2016, Aviram et al. 2016, Chio et al. 2017). Much of the information about the mammalian system of insertion by ASNA1 (TRC40) has been inferred from the Saccharomyces cerevisiae homologue Get3.

Prior to post-translational insertion by ASNA1, SGTA binds the transmembrane domain of the substrate TA protein immediately after translation (Leznicki et al. 2011, Leznicki and High 2012, Xu et al. 2012, Wunderly et al. 2014, Shao et al. 2017), the SGTA:TA protein complex then binds the BAG6 complex (BAG6:GET4:UBL4A) via UBL4A (Winnefeld et al. 2006, Chartron et al. 2012, Xu et al. 2012, Leznicki et al. 2013, Mock et al. 2015, Kuwabara et al. 2015, Shao et al. 2017), and the TA protein is transferred to ASNA1 (Mariappan et al. 2010, Leznicki et al. 2011, Shao et al. 2017), also bound by the BAG6 complex via UBL4A. The ASNA1:TA protein complex then docks at the WRB:CAMLG (WRB:CAML) complex located in the ER membrane and the TA protein is inserted into the ER membrane by an uncharacterized mechanism that involves ATP and the transmembrane domain insertase activity of the WRB:CAML complex (Vilardi et al. 2011, Vilardi et al. 2014, Vogl et al. 2016, and inferred from yeast in Wang et al. 2014).

Misfolded TA proteins, overexpressed TA proteins, and membrane proteins mislocalized in the cytosol bind SGTA but are not efficiently transferred to ASNA1 and, instead, are retained by BAG6 which recruits RNF126 to ubiquitinate them, targeting them for degradation by the proteasome (Wang et al. 2011, Leznicki and High 2012, Xu et al. 2012, Rodrigo-Brenni et al. 2014, Wunderly et al. 2014, Shao et al. 2017, reviewed in Lee and Ye 2013, Casson et al. 2016, Krysztofinska et al. 2016, Guna and Hegde 2018).


#### Edit history

| Date | Action | Author |
|---|---|---|
| 2018-05-28 | Edited | May B |
| 2018-05-28 | Authored | May B |
| 2018-05-29 | Created | May B |
| 2018-11-07 | Reviewed | Farkas Á, DeLaurentiis E, Schwappach B |
| 2021-05-22 | Modified | Shorser S |

#### Entities found in this pathway (2)

| Input | UniProt Id | Input | UniProt Id |
|---|---|---|---|
| OTOF | Q9HC10 | VAMP1 | P63027 |

### 6. Identifiers found

Below is a list of the input identifiers that have been found or mapped to an equivalent element in Reactome, classified by resource.

#### Entities (63)

| Input | UniProt Id | Input | UniProt Id | Input | UniProt Id |
|---|---|---|---|---|---|
| ABLIM2 | Q6H8Q1 | ACY3 | Q96HD9 | ALDH1L1 | O75891 |
| ARHGAP26 | Q9UNA1 | AZGP1 | P25311 | BDNF | P23560 |
| BTG2 | P78543 | CABP1 | Q15084 | CAMK2A | Q9UQM7 |
| CD244 | Q9BZW8 | COL1A2 | P08123 | COL3A1 | P02461 |
| COL8A2 | P25067, Q14050 | CRHBP | P24387 | CRTC1 | Q6UUV9 |
| CXADR | P78310 | DIO2 | Q92813 | EDNRB | P24530 |
| GABRA5 | P31644 | GABRQ | Q9UN88 | GPR183 | P32249 |
| GRIK1 | P39086 | GRIN3A | Q8TCU5 | GSTM3 | P21266, P46439 |
| IER3 | P46695 | IFITM2 | Q01629 | IGF1 | P05019 |
| ITGB4 | P16144 | ITIH2 | P19823 | KLHL13 | Q9P2J3, Q9P2N7 |
| LPL | P06858 | LUM | P51884 | MMD | Q9P0K1 |
| MYBPC1 | Q00872 | NCKIPSD | Q9NZQ3 | NPY | P01303 |
| NRGN | Q92686 | OGN | P20774 | OTOF | Q9HC10 |
| PGAP1 | Q75T13 | PLEKHA2 | Q9HB19 | PNOC | Q13519 |
| PPM1K | Q8N3J5 | PTGIS | Q16647 | PTPRU | Q92729 |
| RAB37 | Q96AX2 | RELN | P78509 | RND3 | P61587 |
| S100A9 | P06702 | SCG2 | O00255, P13521 | SCN4B | Q8IWT1 |
| SELPLG | Q14242 | SH2D1B | O14796 | SLC14A1 | Q13336 |
| ST6GAL2 | Q96JF0 | SUCNR1 | Q9BXA5 | TAGAP | Q8N103 |
| TGFBI | Q15582 | TLR2 | O60603 | TMOD1 | P28289 |
| VAMP1 | P23763, P63027 | VWF | P04275 | WIF1 | Q9Y5W5 |

| Input | Ensembl Id | Input | Ensembl Id | Input | Ensembl Id |
|---|---|---|---|---|---|
| BDNF | ENSG00000176697 | BTG2 | ENSG00000159388 | COL1A2 | ENSG00000164692 |
| IFITM2 | ENSG00000185201 | LPL | ENSG00000175445 | NPY | ENSG00000122585 |

### 7. Identifiers not found

These 36 identifiers were not found neither mapped to any entity in Reactome.

| | | | | | | | |
|---|---|---|---|---|---|---|---|
| ABTB1 | ANKRD2 | APOL2 | C10orf10 | C1QTNF3 | C2CD2 | CARD10 | CDKL1 |
| DHRS4L2 | FILIP1L | GDF10 | KLF10 | LUZP2 | MACROD1 | MCF2L2 | MMD2 |
| MPEG1 | MUM1L1 | NEFH | NEFM | NNAT | NRAP | OLFML3 | PAQR5 |
| PNKD | PRRT2 | RERGL | RN7SKP203 | RNVU1-18 | RP11-179B2.2 | RP11-572C15.6 | SAMD14 |
| SEMA3D | TMEM215 | TRIB2 | TRIM67 | | | | |
