## Supplementary material for "Compartment-specific total RNA profile of Hippocampal and Cortical cells from Mesial Temporal Lobe Epilepsy tissue": Other supplemental files_Reactome: Cx_mTLE non-HS_Nuc_updownspecific_report.pdf

### Pathway Analysis Report

This report contains the pathway analysis results for the submitted sample ''. Analysis was performed against Reactome version 77 on 13/09/2021. The web link to these results is:

https://reactome.org/PathwayBrowser/#/ANALYSIS=MjAyMTA5MTMxMDI4MzNfNTE4NDU%3D

Please keep in mind that analysis results are temporarily stored on our server. The storage period depends on usage of the service but is at least 7 days. As a result, please note that this URL is only valid for a limited time period and it might have expired.

### Table of Contents

1. Introduction
2. Properties
3. Genome-wide overview
4. Most significant pathways
5. Pathways details
6. Identifiers found
7. Identifiers not found

### 1. Introduction

Reactome is a curated database of pathways and reactions in human biology. Reactions can be considered as pathway 'steps'. Reactome defines a 'reaction' as any event in biology that changes the state of a biological molecule. Binding, activation, translocation, degradation and classical biochemical events involving a catalyst are all reactions. Information in the database is authored by expert biologists, entered and maintained by Reactome's team of curators and editorial staff. Reactome content frequently cross-references other resources e.g. NCBI, Ensembl, UniProt, KEGG (Gene and Compound), ChEBI, PubMed and GO. Orthologous reactions inferred from annotation for Homo sapiens are available for 17 non-human species including mouse, rat, chicken, puffer fish, worm, fly, yeast, rice, and Arabidopsis. Pathways are represented by simple diagrams following an SBGN-like format.

Reactome's annotated data describe reactions possible if all annotated proteins and small molecules were present and active simultaneously in a cell. By overlaying an experimental dataset on these annotations, a user can perform a pathway over-representation analysis. By overlaying quantitative expression data or time series, a user can visualize the extent of change in affected pathways and its progression. A binomial test is used to calculate the probability shown for each result, and the p-values are corrected for the multiple testing (Benjamini–Hochberg procedure) that arises from evaluating the submitted list of identifiers against every pathway.

To learn more about our Pathway Analysis, please have a look at our relevant publications:


Fabregat A, Sidiropoulos K, Garapati P, Gillespie M, Hausmann K, Haw R, … D'Eustachio P (2016). The reactome pathway knowledgebase. Nucleic Acids Research, 44(D1), D481–D487. https://doi.org/10.1093/nar/gkv1351.

Fabregat A, Sidiropoulos K, Viteri G, Forner O, Marin-Garcia P, Arnau V, … Hermjakob H (2017). Reactome pathway analysis: a high-performance in-memory approach. BMC Bioinformatics, 18.

#### 2. Properties

- This is an **overrepresentation** analysis: A statistical (hypergeometric distribution) test that determines whether certain Reactome pathways are over-represented (enriched) in the submitted data. It answers the question ‘Does my list contain more proteins for pathway X than would be expected by chance?’ This test produces a probability score, which is corrected for false discovery rate using the Benjamani-Hochberg method.
- 60 out of 138 identifiers in the sample were found in Reactome, where 350 pathways were hit by at least one of them.
- All non-human identifiers have been converted to their human equivalent.
- This report is filtered to show only results for species 'Homo sapiens' and resource 'all resources'.
- The unique ID for this analysis (token) is MjAyMTA5MTMxMDI4MzNfNTE4NDU%3D. This ID is valid for at least 7 days in Reactome’s server. Use it to access Reactome services with your data.

### 3. Genome-wide overview

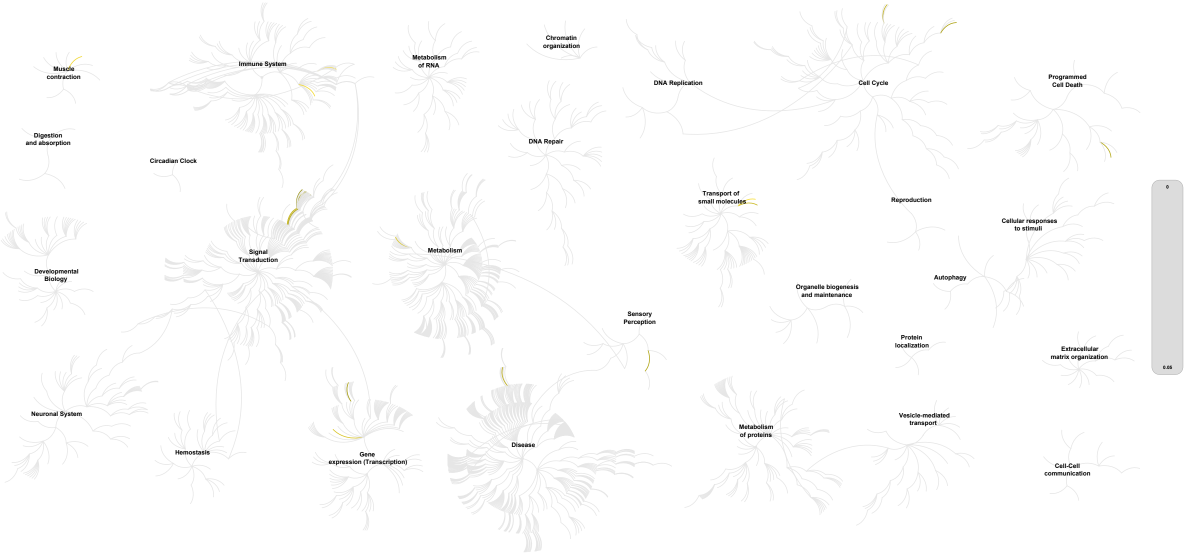


This figure shows a genome-wide overview of the results of your pathway analysis. Reactome pathways are arranged in a hierarchy. The center of each of the circular "bursts" is the root of one top-level pathway, for example "DNA Repair". Each step away from the center represents the next level lower in the pathway hierarchy. The color code denotes over-representation of that pathway in your input dataset. Light grey signifies pathways which are not significantly over-represented.

### 4. Most significant pathways

The following table shows the 25 most relevant pathways sorted by p-value.

| Pathway name | Entities | | | | Reactions | |
|---|---|---|---|---|---|---|
| | found | ratio | p-value | FDR* | found | ratio |
| Transport of glycerol from adipocytes to the liver by Aquaporins | 2 / 3 | 2.06e-04 | 5.30e-04 | 0.167 | 2 / 2 | 1.48e-04 |
| Interleukin-33 signaling | 2 / 4 | 2.75e-04 | 9.35e-04 | 0.167 | 2 / 2 | 1.48e-04 |
| Interleukin-4 and Interleukin-13 signaling | 8 / 211 | 0.015 | 0.003 | 0.298 | 21 / 47 | 0.003 |
| Phase 1 - inactivation of fast Na+ channels | 2 / 8 | 5.50e-04 | 0.004 | 0.323 | 1 / 1 | 7.40e-05 |
| FGFRL1 modulation of FGFR1 signaling | 2 / 14 | 9.63e-04 | 0.011 | 0.417 | 1 / 2 | 1.48e-04 |
| Vitamin B2 (riboflavin) metabolism | 2 / 17 | 0.001 | 0.015 | 0.417 | 2 / 5 | 3.70e-04 |
| Nuclear Receptor transcription pathway | 4 / 86 | 0.006 | 0.016 | 0.417 | 2 / 2 | 1.48e-04 |
| Passive transport by Aquaporins | 2 / 21 | 0.001 | 0.023 | 0.417 | 6 / 8 | 5.92e-04 |
| FGFR1 ligand binding and activation | 2 / 21 | 0.001 | 0.023 | 0.417 | 4 / 7 | 5.18e-04 |
| Phospholipase C-mediated cascade: FGFR1 | 2 / 22 | 0.002 | 0.025 | 0.417 | 3 / 3 | 2.22e-04 |
| Activated point mutants of FGFR2 | 2 / 23 | 0.002 | 0.027 | 0.417 | 6 / 10 | 7.40e-04 |
| Depolymerisation of the Nuclear Lamina | 2 / 23 | 0.002 | 0.027 | 0.417 | 2 / 6 | 4.44e-04 |
| Phospholipase C-mediated cascade; FGFR2 | 2 / 25 | 0.002 | 0.031 | 0.417 | 3 / 3 | 2.22e-04 |
| Breakdown of the nuclear lamina | 1 / 3 | 2.06e-04 | 0.032 | 0.417 | 1 / 3 | 2.22e-04 |
| FGFR2 ligand binding and activation | 2 / 26 | 0.002 | 0.034 | 0.417 | 5 / 5 | 3.70e-04 |
| Sensory processing of sound by outer hair cells of the cochlea | 3 / 64 | 0.004 | 0.034 | 0.417 | 1 / 8 | 5.92e-04 |
| Initiation of Nuclear Envelope (NE) Reformation | 2 / 27 | 0.002 | 0.036 | 0.417 | 1 / 7 | 5.18e-04 |
| PI-3K cascade:FGFR1 | 2 / 28 | 0.002 | 0.039 | 0.417 | 6 / 7 | 5.18e-04 |
| TP53 regulates transcription of additional cell cycle genes whose exact role in the p53 pathway remain uncertain | 2 / 28 | 0.002 | 0.039 | 0.417 | 3 / 14 | 0.001 |
| SHC-mediated cascade:FGFR1 | 2 / 30 | 0.002 | 0.044 | 0.417 | 4 / 4 | 2.96e-04 |
| FRS-mediated FGFR1 signaling | 2 / 31 | 0.002 | 0.046 | 0.417 | 9 / 9 | 6.66e-04 |
| PI-3K cascade:FGFR2 | 2 / 31 | 0.002 | 0.046 | 0.417 | 6 / 7 | 5.18e-04 |
| SHC-mediated cascade:FGFR2 | 2 / 33 | 0.002 | 0.052 | 0.417 | 4 / 4 | 2.96e-04 |

| Pathway name | Entities | | | | Reactions | |
|---|---|---|---|---|---|---|
| | found | ratio | p-value | FDR* | found | ratio |
| Sensory processing of sound by inner hair cells of the cochlea | 3 / 76 | 0.005 | 0.052 | 0.417 | 1 / 7 | 5.18e-04 |
| FRS-mediated FGFR2 signaling | 2 / 34 | 0.002 | 0.055 | 0.417 | 9 / 9 | 6.66e-04 |

* False Discovery Rate

### 5. Pathways details

For every pathway of the most significant pathways, we present its diagram, as well as a short summary, its bibliography and the list of inputs found in it.

#### 1. Transport of glycerol from adipocytes to the liver by Aquaporins (R-HSA-432030)

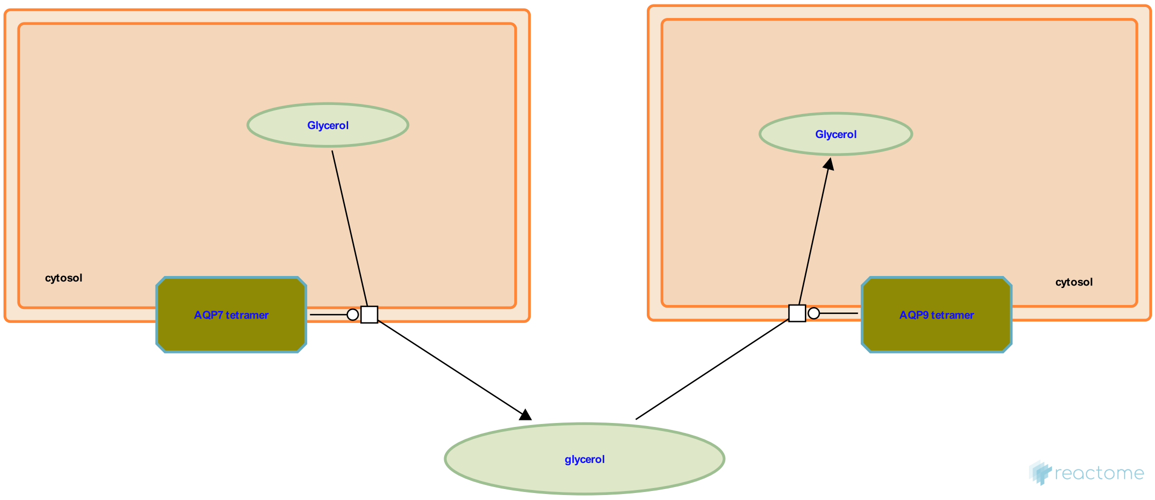


**Cellular compartments:** extracellular region, plasma membrane.

Triglycerides stored in adipocytes are hydrolyzed to yield fatty acids and glycerol. The glycerol is passively transported out of the adipocyte and into the bloodstream by Aquaporin-7 (AQP7) located in the plasma membrane of adipocytes. Glycerol in the bloodstream is passively transported into liver cells by AQP9 located in the plasma membrane of hepatocytes. Once inside the liver cell the glycerol is a substrate for gluconeogenesis.

##### Edit history

| Date | Action | Author |
|---|---|---|
| 2009-08-07 | Edited | May B |
| 2009-08-07 | Authored | May B |
| 2009-08-09 | Created | May B |
| 2010-06-24 | Reviewed | Beitz E |
| 2010-07-15 | Reviewed | Calamita G |
| 2010-07-31 | Reviewed | Mathai JC, MacIver B |
| 2021-05-22 | Modified | Shorser S |

#### Entities found in this pathway (1)

| Input | UniProt Id |
|---|---|
| AQP9 | O14520, O43315 |

#### 2. Interleukin-33 signaling (R-HSA-9014843)

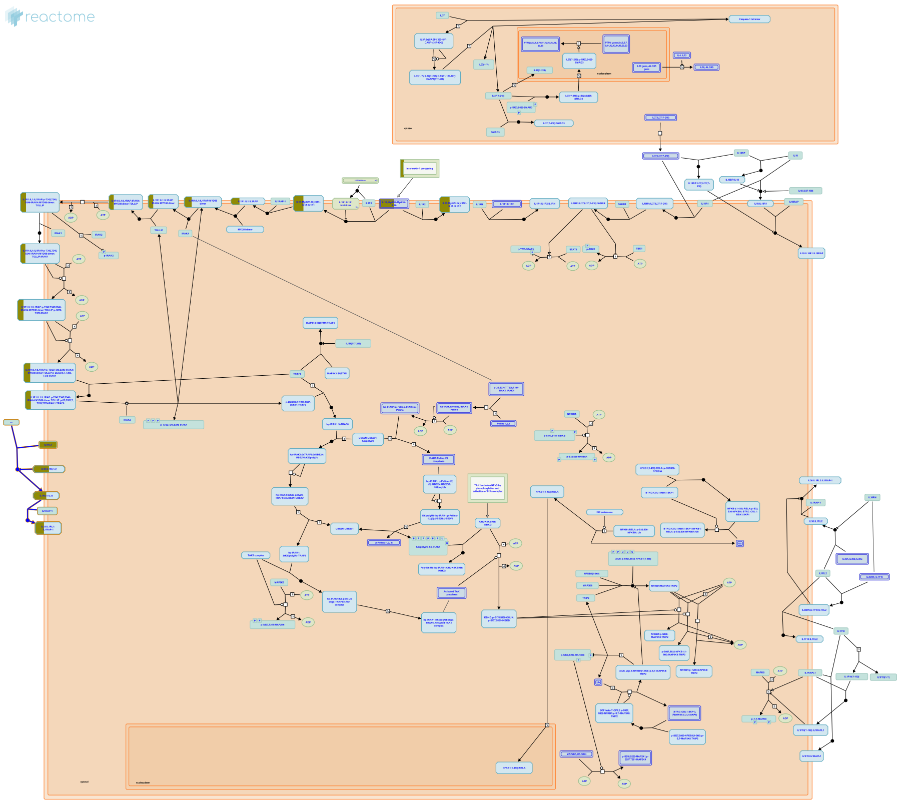

Interleukin-33 (IL33) cytokine is a member of the Interleukin-1 family. It can be classified as an alarmin because it is released into the extracellular space during cell damage. It acts as an endogenous danger signal (Liew et al. 2010).]

The gene product is biologically active (full-length IL33). Its potency has been reported to increase significantly (up to 30x) after cleavage at the N-terminus by inflammatory proteases such as Cathepsin G (CTSG) and Neutrophil elastase (ELANE) (Lefrançais et al. 2012, Lefrançais et al. 2014) but others have suggested that processing inactivates IL33 (Cayrol & Girard 2009). IL33 can act as an extracellular ligand and an intracellular signaling molecule (Martin et al. 2013, 2016). Full-length IL33 has a nuclear localization sequence and can translocate to the nucleus, where it binds heterochromatin (Moussion et al. 2008, Carriere et al. 2007, Roussel et al. 2008, Kuchler et al. 2008, Sundlisaeter et al. 2012, Baekkevold et al. 2003). IL33 that has undergone proteolytic processing is unable to translocate to the nucleus (Martin et al. 2013, Ali et al. 2010).

Binding of extracellular IL33 to its receptor Interleukin-1 receptor-like 1 (IL1RL1, suppression of tumorigenicity 2, ST2) initiates several cellular signaling pathways. Cell injury or death are the dominant mechanisms by which IL33 reaches the extracellular environment, IL33 is not actively secreted by cells (Martin et al. 2016, Kaczmarek et al. 2013, Vancamelbeke et al. 2017). Because IL33 is expressed constitutively by endothelial and epithelial cells it is immediately available to the extracellular microenvironment after cell injury and necrosis (Lefrançais et al. 2012). Increases in extracellular ATP or mechanical stress correlate with increased IL33 secretion by mast cells or cardiomyocytes, respectively (Shimokawa et al. 2017, Kakkar et al. 2012, Zhao et al. 2012, Sanada et al. 2007, Chen et al. 2015).

Soluble IL1RL1 (IL1RL1 Isoform C, ST2V) (Iwahana et al. 2005, Tominaga et al. 1999) shares the extracellular components of IL1RL1, including the ligand binding domain, but lacks the transmembrane and intracellular components of IL1RL1 (Kakkar et al. 2008, Iwahana et al. 1999). The IL33-IL1RL1 complex recruits a co-receptor, most commonly IL1 receptor accessory protein (IL1RAP, IL-1RAcP) (Schmitz et al. 2005, Lingel et al. 2009, Palmer et al. 2008, Liu et al. 2013).


##### Edit history

| Date | Action | Author |
|---|---|---|
| 2014-06-04 | Authored | Jupe S |
| 2016-01-28 | Edited | Jupe S |
| 2016-01-28 | Reviewed | Meldal BH |
| 2017-08-04 | Created | Duenas C |
| 2021-05-21 | Modified | Shorser S |

##### Entities found in this pathway (1)

| Input | UniProt Id |
| --- | --- |
| IL1RL1 | Q01638, Q01638-2 |

#### 3. Interleukin-4 and Interleukin-13 signaling (R-HSA-6785807)

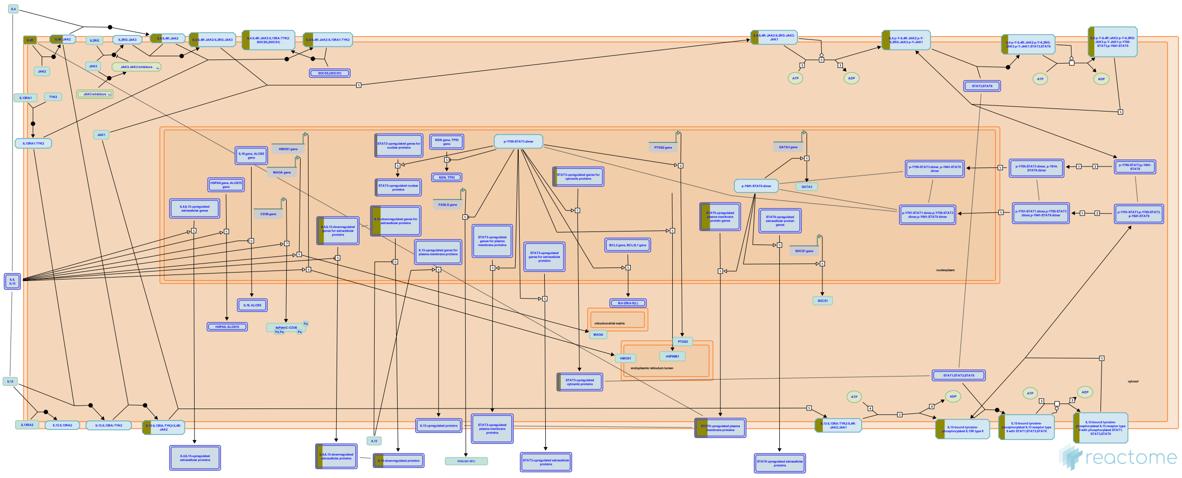

Interleukin-4 (IL4) is a principal regulatory cytokine during the immune response, crucially important in allergy and asthma (Nelms et al. 1999). When resting T cells are antigen-activated and expand in response to Interleukin-2 (IL2), they can differentiate as Type 1 (Th1) or Type 2 (Th2) T helper cells. The outcome is influenced by IL4. Th2 cells secrete IL4, which both stimulates Th2 in an autocrine fashion and acts as a potent B cell growth factor to promote humoral immunity (Nelms et al. 1999).

Interleukin-13 (IL13) is an immunoregulatory cytokine secreted predominantly by activated Th2 cells. It is a key mediator in the pathogenesis of allergic inflammation. IL13 shares many functional properties with IL4, stemming from the fact that they share a common receptor subunit. IL13 receptors are expressed on human B cells, basophils, eosinophils, mast cells, endothelial cells, fibroblasts, monocytes, macrophages, respiratory epithelial cells, and smooth muscle cells, but unlike IL4, not T cells. Thus IL13 does not appear to be important in the initial differentiation of CD4 T cells into Th2 cells, rather it is important in the effector phase of allergic inflammation (Hershey et al. 2003).

IL4 and IL13 induce “alternative activation” of macrophages, inducing an anti-inflammatory phenotype by signaling through IL4R alpha in a STAT6 dependent manner. This signaling plays an important role in the Th2 response, mediating anti-parasitic effects and aiding wound healing (Gordon & Martinez 2010, Loke et al. 2002)

There are two types of IL4 receptor complex (Andrews et al. 2006). Type I IL4R (IL4R1) is predominantly expressed on the surface of hematopoietic cells and consists of IL4R and IL2RG, the common gamma chain. Type II IL4R (IL4R2) is predominantly expressed on the surface of nonhematopoietic cells, it consists of IL4R and IL13RA1 and is also the type II receptor for IL13. (Obiri et al. 1995, Aman et al. 1996, Hilton et al. 1996, Miloux et al. 1997, Zhang et al. 1997). The second receptor for IL13 consists of IL4R and Interleukin-13 receptor alpha 2 (IL13RA2), sometimes called Interleukin-13 binding protein (IL13BP). It has a high affinity receptor for IL13 (Kd = 250 pmol/L) but is not sufficient to render cells responsive to IL13, even in the presence of IL4R (Donaldson et al. 1998). It is reported to exist in soluble form (Zhang et al. 1997) and when overexpressed reduces JAK-STAT signaling (Kawakami et al. 2001). It's function may be to prevent IL13 signalling via the functional IL4R:IL13RA1 receptor. IL13RA2 is overexpressed and enhances cell invasion in some human cancers (Joshi & Puri 2012).

The first step in the formation of IL4R1 (IL4:IL4R:IL2RB) is the binding of IL4 with IL4R (Hoffman et al. 1995, Shen et al. 1996, Hage et al. 1999). This is also the first step in formation of IL4R2 (IL4:IL4R:IL13RA1). After the initial binding of IL4 and IL4R, IL2RB binds (LaPorte et al. 2008), to form IL4R1. Alternatively, IL13RA1 binds, forming IL4R2. In contrast, the type II IL13 complex (IL13R2) forms with IL13 first binding to IL13RA1 followed by recruitment of IL4R (Wang et al. 2009).

Crystal structures of the IL4:IL4R:IL2RG, IL4:IL4R:IL13RA1 and IL13:IL4R:IL13RA1 complexes have been determined (LaPorte et al. 2008). Consistent with these structures, in monocytes IL4R is tyrosine phosphorylated in response to both IL4 and IL13 (Roy et al. 2002, Gordon & Martinez 2010) while IL13RA1 phosphorylation is induced only by IL13 (Roy et al. 2002, LaPorte et al. 2008) and IL2RG phosphorylation is induced only by IL4 (Roy et al. 2002).

Both IL4 receptor complexes signal through Jak/STAT cascades. IL4R is constitutively-associated with JAK2 (Roy et al. 2002) and associates with JAK1 following binding of IL4 (Yin et al. 1994) or IL13 (Roy et al. 2002). IL2RG constitutively associates with JAK3 (Boussiotis et al. 1994, Russell et al. 1994). IL13RA1 constitutively associates with TYK2 (Umeshita-Suyama et al. 2000, Roy et al. 2002, LaPorte et al. 2008, Bhattacharjee et al. 2013).

IL4 binding to IL4R1 leads to phosphorylation of JAK1 (but not JAK2) and STAT6 activation (Takeda et al. 1994, Ratthe et al. 2007, Bhattacharjee et al. 2013).

IL13 binding increases activating tyrosine-99 phosphorylation of IL13RA1 but not that of IL2RG. IL4 binding to IL2RG leads to its tyrosine phosphorylation (Roy et al. 2002). IL13 binding to IL4R2 leads to TYK2 and JAK2 (but not JAK1) phosphorylation (Roy & Cathcart 1998, Roy et al. 2002).

Phosphorylated TYK2 binds and phosphorylates STAT6 and possibly STAT1 (Bhattacharjee et al. 2013).

A second mechanism of signal transduction activated by IL4 and IL13 leads to the insulin receptor substrate (IRS) family (Kelly-Welch et al. 2003). IL4R1 associates with insulin receptor substrate 2 and activates the PI3K/Akt and Ras/MEK/Erk pathways involved in cell proliferation, survival and translational control. IL4R2 does not associate with insulin receptor substrate 2 and consequently the PI3K/Akt and Ras/MEK/Erk pathways are not activated (Busch-Dienstfertig & González-Rodríguez 2013).

#### Edit history

| Date | Action | Author |
|---|---|---|
| 2015-07-01 | Authored | Jupe S |
| 2015-07-01 | Created | Jupe S |
| 2016-09-02 | Edited | Jupe S |
| 2016-09-02 | Reviewed | Leibovich SJ |

| Date | Action | Author |
|---|---|---|
| 2021-05-31 | Modified | Shorser S |

#### Entities found in this pathway (4)

| Input | UniProt Id |
|---|---|
| CEBPD | P49716 |
| IL4R | P24394 |

| Input | UniProt Id |
|---|---|
| IL1A | P01583 |
| MCL1 | Q07820 |

| Input | Ensembl Id |
|---|---|
| CEBPD | ENSG00000221869 |
| IL4R | ENSG00000077238 |

| Input | Ensembl Id |
|---|---|
| IL1A | ENSG00000115008 |
| MCL1 | ENSG00000143384 |

#### 4. Phase 1 - inactivation of fast Na+ channels (R-HSA-5576894)

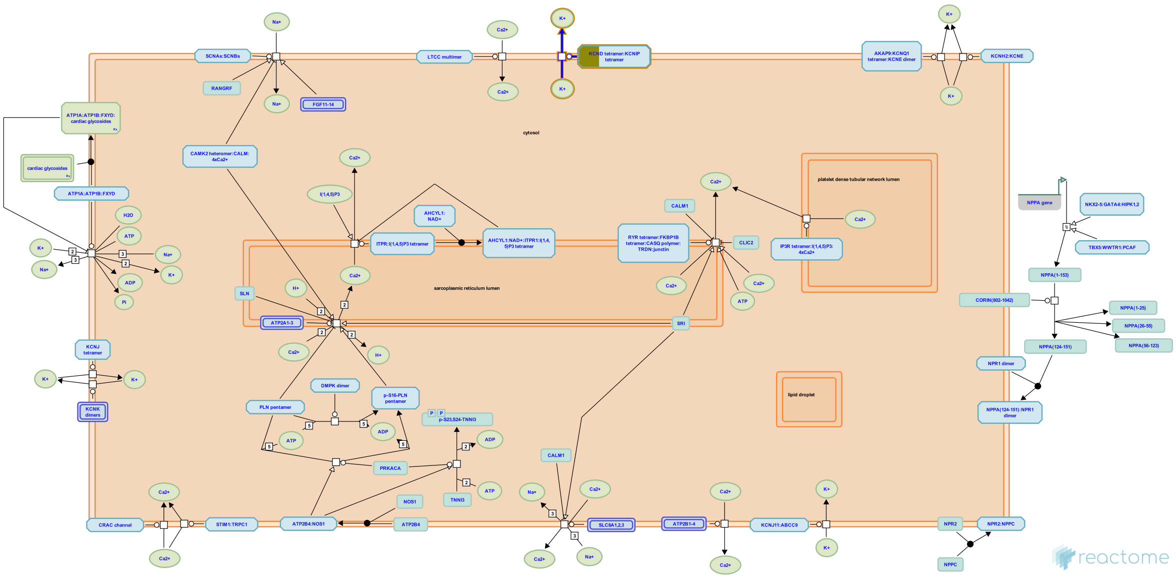

Phase 1 of the cardiac action potential is the inactivation of the fast Na+ channels. The transient net outward current causing the small downward deflection (the "notch" of the action potetial) is due to the movement of K+ and Cl- ions. In pacemaker cells, this phase is due to rapid K+ efflux and closure of L-type Ca2+ channels (Park & Fishman 2011, Grant 2009).


##### Edit history

| Date | Action | Author |
|---|---|---|
| 2014-05-27 | Edited | Jassal B |
| 2014-05-27 | Authored | Jassal B |
| 2014-05-27 | Created | Jassal B |
| 2015-11-09 | Reviewed | Colotti G |
| 2021-05-22 | Modified | Shorser S |

##### Entities found in this pathway (2)

| Input | UniProt Id | Input | UniProt Id |
|---|---|---|---|
| KCND3 | Q9UK17 | KCNIP2 | Q9NS61 |

#### 5. FGFRL1 modulation of FGFR1 signaling (R-HSA-5658623)

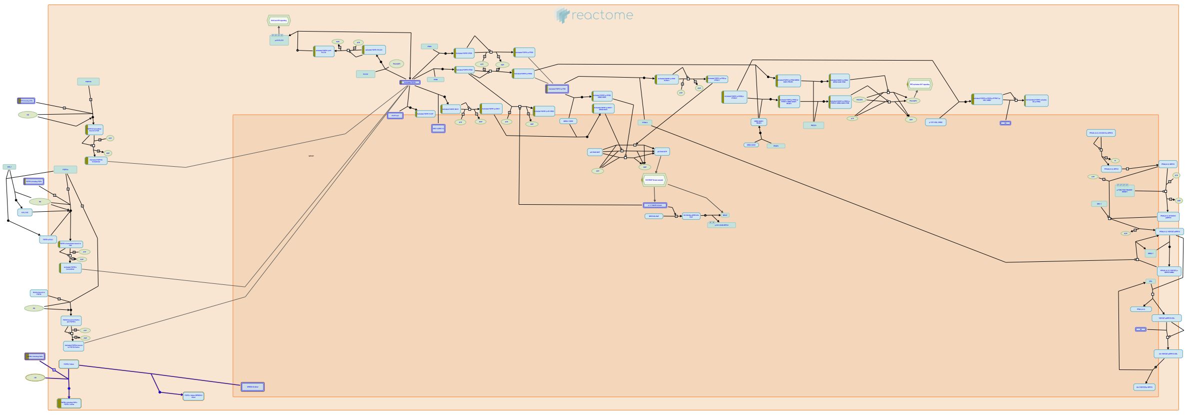

FGFRL1 is a fifth member of the FGFR family of receptors. The extracellular region has 40% sequence similarity with FGFR1-4, but FGFRL1 lacks the internal kinase domain of the other FGF receptors and how it acts in FGFR signaling is unclear. Some models suggest FGFRL1 restricts canonical FGFR signaling by sequestering ligand away from kinase-active receptors, while other models suggest that FGFRL1 may promote canonical signaling by nucleating signaling complexes or enhancing ERK1/2 activation (reviewed in Trueb, 2011; Trueb et al, 2013).


##### Edit history

| Date | Action | Author |
|---|---|---|
| 2014-12-12 | Authored | Rothfels K |
| 2014-12-20 | Created | Rothfels K |
| 2015-01-14 | Edited | Rothfels K |
| 2016-01-06 | Reviewed | Nishimura T, Grose RP |
| 2016-03-18 | Reviewed | Gotoh N |
| 2021-05-22 | Modified | Shorser S |

##### Entities found in this pathway (2)

| Input | UniProt Id | Input | UniProt Id |
|---|---|---|---|
| FGF10 | O15520 | FGF17 | O60258-1 |

#### 6. Vitamin B2 (riboflavin) metabolism (R-HSA-196843)

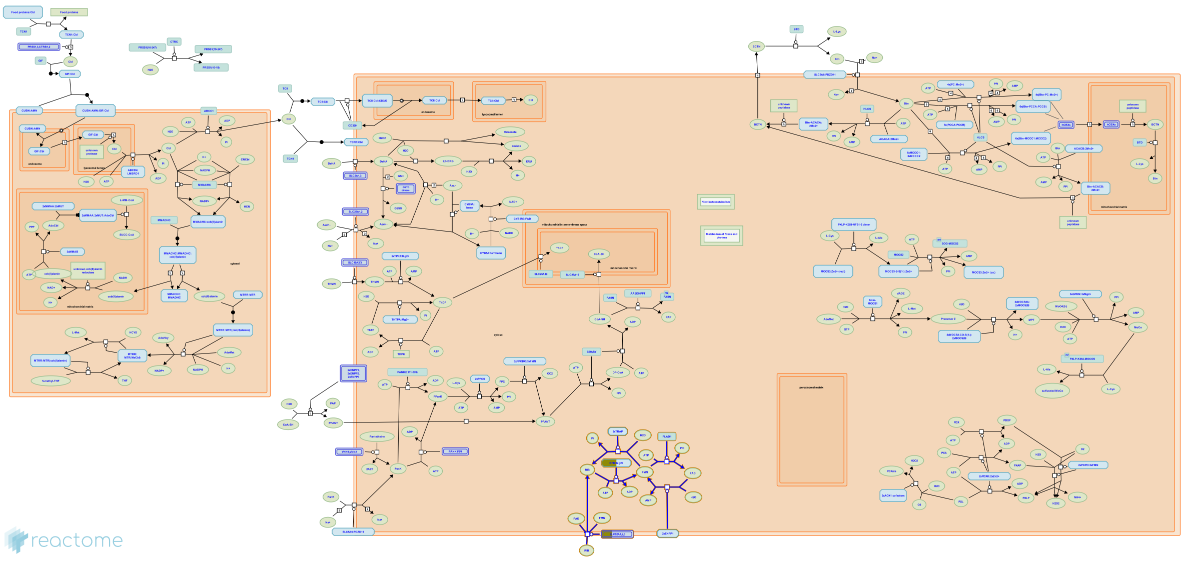

Riboflavin (vitamin B2, E101) is an essential component for the cofactors FAD (flavin-adenine dinucleotide) and FMN (flavin mononucleotide). Together with NAD+ and NADP+, FAD and FMN are important hydrogen carriers and take part in more than 100 redox reactions involved in energy metabolism. Riboflavin is present in many vegetables and meat and during digestion, various flavoproteins from food are degraded and riboflavin is resorbed. The major degradation and excretion product in humans is riboflavin (Rivlin 1970).


##### Edit history

| Date | Action | Author |
|---|---|---|
| 2007-04-24 | Created | Jassal B |
| 2021-05-22 | Modified | Shorser S |

##### Entities found in this pathway (2)

| Input | UniProt Id | Input | UniProt Id |
|---|---|---|---|
| FMN1 | Q969G6 | SLC52A3 | Q9NQ40 |

#### 7. Nuclear Receptor transcription pathway (R-HSA-383280)

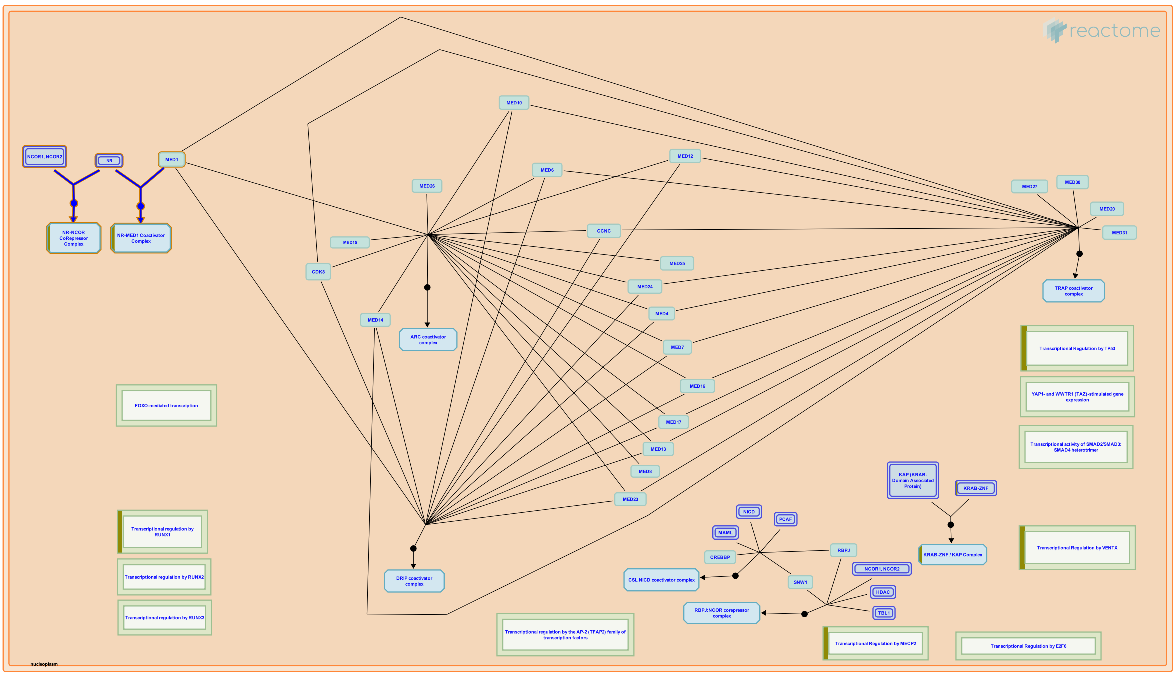


A classic example of bifunctional transcription factors is the family of Nuclear Receptor (NR) proteins. These are DNA-binding transcription factors that bind certain hormones, vitamins, and other small, diffusible signaling molecules. The non-liganded NRs recruit specific corepressor complexes of the NCOR/SMRT type, to mediate transcriptional repression of the target genes to which they are bound. During signaling, ligand binding to a specific domain the NR proteins induces a conformational change that results in the exchange of the associated CoR complex, and its replacement by a specific coactivator complex of the TRAP / DRIP / Mediator type. These coactivator complexes typically nucleate around a MED1 coactivator protein that is directly bound to the NR transcription factor.

A general feature of the 49 human NR proteins is that in the unliganded state, they each bind directly to an NCOR corepressor protein, either NCOR1 or NCOR2 (NCOR2 was previously named "SMRT"). This NCOR protein nucleates the assembly of additional, specific corepressor proteins, depending on the cell and DNA context. The NR-NCOR interaction is mediated by a specific protein interaction domain (PID) present in the NRs that binds to specific cognate PID(s) present in the NCOR proteins. Thus, the human NRs each take part in an NR-NCOR binding reaction in the absence of binding by their ligand.

A second general feature of the NR proteins is that they each contain an additional, but different PID that mediates specific binding interactions with MED1 proteins. In the ligand-bound state, NRs each take part in an NR-MED1 binding reaction to form an NR-MED1 complex. The bound MED1 then functions to nucleate the assembly of additional specific coactivator proteins, depending on the cell and DNA context, such as what specific target gene promoter they are bound to, and in what cell type.

The formation of specific MED1-containing coactivator complexes on specific NR proteins has been well-characterized for a number of the human NR proteins (see Table 1 in (Bourbon, 2004)). For example, binding of thyroid hormone (TH) to the human TH Receptor (THRA or THRB) was found to result in the recruitment of a specific complex of Thyroid Receptor Associated Proteins - the TRAP coactivator complex - of which the TRAP220 subunit was later identified to be the Mediator 1 (MED1) homologue.

Similarly, binding of Vitamin D to the human Vitamin D3 Receptor was found to result in the recruitment of a specific complex of D Receptor Interacting Proteins - the DRIP coactivator complex, of which the DRIP205 subunit was later identified to be human MED1.

##### References

##### Edit history

| Date | Action | Author |
|---|---|---|
| 2008-11-20 | Authored | Caudy M |
| 2008-12-03 | Created | Caudy M |
| 2009-05-27 | Edited | Caudy M |
| 2009-08-29 | Reviewed | Freedman LP |
| 2021-05-31 | Modified | Shorser S |

##### Entities found in this pathway (3)

| Input | UniProt Id | Input | UniProt Id | Input | UniProt Id |
|---|---|---|---|---|---|
| NR4A1 | P22736 | NR4A2 | P43354 | NR4A3 | Q92570-1, Q92570-2 |

#### 8. Passive transport by Aquaporins (R-HSA-432047)

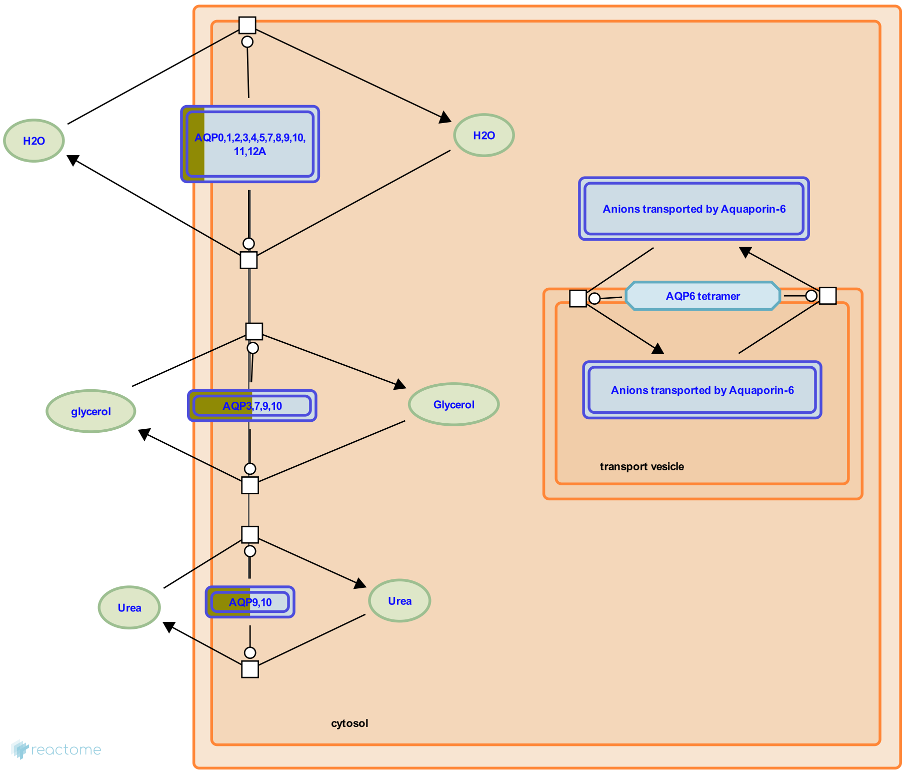


**Cellular compartments:** plasma membrane, transport vesicle membrane.

Aquaporins (AQP's) are six-pass transmembrane proteins that form channels in membranes. Each monomer contains a central channel formed in part by two asparagine-proline-alanine motifs (NPA boxes) that confer selectivity for water and/or solutes. The monomers assemble into tetramers. During passive transport by Aquaporins most aquaporins (i.e. AQP0/MIP, AQP1, AQP2, AQP3, AQP4, AQP5, AQP7, AQP8, AQP9, AQP10) transport water into and out of cells according to the osmotic gradient across the membrane. Four aquaporins (the aquaglyceroporins AQP3, AQP7, AQP9, AQP10) conduct glycerol, three aquaporins (AQP7, AQP9, AQP10) conduct urea, and one aquaporin (AQP6) conducts anions, especially nitrate. AQP8 also conducts ammonia in addition to water.

AQP11 and AQP12, classified as group III aquaporins, were identified as a result of the genome sequencing project and are characterized by having variations in the first NPA box when compared to more traditional aquaporins. Additionally, a conserved cysteine residue is present about 9 amino acids downstream from the second NPA box and this cysteine is considered indicative of group III aquaporins. Purified AQP11 incorporated into liposomes showed water transport. Knockout mice lacking AQP11 had fatal cyst formation in the proximal tubule of the kidney. Exogenously expressed AQP12 showed intracellular localization. AQP12 is expressed exclusively in pancreatic acinar cells.

Aquaporins are important in fluid and solute transport in various tissues. During Transport of glycerol from adipocytes to the liver by Aquaporins, glycerol generated by triglyceride hydrolysis is exported from adipocytes by AQP7 and is imported into liver cells via AQP9. AQP1 plays a role in forming cerebrospinal fluid and AQP1, AQP4, and AQP9 appear to be important in maintaining fluid balance in the brain. AQP0, AQP1, AQP3, AQP4, AQP8, AQP9, and AQP11 play roles in the physiology of the hepatobiliary tract.

In the kidney, water and solutes are passed out of the bloodstream and into the proximal tubule via the slit-like structure formed by nephrin in the glomerulus. Water is reabsorbed from the filtrate during its transit through the proximal tubule, the descending loop of Henle, the distal convoluted tubule, and the collecting duct. Aquaporin-1 (AQP1) in the proximal tubule and the descending thin limb of Henle is responsible for about 90% of reabsorption (as estimated from mouse knockouts of AQP1). AQP1 is located on both the apical and basolateral surface of epithelial cells and thus transports water through the epithelium and back into the bloodstream. In the collecting duct epithelial cells have AQP2 on their apical surfaces and AQP3 and AQP4 on their basolateral surfaces to transport water across the epithelium. Vasopressin regulates renal water homeostasis via Aquaporins by regulating the permeability of the epithelium through activation of a signaling cascade leading to the phosphorylation of AQP2 and its translocation from intracellular vesicles to the apical membrane of collecting duct cells.

Here, three views of aquaporin-mediated transport have been annotated: a generic view of transport mediated by the various families of aquaporins independent of tissue type (Passive transport by Aquaporins), a view of the role of specific aquaporins in maintenance of renal water balance (Vasopressin regulates renal water homeostasis via Aquaporins), and a view of the role of specific aquaporins in glycerol transport from adipocytes to the liver (Transport of glycerol from adipocytes to the liver by Aquaporins).


#### Edit history

| Date | Action | Author |
|---|---|---|
| 2009-08-07 | Edited | May B |
| 2009-08-07 | Authored | May B |
| 2009-08-09 | Created | May B |
| 2010-06-24 | Reviewed | Beitz E |
| 2010-07-15 | Reviewed | Calamita G |
| 2010-07-31 | Reviewed | Mathai JC, MacIver B |
| 2021-05-22 | Modified | Shorser S |

#### Entities found in this pathway (1)

| Input | UniProt Id |
|---|---|
| AQP9 | O14520, O43315 |

#### 9. FGFR1 ligand binding and activation (R-HSA-190242)

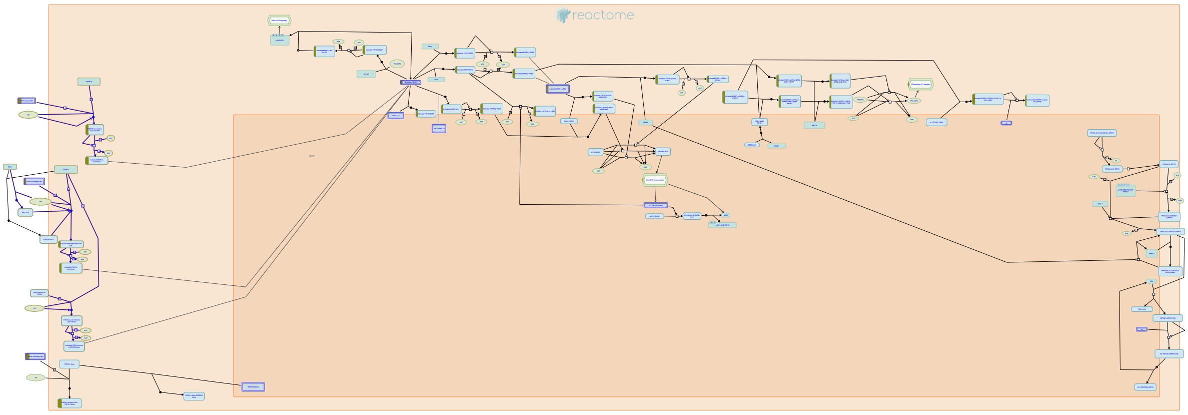

The vertebrate fibroblast growth factor receptor 1 (FGFR1) is alternatively spliced generating multiple variants that are differentially expressed during embryo development and in the adult body. The restricted expression patterns of FGFR1 isoforms, together with differential expression and binding of specific ligands, leads to activation of common FGFR1 signal transduction pathways, but may result in distinctively different biological responses as a result of differences in cellular context. FGFR1 isoforms are also present in the nucleus in complex with various fibroblast growth factors where they function to regulate transcription of target genes.

FGFR is probably activated by NCAM very differently from the way by which it is activated by FGFs, reflecting the different conditions for NCAM-FGFR and FGF-FGFR interactions. The affinity of FGF for FGFR is approximately 10e6 times higher than that of NCAM for FGFR. Moreover, in the brain NCAM is constantly present on the cell surface at a much higher (micromolar) concentration than FGFs, which only appear transiently in the extracellular environment in the nanomolar range.


##### Edit history

| Date | Action | Author |
|---|---|---|
| 2006-12-18 | Created | de Bono B |
| 2007-01-10 | Authored | de Bono B |
| 2007-02-07 | Reviewed | Mohammadi M |
| 2007-02-11 | Edited | D'Eustachio P, de Bono B |
| 2016-01-06 | Reviewed | Grose RP |

| Date | Action | Author |
|---|---|---|
| 2021-05-22 | Modified | Shorser S |

#### Entities found in this pathway (2)

| Input | UniProt Id | Input | UniProt Id |
|---|---|---|---|
| FGF10 | O15520 | FGF17 | O60258-1 |

#### 10. Phospholipase C-mediated cascade: FGFR1 (R-HSA-5654219)

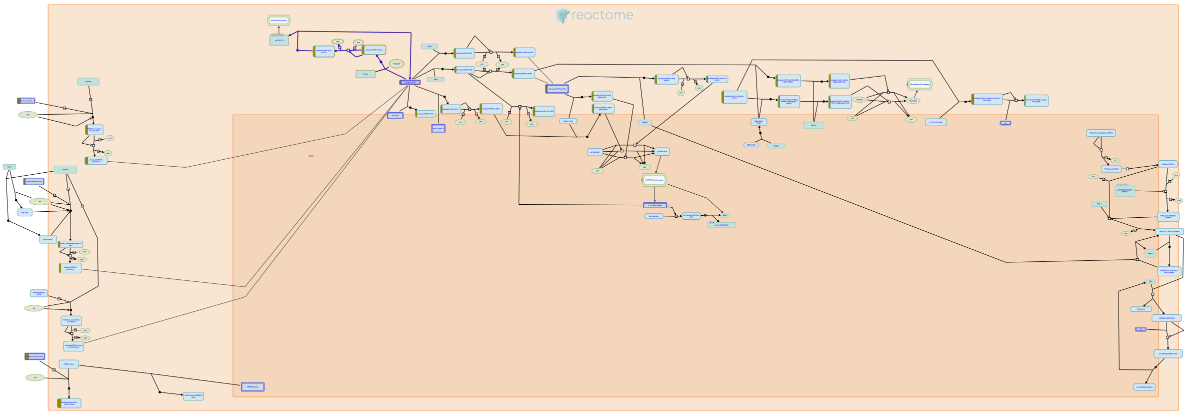

**Cellular compartments:** plasma membrane.

Phospholipase C-gamma (PLC-gamma) is a substrate of the fibroblast growth factor receptor (FGFR) and other receptors with tyrosine kinase activity. It is known that the src homology region 2 (SH2 domain) of PLC-gamma and of other signaling molecules (such as GTPase-activating protein and phosphatidylinositol 3-kinase-associated p85) direct their binding toward autophosphorylated tyrosine residues of the FGFR. Recruitment of PLC-gamma results in its phosphorylation and activation by the receptor.  Activated PLC-gamma hydrolyzes phosphatidyl inositol[4,5] P2 to form the second messengers diacylglycerol (DAG) and Ins [1,4,5]P3, which stimulate calcium release and activation of calcium/calmodulin dependent kinases.


##### Edit history

| Date | Action | Author |
|---|---|---|
| 2007-01-10 | Authored | de Bono B |
| 2007-02-07 | Reviewed | Mohammadi M |
| 2010-02-03 | Edited | Jupe S |
| 2014-12-04 | Created | Rothfels K |
| 2021-05-22 | Modified | Shorser S |

##### Entities found in this pathway (2)

| Input | UniProt Id | Input | UniProt Id |
|---|---|---|---|
| FGF10 | O15520 | FGF17 | O60258-1 |

#### 11. Activated point mutants of FGFR2 (R-HSA-2033519)

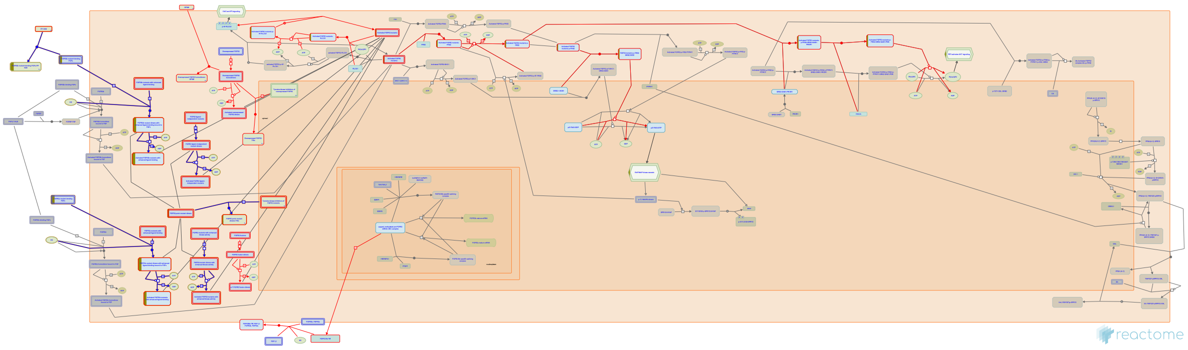

**Cellular compartments:** plasma membrane, cytosol, extracellular region.

**Diseases:** cancer, bone development disease.

Autosomal dominant mutations in FGFR2 are associated with the development of a range of skeletal disorders including Beare-Stevensen cutis gyrata syndrome, Pfeiffer syndrome, Jackson-Weiss syndrome, Crouzon syndrome and Apert Syndrome (reveiwed in Burke, 1998; Webster and Donoghue 1997; Cunningham, 2007). Mutations that give rise to Crouzon, Jackson-Weiss and Pfeiffer syndromes tend to cluster in the third Ig-like domain of the receptor, either in exon IIIa (shared by the IIIb and the IIIc isoforms) or in the FGFR2c-specific exon IIIc. These mutations frequently involve creation or removal of a cysteine residue, leading to the formation of an unpaired cysteine residue that is thought to promote intramolecular dimerization and thus constitutive, ligand-independent activation (reviewed in Burke, 1998; Webster and Donoghue, 1997; Cunningham, 2007). Mutations in FGFR2 that give rise to Apert Syndrome cluster to the highly conserved Pro-Ser dipeptide in the IgII-Ig III linker; mutations in the paralogous residues of FGFR1 and 3 give rise to Pfeiffer and Muenke syndromes, respectively (Muenke, 1994; Wilkie, 1995; Bellus, 1996). Development of Beare-Stevensen cutis gyrata is associated with mutations in the transmembrane-proximal region of the receptor (Przylepa, 1996), and similar mutations in FGFR3 are linked to the development of thanatophoric dysplasia I (Tavormina, 1995a). These mutations all affect FGFR2 signaling without altering the intrinsic kinase activity of the receptor.

Activating point mutations have also been identified in FGFR2 in ~15% of endometrial cancers, as well as to a lesser extent in ovarian and gastric cancers (Dutt, 2008; Pollock, 2007; Byron, 2010; Jang, 2001). These mutations are found largely in the extracellular region and in the kinase domain of the receptor, and parallel activating mutations seen in autosomal dominant disorders described above.

Activating mutations in FGFR2 are thought to contribute to receptor activation through diverse mechanisms, including constitutive ligand-independent dimerization (Robertson, 1998), expanded range and affinity for ligand (Ibrahimi, 2004b; Yu, 2000) and enhanced kinase activity (Byron, 2008; Chen, 2007).

#### Edit history

| Date | Action | Author |
|---|---|---|
| 2012-01-09 | Created | Rothfels K |
| 2012-02-10 | Authored | Rothfels K |
| 2012-05-15 | Reviewed | Ezzat S |
| 2012-05-16 | Edited | Rothfels K |
| 2012-05-26 | Modified | Rothfels K |

#### Entities found in this pathway (2)

| Input | UniProt Id | Input | UniProt Id |
|---|---|---|---|
| FGF10 | O15520 | FGF17 | O60258-1 |

#### 12. Depolymerisation of the Nuclear Lamina (R-HSA-4419969)

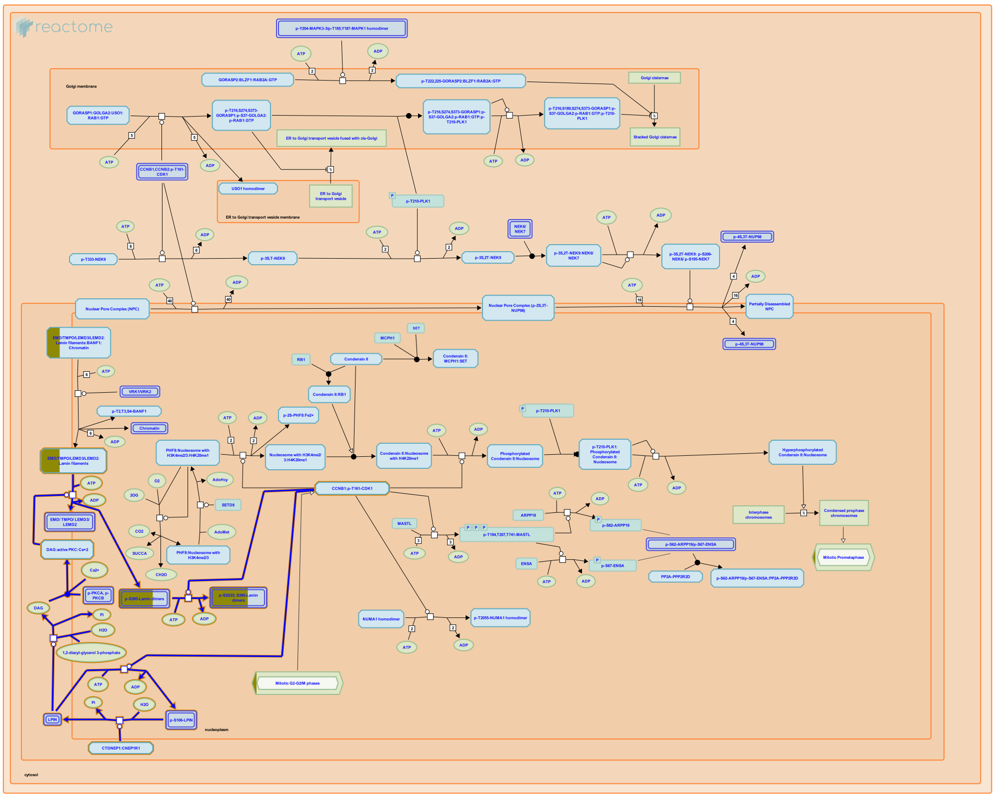

The nuclear envelope breakdown in mitotic prophase involves depolymerisation of lamin filaments, the main constituents of the nuclear lamina. The nuclear lamina is located at the nuclear face of the inner nuclear membrane and plays and important role in the structure and function of the nuclear envelope (reviewed by Burke and Stewart 2012). Depolymerisation of lamin filaments, which consist of lamin homodimers associated through electrostatic interactions in head-to-tail molecular strings, is triggered by phosphorylation of lamins. While CDK1 phosphorylates the N-termini of lamins (Heald and McKeon 1990, Peter et al. 1990, Ward and Kirschner 1990, Mall et al. 2012), PKCs (PRKCA and PRKCB) phosphorylate the C-termini of lamins (Hocevar et al. 1993, Goss et al. 1994, Mall et al. 2012). PKCs are activated by lipid-mediated signaling, where lipins, activated by CTDNEP1:CNEP1R1 serine/threonine protein phosphatase complex, catalyze the formation of DAG (Gorjanacz et al. 2009, Golden et al. 2009, Wu et al. 2011, Han et al. 2012, Mall et al. 2012).

#### Edit history

| Date | Action | Author |
|---|---|---|
| 2013-08-28 | Created | Orlic-Milacic M |
| 2014-02-02 | Edited | Orlic-Milacic M |
| 2014-02-02 | Authored | Orlic-Milacic M |
| 2014-02-10 | Reviewed | Gorjánácz M |
| 2021-05-22 | Modified | Shorser S |

#### Entities found in this pathway (1)

| Input | UniProt Id |
|---|---|
| LMNA | P02545-1, P02545-2 |

#### 13. Phospholipase C-mediated cascade; FGFR2 (R-HSA-5654221)

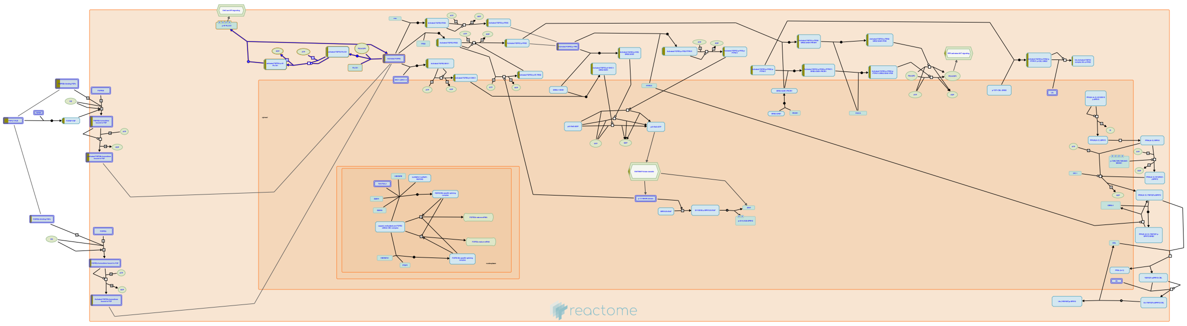

**Cellular compartments:** plasma membrane.

Phospholipase C-gamma (PLC-gamma) is a substrate of the fibroblast growth factor receptor (FGFR) and other receptors with tyrosine kinase activity. It is known that the src homology region 2 (SH2 domain) of PLC-gamma and of other signaling molecules (such as GTPase-activating protein and phosphatidylinositol 3-kinase-associated p85) direct their binding toward autophosphorylated tyrosine residues of the FGFR. Recruitment of PLC-gamma results in its phosphorylation and activation by the receptor. Activated PLC-gamma hydrolyzes phosphatidyl inositol[4,5] P2 to form the second messengers diacylglycerol (DAG) and Ins [1,4,5]P3, which stimulate calcium release and activation of calcium/calmodulin dependent kinases.


##### Edit history

| Date | Action | Author |
| --- | --- | --- |
| 2007-01-10 | Authored | de Bono B |
| 2007-02-07 | Reviewed | Mohammadi M |
| 2010-02-03 | Edited | Jupe S |
| 2014-12-04 | Created | Rothfels K |
| 2021-05-21 | Modified | Shorser S |

##### Entities found in this pathway (2)

| Input | UniProt Id | Input | UniProt Id |
| --- | --- | --- | --- |
| FGF10 | O15520 | FGF17 | O60258-1 |

#### 14. Breakdown of the nuclear lamina (R-HSA-352238)

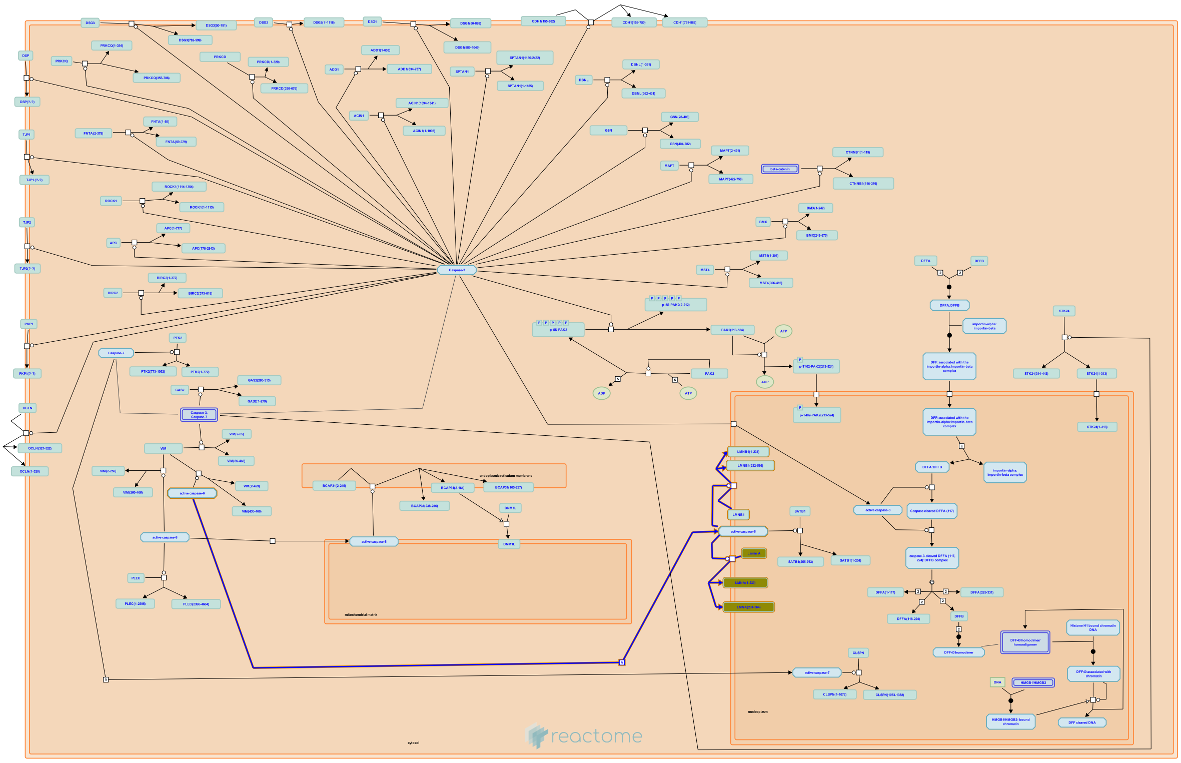

**Cellular compartments:** nuclear envelope.

Activated caspases cleave  nuclear lamins causing the irreversible breakdown of the nuclear lamina.


##### Edit history

| Date | Action | Author |
|---|---|---|
| 2008-05-18 | Authored | Schulze-Osthoff K |
| 2008-05-20 | Edited | Matthews L |
| 2008-06-02 | Created | Matthews L |
| 2008-06-11 | Reviewed | Ranganathan S |
| 2021-05-31 | Modified | Shorser S |

##### Entities found in this pathway (1)

| Input | UniProt Id |
|---|---|
| LMNA | P02545-1 |

#### 15. FGFR2 ligand binding and activation (R-HSA-190241)

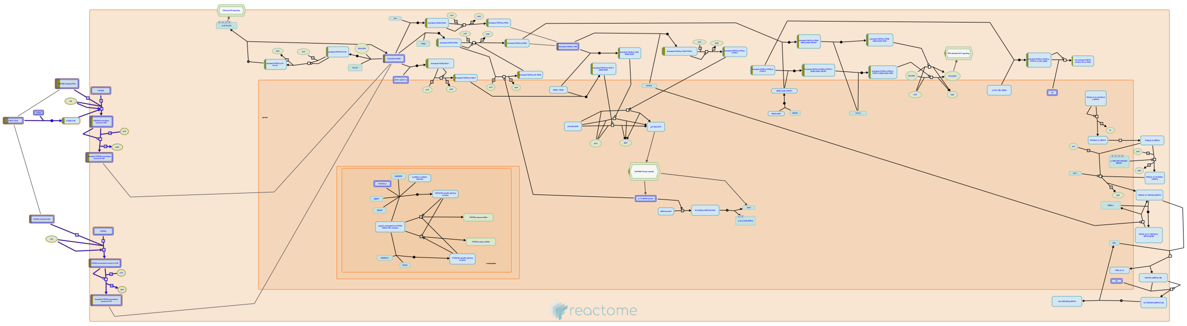

Dominant mutations in the fibroblast growth factor receptor 2 (FGFR2) gene have been identified as causes of four phenotypically distinct craniosynostosis syndromes, including Crouzon, Jackson-Weiss, Pfeiffer, and Apert syndromes. FGFR2 binds a number of different FGFs preferentially, as illustrated in this pathway.

FGFR is probably activated by NCAM very differently from the way by which it is activated by FGFs, reflecting the different conditions for NCAM-FGFR and FGF-FGFR interactions. The affinity of FGF for FGFR is approximately 10e6 times higher than that of NCAM for FGFR. Moreover, in the brain NCAM is constantly present on the cell surface at a much higher (micromolar) concentration than FGFs, which only appear transiently in the extracellular environment in the nanomolar range.


##### Edit history

| Date | Action | Author |
|---|---|---|
| 2006-12-18 | Created | de Bono B |
| 2007-01-10 | Authored | de Bono B |
| 2007-02-07 | Reviewed | Mohammadi M |
| 2007-02-11 | Edited | D'Eustachio P, de Bono B |
| 2021-05-21 | Modified | Shorser S |

##### Entities found in this pathway (2)

| Input | UniProt Id | Input | UniProt Id |
|---|---|---|---|
| FGF10 | O15520 | FGF17 | O60258-1 |

#### 16. Sensory processing of sound by outer hair cells of the cochlea (R-HSA-9662361)

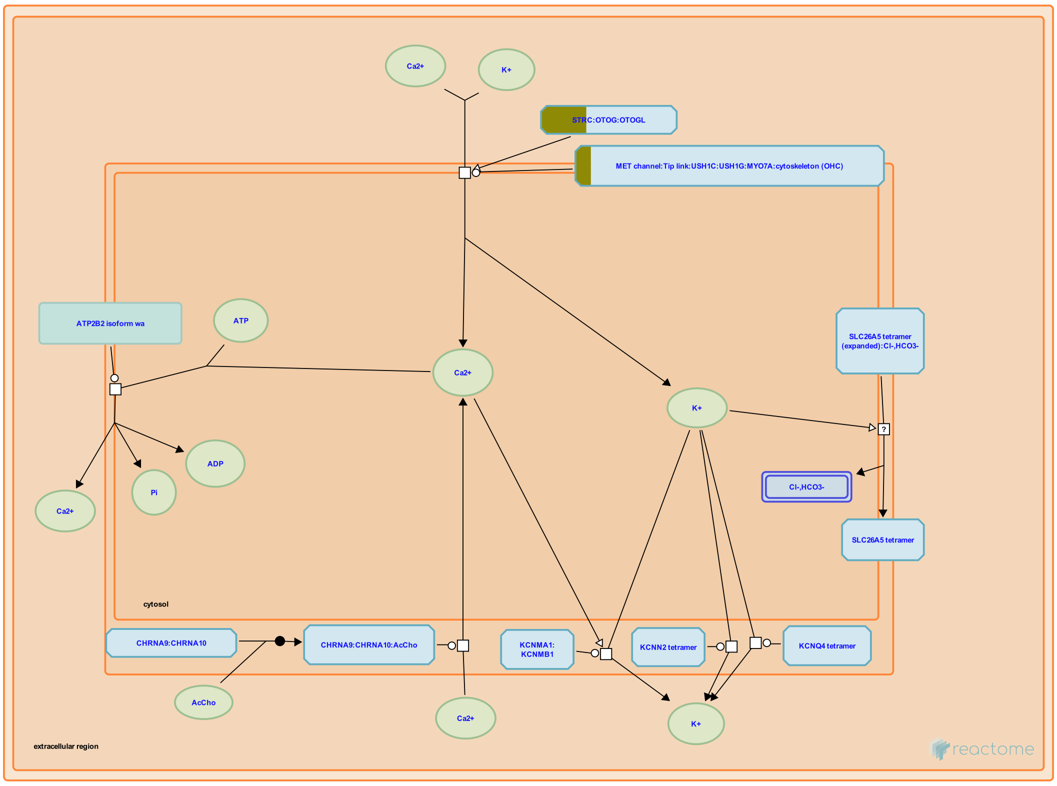


Outer hair cells (OHCs) produce amplification of sound waves in the cochlea by shortening and lengthening in response to sound, a phenomenon called electromotility (reviewed in Kim and Fettiplace 2014, Fettiplace 2016, Fettiplace 2017, Fritzsch et al. 2017, Ashmore 2019). Like inner hair cells, OHCs possess apical stereocilia arranged in rows of ascending height. A taller stereocilium is connected to a shorter stereocilium by a tip link comprising a CDH23 dimer onthe side of the taller stereocilium and a PCDH15 dimer on theapex of the shorter stereocilium. PCDH15 interacts with LHFPL5, a subunit of the mechanoelectrical transduction channel complex (MET channel, also called the mechanotransduction channel), which contains TMC1 or TMC2, TMIE, CIB2, and LHFPL5 (reviewed in Fettiplace 2016). Deflection of the stereocilia in one direction produces tension on the tip link that increases the open probability of the MET channel, resulting in depolarization of the OHC. Deflection of the stereocilia in the opposite direction produces compression on the tip link that decreases the the open probability of the MET channel, resulting in hyperpolarization of the OHC.

Sound causes micromechanical motions of the organ of Corti that result in alternating tension and compression in the tip link that produce excitatory-inhibitory cycles of MET channel openings and closings relative to the MET channel's resting open probability. This causes directionally alternating fluxes of K+ and Ca2+, yielding depolarization-hyperpolarization cycles that cause conformational changes in prestin (SLC26A5). These cycles are asymmetrical, with contraction caused by depolarization dominating elongation caused by hyperpolarization due to the asymmetry of the open probability of MET channels. Stereociliary ATP2B2 (PMCA2) extrudes calcium ions and basally located KCNQ4 extrudes potassium ions to repolarize the OHC.

Depolarization of the OHC causes a decrease in length of the OHC due to a very rapid, voltage-sensitive change in conformation of the membrane protein prestin (SLC26A5), an unusual member of the anion transporter family located in the lateral membrane (Mahendrasingam et al, 2010) that appears to respond to cytosolic chloride by altering its conformation in the plane of the plasma membrane (reviewed in Dallos et al. 2006, Dallos 2008, Hudspeth 2014, Reichenbach and Hudspeth 2014, Ashmore 2019, Santos-Sacchi 2019). Prestin also appears to act as a weak chloride-bicarbonate antiporter (Mistrik et al. 2012). Changes in length of the OHCs cause movement of the reticular lamina toward and away from the basilar membrane.


##### Edit history

| Date | Action | Author |
|---|---|---|
| 2019-09-23 | Edited | May B |
| 2019-09-23 | Authored | May B |
| 2019-09-23 | Created | May B |
| 2020-09-14 | Reviewed | Furness DN, Dallos P |
| 2020-12-12 | Modified | May B |

##### Entities found in this pathway (3)

| Input | UniProt Id | Input | UniProt Id | Input | UniProt Id |
|---|---|---|---|---|---|
| MYO15A | Q9UKN7 | PCDH15 | Q96QU1 | STRC | Q7RTU9 |

#### 17. Initiation of Nuclear Envelope (NE) Reformation (R-HSA-2995383)

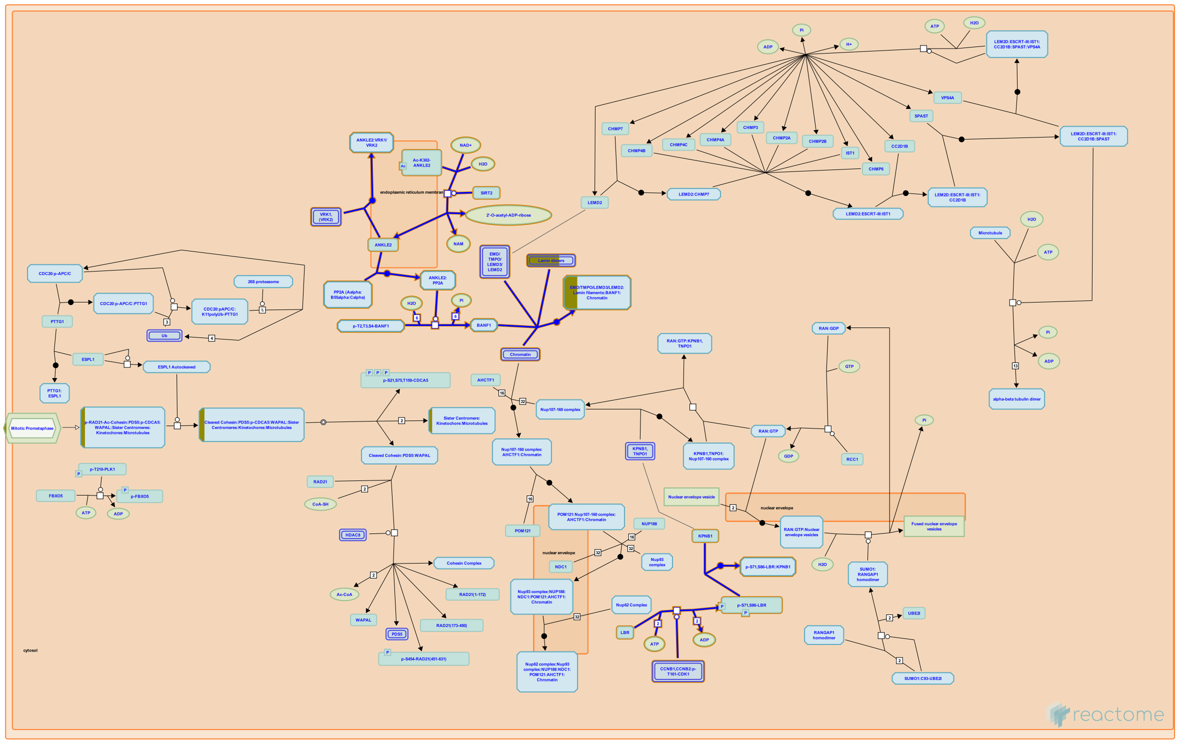

Reassembly of the nuclear envelope (NE) is initiated at late anaphase/early telophase when BANF1 (BAF), which is dispersed throughout the cytoplasm during metaphase, accumulates on the surfaces of coalesced chromosomes. This is coordinated with the chromatin association of membranes and inner nuclear membrane proteins that include EMD (emerin), TMPO (LAP2beta), LEMD3 (MAN1) and LEMD2 (LEM2), and lamins (Haraguchi et al. 2008, reviewed by Wandke and Kutay 2013). The DNA-cross-bridging activity of BANF1 is required for individual chromosomes to properly coalesce for enclosure in a single nucleus (Samwer et al. 2017).


##### Edit history

| Date | Action | Author |
| --- | --- | --- |
| 2013-01-22 | Created | Orlic-Milacic M |
| 2013-01-23 | Edited | Gillespie ME |
| 2013-01-23 | Authored | Orlic-Milacic M |
| 2013-01-30 | Reviewed | Gorjánácz M, Mattaj IW |

| Date | Action | Author |
|---|---|---|
| 2019-11-07 | Revised | Gerace L |
| 2019-11-26 | Edited | Orlic-Milacic M |
| 2021-05-22 | Modified | Shorser S |

#### Entities found in this pathway (1)

| Input | UniProt Id |
|---|---|
| LMNA | P02545-1, P02545-2 |

#### 18. PI-3K cascade:FGFR1 (R-HSA-5654689)

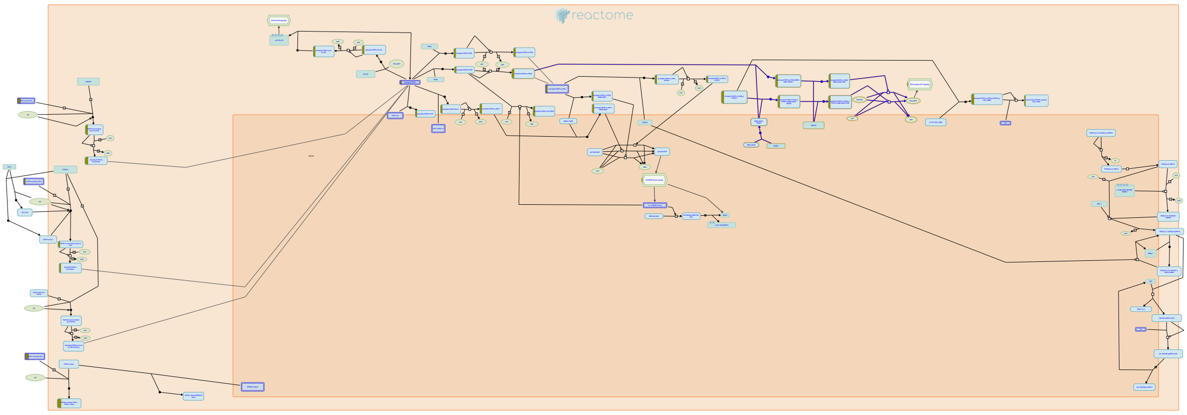

The ability of growth factors to protect from apoptosis is primarily due to the activation of the AKT survival pathway. P-I-3-kinase dependent activation of PDK leads to the activation of AKT which in turn affects the activity or expression of pro-apoptotic factors, which contribute to protection from apoptosis. AKT activation also blocks the activity of GSK-3b which could lead to additional antiapoptotic signals.


##### Edit history

| Date | Action | Author |
|---|---|---|
| 2007-01-10 | Authored | de Bono B |
| 2007-02-07 | Reviewed | Mohammadi M |
| 2010-02-03 | Edited | Jupe S |
| 2014-12-04 | Created | Rothfels K |
| 2021-05-31 | Modified | Shorser S |

##### Entities found in this pathway (2)

| Input | UniProt Id | Input | UniProt Id |
|---|---|---|---|
| FGF10 | O15520 | FGF17 | O60258-1 |

#### 19. TP53 regulates transcription of additional cell cycle genes whose exact role in the p53 pathway remain uncertain (R-HSA-6804115)

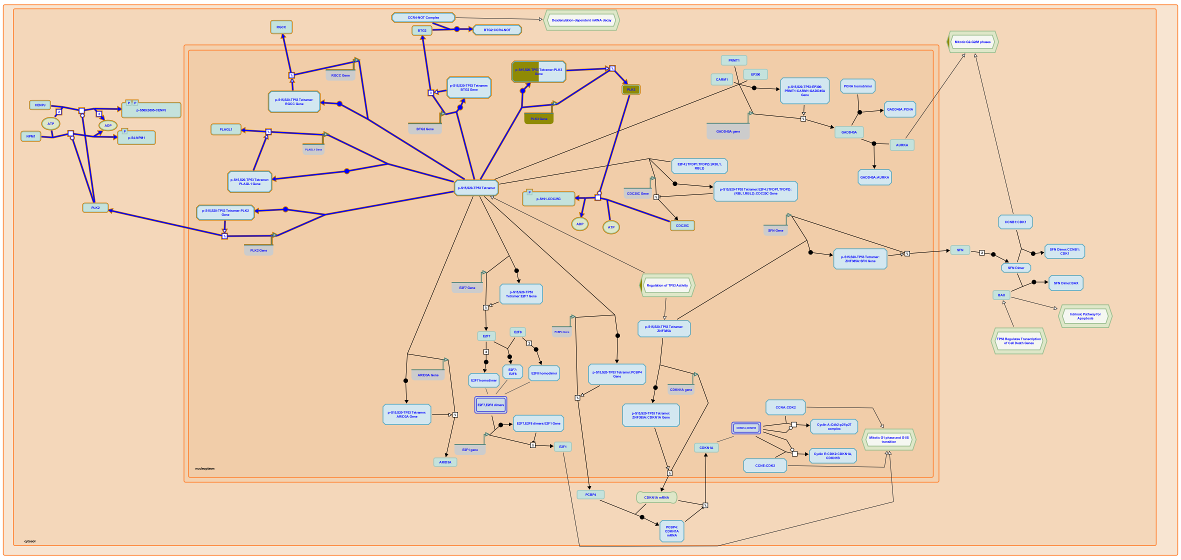

BTG2 is induced by TP53, leading to cessation of cellular proliferation (Rouault et al. 1996, Duriez et al. 2002). BTG2 binds to the CCR4-NOT complex and promotes mRNA deadenylation activity of this complex. Interaction between BTG2 and CCR4-NOT is needed for the antiproliferative activity of BTG2, but the underlying mechanism has not been elucidated (Rouault et al. 1998, Mauxion et al. 2008, Horiuchi et al. 2009, Doidge et al. 2012, Ezzeddine et al. 2012). Two polo-like kinases, PLK2 and PLK3, are direct transcriptional targets of TP53. TP53-mediated induction of PLK2 may be important for prevention of mitotic catastrophe after spindle damage (Burns et al. 2003). PLK2 is involved in the regulation of centrosome duplication through phosphorylation of centrosome-related proteins CENPJ (Chang et al. 2010) and NPM1 (Krause and Hoffmann 2010). PLK2 is frequently transcriptionally silenced through promoter methylation in B-cell malignancies (Syed et al. 2006). Induction of PLK3 transcription by TP53 (Jen and Cheung 2005) may be important for coordination of M phase events through PLK3-mediated nuclear accumulation of CDC25C (Bahassi et al. 2004). RGCC is induced by TP53 and implicated in cell cycle regulation, possibly through its association with PLK1 (Saigusa et al. 2007). PLAGL1 (ZAC1) is a zinc finger protein directly transcriptionally induced by TP53 (Rozenfeld-Granot et al. 2002). PLAGL1 expression is frequently lost in cancer (Varrault et al. 1998) and PLAGL1 has been implicated in both cell cycle arrest and apoptosis (Spengler et al. 1997), but its mechanism of action remains unknown.

#### Edit history

| Date | Action | Author |
|---|---|---|
| 2015-10-08 | Created | Orlic-Milacic M |
| 2015-10-14 | Edited | Orlic-Milacic M |
| 2015-10-14 | Authored | Orlic-Milacic M |
| 2016-02-04 | Reviewed | Zaccara S, Inga A |
| 2021-05-22 | Modified | Shorser S |

#### Entities found in this pathway (1)

| Input | UniProt Id |
|---|---|
| PLK3 | Q9H4B4 |

| Input | Ensembl Id |
|---|---|
| PLK3 | ENSG00000173846 |

#### 20. SHC-mediated cascade:FGFR1 (R-HSA-5654688)

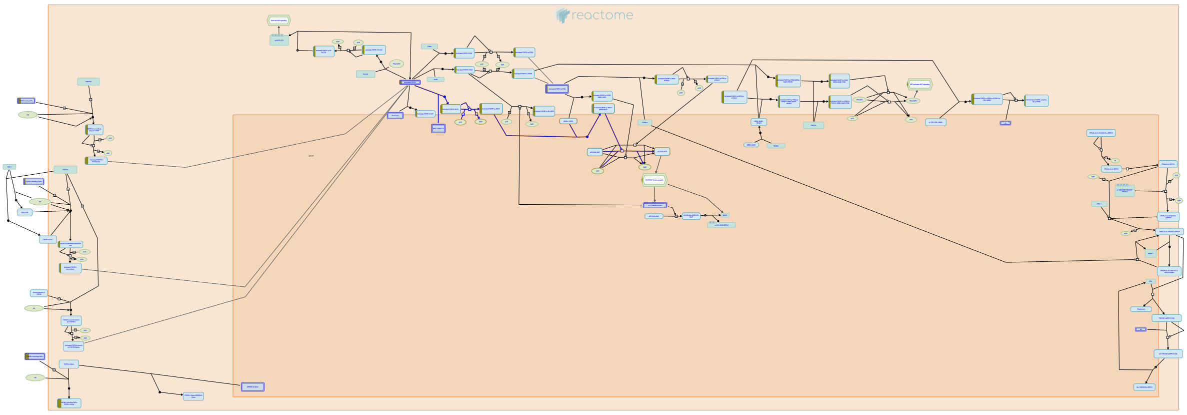

**Cellular compartments:** plasma membrane.

The exact role of SHC1 in FGFR signaling remains unclear. Numerous studies have shown that the p46 and p52 isoforms of SHC1 are phosphorylated in response to FGF stimulation, but direct interaction with the receptor has not been demonstrated. Co-precipitation of p46 and p52 with the FGFR2 IIIc receptor has been reported, but this interaction is thought to be indirect, possibly mediated by SRC. Consistent with this, co-precipitation of SHC1 and FGFR1 IIIc is seen in mammalian cells expressing v-SRC. The p66 isoform of SHC1 has also been co-precipitated with FGFR3, but this occurs independently of receptor stimulation, and the p66 isoform not been shown to undergo FGF-dependent phosphorylation. SHC1 has been shown to associate with GRB2 and SOS1 in response to FGF stimulation, suggesting that the recruitment of SHC1 may contribute to activation of the MAPK cascade downstream of FGFR.


##### Edit history

| Date | Action | Author |
|---|---|---|
| 2007-01-10 | Authored | de Bono B |
| 2007-02-07 | Reviewed | Mohammadi M |
| 2010-02-03 | Edited | Jupe S |

| Date | Action | Author |
|---|---|---|
| 2014-12-04 | Created | Rothfels K |
| 2021-05-31 | Modified | Shorser S |

#### Entities found in this pathway (2)

| Input | UniProt Id | Input | UniProt Id |
|---|---|---|---|
| FGF10 | O15520 | FGF17 | O60258-1 |

#### 21. FRS-mediated FGFR1 signaling (R-HSA-5654693)

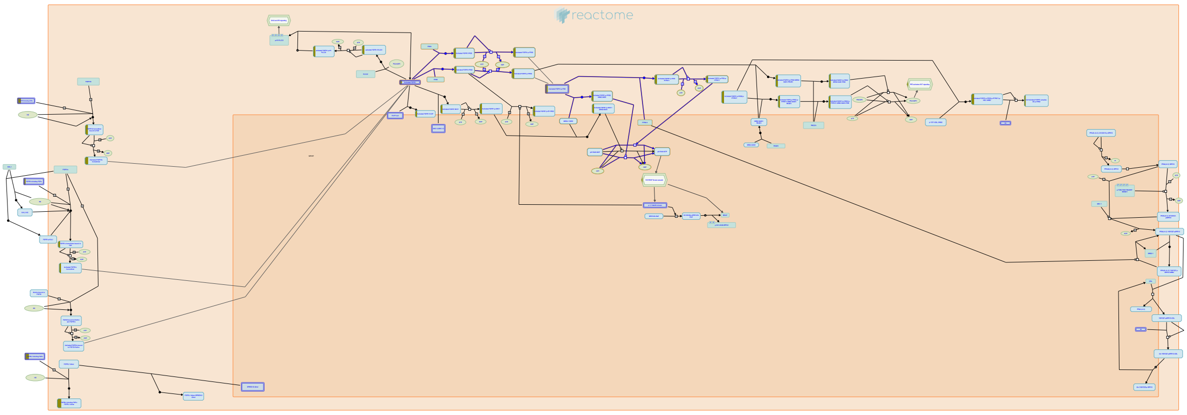

The FRS family of scaffolding adaptor proteins has two members, FRS2 (also known as FRS2 alpha) and FRS3 (also known as FRS2beta or SNT-2). Activation of FGFR tyrosine kinase allows FRS proteins to become phosphorylated on tyrosine residues and then bind to the adaptor GRB2 and the tyrosine phosphatase PPTN11/SHP2. Subsequently, PPTN11 activates the RAS-MAP kinase pathway and GRB2 activates the RAS-MAP kinase , PI-3-kinase and ubiquitinations/degradation pathways by binding to SOS, GAB1 and CBL, respectively, via the SH3 domains of GRB2. FRS2 acts as a central mediator in FGF signaling mainly because it induces sustained levels of activation of ERK with ubiquitous expression.


##### Edit history

| Date | Action | Author |
|---|---|---|
| 2007-01-10 | Authored | de Bono B |
| 2007-02-07 | Reviewed | Mohammadi M |
| 2010-02-03 | Edited | Jupe S |
| 2011-08-26 | Reviewed | Gotoh N |
| 2014-12-04 | Created | Rothfels K |
| 2021-05-22 | Modified | Shorser S |

##### Entities found in this pathway (2)

| Input | UniProt Id | Input | UniProt Id |
|---|---|---|---|
| FGF10 | O15520 | FGF17 | O60258-1 |

#### 22. PI-3K cascade:FGFR2 (R-HSA-5654695)

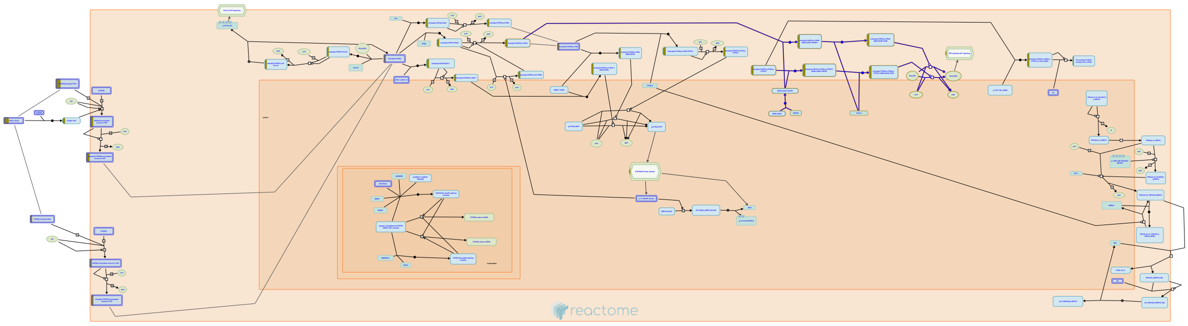

The ability of growth factors to protect from apoptosis is primarily due to the activation of the AKT survival pathway. P-I-3-kinase dependent activation of PDK leads to the activation of AKT which in turn affects the activity or expression of pro-apoptotic factors, which contribute to protection from apoptosis. AKT activation also blocks the activity of GSK-3b which could lead to additional antiapoptotic signals.


##### Edit history

| Date | Action | Author |
|---|---|---|
| 2007-01-10 | Authored | de Bono B |
| 2007-02-07 | Reviewed | Mohammadi M |
| 2010-02-03 | Edited | Jupe S |
| 2014-12-04 | Created | Rothfels K |
| 2021-05-31 | Modified | Shorser S |

##### Entities found in this pathway (2)

| Input | UniProt Id | Input | UniProt Id |
|---|---|---|---|
| FGF10 | O15520 | FGF17 | O60258-1 |

#### 23. SHC-mediated cascade:FGFR2 (R-HSA-5654699)

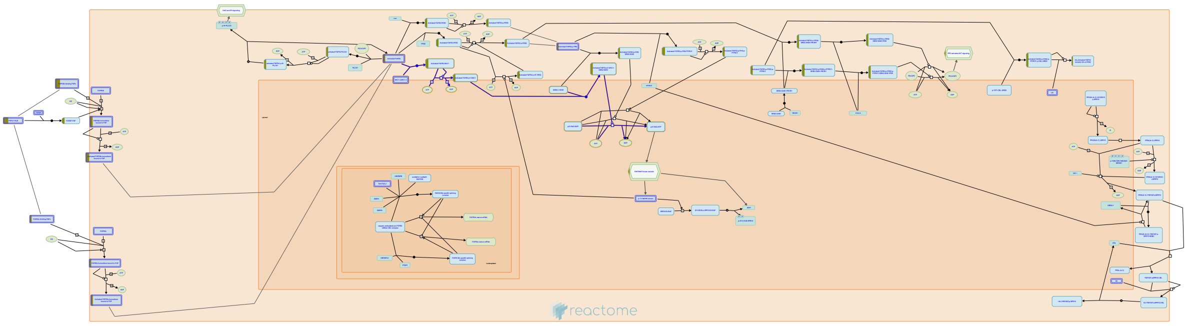

**Cellular compartments:** plasma membrane.

The exact role of SHC1 in FGFR signaling remains unclear. Numerous studies have shown that the p46 and p52 isoforms of SHC1 are phosphorylated in response to FGF stimulation, but direct interaction with the receptor has not been demonstrated. Co-precipitation of p46 and p52 with the FGFR2 IIIc receptor has been reported, but this interaction is thought to be indirect, possibly mediated by SRC. Consistent with this, co-precipitation of SHC1 and FGFR1 IIIc is seen in mammalian cells expressing v-SRC. The p66 isoform of SHC1 has also been co-precipitated with FGFR3, but this occurs independently of receptor stimulation, and the p66 isoform not been shown to undergo FGF-dependent phosphorylation. SHC1 has been shown to associate with GRB2 and SOS1 in response to FGF stimulation, suggesting that the recruitment of SHC1 may contribute to activation of the MAPK cascade downstream of FGFR.


##### Edit history

| Date | Action | Author |
|---|---|---|
| 2007-01-10 | Authored | de Bono B |
| 2007-02-07 | Reviewed | Mohammadi M |
| 2010-02-03 | Edited | Jupe S |
| 2014-12-04 | Created | Rothfels K |
| 2021-05-31 | Modified | Shorser S |

#### Entities found in this pathway (2)

| Input | UniProt Id | Input | UniProt Id |
|---|---|---|---|
| FGF10 | O15520 | FGF17 | O60258-1 |

#### 24. Sensory processing of sound by inner hair cells of the cochlea (R-HSA-9662360)

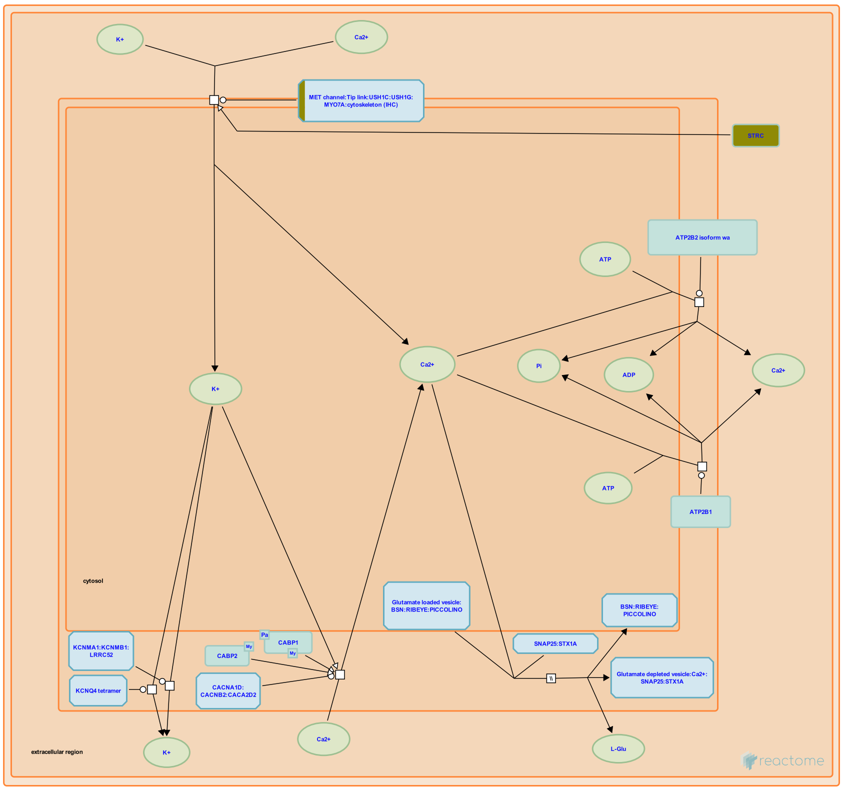


Inner hair cells (IHCs) of the cochlea transduce sound waves into an ionic (mainly potassium) current that leads to exocytosis of glutamate from the IHC and activation of postsynaptic type I afferent fibers of the radial ganglion (reviewed in Meyer and Moser 2010, Moser and Vogl 2016, Fettiplace 2017). IHCs have stereocilia on their apical surface that are arranged in rows of increasing height, a "staircase" arrangement. Stereocilia of different rows are connected by a tip link comprising a CDH23 dimer on the taller stereocilium bound to a PCDH15 dimer on the shorter stereocilium. PCDH15 interacts with LHFPL5, an auxiliary subunit of the mechanoelectrical transduction channel (MET channel, also called the mechanotransduction channel), which contains at least TMC1 (adults) or TMC2 (newborns), TMIE, and the auxiliary subunits LHFPL5 and CIB2 (reviewed in Fettiplace and Kim 2014, Fettiplace 2016).

Deflection of the stereocilia by sound waves creates tension on the tip link that increases the open probability of the MET channel, which then transports calcium and potassium ions from the scala media into the IHC, depolarizing the IHC (reviewed in Fettiplace 2017). The potassium channel KCNQ4 located in the neck region of the cell may also participate in depolarization. The depolarization of the IHC opens voltage-gated Cav1.3 channels (CACNA1D:CACA2D2:CACNB2) located in stripes near ribbon synapses on the basolateral surface of the IHC. The resulting localized influx of calcium ions activates exocytosis of glutamate into the synapse by an interaction between calcium and Otoferlin (OTOF) on glutamate-loaded vesicles in the IHC (reviewed in Wichmann 2015).

Ribbon synapses are characterized by a multiprotein complex, the ribbon, that contains at least BASSOON, RIBEYE (an isoform of CTBP2), and PICCOLINO (a small isoform of PICCOLO) and appears to act to transiently tether vesicles near the synapse and thereby increase the pool of readily releasable vesicles (reviewed in Safieddine et al. 2012, Wichman and Moser 2015, Pangrsic and Vogl 2018, Moser et al. 2020).

ATP2B1 calcium channels, ATP2B2 calcium channels, KCNMA1:KCNMB1:LRRC52 potassium channels, and basolateral KCNQ4 potassium channels transport cations out of the IHC and thereby act to repolarize the cell and limit the duration of the synaptic potentials (reviewed in Patuzzi 2011, Oak and Yi 2014).


#### Edit history

| Date | Action | Author |
|---|---|---|
| 2019-09-23 | Edited | May B |
| 2019-09-23 | Authored | May B |
| 2019-09-23 | Created | May B |
| 2020-09-14 | Reviewed | Furness DN |
| 2020-12-12 | Modified | May B |

#### Entities found in this pathway (3)

| Input | UniProt Id | Input | UniProt Id | Input | UniProt Id |
|---|---|---|---|---|---|
| MYO15A | Q9UKN7 | PCDH15 | Q96QU1 | STRC | Q7RTU9 |

#### 25. FRS-mediated FGFR2 signaling (R-HSA-5654700)

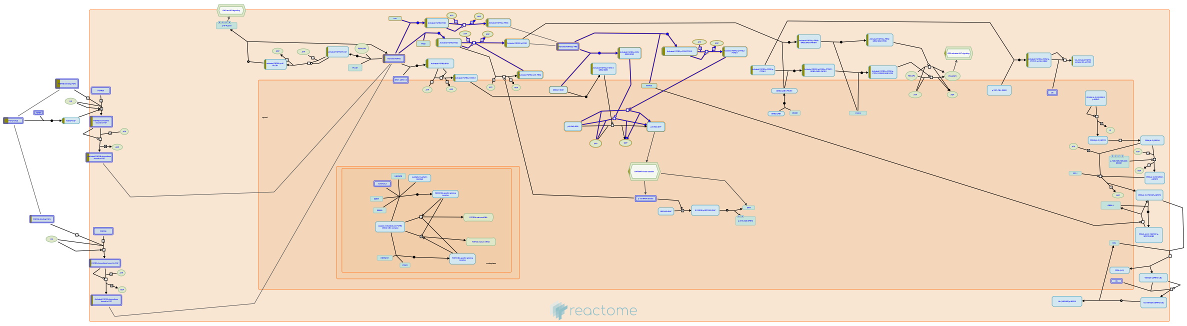

The FRS family of scaffolding adaptor proteins has two members, FRS2 (also known as FRS2 alpha) and FRS3 (also known as FRS2beta or SNT-2). Activation of FGFR tyrosine kinase allows FRS proteins to become phosphorylated on tyrosine residues and then bind to the adaptor GRB2 and the tyrosine phosphatase PPTN11/SHP2. Subsequently, PPTN11 activates the RAS-MAP kinase pathway and GRB2 activates the RAS-MAP kinase , PI-3-kinase and ubiquitinations/degradation pathways by binding to SOS, GAB1 and CBL, respectively, via the SH3 domains of GRB2. FRS2 acts as a central mediator in FGF signaling mainly because it induces sustained levels of activation of ERK with ubiquitous expression.


##### Edit history

| Date | Action | Author |
|---|---|---|
| 2007-01-10 | Authored | de Bono B |
| 2007-02-07 | Reviewed | Mohammadi M |
| 2010-02-03 | Edited | Jupe S |
| 2011-08-26 | Reviewed | Gotoh N |
| 2014-12-04 | Created | Rothfels K |
| 2021-05-22 | Modified | Shorser S |

##### Entities found in this pathway (2)

| Input | UniProt Id | Input | UniProt Id |
|---|---|---|---|
| FGF10 | O15520 | FGF17 | O60258-1 |

| Input | UniProt Id | Input | UniProt Id |
|---|---|---|---|

### 6. Identifiers found

Below is a list of the input identifiers that have been found or mapped to an equivalent element in Reactome, classified by resource.

#### Entities (60)

| Input | UniProt Id | Input | UniProt Id | Input | UniProt Id |
|---|---|---|---|---|---|
| APBB1IP | Q7Z5R6 | AQP9 | O14520, O43315 | ARNTL | O00327 |
| ART3 | O60678 | ATF3 | P18847 | BAMBI | Q13145 |
| CCL4L2 | Q8NHW4 | CEBPD | P49716 | CLEC7A | Q9BXN2 |
| COL22A1 | Q8NFW1 | CX3CR1 | P49238 | CYR61 | O00622 |
| DOCK8 | Q8NF50 | EGR3 | Q06889 | FGF10 | O15520 |
| FGF17 | O60258-1 | FMN1 | Q969G6 | GABRG2 | P18507 |
| GPCPD1 | Q9NPB8 | GPX3 | O75715, P22352 | GRIN1 | Q05586 |
| HTR1E | P28566 | HTR3B | O95264 | IL1A | P01583 |
| IL1RL1 | Q01638, Q01638-2 | IL4R | P24394 | IL7R | P16871 |
| KCND3 | Q9UK17 | KCNIP2 | Q9NS61 | LMNA | P02545-1, P02545-2 |
| MAST1 | Q7Z460 | MCL1 | Q07820 | MLXIPL | Q9NP71 |
| MYO15A | Q9UKN7 | NIPAL2 | Q9H841 | NISCH | Q9Y2I1 |
| NR4A1 | P22736 | NR4A2 | P43354 | NR4A3 | Q92570-1, Q92570-2 |
| OLR1 | P78380 | PCDH15 | Q96QU1 | PLEKHG5 | O94827 |
| PLK3 | Q9H4B4 | PTN | P21246 | PXDN | Q92626 |
| SESN1 | Q9Y6P5-1, Q9Y6P5-3 | SLC17A6 | Q9P2U8 | SLC52A3 | Q9NQ40 |
| SLCO4A1 | Q96BD0 | SOX6 | P35712 | STEAP2 | Q8NFT2 |
| STRC | Q7RTU9 | TAF7L | Q5H9L4 | TIAM2 | Q8IVF5 |
| TNFRSF25 | Q93038 | UNC5A | Q6ZN44 | VCAN | P13611 |
| ZBTB16 | Q05516 | ZNF441 | Q8N8Z8 | | |

| Input | Ensembl Id | Input | Ensembl Id | Input | Ensembl Id |
|---|---|---|---|---|---|
| ARNTL | ENSG00000133794 | ATF3 | ENSG00000162772 | CEBPD | ENSG00000221869 |
| GRIN1 | ENSG00000169258 | IL1A | ENSG00000115008 | IL4R | ENSG00000077238 |
| LMNA | ENSG00000160789 | MCL1 | ENSG00000143384 | NR4A3 | ENSG00000119508 |
| PLK3 | ENSG00000173846 | SESN1 | ENSG00000080546 | TIAM2 | ENSG00000146426 |

| Input | miRBase Id |
|---|---|
| MIR24-2 | MI0000081 |

### 7. Identifiers not found

These 78 identifiers were not found neither mapped to any entity in Reactome.

| | | | | | | | |
|---|---|---|---|---|---|---|---|
| AC003003.5 | AC093590.1 | AL589743.1 | ANKRD20A5P | ANKRD37 | ARHGAP36 | C22orf24 | C9orf91 |
| CADPS2 | CALY | CBLN2 | CD83 | CERKL | CHRD | CMAHP | CTD-2020K17.3 |
| CTD-2201E9.2 | CYP1B1-AS1 | CYP4F35P | DCDC1 | EGFEM1P | FAIM2 | FAM179A | FAM95B1 |
| FAT1 | GSDMB | HCG22 | JPH4 | KCNT1 | KIAA1683 | LIMA1 | LINC00575 |
| LINC00846 | LOC100421166 | MAPK15 | MIR17HG | MIR181A1HG | MIR22HG | NICN1-AS1 | NME5 |
| OGFOD3 | PDLIM3 | PER3 | PKD1L1 | PKHD1L1 | PRRT3 | RHOT1P1 | RNU1-146P |
| RP1-90J20.11 | RP1-90J20.12 | RP11-15H20.6 | RP11-21L23.2 | RP11-284F21.11 | RP11-314P15.2 | RP11-382N13.1 | RP11-41O4.1 |
| RP11-535M15.2 | RP11-5N11.2 | RP11-64P14.7 | RP11-757O6.1 | RP11-79E3.2 | RP5-944M2.2 | RPS16P5 | SAMD3 |
| SAMSN1 | SCARNA2 | SFMBT2 | SNORA79 | TCP10L | TMEM120B | TMEM56 | TTC39A |
| U95743.1 | USP32P1 | VWA3A | WSCD1 | YPEL4 | ZGLP1 | | |
