## Supplementary material for "Compartment-specific total RNA profile of Hippocampal and Cortical cells from Mesial Temporal Lobe Epilepsy tissue": Other supplemental files_Reactome: Cx_mTLE+HS_Cyt_updownspecific_report.pdf

### 3. Genome-wide overview

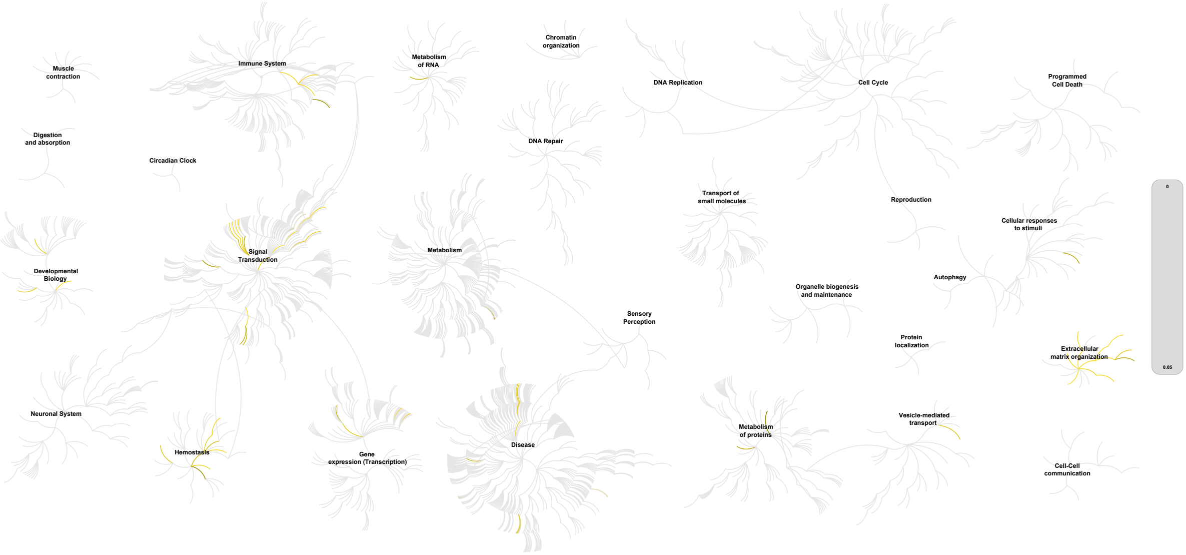


This figure shows a genome-wide overview of the results of your pathway analysis. Reactome pathways are arranged in a hierarchy. The center of each of the circular "bursts" is the root of one top-level pathway, for example "DNA Repair". Each step away from the center represents the next level lower in the pathway hierarchy. The color code denotes over-representation of that pathway in your input dataset. Light grey signifies pathways which are not significantly over-represented.

### 4. Most significant pathways

The following table shows the 25 most relevant pathways sorted by p-value.

| Pathway name | Entities | | | | Reactions | |
|---|---|---|---|---|---|---|
| | found | ratio | p-value | FDR* | found | ratio |
| Interleukin-4 and Interleukin-13 signaling | 16 / 211 | 0.015 | 1.88e-06 | 0.001 | 7 / 47 | 0.003 |
| NGF-stimulated transcription | 8 / 56 | 0.004 | 9.78e-06 | 0.002 | 12 / 37 | 0.003 |
| Collagen biosynthesis and modifying enzymes | 9 / 76 | 0.005 | 1.20e-05 | 0.002 | 26 / 51 | 0.004 |
| NOTCH2 intracellular domain regulates transcription | 5 / 16 | 0.001 | 1.24e-05 | 0.002 | 9 / 9 | 6.66e-04 |
| Nuclear Events (kinase and transcription factor activation) | 9 / 80 | 0.006 | 1.80e-05 | 0.002 | 13 / 48 | 0.004 |
| Assembly of collagen fibrils and other multimeric structures | 8 / 67 | 0.005 | 3.47e-05 | 0.004 | 20 / 26 | 0.002 |
| Degradation of the extracellular matrix | 11 / 148 | 0.01 | 9.22e-05 | 0.009 | 21 / 105 | 0.008 |
| Extracellular matrix organization | 17 / 329 | 0.023 | 1.15e-04 | 0.009 | 102 / 319 | 0.024 |
| NOTCH4 Intracellular Domain Regulates Transcription | 5 / 26 | 0.002 | 1.21e-04 | 0.009 | 8 / 9 | 6.66e-04 |
| Collagen formation | 9 / 104 | 0.007 | 1.31e-04 | 0.009 | 46 / 77 | 0.006 |
| Collagen chain trimerization | 6 / 44 | 0.003 | 1.65e-04 | 0.01 | 6 / 28 | 0.002 |
| Signaling by NTRK1 (TRKA) | 10 / 143 | 0.01 | 3.09e-04 | 0.018 | 14 / 102 | 0.008 |
| NOTCH3 Intracellular Domain Regulates Transcription | 5 / 36 | 0.002 | 5.34e-04 | 0.028 | 20 / 22 | 0.002 |
| Signaling by NOTCH2 | 5 / 38 | 0.003 | 6.80e-04 | 0.033 | 9 / 20 | 0.001 |
| Signaling by NTRKs | 10 / 166 | 0.011 | 9.61e-04 | 0.044 | 14 / 164 | 0.012 |
| Integrin cell surface interactions | 7 / 86 | 0.006 | 0.001 | 0.045 | 10 / 55 | 0.004 |
| GP1b-IX-V activation signalling | 3 / 12 | 8.25e-04 | 0.001 | 0.056 | 5 / 7 | 5.18e-04 |
| Collagen degradation | 6 / 69 | 0.005 | 0.002 | 0.063 | 11 / 34 | 0.003 |
| RAF-independent MAPK1/3 activation | 4 / 28 | 0.002 | 0.002 | 0.063 | 4 / 12 | 8.88e-04 |
| Syndecan interactions | 4 / 29 | 0.002 | 0.002 | 0.068 | 8 / 15 | 0.001 |
| LGI-ADAM interactions | 3 / 14 | 9.63e-04 | 0.002 | 0.07 | 4 / 5 | 3.70e-04 |
| Signaling by Interleukins | 22 / 643 | 0.044 | 0.003 | 0.088 | 70 / 493 | 0.036 |
| RUNX3 regulates NOTCH signaling | 3 / 16 | 0.001 | 0.003 | 0.088 | 5 / 7 | 5.18e-04 |
| Platelet Adhesion to exposed collagen | 3 / 16 | 0.001 | 0.003 | 0.088 | 4 / 6 | 4.44e-04 |
| ECM proteoglycans | 6 / 79 | 0.005 | 0.003 | 0.089 | 9 / 23 | 0.002 |

* False Discovery Rate

### 5. Pathways details

For every pathway of the most significant pathways, we present its diagram, as well as a short summary, its bibliography and the list of inputs found in it.

#### 1. Interleukin-4 and Interleukin-13 signaling (R-HSA-6785807)

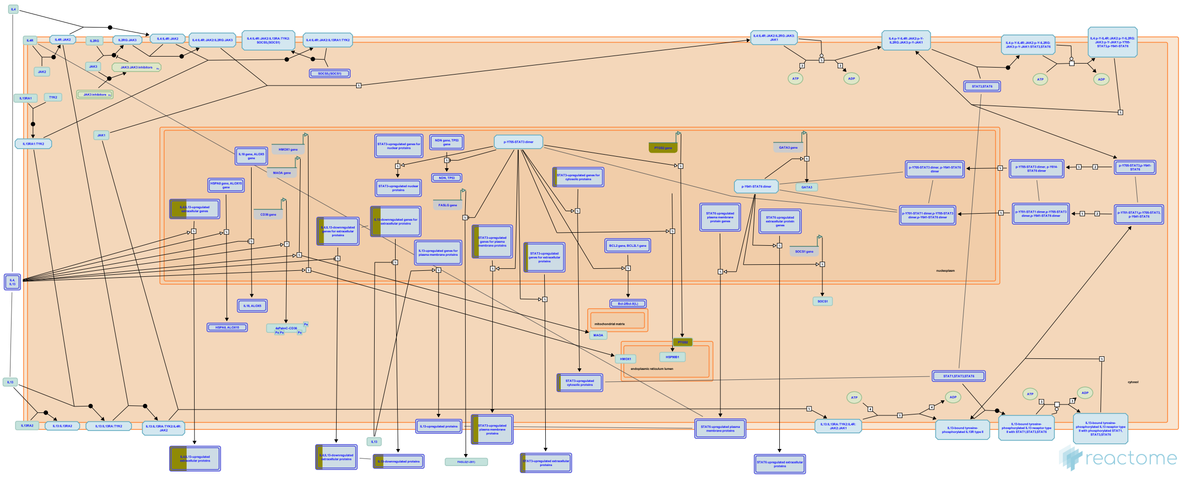

Interleukin-4 (IL4) is a principal regulatory cytokine during the immune response, crucially important in allergy and asthma (Nelms et al. 1999). When resting T cells are antigen-activated and expand in response to Interleukin-2 (IL2), they can differentiate as Type 1 (Th1) or Type 2 (Th2) T helper cells. The outcome is influenced by IL4. Th2 cells secrete IL4, which both stimulates Th2 in an autocrine fashion and acts as a potent B cell growth factor to promote humoral immunity (Nelms et al. 1999).

| Input | UniProt Id | Input | UniProt Id | Input | UniProt Id |
|---|---|---|---|---|---|
| COL1A2 | P08123 | FN1 | P02751 | IL1A | P01583 |
| IL6R | P08887 | NOS2 | P35228 | PTGS2 | P35354 |
| TIMP1 | P01033 | VEGFA | P15692 | | |

| Input | Ensembl Id | Input | Ensembl Id | Input | Ensembl Id |
|---|---|---|---|---|---|
| COL1A2 | ENSG00000164692 | FN1 | ENSG00000115414 | IL1A | ENSG00000115008 |
| IL6R | ENSG00000160712 | NOS2 | ENSG00000007171 | PTGS2 | ENSG00000073756 |
| TIMP1 | ENSG00000102265 | VEGFA | ENSG00000112715 | | |

#### 2. NGF-stimulated transcription (R-HSA-9031628)

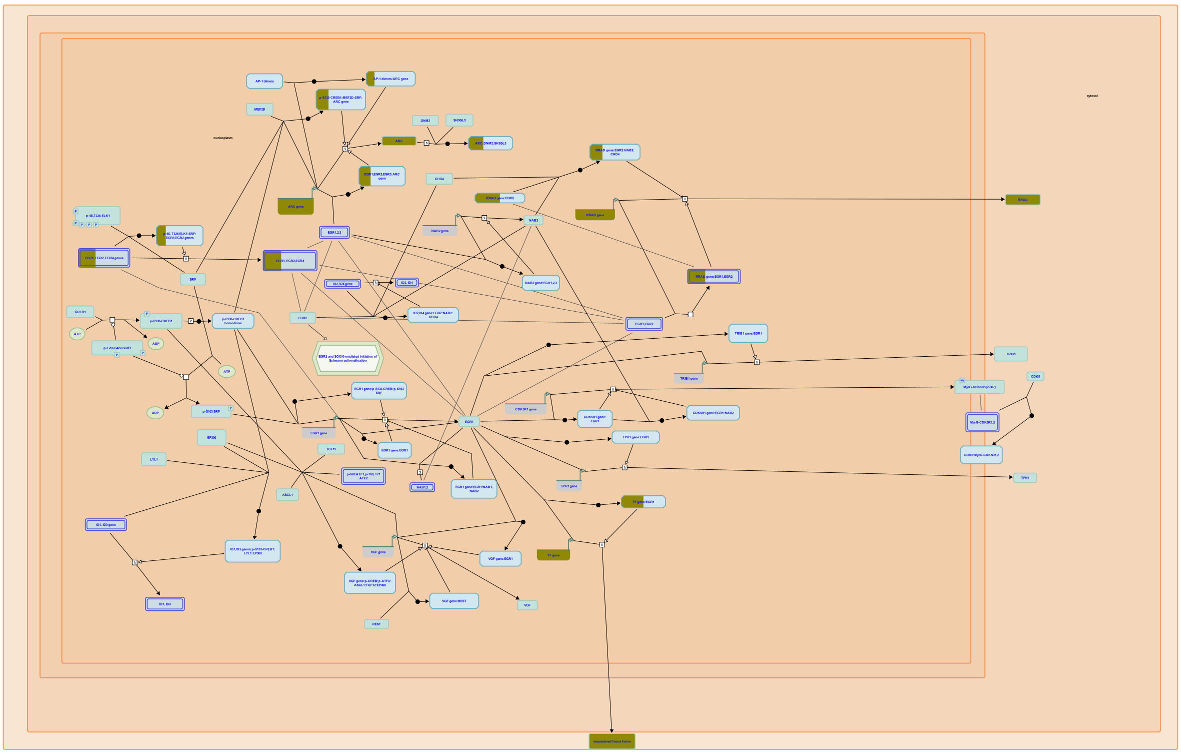

NGF stimulation induces expression of a wide array of transcriptional targets. In rat PC12 cells, a common model for NGF signaling, stimulation with NGF causes cells to exit the cell cycle and undergo a differentiation program leading to neurite outgrowth. This program is driven by the expression of immediate early genes (IEGs), which frequently encode transcription factors regulating the activity of NGF-specific delayed response genes (reviewed in Sheng and Greenberg, 1990; Flavell and Grennberg, 2008; Santiago and Bashaw, 2014).


##### Edit history

| Date | Action | Author |
|---|---|---|
| 2017-12-01 | Created | Rothfels K |
| 2019-08-16 | Authored | Rothfels K |
| 2020-01-17 | Reviewed | Aletta J M |
| 2020-02-24 | Edited | Rothfels K |
| 2021-05-22 | Modified | Shorser S |

#### Entities found in this pathway (4)

| Input | UniProt Id |
|---|---|
| ARC | Q7LC44 |
| F3 | P13726 |

| Input | UniProt Id |
|---|---|
| EGR4 | Q05215 |
| RRAD | P55042 |

| Input | Ensembl Id |
|---|---|
| ARC | ENSG00000198576 |
| F3 | ENSG00000117525 |

| Input | Ensembl Id |
|---|---|
| EGR4 | ENSG00000135625 |
| RRAD | ENSG00000166592 |

#### 3. Collagen biosynthesis and modifying enzymes (R-HSA-1650814)

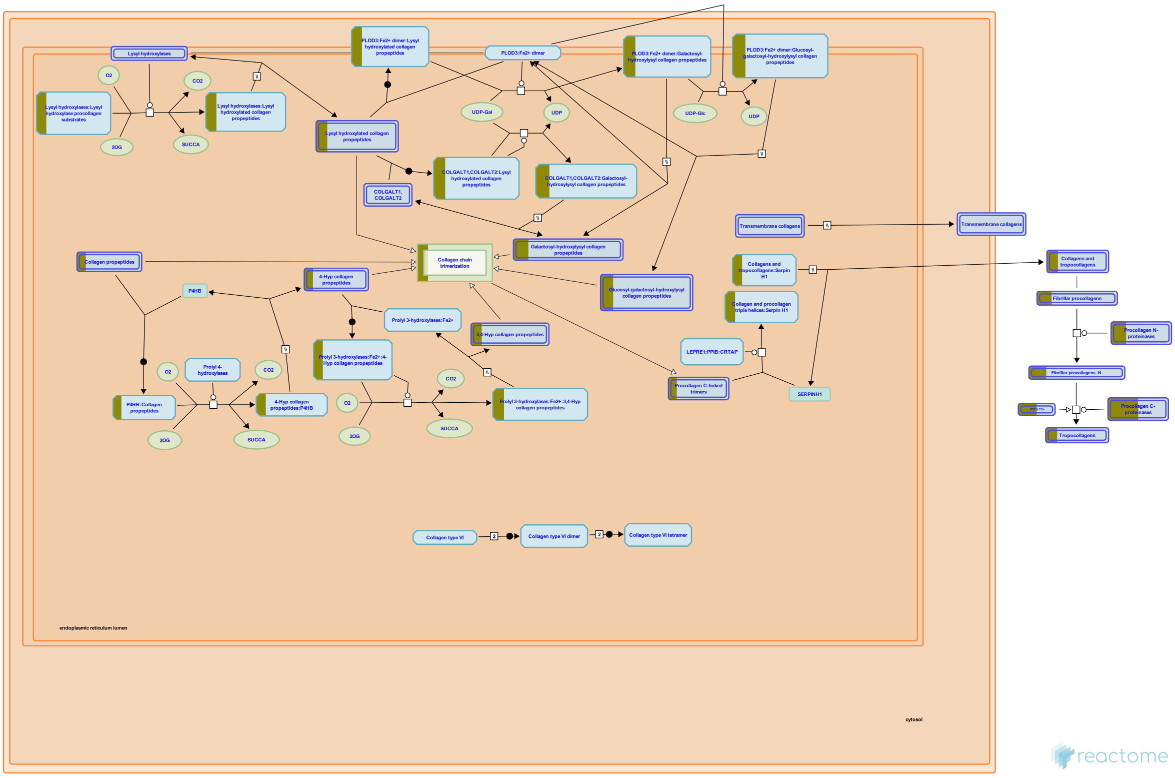


The biosynthesis of collagen is a multistep process. Collagen propeptides are cotranslationally translocated into the ER lumen. Propeptides undergo a number of post-translational modifications. Proline and lysine residues may be hydroxylated by prolyl 3-, prolyl 4- and lysyl hydroxylases. 4-hydroxyproline is essential for intramolecular hydrogen bonding and stability of the triple helical collagenous domain. In fibril forming collagens approximately 50% of prolines are 4-hydroxylated; the extent of this and of 3-hydroxyproline and lysine hydroxylation varies between tissues and collagen types (Kivirikko et al. 1972, 1992). Hydroxylysine molecules can form cross-links between collagen molecules in fibrils, and are sites for glycosyl- and galactosylation. Collagen peptides all have non-collagenous domains; collagens within the subclasses have common chain structures. These non-collagenous domains have regulatory functions; some are biologically active when cleaved from the main peptide chain. Fibrillar collagens all have a large triple helical domain (COL1) bordered by N and C terminal extensions, called the N and C propeptides, which are cleaved prior to formation of the collagen fibril. The C propeptide, also called the NC1 domain, is highly conserved. It directs chain association during intracellular assembly of the procollagen molecule from three collagen propeptide alpha chains (Hulmes 2002). The N-propeptide has a short linker (NC2) connecting the main triple helix to a short minor one (COL2) and a globular N-terminal region NC3. NC3 domains are variable both in size and the domains they contain.

| Input | UniProt Id | Input | UniProt Id | Input | UniProt Id |
|---|---|---|---|---|---|
| ADAMTS2 | O95450 | COL14A1 | Q05707 | COL15A1 | P39059 |
| COL1A2 | P08123 | COL5A1 | P20908 | COL8A2 | P25067, Q14050 |
| PCOLCE | Q15113 | TLL2 | Q9Y6L7 | | |

#### 4. NOTCH2 intracellular domain regulates transcription (R-HSA-2197563)

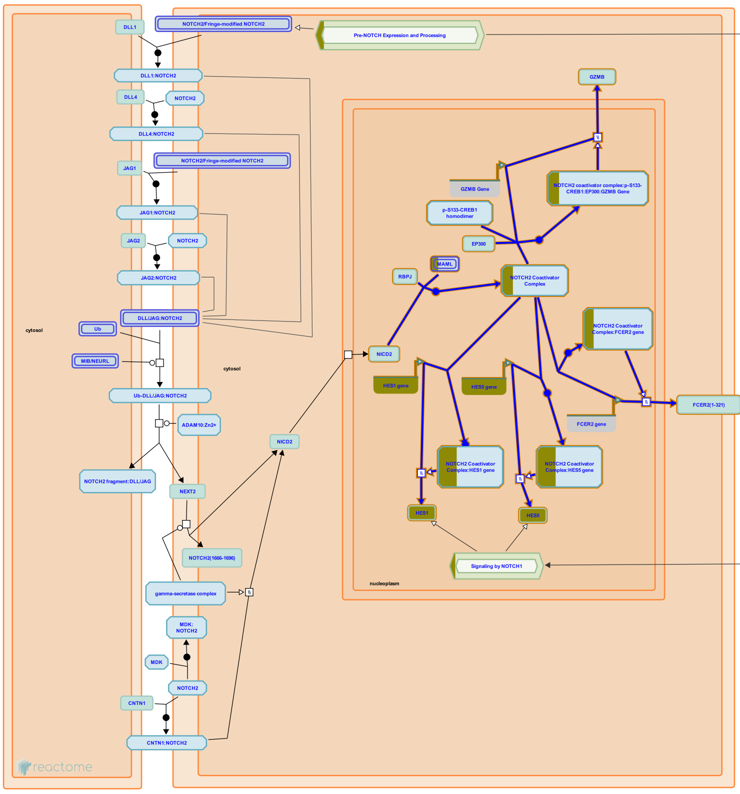


**Cellular compartments:** nucleoplasm.

In the nucleus, NICD2 forms a complex with RBPJ (CBF1, CSL) and MAML (mastermind). NICD2:RBPJ:MAML complex activates transcription from RBPJ-binding promoter elements (RBEs) (Wu et al. 2000). Besides NICD2, RBPJ and MAML, NOTCH2 coactivator complex likely includes other proteins, shown as components of the NOTCH1 coactivator complex.

NOTCH2 coactivator complex directly stimulates transcription of HES1 and HES5 genes (Shimizu et al. 2002), both of which are known NOTCH1 targets.

The promoter of FCER2 (CD23A) contains several RBEs that are occupied by NOTCH2 but not NOTCH1 coactivator complexes, and NOTCH2 activation stimulates FCER2 transcription. Overexpression of FCER2 (CD23A) is a hallmark of B-cell chronic lymphocytic leukemia (B-CLL) and correlates with the malfunction of apoptosis, which is thought be an underlying mechanism of B-CLL development. The Epstein-Barr virus protein EBNA2 can also activate FCER2 transcription through RBEs, possibly by mimicking NOTCH2 signaling (Hubmann et al. 2002).

NOTCH2 coactivator complex occupies the proximal RBE of the GZMB (granzyme B) promoter and at the same time interacts with phosphorylated CREB1, bound to an adjacent CRE site. EP300 transcriptional coactivator is also recruited to this complex through association with CREB1 (Maekawa et al. 2008). NOTCH2 coactivator complex together with CREBP1 and EP300 stimulates transcription of GZMB (granzyme B), which is important for the cytotoxic function of CD8+ T-cells (Maekawa et al. 2008).

There are indications that NOTCH2 genetically interacts with hepatocyte nuclear factor 1-beta (HNF1B) in kidney development (Massa et al. 2013, Heliot et al. 2013) and with hepatocyte nuclear factor 6 (HNF6) in bile duct formation (Vanderpool et al. 2012), but the exact nature of these genetic interactions has not been defined.


#### Edit history

| Date | Action | Author |
|---|---|---|
| 2012-04-12 | Created | Orlic-Milacic M |
| 2013-01-11 | Authored | Orlic-Milacic M |
| 2013-01-14 | Edited | Haw R |
| 2013-04-25 | Reviewed | Ilagan MXG, Boyle S |
| 2021-05-21 | Modified | Shorser S |

#### Entities found in this pathway (3)

| Input | UniProt Id | Input | UniProt Id | Input | UniProt Id |
|---|---|---|---|---|---|
| HES1 | Q14469 | HES5 | Q5TA89 | MAML3 | Q96JK9 |

| Input | Ensembl Id | Input | Ensembl Id |
|---|---|---|---|
| HES1 | ENSG00000114315 | HES5 | ENSG00000197921 |

#### 5. Nuclear Events (kinase and transcription factor activation) (R-HSA-198725)

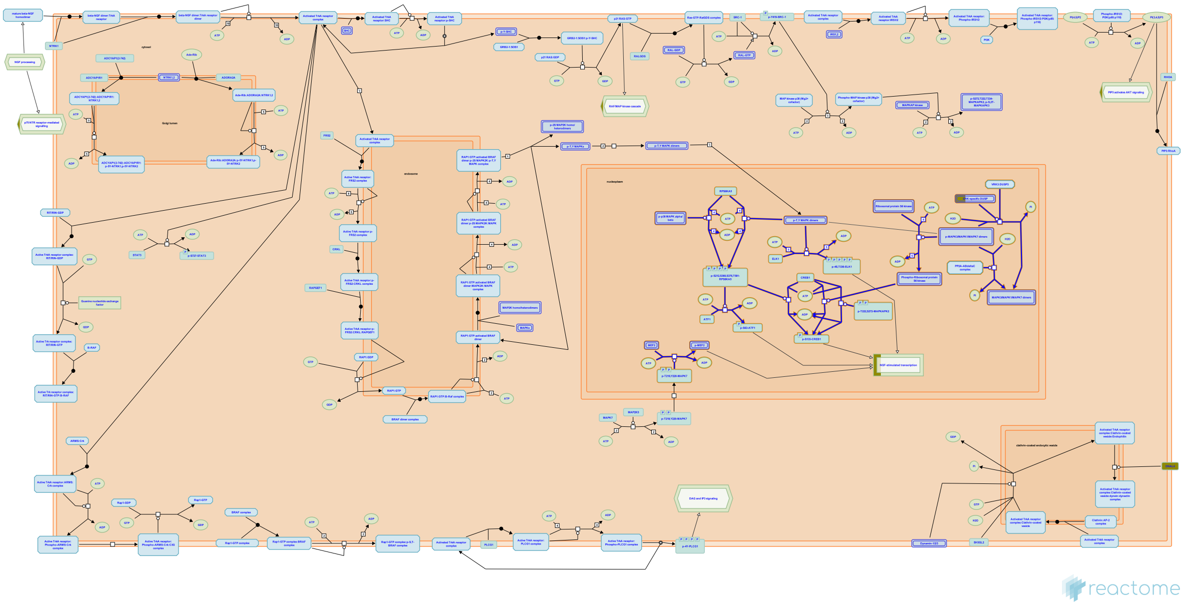

An important function of the kinase cascade triggered by neurotrophins is to induce the phosphorylation and activation of transcription factors in the nucleus to initiate new programs of gene expression. Transcription factors directly activated by neurotrophin signalling are responsible for induction of immediate-early genes, many of which are transcription factors. These in turn are involved in the induction of delayed-early genes.


##### Edit history

| Date | Action | Author |
|---|---|---|
| 2006-10-10 | Authored | Annibali D, Nasi S |
| 2007-07-10 | Created | |
| 2007-11-08 | Reviewed | Greene LA |
| 2021-05-22 | Modified | Shorser S |

##### Entities found in this pathway (5)

| Input | UniProt Id | Input | UniProt Id | Input | UniProt Id |
|---|---|---|---|---|---|
| ARC | Q7LC44 | DUSP6 | Q16828 | EGR4 | Q05215 |
| F3 | P13726 | RRAD | P55042 | | |

| Input | Ensembl Id | Input | Ensembl Id |
|---|---|---|---|
| ARC | ENSG00000198576 | EGR4 | ENSG00000135625 |
| F3 | ENSG00000117525 | RRAD | ENSG00000166592 |

#### 6. Assembly of collagen fibrils and other multimeric structures (R-HSA-2022090)

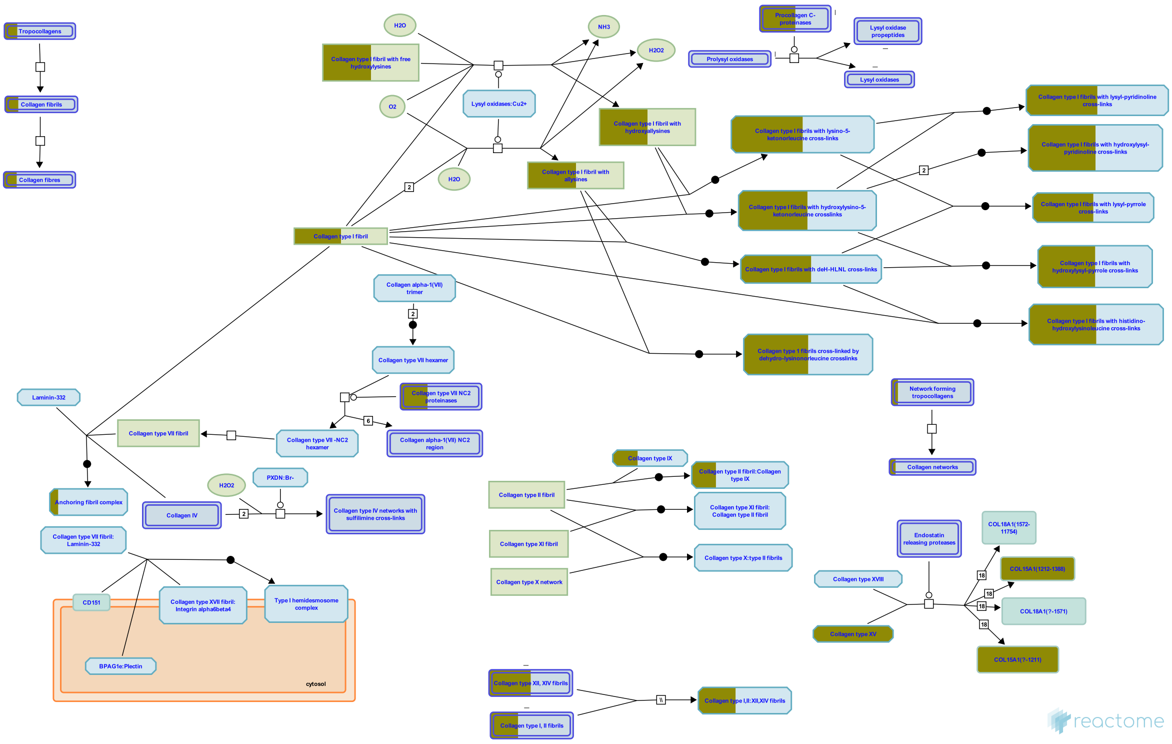


Collagen trimers in triple-helical form, referred to as procollagen or collagen molecules, are exported from the ER and trafficked through the Golgi network before secretion into the extracellular space. For fibrillar collagens namely types I, II, III, V, XI, XXIV and XXVII (Gordon & Hahn 2010, Ricard-Blum 2011) secretion is concomitant with processing of the N and C terminal collagen propeptides. These processed molecules are known as tropocollagens, considered to be the units of higher order collagen structures. They form within the extracellular space via a process that can proceed spontaneously, but in the cellular environment is regulated by many collagen binding proteins such as the FACIT (Fibril Associated Collagens with Interrupted Triple helices) family collagens and Small Leucine-Rich Proteoglycans (SLRPs). The architecture formed ultimately depends on the collagen subtype and the cellular conditions. Structures include the well-known fibrils and fibres formed by the major structural collagens type I and II plus several different types of supramolecular assembly (Bruckner 2010). The mechanical and physical properties of tissues depend on the spatial arrangement and composition of these collagen-containing structures (Kadler et al. 1996, Shoulders & Raines 2009, Birk & Bruckner 2011).

#### Entities found in this pathway (7)

| Input | UniProt Id | Input | UniProt Id | Input | UniProt Id |
|---|---|---|---|---|---|
| COL14A1 | Q05707 | COL15A1 | P39059 | COL1A2 | P08123 |
| COL5A1 | P20908 | COL8A2 | P25067, Q14050 | PCOLCE | Q15113 |
| TLL2 | Q9Y6L7 | | | | |

#### 7. Degradation of the extracellular matrix (R-HSA-1474228)

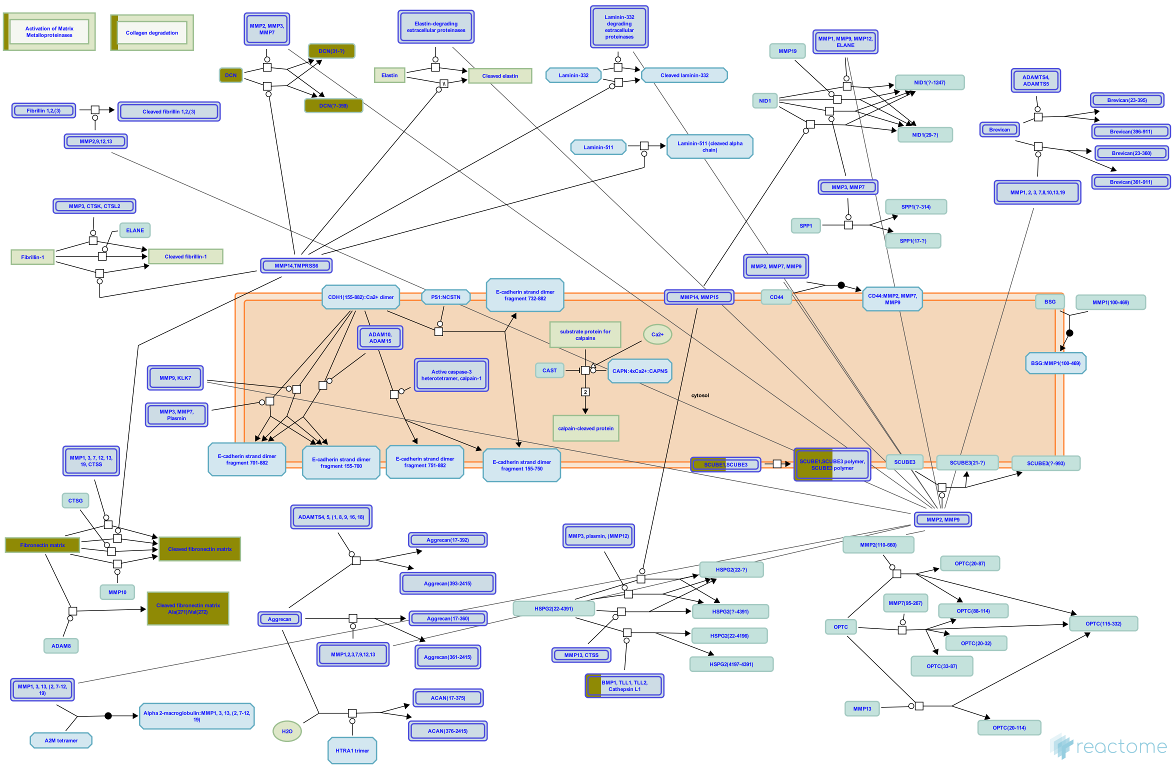


Matrix metalloproteinases (MMPs), previously referred to as matrixins because of their role in degradation of the extracellular matrix (ECM), are zinc and calcium dependent proteases belonging to the metzincin family. They contain a characteristic zinc-binding motif HEXXHXXGXXH (Stocker & Bode 1995) and a conserved Methionine which forms a Met-turn. Humans have 24 MMP genes giving rise to 23 MMP proteins, as MMP23 is encoded by two identical genes. All MMPs contain an N-terminal secretory signal peptide and a prodomain with a conserved PRCGXPD motif that in the inactive enzyme is localized with the catalytic site, the cysteine acting as a fourth unpaired ligand for the catalytic zinc atom. Activation involves delocalization of the domain containing this cysteine by a conformational change or proteolytic cleavage, a mechanism referred to as the cysteine-switch (Van Wart & Birkedal-Hansen 1990). Most MMPs are secreted but the membrane type MT-MMPs are membrane anchored and some MMPs may act on intracellular proteins. Various domains determine substrate specificity, cell localization and activation (Hadler-Olsen et al. 2011). MMPs are regulated by transcription, cellular location (most are not activated until secreted), activating proteinases that can be other MMPs, and by metalloproteinase inhibitors such as the tissue inhibitors of metalloproteinases (TIMPs). MMPs are best known for their role in the degradation and removal of ECM molecules. In addition, cleavage of the ECM and other cell surface molecules can release ECM-bound growth factors, and a number of non-ECM proteins are substrates of MMPs (Nagase et al. 2006). MMPs can be divided into subgroups based on domain structure and substrate specificity but it is clear that these are somewhat artificial, many MMPs belong to more than one functional group (Vise & Nagase 2003, Somerville et al. 2003).


##### Edit history

| Date | Action | Author |
|---|---|---|
| 2011-08-05 | Created | Jupe S |
| 2011-09-09 | Authored | Jupe S |
| 2012-02-21 | Edited | Jupe S |
| 2012-02-28 | Reviewed | D'Eustachio P |
| 2021-05-22 | Modified | Shorser S |

#### Entities found in this pathway (10)

| Input | UniProt Id | Input | UniProt Id | Input | UniProt Id |
|---|---|---|---|---|---|
| COL14A1 | Q05707 | COL15A1 | P39059 | COL1A2 | P08123 |
| COL5A1 | P20908 | COL8A2 | P25067, Q14050 | DCN | P07585 |
| FN1 | P02751 | SCUBE1 | Q8IWY4 | TIMP1 | P01033 |
| TLL2 | Q9Y6L7 | | | | |

#### 8. Extracellular matrix organization (R-HSA-1474244)

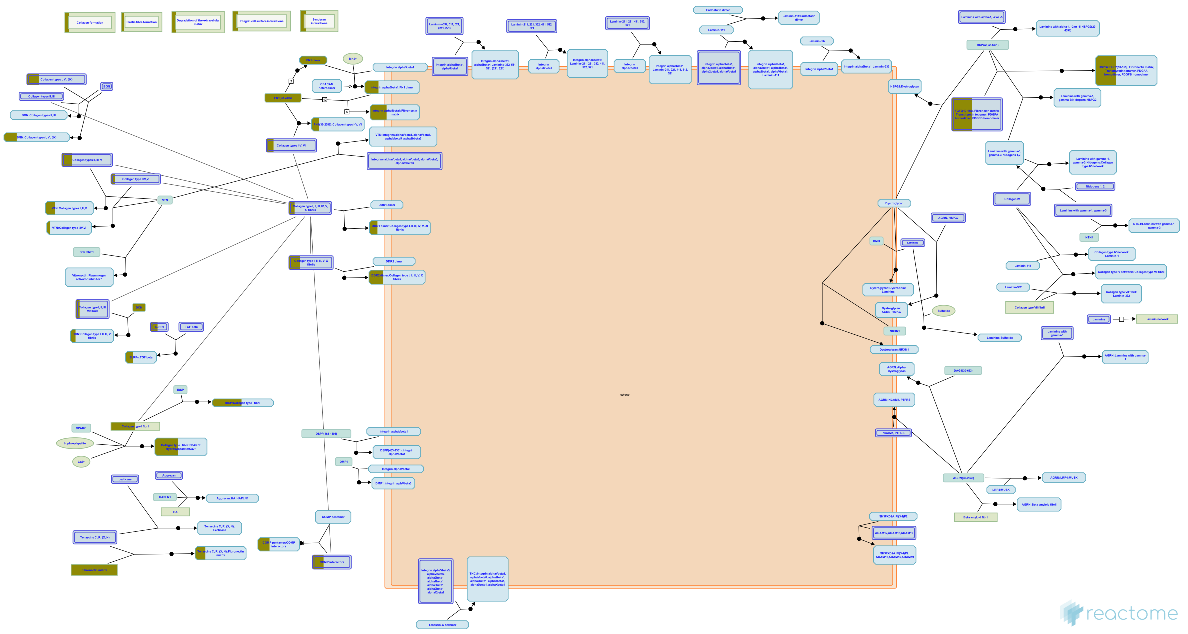

The extracellular matrix is a component of all mammalian tissues, a network consisting largely of the fibrous proteins collagen, elastin and associated-microfibrils, fibronectin and laminins embedded in a viscoelastic gel of anionic proteoglycan polymers. It performs many functions in addition to its structural role; as a major component of the cellular microenvironment it influences cell behaviours such as proliferation, adhesion and migration, and regulates cell differentiation and death (Hynes 2009).

| Date | Action | Author |
|---|---|---|
| 2021-05-22 | Modified | Shorser S |

#### Entities found in this pathway (16)

| Input | UniProt Id | Input | UniProt Id | Input | UniProt Id |
|---|---|---|---|---|---|
| ADAMTS2 | O95450 | COL14A1 | Q05707 | COL15A1 | P39059 |
| COL1A2 | P08123 | COL5A1 | P20908 | COL8A2 | P25067, Q14050 |
| DCN | P07585 | FN1 | P02751 | LUM | P51884 |
| PCOLCE | Q15113 | PDGFB | P01127 | SCUBE1 | Q8IWY4 |
| SDC1 | P18827 | TIMP1 | P01033 | TLL2 | Q9Y6L7 |
| VWF | P04275 | | | | |

#### 9. NOTCH4 Intracellular Domain Regulates Transcription (R-HSA-9013695)

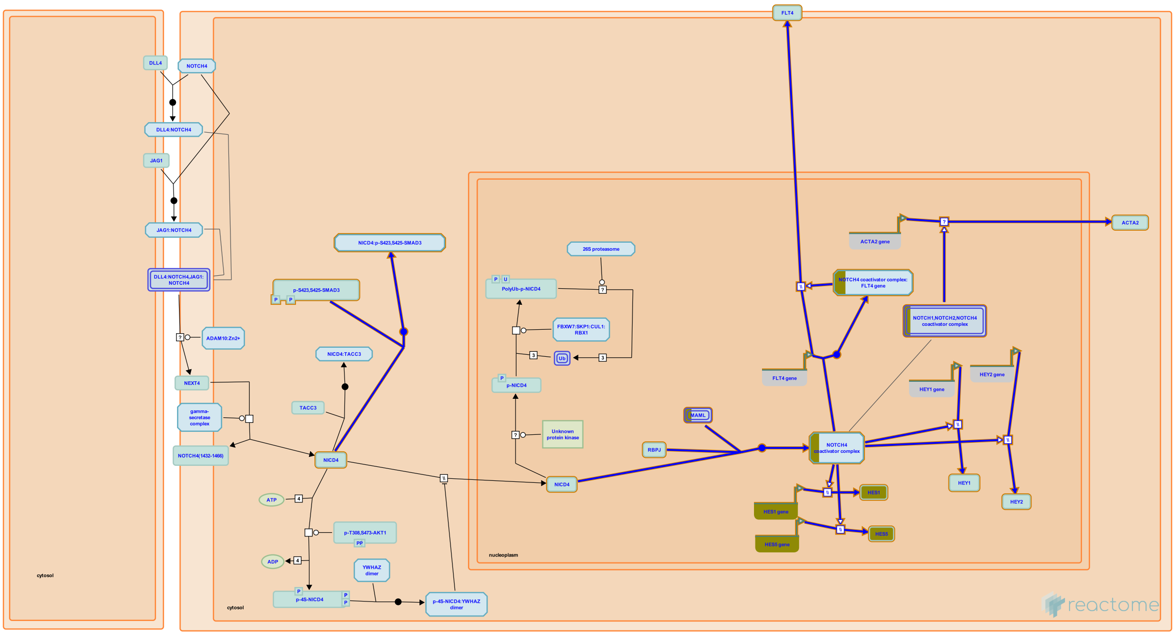


In the nucleus, NOTCH4 intracellular domain fragment (NICD4) binds transcription factors RBPJ (CSL) and mastermind family members (MAML1, MAML2 or MAML3) to form the NOTCH4 co-activator complex (Lin et al. 2002). The NOTCH4 coactivator complex stimulates transcription of well-established NOTCH targets HES1, HES5, HEY1 and HEY2 in a cellular context-dependent manner (Lin et al. 2002, Raafat et al.2004, Tsunematsu et al. 2004, Bargo et al. 2010). NOTCH4 also stimulates transcription of the FLT4 (VEGFR3) gene, encoding vascular endothelial growth factor receptor-3 (Shawber et al. 2007) and ACTA2 gene, encoding smooth muscle alpha actin (Tang et al. 2008).

NICD4 inhibits TGF-beta-induced SMAD-mediated transcription via binding of NICD4 to TGF-beta activated SMAD3 (Sun et al. 2005, Grabias and Konstantopoulos 2013).

##### Edit history

| Date | Action | Author |
|---|---|---|
| 2017-07-25 | Created | Orlic-Milacic M |
| 2018-04-05 | Authored | Orlic-Milacic M |
| 2018-05-01 | Reviewed | Haw R |
| 2018-05-09 | Modified | Orlic-Milacic M |
| 2018-05-09 | Edited | Orlic-Milacic M |

#### Entities found in this pathway (3)

| Input | UniProt Id | Input | UniProt Id | Input | UniProt Id |
|---|---|---|---|---|---|
| HES1 | Q14469 | HES5 | Q5TA89 | MAML3 | Q96JK9 |

| Input | Ensembl Id | Input | Ensembl Id |
|---|---|---|---|
| HES1 | ENSG00000114315 | HES5 | ENSG00000197921 |

#### 10. Collagen formation (R-HSA-1474290)

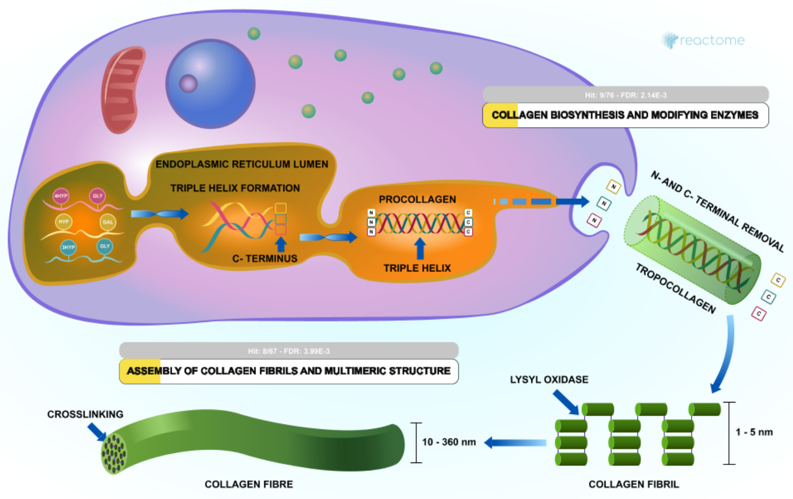


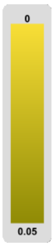

Collagen is a family of at least 29 structural proteins derived from over 40 human genes (Myllyharju & Kivirikko 2004). It is the main component of connective tissue, and the most abundant protein in mammals making up about 25% to 35% of whole-body protein content. A defining feature of collagens is the formation of trimeric left-handed polyproline II-type helical collagenous regions. The packing within these regions is made possible by the presence of the smallest amino acid, glycine, at every third residue, resulting in a repeating motif Gly-X-Y where X is often proline (Pro) and Y often 4-hydroxyproline (4Hyp). Gly-Pro-Hyp is the most common triplet in collagen (Ramshaw et al. 1998). Collagen peptide chains also have non-collagenous domains, with collagen subclasses having common chain structures. Collagen fibrils are mostly found in fibrous tissues such as tendon, ligament and skin. Other forms of collagen are abundant in cornea, cartilage, bone, blood vessels, the gut, and intervertebral disc. In muscle tissue, collagen is a major component of the endomysium, constituting up to 6% of muscle mass. Gelatin, used in food and industry, is collagen that has been irreversibly hydrolyzed.

| Input | UniProt Id | Input | UniProt Id | Input | UniProt Id |
|---|---|---|---|---|---|
| ADAMTS2 | O95450 | COL14A1 | Q05707 | COL15A1 | P39059 |
| COL1A2 | P08123 | COL5A1 | P20908 | COL8A2 | P25067, Q14050 |
| PCOLCE | Q15113 | TLL2 | Q9Y6L7 | | |

#### 11. Collagen chain trimerization (R-HSA-8948216)

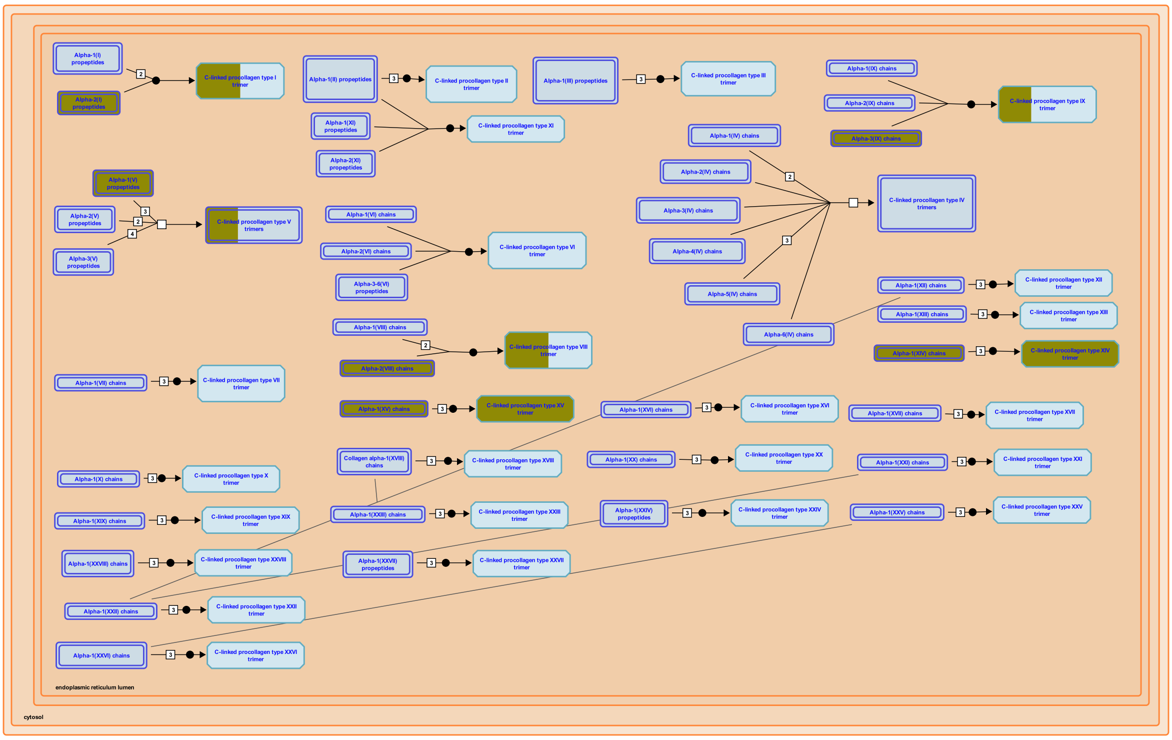


The C-propeptides of collagen propeptide chains are essential for the association of three peptide chains into a trimeric but non-helical procollagen. This initial binding event determines the composition of the trimer, brings the individual chains into the correct register and initiates formation of the triple helix at the C-terminus, which then proceeds towards the N-terminus in a zipper-like fashion (Engel & Prockop 1991). Most early refolding studies were performed with collagen type III, which contains a disulfide linkage at the C-terminus of its triple helix (Bächinger et al. 1978, Bruckner et al. 1978) that acts as a permanent linker even after removal of the non-collagenous domains.

Mutations within the C-propeptides further suggest that they are crucial for the correct interaction of the three polypeptide chains and for subsequent correct folding (refs. in Boudko et al. 2011).


##### Edit history

| Date | Action | Author |
|---|---|---|
| 2012-04-11 | Authored | Jupe S |
| 2012-05-24 | Reviewed | Canty-Laird EG |
| 2016-11-03 | Edited | Jupe S |

| Date | Action | Author |
|---|---|---|
| 2016-11-11 | Created | Jupe S |
| 2021-05-22 | Modified | Shorser S |

#### Entities found in this pathway (5)

| Input | UniProt Id | Input | UniProt Id | Input | UniProt Id |
|---|---|---|---|---|---|
| COL14A1 | Q05707 | COL15A1 | P39059 | COL1A2 | P08123 |
| COL5A1 | P20908 | COL8A2 | P25067, Q14050 | | |

#### 12. Signaling by NTRK1 (TRKA) (R-HSA-187037)

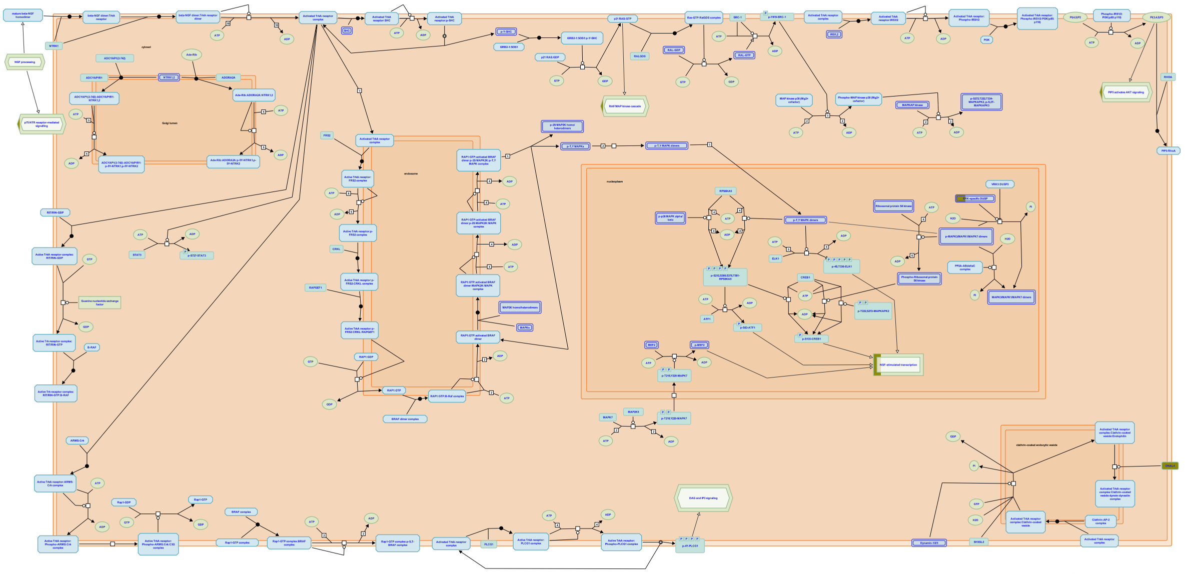

Trk receptors signal from the plasma membrane and from intracellular membranes, particularly from early endosomes. Signalling from the plasma membrane is fast but transient; signalling from endosomes is slower but long lasting. Signalling from the plasma membrane is annotated here. TRK signalling leads to proliferation in some cell types and neuronal differentiation in others. Proliferation is the likely outcome of short term signalling, as observed following stimulation of EGFR (EGF receptor). Long term signalling via TRK receptors, instead, was clearly shown to be required for neuronal differentiation in response to neurotrophins.


##### Edit history

| Date | Action | Author |
|---|---|---|
| 2006-09-07 | Created | Jassal B |
| 2006-10-10 | Edited | Jassal B |
| 2006-10-10 | Authored | Annibali D, Nasi S |
| 2007-11-08 | Reviewed | Greene LA |
| 2021-05-22 | Modified | Shorser S |

##### Entities found in this pathway (6)

| Input | UniProt Id |
| --- | --- |
| ARC | Q7LC44 |
| EGR4 | Q05215 |

| Input | UniProt Id |
| --- | --- |
| DNAL4 | O96015 |
| F3 | P13726 |

| Input | UniProt Id |
| --- | --- |
| DUSP6 | Q16828 |
| RRAD | P55042 |

| Input | Ensembl Id |
| --- | --- |
| ARC | ENSG00000198576 |
| F3 | ENSG00000117525 |

| Input | Ensembl Id |
| --- | --- |
| EGR4 | ENSG00000135625 |
| RRAD | ENSG00000166592 |

#### 13. NOTCH3 Intracellular Domain Regulates Transcription (R-HSA-9013508)

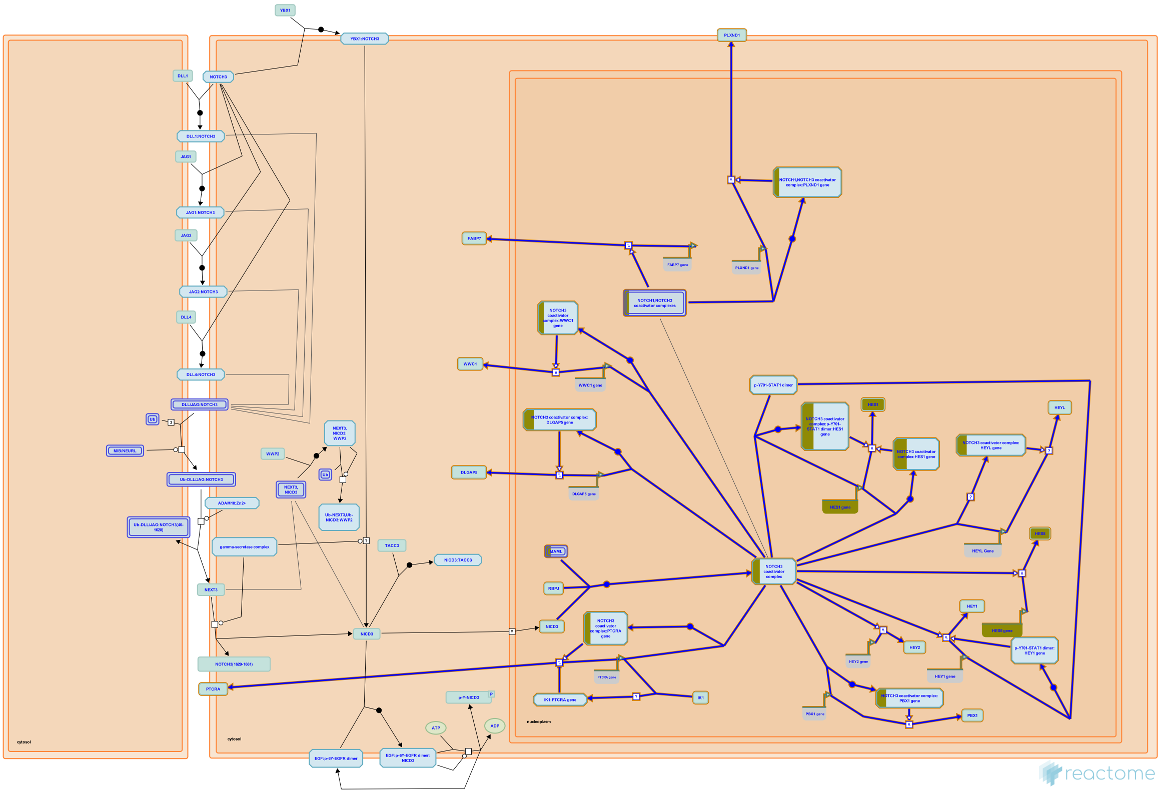

In the nucleus, NICD3 forms a complex with RBPJ (CBF1, CSL) and MAML (mastermind) proteins MAML1, MAML2 or MAML3 (possibly also MAMLD1). NICD3:RBPJ:MAML complex, also known as the NOTCH3 coactivator complex, activates transcription from RBPJ-binding promoter elements (Lin et al. 2002). While NOTCH1 prefers paired RBPJ binding sites, NOTCH3 preferentially binds to single RBPJ binding sites (Ong et al. 2006).

NOTCH3 coactivator complex induces transcription of the well established NOTCH target genes HES1 (Lin et al. 2002, Boelens et al. 2014), HEYL (Maier and Gessler 2000, Geimer Le Lay et al. 2014), HES5 (Lin te al. 2002, Shimizu et al. 2002), and HEY2 (Wang et al. 2002).

NOTCH3 positively regulates transcription of the pre-T-cell receptor alpha chain (PTCRA, commonly known as pT-alpha or pre-TCRalpha) (Talora et al. 2003, Bellavia et al. 2007). IK1, splicing isoform of the transcription factor Ikaros (IKZF1), competes with RBPJ for binding to the PTCRA promoter and inhibits PTCRA transcription. NOTCH3, through pre-TCR signaling, stimulates expression of the RNA binding protein HuD, which promotes splicing of IKZF1 into dominant negative isoforms. These dominant negative isoforms of IKZF1 heterodimerize with IK1, preventing its binding to target DNA sequences and thus contributing to sustained transcription of PTCRA (Bellavia et al. 2007, reviewed by Bellavia, Mecarrozzi, Campese, Grazioli, Gulino and Screpanti 2007).

NOTCH3-triggered pre-TCR-signaling downregulates the activity of the transcription factor TCF3 (E2A), through ERK-dependent induction of ID1. Inhibition of TCF3-mediated transcription downstream of NOTCH3 contributes to development of T-cell lymphomas in transgenic mice expressing NICD3 (Talora et al. 2003). Activation of ERKs downstream of NOTCH3-stimulated pre-TCR signaling leads to phosphorylation of the transcription factor TAL1, formation of the TAL1:SP1 complex, and activation of cyclin D1 (CCND1) transcription, which stimulates cell division (Talora et al. 2006).

NOTCH3 signaling can activate NF-kappaB (NFKB)-mediate transcription either indirectly, through activation of pre-TCR signaling, or directly, through association of NOTCH3 with IKKA. NFKB is constitutively active in T lymphoma cells derived from NOTCH3 transgenic mice (Vacca et al. 2006).

Transcription of the PLXND1 gene, encoding the semaphorin receptor Plexin D1, is directly stimulated by NOTCH1 and NOTCH3 coactivator complexes. PLXND1 is involved in neuronal migration and cancer cell invasiveness (Rehman et al. 2016). Expression of FABP7 (BLBP) in radial glia is positively regulated by NOTCH1 and NOTCH3 during neuronal migration (Anthony et al. 2005, Keilani and Sugaya 2008).

NOTCH3 gene is frequently amplified in ovarian cancer (Park et al. 2006). NOTCH3 coactivator complex directly stimulates DLGAP5 transcription. DLGAP5 is involved in G2/M transition and is overexpressed in ovarian cancer cells. (Chen et al. 2012). Another gene overexpressed in ovarian cancer whose transcription is directly stimulated by NOTCH3 is PBX1 (Park et al. 2008). The NOTCH3 coactivator complex directly stimulates WWC1 gene transcription. WWC1 gene encodes protein Kibra, involved in Hippo signaling. NOTCH3-mediated induction of WWC1 positively regulates Hippo signaling and inhibits epithelial-to-mesenchymal transition (EMT) in triple negative breast cancer cells (Zhang et al. 2016).


#### Edit history

| Date | Action | Author |
|---|---|---|
| 2017-07-25 | Created | Orlic-Milacic M |
| 2017-09-20 | Authored | Orlic-Milacic M |
| 2017-10-30 | Reviewed | Haw R |
| 2017-11-02 | Modified | Orlic-Milacic M |
| 2017-11-02 | Edited | Orlic-Milacic M |

#### Entities found in this pathway (3)

| Input | UniProt Id | Input | UniProt Id | Input | UniProt Id |
|---|---|---|---|---|---|
| HES1 | Q14469 | HES5 | Q5TA89 | MAML3 | Q96JK9 |

| Input | Ensembl Id | Input | Ensembl Id |
|---|---|---|---|
| HES1 | ENSG00000114315 | HES5 | ENSG00000197921 |

#### 14. Signaling by NOTCH2 (R-HSA-1980145)

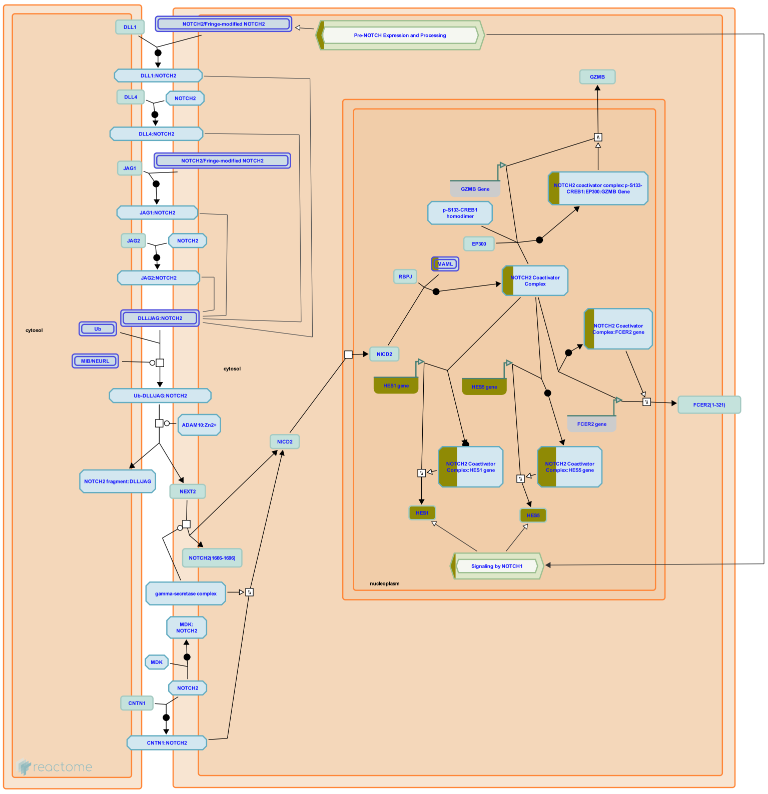


**Cellular compartments:** plasma membrane, cytosol, nucleoplasm.

NOTCH2 is activated by binding Delta-like and Jagged ligands (DLL/JAG) expressed in trans on neighboring cells (Shimizu et al. 1999, Shimizu et al. 2000, Hicks et al. 2000, Ji et al. 2004). In trans ligand-receptor binding is followed by ADAM10 mediated (Gibb et al. 2010, Shimizu et al. 2000) and gamma secretase complex mediated cleavage of NOTCH2 (Saxena et al. 2001, De Strooper et al. 1999), resulting in the release of the intracellular domain of NOTCH2, NICD2, into the cytosol. NICD2 traffics to the nucleus where it acts as a transcriptional regulator. For a recent review of the cannonical NOTCH signaling, please refer to Kopan and Ilagan 2009, D'Souza et al. 2010, Kovall and Blacklow 2010. CNTN1 (contactin 1), a protein involved in oligodendrocyte maturation (Hu et al. 2003) and MDK (midkine) (Huang et al. 2008, Gungor et al. 2011), which plays an important role in epithelial-to-mesenchymal transition, can also bind NOTCH2 and activate NOTCH2 signaling.

In the nucleus, NICD2 forms a complex with RBPJ (CBF1, CSL) and MAML (mastermind). The NICD2:RBPJ:MAML complex activates transcription from RBPJ binding promoter elements (RBEs) (Wu et al. 2000). NOTCH2 coactivator complexes directly stimulate transcription of HES1 and HES5 genes (Shimizu et al. 2002), both of which are known NOTCH1 targets. NOTCH2 but not NOTCH1 coactivator complexes, stimulate FCER2 transcription. Overexpression of FCER2 (CD23A) is a hallmark of B-cell chronic lymphocytic leukemia (B-CLL) and correlates with the malfunction of apoptosis, which is thought be an underlying mechanism of B-CLL development (Hubmann et al. 2002). NOTCH2 coactivator complexes together with CREBP1 and EP300 stimulate transcription of GZMB (granzyme B), which is important for the cytotoxic function of CD8+ T cells (Maekawa et al. 2008).

NOTCH2 gene expression is differentially regulated during human B-cell development, with NOTCH2 transcripts appearing at late developmental stages (Bertrand et al. 2000).

NOTCH2 mutations are a rare cause of Alagille syndrome (AGS). AGS is a dominant congenital multisystem disorder characterized mainly by hepatic bile duct abnormalities. Craniofacial, heart and kidney abnormalities are also frequently observed in the Alagille spectrum (Alagille et al. 1975). AGS is predominantly caused by mutations in JAG1, a NOTCH2 ligand (Oda et al. 1997, Li et al. 1997), but it can also be caused by mutations in NOTCH2 (McDaniell et al. 2006).

Hajdu-Cheney syndrome, an autosomal dominant disorder characterized by severe and progressive bone loss, is caused by NOTCH2 mutations that result in premature C-terminal NOTCH2 truncation, probably leading to increased NOTCH2 signaling (Simpson et al. 2011, Isidor et al. 2011, Majewski et al. 2011).


#### Edit history

| Date | Action | Author |
|---|---|---|
| 2004-12-15 | Reviewed | Joutel A |
| 2004-12-15 | Authored | Jassal B |
| 2011-11-09 | Created | Orlic-Milacic M |
| 2012-02-11 | Edited | Orlic-Milacic M |
| 2013-01-11 | Revised | Orlic-Milacic M |
| 2013-04-25 | Reviewed | Ilagan MXG, Boyle S |
| 2021-05-21 | Modified | Shorser S |

#### Entities found in this pathway (3)

| Input | UniProt Id | Input | UniProt Id | Input | UniProt Id |
|---|---|---|---|---|---|
| HES1 | Q14469 | HES5 | Q5TA89 | MAML3 | Q96JK9 |

| Input | Ensembl Id | Input | Ensembl Id |
|---|---|---|---|
| HES1 | ENSG00000114315 | HES5 | ENSG00000197921 |

#### 15. Signaling by NTRKs (R-HSA-166520)

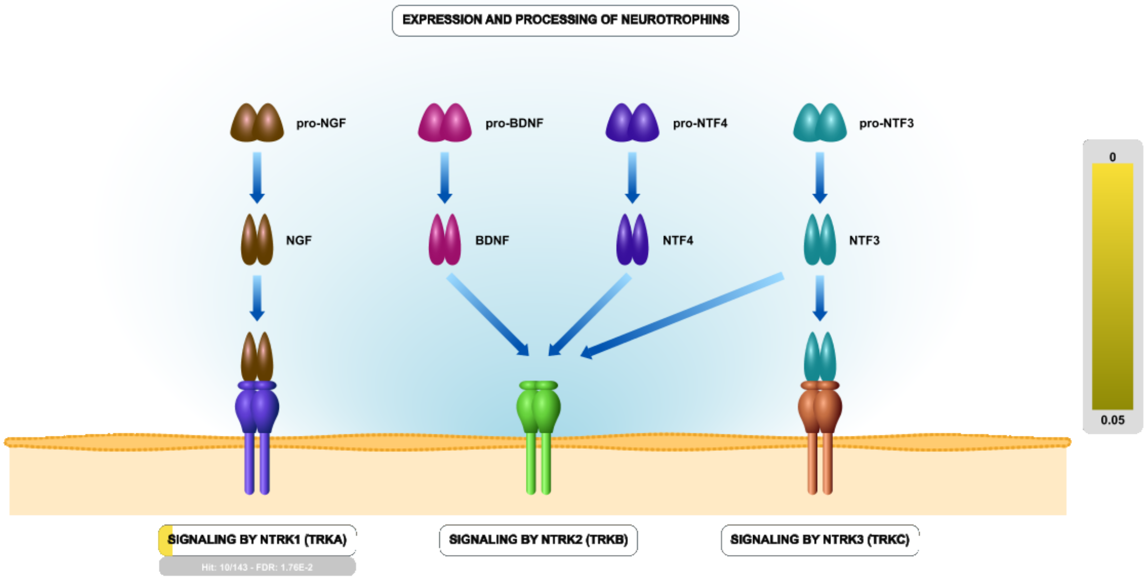


Neurotrophins (NGF, BDNF, NTF3 and NTF4) play pivotal roles in survival, differentiation, and plasticity of neurons in the peripheral and central nervous system. They are produced, and secreted in minute amounts, by a variety of tissues. They signal through two types of receptors: NTRK (TRK) tyrosine kinase receptors (TRKA, TRKB, TRKC), which differ in their preferred neurotrophin ligand, and p75NTR death receptor, which interacts with all neurotrophins. Besides the nervous system, TRK receptors and p75NTR are expressed in a variety of other tissues. For review, please refer to Bibel and Barde 2000, Poo 2001, Lu et al. 2005, Skaper 2012, Park and Poo 2013.

NTRK receptors, NTRK1 (TRKA), NTRK2 (TRKB) and NTRK3 (TRKC) are receptor tyrosine kinases activated by ligand binding to their extracellular domain. Ligand binding induces receptor dimerization, followed by trans-autophosphorylation of dimerized receptors on conserved tyrosine residues in the cytoplasmic region. Phosphorylated tyrosines in the intracellular domain of the receptor serve as docking sites for adapter proteins, triggering downstream signaling cascades.

NTRK1 (TRKA) is the receptor for the nerve growth factor (NGF). NGF is primarily secreted by tissues that are innervated by sensory and sympathetic neurons. NTRK1 signaling promotes growth and survival of neurons during embryonic development and maintenance of neuronal cell integrity in adulthood (reviewed by Marlin and Li 2015).

Brain-derived neurotrophic factor (BDNF) and neurotrophin-4 (NTF4, also known as NT-4) are two high affinity ligands for NTRK2 (TRKB). Neurotrophin-3 (NTF3, also known as NT-3) binds to NTRK2 with low affinity and may not be a physiologically relevant ligand. Nerve growth factor (NGF), a high affinity ligand for NTRK1, does not interact with NTRK2. NTRK2 signaling is implicated in neuronal development in both the peripheral (PNS) and central nervous system (CNS) and may play a role in long-term potentiation (LTP) and learning (reviewed by Minichiello 2009). NTRK2 may modify neuronal excitability and synaptic transmission by directly phosphorylating voltage gated channels (Rogalski et al. 2000).

NTF3 (NT-3) is the ligand for NTRK3 (TRKC). Signaling downstream of activated NTRK3, regulates cell survival, proliferation and motility. In the absence of its ligand, NTRK3 functions as a dependence receptor and triggers BAX and CASP9-dependent cell death (Tauszig-Delamasure et al. 2007, Ichim et al. 2013).


#### Edit history

| Date | Action | Author |
|---|---|---|
| 2005-09-07 | Created | Jassal B |
| 2006-10-10 | Edited | Jassal B |
| 2006-10-10 | Authored | Annibali D, Nasi S |
| 2007-11-08 | Reviewed | Greene LA |
| 2021-05-22 | Modified | Shorser S |

#### Entities found in this pathway (6)

| Input | UniProt Id | Input | UniProt Id | Input | UniProt Id |
|---|---|---|---|---|---|
| ARC | Q7LC44 | DNAL4 | O96015 | DUSP6 | Q16828 |
| EGR4 | Q05215 | F3 | P13726 | RRAD | P55042 |

| Input | Ensembl Id | Input | Ensembl Id |
|---|---|---|---|
| ARC | ENSG00000198576 | EGR4 | ENSG00000135625 |
| F3 | ENSG00000117525 | RRAD | ENSG00000166592 |

#### 16. Integrin cell surface interactions (R-HSA-216083)

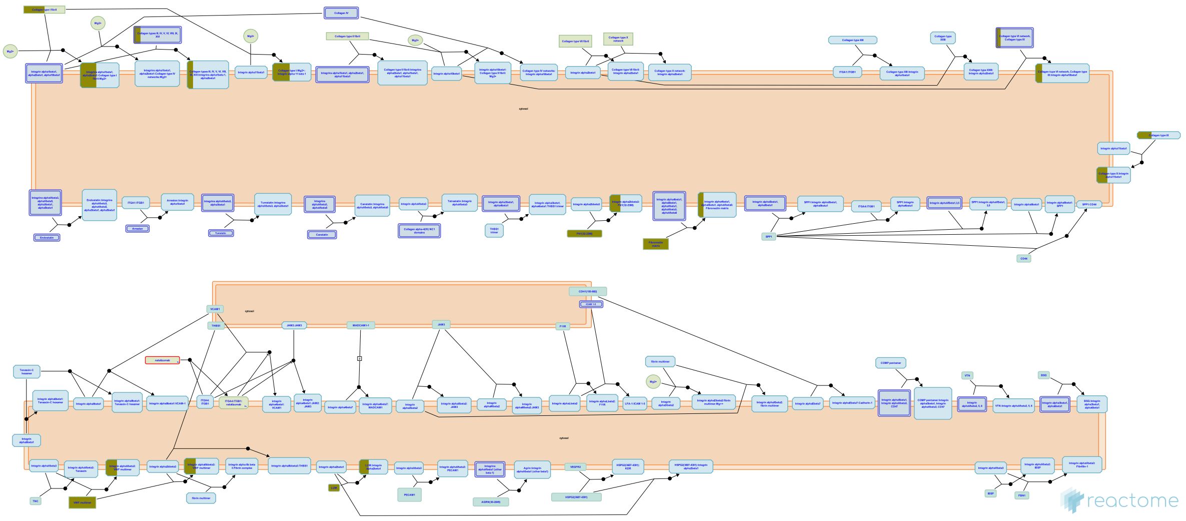

The extracellular matrix (ECM) is a network of macro-molecules that underlies all epithelia and endothelia and that surrounds all connective tissue cells. This matrix provides the mechanical strength and also influences the behavior and differentiation state of cells in contact with it. The ECM are diverse in composition, but they generally comprise a mixture of fibrillar proteins, polysaccharides synthesized, secreted and organized by neighboring cells. Collagens, fibronectin, and laminins are the principal components involved in cell matrix interactions; other components, such as vitronectin, thrombospondin, and osteopontin, although less abundant, are also important adhesive molecules.

Integrins are the receptors that mediate cell adhesion to ECM. Integrins consists of one alpha and one beta subunit forming a noncovalently bound heterodimer. 18 alpha and 8 beta subunits have been identified in humans that combine to form 24 different receptors.

The integrin dimers can be broadly divided into three families consisting of the beta1, beta2/beta7, and beta3/alphaV integrins. beta1 associates with 12 alpha-subunits and can be further divided into RGD-, collagen-, or laminin binding and the related alpha4/alpha9 integrins that recognise both matrix and vascular ligands. beta2/beta7 integrins are restricted to leukocytes and mediate cell-cell rather than cell-matrix interactions, although some recognize fibrinogen. The beta3/alphaV family members are all RGD receptors and comprise aIIbb3, an important receptor on platelets, and the remaining b-subunits, which all associate with alphaV. It is the collagen receptors and leukocyte-specific integrins that contain alpha A-domains.

#### Edit history

| Date | Action | Author |
|---|---|---|
| 2008-03-11 | Edited | Garapati P V |
| 2008-03-11 | Created | Garapati P V |
| 2008-05-07 | Reviewed | Hynes R, Humphries MJ, Yamada KM |
| 2008-05-07 | Authored | Geiger B, Horwitz AR |
| 2012-08-08 | Authored | Jupe S |
| 2013-08-13 | Edited | Jupe S |
| 2013-08-13 | Reviewed | Ricard-Blum S |
| 2021-05-22 | Modified | Shorser S |

#### Entities found in this pathway (6)

| Input | UniProt Id | Input | UniProt Id | Input | UniProt Id |
|---|---|---|---|---|---|
| COL1A2 | P08123 | COL5A1 | P20908 | COL8A2 | P25067, Q14050 |
| FN1 | P02751 | LUM | P51884 | VWF | P04275 |

#### 17. GP1b-IX-V activation signalling (R-HSA-430116)

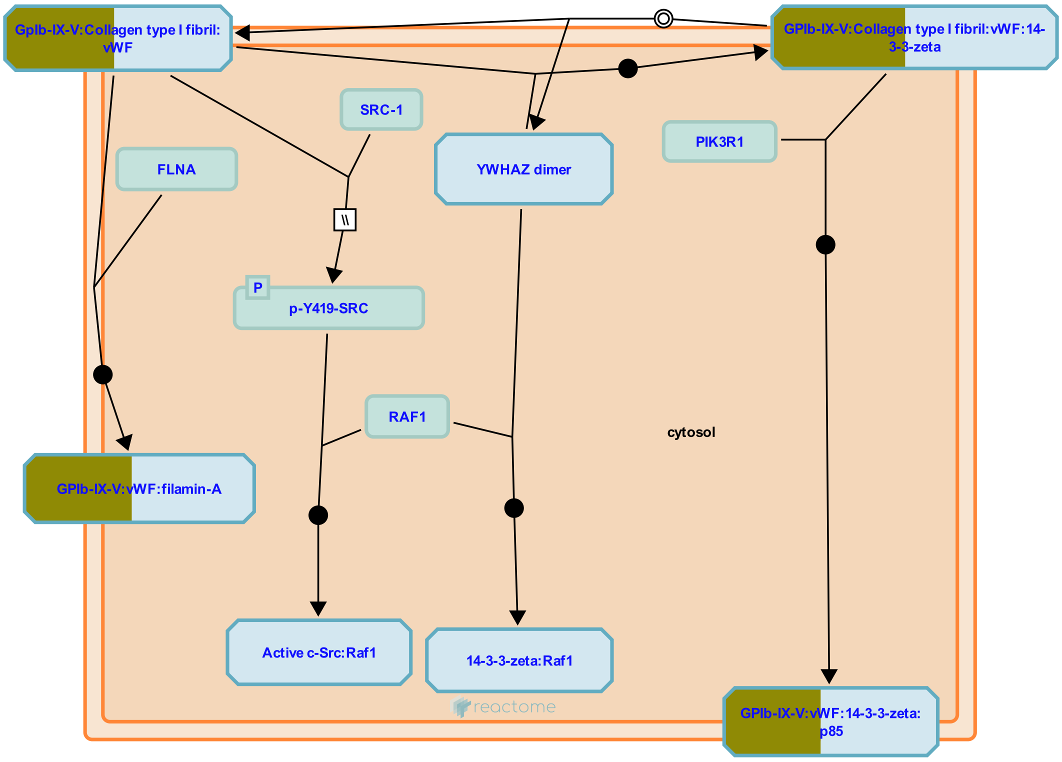


**Cellular compartments:** plasma membrane.

The platelet GPIb complex (GP1b-IX-V) together with GPVI are primarily responsible for regulating the initial adhesion of platelets to the damaged blood vessel and platelet activation. The importance of GPIb is demonstrated by the bleeding problems in patients with Bernard-Soulier syndrome where this receptor is either absent or defective. GP1b-IX-V binds von Willebrand Factor (vWF) to resting platelets, particularly under conditions of high shear stress. This transient interaction is the first stage of the vascular repair process. Activation of GP1b-IX-V on exposure of the fibrous matrix following atherosclerotic plaque rupture, or in occluded arteries, is a major contributory factor leading to thrombus formation leading to heart attack or stroke.
GpIb also binds thrombin (Yamamoto et al. 1986), at a site distinct from the site of vWF binding, acting as a docking site for thrombin which then activates Proteinase Activated Receptors leading to enhanced platelet activation (Dormann et al. 2000).


##### Edit history

| Date | Action | Author |
|---|---|---|
| 2009-06-03 | Authored | Akkerman JW |
| 2009-07-30 | Created | Jupe S |
| 2010-06-07 | Edited | Jupe S |

| Date | Action | Author |
| --- | --- | --- |
| 2010-06-07 | Reviewed | Kunapuli SP |
| 2021-05-22 | Modified | Shorser S |

#### Entities found in this pathway (3)

| Input | UniProt Id | Input | UniProt Id | Input | UniProt Id |
| --- | --- | --- | --- | --- | --- |
| COL1A2 | P08123 | TSKU | P07359 | VWF | P04275 |

#### 18. Collagen degradation (R-HSA-1442490)

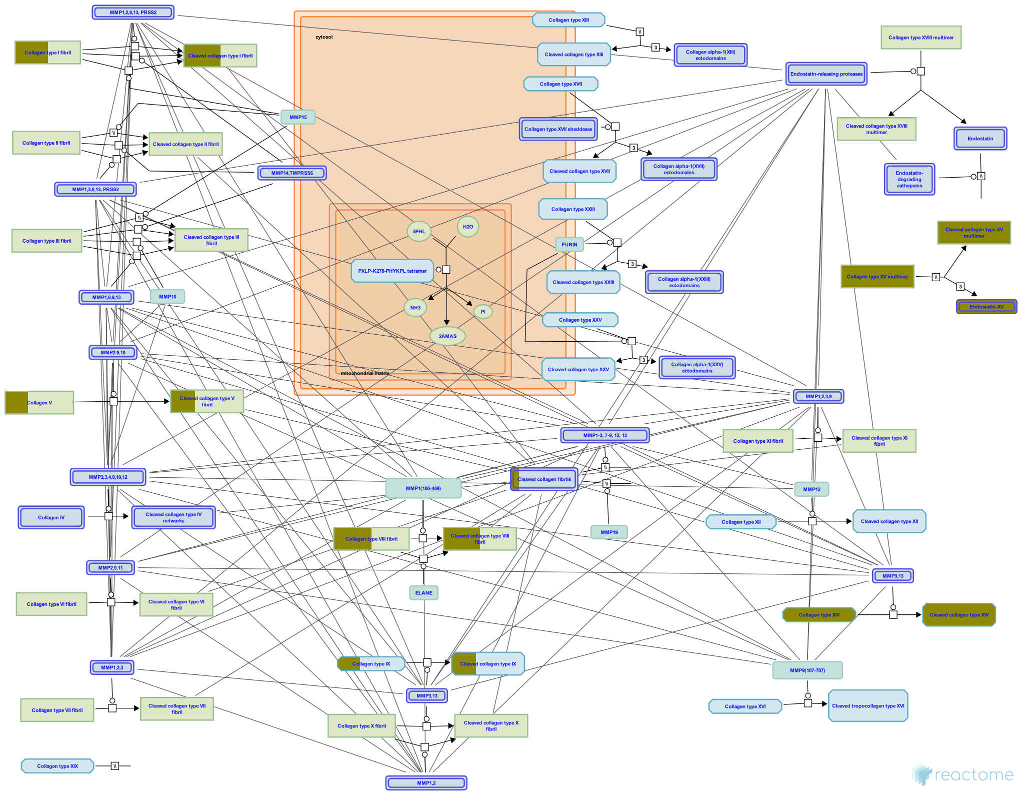

Collagen fibril diameter and spatial organisation are dependent on the species, tissue type and stage of development (Parry 1988). The lengths of collagen fibrils in mature tissues are largely unknown but in tendon can be measured in millimetres (Craig et al. 1989). Collagen fibrils isolated from adult bovine corneal stroma had ~350 collagen molecules in transverse section, tapering down to three molecules at the growing tip (Holmes & Kadler 2005).


#### Edit history

| Date | Action | Author |
|---|---|---|
| 2011-07-12 | Authored | Jupe S |
| 2011-07-12 | Created | Jupe S |
| 2012-10-08 | Reviewed | Sorsa T |
| 2012-11-12 | Edited | Jupe S |
| 2021-05-22 | Modified | Shorser S |

#### Entities found in this pathway (5)

| Input | UniProt Id | Input | UniProt Id | Input | UniProt Id |
|---|---|---|---|---|---|
| COL14A1 | Q05707 | COL15A1 | P39059 | COL1A2 | P08123 |
| COL5A1 | P20908 | COL8A2 | P25067, Q14050 | | |

#### 19. RAF-independent MAPK1/3 activation (R-HSA-112409)

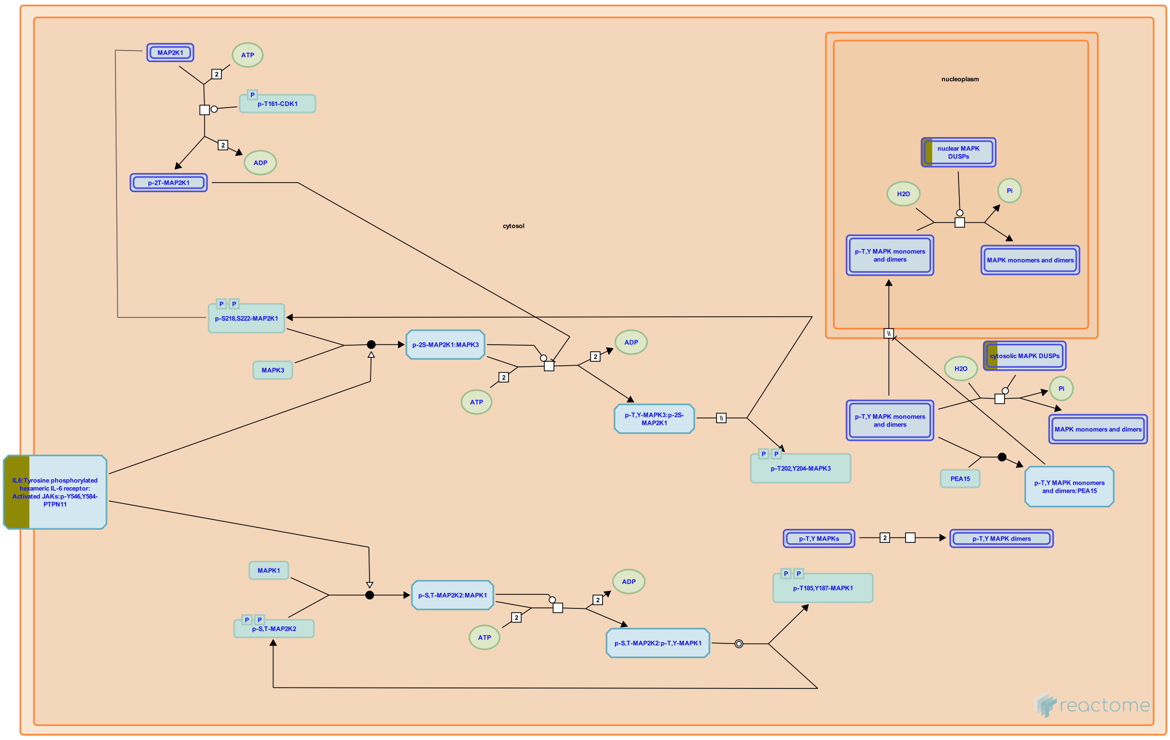


**Cellular compartments:** cytosol, nucleoplasm.

Depending upon the stimulus and cell type mitogen-activated protein kinases (MAPK) signaling pathway can transmit signals to regulate many different biological processes by virtue of their ability to target multiple effector proteins (Kyriakis JM & Avruch J 2012; Yoon and Seger 2006; Shaul YD & Seger R 2007; Arthur JS & Ley SC 2013). In particular, the extracellular signal-regulated kinases MAPK3(ERK1) and MAPK1 (ERK2) are involved in diverse cellular processes such as proliferation, differentiation, regulation of inflammatory responses, cytoskeletal remodeling, cell motility and invasion through the increase of matrix metalloproteinase production (Viala E & Pouyssegur J 2004; Hsu MC et al. 2006; Dawson CW et al.2008; Kuriakose T et al. 2014).The canonical RAF:MAP2K:MAPK1/3 cascade is stimulated by various extracellular stimuli including hormones, cytokines, growth factors, heat shock and UV irradiation triggering the GEF-mediated activation of RAS at the plasma membrane and leading to the activation of the RAF MAP3 kinases. However, many physiological and pathological stimuli have been found to activate MAPK1/3 independently of RAF and RAS (Dawson CW et al. 2008; Wang J et al. 2009; Kuriakose T et al. 2014). For example, AMP-activated protein kinase (AMPK), but not RAF1, was reported to regulate MAP2K1/2 and MAPK1/3 (MEK and ERK) activation in rat hepatoma H4IIE and human erythroleukemia K562 cells in response to autophagy stimuli (Wang J et al. 2009). Tumor progression locus 2 (TPL2, also known as MAP3K8 and COT) is another MAP3 kinase which promotes MAPK1/3 (ERK)-regulated immune responses downstream of toll-like receptors (TLR), TNF receptor and IL1beta signaling pathways (Gantke T et al. 2011).

In response to stimuli the cell surface receptors transmit signals inducing MAP3 kinases, e.g., TPL2, MEKK1, which in turn phosphorylate MAP2Ks (MEK1/2). MAP2K then phosphorylate and activate the MAPK1/3 (ERK1 and ERK2 MAPKs). Activated MAPK1/3 phosphorylate and regulate the activities of an ever growing pool of substrates that are estimated to comprise over 160 proteins (Yoon and Seger 2006). The majority of ERK substrates are nuclear proteins, but others are found in the cytoplasm and other organelles. Activated MAPK1/3 can translocate to the nucleus, where they phosphorylate and regulate various transcription factors, such as Ets family transcription factors (e.g., ELK1), ultimately leading to changes in gene expression (Zuber J et al. 2000).


#### Edit history

| Date | Action | Author |
|---|---|---|
| 2004-04-29 | Created | Charalambous M |
| 2007-11-08 | Reviewed | Greene LA |
| 2021-05-31 | Modified | Shorser S |

#### Entities found in this pathway (3)

| Input | UniProt Id | Input | UniProt Id | Input | UniProt Id |
|---|---|---|---|---|---|
| DUSP1 | P28562 | DUSP6 | Q16828 | IL6R | P08887, P08887-2 |

#### 20. Syndecan interactions (R-HSA-3000170)

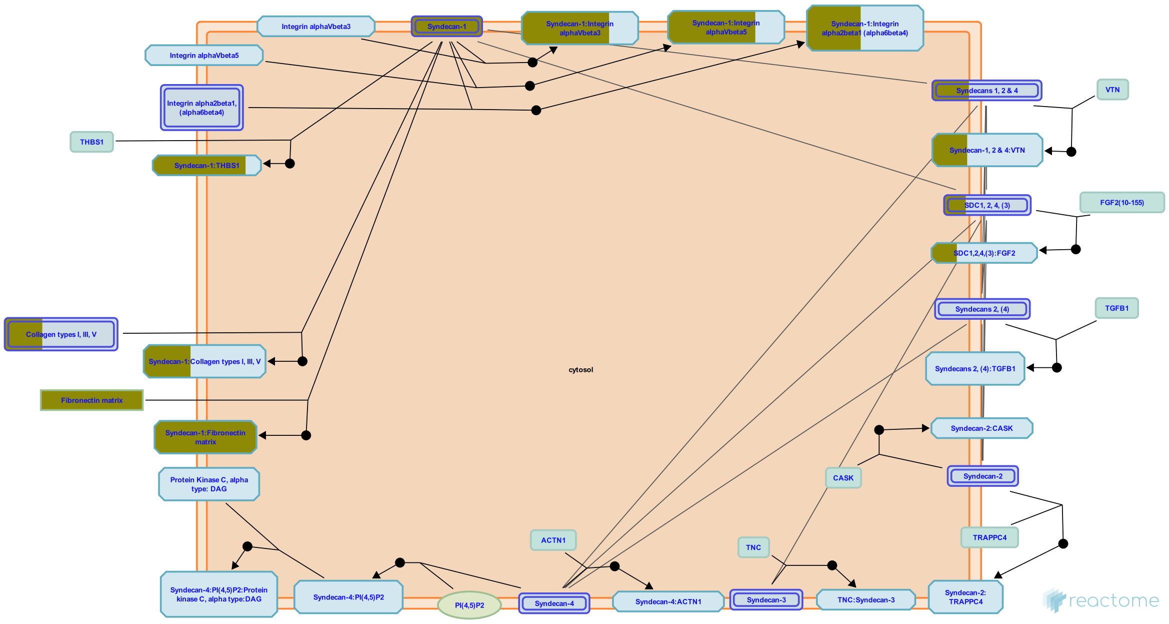


Syndecans are type I transmembrane proteins, with an N-terminal ectodomain that contains several consensus sequences for glycosaminoglycan (GAG) attachment and a short C-terminal cytoplasmic domain. Syndecan-1 and -3 GAG attachment sites occur in two distinct clusters, one near the N-terminus and the other near the membrane-attachment site, separated by a proline and threonine-rich 'spacer'. Syndecan ectodomain sequences are poorly conserved in the family and between species, but the transmembrane and cytoplasmic domains are highly conserved. Syndecan-1 and -3 form a subfamily. Syndecan core proteins form dimers (Choi et al. 2007) and at least syndecan-3 and -4 form oligomers (Asundi & Carey 1995, Shin et al. 2012). Syndecan-1 is the major syndecan of epithelial cells including vascular endothelium. Syndecan-2 is present mostly in mesenchymal, neuronal and smooth muscle cells. Syndecan-3 is the major syndecan of the nervous system, while syndecan-4 is ubiquitously expressed but at lower levels than the other syndecans (refs in Alexopoulou et al. 2007). The core syndecan protein has three to five heparan sulfate or chondroitin sulfate chains, which interact with a variety of ligands including fibroblast growth factors, vascular endothelial growth factor, transforming growth factor-beta, fibronectin, collagen, vitronectin and several integrins. Syndecans may act as integrin coreceptors. Interactions between fibronectin and syndecans are modulated by tenascin-C.

Syndecans bind a wide variety of soluble and insoluble ligands, inckluding extracellular matrix components, cell adhesion molecules, growth factors, cytokines, and proteinases. As the cleaved ectodomains of syndecans retain the ability to bind ligands, ectodomain shedding is a mechanism for releasing soluble effectors that may compete for ligands with their cell-bound counterparts (Kainulainen et al. 1998). Shed ectodomains are found in inflammatory fluids (Subramanian et al. 1997) and may induce the proliferation of cancer cells (Maeda et al. 2004).

#### Edit history

| Date | Action | Author |
|---|---|---|
| 2012-07-31 | Authored | Jupe S |
| 2013-01-24 | Created | Jupe S |
| 2013-04-26 | Edited | Jupe S |
| 2013-05-22 | Reviewed | Ricard-Blum S, Fuentes J |
| 2021-05-22 | Modified | Shorser S |

#### Entities found in this pathway (4)

| Input | UniProt Id | Input | UniProt Id |
|---|---|---|---|
| COL1A2 | P08123 | COL5A1 | P20908 |
| FN1 | P02751 | SDC1 | P18827 |

#### 21. LGI-ADAM interactions (R-HSA-5682910)

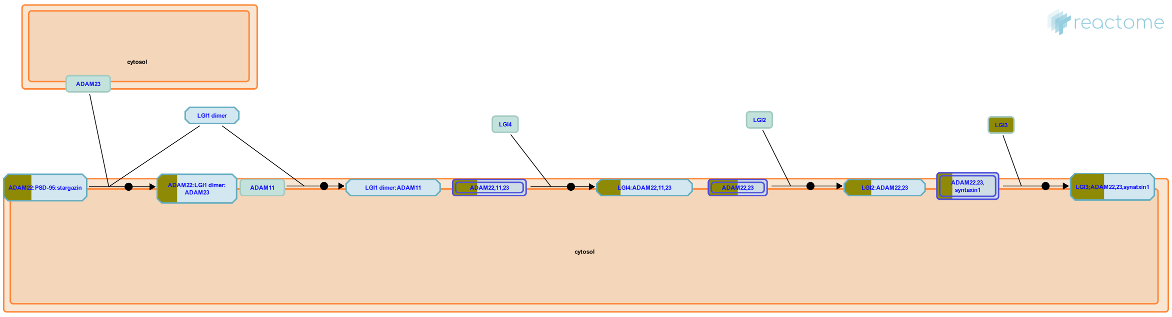


**Cellular compartments:** extracellular region.

Synapse formation and maturation require multiple interactions between presynaptic and postsynaptic neurons. These interactions are mediated by a diverse set of synaptogenic proteins (Kegel et al. 2013, Siddiqui & Craig 2011). Initial synapse formation needs both the binding of secreted proteins to presynaptic and postsynaptic receptors, and the direct binding between presynaptic and postsynaptic transmembrane proteins. One class of molecules that plays an important role in cellular interactions in nervous system development and function is the leucine-rich glioma inactivated (LGI) protein family. These are secreted synaptogenic proteins consisting of an LRR (leucine-rich repeat) domain and a epilepsy-associated or EPTP (epitempin) domain (Gu et al. 2002). Both protein domains are generally involved in protein-protein interactions. Genetic and biochemical evidence suggests that the mechanism of action of LGI proteins involves binding to a subset of cell surface receptors belonging to the ADAM (a disintegrin and metalloproteinase) family, i.e. ADAM11, ADAM22 and ADAM23. These interactions play crucial role in the development and function of the vertebrate nervous system mainly mediating synaptic transmission and myelination (Kegel et al. 2013, Novak 2004, Seals & Courtneidge 2003).

##### Edit history

| Date | Action | Author |
|---|---|---|
| 2015-03-11 | Edited | Garapati P V |
| 2015-03-11 | Authored | Garapati P V |
| 2015-03-11 | Created | Garapati P V |
| 2015-04-20 | Reviewed | Meijer D |
| 2021-05-21 | Modified | Shorser S |

##### Entities found in this pathway (3)

| Input | UniProt Id | Input | UniProt Id | Input | UniProt Id |
|---|---|---|---|---|---|
| CACNG4 | Q9UBN1 | LGI3 | Q8N145 | MMD | Q9P0K1 |

#### 22. Signaling by Interleukins (R-HSA-449147)

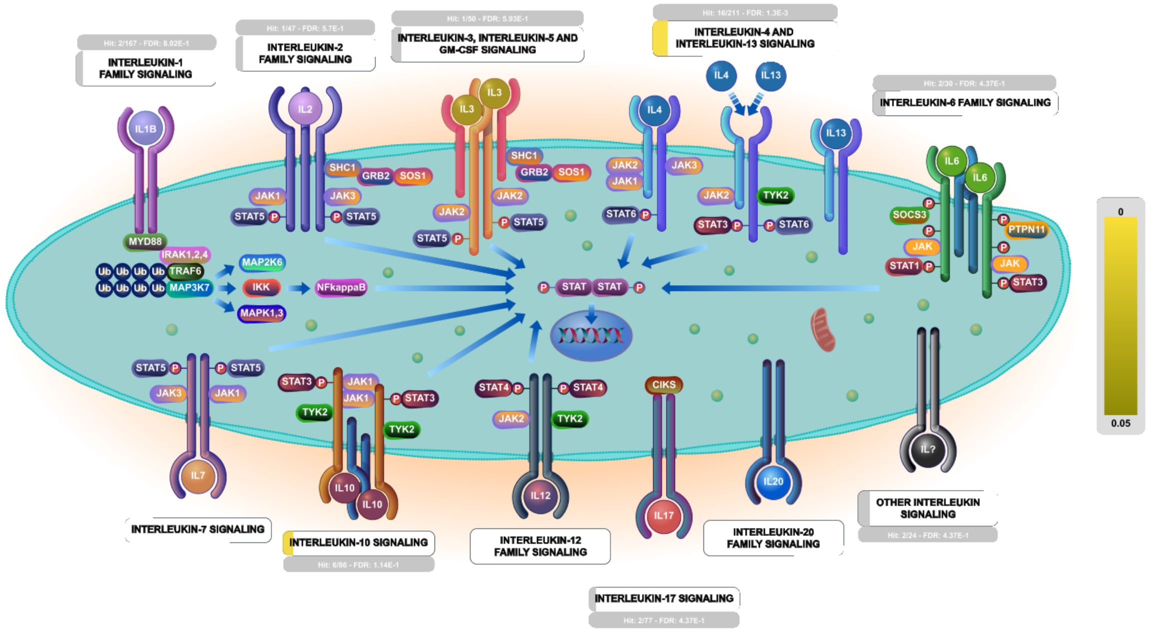


**Cellular compartments:** plasma membrane.

Interleukins are low molecular weight proteins that bind to cell surface receptors and act in an autocrine and/or paracrine fashion. They were first identified as factors produced by leukocytes but are now known to be produced by many other cells throughout the body. They have pleiotropic effects on cells which bind them, impacting processes such as tissue growth and repair, hematopoietic homeostasis, and multiple levels of the host defense against pathogens where they are an essential part of the immune system.

##### Edit history

| Date | Action | Author |
|---|---|---|
| 2009-11-27 | Created | Jupe S |
| 2010-05-17 | Reviewed | Pinteaux E |
| 2010-05-17 | Authored | Ray KP |
| 2010-05-26 | Edited | Jupe S |
| 2021-05-22 | Modified | Shorser S |

##### Entities found in this pathway (13)

| Input | UniProt Id | Input | UniProt Id | Input | UniProt Id |
|---|---|---|---|---|---|
| COL1A2 | P08123 | CSF2RA | P15509 | DUSP6 | Q16828 |
| FN1 | P02751 | IL1A | P01583 | IL6R | P08887, P08887-2 |
| MAP2K6 | P52564 | NOS2 | P35228 | PTGS2 | P35354 |
| SDC1 | P18827 | TIMP1 | P01033 | VAMP1 | P63027 |
| VEGFA | P15692 | | | | |

| Input | Ensembl Id | Input | Ensembl Id | Input | Ensembl Id |
|---|---|---|---|---|---|
| COL1A2 | ENSG00000164692 | FN1 | ENSG00000115414 | IL1A | ENSG00000115008 |
| IL6R | ENSG00000160712 | NOS2 | ENSG00000007171 | PTGS2 | ENSG00000073756 |
| TIMP1 | ENSG00000102265 | VEGFA | ENSG00000112715 | | |

#### 23. RUNX3 regulates NOTCH signaling (R-HSA-8941856)

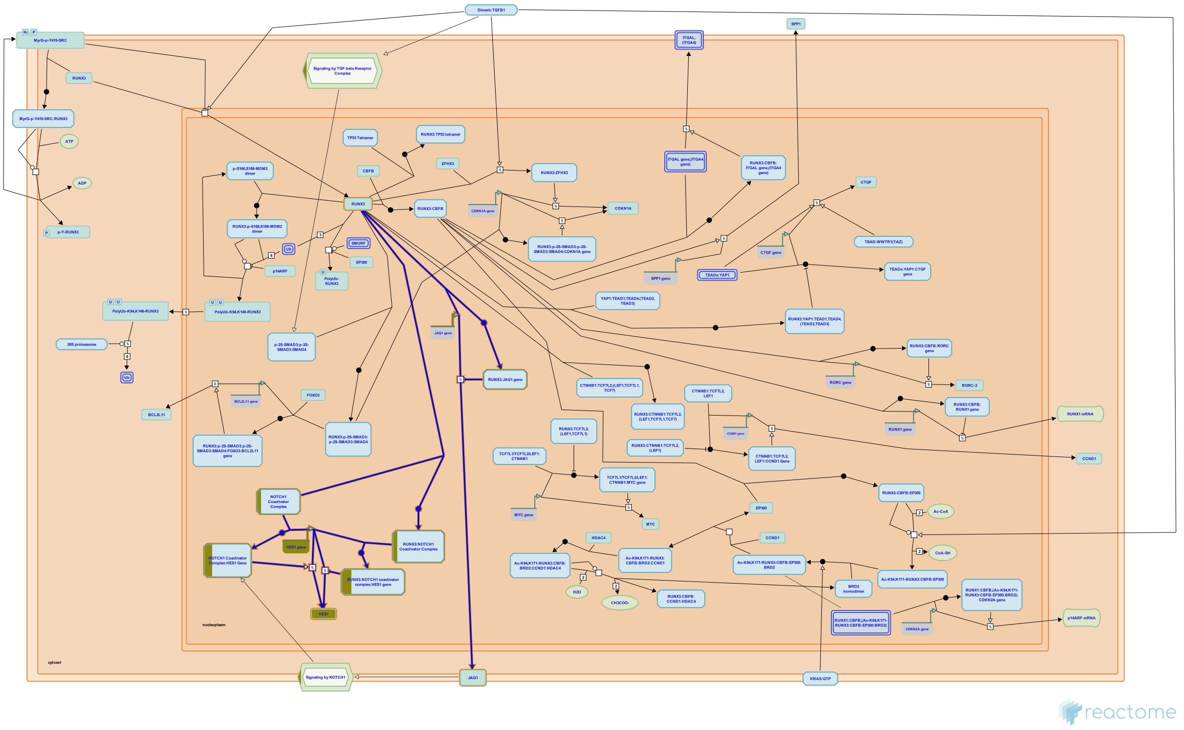

RUNX3 negatively regulates NOTCH signaling, which contributes to the tumor suppressor role of RUNX3 in hepatocellular carcinoma. RUNX3 binds the promoter of the JAG1 gene, encoding NOTCH ligand JAG1 and inhibits its transcription (Nishina et al. 2011). In addition, RUNX3 also binds to the NOTCH1 coactivator complex at the promoter of HES1, a NOTCH target gene, and inhibits HES1 transcription (Gao et al. 2010).


##### Edit history

| Date | Action | Author |
|---|---|---|
| 2016-10-06 | Created | Orlic-Milacic M |
| 2016-12-13 | Authored | Orlic-Milacic M |
| 2017-01-31 | Edited | Orlic-Milacic M |
| 2017-01-31 | Reviewed | Ito Y, Chuang LS |
| 2021-05-21 | Modified | Shorser S |

##### Entities found in this pathway (2)

| Input | UniProt Id |
|---|---|
| HES1 | Q14469 |

| Input | UniProt Id |
|---|---|
| MAML3 | Q96JK9 |

| Input | Ensembl Id |
|---|---|
| HES1 | ENSG00000114315 |

#### 24. Platelet Adhesion to exposed collagen (R-HSA-75892)

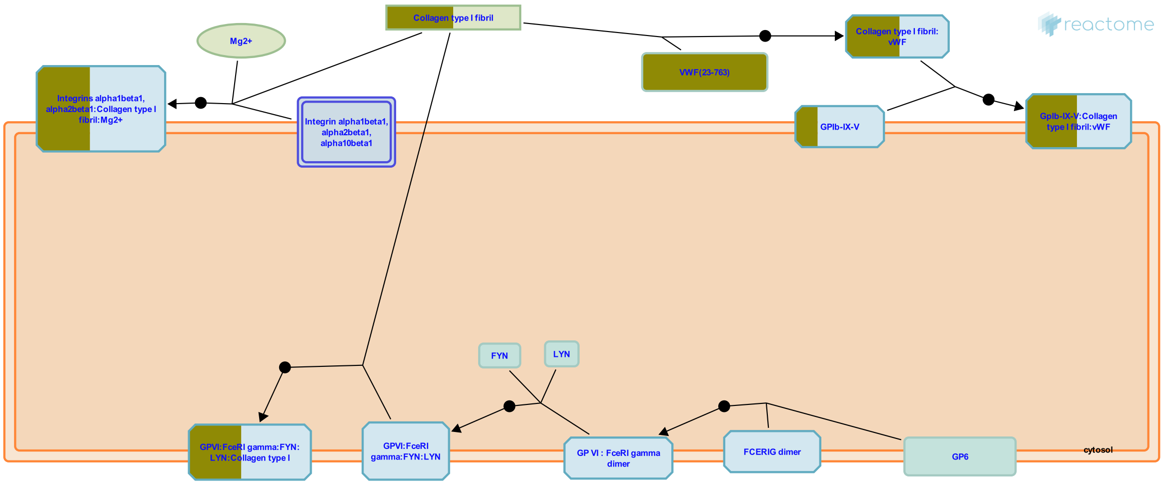


Initiation of platelet adhesion is the first step in the formation of the platelet plug. Circulating platelets are arrested and subsequently activated by exposed collagen and vWF. It is not entirely clear which type of collagen is responsible for adhesion and activation; collagen types I and III are abundant in vascular epithelia but several other types incluing IV are present (Farndale 2006). Several collagen binding proteins are expressed on platelets, including integrin alpha2 beta1, GPVI, and GPIV. Integrin alpha2 beta1, known on leukocytes as VLA-2, is the major platelet collagen receptor (Kunicki et al. 1988). It requires Mg2+ to interact with collagen and may require initiation mediated by the activation of integrin alphaIIb beta3 (van de Walle 2007). Binding occurs via the alpha2 subunit I domain to a collagen motif with the sequence Gly-Phe-Hyp-Gly-Glu-Arg (Emsley 2000). Binding of collagen to alpha2 beta1 generates intracellular signals that contribute to platelet activation. These facilitate the engagement of the lower-affinity collagen receptor, GPVI (Keely 1996), the key receptor involved in collagen-induced platelet activation. The GPVI receptor is a complex of the GPVI protein with a dimer of Fc epsilon R1 gamma (FceRI gamma). The Src family kinases Fyn and Lyn constitutively associate with the GPVI:FceRIgamma complex in platelets and initiate platelet activation through phosphorylation of the immunoreceptor tyrosine-based activation motif (ITAM) in FceRI gamma, leading to binding and activation of the tyrosine kinase Syk. Downstream of Syk, a series of adapter molecules and effectors lead to platelet activation. vWF protein is a polymeric structure of variable size. It is secreted in two directions, by the endothelium basolaterally and into the bloodstream. Shear-induced aggregation is achieved when vWF binds via its A1 domain to GPIb (part of GPIb-IX-V), and via its A3 domain mediating collagen binding to the subendothelium. The interaction between vWF and GPIb is regulated by shear force; an increase in the shear stress results in a corresponding increase in the affinity of vWF for GPIb.


##### Edit history

| Date | Action | Author |
| --- | --- | --- |
| 2004-08-13 | Authored | de Bono B |
| 2004-09-25 | Created | Farndale R, Pace NP, de Bono B |
| 2021-05-21 | Modified | Shorser S |

#### Entities found in this pathway (3)

| Input | UniProt Id | Input | UniProt Id | Input | UniProt Id |
| --- | --- | --- | --- | --- | --- |
| COL1A2 | P08123 | TSKU | P07359 | VWF | P04275 |

#### 25. ECM proteoglycans (R-HSA-3000178)

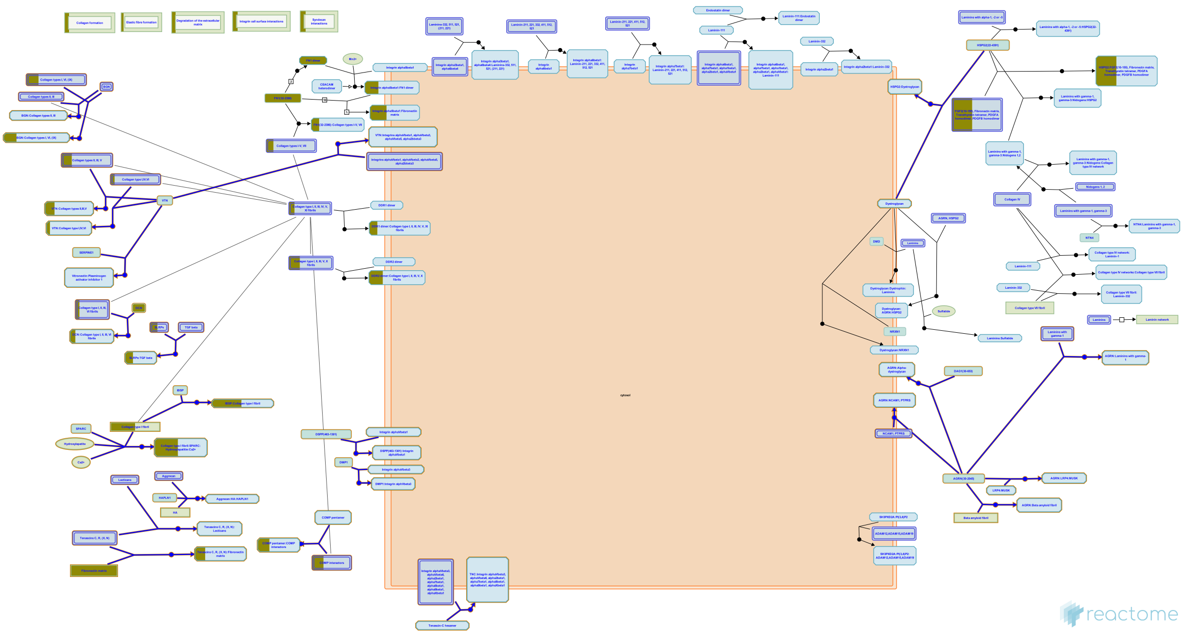

Proteoglycans are major components of the extracellular matrix. In cartilage the matrix constitutes more than 90% of tissue dry weight. Proteoglycans are proteins substituted with glycosaminoglycans (GAGs), linear polysaccharides consisting of a repeating disaccharide, generally of an acetylated amino sugar alternating

with a uronic acid. Most proteoglycans are located in the extracellular

space. Proteoglycans are highly diverse, both in terms of the core proteins and the subtypes of GAG chains, namely chondroitin sulfate (CS), keratan sulfate (KS), dermatan sulfate (DS) and heparan sulfate (HS). Hyaluronan is a non-sulfated GAG whose molecular weight runs into millions of Dalton; in articular cartilage, a single hyaluronan molecule can hold upto 100 aggrecan molecules and these aggregates are stabilized by a link protein.


##### Edit history

| Date | Action | Author |
|---|---|---|
| 2013-01-10 | Authored | Jupe S |
| 2013-01-24 | Created | Jupe S |
| 2013-04-26 | Edited | Jupe S |
| 2013-05-21 | Reviewed | Venkatesan N |
| 2013-05-22 | Reviewed | Ricard-Blum S |

| Date | Action | Author |
|---|---|---|
| 2021-05-22 | Modified | Shorser S |

#### Entities found in this pathway (6)

| Input | UniProt Id | Input | UniProt Id | Input | UniProt Id |
|---|---|---|---|---|---|
| COL1A2 | P08123 | COL5A1 | P20908 | COL8A2 | Q14050 |
| DCN | P07585 | FN1 | P02751 | LUM | P51884 |

### 6. Identifiers found

Below is a list of the input identifiers that have been found or mapped to an equivalent element in Reactome, classified by resource.

#### Entities (136)

| Input | UniProt Id | Input | UniProt Id | Input | UniProt Id |
|---|---|---|---|---|---|
| ACY3 | Q96HD9 | ADAMTS2 | O95450 | AHRR | A9YTQ3 |
| ALDH1L1 | O75891 | ARC | Q7LC44 | ATAD2 | Q6PL18 |
| AZGP1 | P25311 | B3GNT7 | Q8NFL0 | B4GALT1 | P15291 |
| CACNG4 | Q9UBN1 | CAMK1 | Q14012 | CD244 | Q9BZW8 |
| CDH13 | P55290 | CLDN10 | P78369 | COL14A1 | Q05707 |
| COL15A1 | P39059 | COL1A2 | P08123 | COL5A1 | P20908 |
| COL8A2 | P25067, Q14050 | COLEC12 | Q5KU26 | CRH | P06850 |
| CRHBP | P24387 | CSF2RA | P15509 | CUBN | O60494 |
| CUL4B | Q13620 | CXADR | P78310 | DAB1 | O75553 |
| DCN | P07585 | DENND2C | Q68D51 | DNAL4 | O96015 |
| DOT1L | Q8TEK3 | DUSP1 | P28562 | DUSP6 | Q16828 |
| EDNRB | P24530 | EFNA3 | P52797 | EGR4 | Q05215 |
| ELOVL2 | Q9NXB9, Q9NYP7 | EPHB3 | P54753 | F3 | P13726 |
| FGFR1OP2 | Q9NVK5 | FLRT1 | Q9NZU1 | FN1 | P02751 |
| GNG4 | P50150 | GPER1 | Q99527 | GPR17 | Q13304 |
| GPR183 | P32249 | GRIK1 | P39086 | GRIN3A | Q8TCU5 |
| GSTT1 | P0CG30, P30711 | HAS2 | Q92819 | HES1 | Q14469 |
| HES5 | Q5TA89 | HRH1 | P35367 | HSD17B12 | Q53GQ0 |
| IER3 | P46695 | IFITM2 | Q01629 | IGF2 | P01344 |
| IL1A | P01583 | IL6R | P08887 | KCNJ9 | Q92806 |
| KIF26A | Q9ULI4 | LGI3 | Q8N145 | LPL | P06858 |
| LRP8 | Q14114 | LUM | P51884 | LYPD1 | Q8N2G4 |
| MAML3 | Q96JK9 | MAP2K6 | P52564 | MAP3K1 | Q13233 |
| MCF2 | P10911 | MCHR1 | Q99705 | MMD | Q9P0K1 |
| MYBPC1 | Q00872 | NAPEPLD | Q6IQ20 | NEFL | P07196 |
| NIPAL2 | Q9H841 | NOS2 | P35228 | NPY | P01298, P01303 |
| NPY2R | P49146 | ORAI2 | Q96SN7 | OTOGL | Q3ZCN5 |
| PCOLCE | Q15113 | PDGFB | P01127 | PDIA5 | Q14554 |
| PDK4 | Q16654 | PENK | P01210 | PGAP1 | Q75T13 |
| PLCXD2 | Q5T2D3 | PLD6 | Q8N2A8 | PMEPA1 | Q969W9 |
| PPM1K | Q8N3J5 | PPP1R3C | Q9UQK1 | PTGER4 | P35408 |
| PTGS2 | P35354 | PTPRU | Q92729 | PVALB | P20472 |
| RAB37 | Q96AX2 | RAB39A | Q14964 | RELN | P78509 |
| RRAD | P55042 | RXRB | P28702 | SCARA5 | P21757, Q6ZMJ2 |
| SCG2 | O00255, P13521 | SCUBE1 | Q8IWY4 | SDC1 | P18827 |
| SEC61A2 | Q9H9S3 | SH2D1B | O14796 | SH3BP1 | Q9Y3L3 |
| SIK1 | O00567 | SLC13A4 | Q9UKG4 | SLC14A1 | Q13336 |
| SLC16A10 | Q8TF71 | SLC25A12 | O75746 | SLC2A3 | P11169 |
| SLC4A4 | Q9Y6R1 | SLC7A10 | Q9NS82 | SLC7A5 | Q01650 |
| SLN | O15427 | SPC24 | Q8NBT2 | SPTSSB | Q8NFR3 |
| STEAP2 | Q8NFT2 | TAGAP | Q8N103 | TIFA | P38919 |

| Input | UniProt Id |
|---|---|
| TIMP1 | P01033 |
| TNFSF18 | Q9UNG2 |
| TYW3 | Q6IPR3 |
| VWF | P04275 |
| WNT7A | O00755 |

| Input | UniProt Id |
|---|---|
| TIMP3 | P35625 |
| TSC22D3 | Q99576 |
| VAMP1 | P63027 |
| WIF1 | Q9Y5W5 |

| Input | UniProt Id |
|---|---|
| TLL2 | Q9Y6L7 |
| TSKU | P07359 |
| VEGFA | P15692 |
| WNT2 | O00755, P09544 |

| Input | Ensembl Id |
|---|---|
| ARC | ENSG00000198576 |
| EGR4 | ENSG00000135625 |
| HES1 | ENSG00000114315 |
| IGF2 | ENST00000337883, ENST00000381406 |
| LPL | ENSG00000175445 |
| PDIA5 | ENSG00000065485 |
| RRAD | ENSG00000166592 |
| VEGFA | ENSG00000112715 |

| Input | Ensembl Id |
|---|---|
| COL1A2 | ENSG00000164692 |
| F3 | ENSG00000117525 |
| HES5 | ENSG00000197921 |
| IL1A | ENSG00000115008 |
| NOS2 | ENSG00000007171 |
| PTGS2 | ENSG00000073756 |
| SLC2A3 | ENSG00000059804 |

| Input | Ensembl Id |
|---|---|
| CRH | ENSG00000147571 |
| FN1 | ENSG00000115414 |
| IFITM2 | ENSG00000185201 |
| IL6R | ENSG00000160712 |
| NPY | ENSG00000122585 |
| PVALB | ENSG00000100362 |
| TIMP1 | ENSG00000102265 |

### 7. Identifiers not found

These 87 identifiers were not found neither mapped to any entity in Reactome.

| | | | | | | | |
|---|---|---|---|---|---|---|---|
| AC117395.1 | ACVR1 | ADAP2 | ANKRD24 | BCL7A | BCOR | BMP6 | BTG3 |
| C12orf75 | C1QTNF3 | C1orf51 | CABP7 | CBLN1 | CD24P4 | CLN8 | DCDC1 |
| DLX6-AS1 | DNAJB4 | ELFN1 | FAM149A | FAM217B | FAM84A | FAM89A | FILIP1L |
| FIZ1 | GOLPH3L | GPR125 | GPR153 | GPR3 | GRID2 | IGFBPL1 | IGSF9B |
| ISM1 | ITPRIPL2 | LHFPL3 | LINC00925 | LINC00969 | LUZP2 | MCTP1 | MEG3 |
| MUM1L1 | MXRA5 | NDNF | NECAB1 | NEFH | NPNT | NXPH4 | PCSK7 |
| PHF21B | PHLDB2 | PLVAP | PNKD | PRUNE2 | PTPRE | RASD1 | RN7SKP203 |
| RNVU1-18 | RP1-161N10.1 | RP11-137H2.6 | RP11-150O12.1 | RP11-247L20.4 | RP11-2B6.2 | RP11-411B10.4 | RP11-432J22.2 |
| RP11-53O19.3 | RP11-676J15.1 | RP13-514E23.1 | RP4-555D20.2 | RP6-91H8.5 | RREB1 | SEL1L3 | SERTAD1 |
| SFXN5 | SH3BP2 | SLC39A12 | SYNDIG1 | TENM3 | TMEM255A | TRIB2 | TRIM67 |
| VSTM2B | WDR49 | XIRP1 | XKR7 | ZMIZ1 | ZNF365 | ZSCAN31 | |
