## Supplementary material for "Compartment-specific total RNA profile of Hippocampal and Cortical cells from Mesial Temporal Lobe Epilepsy tissue": Other supplemental files_Reactome: Cx_mTLE+HS_Nuc_updownspecific_report.pdf

### 3. Genome-wide overview

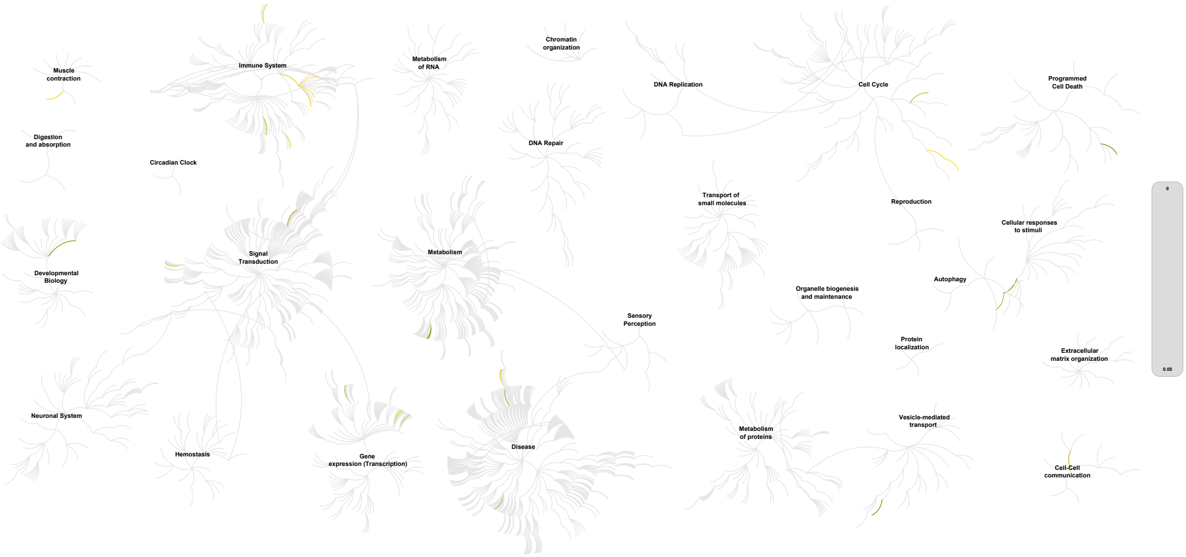


This figure shows a genome-wide overview of the results of your pathway analysis. Reactome pathways are arranged in a hierarchy. The center of each of the circular "bursts" is the root of one top-level pathway, for example "DNA Repair". Each step away from the center represents the next level lower in the pathway hierarchy. The color code denotes over-representation of that pathway in your input dataset. Light grey signifies pathways which are not significantly over-represented.

### 4. Most significant pathways

The following table shows the 25 most relevant pathways sorted by p-value.

| Pathway name | Entities | | | | Reactions | |
|---|---|---|---|---|---|---|
| | found | ratio | p-value | FDR* | found | ratio |
| Interleukin-10 signaling | 11 / 86 | 0.006 | 1.26e-04 | 0.087 | 2 / 15 | 0.001 |
| Interleukin-4 and Interleukin-13 signaling | 18 / 211 | 0.015 | 1.96e-04 | 0.087 | 8 / 47 | 0.003 |
| Transcriptional activation of cell cycle inhibitor p21 | 3 / 6 | 4.13e-04 | 0.001 | 0.222 | 4 / 5 | 3.70e-04 |
| Transcriptional activation of p53 responsive genes | 3 / 6 | 4.13e-04 | 0.001 | 0.222 | 4 / 5 | 3.70e-04 |
| RUNX3 regulates CDKN1A transcription | 3 / 8 | 5.50e-04 | 0.002 | 0.403 | 4 / 6 | 4.44e-04 |
| CLEC7A (Dectin-1) induces NFAT activation | 4 / 18 | 0.001 | 0.003 | 0.419 | 4 / 6 | 4.44e-04 |
| Smooth Muscle Contraction | 6 / 49 | 0.003 | 0.005 | 0.603 | 6 / 11 | 8.14e-04 |
| Calcineurin activates NFAT | 3 / 12 | 8.25e-04 | 0.007 | 0.603 | 2 / 3 | 2.22e-04 |
| RUNX3 regulates RUNX1-mediated transcription | 2 / 4 | 2.75e-04 | 0.007 | 0.603 | 2 / 2 | 1.48e-04 |
| Other interleukin signaling | 4 / 24 | 0.002 | 0.008 | 0.603 | 6 / 17 | 0.001 |
| Signaling by Interleukins | 32 / 643 | 0.044 | 0.009 | 0.603 | 50 / 493 | 0.036 |
| Chemokine receptors bind chemokines | 6 / 57 | 0.004 | 0.011 | 0.603 | 3 / 19 | 0.001 |
| Signaling by FGFR1 amplification mutants | 2 / 5 | 3.44e-04 | 0.011 | 0.603 | 5 / 5 | 3.70e-04 |
| Signaling by activated point mutants of FGFR1 | 3 / 15 | 0.001 | 0.013 | 0.603 | 4 / 4 | 2.96e-04 |
| TFAP2 (AP-2) family regulates transcription of cell cycle factors | 2 / 6 | 4.13e-04 | 0.016 | 0.603 | 3 / 4 | 2.96e-04 |
| G2/M DNA replication checkpoint | 2 / 7 | 4.81e-04 | 0.021 | 0.603 | 2 / 2 | 1.48e-04 |
| DEx/H-box helicases activate type I IFN and inflammatory cytokines production | 2 / 7 | 4.81e-04 | 0.021 | 0.603 | 1 / 5 | 3.70e-04 |
| Cell-extracellular matrix interactions | 3 / 19 | 0.001 | 0.024 | 0.603 | 2 / 10 | 7.40e-04 |
| ATF4 activates genes in response to endoplasmic reticulum stress | 4 / 34 | 0.002 | 0.024 | 0.603 | 2 / 7 | 5.18e-04 |
| TP53 Regulates Transcription of Genes Involved in G1 Cell Cycle Arrest | 3 / 20 | 0.001 | 0.027 | 0.603 | 6 / 17 | 0.001 |
| IkBA variant leads to EDA-ID | 2 / 8 | 5.50e-04 | 0.027 | 0.603 | 2 / 2 | 1.48e-04 |
| FGFR1 ligand binding and activation | 3 / 21 | 0.001 | 0.03 | 0.603 | 6 / 7 | 5.18e-04 |

| Pathway name | Entities | | | | Reactions | |
|---|---|---|---|---|---|---|
| | found | ratio | p-value | FDR* | found | ratio |
| Phospholipase C-mediated cascade: FGFR1 | 3 / 22 | 0.002 | 0.034 | 0.603 | 3 / 3 | 2.22e-04 |
| Negative regulation of FGFR1 signaling | 4 / 39 | 0.003 | 0.037 | 0.603 | 15 / 15 | 0.001 |
| FGFR1 mutant receptor activation | 4 / 39 | 0.003 | 0.037 | 0.603 | 11 / 25 | 0.002 |

* False Discovery Rate

### 5. Pathways details

For every pathway of the most significant pathways, we present its diagram, as well as a short summary, its bibliography and the list of inputs found in it.

#### 1. Interleukin-10 signaling (R-HSA-6783783)

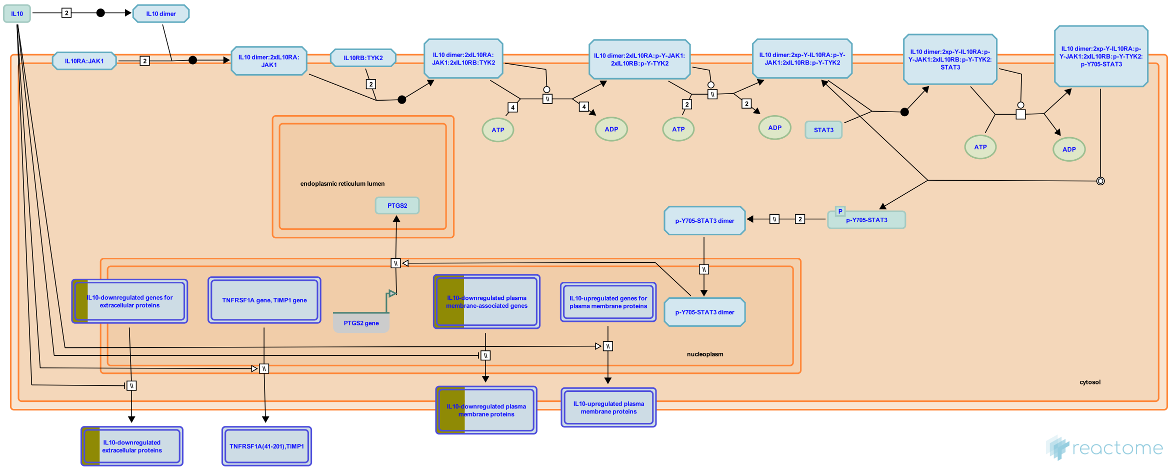


Interleukin-10 (IL10) was originally described as a factor named cytokine synthesis inhibitory factor that inhibited T-helper (Th) 1 activation and Th1 cytokine production (Fiorentino et al. 1989). It was found to be expressed by a variety of cell types including macrophages, dendritic cell subsets, B cells, several T-cell subpopulations including Th2 and T-regulatory cells (Tregs) and Natural Killer (NK) cells (Moore et al. 2001). It is now recognized that the biological effects of IL10 are directed at antigen-presenting cells (APCs) such as macrophages and dendritic cells (DCs), its effects on T-cell development and differentiation are largely indirect via inhibition of macrophage/dendritic cell activation and maturation (Pestka et al. 2004, Mocellin et al. 2004). T cells are thought to be the main source of IL10 (Hedrich & Bream 2010). IL10 inhibits a broad spectrum of activated macrophage/monocyte functions including monokine synthesis, NO production, and expression of class II MHC and costimulatory molecules such as IL12 and CD80/CD86 (de Waal Malefyt et al. 1991, Gazzinelli et al. 1992). Studies with recombinant cytokine and neutralizing antibodies revealed pleiotropic activities of IL10 on B, T, and mast cells (de Waal Malefyt et al. 1993, Rousset et al. 1992, Thompson-Snipes et al. 1991) and provided evidence for the in vivo significance of IL10 activities (Ishida et al. 1992, 1993). IL10 antagonizes the expression of MHC class II and the co-stimulatory molecules CD80/CD86 as well as the pro-inflammatory cytokines IL1Beta, IL6, IL8, TNFalpha and especially IL12 (Fiorentino et al. 1991, D'Andrea et al. 1993). The biological role of IL10 is not limited to inactivation of APCs, it also enhances B cell, granulocyte, mast cell, and keratinocyte growth/differentiation, as well as NK-cell and CD8+ cytotoxic T-cell activation (Moore et al. 2001, Hedrich & Bream 2010). IL10 also enhances NK-cell proliferation and/or production of IFN-gamma (Cai et al. 1999).

IL10-deficient mice exhibited inflammatory bowel disease (IBD) and other exaggerated inflammatory responses (Kuhn et al. 1993, Berg et al. 1995) indicating a critical role for IL10 in limiting inflammatory responses. Dysregulation of IL10 is linked with susceptibility to numerous infectious and autoimmune diseases in humans and mouse models (Hedrich & Bream 2010).

IL10 signaling is initiated by binding of homodimeric IL10 to the extracellular domains of two adjoining IL10RA molecules. This tetramer then binds two IL10RB chains. IL10RB cannot bind to IL10 unless bound to IL10RA (Ding et al. 2001, Yoon et al. 2006); binding of IL10 to IL10RA without the co-presence of IL10RB fails to initiate signal transduction (Kotenko et al. 1997).

IL10 binding activates the receptor-associated Janus tyrosine kinases, JAK1 and TYK2, which are constitutively bound to IL10R1 and IL10R2 respectively. In the classic model of receptor activation assembly of the receptor complex is believed to enable JAK1/TYK2 to phosphorylate and activate each other. Alternatively the binding of IL10 may cause conformational changes that allow the pseudokinase inhibitory domain of one JAK kinase to move away from the kinase domain of the other JAK within the receptor dimer-JAK complex, allowing the two kinase domains to interact and trans-activate (Waters & Brooks 2015).

The activated JAK kinases phosphorylate the intracellular domains of the IL10R1 chains on specific tyrosine residues. These phosphorylated tyrosine residues and their flanking peptide sequences serve as temporary docking sites for the latent, cytosolic, transcription factor, STAT3. STAT3 transiently docks on the IL10R1 chain via its SH2 domain, and is in turn tyrosine phosphorylated by the receptor-associated JAKs. Once activated, it dissociates from the receptor, dimerizes with other STAT3 molecules, and translocates to the nucleus where it binds with high affinity to STAT-binding elements (SBEs) in the promoters of IL-10-inducible genes (Donnelly et al. 1999).

#### Edit history

| Date | Action | Author |
|---|---|---|
| 2015-06-17 | Authored | Jupe S |
| 2015-06-17 | Created | Jupe S |
| 2016-09-05 | Reviewed | Meldal BH |
| 2016-11-14 | Edited | Jupe S |
| 2021-05-31 | Modified | Shorser S |

#### Entities found in this pathway (5)

| Input | UniProt Id | Input | UniProt Id | Input | UniProt Id |
|---|---|---|---|---|---|
| CCL4 | O00626, P13236 | CSF1 | P09603 | CXCL1 | P09341 |
| ICAM1 | P05362 | IL8 | P10145 | | |

| Input | Ensembl Id | Input | Ensembl Id | Input | Ensembl Id |
|---|---|---|---|---|---|
| CCL4 | ENSG00000275302 | CSF1 | ENSG00000184371 | CXCL1 | ENSG00000163739 |
| ICAM1 | ENSG00000090339 | IL8 | ENSG00000169429 | | |

#### 2. Interleukin-4 and Interleukin-13 signaling (R-HSA-6785807)

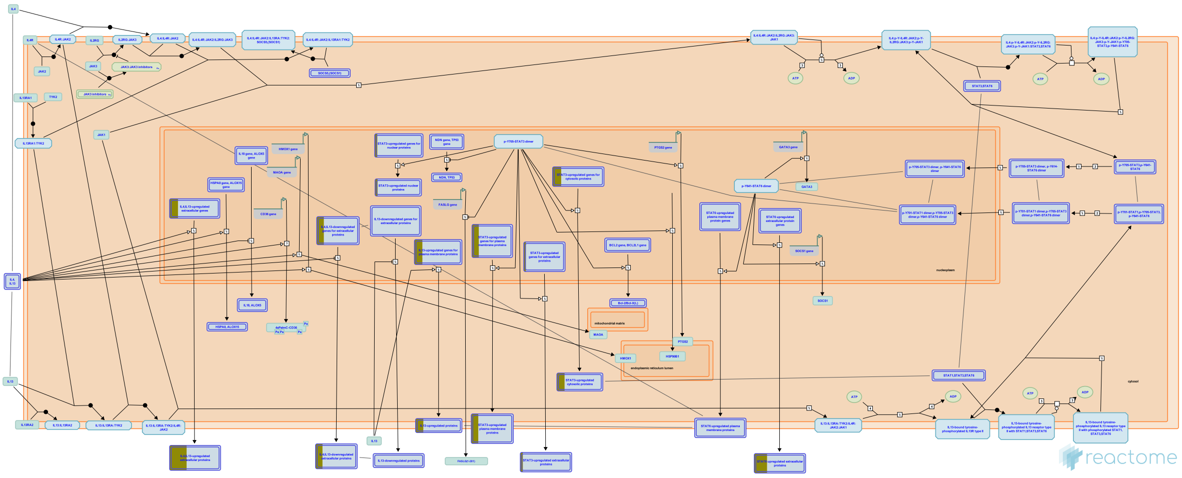

Interleukin-4 (IL4) is a principal regulatory cytokine during the immune response, crucially important in allergy and asthma (Nelms et al. 1999). When resting T cells are antigen-activated and expand in response to Interleukin-2 (IL2), they can differentiate as Type 1 (Th1) or Type 2 (Th2) T helper cells. The outcome is influenced by IL4. Th2 cells secrete IL4, which both stimulates Th2 in an autocrine fashion and acts as a potent B cell growth factor to promote humoral immunity (Nelms et al. 1999).

| Input | UniProt Id | Input | UniProt Id | Input | UniProt Id |
|---|---|---|---|---|---|
| ANXA1 | P04083 | CCL4 | O00626, P51671 | CDKN1A | P38936 |
| ICAM1 | P05362 | IL8 | P10145 | ITGAX | P20702 |
| JUNB | P17275 | MCL1 | Q07820 | TGFB1 | P01137 |

| Input | Ensembl Id | Input | Ensembl Id | Input | Ensembl Id |
|---|---|---|---|---|---|
| ANXA1 | ENSG00000135046 | CDKN1A | ENSG00000124762 | ICAM1 | ENSG00000090339 |
| IL8 | ENSG00000169429 | ITGAX | ENSG00000140678 | JUNB | ENSG00000171223 |
| MCL1 | ENSG00000143384 | TGFB1 | ENSG00000105329 | | |

#### 3. Transcriptional activation of cell cycle inhibitor p21 (R-HSA-69895)

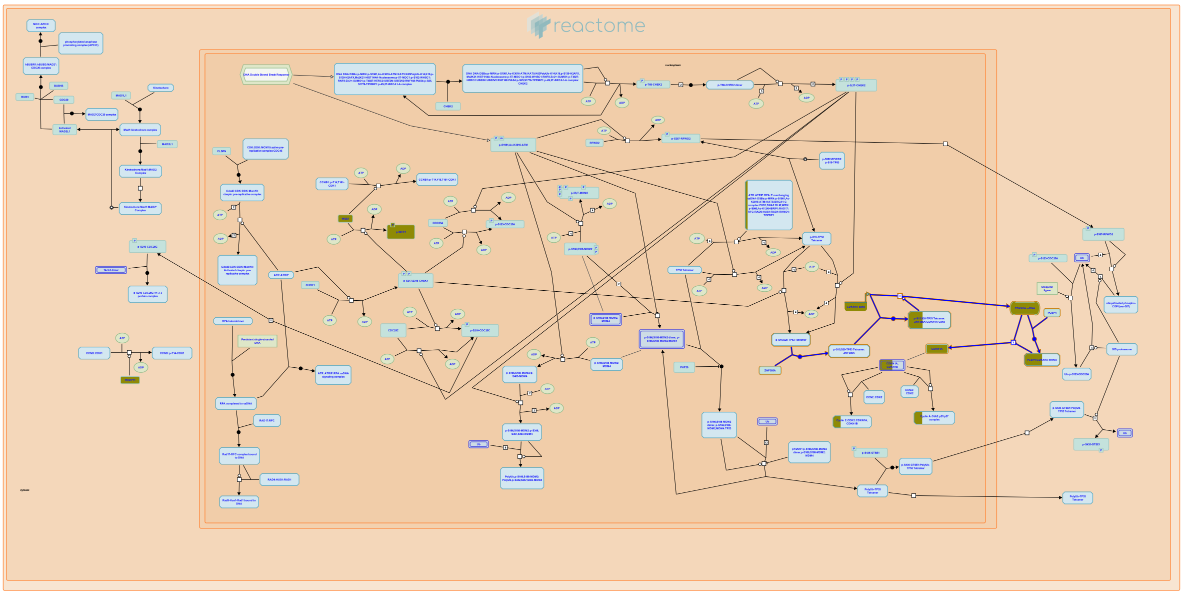

Both p53-independent and p53-dependent mechanisms of induction of p21 mRNA have been demonstrated. p21 is transcriptionally activated by p53 after DNA damage (el-Deiry et al., 1993).


##### Edit history

| Date | Action | Author |
|---|---|---|
| 2003-06-05 | Created | Khanna KK |
| 2021-05-31 | Modified | Shorser S |

##### Entities found in this pathway (1)

| Input | UniProt Id |
|---|---|
| CDKN1A | P38936 |

| Input | Ensembl Id |
|---|---|
| CDKN1A | ENSG00000124762, ENST00000244741 |

#### 4. Transcriptional activation of p53 responsive genes (R-HSA-69560)

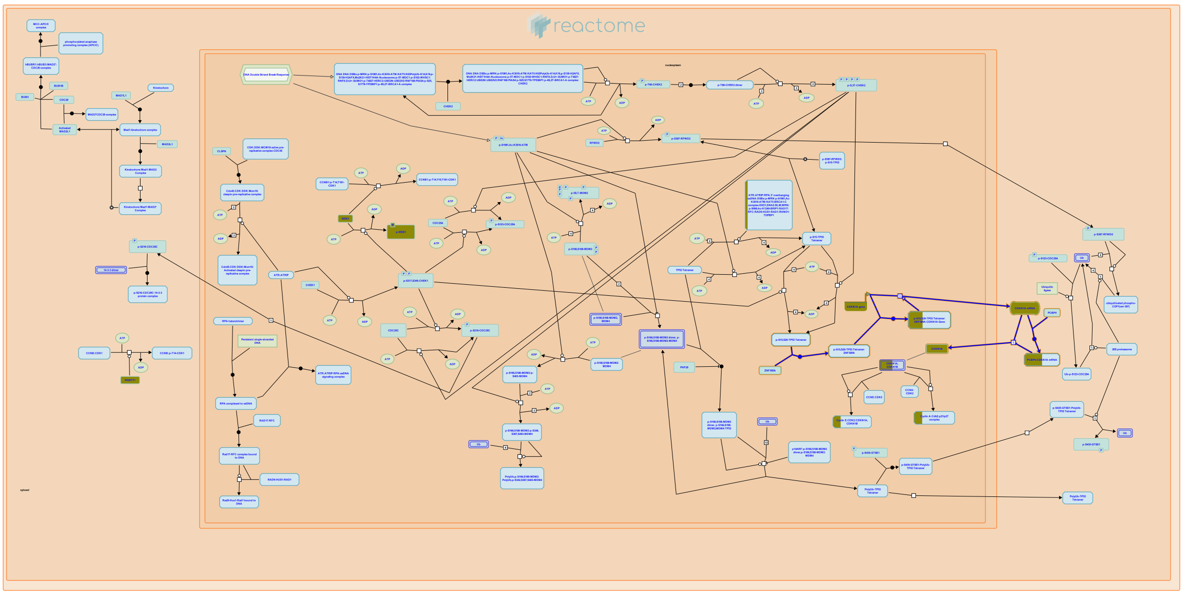

p53 causes G1 arrest by inducing the expression of a cell cycle inhibitor, p21 (El-Deiry et al, 1993; Harper et al, 1993; Xiong et al, 1993). P21 binds and inactivates Cyclin-Cdk complexes that mediate G1/S progression, resulting in lack of phosphorylation of Rb, E2F sequestration and cell cycle arrest at the G1/S transition. Mice with a homozygous deletion of p21 gene are deficient in their ability to undergo a G1/S arrest in response to DNA damage (Deng et al, 1995).


##### Edit history

| Date | Action | Author |
|---|---|---|
| 2003-06-05 | Created | Khanna KK |
| 2021-05-21 | Modified | Shorser S |

##### Entities found in this pathway (1)

| Input | UniProt Id |
|---|---|
| CDKN1A | P38936 |

| Input | Ensembl Id |
|---|---|
| CDKN1A | ENSG00000124762, ENST00000244741 |

#### 5. RUNX3 regulates CDKN1A transcription (R-HSA-8941855)

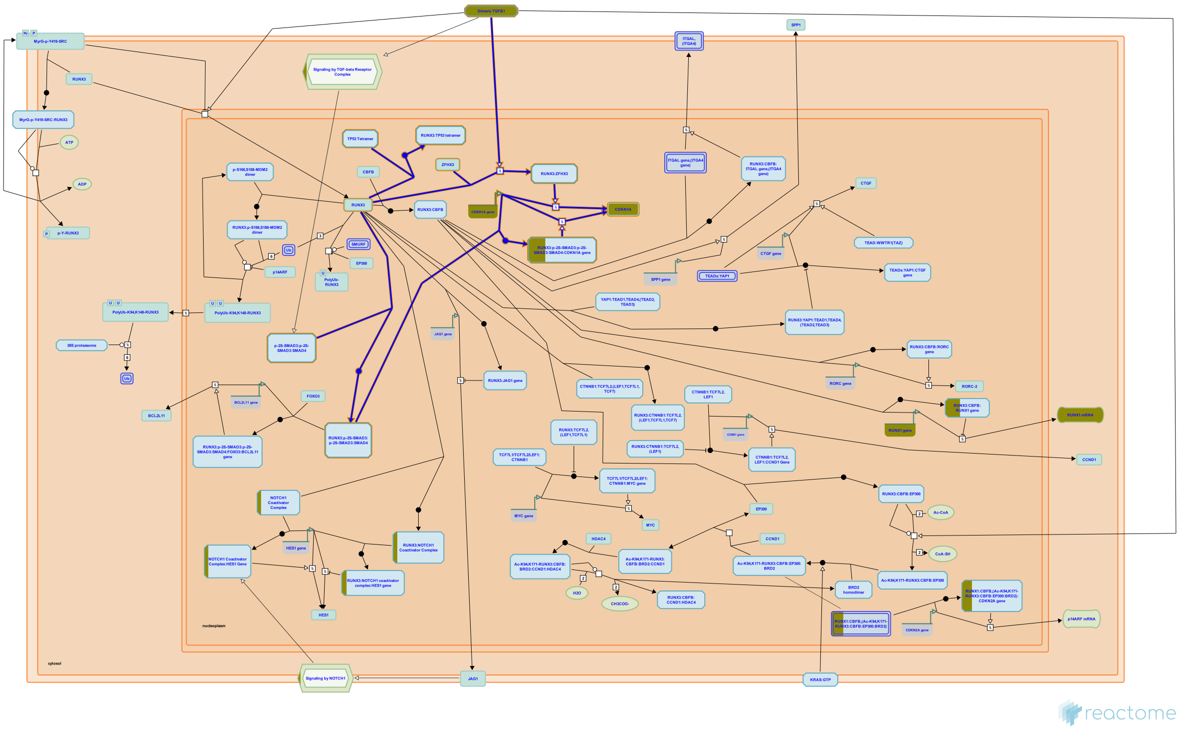

RUNX3 contributes to the upregulation of the CDKN1A (p21) gene transcription in response to TGF-beta (TGFB1) signaling. RUNX3 binds to SMAD3 and SMAD4, and cooperates with the activated SMAD3:SMAD4 complex in transactivation of CDKN1A. Runx3 knockout mice exhibit decreased sensitivity to TGF-beta and develop gastric epithelial hyperplasia (Chi et al. 2005). In response to TGF-beta signaling, the CBFB:RUNX3 complex binds to the tumor suppressor ZFHX3 (ATBF1) and, through an unknown mechanism, this complex positively regulates the CDKN1A transcription (Mabuchi et al. 2010).

In addition, RUNX3 may act as a TP53 co-factor, stimulating TP53-mediated transcription of target genes, including CDKN1A (p21) (Yamada et al. 2010).


##### Edit history

| Date | Action | Author |
|---|---|---|
| 2016-10-06 | Created | Orlic-Milacic M |

| Date | Action | Author |
|---|---|---|
| 2016-12-13 | Authored | Orlic-Milacic M |
| 2017-01-31 | Edited | Orlic-Milacic M |
| 2017-01-31 | Reviewed | Ito Y, Chuang LS |
| 2021-05-31 | Modified | Shorser S |

#### Entities found in this pathway (2)

| Input | UniProt Id | Input | UniProt Id |
|---|---|---|---|
| CDKN1A | P38936 | TGFB1 | P01137 |

| Input | Ensembl Id |
|---|---|
| CDKN1A | ENSG00000124762 |

#### 6. CLEC7A (Dectin-1) induces NFAT activation (R-HSA-5607763)

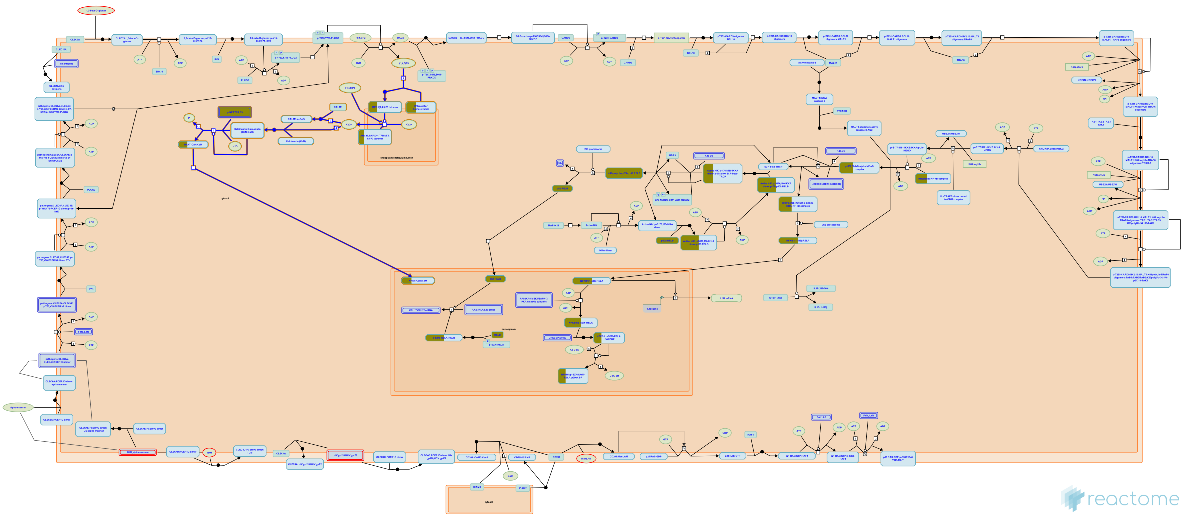

**Cellular compartments:** plasma membrane, cytosol.

CLEC7A (Dectin-1) signals through the classic calcineurin/NFAT pathway through Syk activation phospholipase C-gamma 2 (PLCG2) leading to increased soluble IP3 (inositol trisphosphate). IP3 is able to bind endoplasmic Ca2+ channels, resulting in an influx of Ca2+ into the cytoplasm. This increase in calcium concentration induces calcineurin activation and consequently, dephosphorylation of NFAT and its translocation into the nucleus, triggering gene transcription and extracellular release of Interleukin-2 (Plato et al. 2013, Goodridge et al. 2007, Mourao-Sa et al. 2011).


##### Edit history

| Date | Action | Author |
|---|---|---|
| 2014-07-14 | Edited | Garapati P V |
| 2014-07-14 | Authored | Garapati P V |
| 2014-07-14 | Created | Garapati P V |
| 2014-09-02 | Reviewed | Geijtenbeek TB |
| 2021-05-31 | Modified | Shorser S |

##### Entities found in this pathway (3)

| Input | UniProt Id | Input | UniProt Id | Input | UniProt Id |
|---|---|---|---|---|---|
| ITPR1 | Q14643 | NFATC1 | O95644 | NFATC2 | Q12968, Q13469 |

#### 7. Smooth Muscle Contraction (R-HSA-445355)

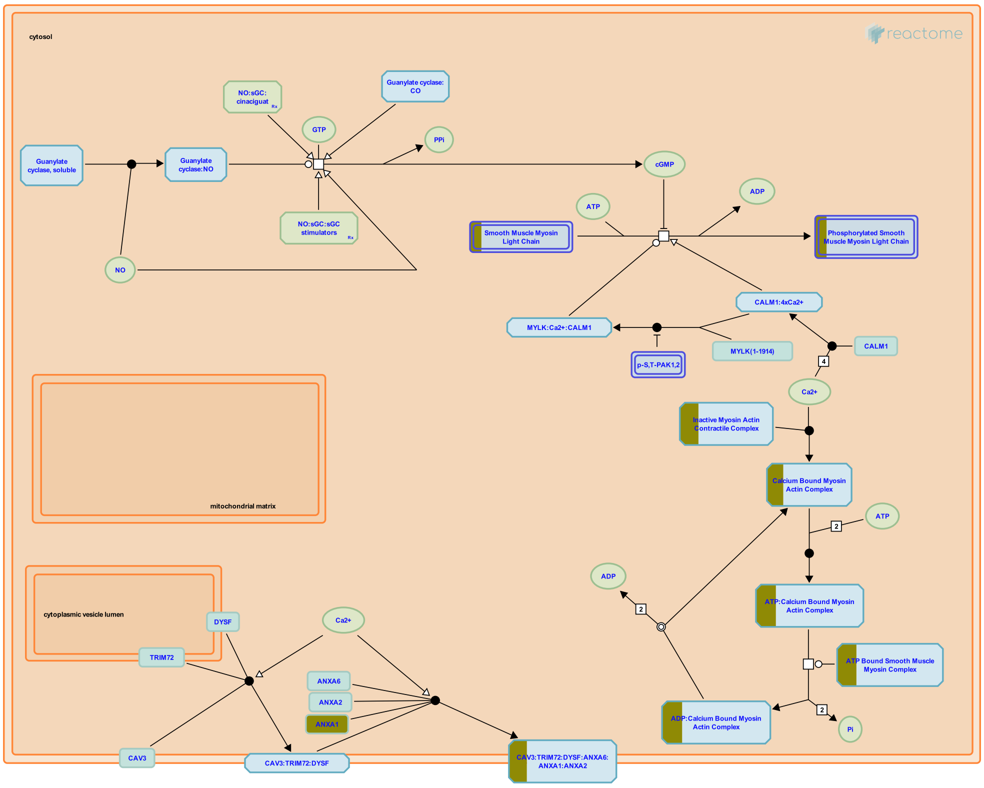


**Cellular compartments:** cytosol, plasma membrane.

Layers of smooth muscle cells can be found in the walls of numerous organs and tissues within the body. Smooth muscle tissue lacks the striated banding pattern characteristic of skeletal and cardiac muscle. Smooth muscle is triggered to contract by the autonomic nervous system, hormones, autocrine/paracrine agents, local chemical signals, and changes in load or length.

Actin:myosin cross bridging is used to develop force with the influx of calcium ions (Ca2+) initiating contraction. Two separate protein pathways, both triggered by calcium influx contribute to contraction, a calmodulin driven kinase pathway, and a caldesmon driven pathway.

Recent evidence suggests that actin, myosin, and intermediate filaments may be far more volatile then previously suspected, and that changes in these cytoskeletal elements along with alterations of the focal adhesions that anchor these proteins may contribute to the contractile cycle.

Contraction in smooth muscle generally uses a variant of the same sliding filament model found in striated muscle, except in smooth muscle the actin and myosin filaments are anchored to focal adhesions, and dense bodies, spread over the surface of the smooth muscle cell. When actin and myosin move across one another focal adhesions are drawn towards dense bodies, effectively squeezing the cell into a smaller conformation. The sliding is triggered by calcium:caldesmon binding, caldesmon acting in an analogous fashion to troponin in striated muscle. Phosphorylation of myosin light chains also is involved in the initiation of an effective contraction.

##### Edit history

| Date | Action | Author |
|---|---|---|
| 2008-01-11 | Reviewed | Rush MG |
| 2009-03-09 | Authored | Gillespie ME |
| 2009-10-30 | Created | Gillespie ME |
| 2009-11-18 | Edited | Gillespie ME |
| 2021-05-31 | Modified | Shorser S |

#### Entities found in this pathway (5)

| Input | UniProt Id | Input | UniProt Id | Input | UniProt Id |
|---|---|---|---|---|---|
| ACTA2 | P62736, P63267 | ANXA1 | P04083 | MYH11 | P35749 |
| MYL6 | P60660 | TPM4 | P67936 | | |

#### 8. Calcineurin activates NFAT (R-HSA-2025928)

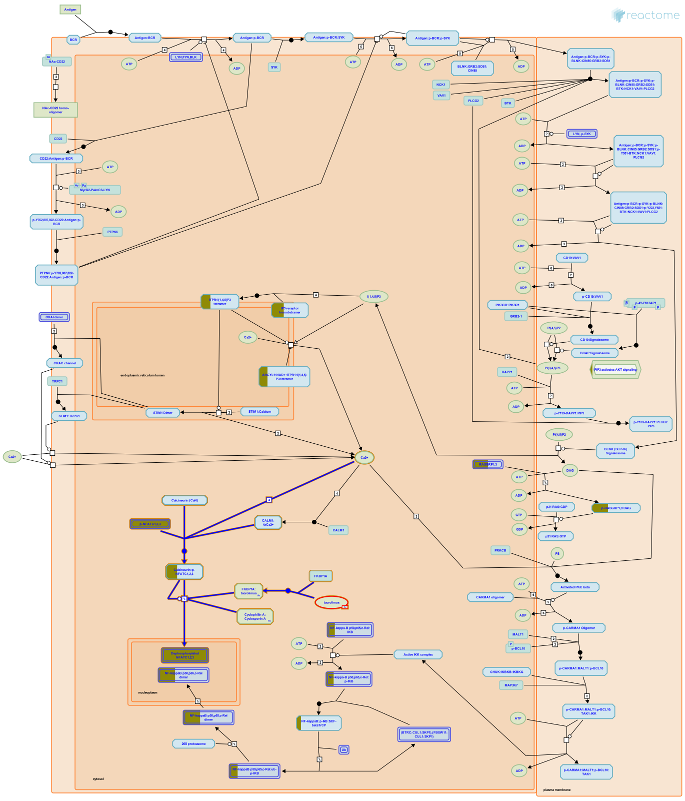

**Cellular compartments:** cytosol, nucleoplasm.

Signaling by the B cell receptor and the T cell receptor stimulate transcription by NFAT factors via calcium (reviewed in Gwack et al. 2007). Cytosolic calcium from intracellular stores and extracellular sources binds calmodulin and activates the protein phosphatase calcineurin. Activated calcineurin dephosphorylates NFATs in the cytosol, exposing nuclear localization sequences on the NFATs and causing the NFATs to be imported into the nucleus where they regulate transcription of target genes in complexes with other transcription factors such as AP-1 and JUN. Calcineurin in the target of the immunosuppressive drugs cyclosporin A and FK-506 (reviewed in Lee and Park 2006).


##### Edit history

| Date | Action | Author |
|---|---|---|
| 2011-12-11 | Edited | May B |
| 2011-12-11 | Authored | May B |
| 2011-12-20 | Created | May B |
| 2012-02-12 | Reviewed | Wienands J |
| 2021-05-31 | Modified | Shorser S |

#### Entities found in this pathway (2)

| Input | UniProt Id | Input | UniProt Id |
|---|---|---|---|
| NFATC1 | O95644 | NFATC2 | Q12968, Q13469 |

#### 9. RUNX3 regulates RUNX1-mediated transcription (R-HSA-8951911)

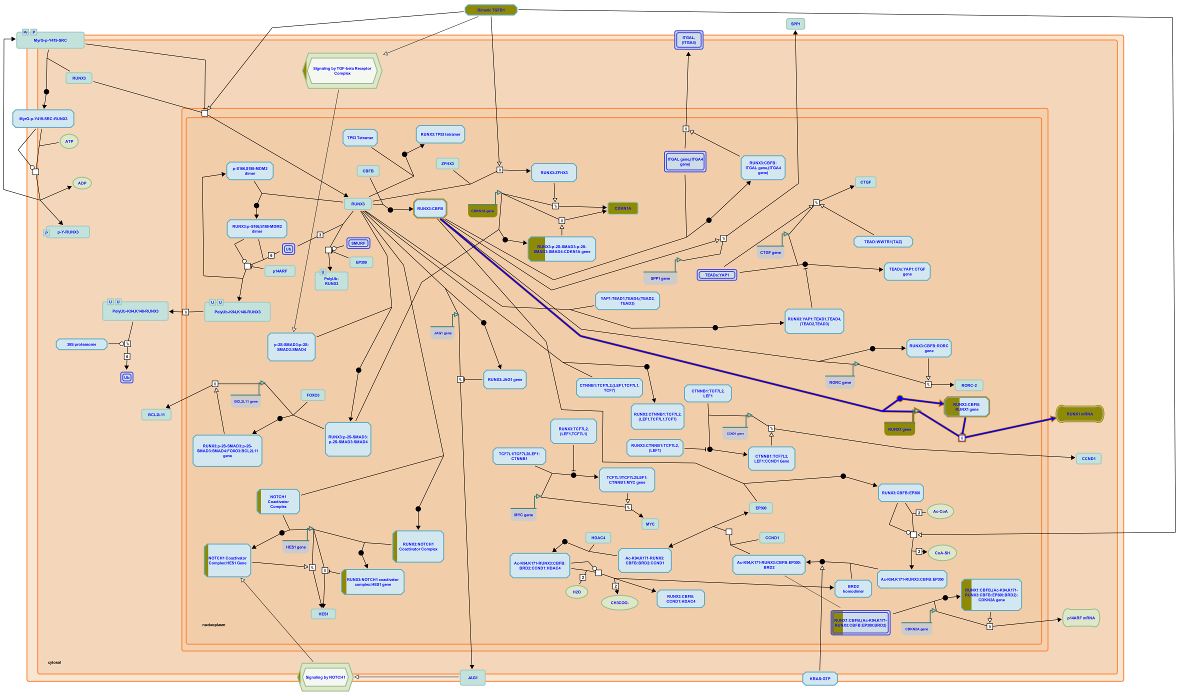

RUNX3 binds to Runx response elements in the distal (P1) promoter of the RUNX1 gene, repressing RUNX1 transcription (Spender et al. 2005).


##### Edit history

| Date | Action | Author |
|---|---|---|
| 2016-12-12 | Created | Orlic-Milacic M |
| 2016-12-13 | Authored | Orlic-Milacic M |
| 2017-01-31 | Edited | Orlic-Milacic M |
| 2017-01-31 | Reviewed | Ito Y, Chuang LS |
| 2017-02-28 | Modified | Orlic-Milacic M |

##### Entities found in this pathway (1)

| Input | Ensembl Id |
|---|---|
| RUNX1 | ENSG00000159216, ENST00000344691 |

#### 10. Other interleukin signaling (R-HSA-449836)

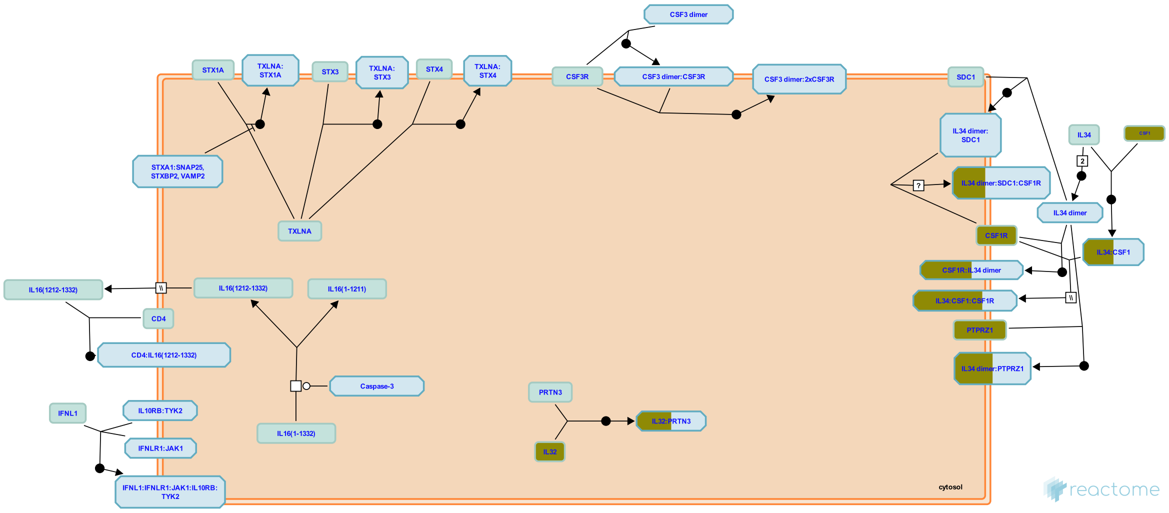


Interleukins are low molecular weight proteins that bind to cell surface receptors and act in an autocrine and/or paracrine fashion. They were first identified as factors produced by leukocytes but are now known to be produced by many other cells throughout the body. They have pleiotropic effects on cells which bind them, impacting processes such as tissue growth and repair, hematopoietic homeostasis, and multiple levels of the host defense against pathogens where they are an essential part of the immune system.


##### Edit history

| Date | Action | Author |
|---|---|---|
| 2009-12-07 | Created | Jupe S |
| 2014-06-04 | Authored | Jupe S |
| 2016-01-28 | Edited | Jupe S |
| 2016-01-28 | Reviewed | Meldal BH |
| 2021-05-22 | Modified | Shorser S |

##### Entities found in this pathway (4)

| Input | UniProt Id |
|---|---|
| CSF1 | P09603 |
| IL32 | P24001 |

| Input | UniProt Id |
|---|---|
| CSF1R | P07333 |
| PTPRZ1 | P23471 |

#### 11. Signaling by Interleukins (R-HSA-449147)

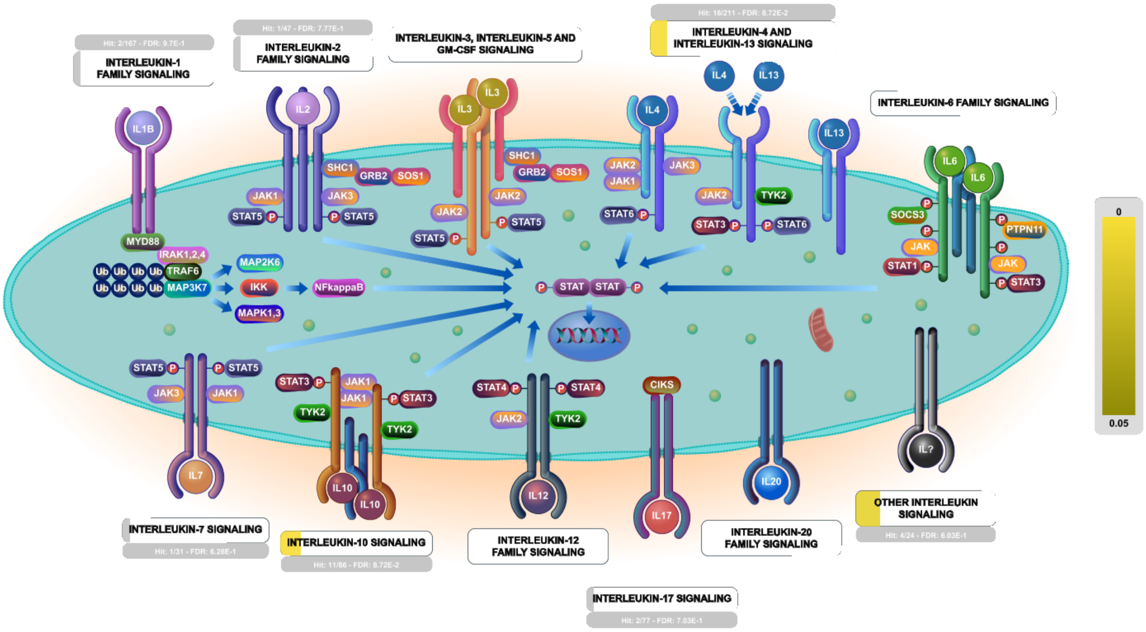


**Cellular compartments:** plasma membrane.

Interleukins are low molecular weight proteins that bind to cell surface receptors and act in an autocrine and/or paracrine fashion. They were first identified as factors produced by leukocytes but are now known to be produced by many other cells throughout the body. They have pleiotropic effects on cells which bind them, impacting processes such as tissue growth and repair, hematopoietic homeostasis, and multiple levels of the host defense against pathogens where they are an essential part of the immune system.

##### Edit history

| Date | Action | Author |
|---|---|---|
| 2009-11-27 | Created | Jupe S |
| 2010-05-17 | Reviewed | Pinteaux E |
| 2010-05-17 | Authored | Ray KP |
| 2010-05-26 | Edited | Jupe S |
| 2021-05-22 | Modified | Shorser S |

##### Entities found in this pathway (19)

| Input | UniProt Id |
| --- | --- |
| ANXA1 | P04083 |
| CSF1 | P09603 |
| ICAM1 | P05362 |
| IL8 | P10145 |
| JUNB | P17275 |
| NFKB2 | Q00653 |
| TGFB1 | P01137 |

| Input | UniProt Id |
| --- | --- |
| CCL4 | O00626, P13236, P51671 |
| CSF1R | P07333 |
| IL32 | P24001 |
| ITGAX | P20702 |
| MCL1 | Q07820 |
| PTK2B | Q14289 |

| Input | UniProt Id |
| --- | --- |
| CDKN1A | P38936 |
| CXCL1 | P09341 |
| IL7R | P16871 |
| JUN | P05412 |
| NFKB1 | P19838 |
| PTPRZ1 | P23471 |

| Input | Ensembl Id |
| --- | --- |
| ANXA1 | ENSG00000135046 |
| CSF1 | ENSG00000184371 |
| IL8 | ENSG00000169429 |
| MCL1 | ENSG00000143384 |

| Input | Ensembl Id |
| --- | --- |
| CCL4 | ENSG00000275302 |
| CXCL1 | ENSG00000163739 |
| ITGAX | ENSG00000140678 |
| TGFB1 | ENSG00000105329 |

| Input | Ensembl Id |
| --- | --- |
| CDKN1A | ENSG00000124762 |
| ICAM1 | ENSG00000090339 |
| JUNB | ENSG00000171223 |

#### 12. Chemokine receptors bind chemokines (R-HSA-380108)

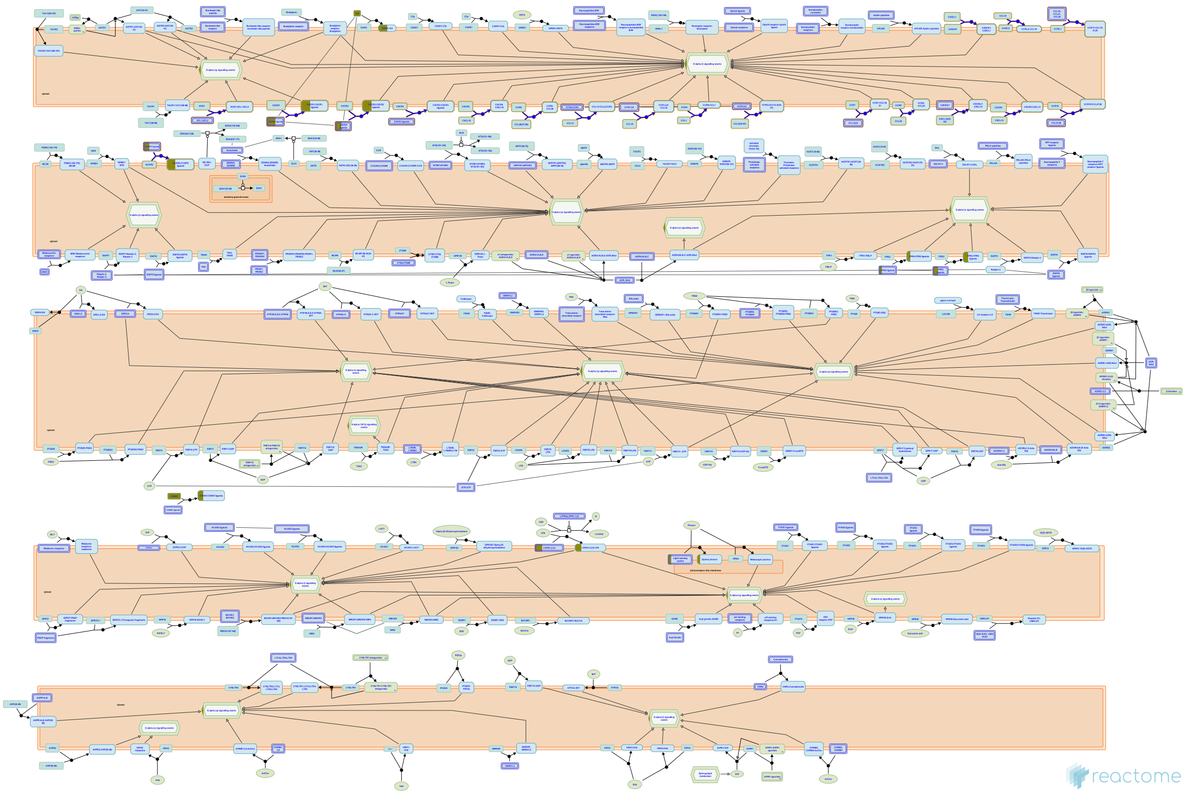

Chemokine receptors are cytokine receptors found on the surface of certain cells, which interact with a type of cytokine called a chemokine. Following interaction, these receptors trigger a flux of intracellular calcium which leads to chemotaxis. Chemokine receptors are divided into different families, CXC chemokine receptors, CC chemokine receptors, CX3C chemokine receptors and XC chemokine receptors that correspond to the 4 distinct subfamilies of chemokines they bind.


##### Edit history

| Date | Action | Author |
| --- | --- | --- |
| 2008-11-07 | Authored | Jassal B |
| 2008-11-07 | Created | Jassal B |
| 2021-05-21 | Modified | Shorser S |

##### Entities found in this pathway (3)

| Input | UniProt Id | Input | UniProt Id | Input | UniProt Id |
| --- | --- | --- | --- | --- | --- |
| CCL4 | O00626, P13236, P51671 | CXCL1 | P09341, P19876 | IL8 | P10145 |

#### 13. Signaling by FGFR1 amplification mutants (R-HSA-1839120)

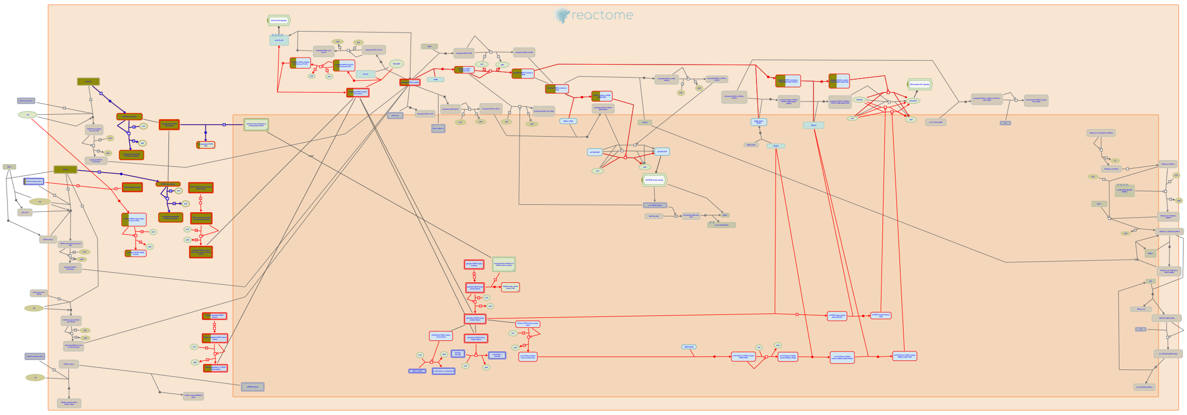

**Cellular compartments:** cytosol, extracellular region, plasma membrane, nucleoplasm.

**Diseases:** cancer.

Amplification or activation of FGFR1 has been reported in lung cancer (Weiss, 2001; Marek, 2009; Dutt, 2011), breast cancer (Reis-Filho, 2006; Turner, 2010), oral squamous carcinoma (Freier, 2007), esophageal squamous cell carcinomas (Ishizuka, 2002), ovarian cancer (Gorringe, 2007), bladder cancer (Simon, 2001), prostate cancer (Edwards, 2003; Acevedo, 2007) and rhabodomyosarcoma (Missiaglia, 2009). Unlike the case for FGFR2 amplifications, FGFR1 amplifications are not associated with additional point mutations and affect signaling without altering the intrinsic kinase activity of the receptor. Overexpressed FGFR1 appears to signal at a basal level in a ligand-independent fashion, but is also able to be stimulated by exogenous ligand. Downstream activation may be the result of aberrant paracrine or autocrine stimulation (reviewed in Turner and Gross, 2010; Greulich and Pollock, 2011). FGFR1 amplification has not been conclusively demonstrated to be the causative oncogenic agent in all of the cancer types mentioned above, and other genes in the 8p11 region may also be candidates in some cases (Bass, 2009; Bernard-Pierrot, 2008; Ray, 2004).


##### Edit history

| Date | Action | Author |
| --- | --- | --- |
| 2011-10-27 | Created | Rothfels K |
| 2012-02-10 | Authored | Rothfels K |
| 2012-05-15 | Reviewed | Ezzat S |
| 2012-05-16 | Edited | Rothfels K |
| 2014-12-04 | Modified | Rothfels K |

#### Entities found in this pathway (1)

| Input | UniProt Id |
| --- | --- |
| FGFR1 | P11362-1, P11362-19 |

#### 14. Signaling by activated point mutants of FGFR1 (R-HSA-1839122)

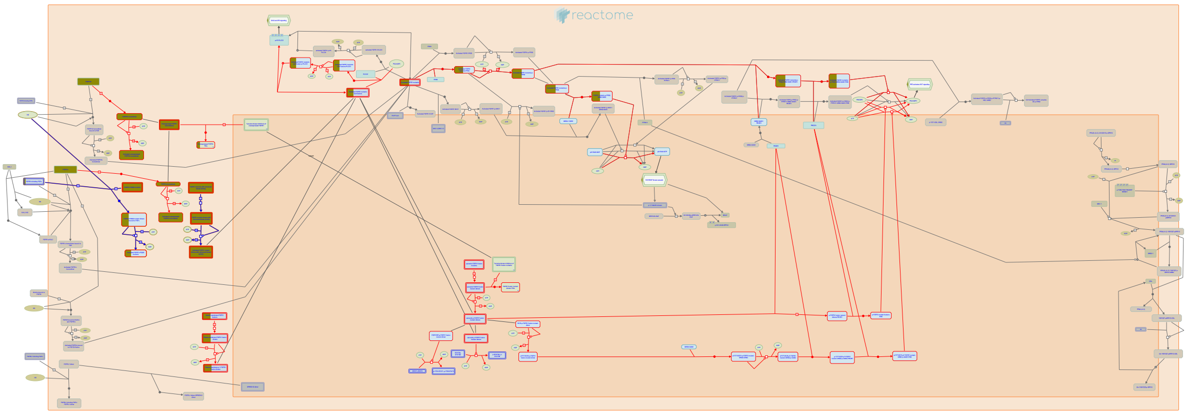

**Cellular compartments:** cytosol, extracellular region, plasma membrane.

**Diseases:** bone development disease, cancer.

Unlike FGFR2 and FGFR3, FGFR1 appears not to be a frequent target of activating point mutations (reviewed in Wesche, 2011; Turner and Grose, 2010). Germline point mutations at residue P252 have been identified in Pfeiffer syndrome (reviewed in Webster and Donoghue, 1997; Burke, 1998; Cunningham, 2007) while mutation of the same residue arising somatically has been identified in melanoma and lung cancer (Ruhe, 2007; Davies, 2005). Two kinase domain mutations have been characterized in glioblastoma (Rand, 2005; Network TCGA, 2008), both at positions that are also mutated in an autosomal disorder in one of the FGFR family members (Muenke, 1994; Bellus, 1995a; Bellus, 2000; Tavormina, 1995a; Tavormina, 1999).


##### Edit history

| Date | Action | Author |
|---|---|---|
| 2011-10-27 | Created | Rothfels K |

| Date | Action | Author |
|---|---|---|
| 2012-02-10 | Authored | Rothfels K |
| 2012-05-15 | Reviewed | Ezzat S |
| 2012-05-16 | Edited | Rothfels K |
| 2012-05-26 | Modified | Rothfels K |

#### Entities found in this pathway (2)

| Input | UniProt Id |
|---|---|
| FGF17 | O60258-1 |

| Input | UniProt Id |
|---|---|
| FGFR1 | P11362, P11362-1 |

#### 15. TFAP2 (AP-2) family regulates transcription of cell cycle factors (R-HSA-8866911)

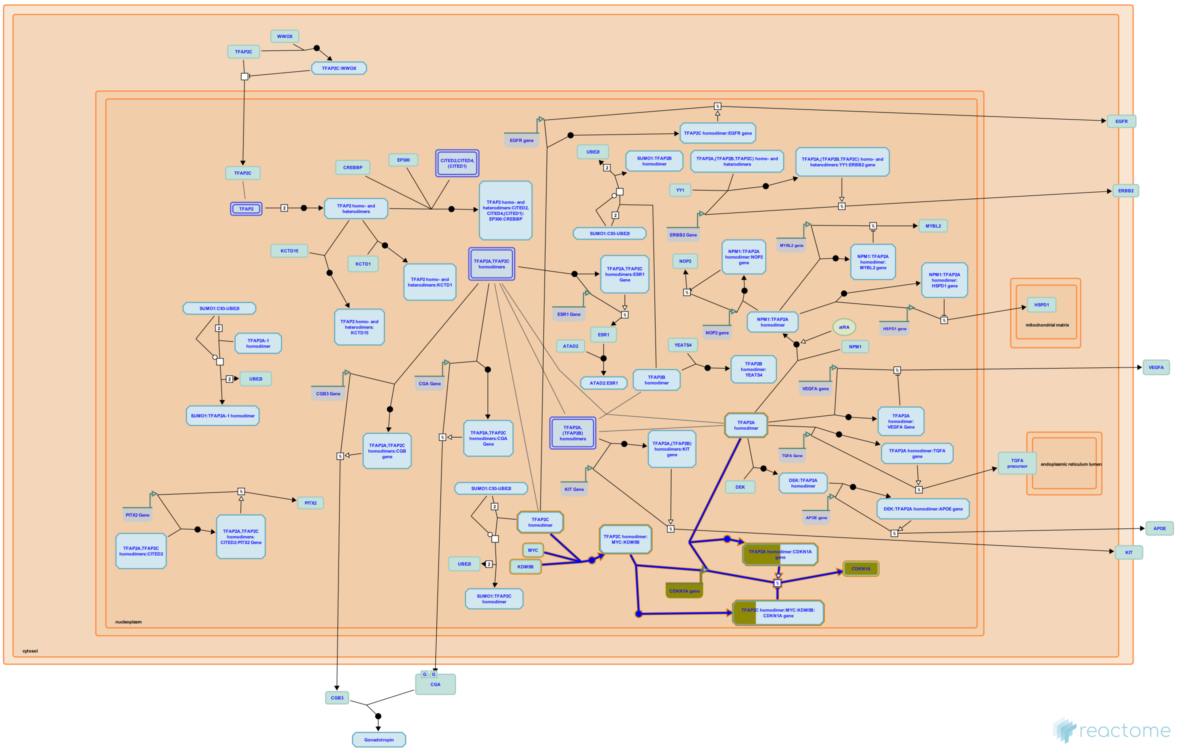

TFAP2A and TFAP2C play opposing roles in transcriptional regulation of the CDKN1A (p21) gene locus. While TFAP2A stimulates transcription of the CDKN1A cyclin-dependent kinase inhibitor (Zeng et al. 1997, Williams et al. 2009, Scibetta et al. 2010), TFAP2C, in cooperation with MYC and histone demethylase KDM5B, represses CDKN1A transcription (Williams et al. 2009, Scibetta et al. 2010, Wong et al. 2012).


##### Edit history

| Date | Action | Author |
|---|---|---|
| 2016-03-14 | Edited | Orlic-Milacic M |
| 2016-03-14 | Authored | Orlic-Milacic M |
| 2016-04-04 | Created | Orlic-Milacic M |

| Date | Action | Author |
|---|---|---|
| 2016-05-04 | Reviewed | Dawid IB, Zarelli VE |
| 2016-05-17 | Reviewed | Bogachek MV, Weigel RJ |
| 2021-05-22 | Modified | Shorser S |

#### Entities found in this pathway (1)

| Input | UniProt Id |
|---|---|
| CDKN1A | P38936 |

| Input | Ensembl Id |
|---|---|
| CDKN1A | ENSG00000124762 |

#### 16. G2/M DNA replication checkpoint (R-HSA-69478)

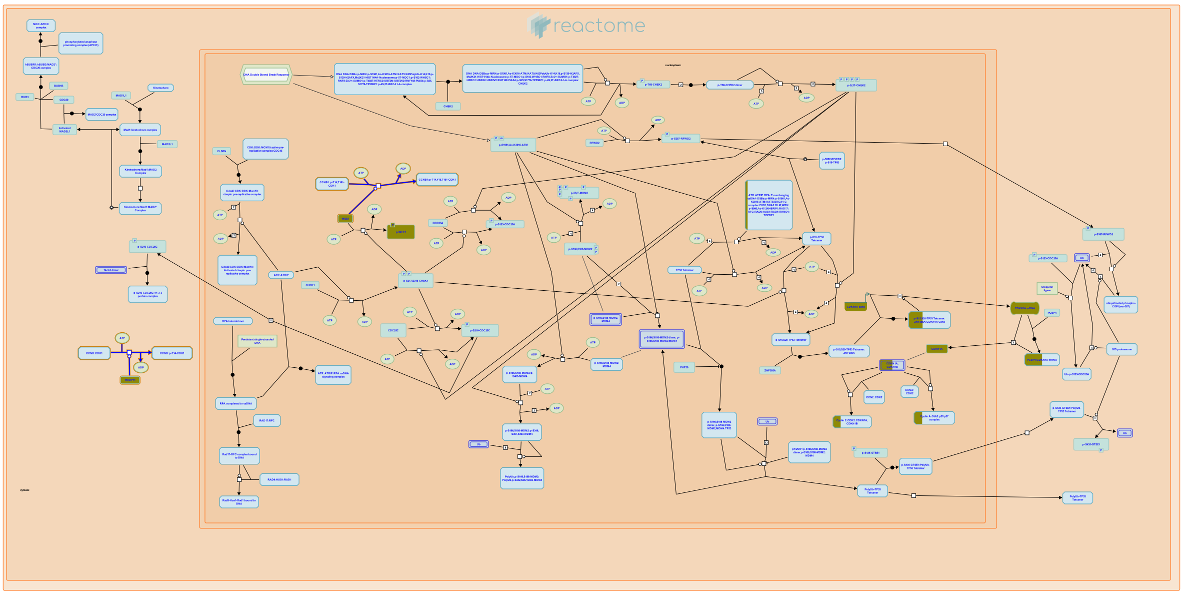

The G2/M DNA replication checkpoint ensures that mitosis is not initiated until DNA replication is complete. If replication is blocked, the DNA replication checkpoint signals to maintain Cyclin B - Cdc2 complexes in their T14Y15 phosphorylated and inactive state. This prevents the phosphorylation of proteins involved in G2/M transition, and prevents mitotic entry.

Failure of these checkpoints results in changes of ploidy: in the case of mitosis without completion of DNA replication, aneuploidy of <2C will result, and the opposite is true if DNA replication is completed more than once in a single cell cycle with an overall increase in ploidy. The mechanism by which unreplicated DNA is first detected by the cell is unknown.

##### References

##### Edit history

| Date | Action | Author |
|---|---|---|
| 2003-06-05 | Created | Walworth N, O'Donnell M |
| 2021-05-22 | Modified | Shorser S |

##### Entities found in this pathway (1)

| Input | UniProt Id |
|---|---|
| WEE1 | P30291, Q99640 |

#### 17. DEx/H-box helicases activate type I IFN and inflammatory cytokines production (R-HSA-3134963)

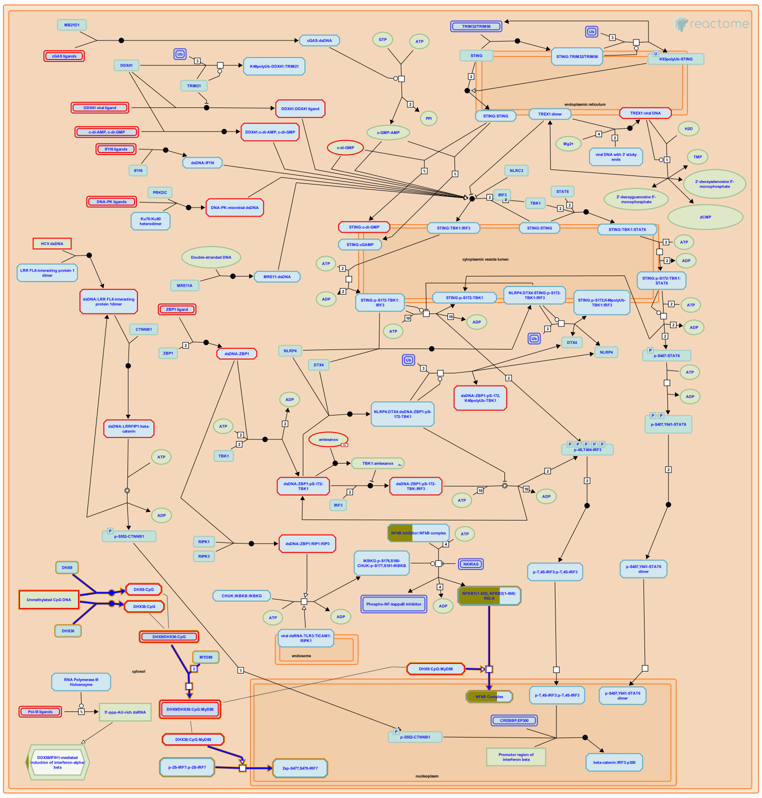

DHX36 and DHX9 are aspartate-glutamate-any amino acid aspartate/histidine (DExD/H) box helicase (DHX) proteins that localize in the cytosol. The DHX RNA helicases family includes a large number of proteins that are implicated in RNA metabolism. Members of this family, RIG-1 and MDA5, have been shown to sense a non-self RNA leading to type I IFN production. RNA helicases DHX36 and DHX9 were found to trigger host responses to non-self DNA in MyD88-dependent manner. DHX36 sensed CpG class A, while DHX9 sensed CpG class B. Both DHX36 and DHX9 were critical for antiviral immune responses in viral DNA-stimulated human plasmacytoid dendritic cells (pDC) (Kim T et al. 2010).


##### Edit history

| Date | Action | Author |
|---|---|---|
| 2013-02-06 | Authored | Shamovsky V |
| 2013-02-11 | Reviewed | D'Eustachio P |
| 2013-02-13 | Created | Shamovsky V |
| 2013-05-17 | Edited | Shamovsky V |
| 2013-05-22 | Reviewed | Wu J, Jin L |
| 2021-05-31 | Modified | Shorser S |

#### Entities found in this pathway (2)

| Input | UniProt Id | Input | UniProt Id |
|---|---|---|---|
| NFKB1 | P19838 | NFKB2 | Q00653 |

#### 18. Cell-extracellular matrix interactions (R-HSA-446353)

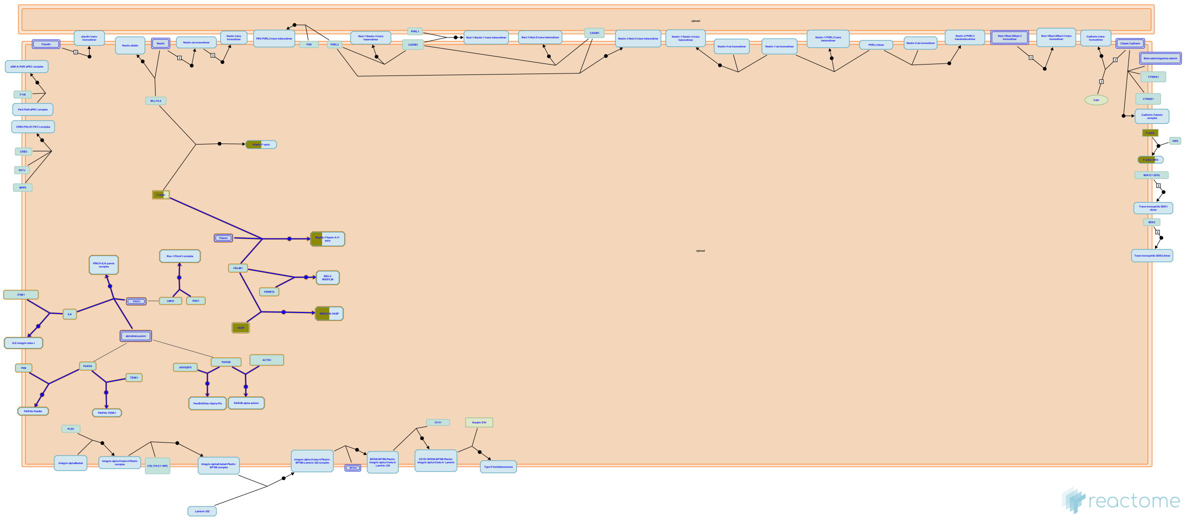

Cell-extracellular matrix (ECM) interactions play a critical role in regulating a variety of cellular processes in multicellular organisms including motility, shape change, survival, proliferation and differentiation. Cell-ECM contact is mediated by transmembrane cell adhesion receptors, such as integrins, that interact with extracellular matrix proteins as well as a number of cytoplasmic adaptor proteins. Many of these adaptor proteins physically interact with the actin cytoskeleton or function in signal transduction.

Several protein complexes interact with the cytoplasmic tail of integrins and function in transducing bi-directional signals between the ECM and intracellular signaling pathways (reviewed in Sepulveda et al., 2005).

Early events that are triggered by interactions with ECM, such as formation/turnover of Focal Adhesions, regulation of actin dynamics and protrusion of lamellipodia to promote cellular spreading and motility are modulated by PINCH- ILK- parvin complexes (see Sepulveda et al., 2005). A number of partners of the PINCH-ILK-parvin complex components have been identified that regulate and/or mediate the functions of these complexes (reviewed in Wu, 2004). Interactions with some of these partners modulate cytoskeletal remodeling and cell spreading.


##### Edit history

| Date | Action | Author |
|---|---|---|
| 2009-10-12 | Authored | Matthews L |
| 2009-11-10 | Edited | Matthews L |
| 2009-11-12 | Reviewed | Wu C |
| 2009-11-12 | Created | Matthews L |
| 2021-05-22 | Modified | Shorser S |

##### Entities found in this pathway (2)

| Input | UniProt Id |
|---|---|
| ACTG1 | P60709, P63261 |
| VASP | P50552 |

#### 19. ATF4 activates genes in response to endoplasmic reticulum stress (R-HSA-380994)

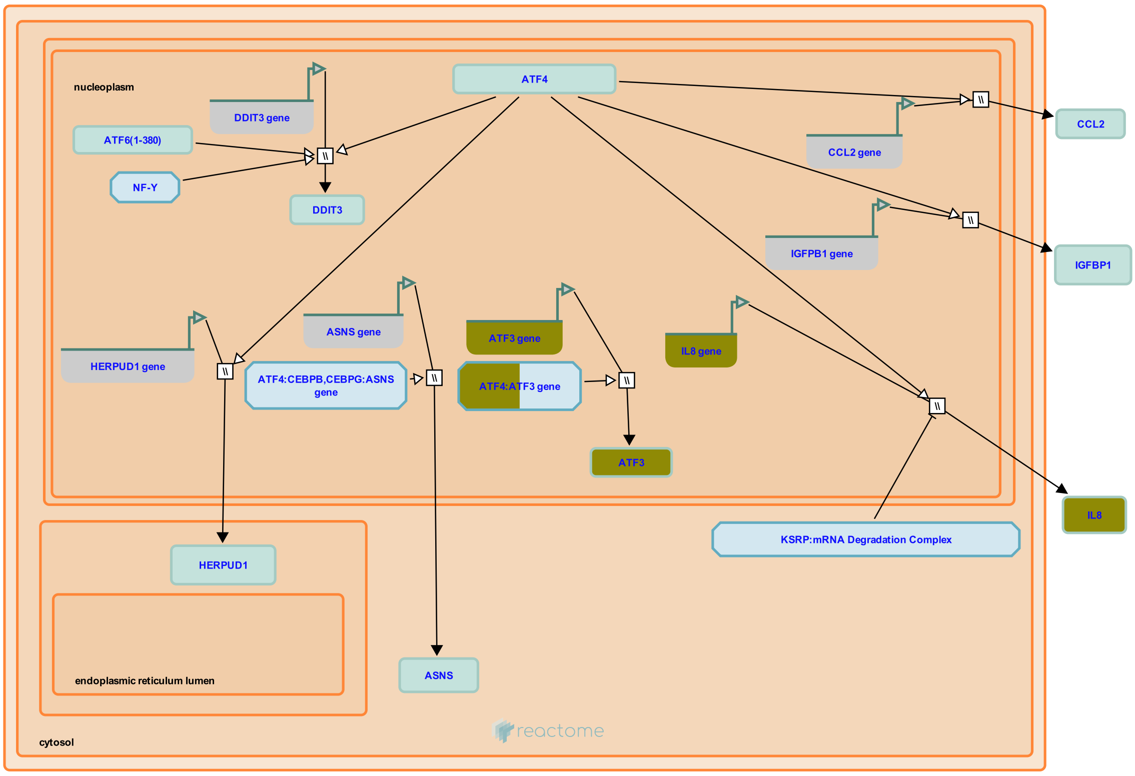


ATF4 is a transcription factor and activates expression of IL-8, MCP1, IGFBP-1, CHOP, HERP1 and ATF3.


##### Edit history

| Date | Action | Author |
|---|---|---|
| 2008-11-19 | Created | May B |

| Date | Action | Author |
|---|---|---|
| 2008-12-02 | Reviewed | Matthews L, D'Eustachio P, Gillespie ME |
| 2009-06-02 | Edited | May B |
| 2009-06-02 | Authored | May B |
| 2010-04-30 | Reviewed | Urano F |
| 2020-11-11 | Modified | D'Eustachio P |

#### Entities found in this pathway (2)

| Input | UniProt Id |
|---|---|
| ATF3 | P18847 |

| Input | UniProt Id |
|---|---|
| IL8 | P10145 |

| Input | Ensembl Id |
|---|---|
| ATF3 | ENSG00000162772 |

| Input | Ensembl Id |
|---|---|
| IL8 | ENSG00000169429 |

#### 20. TP53 Regulates Transcription of Genes Involved in G1 Cell Cycle Arrest (R-HSA-6804116)

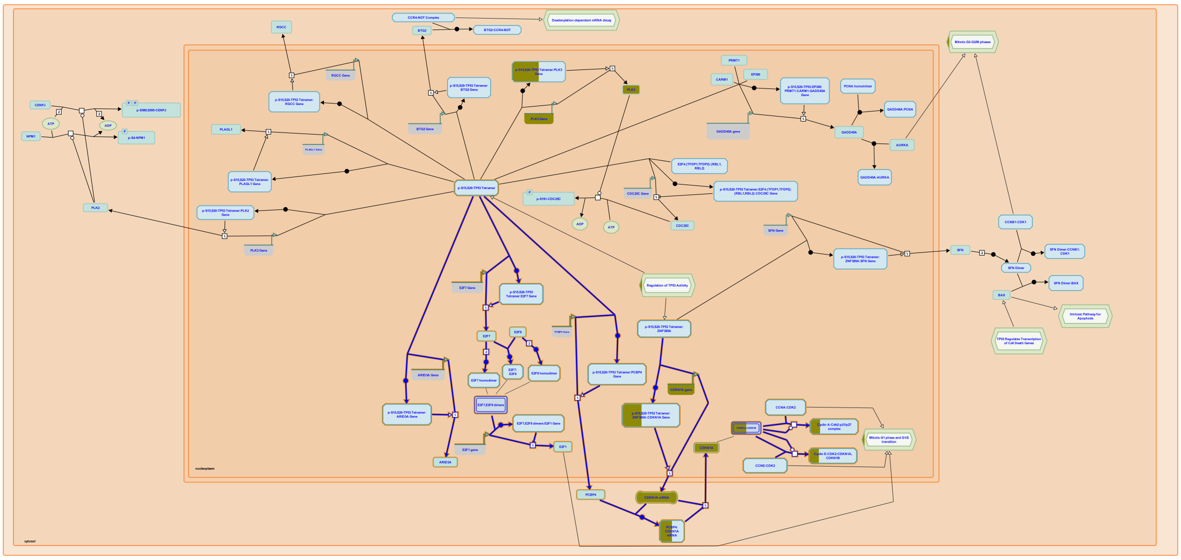

The most prominent TP53 target involved in G1 arrest is the inhibitor of cyclin-dependent kinases CDKN1A (p21). CDKN1A is one of the earliest genes induced by TP53 (El-Deiry et al. 1993). CDKN1A binds and inactivates CDK2 in complex with cyclin A (CCNA) or E (CCNE), thus preventing G1/S transition (Harper et al. 1993). Considering its impact on the cell cycle outcome, CDKN1A expression levels are tightly regulated. For instance, under prolonged stress, TP53 can induce the transcription of an RNA binding protein PCBP4, which can bind and destabilize CDKN1A mRNA, thus alleviating G1 arrest and directing the affected cell towards G2 arrest and, possibly, apoptosis (Zhu and Chen 2000, Scoumanne et al. 2011). Expression of E2F7 is directly induced by TP53. E2F7 contributes to G1 cell cycle arrest by repressing transcription of E2F1, a transcription factor that promotes expression of many genes needed for G1/S transition (Aksoy et al. 2012, Carvajal et al. 2012). ARID3A is a direct transcriptional target of TP53 (Ma et al. 2003) that may promote G1 arrest by cooperating with TP53 in induction of CDKN1A transcription (Lestari et al. 2012). However, ARID3A may also promote G1/S transition by stimulating transcriptional activity of E2F1 (Suzuki et al. 1998, Peeper et al. 2002).

TP53 has co-factors that are key determinants of transcriptional selectivity within the p53 network. For instance, the zinc finger transcription factor ZNF385A (HZF) is a direct transcriptional target of TP53 that can form a complex with TP53 and facilitate TP53-mediated induction of CDKN1A, strongly favouring cell cycle arrest over apoptosis (Das et al. 2007).

#### Edit history

| Date | Action | Author |
|---|---|---|
| 2015-10-08 | Created | Orlic-Milacic M |
| 2015-10-14 | Edited | Orlic-Milacic M |
| 2015-10-14 | Authored | Orlic-Milacic M |
| 2016-02-04 | Reviewed | Zaccara S, Inga A |
| 2017-01-03 | Revised | Orlic-Milacic M |
| 2021-05-31 | Modified | Shorser S |

#### Entities found in this pathway (1)

| Input | UniProt Id |
|---|---|
| CDKN1A | P38936 |

| Input | Ensembl Id |
|---|---|
| CDKN1A | ENSG00000124762, ENST00000244741 |

#### 21. IkBA variant leads to EDA-ID (R-HSA-5603029)

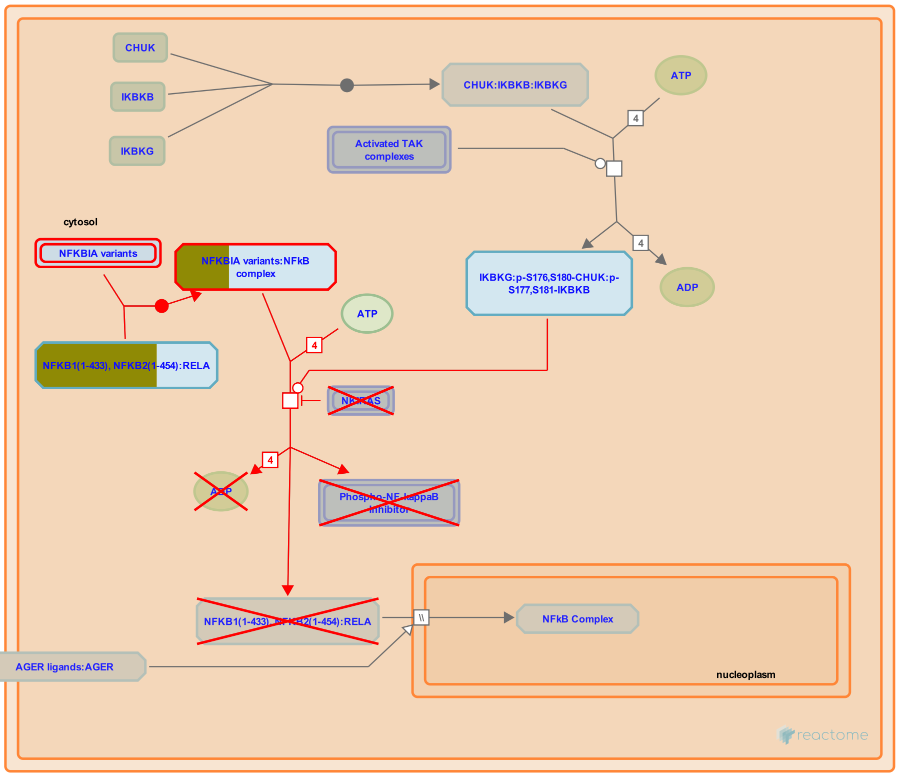


**Diseases:** primary immunodeficiency disease.

The nuclear factor kappa B (NFkB) family of transcription factors is kept inactive in the cytoplasm by the inhibitor of kappa B (IkB) family members IKBA (IkB alpha, NFKBIA), IKBB (IkB beta, NFKBIB) and IKBE (IkB epsilon, NFKBIE) (Oeckinghaus A and Ghosh S 2009). Multiple stimuli such as inflammatory cytokines, microbial products or various types of stress activate NFkB signaling leading to stimuli-induced phosphorylation of IkB molecule (Scherer DC et al. 1995; Alkalay I et al. 1995; Lawrence T 2009; Hoesel B and Schmid JA 2013). The phosphorylation of IkB proteins triggers their polyubiquitination and subsequent degradation by 26S proteasome, allowing free NFkB dimer to translocate to the nucleus where it directs the expression of target genes. Studies have identified an autosomal dominant form of ectodermal dysplasia with immunodeficiency (AD-EDA-ID) caused by a hypermorphic heterozygous mutation of NFKBIA/IKBA gene. The IKBA defects prevent the phosphorylation and degradation of IKBA protein resulting in gain-of-function condition with the enhanced inhibitory capacity of IKBA in sequestering NF?B dimers in the cytoplasm (Courtois G et al. 2003; Lopes-Granados E et al. 2008; Schimke LF et al. 2013).


#### Edit history

| Date | Action | Author |
|---|---|---|
| 2014-05-21 | Authored | Shamovsky V |
| 2014-06-26 | Created | Shamovsky V |
| 2014-09-06 | Reviewed | D'Eustachio P |
| 2015-02-10 | Edited | Shamovsky V |
| 2015-02-15 | Reviewed | McDonald DR |
| 2021-05-31 | Modified | Shorser S |

#### Entities found in this pathway (2)

| Input | UniProt Id | Input | UniProt Id |
|---|---|---|---|
| NFKB1 | P19838 | NFKB2 | Q00653 |

#### 22. FGFR1 ligand binding and activation (R-HSA-190242)

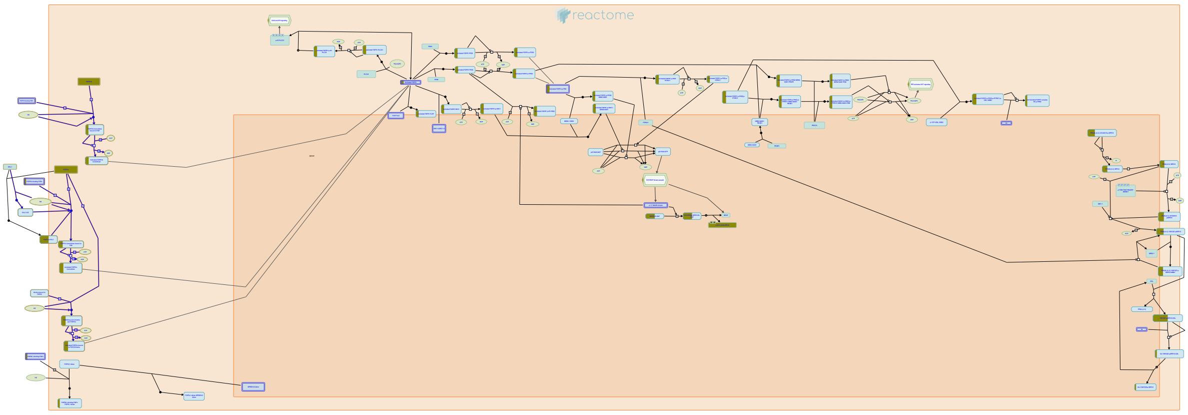

The vertebrate fibroblast growth factor receptor 1 (FGFR1) is alternatively spliced generating multiple variants that are differentially expressed during embryo development and in the adult body. The restricted expression patterns of FGFR1 isoforms, together with differential expression and binding of specific ligands, leads to activation of common FGFR1 signal transduction pathways, but may result in distinctively different biological responses as a result of differences in cellular context. FGFR1 isoforms are also present in the nucleus in complex with various fibroblast growth factors where they function to regulate transcription of target genes.

FGFR is probably activated by NCAM very differently from the way by which it is activated by FGFs, reflecting the different conditions for NCAM-FGFR and FGF-FGFR interactions. The affinity of FGF for FGFR is approximately 10e6 times higher than that of NCAM for FGFR. Moreover, in the brain NCAM is constantly present on the cell surface at a much higher (micromolar) concentration than FGFs, which only appear transiently in the extracellular environment in the nanomolar range.


##### Edit history

| Date | Action | Author |
|---|---|---|
| 2006-12-18 | Created | de Bono B |
| 2007-01-10 | Authored | de Bono B |
| 2007-02-07 | Reviewed | Mohammadi M |
| 2007-02-11 | Edited | D'Eustachio P, de Bono B |
| 2016-01-06 | Reviewed | Grose RP |

| Date | Action | Author |
|---|---|---|
| 2021-05-22 | Modified | Shorser S |

#### Entities found in this pathway (2)

| Input | UniProt Id | Input | UniProt Id |
|---|---|---|---|
| FGF17 | O60258-1 | FGFR1 | P11362-1, P11362-19 |

#### 23. Phospholipase C-mediated cascade: FGFR1 (R-HSA-5654219)

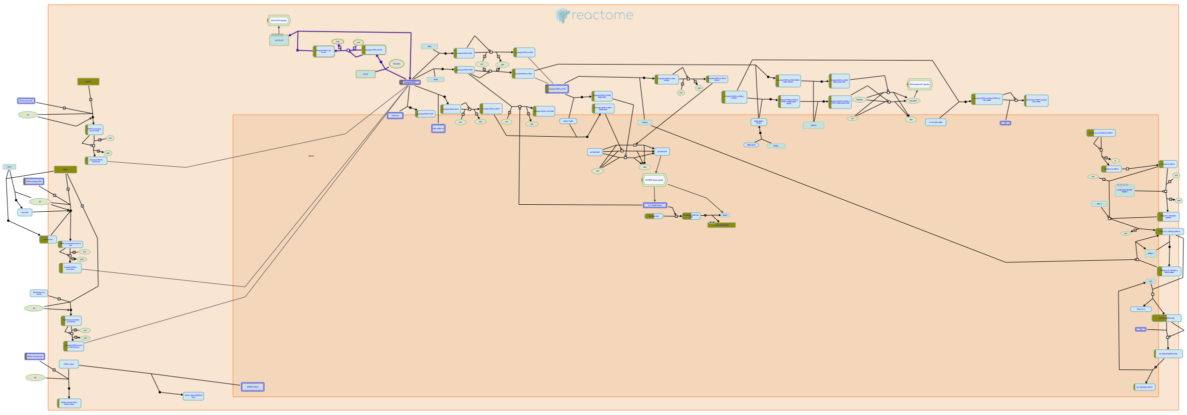

**Cellular compartments:** plasma membrane.

Phospholipase C-gamma (PLC-gamma) is a substrate of the fibroblast growth factor receptor (FGFR) and other receptors with tyrosine kinase activity. It is known that the src homology region 2 (SH2 domain) of PLC-gamma and of other signaling molecules (such as GTPase-activating protein and phosphatidylinositol 3-kinase-associated p85) direct their binding toward autophosphorylated tyrosine residues of the FGFR. Recruitment of PLC-gamma results in its phosphorylation and activation by the receptor. Activated PLC-gamma hydrolyzes phosphatidyl inositol[4,5] P2 to form the second messengers diacylglycerol (DAG) and Ins [1,4,5]P3, which stimulate calcium release and activation of calcium/calmodulin dependent kinases.


##### Edit history

| Date | Action | Author |
|---|---|---|
| 2007-01-10 | Authored | de Bono B |
| 2007-02-07 | Reviewed | Mohammadi M |
| 2010-02-03 | Edited | Jupe S |
| 2014-12-04 | Created | Rothfels K |
| 2021-05-22 | Modified | Shorser S |

##### Entities found in this pathway (2)

| Input | UniProt Id | Input | UniProt Id |
|---|---|---|---|
| FGF17 | O60258-1 | FGFR1 | P11362-1, P11362-19 |

#### 24. Negative regulation of FGFR1 signaling (R-HSA-5654726)

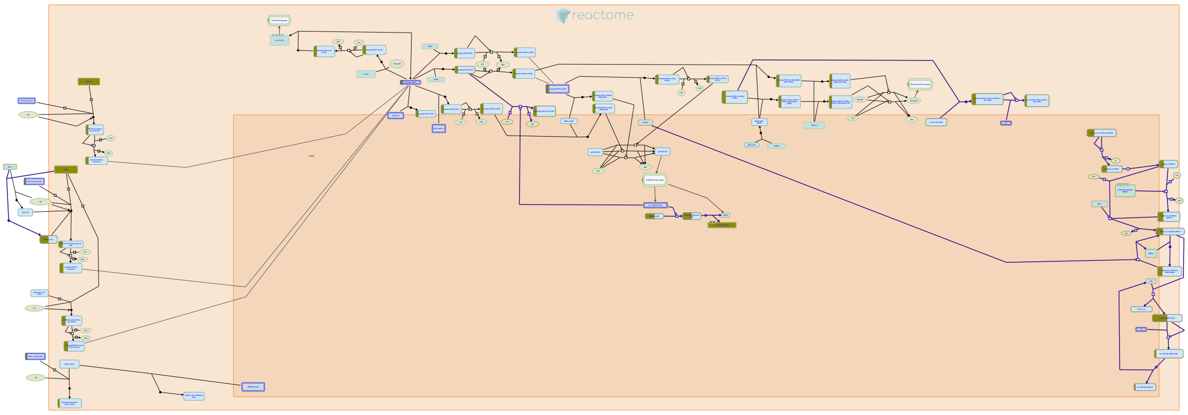

**Cellular compartments:** cytosol, extracellular region, plasma membrane.

Once activated, the FGFR signaling pathway is regulated by numerous negative feedback mechanisms. These include downregulation of receptors through CBL-mediated ubiquitination and endocytosis, ERK-mediated inhibition of FRS2-tyrosine phosphorylation and the attenuation of ERK signaling through the action of dual-specificity phosphatases, IL17RD/SEF, Sprouty and Spred proteins. A number of these inhibitors are themselves transcriptional targets of the activated FGFR pathway.


##### Edit history

| Date | Action | Author |
|---|---|---|
| 2011-08-15 | Authored | Rothfels K |
| 2011-08-26 | Reviewed | Gotoh N |
| 2014-12-04 | Created | Rothfels K |
| 2016-01-06 | Reviewed | Grose RP |
| 2021-05-22 | Modified | Shorser S |

##### Entities found in this pathway (3)

| Input | UniProt Id | Input | UniProt Id | Input | UniProt Id |
|---|---|---|---|---|---|
| FGF17 | O60258-1 | FGFR1 | P11362-1, P11362-19 | SPRY2 | O43597 |

#### 25. FGFR1 mutant receptor activation (R-HSA-1839124)

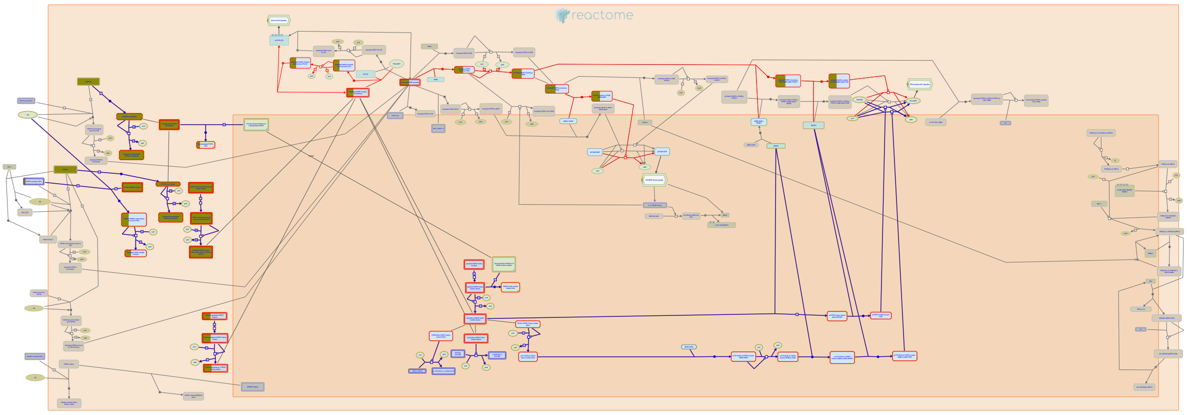

**Cellular compartments:** plasma membrane, cytosol, extracellular region.

**Diseases:** cancer, bone development disease.

The FGFR1 gene has been shown to be subject to activating mutations, chromosomal rearrangements and gene amplification leading to a variety of proliferative and developmental disorders depending on whether these events occur in the germline or arise somatically (reviewed in Webster and Donoghue, 1997; Burke, 1998; Cunningham, 2007; Wesche, 2011; Greulich and Pollock, 2011). Many of the resulting mutant FGFR1 proteins can dimerize and promote signaling in a ligand-independent fashion, although signal transduction may still be amplified in the presence of ligand (reviewed in Turner and Gross, 2010; Greulich and Pollock, 2011; Wesche et al, 2011).


##### Edit history

| Date | Action | Author |
|---|---|---|
| 2011-10-27 | Created | Rothfels K |
| 2012-02-10 | Authored | Rothfels K |
| 2012-05-15 | Reviewed | Ezzat S |
| 2012-05-16 | Edited | Rothfels K |
| 2016-01-09 | Revised | Rothfels K |
| 2016-01-22 | Modified | Rothfels K |

#### Entities found in this pathway (2)

| Input | UniProt Id | Input | UniProt Id |
|---|---|---|---|
| FGF17 | O60258-1 | FGFR1 | P11362, P11362-1, P11362-19 |

### 6. Identifiers found

Below is a list of the input identifiers that have been found or mapped to an equivalent element in Reactome, classified by resource.

#### Entities (183)

| Input | UniProt Id | Input | UniProt Id | Input | UniProt Id |
|---|---|---|---|---|---|
| ABCC3 | O15438 | ACTA2 | P62736, P63267 | ACTG1 | P60709, P63261 |
| ACTN2 | P35609 | ADAMTS12 | P58397 | ADAMTS3 | O15072 |
| ALS2CL | Q60I27 | ANXA1 | P04083 | AOC3 | O75106, Q16853 |
| AP1G2 | O75843 | APBB1IP | Q7Z5R6 | ARHGAP24 | Q8N264 |
| ARHGEF25 | Q86VW2 | ARHGEF28 | Q8N1W1 | ARHGEF40 | Q8TER5 |
| ARNTL | O00327 | ART3 | O60678 | ASIC3 | Q9UHC3 |
| ATF3 | P18847 | BBS12 | Q6ZW61 | C5AR2 | Q9P296 |
| CACNA1H | O95180 | CACNA2D3 | Q8IZS8 | CCL4 | O00626, P13236 |
| CCL4L2 | Q8NHW4 | CD84 | Q9UIB8 | CDKN1A | P38936 |
| CHKA | P35790, Q9Y259 | COL13A1 | Q5TAT6 | COL22A1 | Q8NFW1 |
| COL5A2 | P05997 | CRABP1 | P29762 | CREM | Q03060-6 |
| CSF1 | P09603 | CSF1R | P07333 | CXCL1 | P09341 |
| DDAH2 | O95865 | DNA2 | P51530 | DOCK8 | Q8NF50 |
| DPH1 | Q9BZG8 | DRP2 | Q13474 | DSC2 | Q02487 |
| DSG2 | Q14126 | DSP | P15924 | DUSP2 | Q05923 |
| EFNA5 | P52803 | EMILIN3 | Q9NT22 | ENPP1 | P22413 |
| ENTPD1 | P49961 | EPB41 | P11171 | FBXL21 | Q9UKT6 |
| FGF17 | O60258-1 | FGFR1 | P11362-1, P11362-19 | FMNL3 | Q8IVF7 |
| FOSL1 | P15407 | FTCD | O95954 | GALNT12 | Q8IXK2 |
| GBP1 | P32455 | GFPT2 | O94808 | GPX3 | O75715, P22352 |
| GRIK3 | Q13003 | GRIN1 | Q05586 | HBEGF | Q99075 |
| HECW1 | Q76N89 | HMGA1 | P17096 | HMGN1 | P05114 |
| HTR3B | O95264 | ICAM1 | P05362 | IGF1 | P05019 |
| IGF2BP2 | Q9Y6M1 | IKZF1 | Q13422-1 | IL32 | P24001 |
| IL7R | P16871 | IL8 | P10145 | IRF1 | P10914 |
| ITGA2B | P08514 | ITGAX | P20702 | ITK | Q08881 |
| ITPR1 | Q14643 | JUN | P05412 | JUNB | P17275 |
| KCNJ1 | P48048 | KCNMB1 | Q16558 | KDM6B | O15054 |
| LCP2 | Q13094 | LGR6 | Q9HBX8 | LILRB4 | Q8N423 |
| LMNA | P02545 | LPAR5 | Q9H1C0 | LPCAT4 | Q643R3, Q6ZWT7 |
| LTV1 | Q96GA3 | MAFF | Q9ULX9 | MAMLD1 | Q13495 |
| MCL1 | Q07820 | MDK | P21741 | MEPE | Q9NQ76 |
| MNDA | P41218 | MYH11 | P35749 | MYL6 | P60660 |
| MYO15A | Q9UKN7 | NAALAD2 | Q9Y3Q0 | NFATC1 | O95644 |
| NFATC2 | Q12968, Q13469 | NFKB1 | P19838 | NFKB2 | Q00653 |
| NISCH | Q9Y2I1 | NOS1 | P29475 | NR4A2 | P43354 |
| NRG4 | Q8WWG1 | ODC1 | Q9H936 | OPN3 | Q9H1Y3 |
| OR2AK2 | Q8NG84 | OR2L13 | Q8N349 | OR2M3 | Q8NG83 |
| OR2M5 | A3KFT3 | OSBPL3 | Q9H4L5 | PAPPA | Q13219 |
| PHLDA1 | Q8WV24 | PIK3R5 | Q8WYR1 | PIP4K2C | Q8TBX8 |
| PLA2G4B | P0C869, Q86XP0 | PLAUR | Q03405 | PLEKHG5 | O94827 |

| Input | UniProt Id | Input | UniProt Id | Input | UniProt Id |
|---|---|---|---|---|---|
| PLEKHO2 | Q8TD55 | PLK3 | Q9H4B4 | PLN | P26678 |
| PNPLA5 | Q7Z6Z6 | POLQ | O75417 | PPIL6 | Q8IXY8 |
| PTK2B | Q14289 | PTPRZ1 | P23471 | PXDN | Q92626 |
| RASAL1 | O95294 | RASGRP1 | O95267 | RELB | Q01201 |
| REM1 | P18074 | RGS16 | O15492 | RHEB | Q15382 |
| RUNX1 | Q01196 | SEC16B | Q96JE7 | SEMA3A | Q14563 |
| SERPINA3 | P01011, P29622 | SERPIND1 | P05546 | SIRT3 | Q9NTG7 |
| SKIL | P12757 | SLC16A1 | P53985 | SLC22A6 | Q4U2R8 |
| SLC25A2 | Q9BXI2 | SLC29A1 | Q99808 | SLC4A7 | Q9Y6M7 |
| SLITRK5 | O94991 | SOX6 | P35712 | SPRY2 | O43597 |
| ST5 | P78524 | ST6GALNAC5 | Q9BVH7 | STON1 | Q9Y6Q2 |
| SYCP2 | Q9BX26 | SYN3 | O14994 | SYT10 | Q6XYQ8 |
| TGFB1 | P01137 | THBS2 | P35442 | TLE3 | Q04726 |
| TMC2 | Q8TDI7 | TMSB4X | P62328 | TNFAIP6 | P98066 |
| TNFRSF12A | Q9NP84 | TNFRSF25 | Q93038 | TPM4 | P67936 |
| TRIM38 | O00635 | TYROBP | O43914 | USP28 | Q96RU2 |
| VASP | P50552 | WDR1 | O75083 | WEE1 | P30291, Q99640 |
| WFDC2 | P19957 | WNT16 | Q9UBV4 | ZFP36 | P26651 |
| ZIM2 | Q9NZV7 | ZNF267 | Q14586 | ZNF273 | Q14593 |
| ZNF436 | Q9C0F3 | | | | |

| Input | Ensembl Id | Input | Ensembl Id | Input | Ensembl Id |
|---|---|---|---|---|---|
| ACTA2 | ENSG00000107796 | ANXA1 | ENSG00000135046 | ARNTL | ENSG00000133794 |
| ATF3 | ENSG00000162772 | CCL4 | ENSG00000275302 | CDKN1A | ENSG00000124762 |
| CSF1 | ENSG00000184371 | CSF1R | ENSG00000182578 | CXCL1 | ENSG00000163739 |
| GBP1 | ENSG00000117228 | GPRC5A | ENST00000014914 | GRIN1 | ENSG00000169258 |
| ICAM1 | ENSG00000090339 | IL8 | ENSG00000169429 | IRF1 | ENSG00000125347 |
| ITGA2B | ENSG00000005961 | ITGAX | ENSG00000140678 | JUNB | ENSG00000171223 |
| KDM6B | ENSG00000132510 | LMNA | ENSG00000160789 | MCL1 | ENSG00000143384 |
| PLA2G4B | ENSG00000243708 | PLK3 | ENSG00000173846 | RUNX1 | ENSG00000159216, ENST00000344691 |
| SIRT3 | ENSG00000142082 | TGFB1 | ENSG00000105329 | TRIM38 | ENSG00000112343 |

| Input | miRBase Id |
|---|---|
| MIR24-2 | MI0000081 |

### 7. Identifiers not found

These 226 identifiers were not found neither mapped to any entity in Reactome.

| | | | | | | | |
|---|---|---|---|---|---|---|---|
| ABTB1 | AC003102.3 | AC005537.2 | AC007277.3 | AC009878.2 | AC011288.2 | AC011995.3 | AC012065.7 |
| AC017002.2 | AC019186.1 | AC092661.1 | AC106827.1 | AC117947.1 | ADAMTSL4-AS1 | AIRE | AJ006998.2 |
| AL022393.7 | AL589743.1 | ANGPTL1 | ANKRD20A11P | ANKRD20A8P | AP000525.9 | ARID5A | ATP13A3 |
| ATP6V1G1P4 | ATRNL1 | BCL6B | BMP5 | C11orf96 | C21orf128 | C2orf61 | C4orf19 |
| CCBE1 | CCDC144A | CCNYL2 | CELSR1 | CEP164P1 | CERKL | CLEC19A | CLEC1A |
| CNN1 | CNNM1 | CORO6 | CPED1 | CPXM1 | CSDC2 | CTB-111F10.1 | CTC-451A6.4 |
| CTD-2050E21.1 | CTD-2262B20.1 | CTD-2303H24.2 | CTD-2554C21.3 | CTD-2562J15.6 | CYP1B1-AS1 | CYP4F29P | CYP4F35P |
| CYTIP | DPP10-AS1 | EGFEM1P | EIF4BP6 | EMP1 | EPHX4 | ERRFI1 | ERVK3-1 |
| ERVMER34-1 | ETV3 | FAIM2 | FAM101B | FAM106B | FAM106CP | FAM126A | FAM179A |
| FAM211A | FAM46C | FAM83G | FAM95B1 | FHOD1 | FREM2 | FRZB | GDF7 |
| GEM | GPR146 | GPR64 | GPR78 | HABP2 | HLA-DPB2 | HLA-L | HYDIN2 |
| IGFN1 | JPH4 | KCNT1 | KLF6 | LIMA1 | LINC00152 | LINC00284 | LINC00575 |
| LINC00607 | LINC00702 | LINC00943 | LL22NC03-N14H11.1 | LRRC39 | LYNX1 | LZTS3 | MAPK15 |
| MAPK8IP2 | MIR17HG | MIR4482-1 | MIR4697 | MOB3C | MTUS2 | MYRFL | NFKBIZ |
| NIFK | NKAIN4 | NME5 | NME9 | NPM2 | OGFOD3 | OR1H1P | OR2L9P |
| OSR1 | PAMR1 | PAWR | PCDH12 | PDLIM3 | PDLIM4 | PLS3 | PRRT3 |
| PRSS27 | PTP4A1 | PTPRQ | RCBTB2 | RHOT1P1 | RMRP | RN7SKP203 | RNF148 |
| RNU6-316P | RNVU1-15 | RP1-140K8.5 | RP1-144F13.4 | RP1-28C20.1 | RP1-90J20.12 | RP11-138A9.1 | RP11-14I17.3 |
| RP11-153K16.1 | RP11-15H20.5 | RP11-15H20.6 | RP11-16P6.1 | RP11-170N16.2 | RP11-195B21.3 | RP11-21L23.2 | RP11-264E23.1 |
| RP11-282E4.1 | RP11-313F23.5 | RP11-314P15.2 | RP11-341A11.2 | RP11-347C12.2 | RP11-34P13.13 | RP11-382N13.3 | RP11-41O4.1 |
| RP11-430H10.4 | RP11-435B5.5 | RP11-439C15.4 | RP11-448G15.3 | RP11-497D6.5 | RP11-531A24.5 | RP11-539I5.1 | RP11-551L14.1 |
| RP11-561E1.1 | RP11-594C13.1 | RP11-5N11.7 | RP11-638F5.1 | RP11-664H17.1 | RP11-667K14.3 | RP11-667K14.4 | RP11-66B24.4 |
| RP11-757O6.1 | RP11-76E17.4 | RP11-79E3.2 | RP11-894J14.5 | RP11-958J22.1 | RP3-467K16.4 | RP4-564M11.2 | RP4-809F18.1 |
| RP5-944M2.2 | RP5-944M2.3 | RPL24P8 | RPS16P5 | RPS20P22 | RPS26P21 | RTBDN | SAMD3 |
| SDHCP4 | SERPINI1 | SETP12 | SGCD | SLC38A11 | SNORA22 | SNX18P13 | SNX18P26 |
| SNX18P4 | SNX32 | SOGA2 | STK17A | STK36 | SYNC | SYT17 | TCP10L |
| TFAP4 | TFPI2 | TMEM120B | TMEM63C | TMEM74B | TRANK1 | TSPAN2 | TTC39A |
| U95743.1 | USP32P1 | VWA3A | Y_RNA | ZC3H12A | ZFP36L2 | ZNF192P1 | ZNF385B |
| ZNF572 | snoMe28S-Am2634 | | | | | | |
