## Supplementary material for "Compartment-specific total RNA profile of Hippocampal and Cortical cells from Mesial Temporal Lobe Epilepsy tissue": Other supplemental files_Reactome: HC_mTLE non-HS_Cytoplasm_DOWNspecific_report.pdf

### Pathway Analysis Report

This report contains the pathway analysis results for the submitted sample ''. Analysis was performed against Reactome version 77 on 14/09/2021. The web link to these results is:

### 3. Genome-wide overview

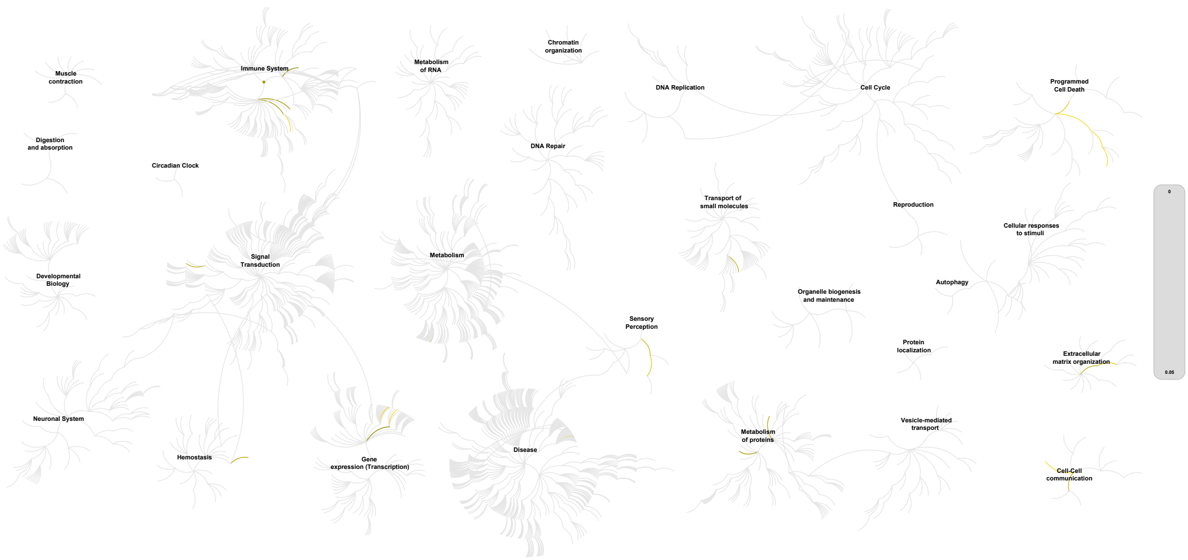


This figure shows a genome-wide overview of the results of your pathway analysis. Reactome pathways are arranged in a hierarchy. The center of each of the circular "bursts" is the root of one top-level pathway, for example "DNA Repair". Each step away from the center represents the next level lower in the pathway hierarchy. The color code denotes over-representation of that pathway in your input dataset. Light grey signifies pathways which are not significantly over-represented.

### 4. Most significant pathways

The following table shows the 25 most relevant pathways sorted by p-value.

| Pathway name | Entities | | | | Reactions | |
|---|---|---|---|---|---|---|
| | found | ratio | p-value | FDR* | found | ratio |
| Caspase-mediated cleavage of cytoskeletal proteins | 3 / 12 | 8.25e-04 | 3.61e-06 | 5.12e-04 | 5 / 10 | 7.40e-04 |
| RUNX2 regulates genes involved in differentiation of myeloid cells | 2 / 6 | 4.13e-04 | 1.00e-04 | 0.005 | 2 / 2 | 1.48e-04 |
| Apoptotic cleavage of cellular proteins | 3 / 38 | 0.003 | 1.10e-04 | 0.005 | 5 / 38 | 0.003 |
| Apoptotic execution phase | 3 / 54 | 0.004 | 3.07e-04 | 0.008 | 5 / 57 | 0.004 |
| Type I hemidesmosome assembly | 2 / 11 | 7.56e-04 | 3.35e-04 | 0.008 | 6 / 6 | 4.44e-04 |
| RUNX1 regulates transcription of genes involved in differentiation of myeloid cells | 2 / 11 | 7.56e-04 | 3.35e-04 | 0.008 | 2 / 7 | 5.18e-04 |
| Metal sequestration by antimicrobial proteins | 2 / 13 | 8.94e-04 | 4.66e-04 | 0.009 | 1 / 5 | 3.70e-04 |
| Defective SLC40A1 causes hemochromatosis 4 (HFE4) (macrophages) | 1 / 4 | 2.75e-04 | 0.01 | 0.137 | 1 / 1 | 7.40e-05 |
| Sensory processing of sound by outer hair cells of the cochlea | 2 / 64 | 0.004 | 0.01 | 0.137 | 1 / 8 | 5.92e-04 |
| Apoptosis | 3 / 192 | 0.013 | 0.011 | 0.137 | 5 / 141 | 0.01 |
| Assembly of collagen fibrils and other multimeric structures | 2 / 67 | 0.005 | 0.011 | 0.137 | 1 / 26 | 0.002 |
| Defective CP causes aceruloplasminemia (ACERULOP) | 1 / 6 | 4.13e-04 | 0.014 | 0.158 | 1 / 1 | 7.40e-05 |
| Sensory processing of sound | 2 / 87 | 0.006 | 0.019 | 0.173 | 1 / 13 | 9.62e-04 |
| Programmed Cell Death | 3 / 238 | 0.016 | 0.019 | 0.173 | 5 / 197 | 0.015 |
| Sodium-coupled sulphate, di- and tri-carboxylate transporters | 1 / 9 | 6.19e-04 | 0.021 | 0.173 | 1 / 5 | 3.70e-04 |
| Cell junction organization | 2 / 94 | 0.006 | 0.022 | 0.173 | 6 / 37 | 0.003 |
| Collagen formation | 2 / 104 | 0.007 | 0.026 | 0.179 | 1 / 77 | 0.006 |
| Calcitonin-like ligand receptors | 1 / 11 | 7.56e-04 | 0.026 | 0.179 | 2 / 4 | 2.96e-04 |
| Post-translational protein phosphorylation | 2 / 109 | 0.007 | 0.028 | 0.179 | 1 / 1 | 7.40e-05 |
| Arachidonate production from DAG | 1 / 13 | 8.94e-04 | 0.031 | 0.179 | 1 / 3 | 2.22e-04 |
| Antimicrobial peptides | 2 / 123 | 0.008 | 0.035 | 0.179 | 1 / 58 | 0.004 |
| Regulation of Insulin-like Growth Factor (IGF) transport and uptake by Insulin-like Growth Factor Binding Proteins (IGFBPs) | 2 / 127 | 0.009 | 0.038 | 0.179 | 1 / 14 | 0.001 |

| Pathway name | Entities | | | | Reactions | |
|---|---|---|---|---|---|---|
| | found | ratio | p-value | FDR* | found | ratio |
| Advanced glycosylation endproduct receptor signaling | 1 / 16 | 0.001 | 0.038 | 0.179 | 1 / 4 | 2.96e-04 |
| Cell-Cell communication | 2 / 133 | 0.009 | 0.041 | 0.179 | 6 / 60 | 0.004 |
| Prolactin receptor signaling | 1 / 18 | 0.001 | 0.042 | 0.179 | 14 / 14 | 0.001 |

* False Discovery Rate

### 5. Pathways details

For every pathway of the most significant pathways, we present its diagram, as well as a short summary, its bibliography and the list of inputs found in it.

#### 1. Caspase-mediated cleavage of cytoskeletal proteins (R-HSA-264870)

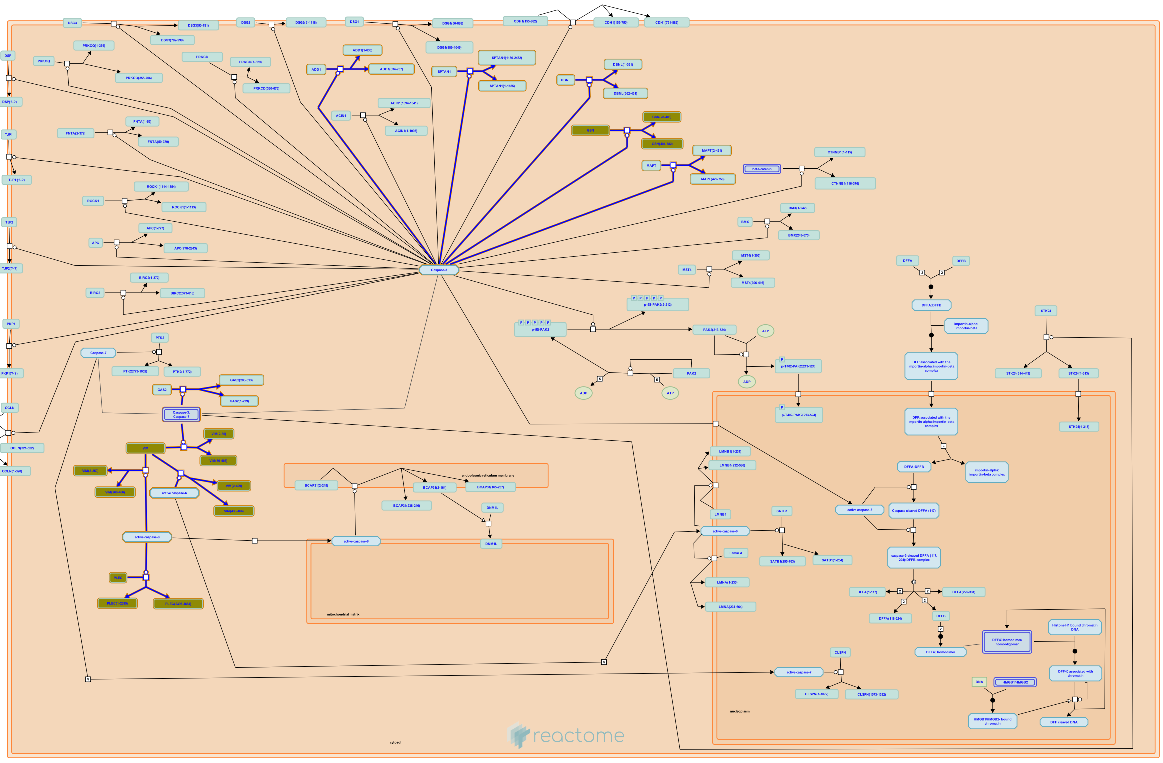

Caspase-mediated cleavage of a number of proteins in the cortical actin network ( ) microfilament system and others involved in maintenance of the cytoskeletal architecture (vimentin, or Gas2 and plectin) may directly contribute to apoptotic changes in cell shape.


##### Edit history

| Date | Action | Author |
|---|---|---|
| 2007-09-03 | Authored | Schulze-Osthoff K |
| 2008-04-14 | Edited | Matthews L |
| 2008-04-14 | Created | Matthews L |
| 2008-06-11 | Reviewed | Ranganathan S |

| Date | Action | Author |
| --- | --- | --- |
| 2008-06-12 | Edited | Matthews L |
| 2021-05-31 | Modified | Shorser S |

#### Entities found in this pathway (3)

| Input | UniProt Id | Input | UniProt Id | Input | UniProt Id |
| --- | --- | --- | --- | --- | --- |
| GSN | P06396 | PLEC | Q15149 | VIM | P08670 |

#### 2. RUNX2 regulates genes involved in differentiation of myeloid cells (R-HSA-8941333)

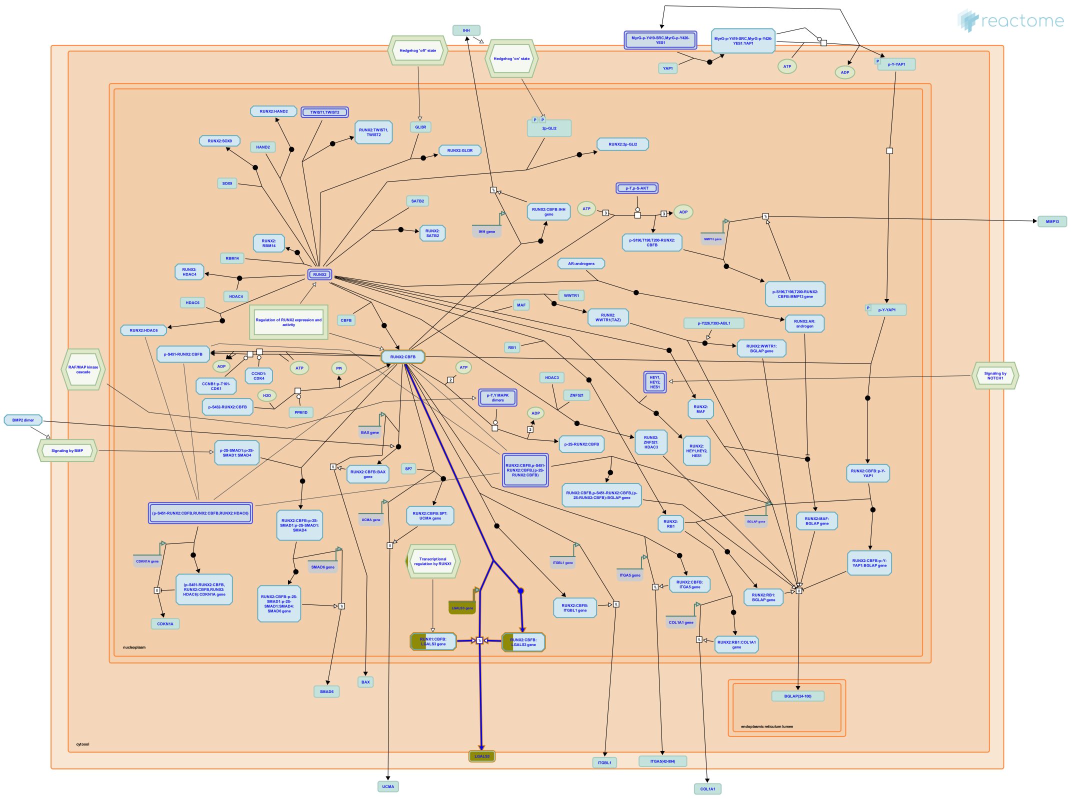

Both RUNX2 and RUNX1 can stimulate transcription of the LGALS3 gene, encoding Galectin-3 (Vladimirova et al. 2008, Zhang et al. 2009). Galectin 3 is expressed in myeloid progenitors and its levels increase during the maturation process (Le Marer 2000). Galectin 3 is highly expressed in pituitary tumors and glioma (Vladimirova et al. 2008, Zhang et al. 2009).


##### Edit history

| Date | Action | Author |
|---|---|---|
| 2016-09-30 | Authored | Orlic-Milacic M |
| 2016-09-30 | Created | Orlic-Milacic M |
| 2017-08-04 | Reviewed | Ducy P |
| 2017-08-09 | Edited | Orlic-Milacic M |
| 2021-05-31 | Modified | Shorser S |

##### Entities found in this pathway (1)

| Input | UniProt Id |
|---|---|
| LGALS3 | P17931 |

| Input | Ensembl Id |
|---|---|
| LGALS3 | ENSG00000131981 |

#### 3. Apoptotic cleavage of cellular proteins (R-HSA-111465)

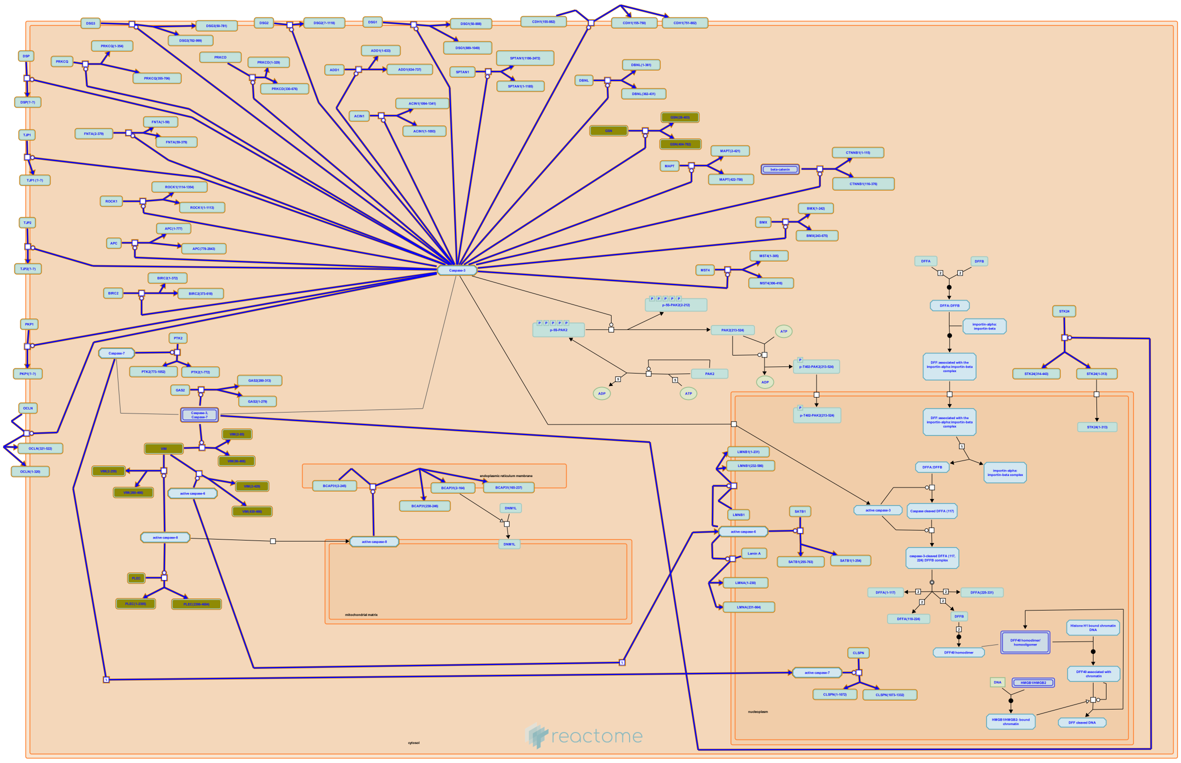

**Cellular compartments:** cytosol.

Apoptotic cell death is achieved by the caspase-mediated
cleavage of various vital proteins. Among caspase targets are proteins such as E-cadherin, Beta-catenin, alpha fodrin, GAS2, FADK, alpha adducin, HIP-55, and desmoglein involved in cell adhesion and maintenance of the cytoskeletal architecture. Cleavage of proteins such as APC and CIAP1 can further stimulate apoptosis by produce proapoptotic proteins (reviewed in Fischer et al., 2003. See also Wee et al., 2006 and the CASVM Caspase Substrates Database: http://www.casbase.org/casvm/squery/index.html ).


##### Edit history

| Date | Action | Author |
|---|---|---|
| 2004-02-17 | Created | Alnemri E |
| 2007-09-03 | Authored | Schulze-Osthoff K |
| 2007-11-23 | Reviewed | Ranganathan S |
| 2008-02-08 | Edited | Matthews L |

| Date | Action | Author |
| --- | --- | --- |
| 2008-05-18 | Revised | Matthews L |
| 2021-05-31 | Modified | Shorser S |

#### Entities found in this pathway (3)

| Input | UniProt Id | Input | UniProt Id | Input | UniProt Id |
| --- | --- | --- | --- | --- | --- |
| GSN | P06396 | PLEC | Q15149 | VIM | P08670 |

#### 4. Apoptotic execution phase (R-HSA-75153)

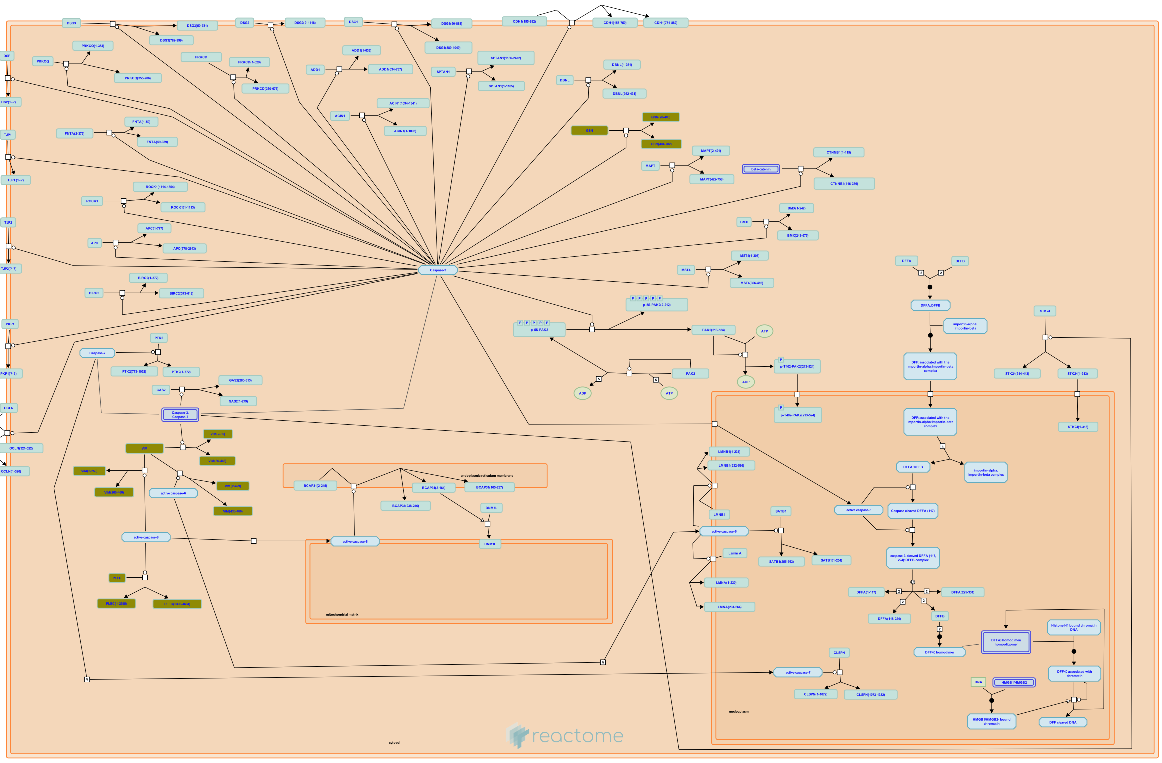

In the execution phase of apoptosis, effector caspases cleave vital cellular proteins leading to the morphological changes that characterize apoptosis. These changes include destruction of the nucleus and other organelles, DNA fragmentation, chromatin condensation, cell shrinkage and cell detachment and membrane blebbing (reviewed in Fischer et al., 2003).


##### Edit history

| Date | Action | Author |
|---|---|---|
| 2004-01-28 | Created | Tschopp J |
| 2004-02-17 | Authored | Alnemri E |
| 2007-11-23 | Reviewed | Ranganathan S |
| 2008-02-12 | Edited | Matthews L |
| 2008-05-20 | Revised | Matthews L |
| 2021-05-31 | Modified | Shorser S |

##### Entities found in this pathway (3)

| Input | UniProt Id | Input | UniProt Id | Input | UniProt Id |
|---|---|---|---|---|---|
| GSN | P06396 | PLEC | Q15149 | VIM | P08670 |

#### 5. Type I hemidesmosome assembly (R-HSA-446107)

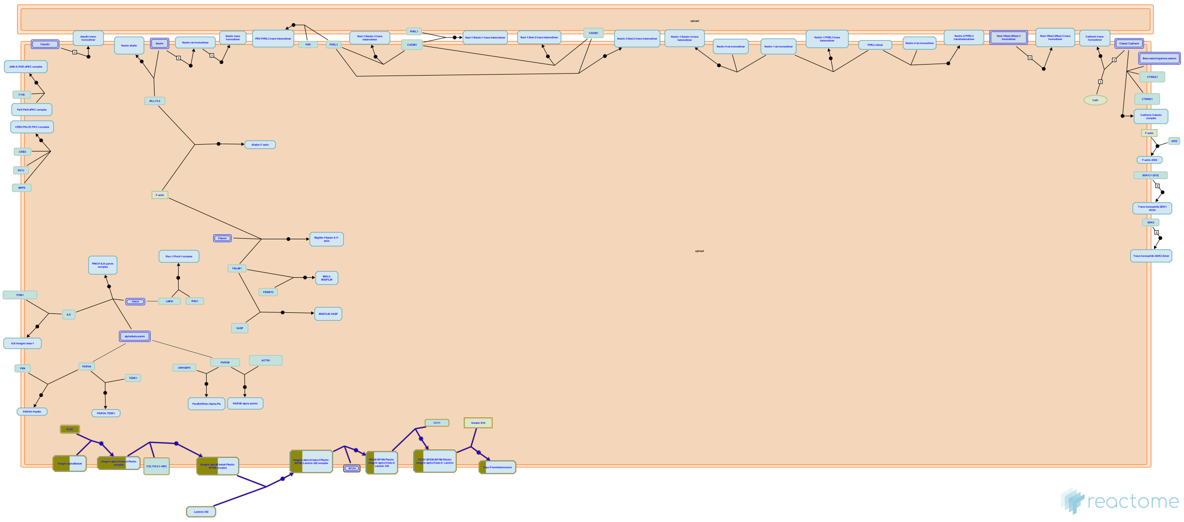

Hemidesmosomes (HDs) are specialized multiprotein junctional complexes that connect the keratin cytoskeleton of epithelial cells to the extracellular matrix and play a critical role in the maintenance of tissue structure and integrity (reviewed in Litjens et al., 2006). HDs mediate adhesion of epithelial cells to the underlying basement membrane in stratified squamous, transitional and pseudostratified epithelia (Jones et al., 1994 ; Borradori and Sonnenberg, 1996). Classical Type I HDs are found in stratified and pseudo-stratified epithelia, such as the skin, and contain a6b4, plectin, tetraspanin CD151 and the bullous pemphigoid (BP) antigens BP180 and BP230 (reviewed in Litjens et al., 2006). While HDs function in promoting stable adhesion, they are highly dynamic structures that are able to disassemble quickly, for example, during cell division, differentiation, or migration (see Margadant et al, 2008).


##### Edit history

| Date | Action | Author |
|---|---|---|
| 2009-11-04 | Edited | Matthews L |
| 2009-11-04 | Authored | Matthews L |
| 2009-11-09 | Created | Matthews L |
| 2009-11-15 | Reviewed | Sonnenberg A |
| 2021-05-22 | Modified | Shorser S |

##### Entities found in this pathway (2)

| Input | UniProt Id | Input | UniProt Id |
|---|---|---|---|
| ITGB4 | P16144 | PLEC | Q15149 |

#### 6. RUNX1 regulates transcription of genes involved in differentiation of myeloid cells (R-HSA-8939246)

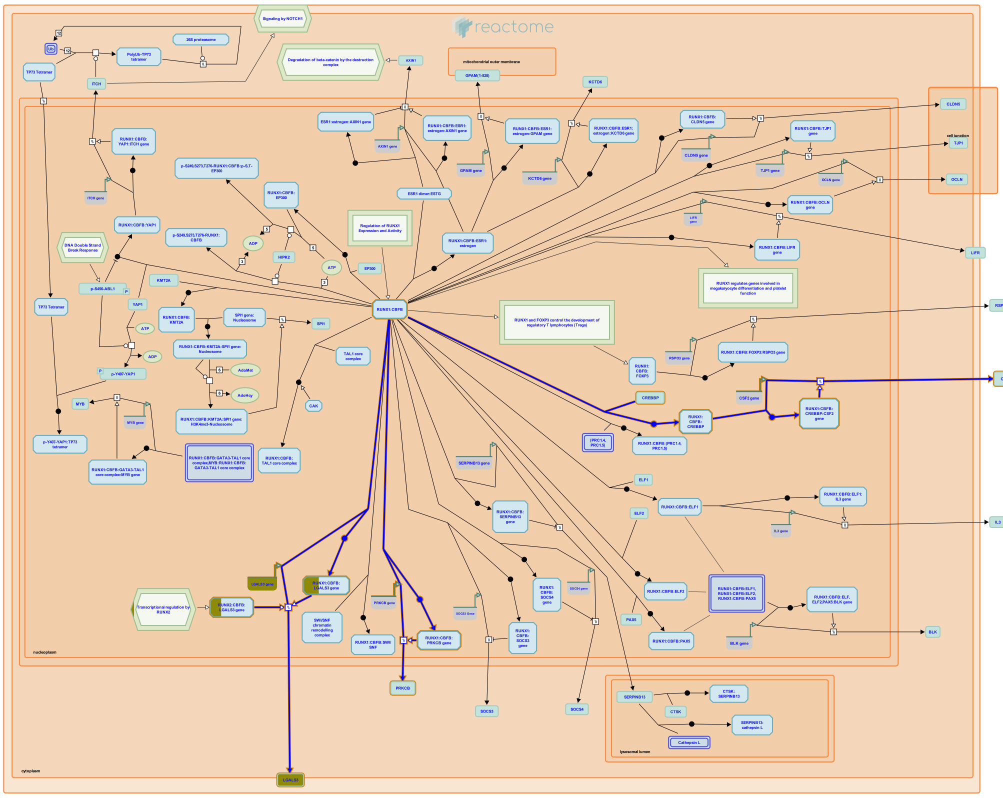

The RUNX1:CBFB complex regulates expression of genes involved in differentiation of myeloid progenitors which can commit to hematopoietic lineages that lead to generation of platelets, erythrocytes, leukocytes or monocytes.

The RUNX1:CBFB complex recruits histone acetyltransferase CREBBP (CBP) to the promoter of the CSF2 gene, encoding Granulocyte-macrophage colony stimulating factor (GM-CSF), thus inducing GM-CSF expression (Oakford et al. 2010). GM-CSF induces growth, differentiation and survival of macrophages, granulocytes, erythrocytes and megakaryocytes from myeloid progenitors (Barreda et al. 2004).

The RUNX1:CBFB complex directly stimulates transcription of the LGALS3 gene, encoding galectin-3 (Zhang et al. 2009). Galectin-3 is expressed in myeloid progenitors and its levels increase during the maturation process (Le Marer 2000).

The PRKCB gene, encoding protein kinase C-beta, which regulates apoptosis of myeloid cells, is directly transactivated by the RUNX1:CBFB complex (Hu et al. 2004).

#### Edit history

| Date | Action | Author |
|---|---|---|
| 2016-09-14 | Authored | Orlic-Milacic M |
| 2016-09-16 | Created | Orlic-Milacic M |
| 2016-12-20 | Reviewed | Ito Y, Chuang LS |
| 2017-05-09 | Edited | Orlic-Milacic M |
| 2021-05-22 | Modified | Shorser S |

#### Entities found in this pathway (1)

| Input | UniProt Id |
|---|---|
| LGALS3 | P17931 |

| Input | Ensembl Id |
|---|---|
| LGALS3 | ENSG00000131981 |

#### 7. Metal sequestration by antimicrobial proteins (R-HSA-6799990)

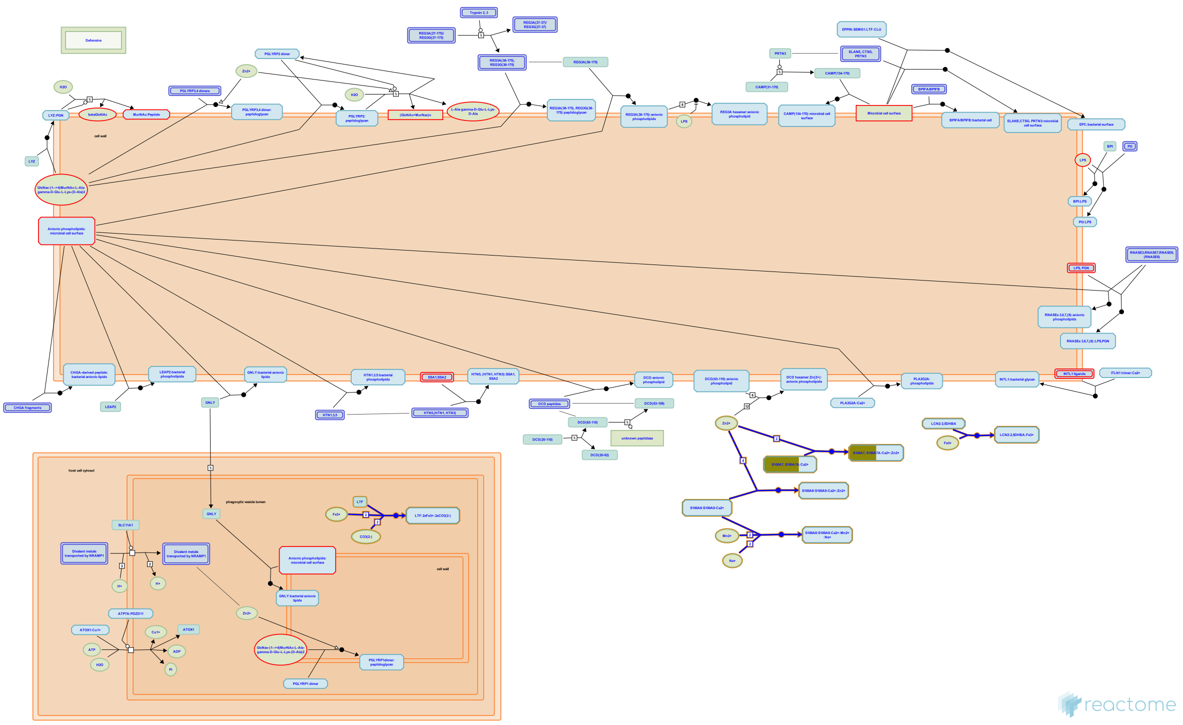

Metals are necessary for all forms of life including microorganisms, evidenced by the fact that metal cations are constituents of approximately 40% of all proteins crystallized to date (Waldron KJ et al. 2009; Foster AW et al. 2014; Guengerich FP 2014, 2015). The ability of microorganisms to maintain the intracellular metal quota is essential and allows microorganisms to adapt to a variety of environments. Accordingly, the ability of the host to control metal quota at inflammation sites can influence host-pathogen interactions. The host may restrict microbial growth either by excluding essential metals from the microbes, by delivery of excess metals to cause toxicity, or by complexing metals in microorganisms (Becker KW & Skaar EP 2014).


##### Edit history

| Date | Action | Author |
|---|---|---|
| 2015-09-26 | Created | Shamovsky V |
| 2015-10-05 | Authored | Shamovsky V |
| 2016-04-15 | Reviewed | Jupe S |
| 2016-08-02 | Reviewed | Hains DS |
| 2016-08-15 | Edited | Shamovsky V |
| 2021-05-22 | Modified | Shorser S |

##### Entities found in this pathway (1)

| Input | UniProt Id |
| --- | --- |
| S100A1 | P31151, Q86SG5 |

#### 8. Defective SLC40A1 causes hemochromatosis 4 (HFE4) (macrophages) (R-HSA-5619049)

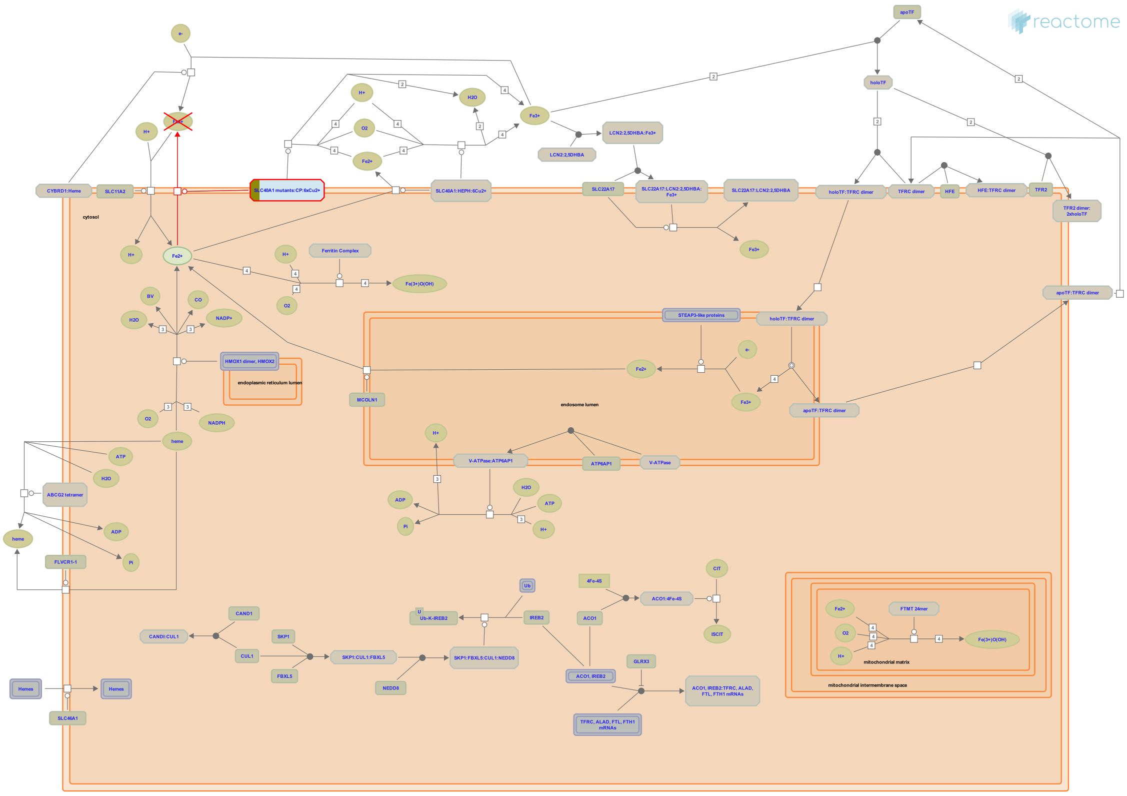

**Diseases:** hemochromatosis.

SLC40A1 (MTP1 aka ferroportin or IREG1) is highly expressed on macrophages where it mediates iron efflux from the breakdown of haem. SLC40A1 colocalises with ceruloplasmin (CP) which stablizes SLC40A1 and is necessary for the efflux reaction to occur. Six copper ions are required by ceruloplasmin as a cofactor.

Defects in SLC40A1 can cause hemochromatosis 4 (HFE4; MIM:606069), a disorder of iron metabolism characterised by iron overload. Excess iron is deposited in a variety of organs leading to their failure, resulting in serious illnesses including cirrhosis, hepatomas, diabetes, cardiomyopathy, arthritis and hypogonadotropic hypogonadism. Severe effects of the disease don't usually appear until after decades of progressive iron overloading (De Domenico et al. 2005, 2006, 2011, Kaplan et al. 2011).

#### Edit history

| Date | Action | Author |
|---|---|---|
| 2014-08-22 | Edited | Jassal B |
| 2014-08-22 | Authored | Jassal B |
| 2014-08-22 | Created | Jassal B |
| 2015-08-04 | Modified | Jassal B |
| 2015-08-04 | Reviewed | Broer S |

#### Entities found in this pathway (1)

| Input | UniProt Id |
|---|---|
| CP | P00450 |

#### 9. Sensory processing of sound by outer hair cells of the cochlea (R-HSA-9662361)

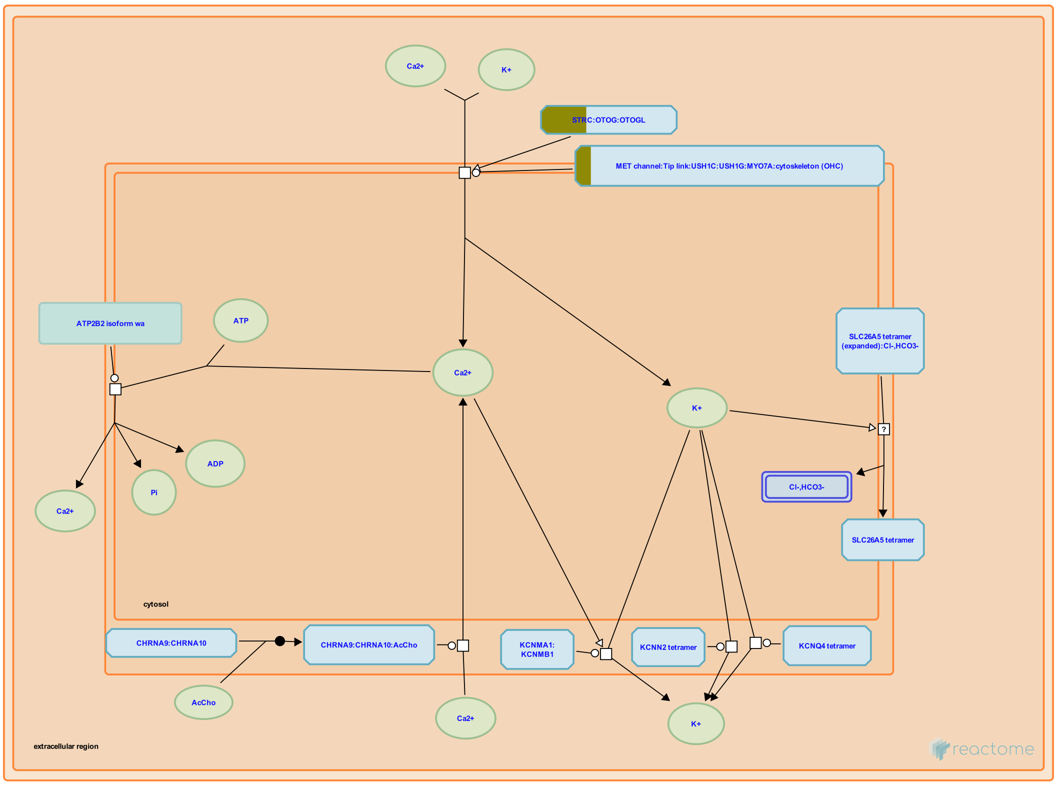


Outer hair cells (OHCs) produce amplification of sound waves in the cochlea by shortening and lengthening in response to sound, a phenomenon called electromotility (reviewed in Kim and Fettiplace 2014, Fettiplace 2016, Fettiplace 2017, Fritzsch et al. 2017, Ashmore 2019). Like inner hair cells, OHCs possess apical stereocilia arranged in rows of ascending height. A taller stereocilium is connected to a shorter stereocilium by a tip link comprising a CDH23 dimer onthe side of the taller stereocilium and a PCDH15 dimer on theapex of the shorter stereocilium. PCDH15 interacts with LHFPL5, a subunit of the mechanoelectrical transduction channel complex (MET channel, also called the mechanotransduction channel), which contains TMC1 or TMC2, TMIE, CIB2, and LHFPL5 (reviewed in Fettiplace 2016). Deflection of the stereocilia in one direction produces tension on the tip link that increases the open probability of the MET channel, resulting in depolarization of the OHC. Deflection of the stereocilia in the opposite direction produces compression on the tip link that decreases the the open probability of the MET channel, resulting in hyperpolarization of the OHC.

Sound causes micromechanical motions of the organ of Corti that result in alternating tension and compression in the tip link that produce excitatory-inhibitory cycles of MET channel openings and closings relative to the MET channel's resting open probability. This causes directionally alternating fluxes of K+ and Ca2+, yielding depolarization-hyperpolarization cycles that cause conformational changes in prestin (SLC26A5). These cycles are asymmetrical, with contraction caused by depolarization dominating elongation caused by hyperpolarization due to the asymmetry of the open probability of MET channels. Stereociliary ATP2B2 (PMCA2) extrudes calcium ions and basally located KCNQ4 extrudes potassium ions to repolarize the OHC.

Depolarization of the OHC causes a decrease in length of the OHC due to a very rapid, voltage-sensitive change in conformation of the membrane protein prestin (SLC26A5), an unusual member of the anion transporter family located in the lateral membrane (Mahendrasingam et al, 2010) that appears to respond to cytosolic chloride by altering its conformation in the plane of the plasma membrane (reviewed in Dallos et al. 2006, Dallos 2008, Hudspeth 2014, Reichenbach and Hudspeth 2014, Ashmore 2019, Santos-Sacchi 2019). Prestin also appears to act as a weak chloride-bicarbonate antiporter (Mistrik et al. 2012). Changes in length of the OHCs cause movement of the reticular lamina toward and away from the basilar membrane.


#### Edit history

| Date | Action | Author |
|---|---|---|
| 2019-09-23 | Edited | May B |
| 2019-09-23 | Authored | May B |
| 2019-09-23 | Created | May B |
| 2020-09-14 | Reviewed | Furness DN, Dallos P |
| 2020-12-12 | Modified | May B |

#### Entities found in this pathway (2)

| Input | UniProt Id | Input | UniProt Id |
|---|---|---|---|
| GSN | P06396 | OTOGL | Q3ZCN5 |

#### 10. Apoptosis (R-HSA-109581)

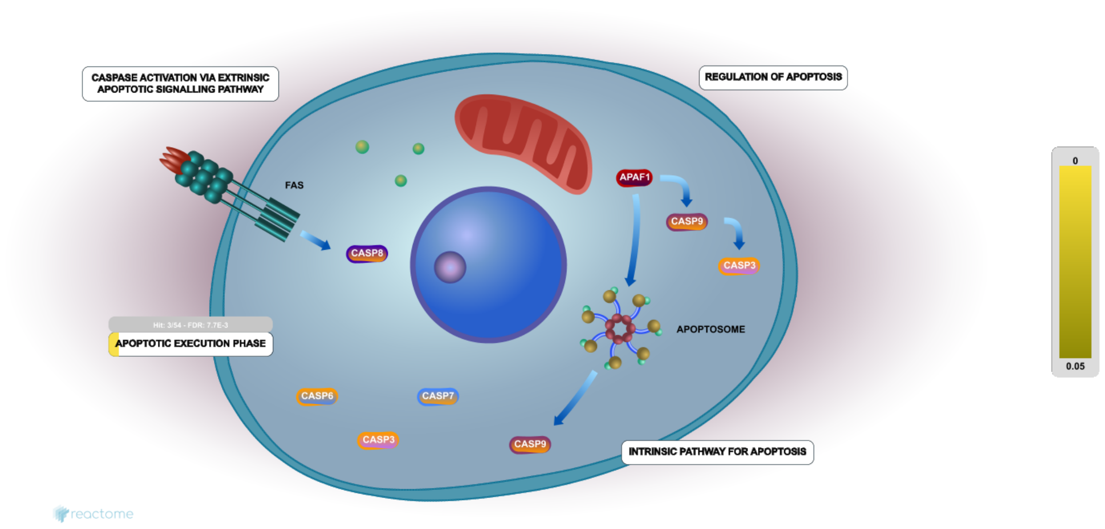


Apoptosis is a distinct form of cell death that is functionally and morphologically different from necrosis. Nuclear chromatin condensation, cytoplasmic shrinking, dilated endoplasmic reticulum, and membrane blebbing characterize apoptosis in general. Mitochondria remain morphologically unchanged. In 1972 Kerr et al introduced the concept of apoptosis as a distinct form of "cell-death", and the mechanisms of various apoptotic pathways are still being revealed today.

The two principal pathways of apoptosis are (1) the Bcl-2 inhibitable or intrinsic pathway induced by various forms of stress like intracellular damage, developmental cues, and external stimuli and (2) the caspase 8/10 dependent or extrinsic pathway initiated by the engagement of death receptors

The caspase 8/10 dependent or extrinsic pathway is a death receptor mediated mechanism that results in the activation of caspase-8 and caspase-10. Activation of death receptors like Fas/CD95, TNFR1, and the TRAIL receptor is promoted by the TNF family of ligands including FASL (APO1L OR CD95L), TNF, LT-alpha, LT-beta, CD40L, LIGHT, RANKL, BLYS/BAFF, and APO2L/TRAIL. These ligands are released in response to microbial infection, or as part of the cellular, humoral immunity responses during the formation of lymphoid organs, activation of dendritic cells, stimulation or survival of T, B, and natural killer (NK) cells, cytotoxic response to viral infection or oncogenic transformation.

The Bcl-2 inhibitable or intrinsic pathway of apoptosis is a stress-inducible process, and acts through the activation of caspase-9 via Apaf-1 and cytochrome c. The rupture of the mitochondrial membrane, a rapid process involving some of the Bcl-2 family proteins, releases these molecules into the cytoplasm. Examples of cellular processes that may induce the intrinsic pathway in response to various damage signals include: auto reactivity in lymphocytes, cytokine deprivation, calcium flux or cellular damage by cytotoxic drugs like taxol, deprivation of nutrients like glucose and growth factors like EGF, anoikis, transactivation of target genes by tumor suppressors including p53.

In many non-immune cells, death signals initiated by the extrinsic pathway are amplified by connections to the intrinsic pathway. The connecting link appears to be the truncated BID (tBID) protein a proteolytic cleavage product mediated by caspase-8 or other enzymes.


#### Edit history

| Date | Action | Author |
|---|---|---|
| 2004-01-16 | Authored | Tsujimoto Y, Hengartner M, Hardwick JM, Tschopp J, Alnemri E |
| 2004-01-16 | Created | Tsujimoto Y, Hengartner M, Hardwick JM, Tschopp J, Alnemri E |
| 2013-11-25 | Edited | Joshi-Tope G, Matthews L, Gopinathrao G, Gillespie ME |
| 2021-05-18 | Reviewed | Ranganathan S, Hengartner M, Vaux DL |
| 2021-05-22 | Modified | Shorser S |

#### Entities found in this pathway (3)

| Input | UniProt Id | Input | UniProt Id | Input | UniProt Id |
|---|---|---|---|---|---|
| GSN | P06396 | PLEC | Q15149 | VIM | P08670 |

### 11. Assembly of collagen fibrils and other multimeric structures (R-HSA-2022090 )

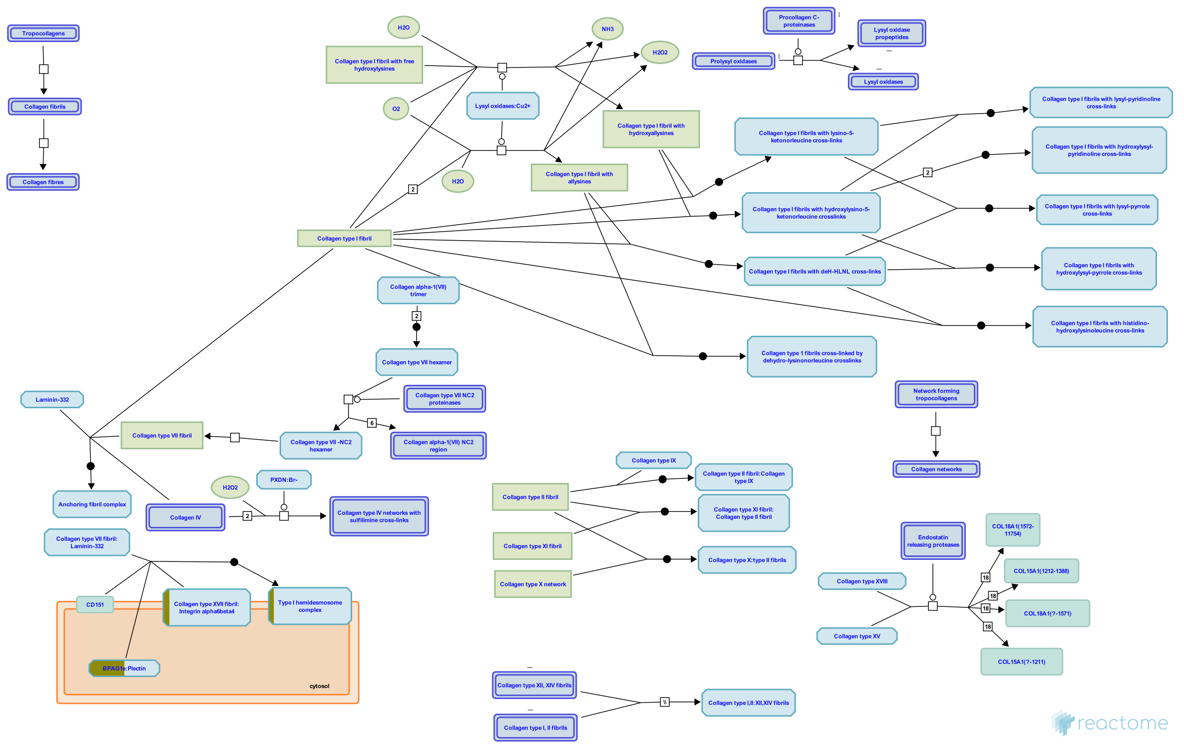


Collagen trimers in triple-helical form, referred to as procollagen or collagen molecules, are exported from the ER and trafficked through the Golgi network before secretion into the extracellular space. For fibrillar collagens namely types I, II, III, V, XI, XXIV and XXVII (Gordon & Hahn 2010, Ricard-Blum 2011) secretion is concomitant with processing of the N and C terminal collagen propeptides. These processed molecules are known as tropocollagens, considered to be the units of higher order collagen structures. They form within the extracellular space via a process that can proceed spontaneously, but in the cellular environment is regulated by many collagen binding proteins such as the FACIT (Fibril Associated Collagens with Interrupted Triple helices) family collagens and Small Leucine-Rich Proteoglycans (SLRPs). The architecture formed ultimately depends on the collagen subtype and the cellular conditions. Structures include the well-known fibrils and fibres formed by the major structural collagens type I and II plus several different types of supramolecular assembly (Bruckner 2010). The mechanical and physical properties of tissues depend on the spatial arrangement and composition of these collagen-containing structures (Kadler et al. 1996, Shoulders & Raines 2009, Birk & Bruckner 2011).

#### Entities found in this pathway (2)

| Input | UniProt Id | Input | UniProt Id |
|---|---|---|---|
| ITGB4 | P16144 | PLEC | Q15149 |

#### 12. Defective CP causes aceruloplasminemia (ACERULOP) (R-HSA-5619060)

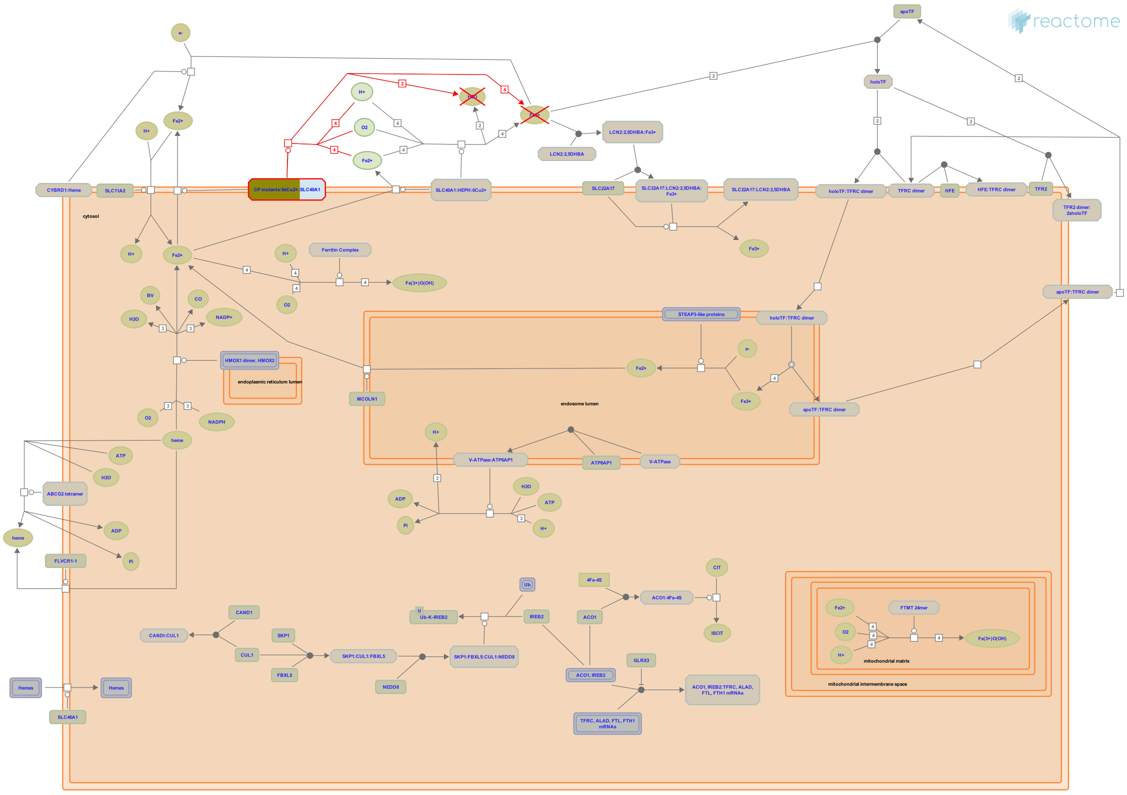

**Diseases:** aceruloplasminemia.

Ceruloplasmin (CP), synthesised in the liver and secreted into plasma, is a copper-binding (6-7 atoms per molecule) glycoprotein involved in iron trafficking in vertebrates. CP is essential for SLC40A1 (ferroportin) stability at the cell surface, the protein that mediates iron efflux from cells. CP also possesses ferroxidase activity, which oxidises ferrous iron (Fe2+) to ferric iron (Fe3+) following its transfer out of the cell. Fe3+ can then be loaded on to extracellular transferrin which transports it around the body to sites where it is required. Iron is vital for many metabolic processes such as electron transport and the transport and storage of oxygen.

Defects in CP (or indeed SLC40A1) can lead to the phenotype of iron overload as seen in the disorder aceruloplasminemia (ACERULOP; MIM:604290). It is a rare autosomal recessive disorder of iron metabolism characterised by iron accumulation mainly in the brain, but also in liver, pancreas and retina. Patients develop retinal degeneration, diabetes mellitus and neurological disturbance. ACERULOP belongs to a group of disorders known as NBIA (neurodegeneration with brain iron accumulation), distinguishing it from hereditary hemochromatosis (serum iron is high but the brain is usually not affected) and from disorders of copper metabolism such as Menkes and Wilson disease (Harris et al. 1995, Kono 2012, Musci et al. 2014).

#### Edit history

| Date | Action | Author |
|---|---|---|
| 2014-08-22 | Edited | Jassal B |
| 2014-08-22 | Authored | Jassal B |
| 2014-08-22 | Created | Jassal B |
| 2015-08-04 | Modified | Jassal B |
| 2015-08-04 | Reviewed | Broer S |

#### Entities found in this pathway (1)

| Input | UniProt Id |
|---|---|
| CP | P00450 |

#### 13. Sensory processing of sound (R-HSA-9659379)

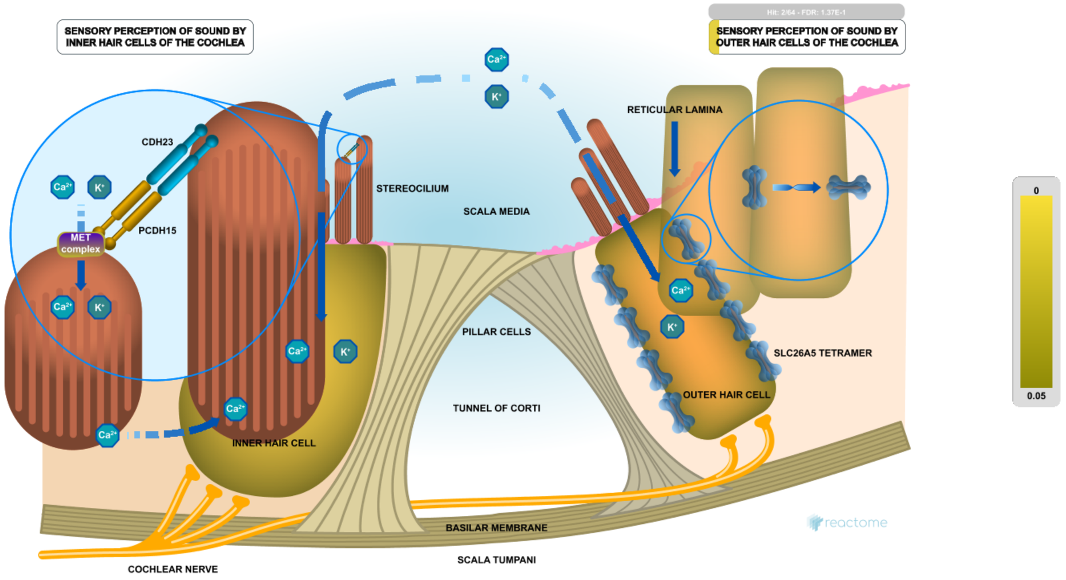


In mammals, sounds are processed in the cochlea, a spiral-shaped organ in the inner ear (reviewed in Basch et al. 2016, Fettiplace 2017, Koppl and Manley 2019). Low frequency sounds are sensed at the distal end (apex) of the cochlea; high frequency sounds are sensed at the proximal end (base) of the cochlea (reviewed in Dallos 1992, Manley 2018). Sound vibrations are transmitted from the eardrum through the three bones of the inner ear (malleus, incus, stapes) and the oval window of the cochlea to the fluids within the cochlea. Within the organ of Corti in the cochlea there are 3 rows of outer hair cells (OHCs) on the external side of the tunnel of Corti and 1 row of inner hair cells (IHCs) on the internal side (Spoendlin 1967). Each IHC synapses with approximately 20 afferent myelinated type I spiral ganglion neurons and functions as a sensory receptor to convert the energy of sound waves to secretion of glutamate neurotransmitter. Multiple OHCs synapse with each unmyelinated type II afferent neuron and OHCs are also synapsed with efferent medial olivocochlear fibers (Spoendlin 1967). The primary function of OHCs, however, is amplification of organ of Corti motions in response to sound (Ryan and Dallos 1975). Amplification is produced by changes in receptor-potential driven cell length caused by changes in the conformation of the unusual membrane protein prestin (SLC26A5, Zheng et al. 2000).

IHCs and OHCs sense the sonic vibrations by deflection of stereocilia on their apical surfaces (reviewed in Fettiplace et al. 2017, McPherson 2018). The stereocilia are arranged in rows of increasing height, with a stereocilium of one row connected to a stereocilium of another row by a tip link composed of a CDH23 dimer on the taller stereocilium joined at its N-termini to the N-termini of a PCDH15 dimer on the shorter stereocilium. CDH23 is connected to the cytoskeleton of the taller stereocilium via MYO7A (MyoVIIa), USH1C (Harmonin), and USH1G (Sans) (reviewed in Peng et al. 2011, Cosgrove and Zallocchi 2014, Barr-Gillespie 2015, Fettiplace 2017, McGrath et al. 2017, Cunningham and Müller 2019, Ó Maoiléidigh and Ricci 2019, Velez-Ortega and Frolenkov 2019) while PCDH15 on the shorter stereocilium interacts with LHFPL5, an auxiliary subunit of the mechanoelectrical transduction channel (MET channel, also known as the mechanotransduction channel), which contains at least TMC1 or TMC2, TMIE, and the auxiliary subunits LHFPL5 and CIB2 (reviewed in Fettiplace 2016, Qiu and Müller 2018, Corey et al. 2019). Deflection of stereocilia in the direction that increases tension on the tip link causes depolarization of the cell by increasing the open probability of the MET channel, which then transports calcium and potassium into the hair cell according to the gradient of those ions between the scala media (containing endolymph at 154 mM K+ and <1 mM Ca2+) at the apex of the cell and the scala tympani (containing perilymph at 7 mM K+) at the base (reviewed in Fettiplace and Kim 2014). Similarly, compression of the tip link by deflection of the stereocilia in the opposite direction decreases the open probability of the MET channel and causes hyperpolarization of the cell.

Depolarization of IHCs causes opening of voltage-gated calcium channels arrayed in stripes on the basolateral membrane close to ribbon synapses formed between the IHC and the afferent fiber of a myelinated type I spiral ganglion neuron. This results in a localized increase in cytosolic calcium ions which interact with Otoferlin (OTOF) on glutamate-containing synaptic vesicles at the ribbon structure to activate exocytosis of glutamate into the synapse formed with the afferent neuron (reviewed in Wichmann 2015, Pangrsic and Vogl 2018). Ribbon synapses are distinguished by electron-dense ribbon structures projecting from the presynaptic membrane into the cytosol and comprising at least BASSOON, RIBEYE (an isoform of CTBP2), and PICCOLINO (an isoform of PICCOLO). The ribbon structures appear to transiently bind synaptic vesicles and facilitate resupply of synaptic vesicles at active zones to refill the pool of readily releasable vesicles (reviewed in Moser et al. 2006, Moser et al. 2020).

In contrast with IHCs, OHCs mainly function in sound amplification by decreasing up to about 4% in length in response to depolarization caused by opening of the MET channel and increasing in length in response to hyperpolarization caused by channel closing, resulting in alternating compression and decompression between the reticular lamina and the basilar membrane. The changes in the length of the OHC are caused by very rapid (microseconds), voltage-sensitive changes in the conformation of the membrane protein prestin (SLC26A5). Stereociliary ATP2B2 (PMCA2) extrudes calcium ions and basally located KCNQ4 extrudes potassium ions to repolarize the OHC.

OHCs are synapsed with efferent cholinergic medial olivocochlear fibers (reviewed in Fritzsch and Elliott 2017, Fuchs and Lauer 2019). Acetylcholine released at the synapse binds an unusual, nicotine-antagonized, nicotinic receptor comprising CHRNA9 and CHRNA10. Upon binding acetylcholine, CHRNA9:CHRNA10 transports calcium ions into the OHC. The calcium activates SK2 potassium channels (KCNN2) and BK potassium channels (KCNMA1:KCNMB1) which extrude potassium ions, hyperpolarize the OHC, and inhibit activation of the OHC.

Loud sounds can cause a temporary threshold shift (temporary loss of hearing) caused by damage to stereocilia and synapses or permanent threshold shift (permanent loss of hearing) caused by damage or death of hair cells and neurons (reviewed in Kurabi et al. 2017).


##### Edit history

| Date | Action | Author |
|---|---|---|
| 2019-08-27 | Edited | May B |
| 2019-08-27 | Authored | May B |
| 2019-08-27 | Created | May B |
| 2020-09-14 | Reviewed | Furness DN |
| 2021-03-04 | Modified | Shorser S |

##### Entities found in this pathway (2)

| Input | UniProt Id | Input | UniProt Id |
|---|---|---|---|
| GSN | P06396 | OTOGL | Q3ZCN5 |

#### 14. Programmed Cell Death (R-HSA-5357801)

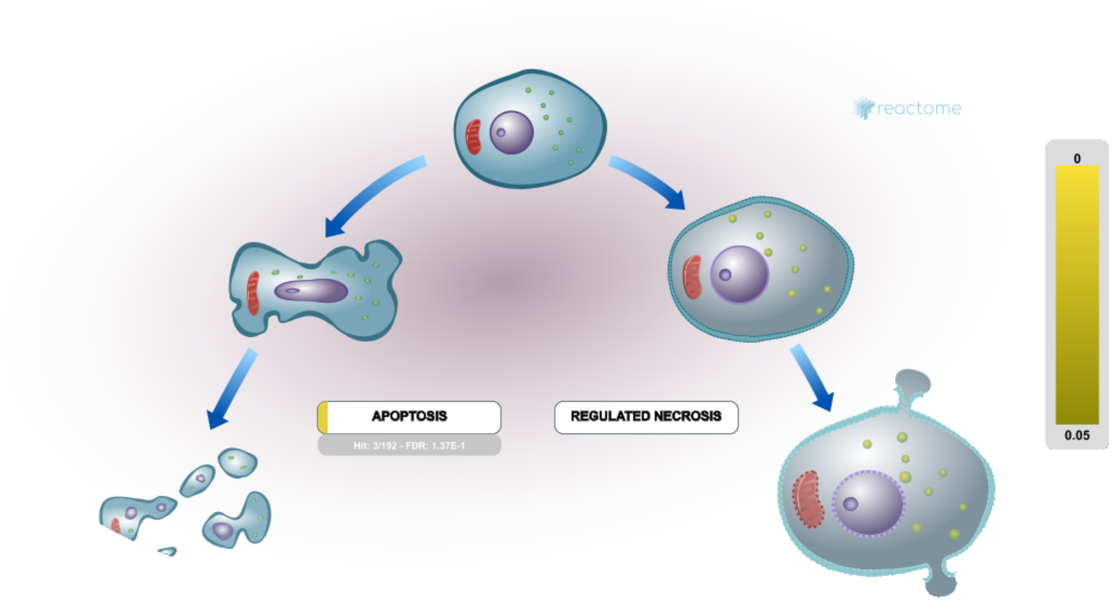


Cell death is a fundamental cellular response that has a crucial role in shaping our bodies during development and in regulating tissue homeostasis by eliminating unwanted cells. There are a number of different forms of cell death, each with a corresponding number of complex subprocesses. The first form of regulated or programmed cell death to be characterized was apoptosis. Evidence has emerged for a number of regulated non-apoptotic cell death pathways, including some with morphological features that were previously attributed to necrosis. More recently necrosis has been subdivided into parts including programmed necrotic cell death processes, such as RIP1-mediated regulated necrosis or pyroptosis.

Reactome currently represents programmed cell death using the model of extrinsic signalling that leads to a molecular decision point pivoting on caspase-8 activation or inhibition. Caspase-8 activation tilts the cell towards apoptosis, while caspase-8 inhibition tilts the cell towards Regulated Necrosis.

The terminology and molecular definitions of cell death-related events annotated here are consistent with the 2015 recommendations of the Nomenclature Committee on Cell Death (NCCD) (Galluzzi L et al. 2015).


##### Edit history

| Date | Action | Author |
|---|---|---|
| 2014-03-26 | Created | Shamovsky V |
| 2014-11-18 | Edited | Shamovsky V |
| 2014-11-18 | Authored | Shamovsky V |

| Date | Action | Author |
|---|---|---|
| 2021-05-22 | Modified | Shorser S |

#### Entities found in this pathway (3)

| Input | UniProt Id | Input | UniProt Id | Input | UniProt Id |
|---|---|---|---|---|---|
| GSN | P06396 | PLEC | Q15149 | VIM | P08670 |

#### 15. Sodium-coupled sulphate, di- and tri-carboxylate transporters (R-HSA-433137 )

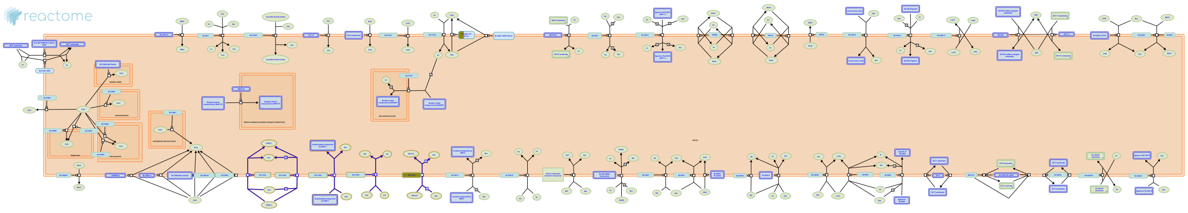


Five human SLC13 genes encode sodium-coupled sulphate, di- and tri-carboxylate transporters located on the plasma membrane. Two transporters (NaS1 and NaS2) co-transport sulphate with sodium. The other members (NaDC1, NaDC3, and NaCT) co-transport sodium with di- and tri-carboxylates such as succinate, citrate and alpha-ketoglutarate (Pajor AM, 2006).

##### Edit history

| Date | Action | Author |
|---|---|---|
| 2009-08-21 | Edited | Jassal B |
| 2009-08-21 | Authored | Jassal B |
| 2009-08-21 | Created | Jassal B |
| 2009-11-12 | Reviewed | He L |
| 2021-05-22 | Modified | Shorser S |

##### Entities found in this pathway (1)

| Input | UniProt Id |
|---|---|
| SLC13A3 | Q8WWT9 |

#### 16. Cell junction organization (R-HSA-446728)

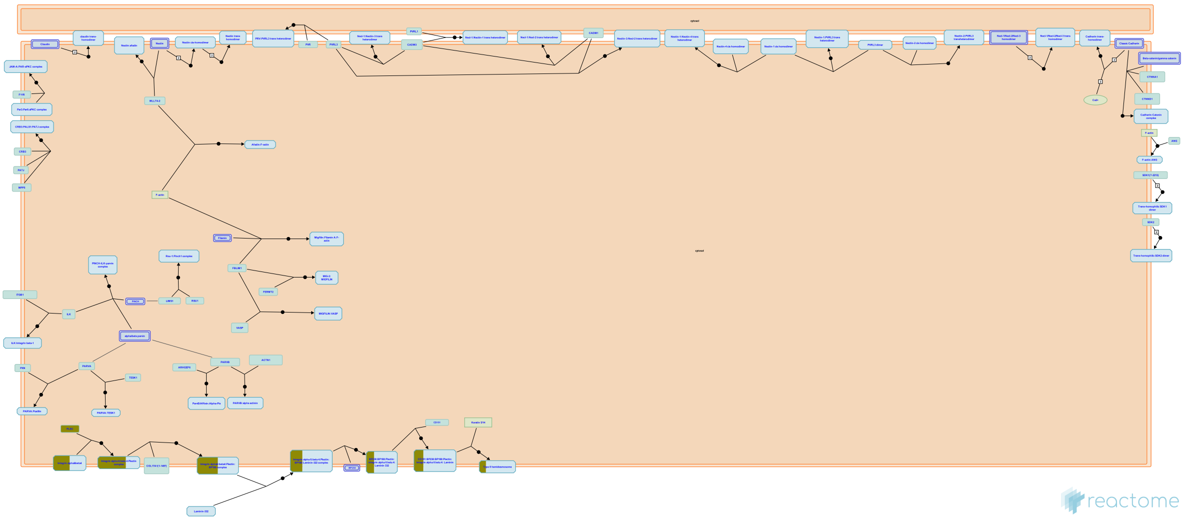

Cell junction organization in Reactome currently covers aspects of cell-cell junction organization, cell-extracellular matrix interactions, and Type I hemidesmosome assembly.

##### References

##### Edit history

| Date | Action | Author |
|---|---|---|
| 2009-11-17 | Edited | Matthews L |
| 2009-11-17 | Created | Matthews L |
| 2021-05-22 | Modified | Shorser S |

##### Entities found in this pathway (2)

| Input | UniProt Id | Input | UniProt Id |
|---|---|---|---|
| ITGB4 | P16144 | PLEC | Q15149 |

### 17. Collagen formation (R-HSA-1474290)

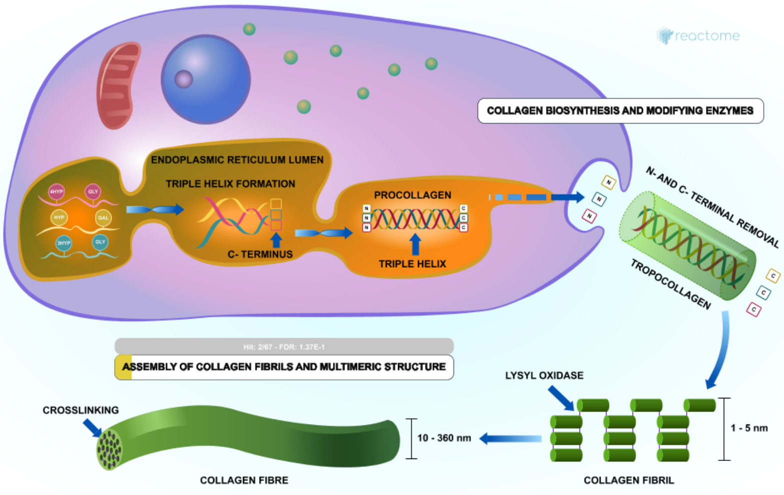


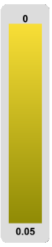

Collagen is a family of at least 29 structural proteins derived from over 40 human genes (Myllyharju & Kivirikko 2004). It is the main component of connective tissue, and the most abundant protein in mammals making up about 25% to 35% of whole-body protein content. A defining feature of collagens is the formation of trimeric left-handed polyproline II-type helical collagenous regions. The packing within these regions is made possible by the presence of the smallest amino acid, glycine, at every third residue, resulting in a repeating motif Gly-X-Y where X is often proline (Pro) and Y often 4-hydroxyproline (4Hyp). Gly-Pro-Hyp is the most common triplet in collagen (Ramshaw et al. 1998). Collagen peptide chains also have non-collagenous domains, with collagen subclasses having common chain structures. Collagen fibrils are mostly found in fibrous tissues such as tendon, ligament and skin. Other forms of collagen are abundant in cornea, cartilage, bone, blood vessels, the gut, and intervertebral disc. In muscle tissue, collagen is a major component of the endomysium, constituting up to 6% of muscle mass. Gelatin, used in food and industry, is collagen that has been irreversibly hydrolyzed.

| Input | UniProt Id | Input | UniProt Id |
|---|---|---|---|
| ITGB4 | P16144 | PLEC | Q15149 |

#### 18. Calcitonin-like ligand receptors (R-HSA-419812)

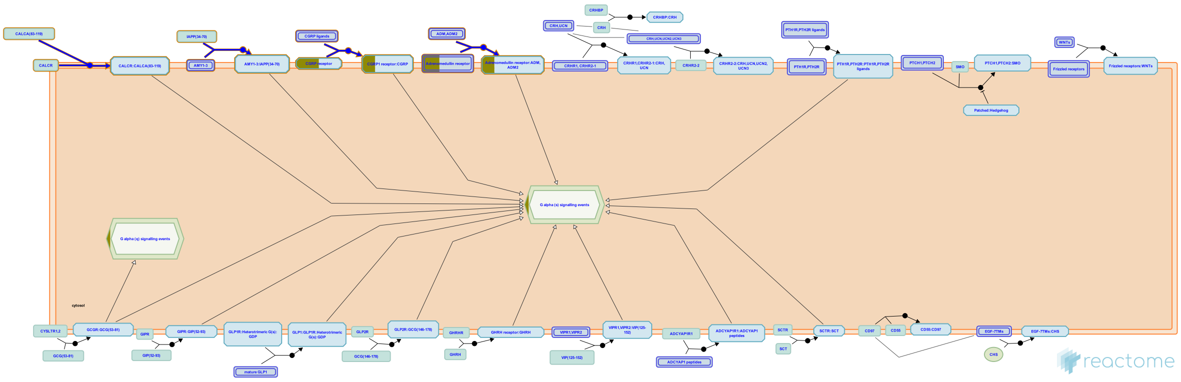

The calcitonin peptide family comprises calcitonin, amylin, calcitonin gene-related peptide (CGRP), adrenomedullin (AM) and intermedin (AM2). Calcitonin is a 32 amino acid peptide, involved in bone homeostasis (Sexton PM et al, 1999). Amylin is a product of the islet beta-cell (Cooper GJ et al, 1987), along with insulin and probably has a hormonal role in the regulation of nutrient intake (Young A and Denaro M, 1998). Adrenomedullin (AM) is a ubiquitously expressed peptide initially isolated from phaechromocytoma (a tumour of the adrenal medulla) (Kitamura K et al, 1993). Both AM and AM2 (Takei Y et al, 2004) belong to a family of calcitonin-related peptide hormones important for regulating diverse physiologic functions and the chemical composition of fluids and tissues.

The receptor family for these peptides consists of two class B GPCRs, the calcitonin receptor (CT) and calcitonin receptor-like receptor (CL) (Poyner DR er al, 2002). Whilst the receptor for calcitonin is a conventional class B GPCR, the receptors for CGRP, AM and amylin require additional proteins, called the receptor activity modifying proteins (RAMPs). There are three RAMPs in mammals; they interact with the CT receptor to convert it to receptors for amylin. For CGRP and AM, the related CL interacts with RAMP1 to give a CGRP receptor and RAMP2 or 3 to give AM receptors. CL by itself will bind no known endogenous ligand.


##### Edit history

| Date | Action | Author |
| --- | --- | --- |
| 2009-05-07 | Edited | Jassal B |
| 2009-05-07 | Authored | Jassal B |
| 2009-05-07 | Created | Jassal B |
| 2009-05-29 | Reviewed | D'Eustachio P |
| 2021-05-21 | Modified | Shorser S |

#### Entities found in this pathway (1)

| Input | UniProt Id |
| --- | --- |
| CALCRL | Q16602 |

#### 19. Post-translational protein phosphorylation (R-HSA-8957275)

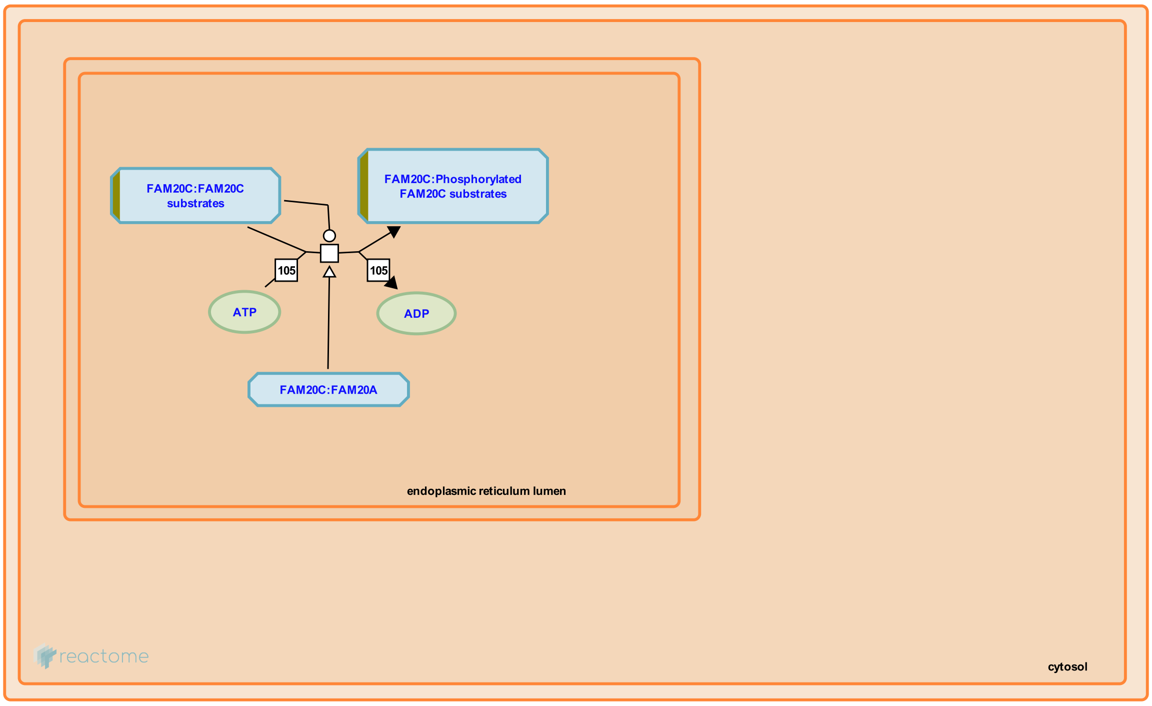


Secretory pathway kinases phosphorylate a diverse array of substrates involved in many physiological processes.


##### Edit history

| Date | Action | Author |
|---|---|---|
| 2016-12-08 | Authored | Jupe S |
| 2017-01-23 | Reviewed | Wiley SE |
| 2017-01-24 | Edited | Jupe S |
| 2017-01-24 | Created | Jupe S |
| 2021-05-31 | Modified | Shorser S |

##### Entities found in this pathway (2)

| Input | UniProt Id | Input | UniProt Id |
|---|---|---|---|
| CP | P00450 | TMEM132E | Q24JP5 |

#### 20. Arachidonate production from DAG (R-HSA-426048)

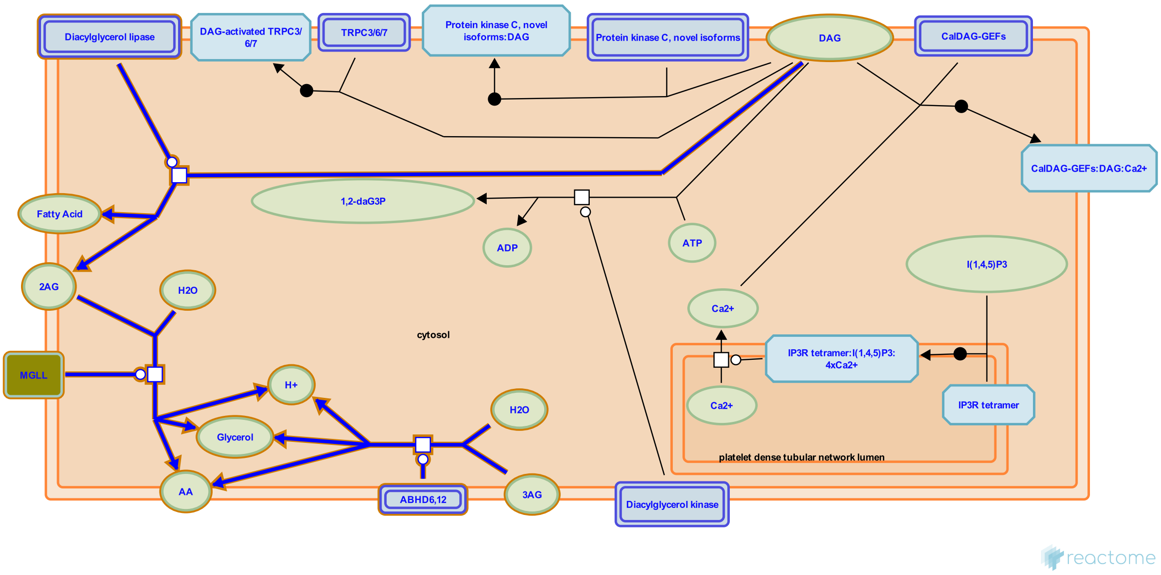


**Cellular compartments:** plasma membrane.

Diacylglycerol (DAG) is an important source of arachidonic acid, a signalling molecule and the precursor of the prostaglandins. In human platelet almost all the DAG produced from phosphatidylinositol degradation contains arachidonate (Takamura et al. 1987). DAG is hydrolysed by DAG lipase to 2-arachidonylglycerol (2-AG) which is further hydrolysed by monoacylglycerol lipase. 2-AG is an agonist of cannabinoid receptor 1.

##### Edit history

| Date | Action | Author |
|---|---|---|
| 2009-06-11 | Authored | Jupe S |
| 2009-06-11 | Created | Jupe S |
| 2009-09-04 | Reviewed | Akkerman JW |
| 2009-09-09 | Edited | Jupe S |
| 2021-05-22 | Modified | Shorser S |

##### Entities found in this pathway (1)

| Input | UniProt Id |
|---|---|
| MGLL | Q99685 |

#### 21. Antimicrobial peptides (R-HSA-6803157)

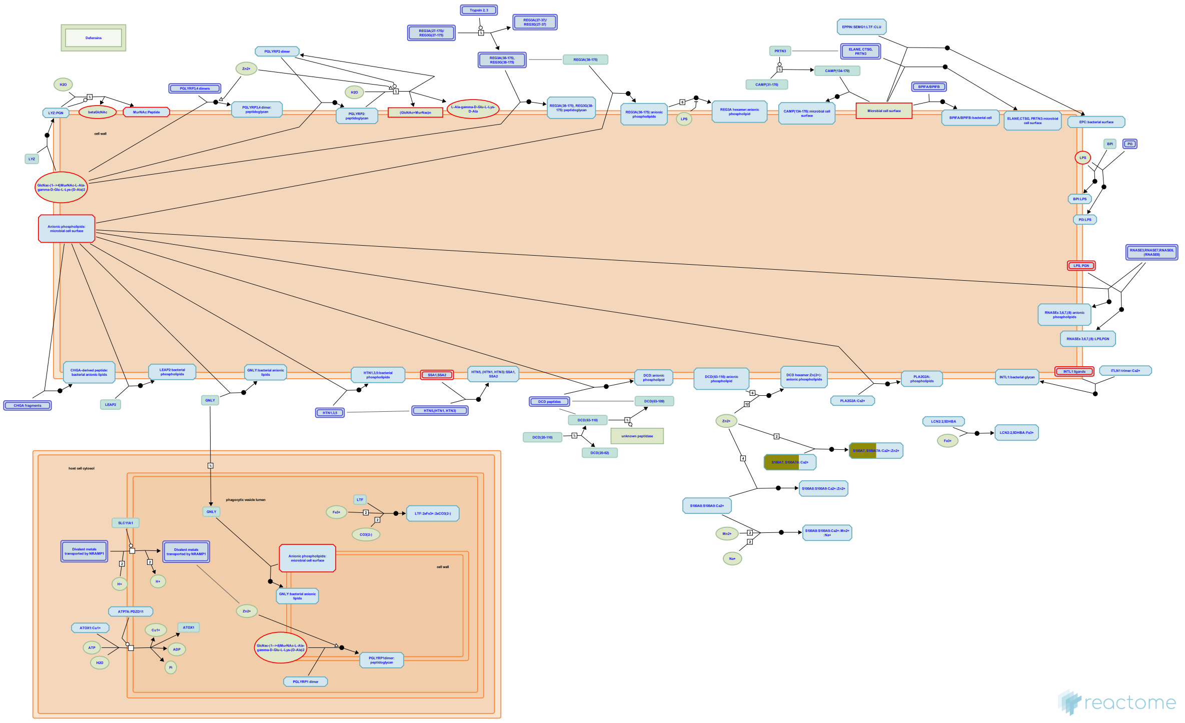

Antimicrobial peptides (AMPs) are small molecular weight proteins with broad spectrum of antimicrobial activity against bacteria, viruses, and fungi (Zasloff M 2002; Radek K & Gallo R 2007). The majority of known AMPs are cationic peptides with common structural characteristics where domains of hydrophobic and cationic amino acids are spatially arranged into an amphipathic design, which facilitates their interaction with bacterial membranes (Shai Y 2002; Yeaman MR & Yount NY 2003; Brown KL & Hancock RE 2006; Dennison SR et al. 2005; Zelezetsky I & Tossi A 2006). It is generally excepted that the electrostatic interaction facilitates the initial binding of the positively charged peptides to the negatively charged bacterial membrane. Moreover, the structural amphiphilicity of AMPs is thought to promote their integration into lipid bilayers of pathogenic cells, leading to membrane disintegration and finally to the microbial cell death. In addition to cationic AMPs a few anionic antimicrobial peptides have been found in humans, however their mechanism of action remains to be clarified (Lai Y et al. 2007; Harris F et al. 2009; Paulmann M et al. 2012). Besides the direct neutralizing effects on bacteria AMPs may modulate cells of the adaptive immunity (neutrophils, T-cells, macrophages) to control inflammation and/or to increase bacterial clearance.

AMPs have also been referred to as cationic host defense peptides, anionic antimicrobial peptides/proteins, cationic amphipathic peptides, cationic AMPs, host defense peptides and alpha-helical antimicrobial peptides (Brown KL & Hancock RE 2006; Harris F et al. 2009; Groenink J et al. 1999; Bradshaw J 2003; Riedl S et al. 2011; Huang Y et al. 2010).

The Reactome module describes the interaction events of various types of human AMPs, such as cathelicidin, histatins and neutrophil serine proteases, with conserved patterns of microbial membranes at the host-pathogen interface. The module includes also proteolytic processing events for dermcidin (DCD) and cathelicidin (CAMP) that become functional upon cleavage. In addition, the module highlights an AMP-associated ability of the host to control metal quota at inflammation sites to influence host-pathogen interactions.

#### Edit history

| Date | Action | Author |
|---|---|---|
| 2015-10-05 | Authored | Shamovsky V |
| 2015-10-05 | Created | Shamovsky V |
| 2016-04-15 | Reviewed | Jupe S |
| 2016-08-02 | Reviewed | Hains DS |
| 2016-08-15 | Edited | Shamovsky V |
| 2021-05-22 | Modified | Shorser S |

#### Entities found in this pathway (1)

| Input | UniProt Id |
|---|---|
| S100A1 | P31151, Q86SG5 |

#### 22. Regulation of Insulin-like Growth Factor (IGF) transport and uptake by Insulin-like Growth Factor Binding Proteins (IGFBPs) (R-HSA-381426)

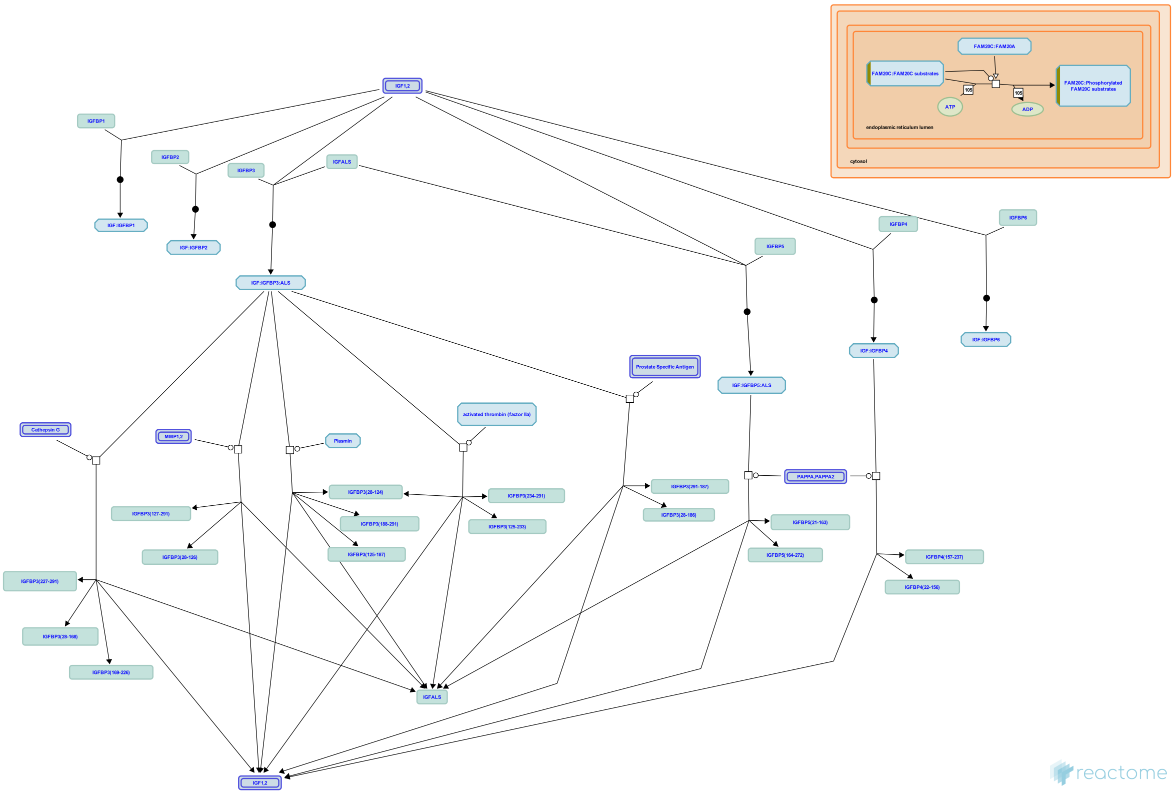


**Cellular compartments:** extracellular region.

The family of Insulin like Growth Factor Binding Proteins (IGFBPs) share 50% amino acid identity with conserved N terminal and C terminal regions responsible for binding Insulin like Growth Factors I and II (IGF I and IGF II). Most circulating IGFs are in complexes with IGFBPs, which are believed to increase the residence of IGFs in the body, modulate availability of IGFs to target receptors for IGFs, reduce insulin like effects of IGFs, and act as signaling molecules independently of IGFs.
About 75% of circulating IGFs are in 1500 220 KDa complexes with IGFBP3 and ALS. Such complexes are too large to pass the endothelial barrier. The remaining 20 25% of IGFs are bound to other IGFBPs in 40 50 KDa complexes. IGFs are released from IGF:IGFBP complexes by proteolysis of the IGFBP. IGFs become active after release, however IGFs may also have activity when still bound to some IGFBPs.
IGFBP1 is enriched in amniotic fluid and is produced in the liver under control of insulin (insulin suppresses production). IGFBP1 binding stimulates IGF function. It is unknown which if any protease degrades IGFBP1.
IGFBP2 is enriched in cerebrospinal fluid; its binding inhibits IGF function. IGFBP2 is not significantly degraded in circulation.
IGFB3, which binds most IGF in the body is enriched in follicular fluid and found in many other tissues. IGFBP 3 may be cleaved by plasmin, thrombin, Prostate specific Antigen (PSA, KLK3), Matrix Metalloprotease-1 (MMP1), and Matrix Metalloprotease-2 (MMP2). IGFBP3 also binds extracellular matrix and binding lowers its affinity for IGFs. IGFBP3 binding stimulates the effects of IGFs.
IGFBP4 acts to inhibit IGF function and is cleaved by Pregnancy associated Plasma Protein A (PAPPA) to release IGF.
IGFBP5 is enriched in bone matrix; its binding stimulates IGF function. IGFBP5 is cleaved by Pregnancy Associated Plasma Protein A2 (PAPPA2), ADAM9, complement C1s from smooth muscle, and thrombin. Only the cleavage site for PAPPA2 is known.
IGFBP6 is enriched in cerebrospinal fluid. It is unknown which if any protease degrades IGFBP6.


#### Edit history

| Date | Action | Author |
|---|---|---|
| 2008-11-20 | Edited | May B, Gopinathrao G |
| 2008-11-20 | Created | May B |
| 2008-12-02 | Reviewed | Matthews L, D'Eustachio P, Gillespie ME |

| Date | Action | Author |
|---|---|---|
| 2021-05-22 | Modified | Shorser S |

#### Entities found in this pathway (2)

| Input | UniProt Id | Input | UniProt Id |
|---|---|---|---|
| CP | P00450 | TMEM132E | Q24JP5 |

#### 23. Advanced glycosylation endproduct receptor signaling (R-HSA-879415)

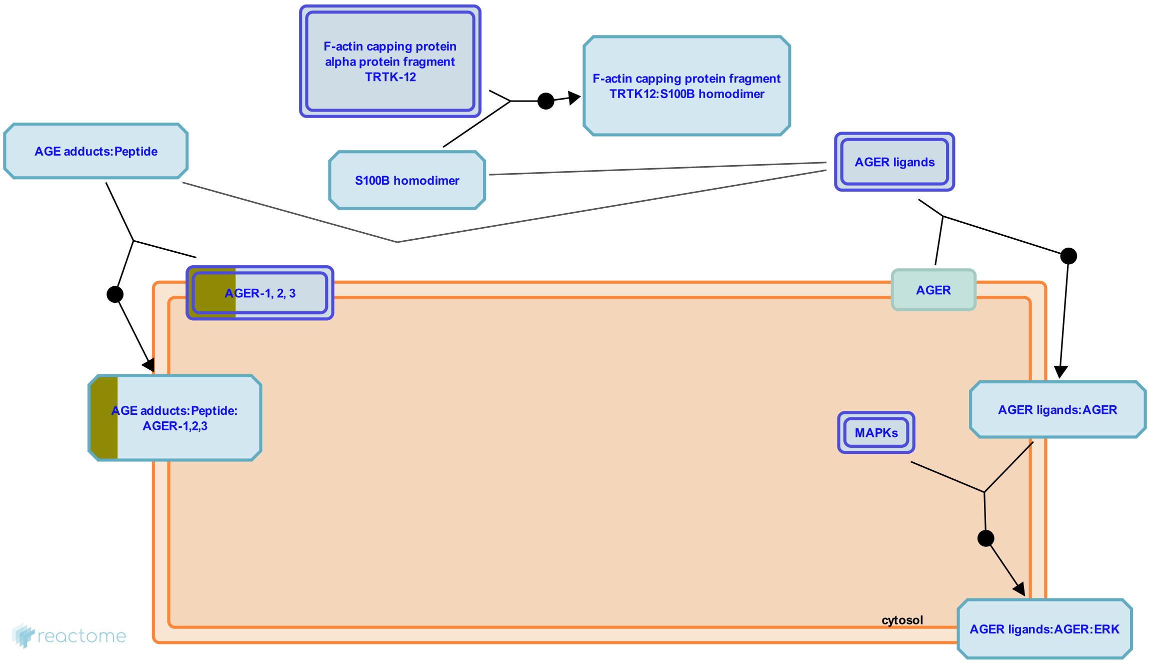


**Cellular compartments:** plasma membrane, extracellular region.

Advanced Glycosylation End- product-specific Receptor (AGER) also known as Receptor for Advanced Glycation End-products (RAGE) is a multi-ligand membrane receptor belonging to the immunoglobulin superfamily. It is considered to be a Pattern Recognition Receptor (Liliensiek et al. 2004). It recognizes a large variety of modified proteins known as advanced glycation/glycosylation endproducts (AGEs), a heterogenous group of structures that are generated by the Maillard reaction, a consequence of long-term incubation of proteins with glucose (Ikeda et al. 1996). Their accumulation is associated with diabetes, atherosclerosis, renal failure and ageing (Schmidt et al. 1999). The most prevalent class of AGE in vivo are N(6)-carboxymethyllysine (NECML) adducts (Kislinger et al. 1991). In addition to AGEs, AGER is a signal transduction receptor for amyloid-beta peptide (Ab) (Yan et al. 1996), mediating Ab neurotoxicity and promoting Ab influx into the brain. AGER also responds to the proinflammatory S100/calgranulins (Hofmann et al. 1999) and High mobility group protein B1 (HMGB1/Amphoterin/DEF), a protein linked to neurite outgrowth and cellular motility (Hori et al. 1995).

The major inflammatory pathway stimulated by AGER activation is NFkappaB. Though the signaling cascade is unclear, several pieces of experimental data suggest that activation of AGER leads to sustained activation and upregulation of NFkappaB, measured as NFkappaB translocation to the nucleus, and increased levels of de novo synthesized NFkappaB (Bierhaus et al. 2001). As this is clearly an indirect effect it is represented here as positive regulation of NFkappaB translocation to the nucleus. AGER can bind ERK1/2 and thereby activate the MAPK and JNK cascades (Bierhaus et al. 2005).

##### Edit history

| Date | Action | Author |
|---|---|---|
| 2010-06-01 | Authored | Jupe S |
| 2010-06-17 | Created | Jupe S |
| 2010-09-01 | Edited | Jupe S |
| 2010-11-09 | Reviewed | Yan SD |
| 2021-05-22 | Modified | Shorser S |

#### Entities found in this pathway (1)

| Input | UniProt Id |
|---|---|
| LGALS3 | P17931 |

#### 24. Cell-Cell communication (R-HSA-1500931)

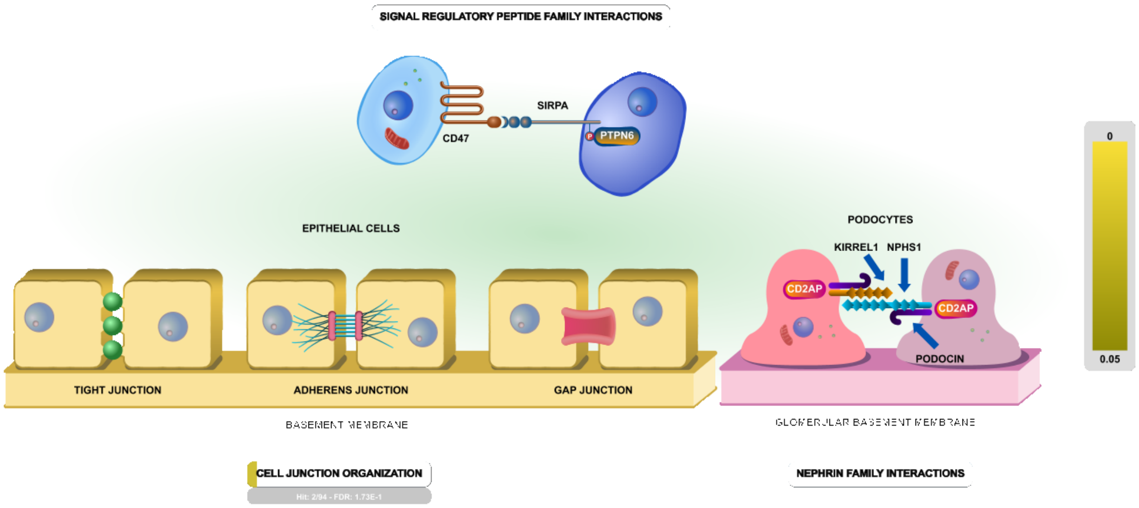


Cell-to-Cell communication is crucial for multicellular organisms because it allows organisms to coordinate the activity of their cells. Some cell-to-cell communication requires direct cell-cell contacts mediated by receptors on their cell surfaces. Members of the immunoglobulin superfamily (IgSF) proteins are some of the cell surface receptors involved in cell-cell recognition, communication and many aspects of the axon guidance and synapse formation-the crucial processes during embryonal development (Rougon & Hobert 2003).

Processes annotated here as aspects of **cell junction organization** mediate the formation and maintenance of adherens junctions, tight junctions, and gap junctions, as well as aspects of cellular interactions with extracellular matrix and hemidesmosome assembly. **Nephrin protein family interactions** are central to the formation of the slit diaphragm, a modified adherens junction. Interactions among members of the **signal regulatory protein family** are important for the regulation of migration and phagocytosis by myeloid cells.

##### Edit history

| Date | Action | Author |
| --- | --- | --- |
| 2011-08-23 | Edited | Garapati P V |
| 2011-08-23 | Authored | Garapati P V |

| Date | Action | Author |
|---|---|---|
| 2011-08-23 | Created | Matthews L |
| 2021-05-22 | Modified | Shorser S |

#### Entities found in this pathway (2)

| Input | UniProt Id | Input | UniProt Id |
|---|---|---|---|
| ITGB4 | P16144 | PLEC | Q15149 |

#### 25. Prolactin receptor signaling (R-HSA-1170546)

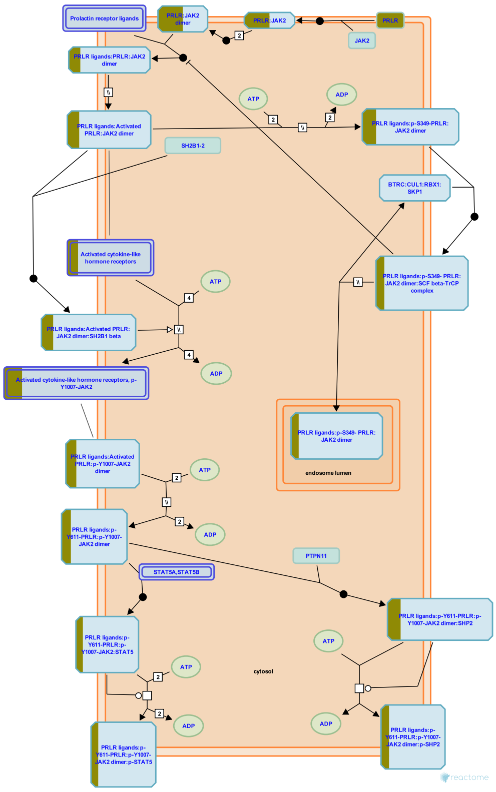


Prolactin (PRL) is a hormone secreted mainly by the anterior pituitary gland. It was originally identified by its ability to stimulate the development of the mammary gland and lactation, but is now known to have numerous and varied functions (Bole-Feysot et al. 1998). Despite this, few pathologies have been associated with abnormalities in prolactin receptor (PRLR) signaling, though roles in various forms of cancer and certain autoimmune disorders have been suggested (Goffin et al. 2002). A vast body of literature suggests effects of PRL in immune cells (Matera 1996) but PRLR KO mice have unaltered immune system development and function (Bouchard et al. 1999). In addition to the pituitary, numerous other tissues produce PRL, including the decidua and myometrium, certain cells of the immune system, brain, skin and exocrine glands such as the mammary, sweat and lacrimal glands (Ben-Jonathan et al. 1996). Pituitary PRL secretion is negatively regulated by inhibitory factors originating from the hypothalamus, the most important of which is dopamine, acting through the D2 subclass of dopamine receptors present in lactotrophs (Freeman et al. 2000). PRL-binding sites or receptors have been identified in numerous cells and tissues of adult mammals. Various forms of PRLR, generated by alternative splicing, have been reported in several species including humans (Kelly et al. 1991, Clevenger et al. 2003).

PRLR is a member of the cytokine receptor superfamily. Like many other members of this family, the first step in receptor activation was generally believed to be ligand-induced dimerization whereby one molecule of PRL bound to two molecules of receptor (Elkins et al. 2000). Recent reports suggest that PRLR pre-assembles at the plasma membrane in the absence of ligand (Gadd & Clevenger 2006, Tallet et al. 2011), suggesting that ligand-induced activation involves conformational changes in preformed PRLR dimers (Broutin et al. 2010).

PRLR has no intrinsic kinase activity but associates (Lebrun et al. 1994, 1995) with Janus kinase 2 (JAK2) which is activated following receptor activation (Campbell et al. 1994, Rui et al. 1994, Carter-Su et al. 2000, Barua et al. 2009). JAK2-dependent activation of JAK1 has also been reported (Neilson et al. 2007). It is generally accepted that activation of JAK2 occurs by transphosphorylation upon ligand-induced receptor activation, based on JAK activation by chimeric receptors in which various extracellular domains of cytokine or tyrosine kinase receptors were fused to the IL-2 receptor beta chain (see Ihle et al. 1994). This activation step involves the tyrosine phosphorylation of JAK2, which in turn phosphorylates PRLR on specific intracellular tyrosine residues leading to STAT5 recruitment and signaling, considered to be the most important signaling cascade for PRLR. STAT1 and STAT3 activation have also been reported (DaSilva et al. 1996) as have many other signaling pathways; signaling through MAP kinases (Shc/SOS/Grb2/Ras/Raf/MAPK) has been reported as a consequence of PRL stimuilation in many different cellular systems (see Bole-Feysot et al. 1998) though it is not clear how this signal is propagated. Other cascades non exhaustively include Src kinases, Focal adhesion kinase, phospholipase C gamma, PI3 kinase/Akt and Nek3 (Clevenger et al. 2003, Miller et al. 2007). The protein tyrosine phosphatase SHP2 is recruited to the C terminal tyrosine of PRLR and may have a regulatory role (Ali & Ali 2000). PRLR phosphotyrosines can recruit insulin receptor substrates (IRS) and other adaptor proteins to the receptor complex (Bole-Feysot et al. 1998).

Female homozygous PRLR knockout mice are completely infertile and show a lack of mammary development (Ormandy et al. 1997). Hemizogotes are unable to lactate following their first pregnancy and depending on the genetic background, this phenotype can persist through subsequent pregnancies (Kelly et al. 2001).


#### Edit history

| Date | Action | Author |
|---|---|---|
| 2011-01-19 | Created | Jupe S |
| 2011-06-13 | Authored | Jupe S |
| 2011-10-17 | Edited | Jupe S |
| 2011-11-08 | Reviewed | Goffin V |
| 2021-05-31 | Modified | Shorser S |

#### Entities found in this pathway (1)

| Input | UniProt Id |
|---|---|
| PRLR | P16471 |

### 6. Identifiers found

Below is a list of the input identifiers that have been found or mapped to an equivalent element in Reactome, classified by resource.

#### Entities (22)

| Input | UniProt Id | Input | UniProt Id | Input | UniProt Id |
|---|---|---|---|---|---|
| AKAP12 | Q02952 | AZGP1 | P25311 | CALCRL | Q16602 |
| CP | P00450 | CRYAB | P02511 | EFEMP1 | Q12805 |
| GSN | P06396 | IFITM1 | P13164 | ITGB4 | P16144 |
| KIF19 | Q2TAC6 | LGALS3 | P17931 | MGLL | Q99685 |
| OTOGL | Q3ZCN5 | PLCD1 | P51178 | PLEC | Q15149 |
| PPAP2B | O14495 | PRLR | P16471 | S100A1 | P31151, Q86SG5 |
| SEC24D | O94855 | SLC13A3 | Q8WWT9 | TMEM132E | Q24JP5 |
| VIM | P08670 | | | | |

| Input | Ensembl Id | Input | Ensembl Id | Input | Ensembl Id |
|---|---|---|---|---|---|
| IFITM1 | ENSG00000185885 | LGALS3 | ENSG00000131981 | VIM | ENSG00000026025 |

### 7. Identifiers not found

These 8 identifiers were not found neither mapped to any entity in Reactome.

| | | | | | | | |
|---|---|---|---|---|---|---|---|
| AK4P1 | PRELID2 | RN7SKP203 | SAMD14 | SLC16A9 | SULF1 | SYNM | TOX |
