## Supplementary material for "Compartment-specific total RNA profile of Hippocampal and Cortical cells from Mesial Temporal Lobe Epilepsy tissue": Other supplemental files_Reactome: HC_mTLE non-HS_Cytoplasm_UPspecific_report.pdf

### Pathway Analysis Report

This report contains the pathway analysis results for the submitted sample ''. Analysis was performed against Reactome version 77 on 14/09/2021. The web link to these results is:

https://reactome.org/PathwayBrowser/#/ANALYSIS=MjAyMTA5MTQwODEzMTlfNTM3ODU%3D

Please keep in mind that analysis results are temporarily stored on our server. The storage period depends on usage of the service but is at least 7 days. As a result, please note that this URL is only valid for a limited time period and it might have expired.

### Table of Contents

1. Introduction
2. Properties
3. Genome-wide overview
4. Most significant pathways
5. Pathways details
6. Identifiers found
7. Identifiers not found

### 1. Introduction

Reactome is a curated database of pathways and reactions in human biology. Reactions can be considered as pathway 'steps'. Reactome defines a 'reaction' as any event in biology that changes the state of a biological molecule. Binding, activation, translocation, degradation and classical biochemical events involving a catalyst are all reactions. Information in the database is authored by expert biologists, entered and maintained by Reactome's team of curators and editorial staff. Reactome content frequently cross-references other resources e.g. NCBI, Ensembl, UniProt, KEGG (Gene and Compound), ChEBI, PubMed and GO. Orthologous reactions inferred from annotation for Homo sapiens are available for 17 non-human species including mouse, rat, chicken, puffer fish, worm, fly, yeast, rice, and Arabidopsis. Pathways are represented by simple diagrams following an SBGN-like format.

Reactome's annotated data describe reactions possible if all annotated proteins and small molecules were present and active simultaneously in a cell. By overlaying an experimental dataset on these annotations, a user can perform a pathway over-representation analysis. By overlaying quantitative expression data or time series, a user can visualize the extent of change in affected pathways and its progression. A binomial test is used to calculate the probability shown for each result, and the p-values are corrected for the multiple testing (Benjamini–Hochberg procedure) that arises from evaluating the submitted list of identifiers against every pathway.

To learn more about our Pathway Analysis, please have a look at our relevant publications:


Fabregat A, Sidiropoulos K, Garapati P, Gillespie M, Hausmann K, Haw R, … D'Eustachio P (2016). The reactome pathway knowledgebase. Nucleic Acids Research, 44(D1), D481–D487. https://doi.org/10.1093/nar/gkv1351.

Fabregat A, Sidiropoulos K, Viteri G, Forner O, Marin-Garcia P, Arnau V, … Hermjakob H (2017). Reactome pathway analysis: a high-performance in-memory approach. BMC Bioinformatics, 18.

#### 2. Properties

- This is an **overrepresentation** analysis: A statistical (hypergeometric distribution) test that determines whether certain Reactome pathways are over-represented (enriched) in the submitted data. It answers the question 'Does my list contain more proteins for pathway X than would be expected by chance?' This test produces a probability score, which is corrected for false discovery rate using the Benjamani-Hochberg method.
- 87 out of 144 identifiers in the sample were found in Reactome, where 393 pathways were hit by at least one of them.
- All non-human identifiers have been converted to their human equivalent.
- This report is filtered to show only results for species 'Homo sapiens' and resource 'all resources'.
- The unique ID for this analysis (token) is MjAyMTA5MTQwODEzMTlfNTM3ODU%3D. This ID is valid for at least 7 days in Reactome's server. Use it to access Reactome services with your data.

### 3. Genome-wide overview

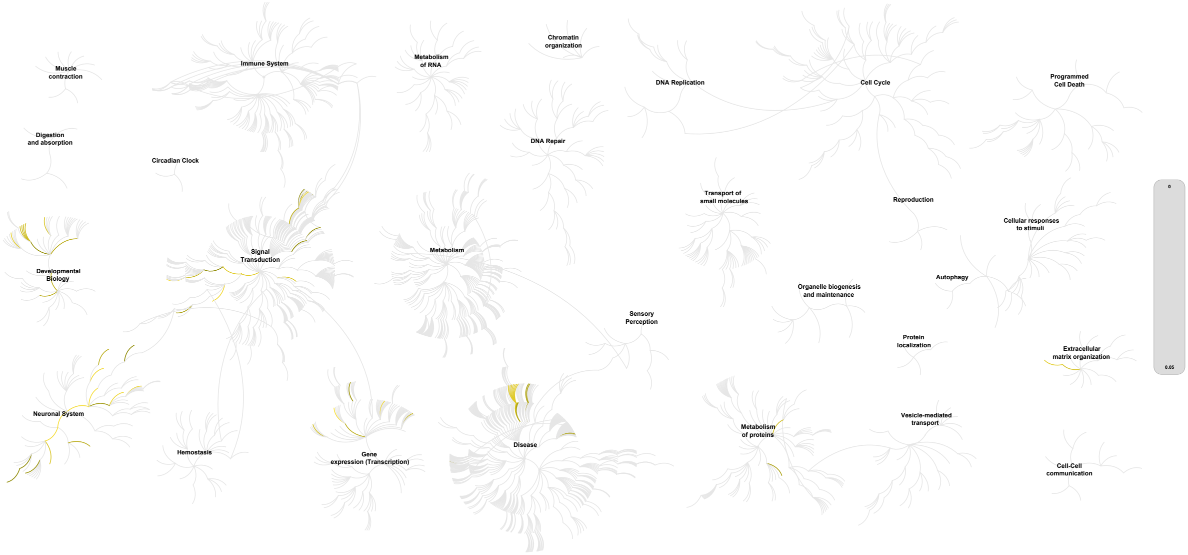


This figure shows a genome-wide overview of the results of your pathway analysis. Reactome pathways are arranged in a hierarchy. The center of each of the circular "bursts" is the root of one top-level pathway, for example "DNA Repair". Each step away from the center represents the next level lower in the pathway hierarchy. The color code denotes over-representation of that pathway in your input dataset. Light grey signifies pathways which are not significantly over-represented.

### 4. Most significant pathways

The following table shows the 25 most relevant pathways sorted by p-value.

| Pathway name | Entities | | | | Reactions | |
|---|---|---|---|---|---|---|
| | found | ratio | p-value | FDR* | found | ratio |
| Neuronal System | 17 / 487 | 0.033 | 5.22e-05 | 0.021 | 55 / 216 | 0.016 |
| Adrenoceptors | 3 / 11 | 7.56e-04 | 3.01e-04 | 0.061 | 1 / 7 | 5.18e-04 |
| MECP2 regulates neuronal receptors and channels | 4 / 32 | 0.002 | 5.51e-04 | 0.065 | 4 / 26 | 0.002 |
| Transmission across Chemical Synapses | 12 / 341 | 0.023 | 6.35e-04 | 0.065 | 45 / 163 | 0.012 |
| GABA receptor activation | 5 / 67 | 0.005 | 0.001 | 0.092 | 5 / 12 | 8.88e-04 |
| Neurotransmitter receptors and postsynaptic signal transmission | 9 / 231 | 0.016 | 0.002 | 0.092 | 41 / 109 | 0.008 |
| Potassium Channels | 6 / 107 | 0.007 | 0.002 | 0.092 | 5 / 19 | 0.001 |
| Activation of AMPA receptors | 2 / 7 | 4.81e-04 | 0.003 | 0.127 | 5 / 5 | 3.70e-04 |
| FGFR1c and Klotho ligand binding and activation | 2 / 7 | 4.81e-04 | 0.003 | 0.127 | 2 / 2 | 1.48e-04 |
| G alpha (12/13) signalling events | 5 / 85 | 0.006 | 0.003 | 0.127 | 5 / 15 | 0.001 |
| Unblocking of NMDA receptors, glutamate binding and activation | 3 / 27 | 0.002 | 0.004 | 0.144 | 5 / 5 | 3.70e-04 |
| RUNX1 regulates transcription of genes involved in WNT signaling | 2 / 9 | 6.19e-04 | 0.005 | 0.169 | 2 / 4 | 2.96e-04 |
| Long-term potentiation | 3 / 31 | 0.002 | 0.006 | 0.171 | 3 / 7 | 5.18e-04 |
| Regulation of commissural axon pathfinding by SLIT and ROBO | 2 / 12 | 8.25e-04 | 0.009 | 0.171 | 4 / 5 | 3.70e-04 |
| Molecules associated with elastic fibres | 3 / 38 | 0.003 | 0.01 | 0.171 | 4 / 10 | 7.40e-04 |
| GPCR ligand binding | 14 / 606 | 0.042 | 0.011 | 0.171 | 10 / 186 | 0.014 |
| Signaling by GPCR | 18 / 864 | 0.059 | 0.011 | 0.171 | 59 / 354 | 0.026 |
| Drug resistance of KIT mutants | 1 / 1 | 6.88e-05 | 0.011 | 0.171 | 7 / 7 | 5.18e-04 |
| KIT mutants bind TKIs | 1 / 1 | 6.88e-05 | 0.011 | 0.171 | 2 / 2 | 1.48e-04 |
| Imatinib-resistant KIT mutants | 1 / 1 | 6.88e-05 | 0.011 | 0.171 | 1 / 1 | 7.40e-05 |
| Sunitinib-resistant KIT mutants | 1 / 1 | 6.88e-05 | 0.011 | 0.171 | 1 / 1 | 7.40e-05 |
| Sorafenib-resistant KIT mutants | 1 / 1 | 6.88e-05 | 0.011 | 0.171 | 1 / 1 | 7.40e-05 |
| Dasatinib-resistant KIT mutants | 1 / 1 | 6.88e-05 | 0.011 | 0.171 | 1 / 1 | 7.40e-05 |
| Nilotinib-resistant KIT mutants | 1 / 1 | 6.88e-05 | 0.011 | 0.171 | 1 / 1 | 7.40e-05 |
| Masitinib-resistant KIT mutants | 1 / 1 | 6.88e-05 | 0.011 | 0.171 | 1 / 1 | 7.40e-05 |

* False Discovery Rate

### 5. Pathways details

For every pathway of the most significant pathways, we present its diagram, as well as a short summary, its bibliography and the list of inputs found in it.

#### 1. Neuronal System (R-HSA-112316)

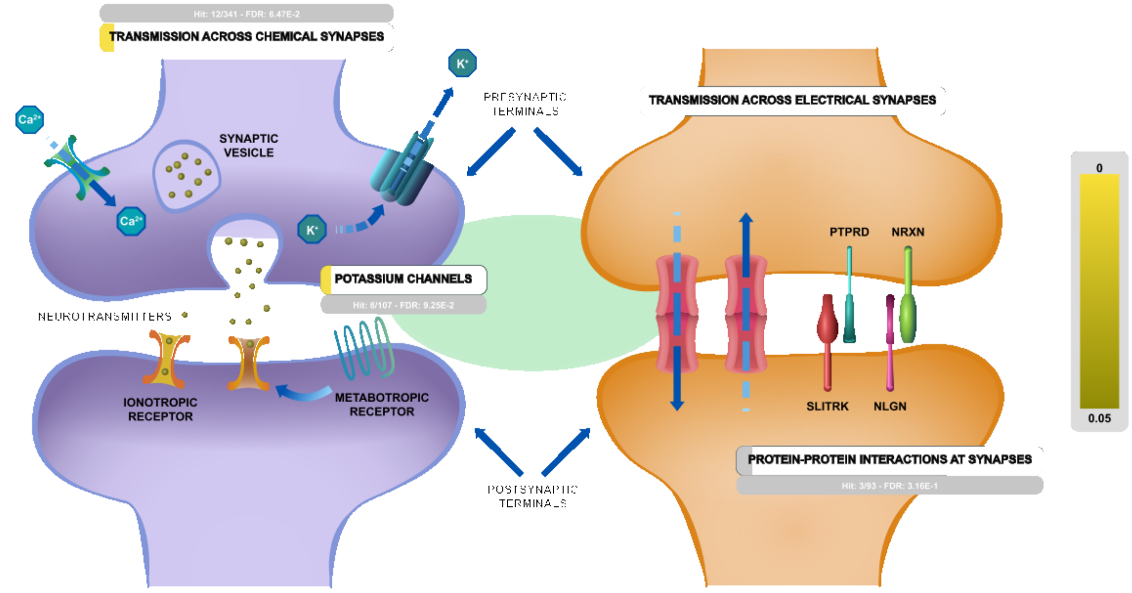


The human brain contains at least 100 billion neurons, each with the ability to influence many other cells. Clearly, highly sophisticated and efficient mechanisms are needed to enable communication among this astronomical number of elements. This communication occurs across synapses, the functional connection between neurons. Synapses can be divided into two general classes: electrical synapses and chemical synapses. Electrical synapses permit direct, passive flow of electrical current from one neuron to another. The current flows through gap junctions, specialized membrane channels that connect the two cells. Chemical synapses enable cell-to-cell communication using neurotransmitter release. Neurotransmitters are chemical agents released by presynaptic neurons that trigger a secondary current flow in postsynaptic neurons by activating specific receptor molecules. Neurotransmitter secretion is triggered by the influx of Ca2+ through voltage-gated channels, which gives rise to a transient increase in Ca2+ concentration within the presynaptic terminal. The rise in Ca2+ concentration causes synaptic vesicles (the presynaptic organelles that store neurotransmitters) to fuse with the presynaptic plasma membrane and release their contents into the space between the pre- and postsynaptic cells.

##### Edit history

| Date | Action | Author |
|---|---|---|
| 2004-04-22 | Created | Joshi-Tope G |
| 2005-11-10 | Edited | Gillespie ME |

| Date | Action | Author |
|---|---|---|
| 2005-11-10 | Authored | Gillespie ME |
| 2021-05-22 | Modified | Shorser S |

#### Entities found in this pathway (15)

| Input | UniProt Id | Input | UniProt Id | Input | UniProt Id |
|---|---|---|---|---|---|
| ACHE | P22303 | CACNA2D1 | Q9UIU0 | GABRA3 | P34903 |
| GABRA5 | P31644 | GABRQ | Q9UN88 | GNG2 | P59768 |
| GRIA1 | P42261 | GRIA2 | P42262 | GRIN2B | Q13224 |
| KCNA3 | P22001, Q09470 | KCNJ6 | P48051 | KCNK2 | O95069, Q13303 |
| LRRTM2 | O43300 | NBEA | Q8NFP9 | SLC1A6 | P48664 |

#### 2. Adrenoceptors (R-HSA-390696)

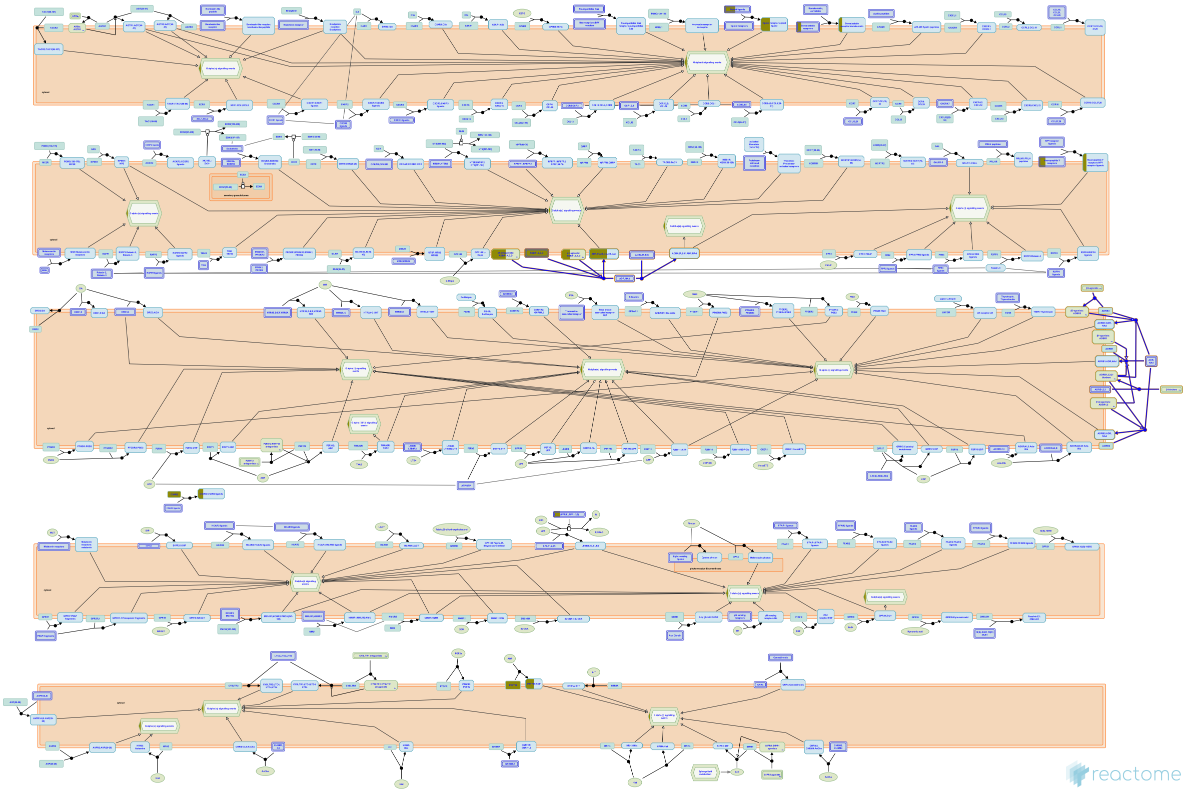

The adrenoceptors (adrenergic receptors) are targets for the catecholamines adrenaline (epinephrine) and noradrenaline (norepinephrine). These receptors are widespread in the body and binding of catecholamines produces a sympathetic response ('flight-or-fight response') resulting in increased heart rate, pupil dilation and energy mobilization amongst other responses. There are three major types of adrenoceptor, alpha1, alpha2 and beta. Each type is thought to have three subtypes; alpha1 (1A,1B,1D), alpha2 (2A,2B,2C) and beta (1,2,3) (Bylund DB et al, 1994).


##### Edit history

| Date | Action | Author |
| --- | --- | --- |
| 2009-02-10 | Edited | Jassal B |
| 2009-02-10 | Authored | Jassal B |
| 2009-02-10 | Created | Jassal B |
| 2009-03-02 | Reviewed | D'Eustachio P |
| 2021-05-22 | Modified | Shorser S |

##### Entities found in this pathway (2)

| Input | UniProt Id | Input | UniProt Id |
| --- | --- | --- | --- |
| ADRA1A | P25100, P35348 | ADRA1B | P35368 |

#### 3. MECP2 regulates neuronal receptors and channels (R-HSA-9022699)

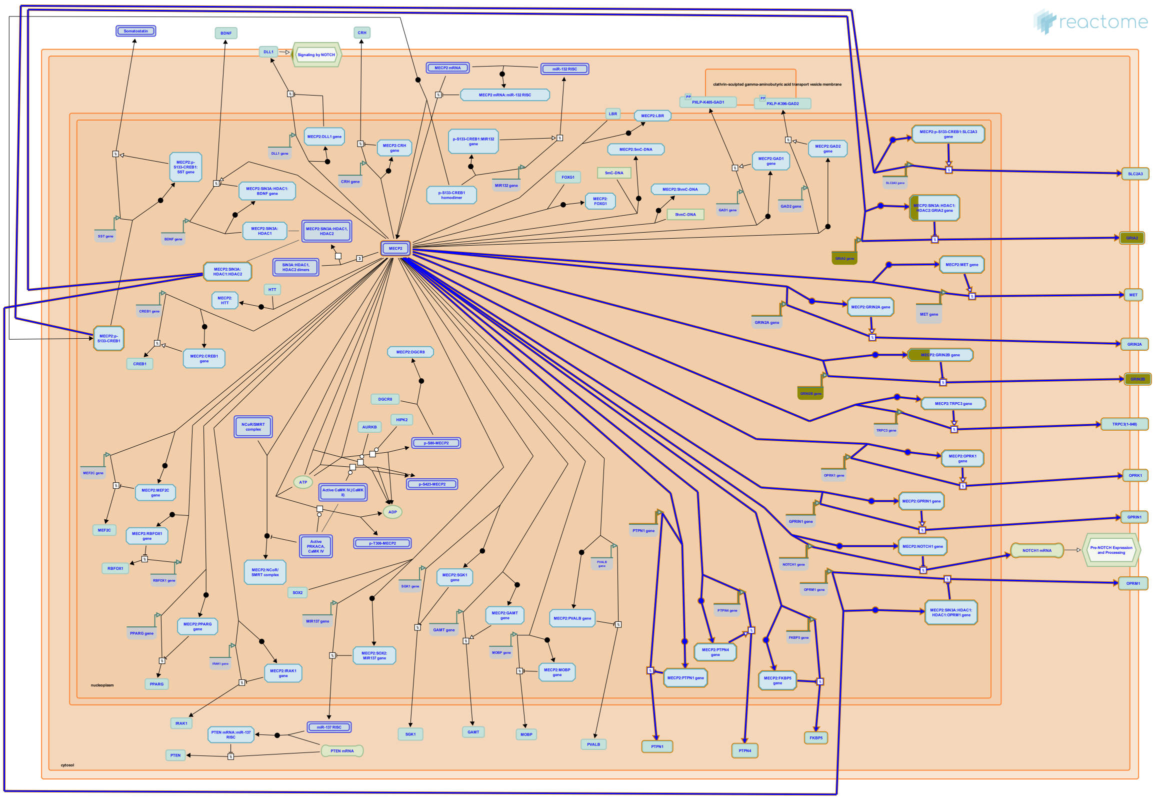

Receptors directly transcriptionally regulated by MECP2 include glutamate receptor GRIA2 (Qiu et al. 2012), NMDA receptor subunits GRIN2A (Durand et al. 2012) and GRIN2B (Lee et al. 2008), opioid receptors OPRK1 (Chahrour et al. 2008) and OPRM1 (Hwang et al. 2009, Hwang et al. 2010, Samaco et al. 2012), GPRIN1 (Chahrour et al. 2008), MET (Plummer et al. 2013), and NOTCH1 (Li et al. 2014). Channels/transporters regulated by MECP2 include TRPC3 (Li et al. 2012) and SLC2A3 (Chen et al. 2013). MECP2 also regulates transcription of FKBP5, involved in trafficking of glucocorticoid receptors (Nuber et al. 2005, Urdinguio et al. 2008) and is implicated in regulation of expression of SEMA3F (semaphorin 3F) in mouse olfactory neurons (Degano et al. 2009). In zebrafish, Mecp2 is implicated in sensory axon guidance by direct stimulation of transcription of Sema5b and Robo2 (Leong et al. 2015). MECP2 may indirectly regulate signaling by neuronal receptor tyrosine kinases by regulating transcription of protein tyrosine phosphatases, PTPN1 (Krishnan et al. 2015) and PTPN4 (Williamson et al. 2015).

#### Edit history

| Date | Action | Author |
|---|---|---|
| 2017-09-25 | Created | Orlic-Milacic M |
| 2017-10-02 | Authored | Orlic-Milacic M |
| 2018-08-07 | Reviewed | Christodoulou J, Krishnaraj R |
| 2018-08-08 | Modified | Orlic-Milacic M |
| 2018-08-08 | Edited | Orlic-Milacic M |

#### Entities found in this pathway (2)

| Input | UniProt Id |
|---|---|
| GRIA2 | P42262 |

| Input | UniProt Id |
|---|---|
| GRIN2B | Q13224 |

| Input | Ensembl Id |
|---|---|
| GRIA2 | ENSG00000120251 |

| Input | Ensembl Id |
|---|---|
| GRIN2B | ENSG00000273079 |

#### 4. Transmission across Chemical Synapses (R-HSA-112315)

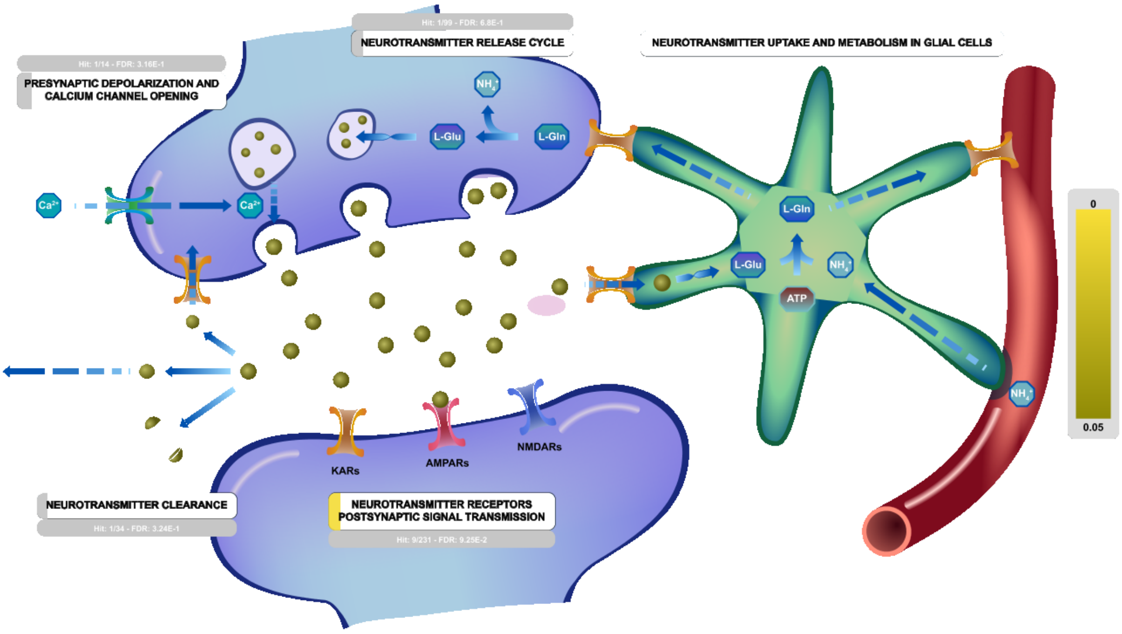


Chemical synapses are specialized junctions that are used for communication between neurons, neurons and muscle or gland cells. The synapse involves a presynaptic neuron and a postsynaptic neuron, muscle cell or glad cell. The pre and the postsynaptic cell are separated by a gap (space) of 20 to 40 nm called the synaptic cleft. The signals pass in a single direction from the presynaptic to postsynaptic neuron (cell). The presynaptic neuron communicates via the release of neurotransmitter which bind the receptors on the postsynaptic cell. The process is initiated when an action potential invades the terminal membrane of the presynaptic neuron.

Action potentials occur in electrically excitable cells such as neurons and muscles and endocrine cells. They are initiated by the transient opening of voltage dependent sodium channels, causing a rapid, large depolarization of membrane potentials that spread along the axon membrane.

When action potentials arrive at the synaptic terminals, depolarization in membrane potential leads to the opening of voltage gated calcium channels located on the presynaptic membrane. The external Ca2+ concentration is approximately 10-3 M while the internal Ca2+ concentration is approximately 10-7 M. Opening of calcium channels causes a rapid influx of Ca2+ into the presynaptic terminal. The elevated presynaptic Ca2+ concentration allows synaptic vesicles to fuse with the plasma membrane of the presynaptic neuron and release their contents, neurotransmitters, into the synaptic cleft. These diffuse across the synaptic cleft and bind to specific receptors on the membrane of the postsynaptic cells. Activation of postsynaptic receptors upon neurotransmitter binding can lead to a multitude of effects in the postsynaptic cell, such as changing the membrane potential and excitability, and triggering intracellular signaling cascades.

##### Edit history

| Date | Action | Author |
| --- | --- | --- |
| 2004-04-22 | Created | Joshi-Tope G |
| 2008-01-14 | Edited | Mahajan SS |
| 2008-01-14 | Authored | Mahajan SS |
| 2008-12-02 | Reviewed | Restituito S, Kavalali E |
| 2020-01-24 | Reviewed | Wen H |
| 2021-05-22 | Modified | Shorser S |

#### Entities found in this pathway (12)

| Input | UniProt Id | Input | UniProt Id | Input | UniProt Id |
| --- | --- | --- | --- | --- | --- |
| ACHE | P22303 | CACNA2D1 | Q9UIU0 | GABRA3 | P34903 |
| GABRA5 | P31644 | GABRQ | Q9UN88 | GNG2 | P59768 |
| GRIA1 | P42261 | GRIA2 | P42262 | GRIN2B | Q13224 |
| KCNJ6 | P48051 | NBEA | Q8NFP9 | SLC1A6 | P48664 |

#### 5. GABA receptor activation (R-HSA-977443)

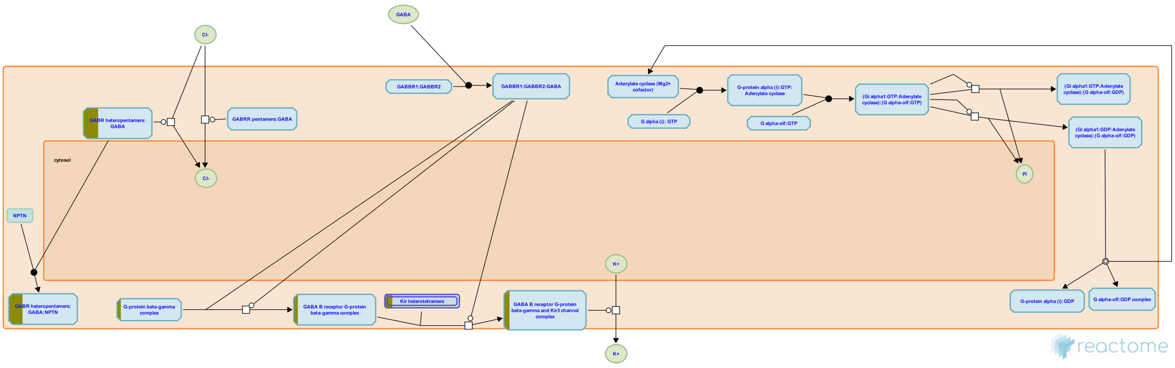

**Cellular compartments:** plasma membrane, cytosol, extracellular region.

Gamma aminobutyric acid (GABA) receptors are the major inhibitory receptors in human synapses. They are of two types. GABA A receptors are fast-acting ligand gated chloride ion channels that mediate membrane depolarization and thus inhibit neurotransmitter release (G Michels et al Crit Rev Biochem Mol Biol 42, 2007, 3-14). GABA B receptors are slow acting metabotropic Gprotein coupled receptors that act via the inhibitory action of their Galpha/Go subunits on adenylate cyclase to attenuate the actions of PKA. In addition, their Gbeta/gamma subunits interact directly with N and P/Q Ca2+ channels to decrease the release of Ca2+. GABA B receptors also interact with Kir3 K+ channels and increase the influx of K+, leading to cell membrane hyperpolarization and inhibition of channels such as NMDA receptors (A Pinard et al Adv Pharmacol, 58, 2010, 231-55).


##### Edit history

| Date | Action | Author |
|---|---|---|
| 2008-11-27 | Reviewed | Restituito S |
| 2010-10-19 | Created | Mahajan SS |
| 2010-11-08 | Authored | Mahajan SS |
| 2010-11-25 | Edited | D'Eustachio P |
| 2021-05-22 | Modified | Shorser S |

##### Entities found in this pathway (5)

| Input | UniProt Id | Input | UniProt Id | Input | UniProt Id |
|---|---|---|---|---|---|
| GABRA3 | P34903 | GABRA5 | P31644 | GABRQ | Q9UN88 |
| GNG2 | P59768 | KCNJ6 | P48051 | | |

#### 6. Neurotransmitter receptors and postsynaptic signal transmission (R-HSA-112314)

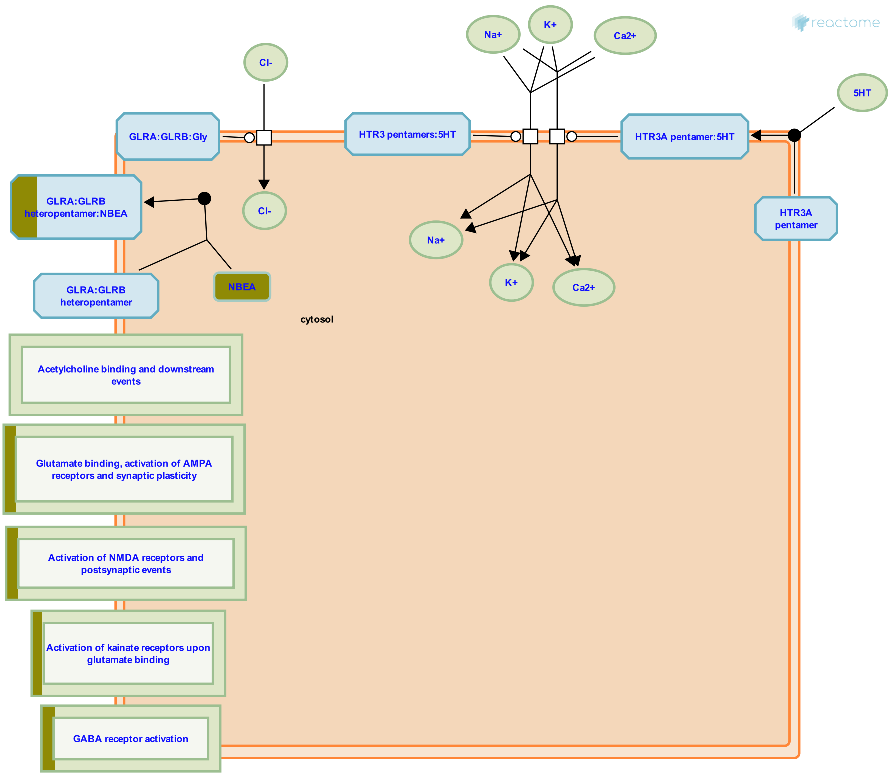


The neurotransmitter in the synaptic cleft released by the pre-synaptic neuron binds specific receptors located on the post-synaptic terminal. These receptors are either ion channels or G protein coupled receptors that function to transmit the signals from the post-synaptic membrane to the cell body.

##### References

##### Edit history

| Date | Action | Author |
|---|---|---|
| 2004-04-22 | Created | Joshi-Tope G |
| 2008-01-14 | Authored | Mahajan SS |
| 2008-12-02 | Reviewed | Restituito S, Kavalali E |
| 2021-05-22 | Modified | Shorser S |

##### Entities found in this pathway (9)

| Input | UniProt Id | Input | UniProt Id | Input | UniProt Id |
|---|---|---|---|---|---|
| GABRA3 | P34903 | GABRA5 | P31644 | GABRQ | Q9UN88 |
| GNG2 | P59768 | GRIA1 | P42261 | GRIA2 | P42262 |
| GRIN2B | Q13224 | KCNJ6 | P48051 | NBEA | Q8NFP9 |

#### 7. Potassium Channels (R-HSA-1296071)

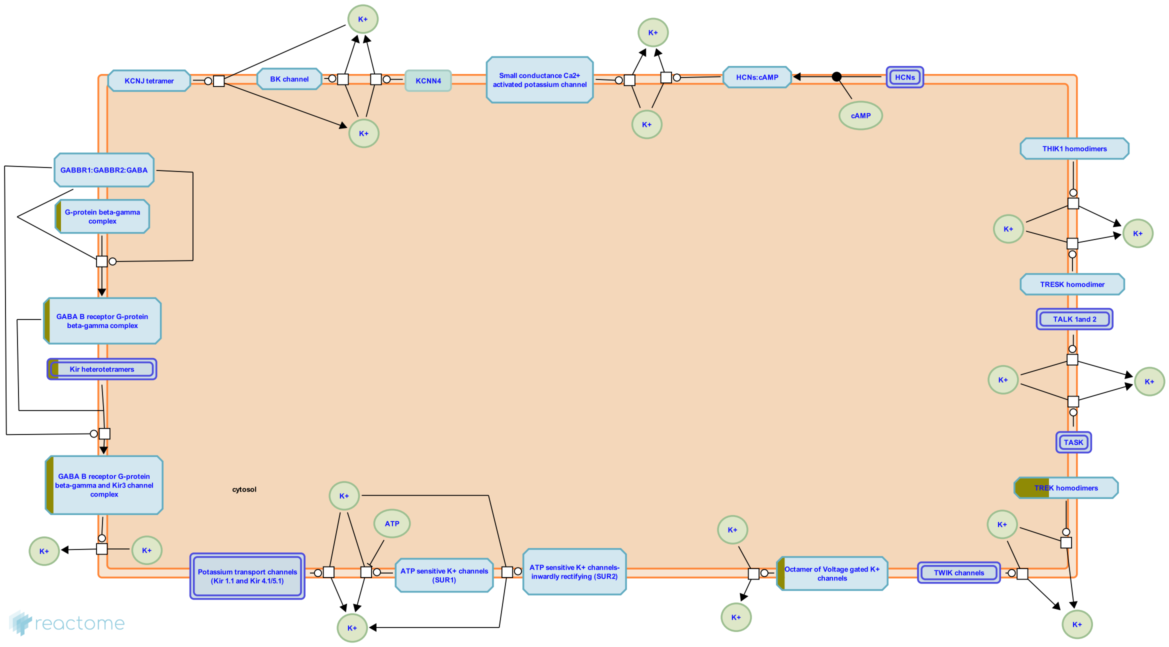


Potassium channels are tetrameric ion channels that are widely distributed and are found in all cell types. Potassium channels control resting membrane potential in neurons, contribute to regulation of action potentials in cardiac muscle and help release of insulin form pancreatic beta cells.

Broadly K+ channels are classified into voltage gated K+ channels, Hyperpolarization activated cyclic nucleotide gated K+ channels (HCN), Tandem pore domain K+ channels, Ca2+ activated K+ channels and inwardly rectifying K+ channels.

##### Edit history

| Date | Action | Author |
| --- | --- | --- |
| 2010-09-23 | Reviewed | Jassal B |
| 2011-05-19 | Authored | Mahajan SS |
| 2011-05-19 | Created | Mahajan SS |
| 2011-05-23 | Edited | Mahajan SS |

| Date | Action | Author |
| --- | --- | --- |
| 2021-05-22 | Modified | Shorser S |

#### Entities found in this pathway (4)

| Input | UniProt Id | Input | UniProt Id |
| --- | --- | --- | --- |
| GNG2 | P59768 | KCNA3 | P22001, Q09470 |
| KCNJ6 | P48051 | KCNK2 | O95069, Q13303 |

#### 8. Activation of AMPA receptors (R-HSA-399710)

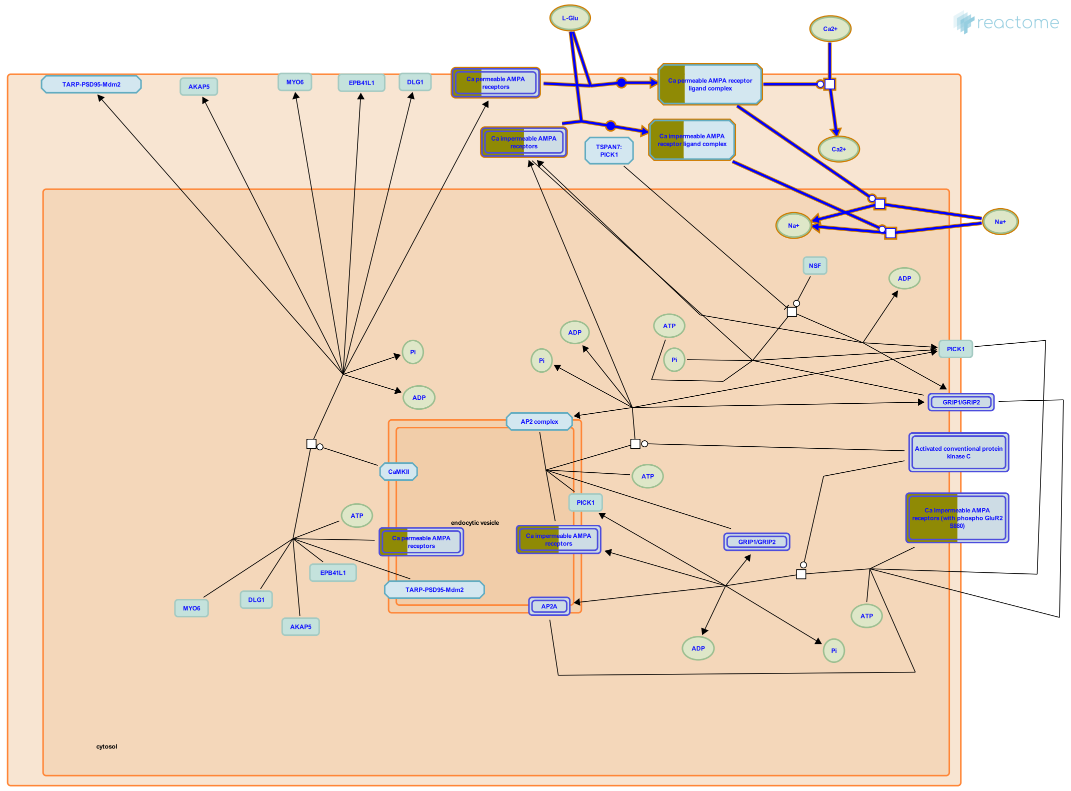


**Cellular compartments:** plasma membrane, extracellular region.

AMPA receptors are functionally either Ca permeable or Ca impermeable based on the subunit composition. Ca permeability is determined by GluR2 subunit which undergoes post-transcriptional RNA editing that changes glutamine (Q) at the pore to arginine (R). Incorporation of even a single subunit in the AMPA receptor confers Ca-limiting properties. Ca permeable AMPA receptors permit Ca and Na whereas Ca impermeable AMPA receptors permit only Na. In general, glutamatergic neurons contain Ca impermeable AMPA receptors and GABAergic interneurons contain Ca permeable AMPA receptors. However, some synapses do contain a mixture of Ca permeable and Ca impermeable AMPA receptors. GluR1-4 are encoded by four genes however, alternative splicing generates several functional subunits namely long and short forms of GluR1 and GluR2. GluR4 has long tail only and GluR3 has short tail only. Besides the differences in the tail length, flip/flop isoforms are generated by an interchangeable exon that codes the fourth membranous domain towards the C terminus. The fip/flop isoforms determine rate of desensitization/resensitization and the rate of channel closing. Receptors homomers or heteromers assembled from the combination of GluR1-4 subunits that vary in C tail length and flip/flop versions generates a whole battery of functionally distinct AMPA receptors.

#### Edit history

| Date | Action | Author |
|---|---|---|
| 2008-01-14 | Edited | Mahajan SS |
| 2008-01-14 | Authored | Mahajan SS |
| 2009-03-19 | Created | Mahajan SS |
| 2009-05-15 | Reviewed | Ziff EB |
| 2021-05-31 | Modified | Shorser S |

#### Entities found in this pathway (2)

| Input | UniProt Id | Input | UniProt Id |
|---|---|---|---|
| GRIA1 | P42261 | GRIA2 | P42262 |

#### 9. FGFR1c and Klotho ligand binding and activation (R-HSA-190374)

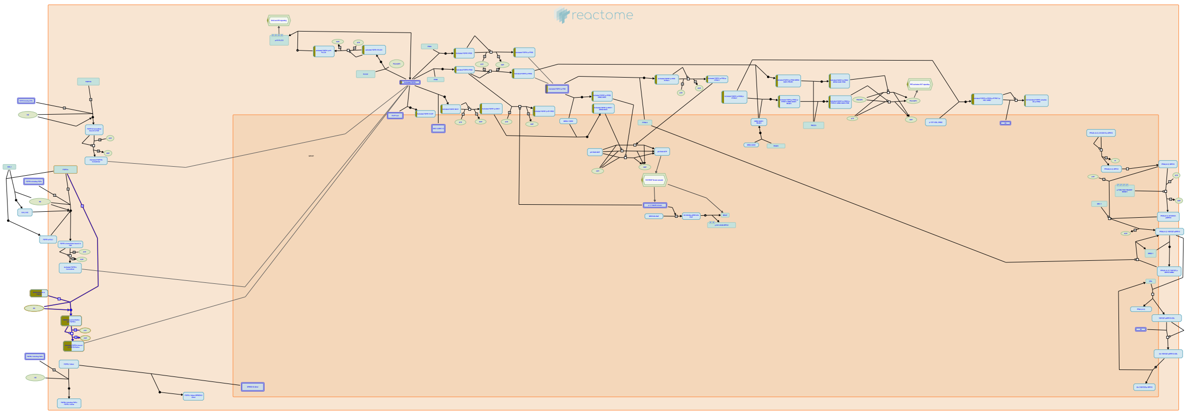

FGF23 is a member of the endocrine subfamily of FGFs. It is produced in bone tissue and regulates kidney functions. Klotho is essential for endogenous FGF23 function as it converts FGFR1c into a specific FGF23 receptor.


##### Edit history

| Date | Action | Author |
|---|---|---|
| 2007-01-02 | Created | de Bono B |
| 2007-01-10 | Authored | de Bono B |
| 2007-02-07 | Reviewed | Mohammadi M |
| 2021-05-21 | Modified | Shorser S |

##### Entities found in this pathway (1)

| Input | UniProt Id |
|---|---|
| KL | Q9UEF7-1, Q9UEF7-2 |

#### 10. G alpha (12/13) signalling events (R-HSA-416482)

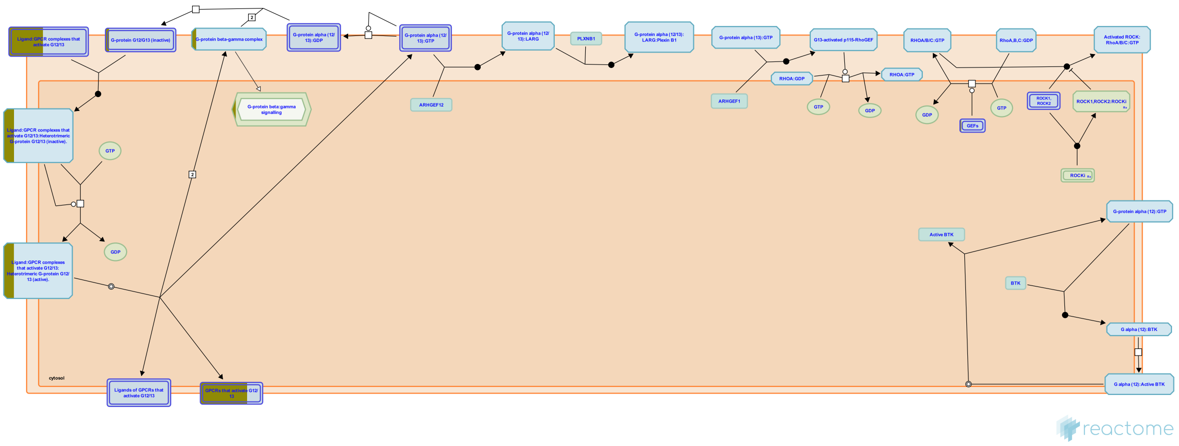

**Cellular compartments:** cytoplasmic side of plasma membrane.

The G12/13 family is probably the least well characterized subtype, partly because G12/13 coupling is difficult to determine when compared with the other subtypes which predominantly rely on assay technologies that measure intracellular calcium. The G12/13 family are best known for their involvement in the processes of cell proliferation and morphology, such as stress fiber and focal adhesion formation. Interactions with Rho guanine nucleotide exchange factors (RhoGEFs) are thought to mediate many of these processes. (Buhl et al.1995, Sugimoto et al. 2003). Activation of Rho or the regulation of events through Rho is often taken as evidence of G12/13 signaling. Receptors that are coupled with G12/13 invariably couple with one or more other G protein subtypes, usually Gq.


##### Edit history

| Date | Action | Author |
|---|---|---|
| 2009-03-18 | Authored | Jupe S |
| 2009-03-27 | Edited | Jupe S |
| 2009-03-27 | Created | Jupe S |
| 2009-06-03 | Reviewed | Akkerman JW |
| 2021-05-31 | Modified | Shorser S |

##### Entities found in this pathway (4)

| Input | UniProt Id | Input | UniProt Id |
|---|---|---|---|
| ADRA1A | P25100, P35348 | ADRA1B | P35368 |
| ARHGEF3 | Q9NR81 | GNG2 | P59768 |

#### 11. Unblocking of NMDA receptors, glutamate binding and activation (R-HSA-438066)

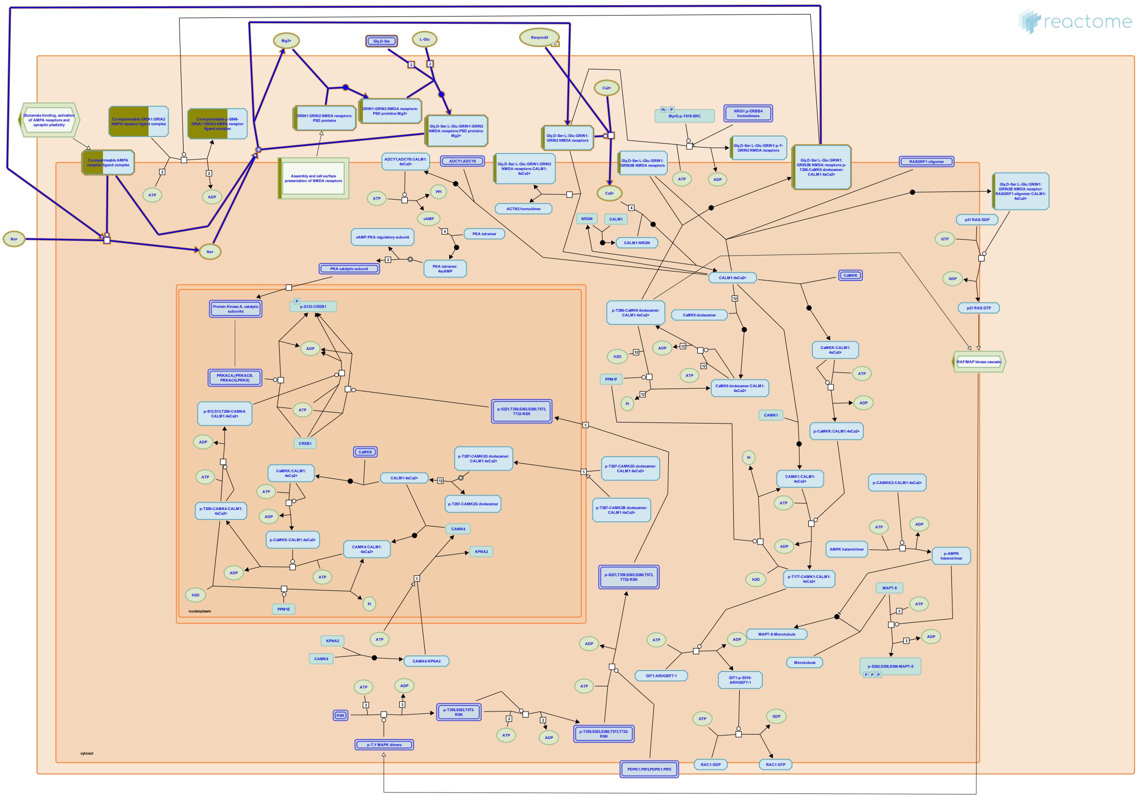

**Cellular compartments:** plasma membrane.

At resting membrane potential, the NMDA receptor ion channel is blocked by extracellular Mg2+ ions and is unable to mediate ion permeation upon binding of ligands (glutamate, glycine, D-serine, NMDA). The voltage block is removed upon depolarization of the post-synaptic cell membrane and Mg2+ is expelled from the NMDA receptor pore (channel), resulting in activated ligand-bound NMDA receptors. The depolarization of the membrane may happen in response to activation of Ca2+ impermeable AMPA receptors, which facilitates Na+ influx, contributing to the unblocking of NMDA receptors. For review, please refer to Traynelis et al. 2010, Paoletti et al. 2013, and Iacobucci and Popescu 2017.


##### Edit history

| Date | Action | Author |
|---|---|---|
| 2009-09-23 | Created | Mahajan SS |

| Date | Action | Author |
|---|---|---|
| 2009-10-29 | Authored | Mahajan SS |
| 2009-11-18 | Reviewed | Tukey D |
| 2009-11-19 | Edited | Gillespie ME |
| 2018-07-31 | Edited | Orlic-Milacic M |
| 2018-10-10 | Revised | Orlic-Milacic M |
| 2018-11-02 | Reviewed | Hansen KB, Yi F |
| 2018-11-07 | Edited | Orlic-Milacic M |
| 2021-05-22 | Modified | Shorser S |

#### Entities found in this pathway (3)

| Input | UniProt Id | Input | UniProt Id | Input | UniProt Id |
|---|---|---|---|---|---|
| GRIA1 | P42261 | GRIA2 | P42262 | GRIN2B | Q13224 |

#### 12. RUNX1 regulates transcription of genes involved in WNT signaling (R-HSA-8939256)

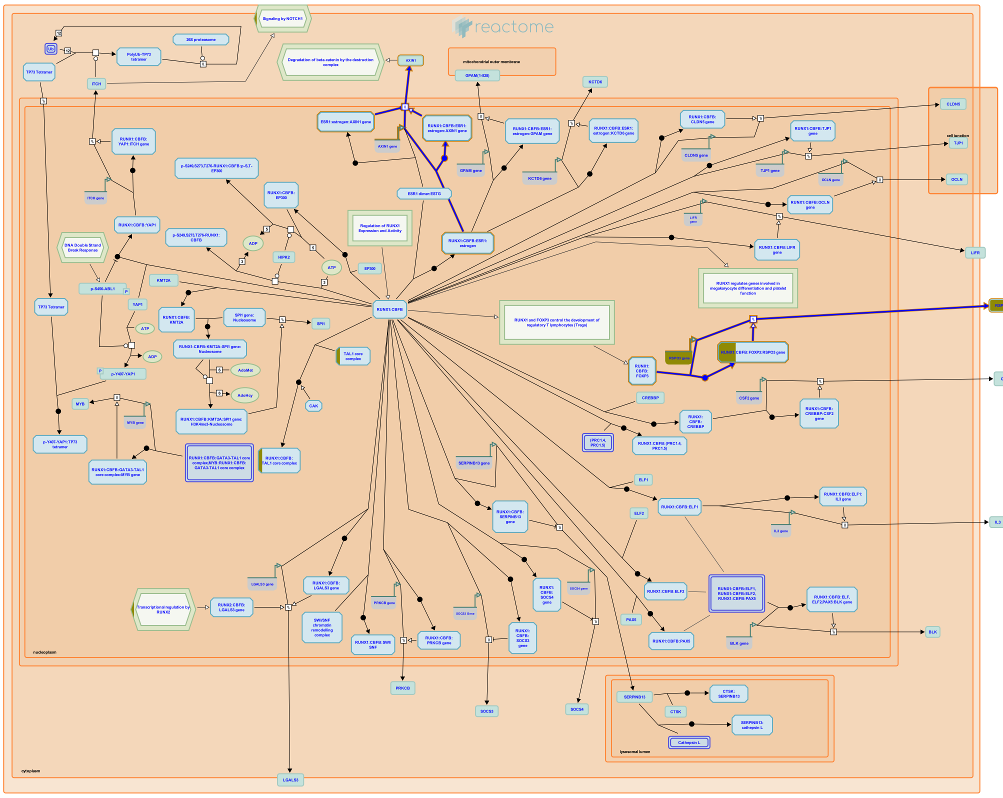


**Cellular compartments:** nucleoplasm.

The RUNX1:CBFB complex directly regulates transcription of at least two components of WNT signaling. In association with its co-factor FOXP3, the RUNX1:CBFB complex stimulates transcription of the RSPO3 gene, encoding a WNT ligand that is implicated as a breast cancer oncogene (Recouvreux et al. 2016). In association with the activated estrogen receptor alpha (ESR1), the RUNX1:CBFB complex stimulates the expression of AXIN1, which functions as a regulator of WNT signaling (Stender et al. 2010).


##### Edit history

| Date | Action | Author |
|---|---|---|
| 2016-09-14 | Authored | Orlic-Milacic M |
| 2016-09-16 | Created | Orlic-Milacic M |
| 2016-12-20 | Reviewed | Ito Y, Chuang LS |

| Date | Action | Author |
|---|---|---|
| 2017-05-09 | Modified | Orlic-Milacic M |
| 2017-05-09 | Edited | Orlic-Milacic M |

#### Entities found in this pathway (1)

| Input | UniProt Id |
|---|---|
| RSPO3 | Q9BXY4 |

| Input | Ensembl Id |
|---|---|
| RSPO3 | ENSG00000146374 |

#### 13. Long-term potentiation (R-HSA-9620244)

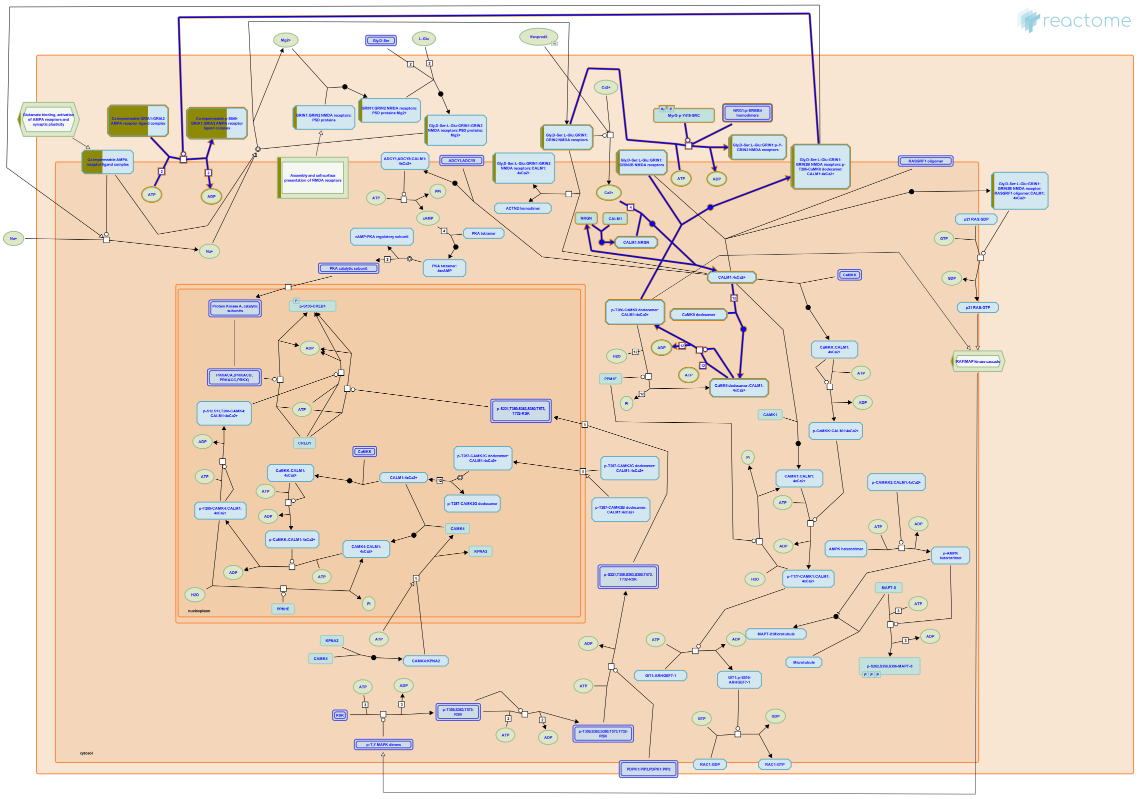

In long-term potentiation (LTP), involved in learning and memory, a brief period of synaptic activity induces a lasting increase in the strength of the synapse. LTP is initiated by NMDA receptor-mediated activation of calcium/calmodulin-dependent protein kinase II (CaMKII), followed by binding of CaMKII to the NMDA receptor and CaMKII-mediated phosphorylation of AMPA receptor subunits (reviewed by Lisman et al. 2012 and Luscher and Malenka 2012).


##### Edit history

| Date | Action | Author |
|---|---|---|
| 2018-09-21 | Created | Orlic-Milacic M |
| 2018-10-10 | Authored | Orlic-Milacic M |
| 2018-11-02 | Reviewed | Hansen KB, Yi F |
| 2018-11-07 | Edited | Orlic-Milacic M |
| 2021-05-31 | Modified | Shorser S |

##### Entities found in this pathway (3)

| Input | UniProt Id | Input | UniProt Id | Input | UniProt Id |
|---|---|---|---|---|---|
| GRIA1 | P42261 | GRIA2 | P42262 | GRIN2B | Q13224 |

#### 14. Regulation of commissural axon pathfinding by SLIT and ROBO (R-HSA-428542)

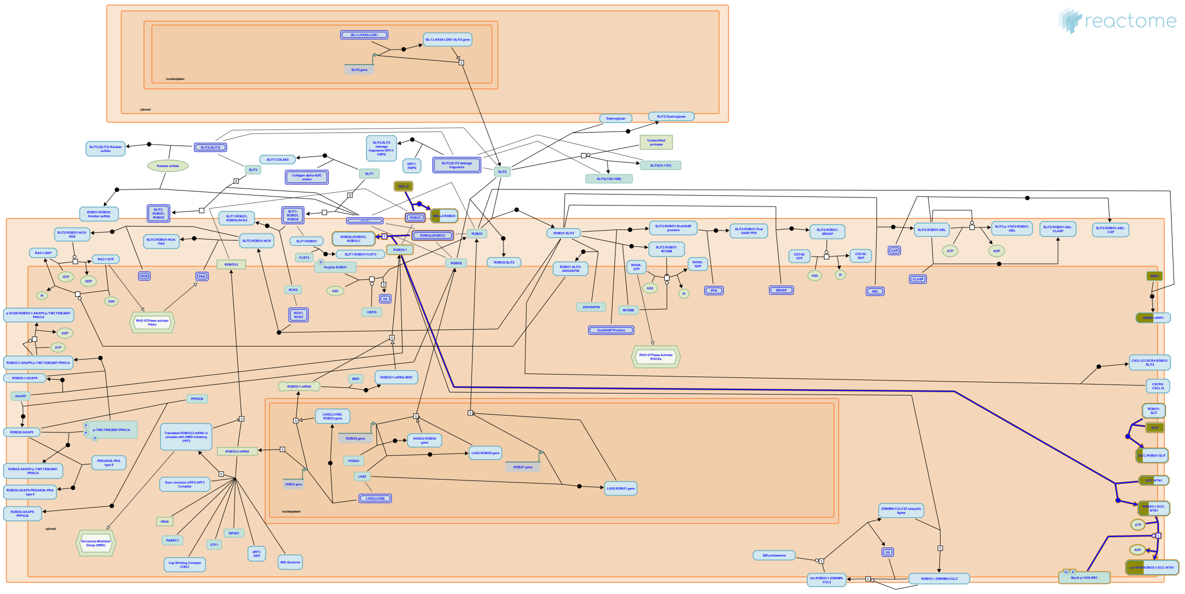

Commissural axons project to the floor plate, attracted by the interaction of their DCC receptors with Netrin-1 (NTN1) produced by floor plate cells (Dickson and Gilestro 2006) and radial glia (Dominici et al. 2017, Varadarajan et al. 2017). Once an axon enters the floor plate, it must be efficiently expelled on the contralateral side. A switch from attraction to repulsion allows commissural axons to enter and then leave the CNS midline. Based on studies in Xenopus neurons and by yeast two hybrid screens, it is observed that the attractive response of axons to netrins is silenced by activation of ROBO. SLIT bound ROBO binds to DCC, preventing it from transducing an attractive response to netrin. The sensitivity of axons to the repulsive action of SLIT does not only depend on repulsive SLIT receptors (ROBO1 and ROBO2), but is also influenced by expression of ROBO3, a SLIT receptor that suppresses the activity of ROBO1 and ROBO2. Upon crossing the midline, commissural axons downregulate expression of ROBO3 and increase expression of ROBO1/ROBO2 (reviewed by Dickson and Gilestro, 2006). Two transcript variants of ROBO3, ROBO3.1 and ROBO3.2 are considered to play different roles in midline crossing. ROBO3.1 is expressed in the pre-crossing and crossing commissural axons, while ROBO3.2, generated by alternative splicing, is expressed after midline crossing and thought to block midline re-crossing (Chen et al. 2008). In addition to SLITs, a secreted ligand NELL2 also acts as an axonal guidance cue that, by acting through ROBO3 receptors, helps to steer commissural axons to the midline. Both ROBO3.1 and ROBO3.2 can bind to a secreted ligand NELL2. Pre-crossing commissural axons, which express ROBO3.1, are repelled by NELL2. Post-crossing axons, which express ROBO3.2 are not repelled by NELL2 (Jaworski et al. 2015). .

#### Edit history

| Date | Action | Author |
|---|---|---|
| 2008-09-05 | Edited | Garapati P V |
| 2008-09-05 | Authored | Garapati P V |
| 2009-07-06 | Created | Garapati P V |
| 2009-08-18 | Reviewed | Kidd T |
| 2017-06-23 | Edited | Orlic-Milacic M |
| 2017-07-31 | Reviewed | Jaworski A |
| 2021-05-22 | Modified | Shorser S |

#### Entities found in this pathway (2)

| Input | UniProt Id | Input | UniProt Id |
|---|---|---|---|
| DCC | P43146 | NELL2 | Q99435 |

#### 15. Molecules associated with elastic fibres (R-HSA-2129379)

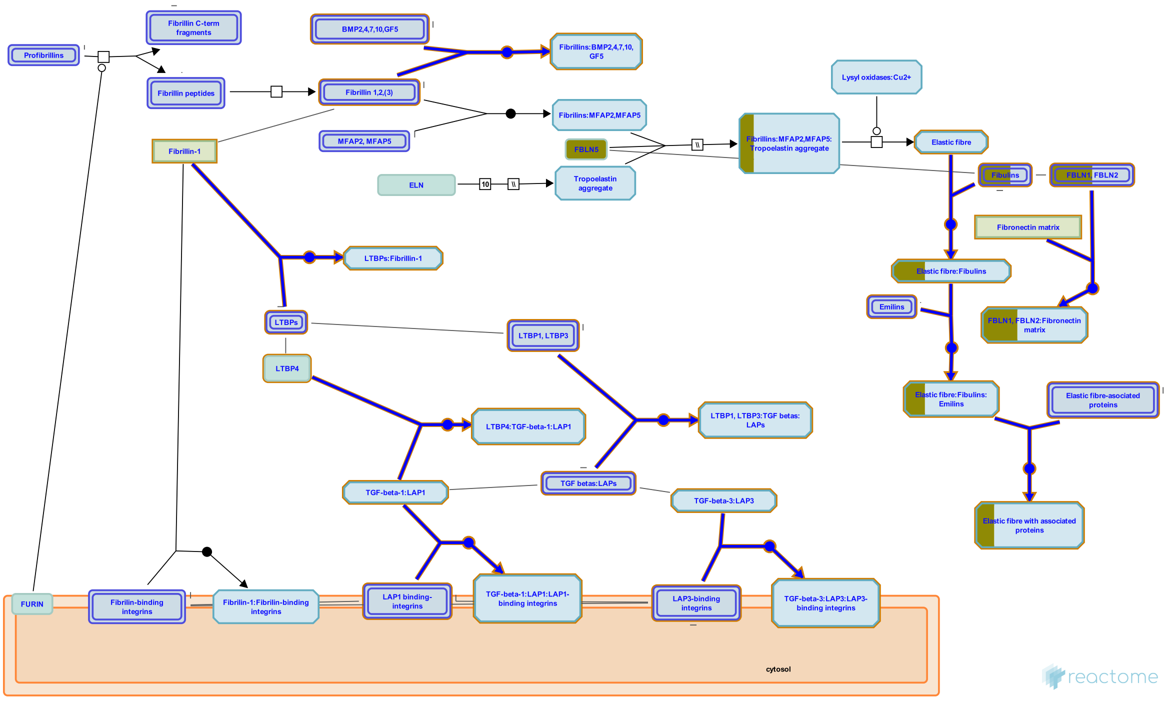


Proteins found associated with microfibrils include vitronectin (Dahlback et al. 1990), latent transforming growth factor beta-binding proteins (Kielty et al. 2002, Munger & Sheppard 2011), emilin (Bressan et al. 1993, Mongiat et al. 2000), members of the microfibrillar-associated proteins (MFAPs, Gibson et al.1996), and fibulins (Roark et al. 1995, Yanagisawa et al. 2002). The significance of these interactions is not well understood but may help mediate elastin-fibrillin interactions during elastic fibre assembly.

Proteoglycans such as versican (Isogai et al. 2002), biglycan, and decorin (Reinboth et al. 2002) can interact with the microfibrils. They confer specific properties including hydration, impact absorption, molecular sieving, regulation of cellular activities, mediation of growth factor association, and release and transport within the extracellular matrix (Buczek-Thomas et al. 2002). In addition, glycosaminoglycans have been shown to interact with tropoelastin through its lysine side chains (Wu et al. 1999) regulating tropoelastin assembly (Tu and Weiss, 2008).


##### Edit history

| Date | Action | Author |
|---|---|---|
| 2012-02-21 | Created | Jupe S |
| 2012-04-30 | Authored | Jupe S |
| 2012-11-02 | Reviewed | Muiznieks LD |
| 2012-11-12 | Edited | Jupe S |

| Date | Action | Author |
|---|---|---|
| 2021-05-22 | Modified | Shorser S |

#### Entities found in this pathway (2)

| Input | UniProt Id | Input | UniProt Id |
|---|---|---|---|
| FBLN2 | P98095 | FBLN5 | Q12805, Q9UBX5 |

#### 16. GPCR ligand binding (R-HSA-500792)

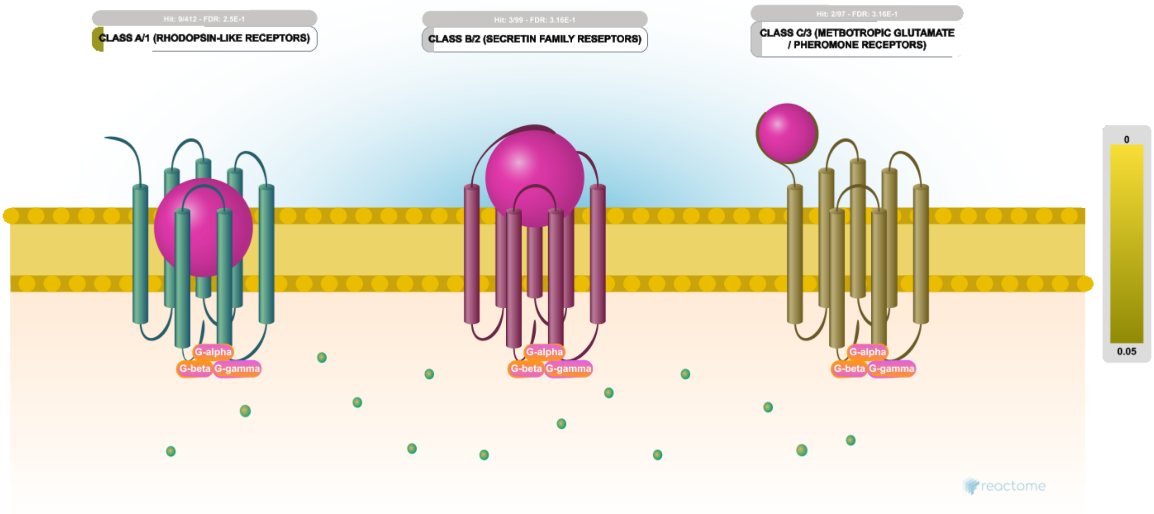


**Cellular compartments:** plasma membrane.

There are more than 800 G-protein coupled receptor (GPCRs) in the human genome, making it the largest receptor superfamily. GPCRs are also the largest class of drug targets, involved in virtually all physiological processes (Frederiksson 2003). GPCRs are receptors for a diverse range of ligands from large proteins to photons (Kristiansen et al. 2004) and have an equal diversity of ligand-binding mechanisms (Gether et al. 2002). Classical GPCR signaling involves signal transduction via heterotrimeric G-proteins, though G-protein independent mechanisms have been reported.

Rhodopsin-like receptors (class A/1) are by far the largest group of GPCRs and the best studied, though a large proportion of the functional and structural studies have focused on a very few members; many remain functionally uncharacterized. This large family can be subdivided into at least 19 subfamilies (Subfamily A1-19) based on phylogenetic analysis (Joost & Methner 2002). Family A includes receptors for a wide variety of ligands including hormones, light and neurotransmitters, encompassing a wide range of functions including many autocrine, paracrine and endocrine processes.

The secretin-like family B/2 GPCRs includes receptors for many hormone-like peptides, such as secretin, calcitonin, parathyroid hormone/parathyroid hormone-related peptides and vasoactive intestinal peptide, which activate adenylyl cyclase and the phosphatidyl-inositol-calcium pathway (Harmar 2001).

The class C/3 GPCRs include the metabotropic glutamate receptors and taste receptors (Brauner-Osborne et al. 2007). All have a large extracellular N-terminus that structurally resembles a clamshell and has an important role in ligand binding.

#### Edit history

| Date | Action | Author |
|---|---|---|
| 2009-12-12 | Reviewed | D'Eustachio P |
| 2010-02-05 | Authored | Jassal B |
| 2010-02-05 | Created | Jassal B |
| 2010-02-10 | Edited | Jupe S |
| 2021-05-22 | Modified | Shorser S |

#### Entities found in this pathway (13)

| Input | UniProt Id | Input | UniProt Id | Input | UniProt Id |
|---|---|---|---|---|---|
| ADRA1A | P25100, P35348 | ADRA1B | P35368 | C5AR2 | Q9P296 |
| FZD1 | Q9UP38 | FZD7 | O75084 | GNG2 | P59768 |
| GRM2 | Q14416 | GRM7 | Q14831 | LPPR4 | Q7Z2D5 |
| NPY2R | P49146 | P2RY13 | Q9BPV8 | PDYN | P01213 |
| SSTR2 | P30874 | | | | |

#### 17. Signaling by GPCR (R-HSA-372790)

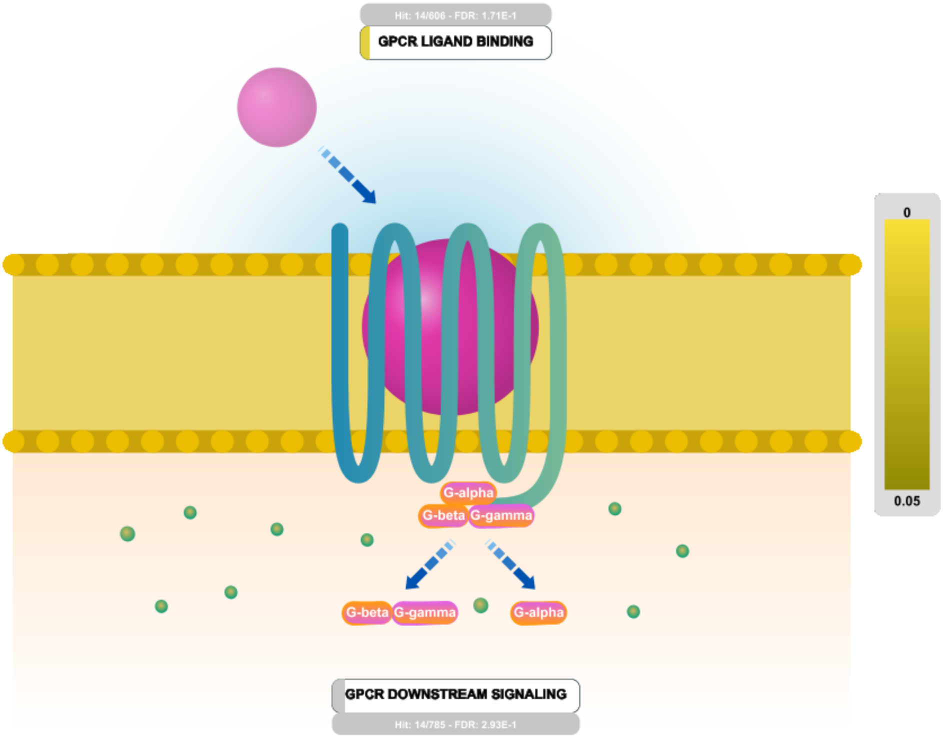


G protein-coupled receptors (GPCRs; 7TM receptors; seven transmembrane domain receptors; heptahelical receptors; G protein-linked receptors [GPLR]) are the largest family of transmembrane receptors in humans, accounting for more than 1% of the protein-coding capacity of the human genome. All known GPCRs share a common architecture of seven membrane-spanning helices connected by intra- and extracellular loops. The extracellular loops contain two highly-conserved cysteine residues that form disulphide bonds to stabilize the structure of the receptor. They recognize diverse messengers such as light, odorants, small molecules, hormones and neurotransmitters. Most GPCRs act as guanine nucleotide exchange factors; activated by ligand binding, they promote GDP-GTP exchange on associated heterotrimeric guanine nucleotide-binding (G) proteins. There are two models for GPCR-G Protein interactions: 1) ligand-GPCR binding first, then binding to G Proteins; 2) "Pre-coupling" of GPCRs and G Proteins before ligand binding (review Oldham WM and Hamm HE, 2008). These in turn activate effector enzymes or ion channels. GPCRs are involved in a range of physiological roles which include the visual sense, smell, behavioural regulation, functions of the autonomic nervous system and regulation of the immune system and inflammation.

GPCRs are divided into classes based on sequence homology and functional similarity. The main mammalian classes, in order of size, are the Rhodopsin-like family A, the Secretin receptor family B, and the Metabotropic glutamate/pheromone receptor family C.

#### Edit history

| Date | Action | Author |
|---|---|---|
| 2008-07-02 | Authored | Jassal B |
| 2008-07-02 | Created | Jassal B |
| 2008-09-01 | Edited | D'Eustachio P |
| 2008-09-01 | Reviewed | Bockaert J |
| 2021-05-22 | Modified | Shorser S |

#### Entities found in this pathway (17)

| Input | UniProt Id | Input | UniProt Id | Input | UniProt Id |
|---|---|---|---|---|---|
| ADRA1A | P25100, P35348 | ADRA1B | P35368 | ARHGEF3 | Q9NR81 |
| C5AR2 | Q9P296 | FZD1 | Q9UP38 | FZD7 | O75084 |
| GNG2 | P59768 | GRM2 | Q14416 | GRM7 | Q14831 |
| LPPR4 | Q7Z2D5 | NBEA | Q8NFP9 | NPY2R | P49146 |
| P2RY13 | Q9BPV8 | PDYN | P01213 | PRKCH | P24723 |
| RASGRP1 | O95267 | SSTR2 | P30874 | | |

#### 18. Drug resistance of KIT mutants (R-HSA-9669937)

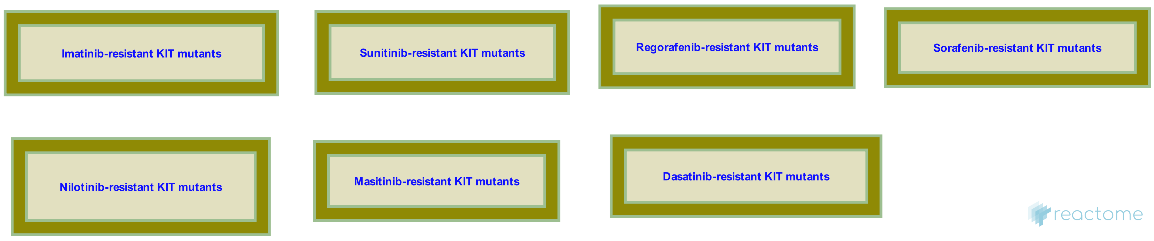


**Diseases:** cancer.

Activating mutations in the juxtamembrane domain of KIT are common in some cancers, including gastrointestinal stromal tumors, melanoma and acute myeloid leukemia (reviewed in Roskoski, 2018). These mutations are sensitive to inhibition with imatinib, which in 2001 was the first tyrosine kinase inhibitor approved for treatment of cancer (Demetri et al, 2002; Corless et al, 2011; reviewed in Zitvogel, 2016). Although highly successful in prolonging survival, imatinib-resistance develops in most patients due to appearance of secondary mutations, often in the ATP-binding pocket or in the activation loop of the kinase domain (Gajiwala et al, 2008; Serrano et al, 2019; reviewed in Roskoski, 2018; Napolitano and Vincenzi, 2019)

##### Edit history

| Date | Action | Author |
|---|---|---|
| 2019-12-03 | Created | Rothfels K |
| 2020-03-13 | Reviewed | García-Valverde A, Pilco-Janeta D, Serrano C |
| 2020-04-01 | Authored | Rothfels K |
| 2020-05-04 | Edited | Rothfels K |
| 2020-05-27 | Modified | Rothfels K |

##### Entities found in this pathway (1)

| Input | UniProt Id |
| --- | --- |
| KIT | P10721 |

#### 19. KIT mutants bind TKIs (R-HSA-9669921)

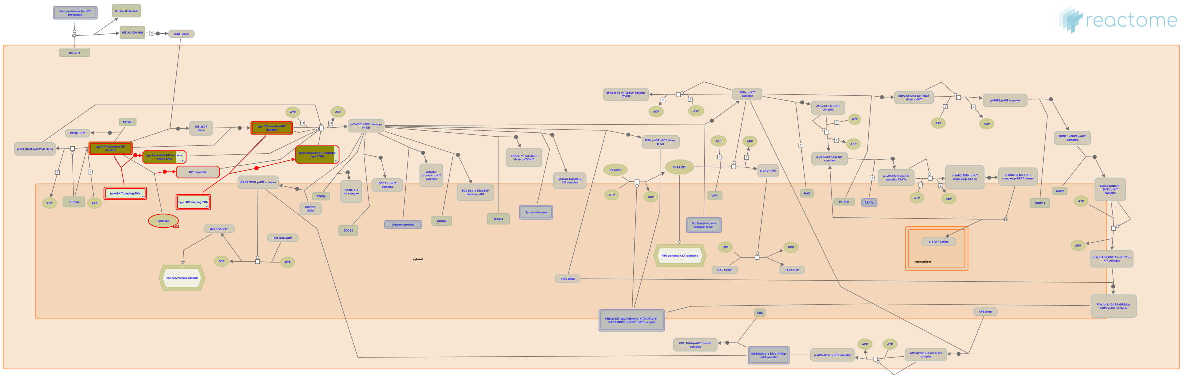

**Diseases:** cancer.

Aberrant signaling by activated forms of KIT can be inhibited by tyrosine kinase inhibitors. Primary mutations in KIT are frequently found in exon 11, encoding the juxtamembrane domain responsible for autoinhibition of the kinase. These mutations are generally sensitive to tyrosine kinase inhibitors such as imatinib. Accumulation of secondary mutations in the ATP-binding pocket and the activation loop of the kinase domain contributes to resistance to first line tyrosine kinase inhibitors. KIT receptors with in these regions are sensitive to a panel of additional tyrosine kinase inhibitors such as sunitinib and regorafenib (Serrano et al, 2019; reviewed in Roskoski, 2018; Klug et al, 2018; Serrano et al, 2017).


##### Edit history

| Date | Action | Author |
|---|---|---|
| 2019-12-03 | Created | Rothfels K |
| 2020-03-13 | Reviewed | García-Valverde A, Pilco-Janeta D, Serrano C |
| 2020-04-01 | Authored | Rothfels K |
| 2020-05-04 | Edited | Rothfels K |
| 2021-04-13 | Modified | Rothfels K |

##### Entities found in this pathway (1)

| Input | UniProt Id |
|---|---|
| KIT | P10721 |

#### 20. Imatinib-resistant KIT mutants (R-HSA-9669917)

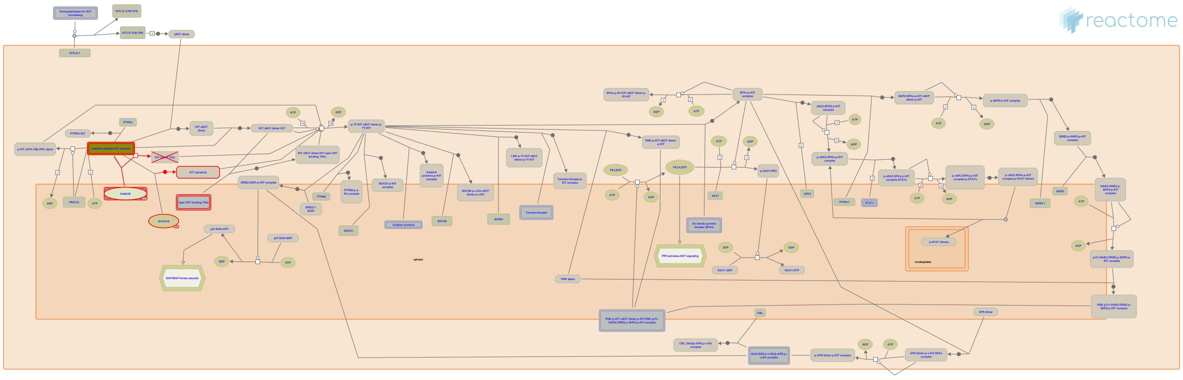

**Diseases:** cancer.

Imatinib is approved for treatment of cancers carrying primary mutations in the KIT receptor. Imatinib binds and inhibits the inactive state of the receptor, including the conformation promoted by exon 11 mutations that relieve the auto-inhibition of the WT protein. Resistance to imatinib arises due to the polyclonal expansion of subpopulations bearing secondary KIT mutations in the ATP binding pocket or the activation loop of the protein (Serrano et al, 2019; reviewed in Roskoski, 2018; Klug et al, 2018; Corless et al, 2011).


##### Edit history

| Date | Action | Author |
|---|---|---|
| 2019-12-03 | Created | Rothfels K |
| 2020-03-13 | Reviewed | García-Valverde A, Pilco-Janeta D, Serrano C |
| 2020-04-01 | Authored | Rothfels K |
| 2020-05-04 | Edited | Rothfels K |
| 2020-05-27 | Modified | Rothfels K |

##### Entities found in this pathway (1)

| Input | UniProt Id |
|---|---|
| KIT | P10721 |

#### 21. Sunitinib-resistant KIT mutants (R-HSA-9669934)

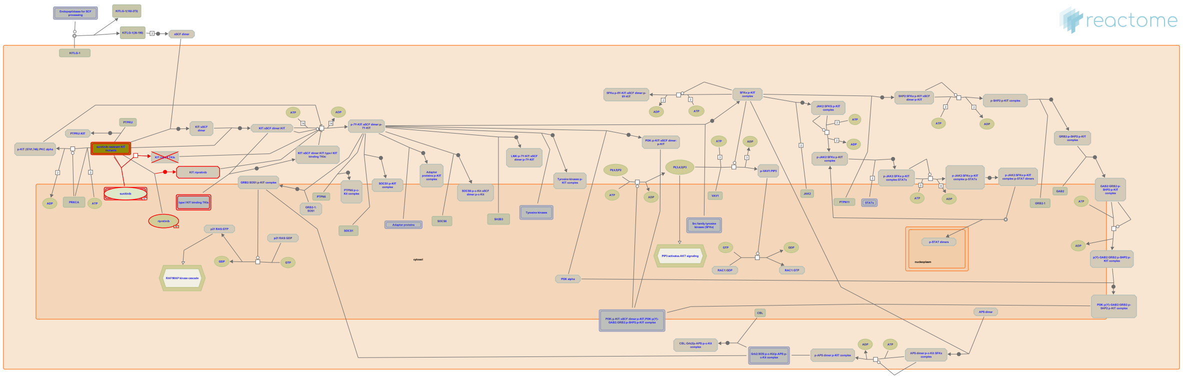

**Diseases:** cancer.

Sunitinib is a class II tyrosine kinase inhibitor that is often used as a second line treatment in KIT-mutated cancers that develop resistance to imatinib (Heinrich et al, 2008; Serrano et al, 2017; reviewed in Roskoski, 2018; Corless et al, 2011).


##### Edit history

| Date | Action | Author |
|---|---|---|
| 2019-12-03 | Created | Rothfels K |
| 2020-03-13 | Reviewed | García-Valverde A, Pilco-Janeta D, Serrano C |
| 2020-04-01 | Authored | Rothfels K |
| 2020-05-04 | Edited | Rothfels K |
| 2020-05-27 | Modified | Rothfels K |

##### Entities found in this pathway (1)

| Input | UniProt Id |
|---|---|
| KIT | P10721 |

#### 22. Sorafenib-resistant KIT mutants (R-HSA-9669936)

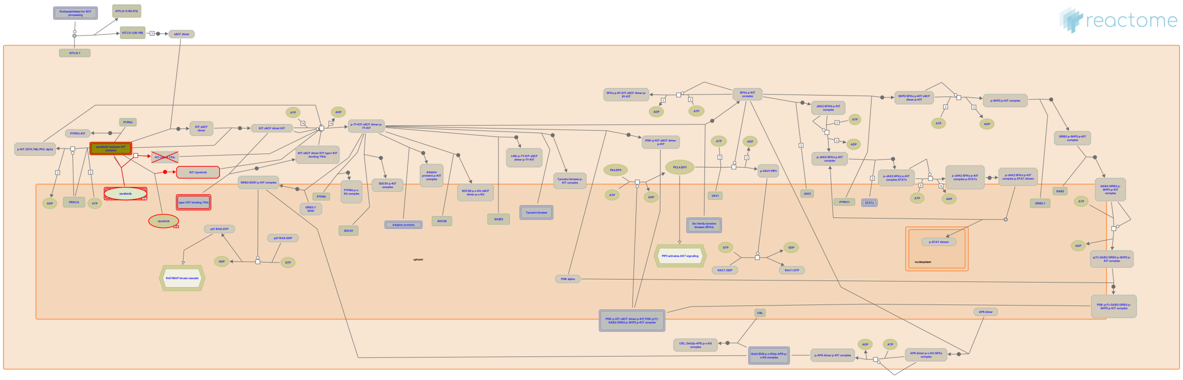

**Diseases:** cancer.

Sorafenib is a type II tyrosine kinase inhibitor that is approved for use in hepatocellular and renal cell carcinoma. It is active against KIT receptors with mutations in the ATP-binding cleft and the activation loop, with the exception of substitutions at D816, which are resistant (Guida et al, 2007; Heinrich et al, 2012; Serrano et al, 2019; Weisberg et al, 2019; reviewed in Roskoski, 2018; Klug et al, 2012).


##### Edit history

| Date | Action | Author |
|---|---|---|
| 2019-12-03 | Created | Rothfels K |
| 2020-03-13 | Reviewed | García-Valverde A, Pilco-Janeta D, Serrano C |
| 2020-04-01 | Authored | Rothfels K |
| 2020-05-04 | Edited | Rothfels K |
| 2020-05-27 | Modified | Rothfels K |

##### Entities found in this pathway (1)

| Input | UniProt Id |
|---|---|
| KIT | P10721 |

#### 23. Dasatinib-resistant KIT mutants (R-HSA-9669914)

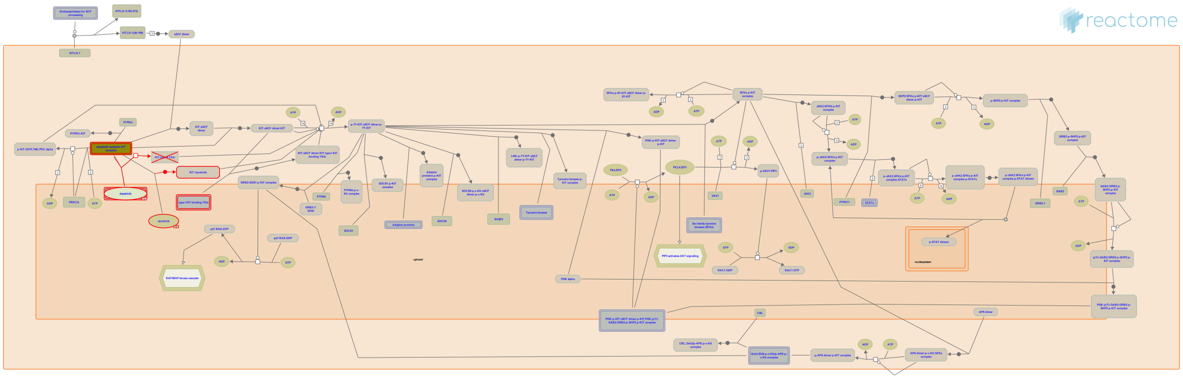

**Diseases:** cancer.

Dasatinib is a type II tyrosine kinase inhibitor that is active against KIT receptors with mutations in the juxtamembrane and activation loop domains, but shows only partial activity against KIT receptors with mutations at residue V654 (Schittenhelm et al, 2006; Serrano et al, 2019).


##### Edit history

| Date | Action | Author |
|---|---|---|
| 2019-12-03 | Created | Rothfels K |
| 2020-03-13 | Reviewed | García-Valverde A, Pilco-Janeta D, Serrano C |
| 2020-04-01 | Authored | Rothfels K |
| 2020-05-04 | Edited | Rothfels K |
| 2020-05-27 | Modified | Rothfels K |

##### Entities found in this pathway (1)

| Input | UniProt Id |
|---|---|
| KIT | P10721 |

#### 24. Nilotinib-resistant KIT mutants (R-HSA-9669926)

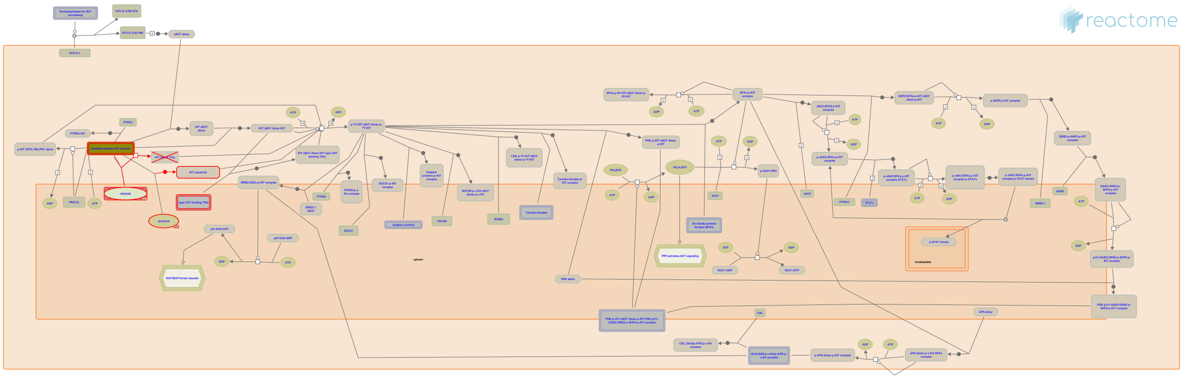

**Diseases:** cancer.

Nilotinib is a type II tyrosine kinase inhibitor currently in clinical trials for treatment of KIT-mutant cancers, and shows variable effectiveness against mutations in exon 11, 13, 17 and 18. Nilotinib is ineffective against the gatekeeper mutation T670I (Kissova et al, 2016; Guo et al, 2007; Roberts et al, 2007; Serrano et al, 2019).


##### Edit history

| Date | Action | Author |
|---|---|---|
| 2019-12-03 | Created | Rothfels K |
| 2020-03-13 | Reviewed | García-Valverde A, Pilco-Janeta D, Serrano C |
| 2020-04-01 | Authored | Rothfels K |
| 2020-05-04 | Edited | Rothfels K |
| 2020-05-27 | Modified | Rothfels K |

##### Entities found in this pathway (1)

| Input | UniProt Id |
|---|---|
| KIT | P10721 |

#### 25. Masitinib-resistant KIT mutants (R-HSA-9669924)

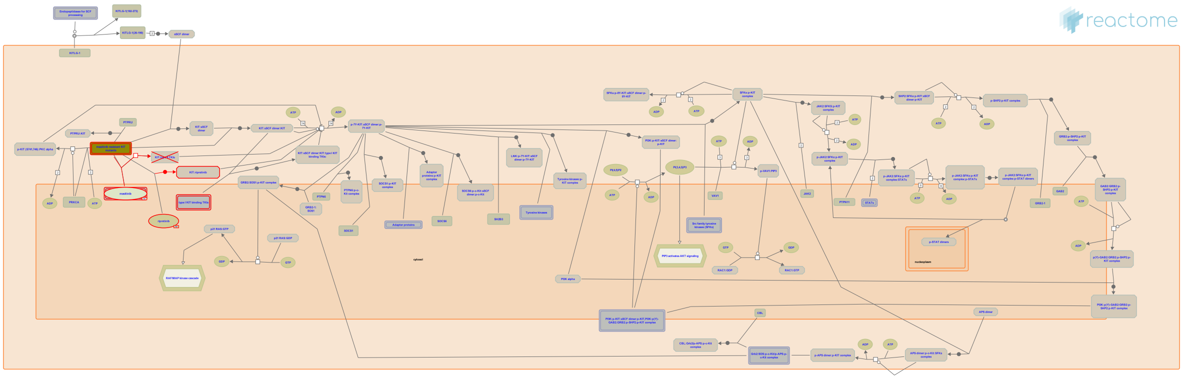

**Diseases:** cancer.

Mastinib is a class II tyrosine kinase inhibitor that targets mutant and wild-type FGFR3, PDGFR and c-KIT (Dubreuil, 2009). Masitinib, like imatinib, is effective in inhibiting the activity of juxtamembrane mutant forms of KIT, but is ineffective against many of the mutations in the activation loop and ATP-binding cleft of the receptor (Dubreuil, 2009; Serrano et al, 2019; reviewed in Demetri, 2011).


##### Edit history

| Date | Action | Author |
|---|---|---|
| 2019-12-03 | Created | Rothfels K |
| 2020-03-13 | Reviewed | García-Valverde A, Pilco-Janeta D, Serrano C |
| 2020-04-01 | Authored | Rothfels K |
| 2020-05-04 | Edited | Rothfels K |
| 2020-05-27 | Modified | Rothfels K |

##### Entities found in this pathway (1)

| Input | UniProt Id |
|---|---|
| KIT | P10721 |

### 6. Identifiers found

Below is a list of the input identifiers that have been found or mapped to an equivalent element in Reactome, classified by resource.

#### Entities (87)

| Input | UniProt Id | Input | UniProt Id | Input | UniProt Id |
|---|---|---|---|---|---|
| ABCG2 | Q9UNQ0 | ACHE | P22303 | ACVR1C | Q8NER5 |
| ADAMTS16 | Q8TE57 | ADRA1A | P25100, P35348 | ADRA1B | P35368 |
| ARHGEF28 | Q8N1W1 | ARHGEF3 | Q9NR81 | ATP8A2 | Q9NTI2 |
| B3GNT1 | O43505, Q9NY97 | BTNL9 | Q6UXG8 | C5AR2 | Q9P296 |
| CACNA2D1 | Q9UIU0 | CDC42EP3 | Q9UKI2 | CDH9 | Q9ULB4 |
| CHGB | P05060 | CHN2 | P52757 | CRYM | Q14894 |
| DCC | P43146 | DIAPH2 | O60879 | DSEL | Q8IZU8 |
| DUSP1 | P28562 | EPHA6 | Q9UF33 | EPHA7 | Q15375 |
| FBLN2 | P98095 | FBLN5 | Q12805, Q9UBX5 | FZD1 | Q9UP38 |
| FZD7 | O75084 | GABRA3 | P34903 | GABRA5 | P31644 |
| GABRQ | Q9UN88 | GALNT8 | Q9NY28 | GATA2 | P23769 |
| GNG2 | P59768 | GPD1 | P21695, Q8N335 | GRIA1 | P42261 |
| GRIA2 | P42262 | GRIN2B | Q13224 | GRM2 | Q14416 |
| GRM7 | Q14831 | HMGCR | P04035 | HS6ST3 | Q8IZP7 |
| IER3 | P46695 | IGF1 | P05019 | IL12RB2 | Q99665 |
| KCNA3 | P22001, Q09470 | KCNJ6 | P48051 | KCNK2 | O95069, Q13303 |
| KIAA0947 | Q9Y2F5 | KIT | P10721 | KL | Q9UEF7-1, Q9UEF7-2 |
| KLHL13 | Q9P2J3, Q9P2N7 | KLHL2 | O95198 | LPPR4 | Q7Z2D5 |
| LRMP | Q12912 | LRP12 | Q14117 | LRRTM2 | O43300 |
| MICA | P01893 | MMD | Q9P0K1 | NBEA | Q8NFP9 |
| NELL2 | Q99435 | NPY2R | P49146 | NRP1 | O14786 |
| OPCML | Q14982 | P2RY13 | Q9BPV8 | PDYN | P01213 |
| PIWIL2 | Q8TC59 | PLK2 | Q9NYY3 | PLXNA4 | Q9HCM2 |
| PRKCH | P24723 | PTPN14 | Q15678 | RASAL1 | O95294 |
| RASGRP1 | O95267 | RSPO3 | Q9BXY4 | SCN3B | Q9NY72 |
| SEMA5A | Q13591 | SLC1A6 | P48664 | SLC24A3 | Q9HC58 |
| SLC4A7 | Q9Y6M7 | SLC6A5 | P23975 | SMAD4 | Q13485 |
| SSTR2 | P30874 | TIE1 | Q02763 | TLL1 | O43897 |
| TRAC | P01848 | TRPC4 | Q9UBN4 | ZNF676 | Q8N7Q3 |

| Input | Ensembl Id | Input | Ensembl Id | Input | Ensembl Id |
|---|---|---|---|---|---|
| GRIA2 | ENSG00000120251 | GRIN2B | ENSG00000273079 | HMGCR | ENSG00000113161 |
| KIT | ENSG00000157404 | PLK2 | ENSG00000145632 | PLXNA4 | ENSG00000221866 |
| PTPN14 | ENSG00000152104 | RSPO3 | ENSG00000146374 | | |

### 7. Identifiers not found

These 57 identifiers were not found neither mapped to any entity in Reactome.

| | | | | | | | |
|---|---|---|---|---|---|---|---|
| AC002480.3 | ACVR1 | APOLD1 | C18orf42 | CCBE1 | CCDC3 | CCNJL | CLEC9A |
| CLSTN2 | CREG2 | CTD-2541J13.1 | DACH2 | FREM2 | GGTA1P | GPR12 | GPR26 |
| GPR85 | HRK | IER2 | IGSF10 | KIAA1244 | KIAA2022 | LINC00643 | LRRC2 |
| LRRN1 | MFSD4 | MGC4294 | MIPEP | MPEG1 | NCAM2 | NETO1 | NEUROD2 |
| NSG2 | OTUD1 | PCDH20 | PCDHGC5 | PRDM8 | RALGPS2 | RN7SL3 | RNVU1-18 |
| RP1-140K8.5 | RP11-143K11.1 | RP11-286B14.1 | RP11-2E11.9 | SEMA3C | SHISA9 | SLC7A14 | SNORD108 |
| STXBP6 | SUSD4 | SYNPR | SYT13 | SYT6 | TMTC1 | XX-CR54.3 | ZDHHC22 |
| ZDHHC23 | | | | | | | |
