## Supplementary material for "Compartment-specific total RNA profile of Hippocampal and Cortical cells from Mesial Temporal Lobe Epilepsy tissue": Other supplemental files_Reactome: HC_mTLE non-HS_Nucleus_DOWNspecific_report.pdf

### Pathway Analysis Report

This report contains the pathway analysis results for the submitted sample ''. Analysis was performed against Reactome version 77 on 14/09/2021. The web link to these results is:

https://reactome.org/PathwayBrowser/#/ANALYSIS=MjAyMTA5MTQwODM2MDFfNTM4MDY%3D

Please keep in mind that analysis results are temporarily stored on our server. The storage period depends on usage of the service but is at least 7 days. As a result, please note that this URL is only valid for a limited time period and it might have expired.

### Table of Contents


1. Introduction
2. Properties
3. Genome-wide overview
4. Most significant pathways
5. Pathways details
6. Identifiers found
7. Identifiers not found

### 1. Introduction

Reactome is a curated database of pathways and reactions in human biology. Reactions can be considered as pathway 'steps'. Reactome defines a 'reaction' as any event in biology that changes the state of a biological molecule. Binding, activation, translocation, degradation and classical biochemical events involving a catalyst are all reactions. Information in the database is authored by expert biologists, entered and maintained by Reactome's team of curators and editorial staff. Reactome content frequently cross-references other resources e.g. NCBI, Ensembl, UniProt, KEGG (Gene and Compound), ChEBI, PubMed and GO. Orthologous reactions inferred from annotation for Homo sapiens are available for 17 non-human species including mouse, rat, chicken, puffer fish, worm, fly, yeast, rice, and Arabidopsis. Pathways are represented by simple diagrams following an SBGN-like format.

Reactome's annotated data describe reactions possible if all annotated proteins and small molecules were present and active simultaneously in a cell. By overlaying an experimental dataset on these annotations, a user can perform a pathway over-representation analysis. By overlaying quantitative expression data or time series, a user can visualize the extent of change in affected pathways and its progression. A binomial test is used to calculate the probability shown for each result, and the p-values are corrected for the multiple testing (Benjamini–Hochberg procedure) that arises from evaluating the submitted list of identifiers against every pathway.

To learn more about our Pathway Analysis, please have a look at our relevant publications:


Fabregat A, Sidiropoulos K, Garapati P, Gillespie M, Hausmann K, Haw R, … D'Eustachio P (2016). The reactome pathway knowledgebase. Nucleic Acids Research, 44(D1), D481–D487. https://doi.org/10.1093/nar/gkv1351.

Fabregat A, Sidiropoulos K, Viteri G, Forner O, Marin-Garcia P, Arnau V, … Hermjakob H (2017). Reactome pathway analysis: a high-performance in-memory approach. BMC Bioinformatics, 18.

### 2. Properties

- This is an **overrepresentation** analysis: A statistical (hypergeometric distribution) test that determines whether certain Reactome pathways are over-represented (enriched) in the submitted data. It answers the question 'Does my list contain more proteins for pathway X than would be expected by chance?' This test produces a probability score, which is corrected for false discovery rate using the Benjamani-Hochberg method.
- 42 out of 81 identifiers in the sample were found in Reactome, where 301 pathways were hit by at least one of them.
- All non-human identifiers have been converted to their human equivalent.
- This report is filtered to show only results for species 'Homo sapiens' and resource 'all resources'.
- The unique ID for this analysis (token) is MjAyMTA5MTQwODM2MDFfNTM4MDY%3D. This ID is valid for at least 7 days in Reactome's server. Use it to access Reactome services with your data.

### 3. Genome-wide overview

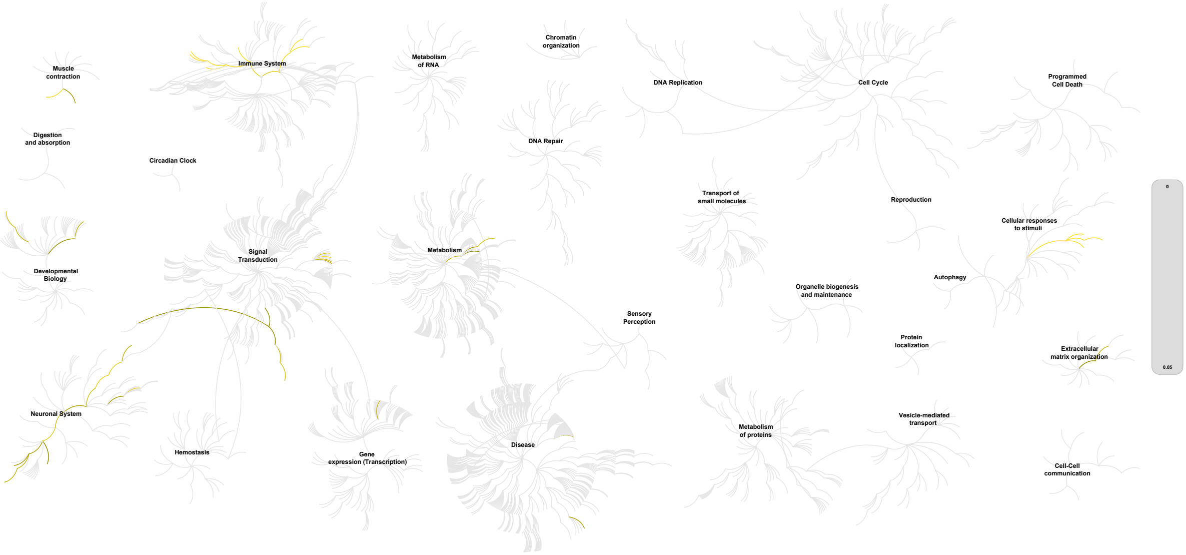


This figure shows a genome-wide overview of the results of your pathway analysis. Reactome pathways are arranged in a hierarchy. The center of each of the circular "bursts" is the root of one top-level pathway, for example "DNA Repair". Each step away from the center represents the next level lower in the pathway hierarchy. The color code denotes over-representation of that pathway in your input dataset. Light grey signifies pathways which are not significantly over-represented.

#### 4. Most significant pathways

The following table shows the 25 most relevant pathways sorted by p-value.

| Pathway name | Entities | | | | Reactions | |
|---|---|---|---|---|---|---|
| | found | ratio | p-value | FDR* | found | ratio |
| Endosomal/Vacuolar pathway | 14 / 82 | 0.006 | 4.11e-15 | 1.27e-12 | 3 / 4 | 2.96e-04 |
| Antigen Presentation: Folding, assembly and peptide loading of class I MHC | 14 / 102 | 0.007 | 7.76e-14 | 1.20e-11 | 13 / 16 | 0.001 |
| Interferon alpha/beta signaling | 15 / 186 | 0.013 | 1.72e-11 | 1.77e-09 | 2 / 22 | 0.002 |
| ER-Phagosome pathway | 14 / 173 | 0.012 | 8.12e-11 | 6.25e-09 | 3 / 10 | 7.40e-04 |
| Antigen processing-Cross presentation | 14 / 195 | 0.013 | 3.78e-10 | 2.31e-08 | 6 / 23 | 0.002 |
| Interferon gamma signaling | 15 / 250 | 0.017 | 9.72e-10 | 4.96e-08 | 2 / 16 | 0.001 |
| Immunoregulatory interactions between a Lymphoid and a non-Lymphoid cell | 15 / 316 | 0.022 | 2.16e-08 | 9.51e-07 | 8 / 44 | 0.003 |
| Attenuation phase | 7 / 47 | 0.003 | 9.33e-08 | 3.54e-06 | 3 / 5 | 3.70e-04 |
| Interferon Signaling | 15 / 394 | 0.027 | 3.63e-07 | 1.23e-05 | 4 / 69 | 0.005 |
| HSF1-dependent transactivation | 7 / 59 | 0.004 | 4.25e-07 | 1.28e-05 | 4 / 8 | 5.92e-04 |
| Class I MHC mediated antigen processing & presentation | 14 / 473 | 0.033 | 1.60e-05 | 4.49e-04 | 19 / 48 | 0.004 |
| HSF1 activation | 5 / 43 | 0.003 | 2.23e-05 | 5.58e-04 | 1 / 7 | 5.18e-04 |
| Regulation of HSF1-mediated heat shock response | 7 / 113 | 0.008 | 2.87e-05 | 6.60e-04 | 7 / 14 | 0.001 |
| RHO GTPases activate CIT | 4 / 23 | 0.002 | 3.27e-05 | 7.20e-04 | 6 / 6 | 4.44e-04 |
| Cellular response to heat stress | 7 / 135 | 0.009 | 8.69e-05 | 0.002 | 12 / 29 | 0.002 |
| Smooth Muscle Contraction | 4 / 49 | 0.003 | 5.79e-04 | 0.011 | 5 / 11 | 8.14e-04 |
| Muscle contraction | 6 / 213 | 0.015 | 0.006 | 0.104 | 10 / 42 | 0.003 |
| GABA B receptor activation | 3 / 49 | 0.003 | 0.006 | 0.104 | 9 / 9 | 6.66e-04 |
| Activation of GABAB receptors | 3 / 49 | 0.003 | 0.006 | 0.104 | 8 / 8 | 5.92e-04 |
| CREB1 phosphorylation through the activation of Adenylate Cyclase | 2 / 17 | 0.001 | 0.008 | 0.115 | 4 / 6 | 4.44e-04 |
| Cytokine Signaling in Immune system | 16 / 1,092 | 0.075 | 0.009 | 0.124 | 7 / 708 | 0.052 |
| Potassium Channels | 4 / 107 | 0.007 | 0.009 | 0.124 | 5 / 19 | 0.001 |
| Adaptive Immune System | 15 / 1,004 | 0.069 | 0.01 | 0.124 | 27 / 264 | 0.02 |
| Neuronal System | 9 / 487 | 0.033 | 0.013 | 0.144 | 30 / 216 | 0.016 |
| PKA activation in glucagon signalling | 2 / 23 | 0.002 | 0.014 | 0.144 | 2 / 2 | 1.48e-04 |

* False Discovery Rate

### 5. Pathways details

For every pathway of the most significant pathways, we present its diagram, as well as a short summary, its bibliography and the list of inputs found in it.

#### 1. Endosomal/Vacuolar pathway (R-HSA-1236977)

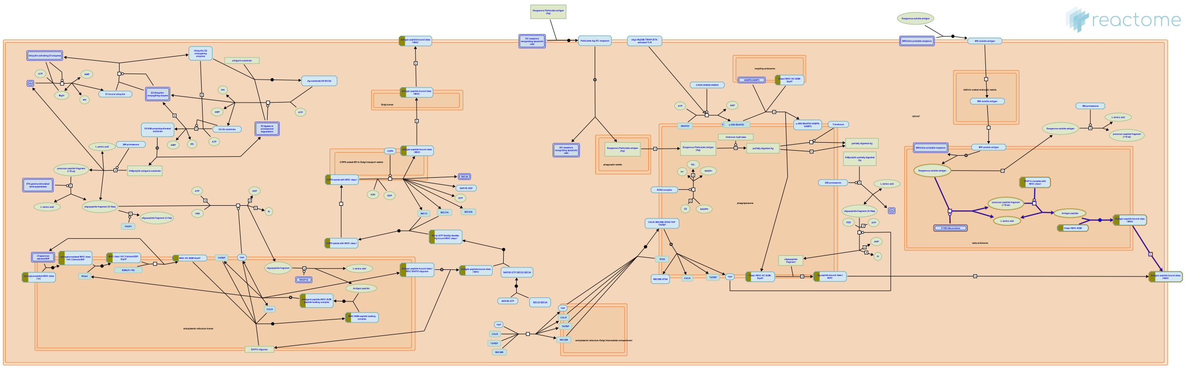

**Cellular compartments:** early endosome.

Some antigens are cross-presented through a vacuolar mechanism that involves generation of antigenic peptides and their loading on to MHC-I molecules within the endosomal compartment in a proteasome and TAP-independent manner. Antigens within the endosome are processed by cathepsin S and other proteases into antigenic peptides. Loading of these peptides onto MHC-I molecules occurs directly within early and late endosomal compartments. Why certain antigens are cross-presented exclusively by the cytosolic pathway while others use the vacuolar pathway is unknown. It may be because some epitopes cannot be generated by endosomal proteolysis, or are completely destroyed. Alternatively, the physical form of the antigen may influence its accessibility to the endosomal or vacuolar pathways (Shen et al. 2004).


##### Edit history

| Date | Action | Author |
|---|---|---|
| 2011-03-28 | Edited | Garapati P V |
| 2011-03-28 | Authored | Garapati P V |
| 2011-03-28 | Created | Garapati P V |
| 2011-05-13 | Reviewed | Desjardins M, English L |
| 2021-05-21 | Modified | Shorser S |

##### Entities found in this pathway (1)

| Input | UniProt Id |
|---|---|
| HLA-C | P04222, P10321, P30499, P30501, P30504, P30505, P30508, P30510, Q07000, Q29865, Q29960, Q29963, Q95604, Q9TNN7 |

#### 2. Antigen Presentation: Folding, assembly and peptide loading of class I MHC (R-HSA-983170)

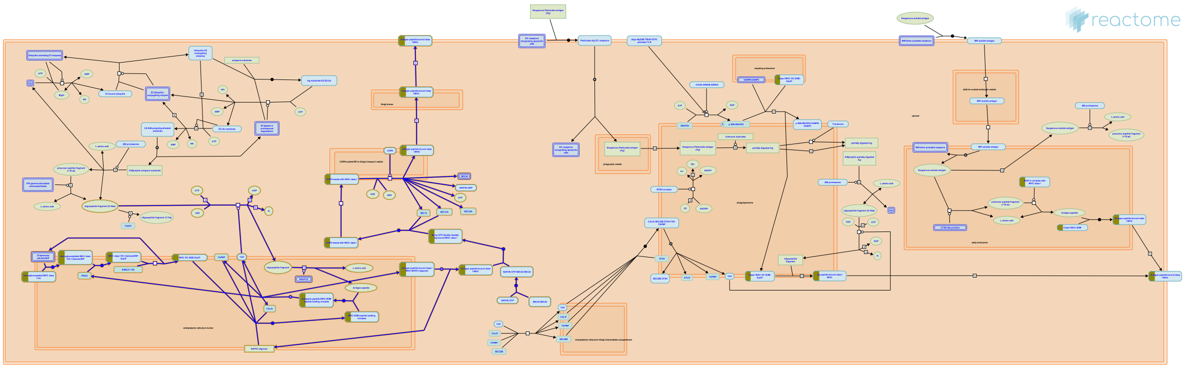

Unlike other glycoproteins, correct folding of MHC class I molecules is not sufficient to trigger their exit from the ER, they exit only after peptide loading. Described here is the process of antigen presentation which consists of the folding, assembly, and peptide loading of MHC class I molecules. The newly synthesized MHC class I Heavy Chain (HC) is initially folded with the help of several chaperones (calnexin, BiP, ERp57) and then binds with Beta-2-microglobulin (B2M). This MHC:B2M heterodimer enters the peptide loading complex (PLC), a multiprotein complex that includes calreticulin, endoplasmic reticulum resident protein 57 (ERp57), transporter associated with antigen processing (TAP) and tapasin. Peptides generated from Ub-proteolysis are transported into the ER through TAP. These peptides are further trimmed by ER-associated aminopeptidase (ERAP) and loaded on to MHC class I molecules. Stable MHC class I trimers with high-affinity peptide are transported from the ER to the cell surface by the Golgi apparatus.


##### Edit history

| Date | Action | Author |
|---|---|---|
| 2010-10-29 | Edited | Garapati P V |
| 2010-10-29 | Authored | Garapati P V |
| 2010-10-29 | Created | Garapati P V |
| 2011-02-11 | Reviewed | Elliott T |
| 2021-05-22 | Modified | Shorser S |

#### Entities found in this pathway (1)

| Input | UniProt Id |
|---|---|
| HLA-C | P04222, P10321, P30499, P30501, P30504, P30505, P30508, P30510, Q07000, Q29865, Q29960, Q29963, Q95604, Q9TNN7 |

#### 3. Interferon alpha/beta signaling (R-HSA-909733)

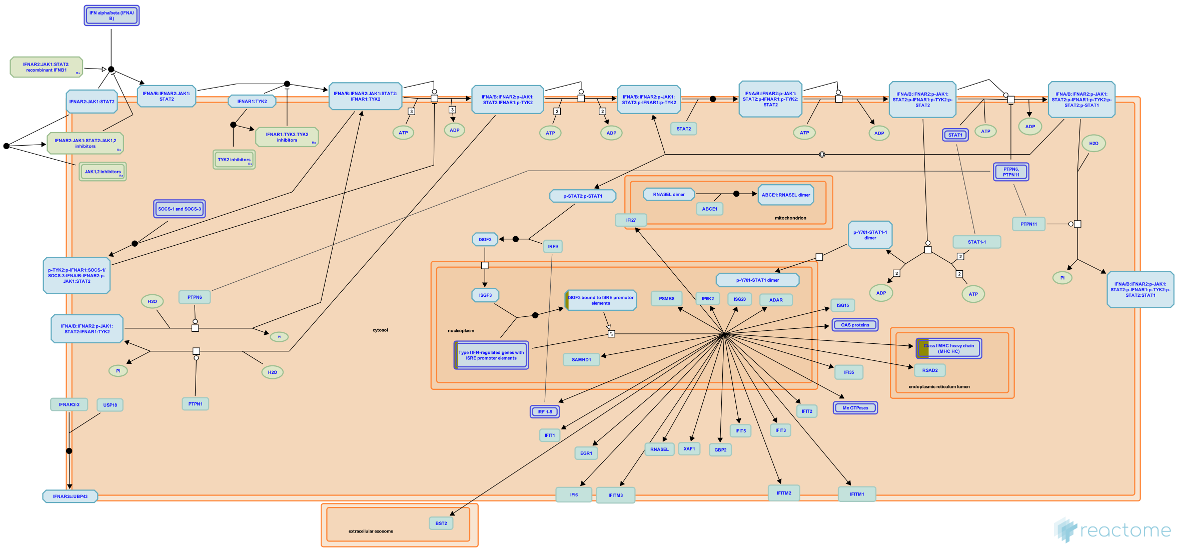

Type I interferons (IFNs) are composed of various genes including IFN alpha (IFNA), beta (IFNB), omega, epsilon, and kappa. In humans the IFNA genes are composed of more than 13 subfamily genes, whereas there is only one IFNB gene. The large family of IFNA/B proteins all bind to a single receptor which is composed of two distinct chains: IFNAR1 and IFNAR2. The IFNA/B stimulation of the IFNA receptor complex leads to the formation of two transcriptional activator complexes: IFNA-activated-factor (AAF), which is a homodimer of STAT1 and IFN-stimulated gene factor 3 (ISGF3), which comprises STAT1, STAT2 and a member of the IRF family, IRF9/P48. AAF mediates activation of the IRF-1 gene by binding to GAS (IFNG-activated site), whereas ISGF3 activates several IFN-inducible genes including IRF3 and IRF7.


##### Edit history

| Date | Action | Author |
|---|---|---|
| 2010-07-07 | Edited | Garapati P V |
| 2010-07-07 | Authored | Garapati P V |
| 2010-07-07 | Created | Garapati P V |
| 2010-08-17 | Reviewed | Abdul-Sater AA, Schindler C |

| Date | Action | Author |
|---|---|---|
| 2021-05-22 | Modified | Shorser S |

#### Entities found in this pathway (1)

| Input | UniProt Id |
|---|---|
| HLA-C | P04222, P10321, P30499, P30501, P30504, P30505, P30508, P30510, Q07000, Q29865, Q29960, Q29963, Q95604, Q9TNN7 |

| Input | Ensembl Id |
|---|---|
| HLA-C | ENSG00000204525 |

#### 4. ER-Phagosome pathway (R-HSA-1236974)

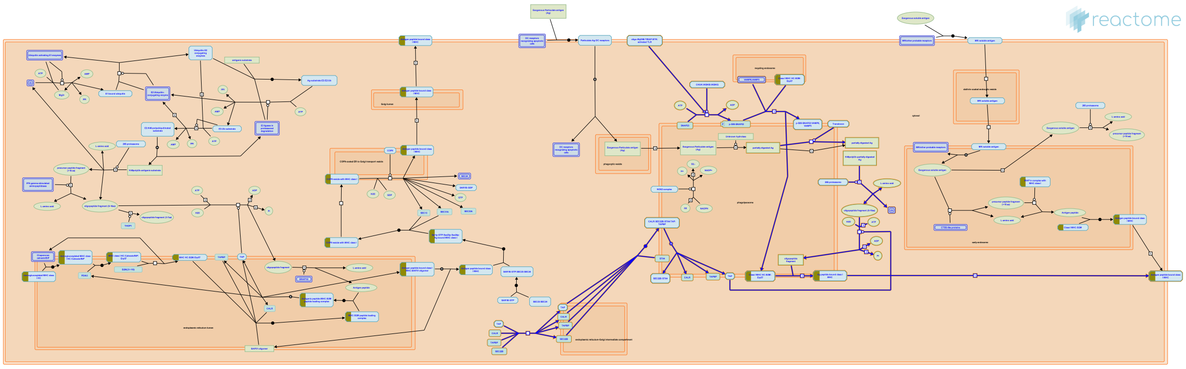

The other TAP-dependent cross-presentation mechanism in phagocytes is the endoplasmic reticulum (ER)-phagosome model. Desjardins  proposed that ER is recruited to the cell surface, where it fuses with the plasma membrane, underneath phagocytic cups, to supply membrane for the formation of nascent phagosomes (Gagnon et al. 2002). Three independent studies simultaneously showed that ER contributes to the vast majority of phagosome membrane (Guermonprez et al. 2003, Houde et al. 2003, Ackerman et al. 2003). The composition of early phagosome membrane contains ER-resident proteins, the components required for cross-presentation. This model is similar to the phagosome-to-cytosol model in that Ag is translocated to cytosol for proteasomal degradation, but differs in that antigenic peptides are translocated back into the phagosome (instead of ER) for peptide:MHC-I complexes. ER fusion with phagosome introduces molecules that are involved in Ag transport to cytosol (Sec61) and proteasome-generated peptides back into the phagosome (TAP) for loading onto MHC-I.

Although the ER-phagosome pathway is controversial, the concept remains attractive as it explains how peptide-receptive MHC-I molecules could intersect with a relatively high concentration of exogenous antigens, presumably a crucial prerequisite for efficient cross-presentation (Basha et al. 2008).


##### Edit history

| Date | Action | Author |
|---|---|---|
| 2011-03-28 | Edited | Garapati P V |
| 2011-03-28 | Authored | Garapati P V |
| 2011-03-28 | Created | Garapati P V |
| 2011-05-13 | Reviewed | Desjardins M, English L |
| 2016-05-16 | Reviewed | Bergeron JJ |
| 2021-05-21 | Modified | Shorser S |

#### Entities found in this pathway (1)

| Input | UniProt Id |
|---|---|
| HLA-C | P04222, P10321, P30499, P30501, P30504, P30505, P30508, P30510, Q07000, Q29865, Q29960, Q29963, Q95604, Q9TNN7 |

#### 5. Antigen processing-Cross presentation (R-HSA-1236975)

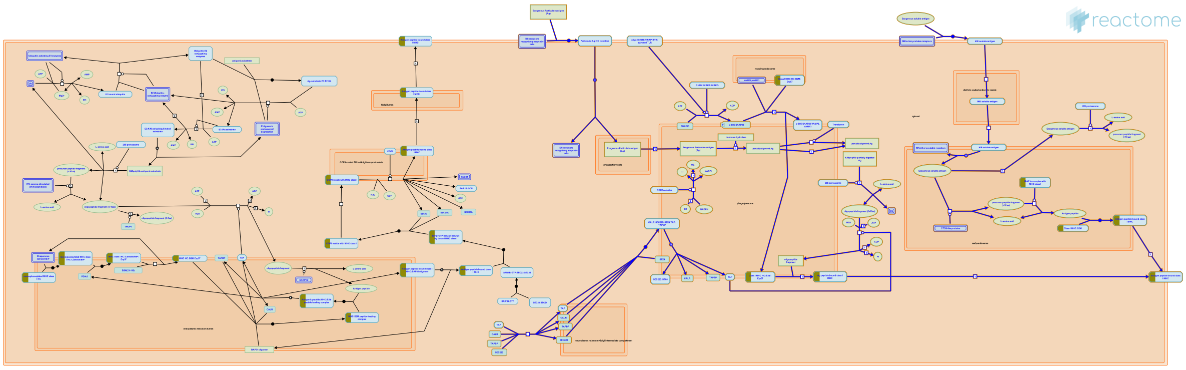

MHC class I molecules generally present peptide antigens derived from proteins synthesized by the cell itself to CD8+ T cells. However, in some circumstances, antigens from extracellular environment can be presented on MHC class I to stimulate CD8+ T cell immunity, a process termed cross-presentation (Rock & Shen. 2005). Cross-presentation/cross-priming is the ability of antigen presenting cells (APCs) to present exogenous antigens on MHC class I molecules to CD8+ T lymphocytes. Among all the APCs, Dendritic cells (DC) are the dominant antigen cross presenting cell types in vivo, although macrophages and B cells appear to cross present model antigens in vitro with a low degree of efficiency (Amigorena & Savina. 2010, Ackermann & Peter Cresswell. 2004). Compared to macrophages, DCs have low levels of lysosomal proteases and exhibit limited lysosomal degradation (Delamarre et al. 2005). This limited proteolysis of internalized antigens by DCs might contribute to their high efficiency for cross presentation (Monua & Trombetta. 2007). APCs acquire the exogenous antigens through endocytic mechanisms, especially phagosomes for particulate/cell-associated antigens and endosomes for soluble protein antigens. There does not seem to be a unique pathway for cross-presentation but rather different potential mechanisms of cross-presentation have been proposed. These proposed pathways can be classified according to the location where two key events occur: 1) processing of the antigenic protein and 2) loading of the processed peptide on to MHC I molecule (Blanchard & Shastri. 2010). Based on the requirement for TAP and cytosolic proteases two mechanisms have been described, a cytosolic pathway (TAP-dependent and proteasome-dependent) or a vacuolar pathway (TAP- and proteasome-independent) (Blanchard & Shastri. 2010, Amigorena & Savina. 2010). Regarding peptide-loading, MHC I could be loaded in the ER or in the phagosome and recycled to cell surface (Blanchard & Shastri. 2010). Exogenous soluble antigens are cross-presented by dendritic cells, albeit with lower efficiency than for particulate substrates. Soluble antigens destined for cross-presentation are taken up by distinct endocytosis mechanisms which route them into stable early endosomes and then to the cytoplasm for proteasomal degradation and peptide loading. The outcome of the cross presentation can be either tolerance or immunity (Rock & Shen. 2005).

#### Edit history

| Date | Action | Author |
|---|---|---|
| 2011-03-28 | Edited | Garapati P V |
| 2011-03-28 | Authored | Garapati P V |
| 2011-03-28 | Created | Garapati P V |
| 2011-05-13 | Reviewed | Desjardins M, English L |
| 2021-05-22 | Modified | Shorser S |

#### Entities found in this pathway (1)

| Input | UniProt Id |
|---|---|
| HLA-C | P04222, P10321, P30499, P30501, P30504, P30505, P30508, P30510, Q07000, Q29865, Q29960, Q29963, Q95604, Q9TNN7 |

#### 6. Interferon gamma signaling (R-HSA-877300)

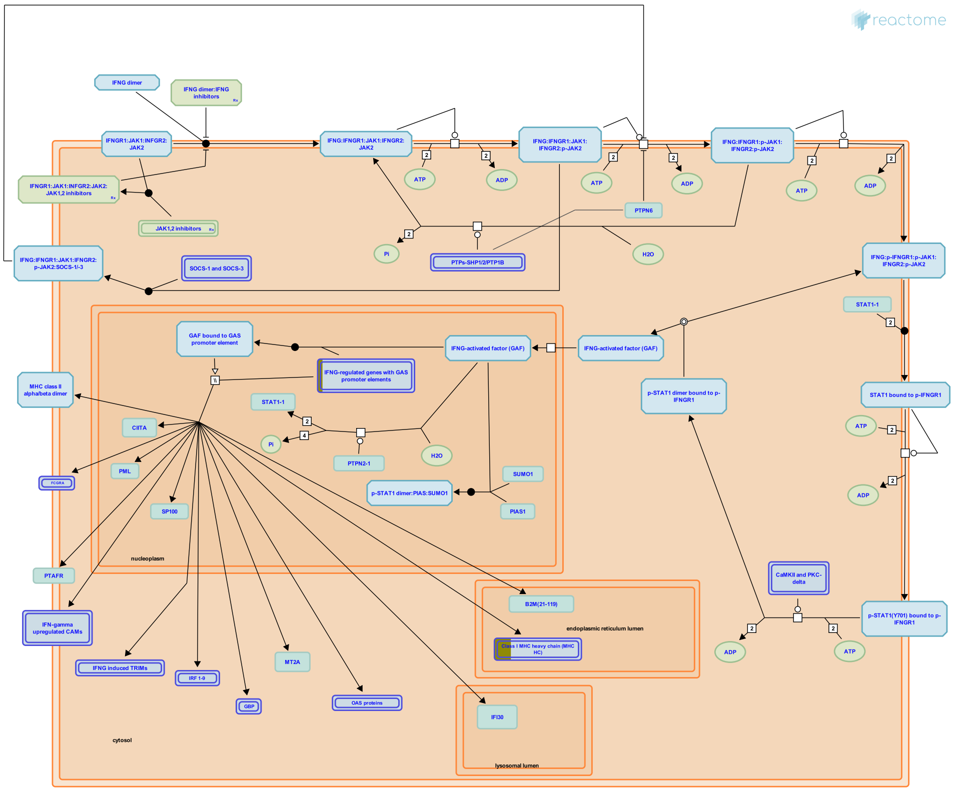


Interferon-gamma (IFN-gamma) belongs to the type II interferon family and is secreted by activated immune cells-primarily T and NK cells, but also B-cells and APC. INFG exerts its effect on cells by interacting with the specific IFN-gamma receptor (IFNGR). IFNGR consists of two chains, namely IFNGR1 (also known as the IFNGR alpha chain) and IFNGR2 (also known as the IFNGR beta chain). IFNGR1 is the ligand binding receptor and is required but not sufficient for signal transduction, whereas IFNGR2 do not bind IFNG independently but mainly plays a role in IFNG signaling and is generally the limiting factor in IFNG responsiveness. Both IFNGR chains lack intrinsic kinase/phosphatase activity and thus rely on other signaling proteins like Janus-activated kinase 1 (JAK1), JAK2 and Signal transducer and activator of transcription 1 (STAT-1) for signal transduction. IFNGR complex in its resting state is a preformed tetramer and upon IFNG association undergoes a conformational change. This conformational change induces the phosphorylation and activation of JAK1, JAK2, and STAT1 which in turn induces genes containing the gamma-interferon activation sequence (GAS) in the promoter.

#### Edit history

| Date | Action | Author |
|---|---|---|
| 2010-06-08 | Edited | Garapati P V |
| 2010-06-08 | Authored | Garapati P V |
| 2010-06-11 | Created | Garapati P V |
| 2010-08-17 | Reviewed | Abdul-Sater AA, Schindler C |
| 2021-05-31 | Modified | Shorser S |

#### Entities found in this pathway (1)

| Input | UniProt Id |
|---|---|
| HLA-C | P04222, P10321, P30499, P30501, P30504, P30505, P30508, P30510, Q07000, Q29865, Q29960, Q29963, Q95604, Q9TNN7 |

| Input | Ensembl Id |
|---|---|
| HLA-C | ENSG00000204525 |

#### 7. Immunoregulatory interactions between a Lymphoid and a non-Lymphoid cell (R-HSA-198933)

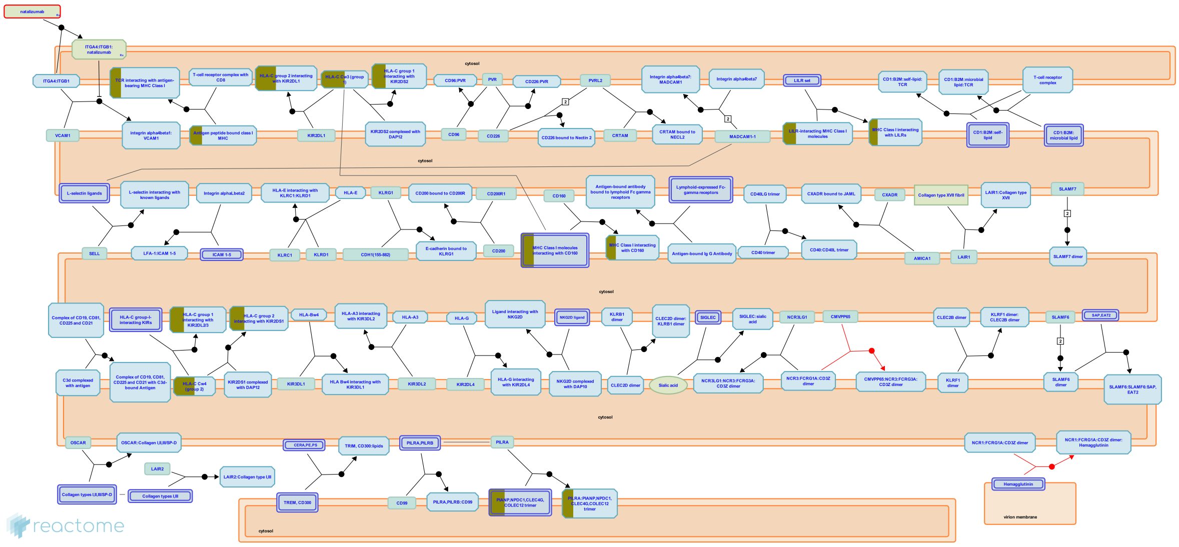

A number of receptors and cell adhesion molecules play a key role in modifying the response of cells of lymphoid origin (such as B-, T- and NK cells) to self and tumor antigens, as well as to pathogenic organisms.

Molecules such as KIRs and LILRs form part of a crucial surveillance system that looks out for any derangement, usually caused by cancer or viral infection, in MHC Class I presentation. Somatic cells are also able to report internal functional impairment by displaying surface stress markers such as MICA. The presence of these molecules on somatic cells is picked up by C-lectin NK immune receptors.

Lymphoid cells are able to regulate their location and movement in accordance to their state of activation, and home in on tissues expressing the appropriate complementary ligands. For example, lymphoid cells may fine tune the presence and concentration of adhesion molecules belonging to the IgSF, Selectin and Integrin class that interact with a number of vascular markers of inflammation.

Furthermore, there are a number of avenues through which lymphoid cells may interact with antigen. This may be presented directly to a specific T-cell receptor in the context of an MHC molecule. Antigen-antibody complexes may anchor to the cell via a small number of lymphoid-specific Fc receptors that may, in turn, influence cell function further. Activated complement factor C3d binds to both antigen and to cell surface receptor CD21. In such cases, the far-reaching influence of CD19 on B-lymphocyte function is tempered by its interaction with CD21.

#### Edit history

| Date | Action | Author |
|---|---|---|
| 2007-07-08 | Authored | de Bono B |
| 2007-07-08 | Created | de Bono B |
| 2007-08-06 | Reviewed | Trowsdale J |
| 2015-03-27 | Authored | Garapati P V |
| 2015-05-13 | Reviewed | Barrow AD |
| 2021-05-22 | Modified | Shorser S |

#### Entities found in this pathway (2)

| Input | UniProt Id | Input | UniProt Id |
|---|---|---|---|
| HLA-C | P04222, P10321, P30499, P30501, P30504, P30505, P30508, P30510, Q07000, Q29865, Q29960, Q29963, Q95604, Q9TNN7 | PIANP | Q8IYJ0 |

#### 8. Attenuation phase (R-HSA-3371568)

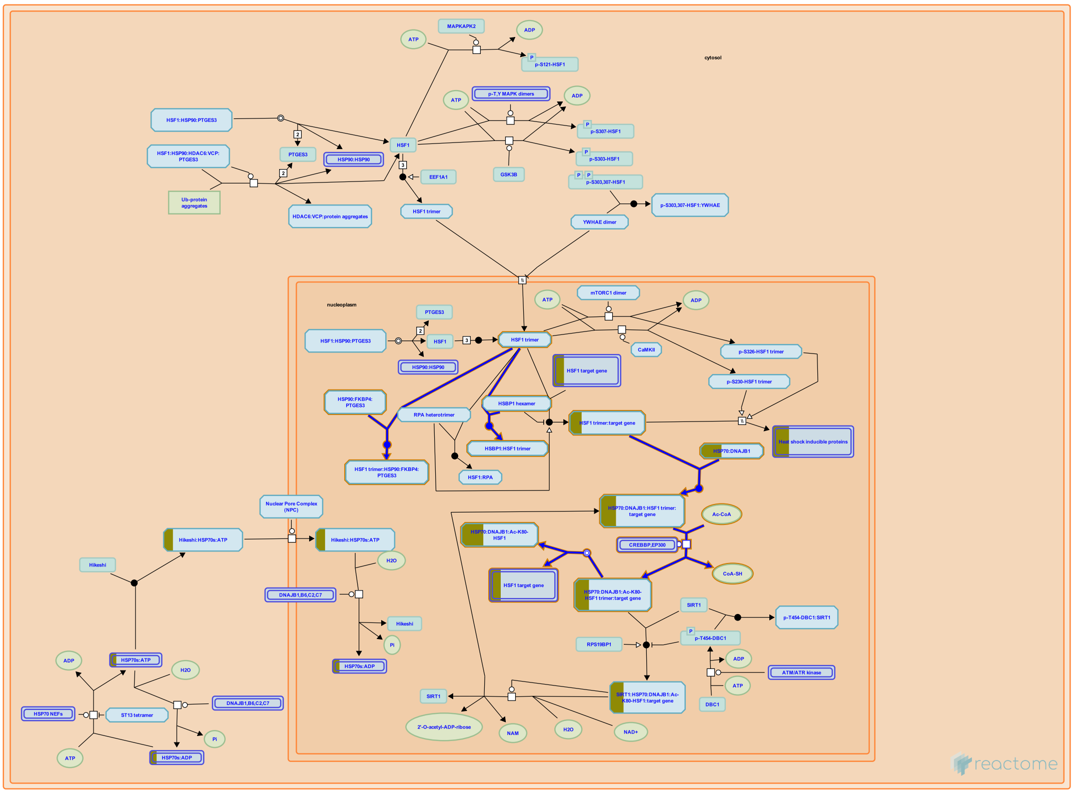

Attenuation of the heat shock transcriptional response occurs during continuous exposure to intermediate heat shock conditions or upon recovery from stress (Abravaya et al. 1991). The attenuation phase of HSF1 cycle involves the transcriptional silencing of HSF1 bound to HSE, the release of HSF1 trimers from HSE and dissociation of HSF1 trimers to monomers. HSF1-driven heat stress associated transcription was shown to depend on inducible and reversible acetylation of HSF1 at Lys80, which negatively regulates DNA binding activity of HSF1 (Westerheide SD et al. 2009). In addition, the attenuation of HSF1 activation takes place when enough HSP70/HSP40 is produced to saturate exposed hydrophobic regions of proteins damaged as a result of heat exposure. The excess HSP70/HSP40 binds to HSF1 trimer, which leads to its dissociation from the promoter and conversion to the inactive monomeric form (Abravaya et al. 1991; Shi Y et al. 1998). Interaction of HSP70 with the transcriptional corepressor repressor element 1-silencing transcription factor corepressor (CoREST) assists in terminating heat-shock response (Gomez AV et al. 2008). HSF1 DNA-binding and transactivation activity were also inhibited upon interaction of HSF1-binding protein (HSBP1) with active trimeric HSF1(Satyal SH et al. 1998).


##### Edit history

| Date | Action | Author |
|---|---|---|
| 2013-05-13 | Created | Shamovsky V |
| 2013-10-29 | Authored | Shamovsky V |

| Date | Action | Author |
| --- | --- | --- |
| 2014-02-17 | Edited | Shamovsky V |
| 2014-02-17 | Reviewed | Pani B |
| 2021-05-22 | Modified | Shorser S |

#### Entities found in this pathway (1)

| Input | UniProt Id |
| --- | --- |
| HSPA1B | P0DMV8, P0DMV9 |

| Input | Ensembl Id |
| --- | --- |
| HSPA1B | ENSG00000204388, ENSG00000212866, ENSG00000224501, ENSG00000231555, ENSG00000232804 |

#### 9. Interferon Signaling (R-HSA-913531)

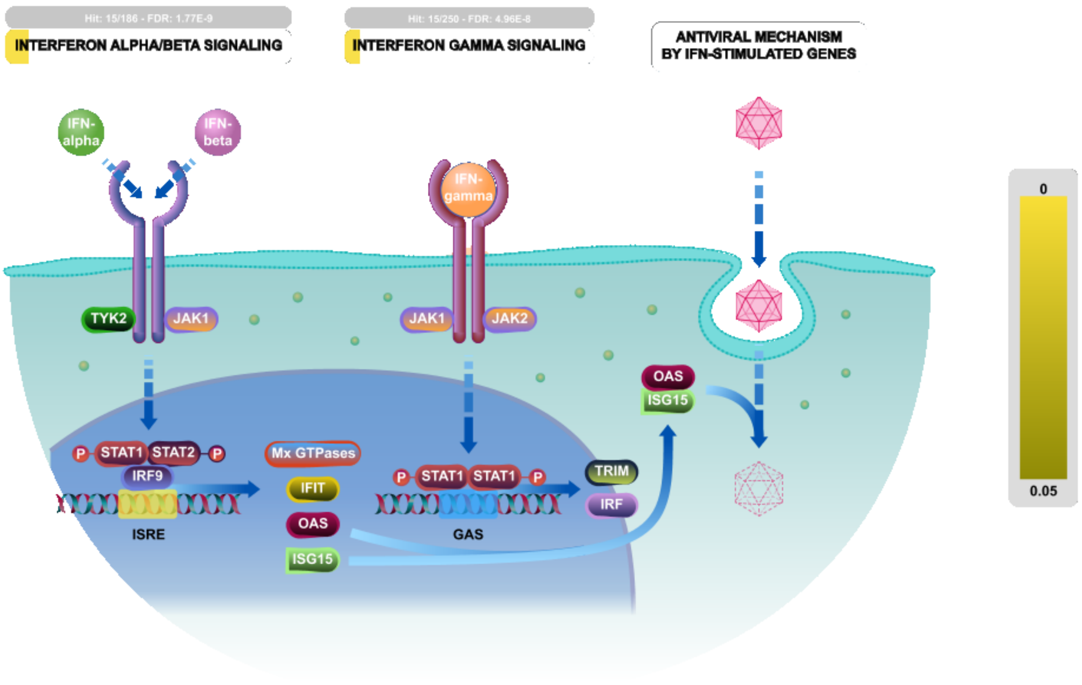


Interferons (IFNs) are cytokines that play a central role in initiating immune responses, especially antiviral and antitumor effects. There are three types of IFNs:Type I (IFN-alpha, -beta and others, such as omega, epsilon, and kappa), Type II (IFN-gamma) and Type III (IFN-lamda). In this module we are mainly focusing on type I IFNs alpha and beta and type II IFN-gamma. Both type I and type II IFNs exert their actions through cognate receptor complexes, IFNAR and IFNGR respectively, present on cell surface membranes. Type I IFNs are broadly expressed heterodimeric receptors composed of the IFNAR1 and IFNAR2 subunits, while the type II IFN receptor consists of IFNGR1 and IFNGR2. Type III interferon lambda has three members: lamda1 (IL-29), lambda2 (IL-28A), and lambda3 (IL-28B) respectively. IFN-lambda signaling is initiated through unique heterodimeric receptor composed of IFN-LR1/IF-28Ralpha and IL10R2 chains.

Type I IFNs typically recruit JAK1 and TYK2 proteins to transduce their signals to STAT1 and 2; in combination with IRF9 (IFN-regulatory factor 9), these proteins form the heterotrimeric complex ISGF3. In nucleus ISGF3 binds to IFN-stimulated response elements (ISRE) to promote gene induction.

Type II IFNs in turn rely upon the activation of JAKs 1 and 2 and STAT1. Once activated, STAT1 dimerizes to form the transcriptional regulator GAF (IFNG activated factor) and this binds to the IFNG activated sequence (GAS) elements and initiate the transcription of IFNG-responsive genes.

Like type I IFNs, IFN-lambda recruits TYK2 and JAK1 kinases and then promote the phosphorylation of STAT1/2, and induce the ISRE3 complex formation.

#### Edit history

| Date | Action | Author |
|---|---|---|
| 2010-07-07 | Edited | Garapati P V |
| 2010-07-07 | Authored | Garapati P V |
| 2010-07-16 | Created | Garapati P V |
| 2010-08-17 | Reviewed | Abdul-Sater AA, Schindler C |
| 2021-05-22 | Modified | Shorser S |

#### Entities found in this pathway (1)

| Input | UniProt Id |
|---|---|
| HLA-C | P04222, P10321, P30499, P30501, P30504, P30505, P30508, P30510, Q07000, Q29865, Q29960, Q29963, Q95604, Q9TNN7 |

| Input | Ensembl Id |
|---|---|
| HLA-C | ENSG00000204525 |

#### 10. HSF1-dependent transactivation (R-HSA-3371571)

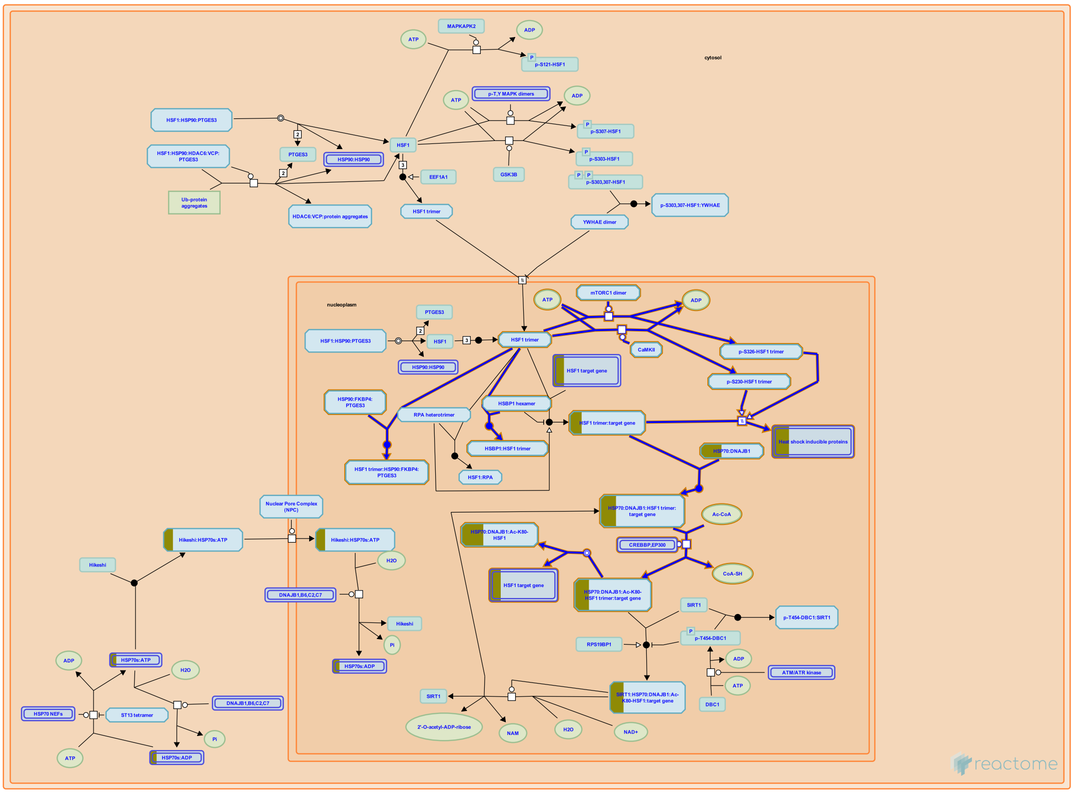


Acquisition of DNA binding activity by HSF1 is necessary but insufficient for transcriptional activation (Cotto JJ et al. 1996; Trinklein ND et al. 2004). In addition to having a sequence-specific DNA binding domain, HSF1 contains a C-terminal region which is involved in activating the transcription of the target genes (Green M et al. 1995). However, the transactivating ability of the transactivation domain itself is not stress sensitive. Rather, it's controled by a regulatory domain of HSF1 (amino acids 221-310), which represses the transactivating ability under normal physiological conditions (Green M et al. 1995; Zuo J et al. 1995; Newton EM et al. 1996). The HSF1 transactivation domain can be divided into two distinct regions, activation domain 1 (AD1) and activation domain 2 (AD2) (Brown SA et al. 1998). AD1 and AD2 each contain residues that are important for both transcriptional initiation and elongation. Mutations in acidic residues in both AD1 and AD2 preferentially affect the ability of HSF1 to stimulate transcriptional initiation, while mutations in phenylalanine residues preferentially affect stimulation of elongation (Brown SA et al. 1998).

Activation of the DNA-bound but transcriptionally incompetent HSF1 is thought to occur upon stress induced HSF1 phosphorylation at several serine residues (Ding XZ et al. 1997; Holmberg CI et al. 2001; Guettouche T et al. 2005). In cells exposed to heat, acquisition of HSE DNA-binding activity was observed to precede phosphorylation of HSF1 (Cotto JJ et al. 1996; Kline MP & Morimoto RI 1997). While there is a sufficient evidence to suggest that phosphorylation of HSF1 is essential to modulate HSF1 transactiviting capacity, mechanisms behind stress stimuli and kinases/phosphatases involved have not been clearly established.


##### Edit history

| Date | Action | Author |
|---|---|---|
| 2013-05-13 | Created | Shamovsky V |
| 2013-10-29 | Authored | Shamovsky V |
| 2014-02-17 | Edited | Shamovsky V |
| 2014-02-17 | Reviewed | Pani B |
| 2021-05-22 | Modified | Shorser S |

#### Entities found in this pathway (1)

| Input | UniProt Id |
|---|---|
| HSPA1B | P0DMV8, P0DMV9 |

| Input | Ensembl Id |
|---|---|
| HSPA1B | ENSG00000204388, ENSG00000212866, ENSG00000224501, ENSG00000231555, ENSG00000232804 |

#### 11. Class I MHC mediated antigen processing & presentation (R-HSA-983169)

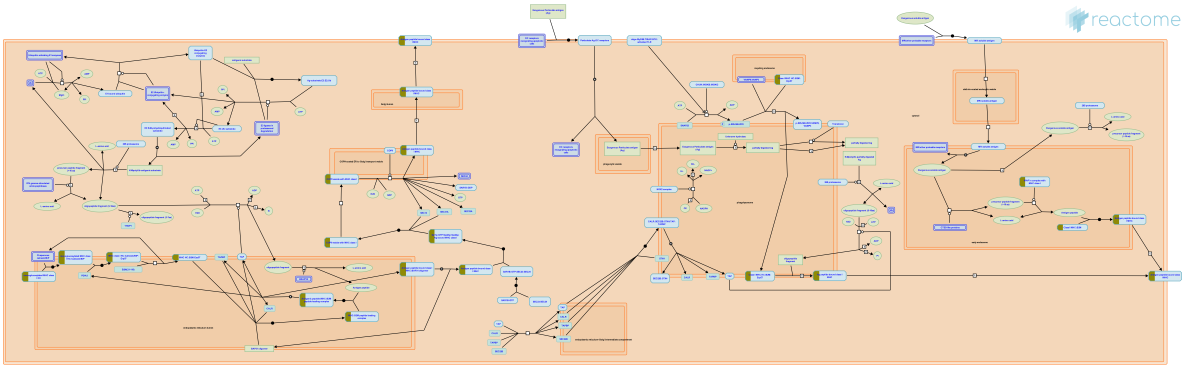

Major histocompatibility complex (MHC) class I molecules play an important role in cell mediated immunity by reporting on intracellular events such as viral infection, the presence of intracellular bacteria or tumor-associated antigens. They bind peptide fragments of these proteins and presenting them to CD8+ T cells at the cell surface. This enables cytotoxic T cells to identify and eliminate cells that are synthesizing abnormal or foreign proteins. MHC class I is a trimeric complex composed of a polymorphic heavy chain (HC or alpha chain) and an invariable light chain, known as beta2-microglobulin (B2M) plus an 8-10 residue peptide ligand. Represented here are the events in the biosynthesis of MHC class I molecules, including generation of antigenic peptides by the ubiquitin/26S-proteasome system, delivery of these peptides to the endoplasmic reticulum (ER), loading of peptides to MHC class I molecules and display of MHC class I complexes on the cell surface.


##### Edit history

| Date | Action | Author |
|---|---|---|
| 2010-10-29 | Edited | Garapati P V |
| 2010-10-29 | Authored | Garapati P V |
| 2010-10-29 | Created | Garapati P V |
| 2011-02-11 | Reviewed | Elliott T |
| 2021-05-22 | Modified | Shorser S |

##### Entities found in this pathway (1)

| Input | UniProt Id |
| --- | --- |
| HLA-C | P04222, P10321, P30499, P30501, P30504, P30505, P30508, P30510, Q07000, Q29865, Q29960, Q29963, Q95604, Q9TNN7 |

#### 12. HSF1 activation (R-HSA-3371511)

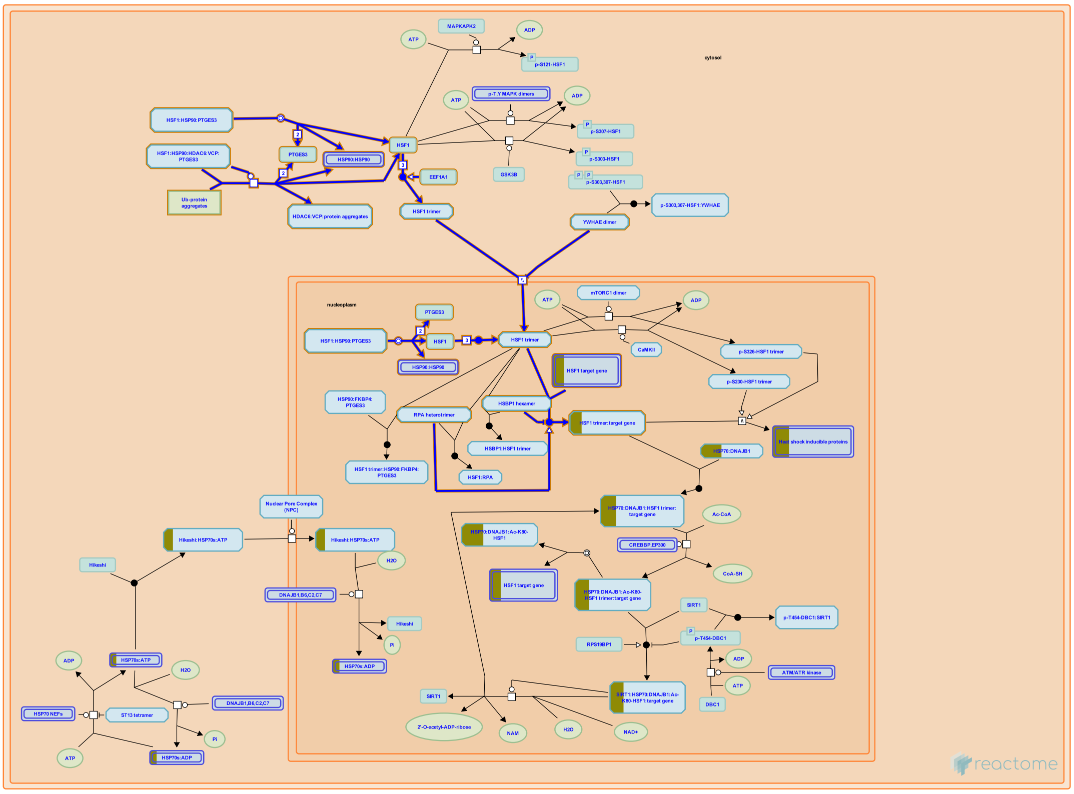

Heat shock factor 1 (HSF1) is a transcription factor that activates gene expression in response to a variety of stresses, including heat shock, oxidative stress, as well as inflammation and infection (Shamovsky I and Nudler E 2008; Akerfelt et al. 2010; Bjork and Sistonen 2010; Anckar and Sistonen 2011).

HSF1 is constitutively present in the cell. In the absence of stress HSF1 is found in both the cytoplasm and the nucleus as an inactive monomer (Sarge KD et al. 1993; Mercier PA et al. 1999; Vujanac M et al. 2005). A physical or chemical proteotoxic stress rapidly induces HSF1 activation, which occurs through a multi?step process, involving HSF1 monomer-to-homotrimer transition, nuclear accumulation, and binding to a promoter element, called the heat shock element (HSE), which leads to the increase in the stress-inducible gene expression (Sarge KD et al. 1993; Baler R et al. 1998; Sonna LA et al. 2002; Shamovsky I and Nudler E 2008; Sakurai H and Enoki Y 2010; Herbomel G et al. 2013). Depending on the type of stress stimulus, the multiple events associated with HSF1 activation might be affected differently (Holmberg CI et al 2000; Bjork and Sistonen 2010).


##### Edit history

| Date | Action | Author |
|---|---|---|
| 2013-05-13 | Created | Shamovsky V |
| 2013-10-29 | Authored | Shamovsky V |
| 2014-02-17 | Edited | Shamovsky V |
| 2014-02-17 | Reviewed | Pani B |
| 2021-05-22 | Modified | Shorser S |

#### Entities found in this pathway (1)

| Input | Ensembl Id |
|---|---|
| HSPA1B | ENSG00000204388, ENSG00000212866, ENSG00000224501, ENSG00000231555, ENSG00000232804 |

#### 13. Regulation of HSF1-mediated heat shock response (R-HSA-3371453)

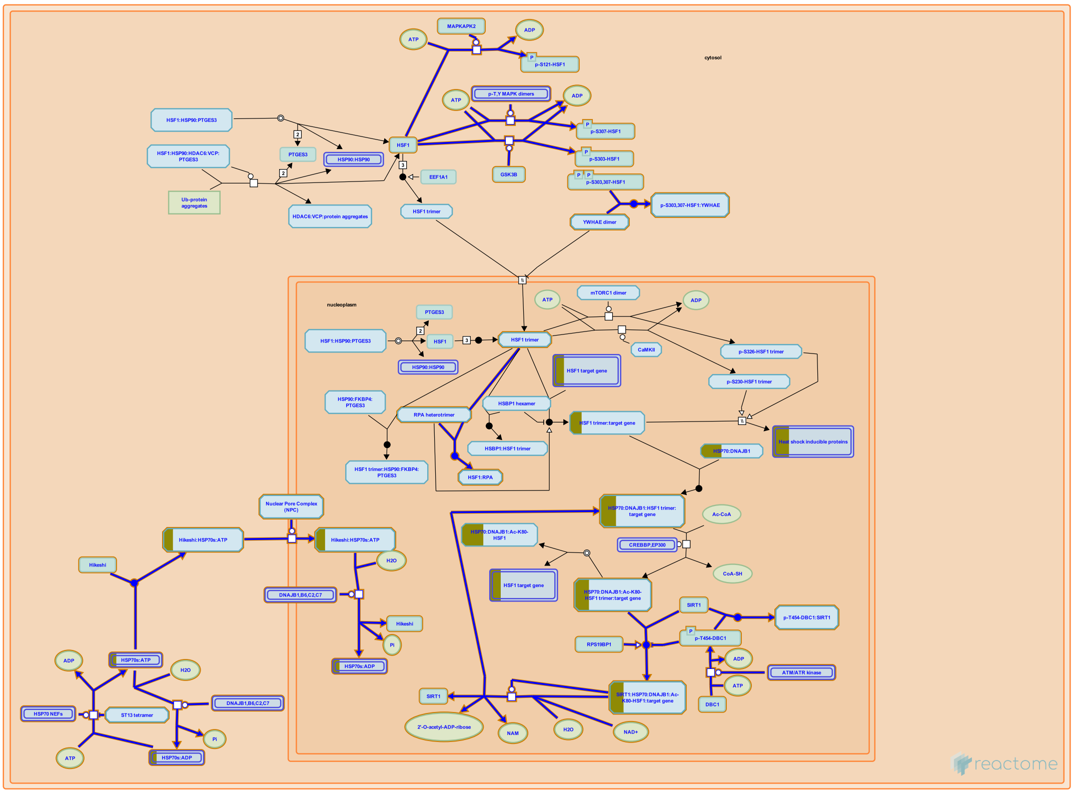

The ability of HSF1 to respond to cellular stresses is under negative regulation by chaperones, modulation of nucleocytoplasmic shuttling, post-translational modifications and transition from monomeric to trimeric state.


##### Edit history

| Date | Action | Author |
|---|---|---|
| 2013-05-13 | Created | Shamovsky V |
| 2013-10-29 | Authored | Shamovsky V |
| 2014-02-17 | Edited | Shamovsky V |
| 2014-02-17 | Reviewed | Pani B |
| 2021-05-22 | Modified | Shorser S |

##### Entities found in this pathway (1)

| Input | UniProt Id |
|---|---|
| HSPA1B | P0DMV8, P0DMV9 |

| Input | Ensembl Id |
|---|---|
| HSPA1B | ENSG00000204388, ENSG00000212866, ENSG00000224501, ENSG00000231555, ENSG00000232804 |

#### 14. RHO GTPases activate CIT (R-HSA-5625900)

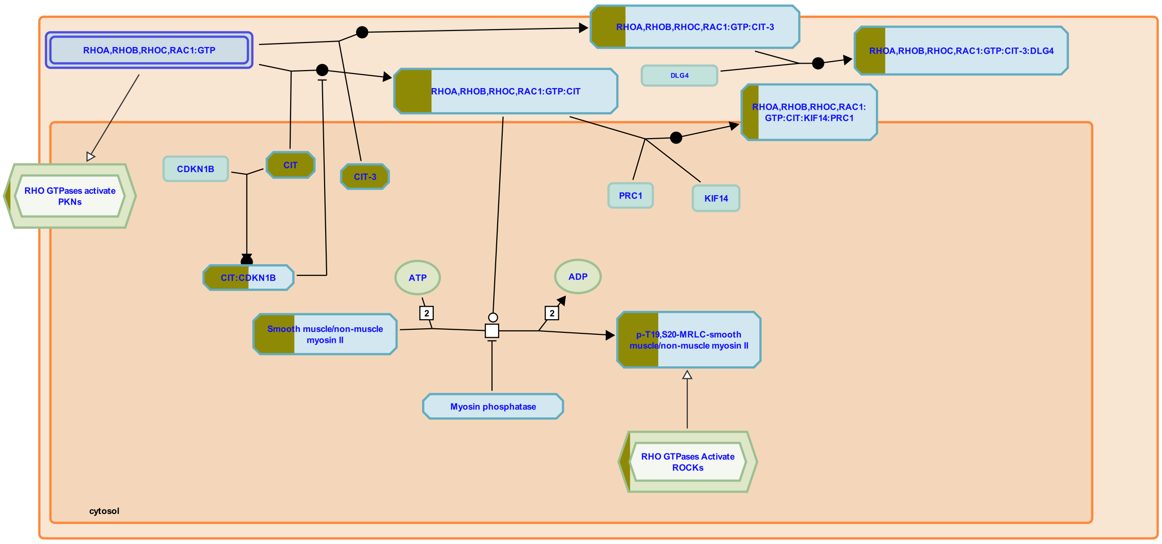


Citron kinase (CIT) or citron RHO-interacting kinase (CRIK) shares similarities with ROCK kinases. Like ROCK, it consists of a serine/threonine kinase domain, a coiled-coil region, a RHO-binding domain, a cysteine rich region and a plekstrin homology (PH) domain, but additionally features a proline-rich region and a PDZ-binding domain. A shorter splicing isoform of CIT, citron-N, is specifically expressed in the nervous system and lacks the kinase domain. Citron-N is a component of the post-synaptic density, where it binds to the PDZ domains of the scaffolding protein PDS-95/SAP90 (Zhang et al. 2006).

While the binding of CIT to RHO GTPases RHOA, RHOB, RHOC and RAC1 is well established (Madaule et al. 1995), the mechanism of CIT activation by GTP-bound RHO GTPases has not been elucidated. There are indications that CIT may be activated through autophosphorylation in the presence of active forms of RHO GTPases (Di Cunto et al. 1998). CIT appears to phosphorylate the myosin regulatory light chain (MRLC), the only substrate identified to date, on the same residues that are phosphorylated by ROCKs, but it has not been established yet how this relates to activation by RHO GTPases (Yamashiro et al. 2003). CIT and RHOA are implicated to act together in Golgi apparatus organization through regulation of the actin cytoskeleton (Camera et al. 2003). CIT is also involved in the regulation of cytokinesis through its interaction with KIF14 (Gruneberg et al. 2006, Bassi et al. 2013, Watanabe et al. 2013) and p27(Kip1) (Serres et al. 2012).

#### Edit history

| Date | Action | Author |
|---|---|---|
| 2014-10-08 | Created | Orlic-Milacic M |
| 2014-10-24 | Authored | Orlic-Milacic M |
| 2014-12-26 | Authored | Rivero Crespo F |
| 2015-02-02 | Edited | Orlic-Milacic M |
| 2021-05-31 | Modified | Shorser S |

#### Entities found in this pathway (3)

| Input | UniProt Id | Input | UniProt Id | Input | UniProt Id |
|---|---|---|---|---|---|
| CIT | O14578, O14578-3 | MYH11 | P35749 | MYL9 | P24844 |

#### 15. Cellular response to heat stress (R-HSA-3371556)

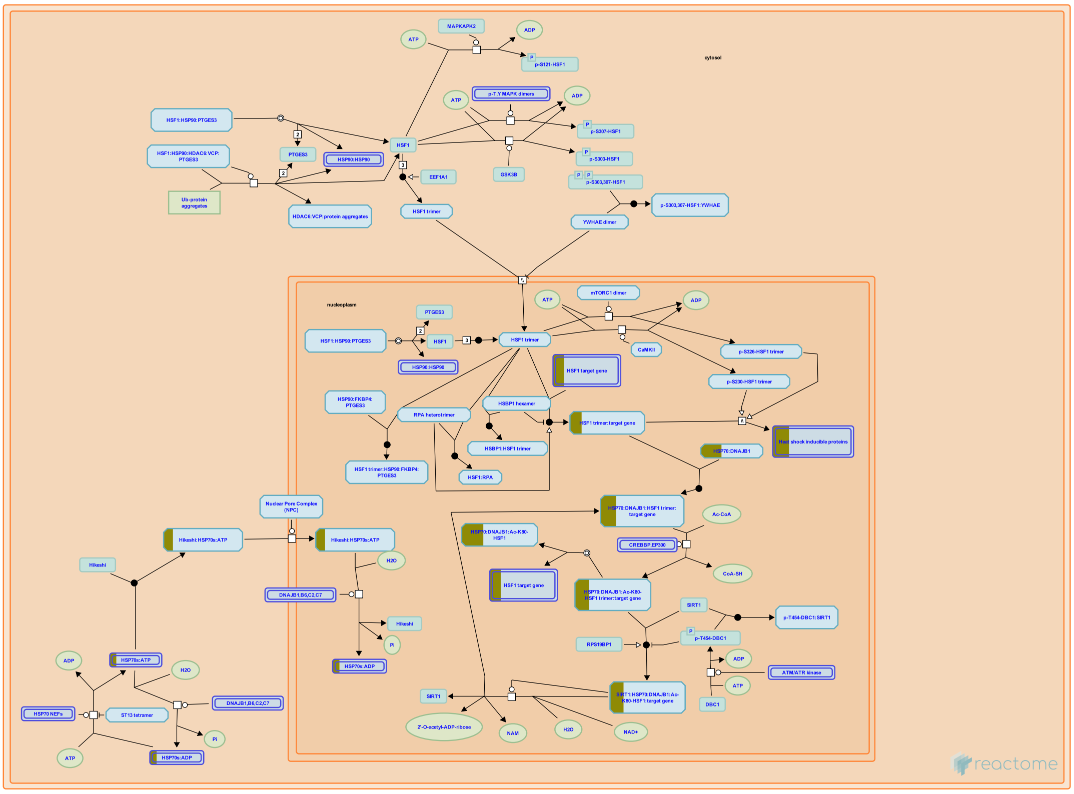

In response to exposure to elevated temperature and certain other proteotoxic stimuli (e.g., hypoxia, free radicals) cells activate a number of cytoprotective mechanisms known collectively as "heat shock response". Major aspects of the heat shock response (HSR) are evolutionarily conserved events that allow cells to recover from protein damage induced by stress (Liu XD et al. 1997; Voellmy R & Boellmann F 2007; Shamovsky I & Nudler E 2008; Anckar J & Sistonen L 2011). The main hallmark of HSR is the dramatic alteration of the gene expression pattern. A diverse group of protein genes is induced by the exposure to temperatures 3-5 degrees higher than physiological. Functionally, most of these genes are molecular chaperones that ensure proper protein folding and quality control to maintain cell proteostasis.

At the same time, heat shock-induced phosphorylation of translation initiation factor eIF2alpha leads to the shutdown of the nascent polypeptide synthesis reducing the burden on the chaperone system that has to deal with the increased amount of misfolded and thermally denatured proteins (Duncan RF & Hershey JWB 1989; Sarkar A et al. 2002; Spriggs KA et al. 2010).

The induction of HS gene expression primarily occurs at the level of transcription and is mediated by heat shock transcription factor HSF1(Sarge KD et al. 1993; Baler R et al. 1993). Human cells express five members of HSF protein family: HSF1, HSF2, HSF4, HSFX and HSFY. HSF1 is the master regulator of the heat inducible gene expression (Zuo J et al. 1995; Akerfelt M et al. 2010). HSF2 is activated in response to certain developmental stimuli in addition to being co-activated with HSF1 to provide promoter-specific fine-tuning of the HS response by forming heterotrimers with HSF1 (Ostling P et al. 2007; Sandqvist A et al. 2009). HSF4 lacks the transcription activation domain and acts as a repressor of certain genes during HS (Nakai A et al. 1997; Tanabe M et al. 1999; Kim SA et al. 2012). Two additional family members HSFX and HSFY, which are located on the X and Y chromosomes respectively, remain to be characterized (Bhowmick BK et al. 2006; Shinka T et al. 2004; Kichine E et al. 2012).

Under normal conditions HSF1 is present in both cytoplasm and nucleus in the form of an inactive monomer. The monomeric state of HSF1 is maintained by an intricate network of protein-protein interactions that include the association with HSP90 multichaperone complex, HSP70/HSP40 chaperone machinery, as well as intramolecular interaction of two conserved hydrophobic repeat regions. Monomeric HSF1 is constitutively phosphorylated on Ser303 and Ser 307 by (Zou J et al. 1998; Knauf U et al. 1996; Kline MP & Moromoto RI 1997; Guettouche T et al. 2005). This phosphorylation plays an essential role in ensuring cytoplasmic localization of at least a subpopulation of HSF1 molecules under normal conditions (Wang X et al. 2004).

Exposure to heat and other proteotoxic stimuli results in the release of HSF1 from the inhibitory complex with chaperones and its subsequent trimerization, which is promoted by its interaction with translation elongation factor eEF1A1 (Baler R et al. 1993; Shamovsky I et al. 2006; Herbomel G et al 2013). The trimerization is believed to involve intermolecular interaction between hydrophobic repeats 1-3 leading to the formation of a triple coil structure. Additional stabilization of the HSF1 trimer is provided by the formation of intermolecular S-S bonds between Cys residues in the DNA binding domain (Lu M et al.2008). Trimeric HSF1 is predominantly localized in the nucleus where it binds the specific sequence in the promoter of hsp genes (Sarge KD et al. 1993; Wang Y and Morgan WD 1994). The binding sequence for HSF1 (HSE, heat shock element) contains series of inverted repeats nGAAn in head-to-tail orientation, with at least three elements being required for the high affinity binding. Binding of the HSF1 trimer to the promoter is not sufficient to induce transcription of the gene (Cotto J et al. 1996). In order to do so, HSF1 needs to undergo inducible phosphorylation on specific Ser residues such as Ser230, Ser326. This phosphorylated form of HSF1 trimer is capable of increasing the promoter initiation rate. HSF1 bound to DNA promotes recruiting components of the transcription mediator complex and relieving promoter-proximal pause of RNA polymerase II through its interaction with TFIIH transcription factor (Yuan CX & Gurley WB 2000).

HSF1 activation is regulated in a precise and tight manner at multiple levels (Zuo J et al. 1995; Cotto J et al. 1996). This allows fast and robust activation of HS response to minimize proteotoxic effects of the stress. The exact set of HSF1 inducible genes is probably cell type specific. Moreover, cells in different pathophysiological states will display different but overlapping profile of HS inducible genes.


#### Edit history

| Date | Action | Author |
| --- | --- | --- |
| 2013-05-13 | Created | Shamovsky V |
| 2013-10-29 | Authored | Shamovsky V |
| 2014-02-17 | Edited | Shamovsky V |
| 2014-02-17 | Reviewed | Pani B |

| Date | Action | Author |
|---|---|---|
| 2021-05-22 | Modified | Shorser S |

#### Entities found in this pathway (1)

| Input | UniProt Id |
|---|---|
| HSPA1B | P0DMV8, P0DMV9 |

| Input | Ensembl Id |
|---|---|
| HSPA1B | ENSG00000204388, ENSG00000212866, ENSG00000224501, ENSG00000231555, ENSG00000232804 |

#### 16. Smooth Muscle Contraction (R-HSA-445355)

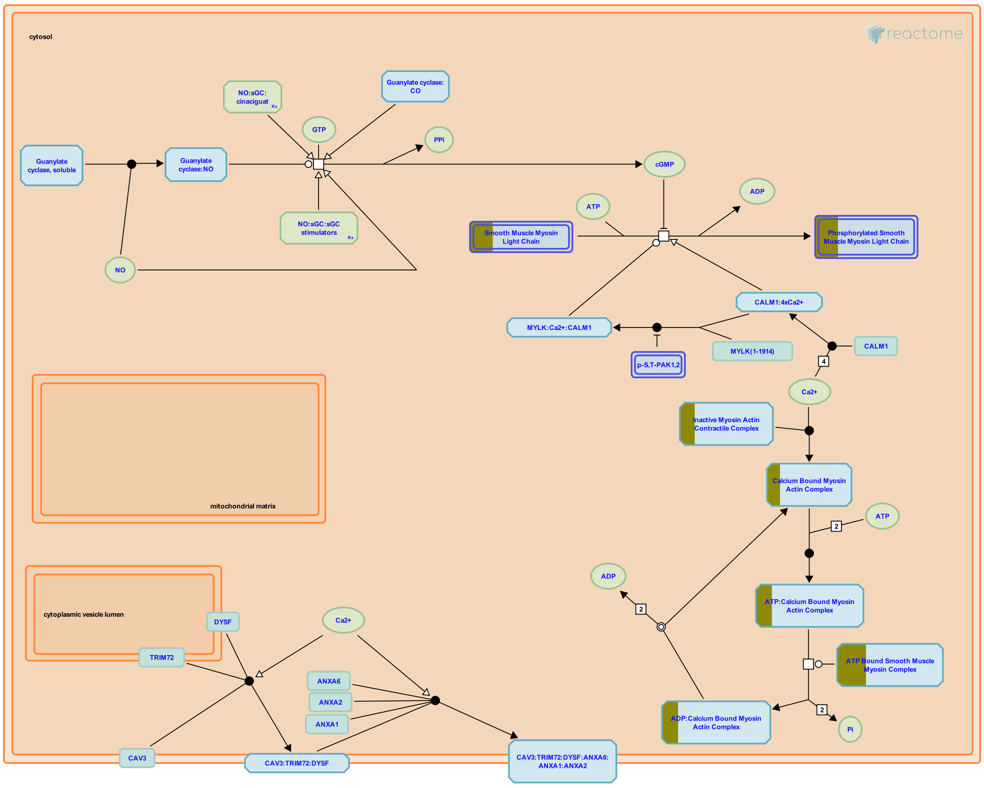


**Cellular compartments:** cytosol, plasma membrane.

Layers of smooth muscle cells can be found in the walls of numerous organs and tissues within the body. Smooth muscle tissue lacks the striated banding pattern characteristic of skeletal and cardiac muscle. Smooth muscle is triggered to contract by the autonomic nervous system, hormones, autocrine/paracrine agents, local chemical signals, and changes in load or length.

Actin:myosin cross bridging is used to develop force with the influx of calcium ions (Ca2+) initiating contraction. Two separate protein pathways, both triggered by calcium influx contribute to contraction, a calmodulin driven kinase pathway, and a caldesmon driven pathway.

Recent evidence suggests that actin, myosin, and intermediate filaments may be far more volatile then previously suspected, and that changes in these cytoskeletal elements along with alterations of the focal adhesions that anchor these proteins may contribute to the contractile cycle.

Contraction in smooth muscle generally uses a variant of the same sliding filament model found in striated muscle, except in smooth muscle the actin and myosin filaments are anchored to focal adhesions, and dense bodies, spread over the surface of the smooth muscle cell. When actin and myosin move across one another focal adhesions are drawn towards dense bodies, effectively squeezing the cell into a smaller conformation. The sliding is triggered by calcium:caldesmon binding, caldesmon acting in an analogous fashion to troponin in striated muscle. Phosphorylation of myosin light chains also is involved in the initiation of an effective contraction.

##### Edit history

| Date | Action | Author |
|---|---|---|
| 2008-01-11 | Reviewed | Rush MG |
| 2009-03-09 | Authored | Gillespie ME |
| 2009-10-30 | Created | Gillespie ME |
| 2009-11-18 | Edited | Gillespie ME |
| 2021-05-31 | Modified | Shorser S |

#### Entities found in this pathway (3)

| Input | UniProt Id | Input | UniProt Id | Input | UniProt Id |
|---|---|---|---|---|---|
| MYH11 | P35749 | MYL9 | P19105, P24844 | TPM2 | P07951 |

#### 17. Muscle contraction (R-HSA-397014)

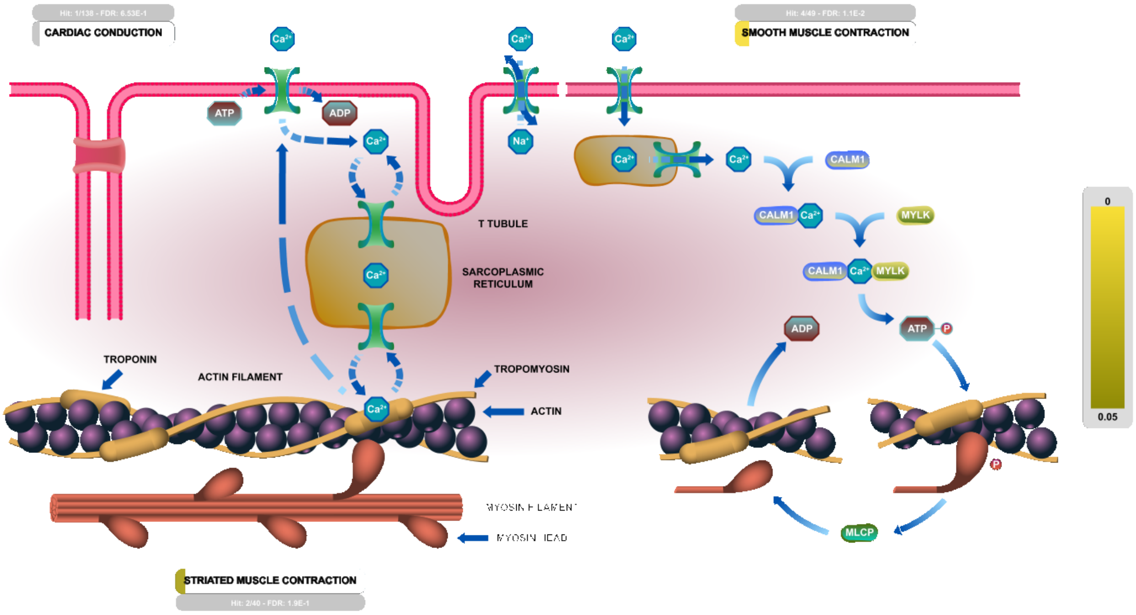


**Cellular compartments:** cytosol, plasma membrane.

In this module, the processes by which calcium binding triggers actin - myosin interactions and force generation in smooth and striated muscle tissues are annotated.

##### References

##### Edit history

| Date | Action | Author |
|---|---|---|
| 2009-02-10 | Authored | Gillespie ME |
| 2009-03-11 | Edited | Gillespie ME |
| 2009-03-11 | Created | May B |
| 2021-05-22 | Modified | Shorser S |

##### Entities found in this pathway (5)

| Input | UniProt Id | Input | UniProt Id | Input | UniProt Id |
|---|---|---|---|---|---|
| DES | P17661 | KCNJ4 | P48050 | MYH11 | P35749 |
| MYL9 | P19105, P24844 | TPM2 | P07951 | | |

#### 18. GABA B receptor activation (R-HSA-977444)

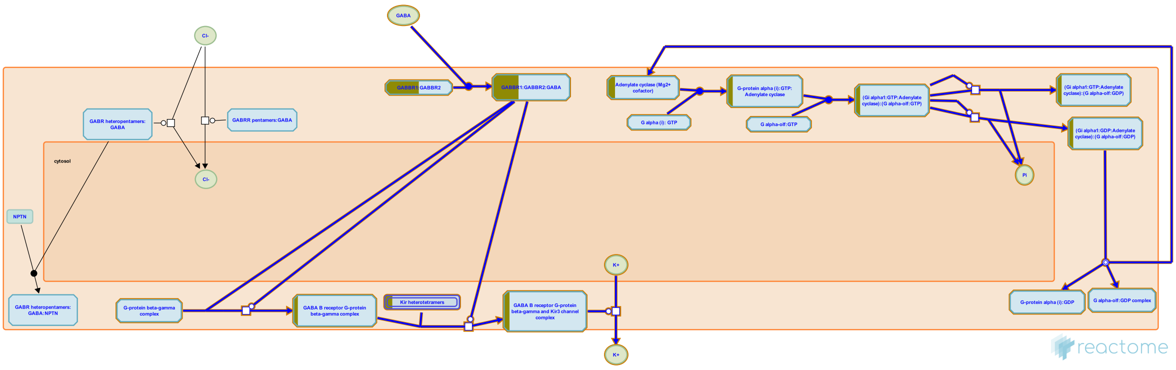


**Cellular compartments:** plasma membrane, cytosol, extracellular region.

Functional GABA B receptors are heteromers of GABA B1 and B2 subunits, complexed with G protein alpha-i, 0, beta, and gamma subunits. They function as metabotropic receptors. When GABA is bound to the B1 sub-unit, the B2 subunit undergoes a conformational change that releases the G alpha-i G0 dimer (which binds and inactivates cytosolic adenylate cyclase) and the G beta G gamma dimer (which activates the GIRK (KIR3) potassium channel) (Pinard et al. 2010).

##### Edit history

| Date | Action | Author |
|---|---|---|
| 2008-11-27 | Reviewed | Restituito S |
| 2010-10-19 | Created | Mahajan SS |
| 2010-11-08 | Authored | Mahajan SS |
| 2010-11-25 | Edited | D'Eustachio P |
| 2021-05-22 | Modified | Shorser S |

##### Entities found in this pathway (3)

| Input | UniProt Id | Input | UniProt Id | Input | UniProt Id |
|---|---|---|---|---|---|
| ADCY1 | Q08828 | GABBR2 | O75899 | KCNJ4 | P48050 |

#### 19. Activation of GABAB receptors (R-HSA-991365)

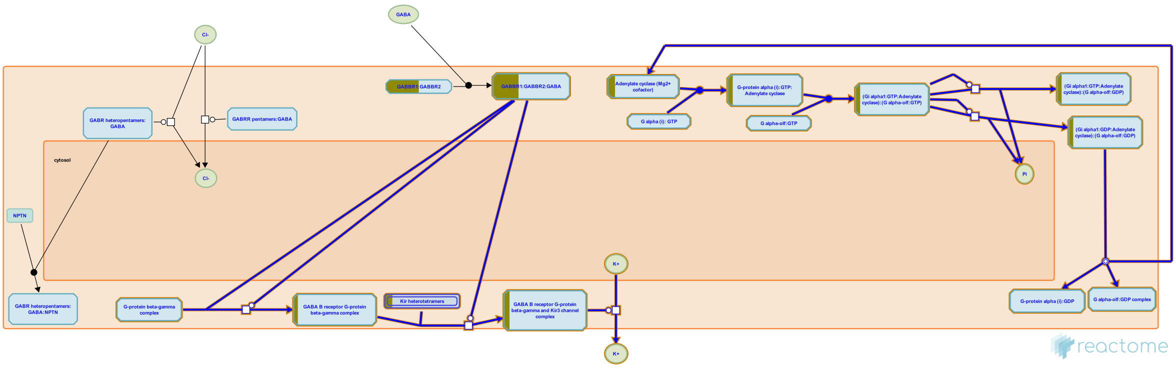


**Cellular compartments:** cytosol, extracellular region.

GABA B receptors are metabotropic receptors that are functionally linked to C type G protein coupled receptors.?
GABA B receptors are activated upon ligand binding. The GABA B1 subunit binds ligand and GABA B2 subunit modulates the activity of adenylate cyclase via the intracellular loop.?
GABA B receptors show inhibitory activity via Galpha/G0 subunits via the inhibition of adenylate cyclase or via the activity of Gbeta/gamma subunits that mediate the inhibition of voltage gated Ca2+ channels.


##### Edit history

| Date | Action | Author |
|---|---|---|
| 2008-11-27 | Reviewed | Restituito S |
| 2010-11-03 | Created | Mahajan SS |
| 2010-11-08 | Authored | Mahajan SS |
| 2010-11-25 | Edited | D'Eustachio P |
| 2021-05-22 | Modified | Shorser S |

##### Entities found in this pathway (3)

| Input | UniProt Id | Input | UniProt Id | Input | UniProt Id |
|---|---|---|---|---|---|
| ADCY1 | Q08828 | GABBR2 | O75899 | KCNJ4 | P48050 |

#### 20. CREB1 phosphorylation through the activation of Adenylate Cyclase (R-HSA-442720)

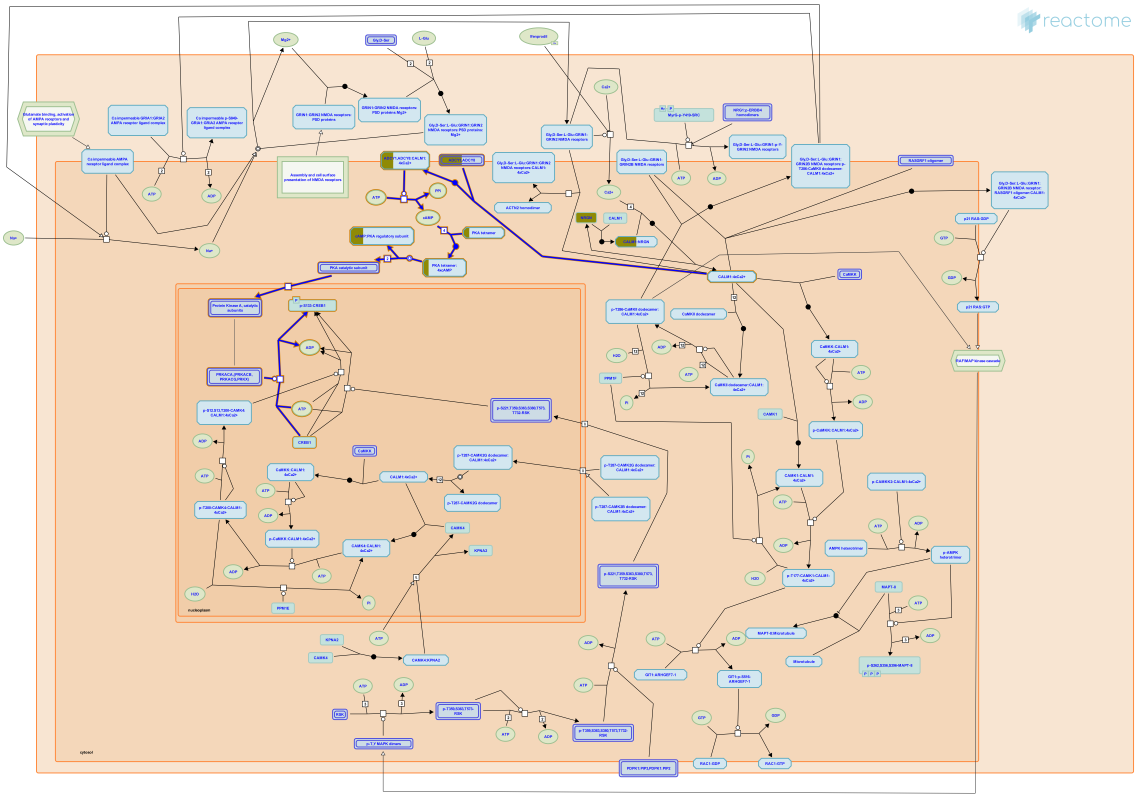

**Cellular compartments:** plasma membrane, cytosol, nucleoplasm.

Ca2+ influx through activated NMDA receptors in the post synaptic neurons activates adenylate cyclase-mediated signal transduction, leading to the activation of PKA and phosphorylation and activation of CREB1 induced transcription (Masada et al. 2012, Chetkovich et al. 1991, Chetkovich and Sweatt 1993


##### Edit history

| Date | Action | Author |
|---|---|---|
| 2009-09-29 | Created | Mahajan SS |
| 2009-10-29 | Authored | Mahajan SS |
| 2009-11-18 | Reviewed | Tukey D |
| 2009-11-19 | Edited | Gillespie ME |
| 2018-10-10 | Revised | Orlic-Milacic M |
| 2018-11-02 | Reviewed | Hansen KB, Yi F |
| 2018-11-07 | Edited | Orlic-Milacic M |
| 2021-05-31 | Modified | Shorser S |

#### Entities found in this pathway (2)

| Input | UniProt Id | Input | UniProt Id |
|---|---|---|---|
| ADCY1 | Q08828 | PRKAR1B | P31321 |

#### 21. Cytokine Signaling in Immune system (R-HSA-1280215)

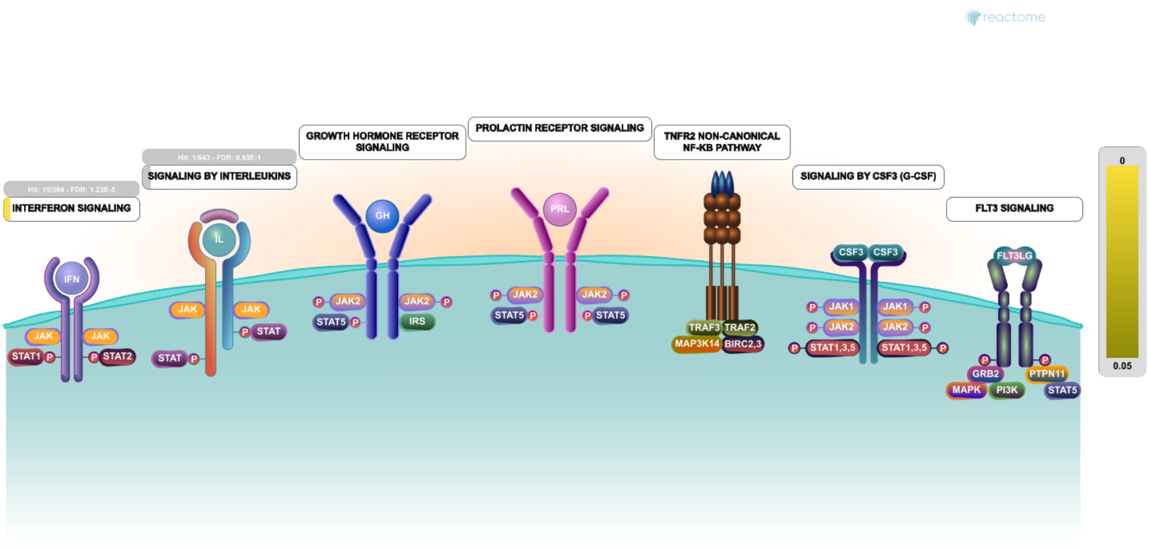


Cytokines are small proteins that regulate and mediate immunity, inflammation, and hematopoiesis. They are secreted in response to immune stimuli, and usually act briefly, locally, at very low concentrations. Cytokines bind to specific membrane receptors, which then signal the cell via second messengers, to regulate cellular activity.

IMMPORT:Bioinformatics for the future of immunology. Retrieved from https://www.immport.org/immportWeb/queryref/geneListSummary.do

COPE. Retrieved from http://www.copewithcytokines.org/cope.cgi

Santamaria P (2003). Cytokines and chemokines in autoimmune disease: an overview. Adv Exp Med Biol, 520, 1-7.


##### Edit history

| Date | Action | Author |
|---|---|---|
| 2011-05-12 | Created | Garapati P V |
| 2011-05-22 | Edited | Ray KP, Jupe S, Garapati P V |
| 2011-05-22 | Authored | Ray KP, Jupe S, Garapati P V |
| 2011-05-29 | Reviewed | Abdul-Sater AA, Schindler C, Pinteaux E |
| 2021-05-22 | Modified | Shorser S |

##### Entities found in this pathway (2)

| Input | UniProt Id | Input | UniProt Id |
|---|---|---|---|
| HLA-C | P04222, P10321, P30499, P30501, P30504, P30505, P30508, P30510, Q07000, Q29865, Q29960, Q29963, Q95604, Q9TNN7 | IL16 | Q14005 |

| Input | Ensembl Id |
|---|---|
| HLA-C | ENSG00000204525 |

#### 22. Potassium Channels (R-HSA-1296071)

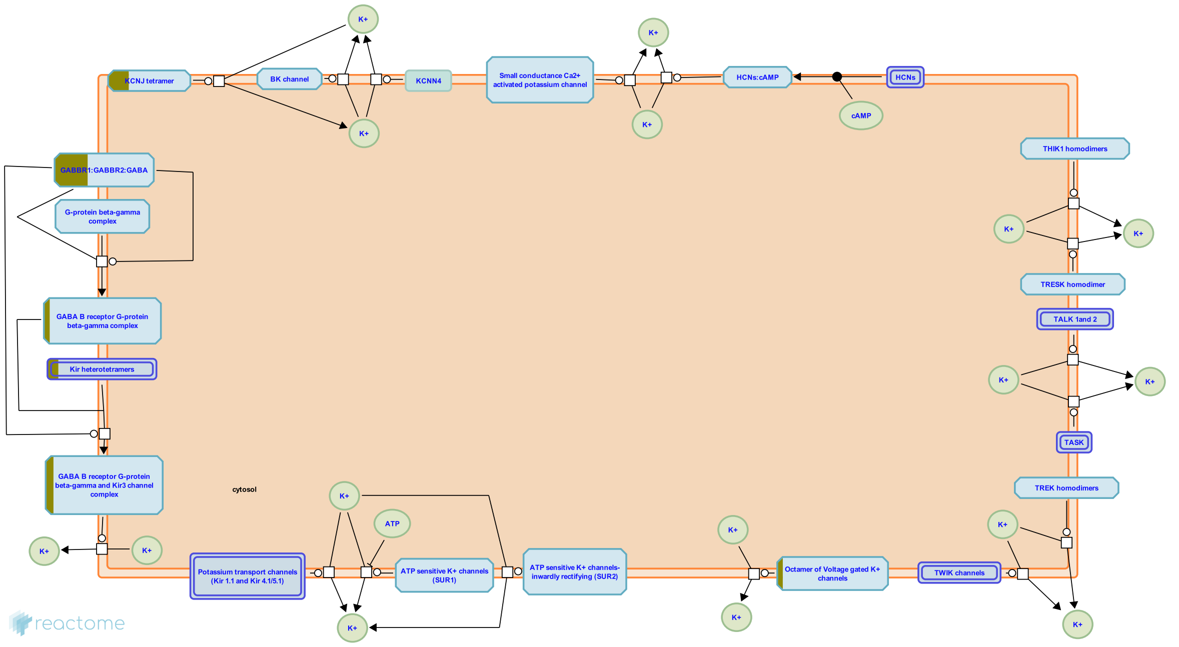


Potassium channels are tetrameric ion channels that are widely distributed and are found in all cell types. Potassium channels control resting membrane potential in neurons, contribute to regulation of action potentials in cardiac muscle and help release of insulin form pancreatic beta cells.

Broadly K+ channels are classified into voltage gated K+ channels, Hyperpolarization activated cyclic nucleotide gated K+ channels (HCN), Tandem pore domain K+ channels, Ca2+ activated K+ channels and inwardly rectifying K+ channels.


##### Edit history

| Date | Action | Author |
|---|---|---|
| 2010-09-23 | Reviewed | Jassal B |
| 2011-05-19 | Authored | Mahajan SS |
| 2011-05-19 | Created | Mahajan SS |
| 2011-05-23 | Edited | Mahajan SS |

| Date | Action | Author |
|---|---|---|
| 2021-05-22 | Modified | Shorser S |

#### Entities found in this pathway (3)

| Input | UniProt Id | Input | UniProt Id | Input | UniProt Id |
|---|---|---|---|---|---|
| GABBR2 | O75899 | KCNB1 | Q14721, Q92953 | KCNJ4 | P48050 |

#### 23. Adaptive Immune System (R-HSA-1280218)

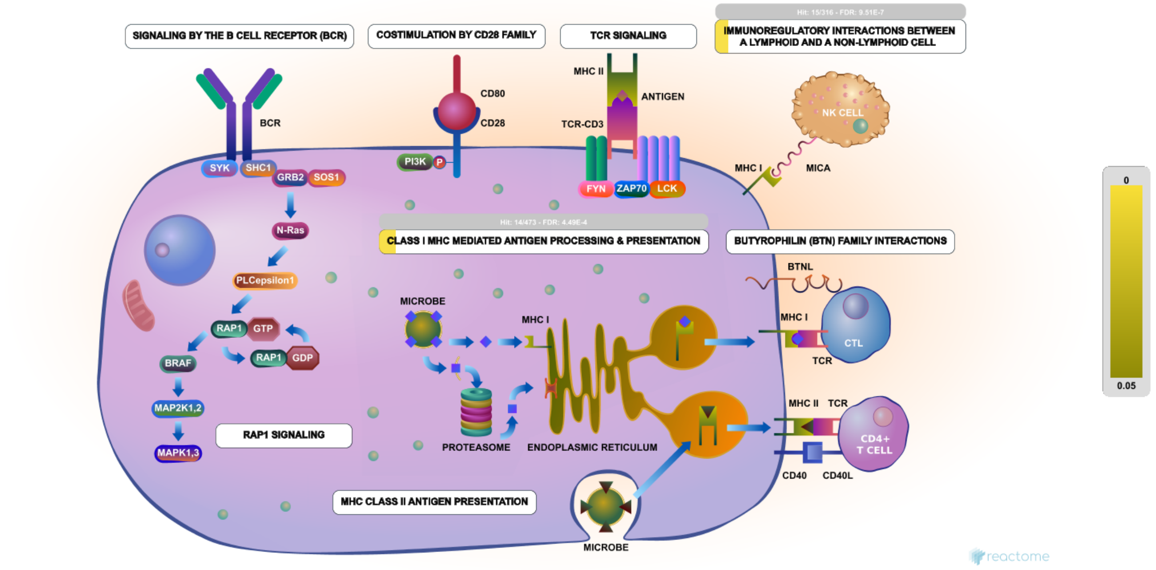


Adaptive immunity refers to antigen-specific immune response efficiently involved in clearing the pathogens. The adaptive immune system is comprised of B and T lymphocytes that express receptors with remarkable diversity tailored to recognize aspects of particular pathogens or antigens. During infection, dendritic cells (DC) which act as sentinels in the peripheral tissues recognize and pick up the pathogen in the form of antigenic determinants and then process these antigens and present them to T cells. These T cells of appropriate specificity respond to the antigen, and either kill the pathogen directly or secrete cytokines that will stimulate B lymphocyte response. B cells provide humoral immunity by secreting antibodies specific for the pathogen or antigen.


##### Edit history

| Date | Action | Author |
|---|---|---|
| 2011-05-12 | Created | Garapati P V |
| 2011-05-22 | Edited | May B, Jupe S, Garapati P V, de Bono B |
| 2011-05-22 | Authored | May B, Jupe S, Garapati P V, de Bono B |
| 2011-05-28 | Reviewed | Heemskerk JW, Bluestone JA, Elliott T, Trowsdale J, Esensten J |
| 2021-05-22 | Modified | Shorser S |

##### Entities found in this pathway (2)

| Input | UniProt Id | Input | UniProt Id |
|---|---|---|---|
| HLA-C | P04222, P10321, P30499, P30501, P30504, P30505, P30508, P30510, Q07000, Q29865, Q29960, Q29963, Q95604, Q9TNN7 | PIANP | Q8IYJ0 |

#### 24. Neuronal System (R-HSA-112316)

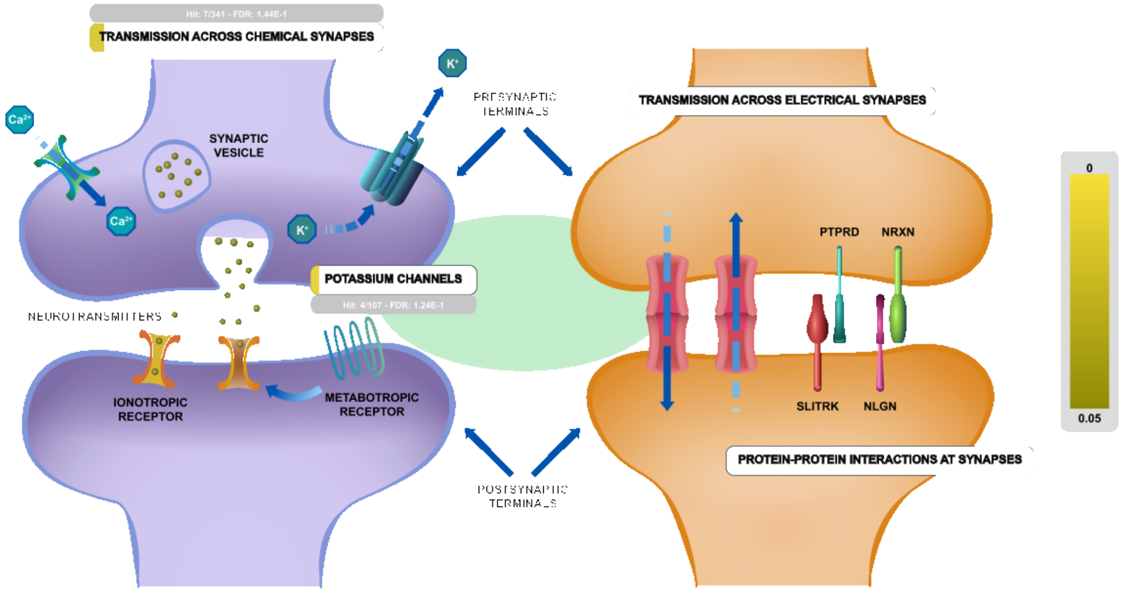


The human brain contains at least 100 billion neurons, each with the ability to influence many other cells. Clearly, highly sophisticated and efficient mechanisms are needed to enable communication among this astronomical number of elements. This communication occurs across synapses, the functional connection between neurons. Synapses can be divided into two general classes: electrical synapses and chemical synapses. Electrical synapses permit direct, passive flow of electrical current from one neuron to another. The current flows through gap junctions, specialized membrane channels that connect the two cells. Chemical synapses enable cell-to-cell communication using neurotransmitter release. Neurotransmitters are chemical agents released by presynaptic neurons that trigger a secondary current flow in postsynaptic neurons by activating specific receptor molecules. Neurotransmitter secretion is triggered by the influx of Ca2+ through voltage-gated channels, which gives rise to a transient increase in Ca2+ concentration within the presynaptic terminal. The rise in Ca2+ concentration causes synaptic vesicles (the presynaptic organelles that store neurotransmitters) to fuse with the presynaptic plasma membrane and release their contents into the space between the pre- and postsynaptic cells.


##### Edit history

| Date | Action | Author |
|---|---|---|
| 2004-04-22 | Created | Joshi-Tope G |
| 2005-11-10 | Edited | Gillespie ME |
| 2005-11-10 | Authored | Gillespie ME |
| 2021-05-22 | Modified | Shorser S |

##### Entities found in this pathway (8)

| Input | UniProt Id |
|---|---|
| ADCY1 | Q08828 |
| GABBR2 | O75899 |
| NRGN | Q92686 |

| Input | UniProt Id |
|---|---|
| CACNA1A | O00555 |
| KCNB1 | Q14721, Q92953 |
| PRKAR1B | P31321 |

| Input | UniProt Id |
|---|---|
| CPLX1 | O14810 |
| KCNJ4 | P48050 |

#### 25. PKA activation in glucagon signalling (R-HSA-164378)

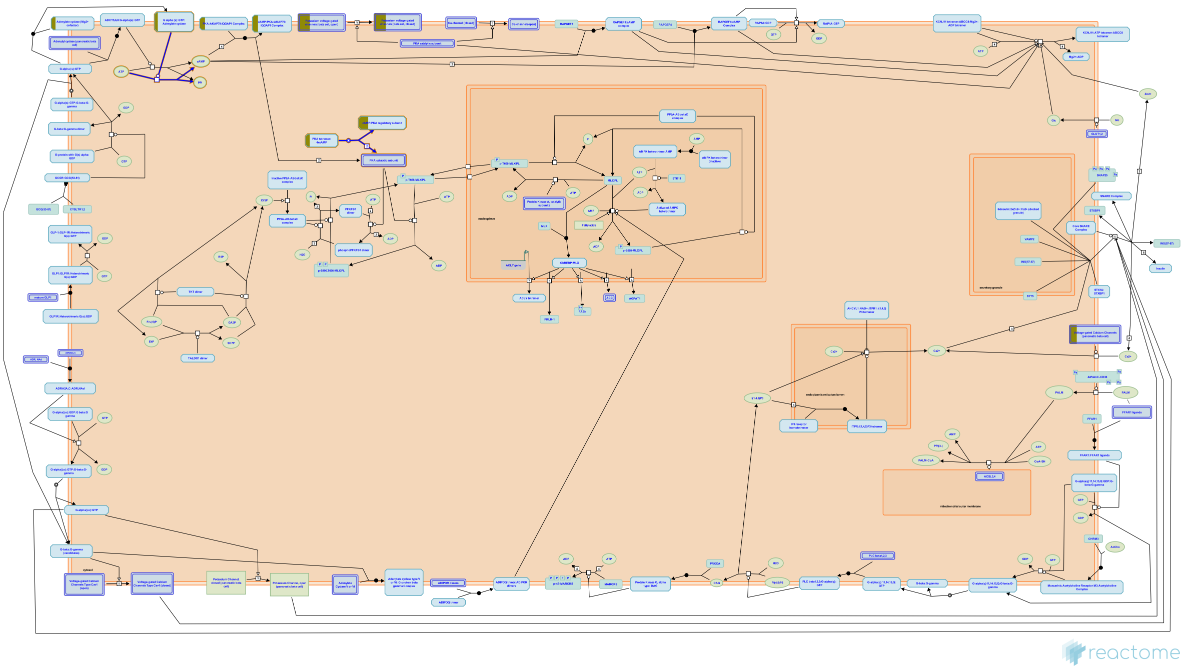

**Cellular compartments:** plasma membrane.

Adenylate cyclase catalyses the synthesis of cyclic AMP (cAMP) from ATP. In the absence of cAMP, protein kinase A (PKA) exists as inactive tetramers of two catalytic subunits and two regulatory subunits. cAMP binding to PKA tetramers causes them to dissociate and release their catalytic subunits as active monomers. Four isoforms of the regulatory subunit are known, that differ in their tissue specificity and functional characteristics, but the specific isoform activated in response to glucagon signaling has not yet been identified.

##### References

##### Edit history

| Date | Action | Author |
|---|---|---|
| 2005-05-19 | Authored | Gopinathrao G, D'Eustachio P |
| 2005-05-19 | Created | Gopinathrao G |
| 2021-05-22 | Modified | Shorser S |

##### Entities found in this pathway (2)

| Input | UniProt Id | Input | UniProt Id |
|---|---|---|---|
| ADCY1 | Q08828 | PRKAR1B | P31321 |

### 6. Identifiers found

Below is a list of the input identifiers that have been found or mapped to an equivalent element in Reactome, classified by resource.

#### Entities (42)

| Input | UniProt Id | Input | UniProt Id | Input | UniProt Id |
|---|---|---|---|---|---|
| ADAMTS14 | Q8WXS8 | ADCY1 | Q08828 | ADCYAP1 | P18509 |
| CACNA1A | O00555 | CCK | P06307 | CFH | P08603, Q03591, Q92496 |
| CIT | O14578, O14578-3 | COLGALT1 | Q8IYK4, Q8NBJ5 | CPLX1 | O14810 |
| CPNE9 | P55201 | DENND3 | A2RUS2 | DES | P17661 |
| EPS8L2 | Q9H6S3 | FAM20A | Q96MK3 | GABBR2 | O75899 |
| GPX3 | O75715, P22352 | HECW1 | Q76N89 | HIST1H2BG | P62807 |
| HLA-C | P04222, P10321, P30499, P30501, P30504, P30505, P30508, P30510, Q07000, Q29865, Q29960, Q29963, Q95604, Q9TNN7 | HSPA1B | P0DMV8, P0DMV9 | IGFBP4 | P22692 |
| IGFBP6 | P24592 | IL16 | Q14005 | KCNB1 | Q14721, Q92953 |
| KCNJ4 | P48050 | LINGO1 | Q96FE5 | MAPK13 | O15264 |
| MYH11 | P35749 | MYL9 | P24844 | NDUFS2 | O75306 |
| NIPAL2 | Q9H841 | NRGN | Q92686 | PCNT | O95613 |
| PIANP | Q8IYJ0 | PRKAR1B | P31321 | RAB17 | Q9H0T7 |
| RXFP1 | Q9HBX9 | SLC6A20 | Q9NP91 | SLCO4A1 | Q96BD0 |
| SYNGR1 | O43759 | TPM2 | P07951 | ZNF697 | Q5TEC3 |

| Input | Ensembl Id | Input | Ensembl Id | Input | Ensembl Id |
|---|---|---|---|---|---|
| HLA-C | ENSG00000204525 | HSPA1B | ENSG00000204388, ENSG00000212866, ENSG00000224501, ENSG00000231555, ENSG00000232804 | MYL9 | ENSG00000101335 |

### 7. Identifiers not found

These 39 identifiers were not found neither mapped to any entity in Reactome.

| | | | | | | | |
|---|---|---|---|---|---|---|---|
| ANKRD18B | ATCAY | BCL7A | C10orf128 | C9orf91 | CALY | CLSTN3 | COCH |
| CPNE5 | CTD-3162L10.1 | CTGLF12P | DIRAS2 | FAT4 | INA | JAKMIP3 | KIAA1107 |
| KIAA1217 | KIAA1549L | LRRC37A2 | NDUFA4L2 | NPIPB3 | PARM1 | PCDH11X | PKNOX2 |
| PTPRT | RP11-197K6.1 | RP11-32B5.1 | RP11-32B5.7 | RP11-583F2.1 | RPH3A | SGSM1 | SH3RF2 |
| SNX18P7 | SORCS1 | SVIL | SYNGR2 | TAGLN | TGM2 | VSTM2L | |
