## Supplementary material for "Compartment-specific total RNA profile of Hippocampal and Cortical cells from Mesial Temporal Lobe Epilepsy tissue": Other supplemental files_Reactome: HC_mTLE non-HS_Nucleus_UPspecific_report.pdf

Fabregat A, Sidiropoulos K, Viteri G, Forner O, Marin-Garcia P, Arnau V, … Hermjakob H (2017). Reactome pathway analysis: a high-performance in-memory approach. BMC Bioinformatics, 18.

### 2. Properties

- This is an **overrepresentation** analysis: A statistical (hypergeometric distribution) test that determines whether certain Reactome pathways are over-represented (enriched) in the submitted data. It answers the question 'Does my list contain more proteins for pathway X than would be expected by chance?' This test produces a probability score, which is corrected for false discovery rate using the Benjamani-Hochberg method.
- 134 out of 256 identifiers in the sample were found in Reactome, where 761 pathways were hit by at least one of them.
- All non-human identifiers have been converted to their human equivalent.
- This report is filtered to show only results for species 'Homo sapiens' and resource 'all resources'.
- The unique ID for this analysis (token) is MjAyMTA5MTQwODMzMTRfNTM4MDU%3D. This ID is valid for at least 7 days in Reactome's server. Use it to access Reactome services with your data.

### 3. Genome-wide overview

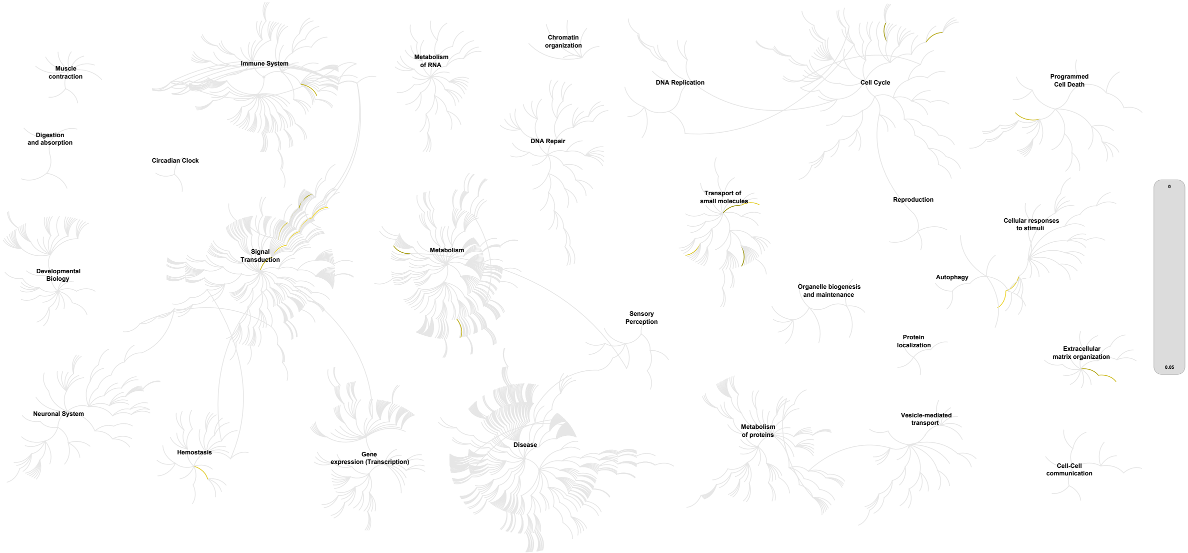


This figure shows a genome-wide overview of the results of your pathway analysis. Reactome pathways are arranged in a hierarchy. The center of each of the circular "bursts" is the root of one top-level pathway, for example "DNA Repair". Each step away from the center represents the next level lower in the pathway hierarchy. The color code denotes over-representation of that pathway in your input dataset. Light grey signifies pathways which are not significantly over-represented.

### 4. Most significant pathways

The following table shows the 25 most relevant pathways sorted by p-value.

| Pathway name | Entities | | | | Reactions | |
|---|---|---|---|---|---|---|
| | found | ratio | p-value | FDR* | found | ratio |
| NGF-stimulated transcription | 8 / 56 | 0.004 | 1.93e-05 | 0.016 | 11 / 37 | 0.003 |
| Nuclear Events (kinase and transcription factor activation) | 8 / 80 | 0.006 | 2.24e-04 | 0.092 | 11 / 48 | 0.004 |
| Signaling by NTRK1 (TRKA) | 9 / 143 | 0.01 | 0.002 | 0.542 | 16 / 102 | 0.008 |
| ATF4 activates genes in response to endoplasmic reticulum stress | 4 / 34 | 0.002 | 0.005 | 0.542 | 2 / 7 | 5.18e-04 |
| Signaling by NTRKs | 9 / 166 | 0.011 | 0.006 | 0.542 | 21 / 164 | 0.012 |
| Passive transport by Aquaporins | 3 / 21 | 0.001 | 0.009 | 0.542 | 4 / 8 | 5.92e-04 |
| PERK regulates gene expression | 4 / 42 | 0.003 | 0.01 | 0.542 | 2 / 11 | 8.14e-04 |
| Formation of Fibrin Clot (Clotting Cascade) | 4 / 43 | 0.003 | 0.011 | 0.542 | 18 / 61 | 0.005 |
| Chylomicron clearance | 2 / 9 | 6.19e-04 | 0.014 | 0.542 | 4 / 4 | 2.96e-04 |
| Nuclear signaling by ERBB4 | 4 / 47 | 0.003 | 0.015 | 0.542 | 4 / 34 | 0.003 |
| Syndecan interactions | 3 / 29 | 0.002 | 0.02 | 0.542 | 7 / 15 | 0.001 |
| BH3-only proteins associate with and inactivate anti-apoptotic BCL-2 members | 2 / 11 | 7.56e-04 | 0.02 | 0.542 | 3 / 4 | 2.96e-04 |
| Signaling by Receptor Tyrosine Kinases | 20 / 617 | 0.042 | 0.022 | 0.542 | 154 / 744 | 0.055 |
| Interleukin-4 and Interleukin-13 signaling | 9 / 211 | 0.015 | 0.026 | 0.542 | 5 / 47 | 0.003 |
| Linoleic acid (LA) metabolism | 3 / 32 | 0.002 | 0.026 | 0.542 | 5 / 10 | 7.40e-04 |
| Nuclear Envelope (NE) Reassembly | 5 / 88 | 0.006 | 0.032 | 0.542 | 3 / 24 | 0.002 |
| Non-integrin membrane-ECM interactions | 4 / 61 | 0.004 | 0.034 | 0.542 | 10 / 22 | 0.002 |
| FGFR1c ligand binding and activation | 2 / 15 | 0.001 | 0.036 | 0.542 | 3 / 3 | 2.22e-04 |
| Vitamin E | 1 / 2 | 1.38e-04 | 0.039 | 0.542 | 2 / 2 | 1.48e-04 |
| Nuclear Envelope Breakdown | 4 / 65 | 0.004 | 0.041 | 0.542 | 5 / 13 | 9.62e-04 |
| Negative regulation of FGFR1 signaling | 3 / 39 | 0.003 | 0.043 | 0.542 | 4 / 15 | 0.001 |
| Aquaporin-mediated transport | 4 / 68 | 0.005 | 0.047 | 0.542 | 9 / 25 | 0.002 |
| Amino acid transport across the plasma membrane | 4 / 68 | 0.005 | 0.047 | 0.542 | 4 / 36 | 0.003 |
| Glycogen metabolism | 3 / 43 | 0.003 | 0.054 | 0.542 | 18 / 39 | 0.003 |

| Pathway name | Entities | | | | Reactions | |
|---|---|---|---|---|---|---|
| | found | ratio | p-value | FDR* | found | ratio |
| **Defective SLC4A4 causes renal tubular acidosis, proximal, with ocular abnormalities and mental retardation (pRTA-OA)** | 1 / 3 | 2.06e-04 | 0.057 | 0.542 | 1 / 1 | 7.40e-05 |

* False Discovery Rate

### 5. Pathways details

For every pathway of the most significant pathways, we present its diagram, as well as a short summary, its bibliography and the list of inputs found in it.

#### 1. NGF-stimulated transcription (R-HSA-9031628)

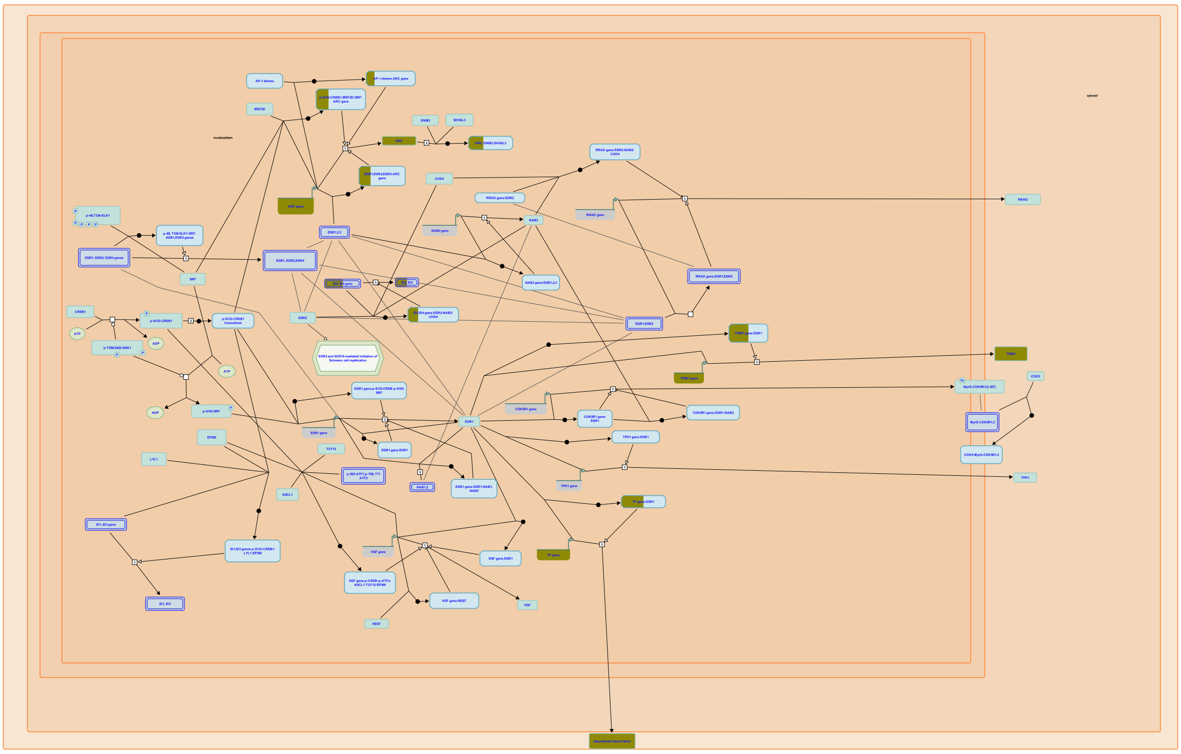

NGF stimulation induces expression of a wide array of transcriptional targets. In rat PC12 cells, a common model for NGF signaling, stimulation with NGF causes cells to exit the cell cycle and undergo a differentiation program leading to neurite outgrowth. This program is driven by the expression of immediate early genes (IEGs), which frequently encode transcription factors regulating the activity of NGF-specific delayed response genes (reviewed in Sheng and Greenberg, 1990; Flavell and Grennberg, 2008; Santiago and Bashaw, 2014).


##### Edit history

| Date | Action | Author |
|---|---|---|
| 2017-12-01 | Created | Rothfels K |

| Date | Action | Author |
|---|---|---|
| 2019-08-16 | Authored | Rothfels K |
| 2020-01-17 | Reviewed | Aletta J M |
| 2020-02-24 | Edited | Rothfels K |
| 2021-05-22 | Modified | Shorser S |

#### Entities found in this pathway (4)

| Input | UniProt Id |
|---|---|
| ARC | Q7LC44 |
| ID4 | P47928 |

| Input | UniProt Id |
|---|---|
| F3 | P13726 |
| TRIB1 | Q96RU8 |

| Input | Ensembl Id |
|---|---|
| ARC | ENSG00000198576 |
| ID4 | ENSG00000172201 |

| Input | Ensembl Id |
|---|---|
| F3 | ENSG00000117525 |
| TRIB1 | ENSG00000173334 |

#### 2. Nuclear Events (kinase and transcription factor activation) (R-HSA-198725)

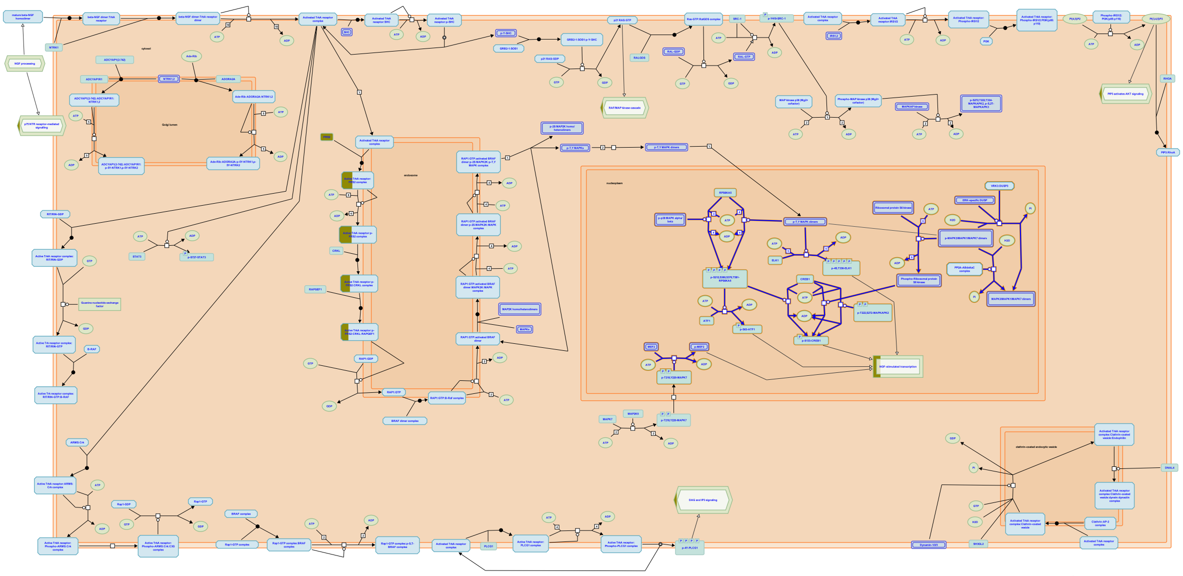

An important function of the kinase cascade triggered by neurotrophins is to induce the phosphorylation and activation of transcription factors in the nucleus to initiate new programs of gene expression. Transcription factors directly activated by neurotrophin signalling are responsible for induction of immediate-early genes, many of which are transcription factors. These in turn are involved in the induction of delayed-early genes.


##### Edit history

| Date | Action | Author |
|---|---|---|
| 2006-10-10 | Authored | Annibali D, Nasi S |
| 2007-07-10 | Created | |
| 2007-11-08 | Reviewed | Greene LA |
| 2021-05-22 | Modified | Shorser S |

##### Entities found in this pathway (4)

| Input | UniProt Id |
|---|---|
| ARC | Q7LC44 |
| ID4 | P47928 |

| Input | UniProt Id |
|---|---|
| F3 | P13726 |
| TRIB1 | Q96RU8 |

| Input | Ensembl Id |
|---|---|
| ARC | ENSG00000198576 |
| ID4 | ENSG00000172201 |

| Input | Ensembl Id |
|---|---|
| F3 | ENSG00000117525 |
| TRIB1 | ENSG00000173334 |

#### 3. Signaling by NTRK1 (TRKA) (R-HSA-187037)

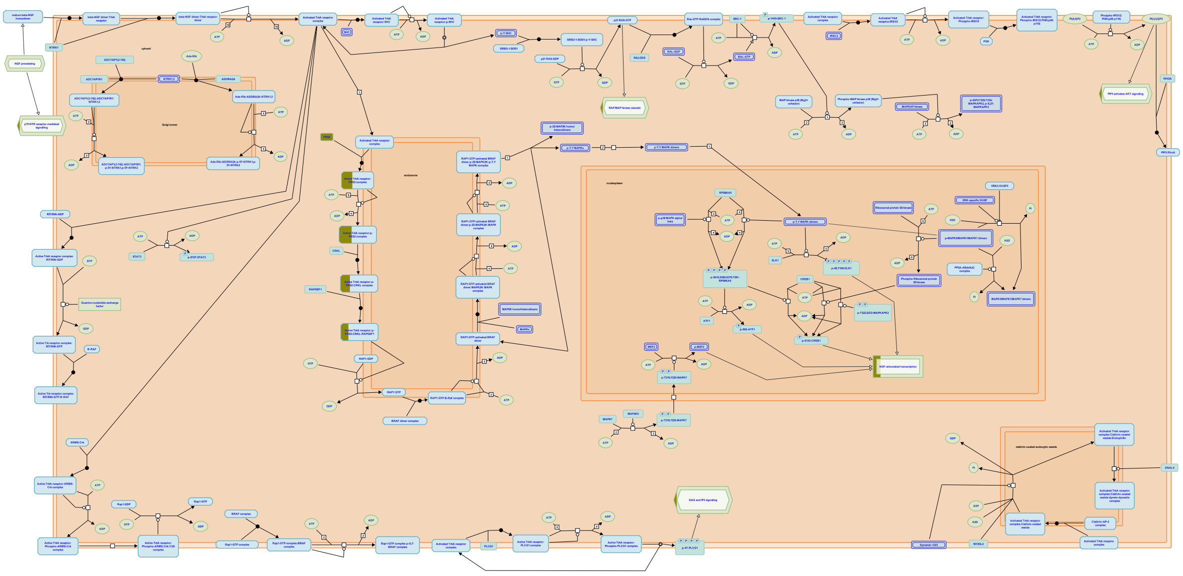

Trk receptors signal from the plasma membrane and from intracellular membranes, particularly from early endosomes. Signalling from the plasma membrane is fast but transient; signalling from endosomes is slower but long lasting. Signalling from the plasma membrane is annotated here. TRK signalling leads to proliferation in some cell types and neuronal differentiation in others. Proliferation is the likely outcome of short term signalling, as observed following stimulation of EGFR (EGF receptor). Long term signalling via TRK receptors, instead, was clearly shown to be required for neuronal differentiation in response to neurotrophins.


##### Edit history

| Date | Action | Author |
|---|---|---|
| 2006-09-07 | Created | Jassal B |
| 2006-10-10 | Edited | Jassal B |
| 2006-10-10 | Authored | Annibali D, Nasi S |
| 2007-11-08 | Reviewed | Greene LA |
| 2021-05-22 | Modified | Shorser S |

##### Entities found in this pathway (5)

| Input | UniProt Id |
|---|---|
| ARC | Q7LC44 |
| ID4 | P47928 |

| Input | UniProt Id |
|---|---|
| F3 | P13726 |
| TRIB1 | Q96RU8 |

| Input | UniProt Id |
|---|---|
| FRS2 | Q8WU20 |

| Input | Ensembl Id |
|---|---|
| ARC | ENSG00000198576 |
| ID4 | ENSG00000172201 |

| Input | Ensembl Id |
|---|---|
| F3 | ENSG00000117525 |
| TRIB1 | ENSG00000173334 |

#### 4. ATF4 activates genes in response to endoplasmic reticulum stress (R-HSA-380994)

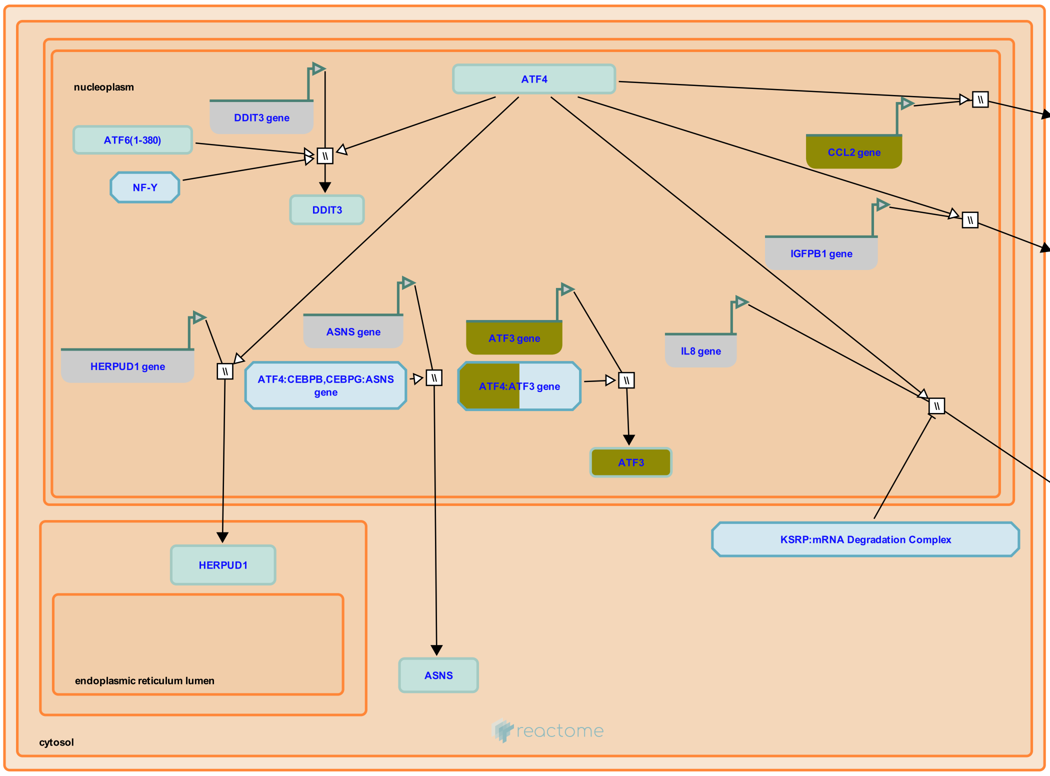


ATF4 is a transcription factor and activates expression of IL-8, MCP1, IGFBP-1, CHOP, HERP1 and ATF3.


##### Edit history

| Date | Action | Author |
|---|---|---|
| 2008-11-19 | Created | May B |

| Date | Action | Author |
|---|---|---|
| 2008-12-02 | Reviewed | Matthews L, D'Eustachio P, Gillespie ME |
| 2009-06-02 | Edited | May B |
| 2009-06-02 | Authored | May B |
| 2010-04-30 | Reviewed | Urano F |
| 2020-11-11 | Modified | D'Eustachio P |

#### Entities found in this pathway (2)

| Input | UniProt Id |
|---|---|
| ATF3 | P18847 |

| Input | UniProt Id |
|---|---|
| CCL2 | P13500 |

| Input | Ensembl Id |
|---|---|
| ATF3 | ENSG00000162772 |

| Input | Ensembl Id |
|---|---|
| CCL2 | ENSG00000108691 |

#### 5. Signaling by NTRKs (R-HSA-166520)

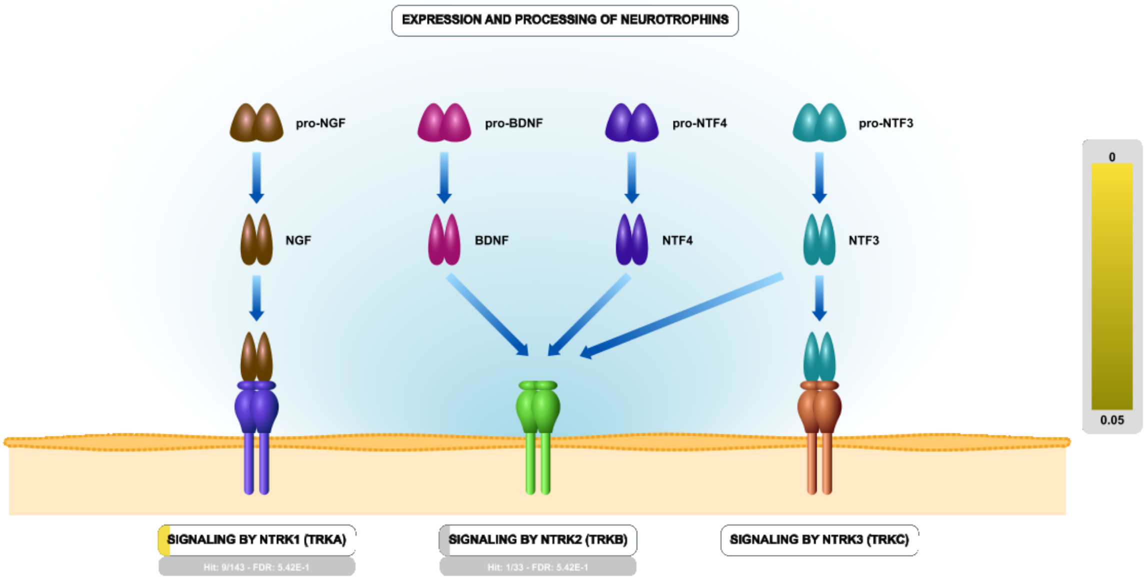


Neurotrophins (NGF, BDNF, NTF3 and NTF4) play pivotal roles in survival, differentiation, and plasticity of neurons in the peripheral and central nervous system. They are produced, and secreted in minute amounts, by a variety of tissues. They signal through two types of receptors: NTRK (TRK) tyrosine kinase receptors (TRKA, TRKB, TRKC), which differ in their preferred neurotrophin ligand, and p75NTR death receptor, which interacts with all neurotrophins. Besides the nervous system, TRK receptors and p75NTR are expressed in a variety of other tissues. For review, please refer to Bibel and Barde 2000, Poo 2001, Lu et al. 2005, Skaper 2012, Park and Poo 2013.

| Input | UniProt Id | Input | UniProt Id | Input | UniProt Id |
|---|---|---|---|---|---|
| ARC | Q7LC44 | F3 | P13726 | FRS2 | Q8WU20 |
| ID4 | P47928 | TRIB1 | Q96RU8 | | |

| Input | Ensembl Id | Input | Ensembl Id |
|---|---|---|---|
| ARC | ENSG00000198576 | F3 | ENSG00000117525 |
| ID4 | ENSG00000172201 | TRIB1 | ENSG00000173334 |

#### 6. Passive transport by Aquaporins (R-HSA-432047)

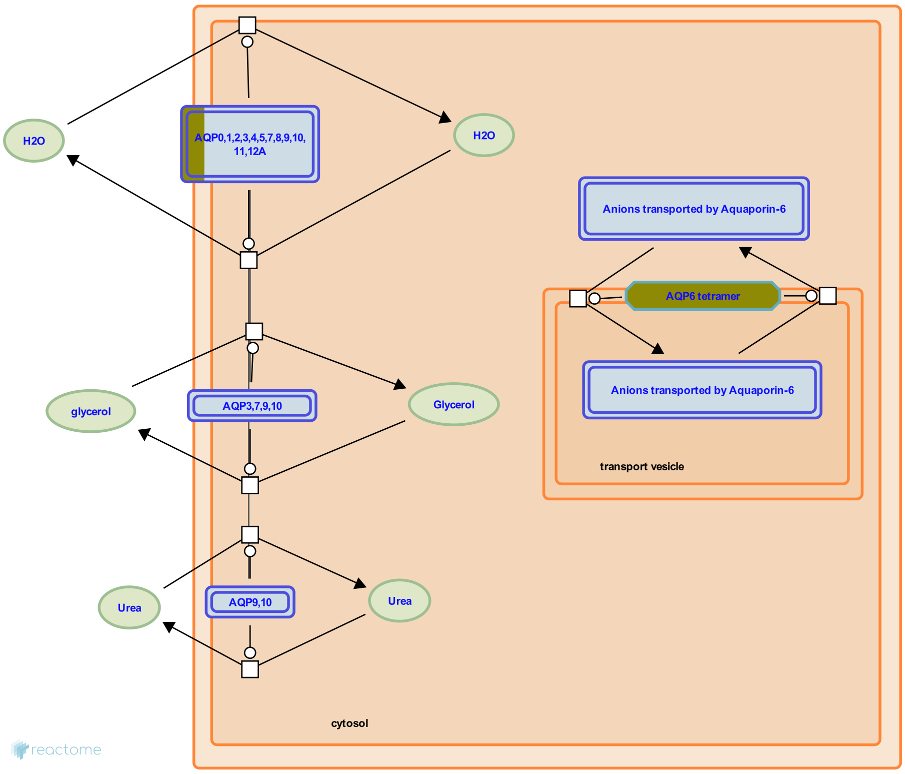


**Cellular compartments:** plasma membrane, transport vesicle membrane.

Aquaporins (AQP's) are six-pass transmembrane proteins that form channels in membranes. Each monomer contains a central channel formed in part by two asparagine-proline-alanine motifs (NPA boxes) that confer selectivity for water and/or solutes. The monomers assemble into tetramers. During passive transport by Aquaporins most aquaporins (i.e. AQP0/MIP, AQP1, AQP2, AQP3, AQP4, AQP5, AQP7, AQP8, AQP9, AQP10) transport water into and out of cells according to the osmotic gradient across the membrane. Four aquaporins (the aquaglyceroporins AQP3, AQP7, AQP9, AQP10) conduct glycerol, three aquaporins (AQP7, AQP9, AQP10) conduct urea, and one aquaporin (AQP6) conducts anions, especially nitrate. AQP8 also conducts ammonia in addition to water.

#### Entities found in this pathway (3)

| Input | UniProt Id | Input | UniProt Id | Input | UniProt Id |
|---|---|---|---|---|---|
| AQP1 | P29972 | AQP4 | P55087 | AQP6 | Q13520 |

#### 7. PERK regulates gene expression (R-HSA-381042)

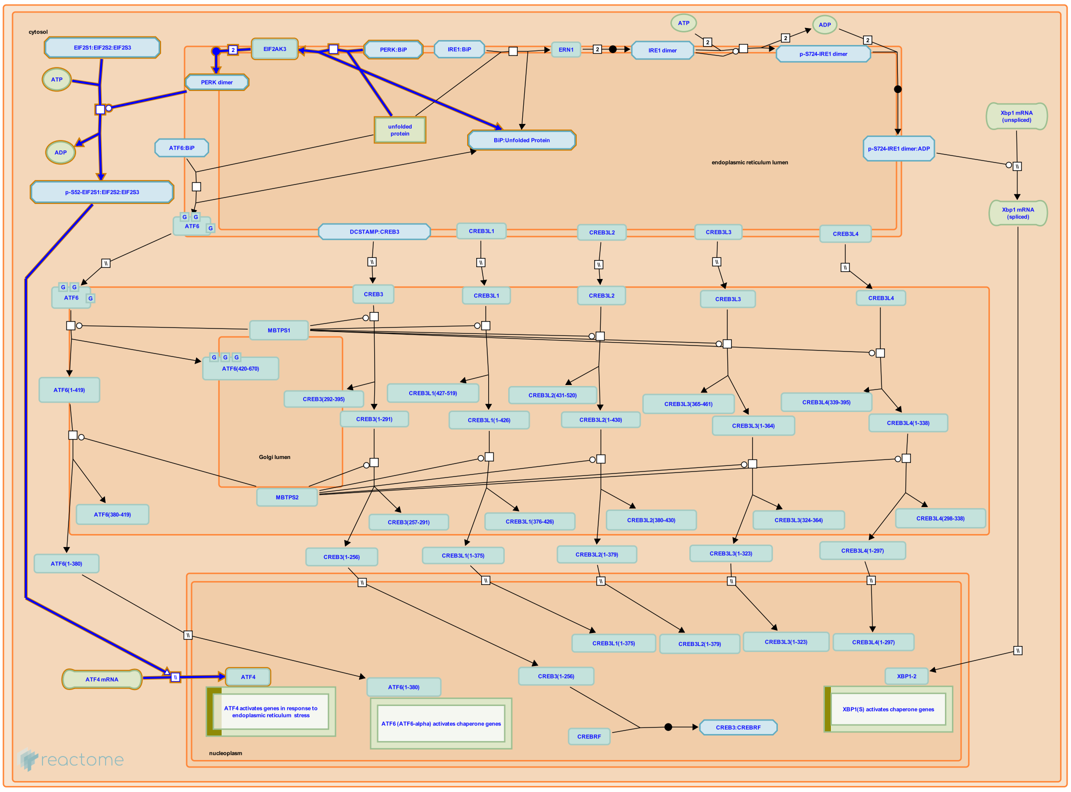


**Cellular compartments:** cytosol, endoplasmic reticulum lumen, endoplasmic reticulum membrane, nucleoplasm.

PERK (EIF2AK3) is a single-pass transmembrane protein located in the endoplasmic reticulum (ER) membrane such that the N-terminus of PERK is luminal and the C-terminus is cytosolic. PERK is maintained in an inactive form by interaction of its luminal domain with BiP, an ER chaperone. BiP also binds unfolded proteins and so BiP dissociates from PERK when unfolded proteins accumulate in the ER. Dissociated PERK monomers spontaneously form homodimers and the homodimeric form of PERK possesses kinase activity in its cytosolic C-terminal domain. The kinase specifically phosphorylates the translation factor eIF2alpha at Ser52, resulting in an arrest of translation. Thus translation of proteins targeted to the ER is downregulated. The translation arrest also causes depletion of Cyclin D1, a rapidly turned over protein. The depletion of Cyclin D1 in turn causes arrest of the cell cycle in G1 phase.


##### Edit history

| Date | Action | Author |
|---|---|---|
| 2008-11-19 | Created | May B |
| 2008-11-20 | Edited | May B, Gopinathrao G |
| 2008-12-02 | Reviewed | Matthews L, D'Eustachio P, Gillespie ME |
| 2009-06-02 | Authored | May B |
| 2010-04-30 | Reviewed | Urano F |
| 2021-05-22 | Modified | Shorser S |

#### Entities found in this pathway (2)

| Input | UniProt Id |
|---|---|
| ATF3 | P18847 |

| Input | UniProt Id |
|---|---|
| CCL2 | P13500 |

| Input | Ensembl Id |
|---|---|
| ATF3 | ENSG00000162772 |

| Input | Ensembl Id |
|---|---|
| CCL2 | ENSG00000108691 |

#### 8. Formation of Fibrin Clot (Clotting Cascade) (R-HSA-140877)

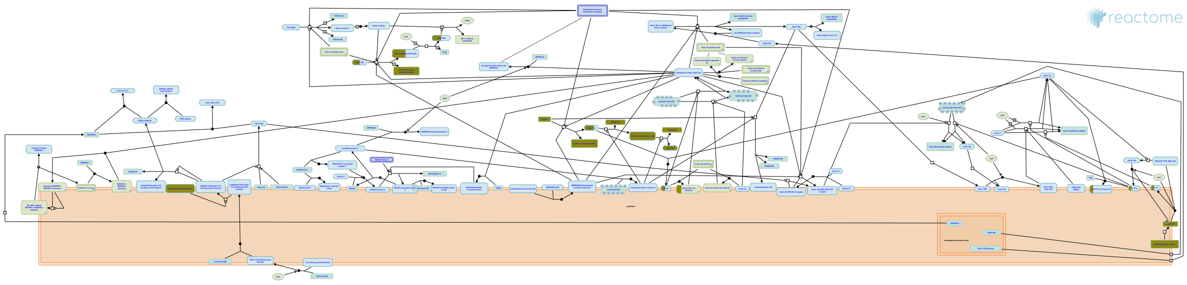

**Cellular compartments:** extracellular region.

The formation of a fibrin clot at the site of an injury to the wall of a normal blood vessel is an essential part of the process to stop blood loss after vascular injury. The reactions that lead to fibrin clot formation are commonly described as a cascade, in which the product of each step is an enzyme or cofactor needed for following reactions to proceed efficiently. The entire clotting cascade can be divided into three portions, the extrinsic pathway, the intrinsic pathway, and the common pathway. The extrinsic pathway begins with the release of tissue factor at the site of vascular injury and leads to the activation of factor X. The intrinsic pathway provides an alternative mechanism for activation of factor X, starting from the activation of factor XII. The common pathway consists of the steps linking the activation of factor X to the formation of a multimeric, cross-linked fibrin clot. Each of these pathways includes not only a cascade of events that generate the catalytic activities needed for clot formation, but also numerous positive and negative regulatory events.


##### Edit history

| Date | Action | Author |
|---|---|---|
| 2004-08-24 | Authored | D'Eustachio P |
| 2004-08-24 | Created | D'Eustachio P |
| 2021-05-22 | Modified | Shorser S |

##### Entities found in this pathway (4)

| Input | UniProt Id | Input | UniProt Id |
|---|---|---|---|
| F3 | P13726 | F5 | P12259 |
| PRCP | P42785 | TG | P00488 |

#### 9. Chylomicron clearance (R-HSA-8964026)

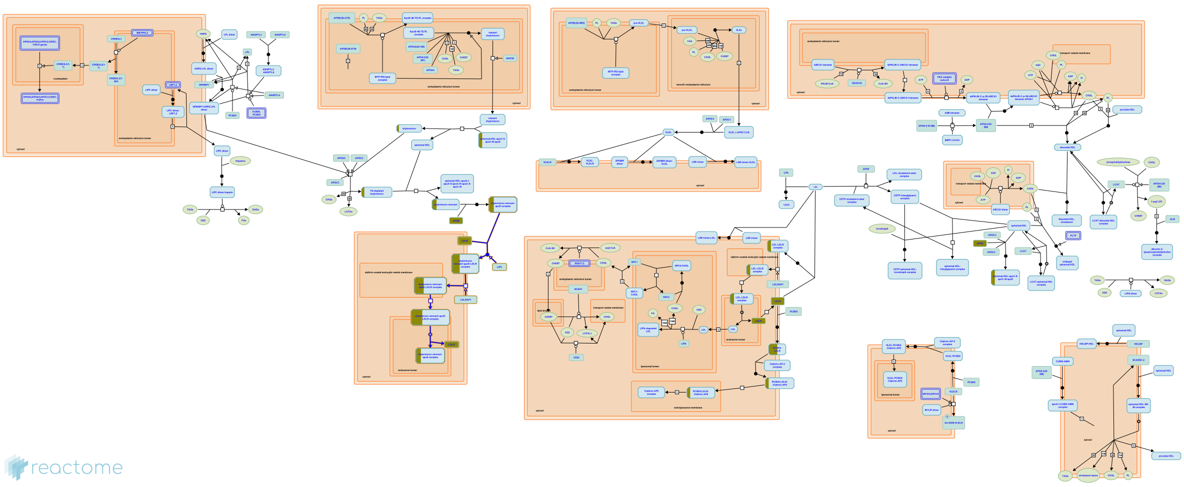

Circulating chylomicrons acquire molecules of apolipoproteins C and E and through interaction with endothelial lipases lose a large fraction of their triacylglycerol. These changes convert them to chylomicron remnants which bind to LDL receptors, primarily on the surfaces of liver cells, clearing them from the circulation.

This binding and clearance process involves several steps and requires the presence of heparan sulfate proteoglycan (HSPG)-associated hepatic lipase (HL). The molecular details of LDLR binding, and of the following steps of remnant endocytosis, are inferred from those of the coorresponding step of LDLR-mediated low-density lipoprotein (LDL) endocytosis (Redgrave 2004).

##### Edit history

| Date | Action | Author |
|---|---|---|
| 2007-04-30 | Edited | D'Eustachio P |
| 2007-04-30 | Authored | D'Eustachio P |
| 2016-01-27 | Reviewed | Jassal B |
| 2017-02-15 | Created | D'Eustachio P |
| 2021-05-31 | Modified | Shorser S |

##### Entities found in this pathway (2)

| Input | UniProt Id | Input | UniProt Id |
|---|---|---|---|
| APOE | P02649 | LDLR | P01130 |

#### 10. Nuclear signaling by ERBB4 (R-HSA-1251985)

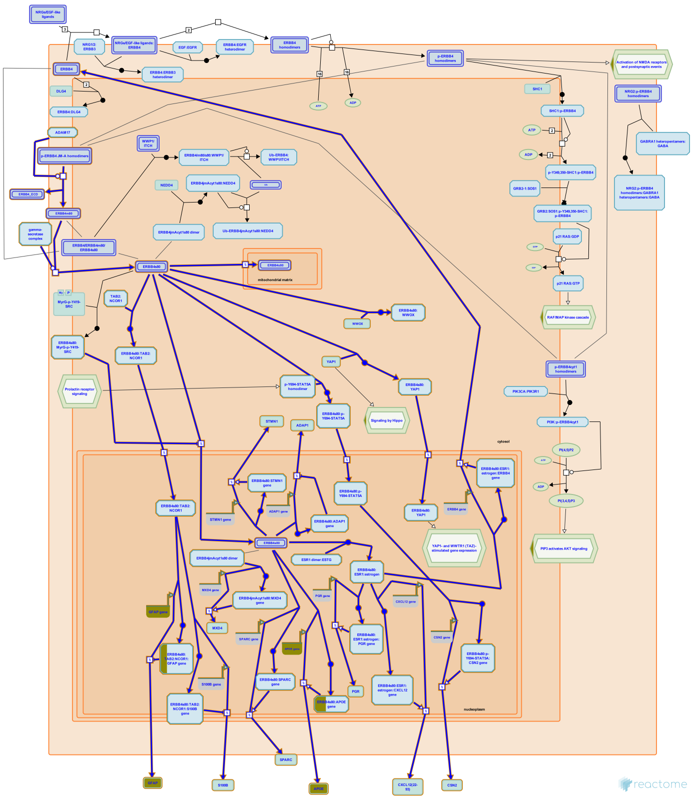

Besides signaling as a transmembrane receptor, ligand activated homodimers of ERBB4 JM-A isoforms (ERBB4 JM-A CYT1 and ERBB4 JM-A CYT2) undergo proteolytic cleavage by ADAM17 (TACE) in the juxtamembrane region, resulting in shedding of the extracellular domain and formation of an 80 kDa membrane bound ERBB4 fragment known as ERBB4 m80 (Rio et al. 2000, Cheng et al. 2003). ERBB4 m80 undergoes further proteolytic cleavage, mediated by the gamma-secretase complex, which releases the soluble 80 kDa ERBB4 intracellular domain, known as ERBB4 s80 or E4ICD, into the cytosol (Ni et al. 2001). ERBB4 s80 is able to translocate to the nucleus, promote nuclear translocation of various transcription factors, and act as a transcription co-factor. In neuronal precursors, ERBB4 s80 binds the complex of TAB and NCOR1, helps to move the complex into the nucleus, and is a co-factor of TAB:NCOR1-mediated inhibition of expression of astrocyte differentiation genes GFAP and S100B (Sardi et al. 2006). In mammary cells, ERBB4 s80 recruits STAT5A transcription factor in the cytosol, shuttles it to the nucleus, and acts as the STAT5A co-factor in binding to and promoting transcription from the beta-casein (CSN2) promoter, and may be involved in the regulation of other lactation-related genes (Williams et al. 2004, Muraoka-Cook et al. 2008). ERBB4 s80 was also shown to bind activated estrogen receptor in the nucleus and act as its transcriptional co-factor in promoting transcription of some estrogen-regulated genes, such as progesterone receptor gene NR3C3 and CXCL12 i.e. SDF1 (Zhu et al. 2006). ERBB4s80 may inhibit transcription of telomerase reverse transcriptase (TERT) by increasing methylation of the TERT gene promoter through an unknown mechanism (Ishibashi et al. 2012).

The C-tail of ERBB4 possesses several WW-domain binding motifs (three in CYT1 isoform and two in CYT2 isoform), which enable interaction of ERBB4 with WW-domain containing proteins. ERBB4 s80, through WW-domain binding motifs, interacts with YAP1 transcription factor, a known proto-oncogene, and may be a co-regulator of YAP1-mediated transcription (Komuro et al. 2003, Omerovic et al. 2004). The tumor suppressor WWOX, another WW-domain containing protein, competes with YAP1 in binding to ERBB4 s80 and prevents translocation of ERBB4 s80 to the nucleus (Aqeilan et al. 2005). ERBB4 s80 is also able to translocate to the mitochondrial matrix, presumably when its nuclear translocation is inhibited. Once in the mitochondrion, the BH3 domain of ERBB4, characteristic of BCL2 family members, may enable it to act as a pro-apoptotic factor (Naresh et al. 2006).


#### Edit history

| Date | Action | Author |
|---|---|---|
| 2011-04-26 | Created | Orlic-Milacic M |
| 2011-11-04 | Authored | Orlic-Milacic M |
| 2011-11-07 | Edited | Matthews L |
| 2011-11-11 | Reviewed | Zeng F, Harris RC |
| 2012-02-20 | Reviewed | Earp HS 3rd, Misior AM |
| 2018-06-28 | Revised | Orlic-Milacic M |
| 2019-02-21 | Revised | Stern DF |
| 2019-02-21 | Authored | Stern DF |
| 2019-03-06 | Edited | Orlic-Milacic M |
| 2021-05-31 | Modified | Shorser S |

#### Entities found in this pathway (2)

| Input | UniProt Id | Input | UniProt Id |
|---|---|---|---|
| APOE | P02649 | GFAP | P14136 |

| Input | Ensembl Id | Input | Ensembl Id |
|---|---|---|---|
| APOE | ENSG00000130203 | GFAP | ENSG00000131095 |

#### 11. Syndecan interactions (R-HSA-3000170)

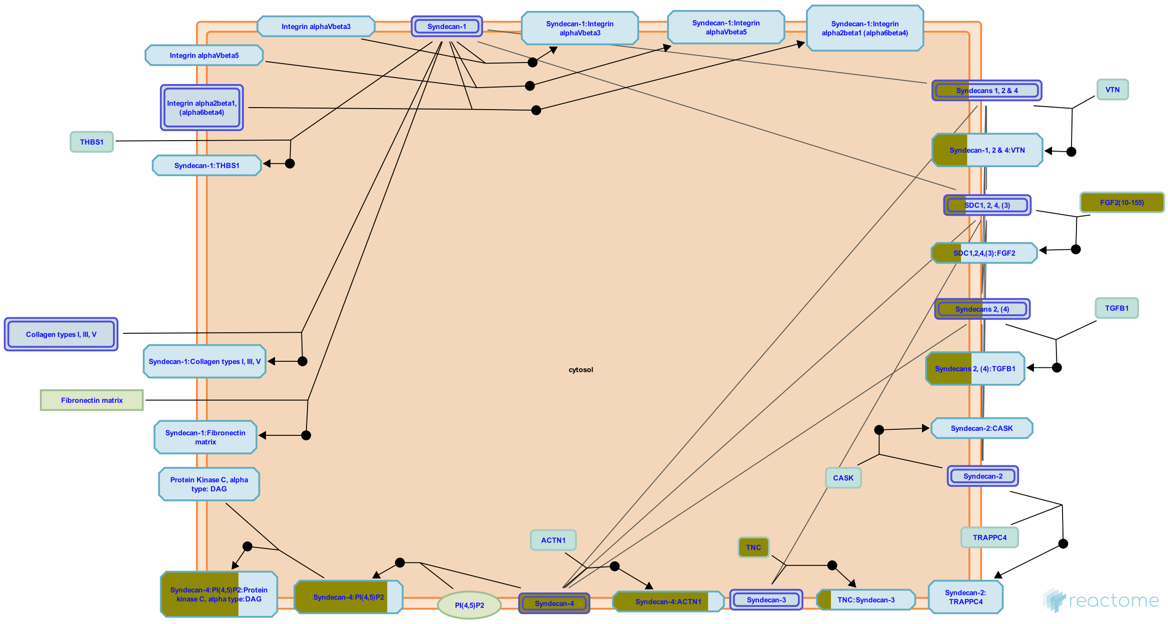


Syndecans are type I transmembrane proteins, with an N-terminal ectodomain that contains several consensus sequences for glycosaminoglycan (GAG) attachment and a short C-terminal cytoplasmic domain. Syndecan-1 and -3 GAG attachment sites occur in two distinct clusters, one near the N-terminus and the other near the membrane-attachment site, separated by a proline and threonine-rich 'spacer'. Syndecan ectodomain sequences are poorly conserved in the family and between species, but the transmembrane and cytoplasmic domains are highly conserved. Syndecan-1 and -3 form a subfamily. Syndecan core proteins form dimers (Choi et al. 2007) and at least syndecan-3 and -4 form oligomers (Asundi & Carey 1995, Shin et al. 2012). Syndecan-1 is the major syndecan of epithelial cells including vascular endothelium. Syndecan-2 is present mostly in mesenchymal, neuronal and smooth muscle cells. Syndecan-3 is the major syndecan of the nervous system, while syndecan-4 is ubiquitously expressed but at lower levels than the other syndecans (refs in Alexopoulou et al. 2007). The core syndecan protein has three to five heparan sulfate or chondroitin sulfate chains, which interact with a variety of ligands including fibroblast growth factors, vascular endothelial growth factor, transforming growth factor-beta, fibronectin, collagen, vitronectin and several integrins. Syndecans may act as integrin coreceptors. Interactions between fibronectin and syndecans are modulated by tenascin-C.

Syndecans bind a wide variety of soluble and insoluble ligands, inckluding extracellular matrix components, cell adhesion molecules, growth factors, cytokines, and proteinases. As the cleaved ectodomains of syndecans retain the ability to bind ligands, ectodomain shedding is a mechanism for releasing soluble effectors that may compete for ligands with their cell-bound counterparts (Kainulainen et al. 1998). Shed ectodomains are found in inflammatory fluids (Subramanian et al. 1997) and may induce the proliferation of cancer cells (Maeda et al. 2004).

#### Edit history

| Date | Action | Author |
|---|---|---|
| 2012-07-31 | Authored | Jupe S |
| 2013-01-24 | Created | Jupe S |
| 2013-04-26 | Edited | Jupe S |
| 2013-05-22 | Reviewed | Ricard-Blum S, Fuentes J |
| 2021-05-22 | Modified | Shorser S |

#### Entities found in this pathway (3)

| Input | UniProt Id | Input | UniProt Id | Input | UniProt Id |
|---|---|---|---|---|---|
| FGF2 | P09038 | SDC4 | P31431 | TNC | P24821 |

#### 12. BH3-only proteins associate with and inactivate anti-apoptotic BCL-2 members (R-HSA-111453)

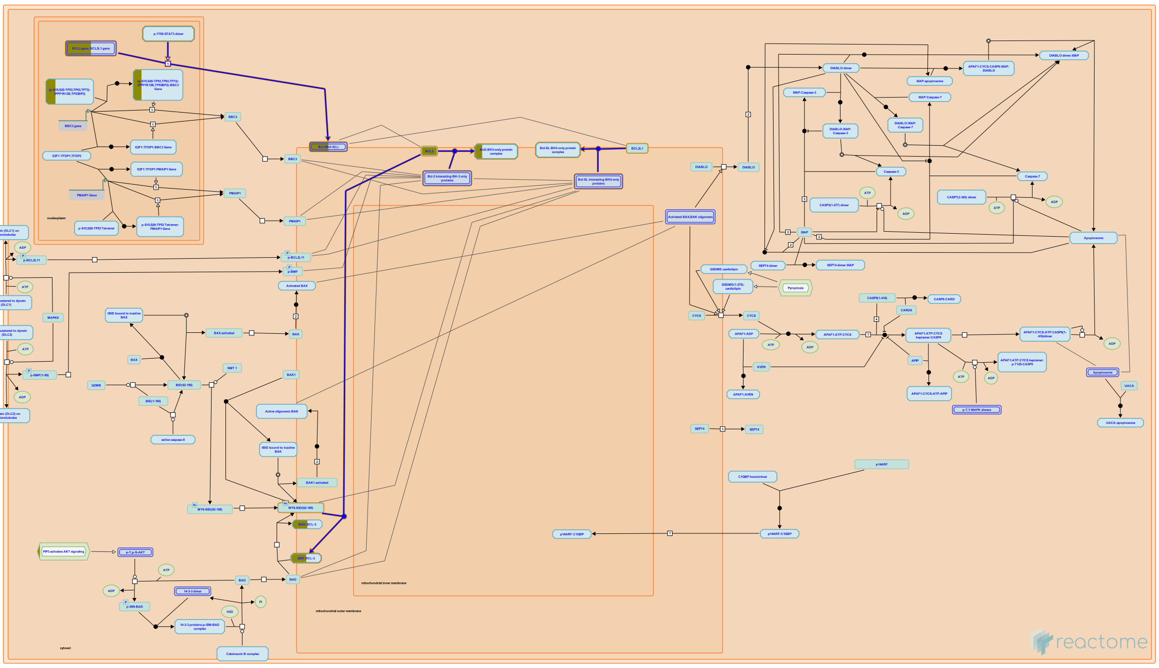

**Cellular compartments:** mitochondrial outer membrane.

Bcl-2 interacts with tBid (Yi et al. 2003), BIM (Puthalakath et al. 1999), PUMA (Nakano and Vousden 2001), NOXA (Oda et al. 2000), BAD (Yang et al. 2005), BMF (Puthalakath et al. 2001), resulting in inactivation of BCL2. Binding of BCL2 to tBID inhibits BID-induced cytochrome C release and apoptosis (Yi et al. 2003). BH3 only proteins associate with and inactivate anti-apoptotic BCL-XL.

##### References

##### Edit history

| Date | Action | Author |
|---|---|---|
| 2004-08-09 | Created | Tsujimoto Y, Hardwick JM |
| 2021-05-31 | Modified | Shorser S |

##### Entities found in this pathway (1)

| Input | UniProt Id |
|---|---|
| BCL2 | P10415 |

| Input | Ensembl Id |
|---|---|
| BCL2 | ENSG00000171791 |

#### 13. Signaling by Receptor Tyrosine Kinases (R-HSA-9006934)

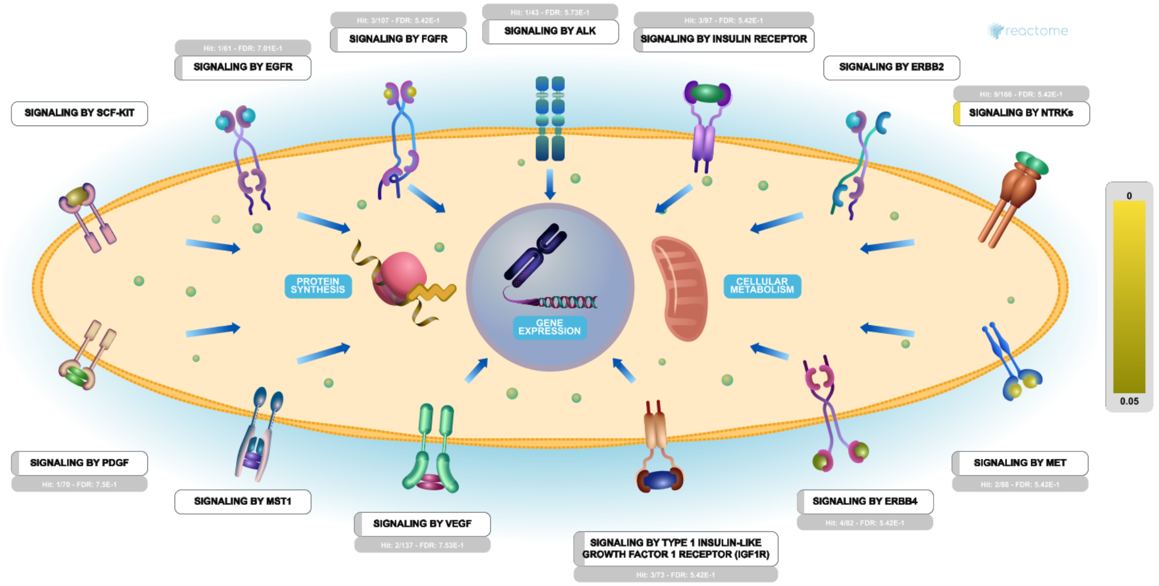


Receptor tyrosine kinases (RTKs) are a major class of cell surface proteins involved in Signal Transduction. Human cells contain ~60 RTKs, grouped into 20 subfamilies based on their domain architecture. All RTK subfamilies are characterized by an extracellular ligand-binding domain, a single transmembrane region and an intracellular region consisting of the tyrosine kinase domain and additional regulatory and protein interaction domains. In general, RTKs associate into dimers upon ligand binding and are activated by autophosphorylation on conserved intracellular tyrosine residues. Autophosphorylation increases the catalytic efficiency of the receptor and provides binding sites for the assembly of downstream signaling complexes (reveiwed in Lemmon and Schlessinger, 2010). Common signaling pathways activated downstream of RTK activation include RAF/MAP kinase cascades (reviewed in McKay and Morrison, 2007 and Wellbrock et al 2004), AKT signaling (reviewed in Manning and Cantley, 2007) and PLC-gamma mediated signaling (reviewed in Patterson et al). Activation of these pathways ultimately results in changes in gene expression and cellular metabolism.


##### Edit history

| Date | Action | Author |
|---|---|---|
| 2017-05-24 | Edited | Rothfels K |
| 2017-05-24 | Authored | Rothfels K |
| 2017-05-24 | Created | Rothfels K |
| 2017-06-22 | Reviewed | D'Eustachio P |
| 2021-05-22 | Modified | Shorser S |

#### Entities found in this pathway (13)

| Input | UniProt Id | Input | UniProt Id | Input | UniProt Id |
|---|---|---|---|---|---|
| ABI1 | Q8IZP0 | APOE | P02649 | ARC | Q7LC44 |
| COL6A3 | P12111 | F3 | P13726 | FGF2 | P09038 |
| FRS2 | Q8WU20 | GFAP | P14136 | ID4 | P47928 |
| KAL1 | P23352 | LAMA1 | P25391 | LRIG1 | Q96JA1 |
| TRIB1 | Q96RU7, Q96RU8 | | | | |

| Input | Ensembl Id | Input | Ensembl Id | Input | Ensembl Id |
|---|---|---|---|---|---|
| APOE | ENSG00000130203 | ARC | ENSG00000198576 | F3 | ENSG00000117525 |
| GFAP | ENSG00000131095 | ID4 | ENSG00000172201 | TRIB1 | ENSG00000173334 |

#### 14. Interleukin-4 and Interleukin-13 signaling (R-HSA-6785807)

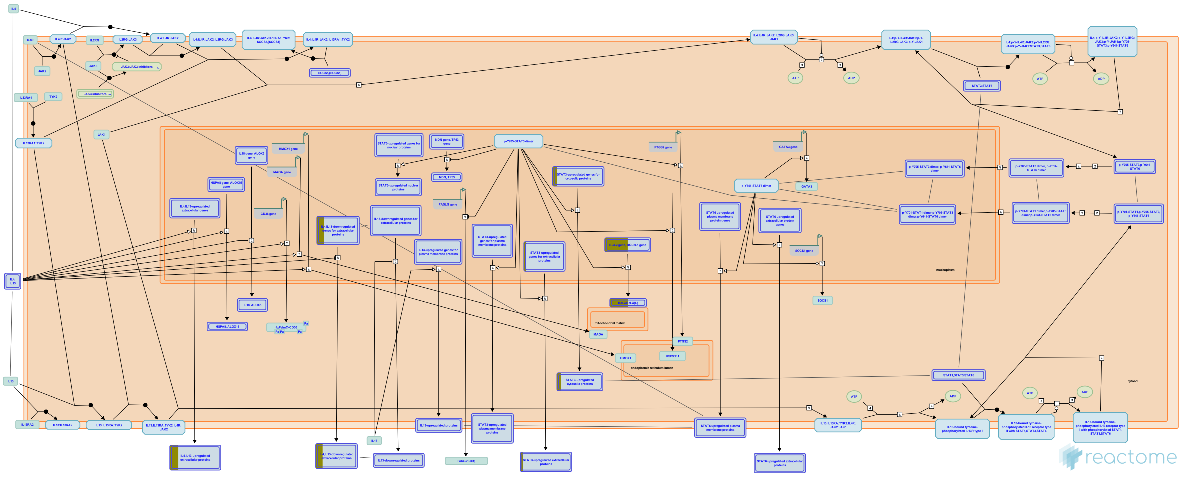

Interleukin-4 (IL4) is a principal regulatory cytokine during the immune response, crucially important in allergy and asthma (Nelms et al. 1999). When resting T cells are antigen-activated and expand in response to Interleukin-2 (IL2), they can differentiate as Type 1 (Th1) or Type 2 (Th2) T helper cells. The outcome is influenced by IL4. Th2 cells secrete IL4, which both stimulates Th2 in an autocrine fashion and acts as a potent B cell growth factor to promote humoral immunity (Nelms et al. 1999).

| Input | UniProt Id |
|---|---|
| BCL2 | P10415 |
| MCL1 | Q07820 |

| Input | UniProt Id |
|---|---|
| CCL2 | P13500 |
| TG | P00488 |

| Input | UniProt Id |
|---|---|
| FGF2 | P09038 |

| Input | Ensembl Id |
|---|---|
| BCL2 | ENSG00000171791 |
| FGF2 | ENSG00000138685 |

| Input | Ensembl Id |
|---|---|
| CCL2 | ENSG00000108691 |
| MCL1 | ENSG00000143384 |

#### 15. Linoleic acid (LA) metabolism (R-HSA-2046105)

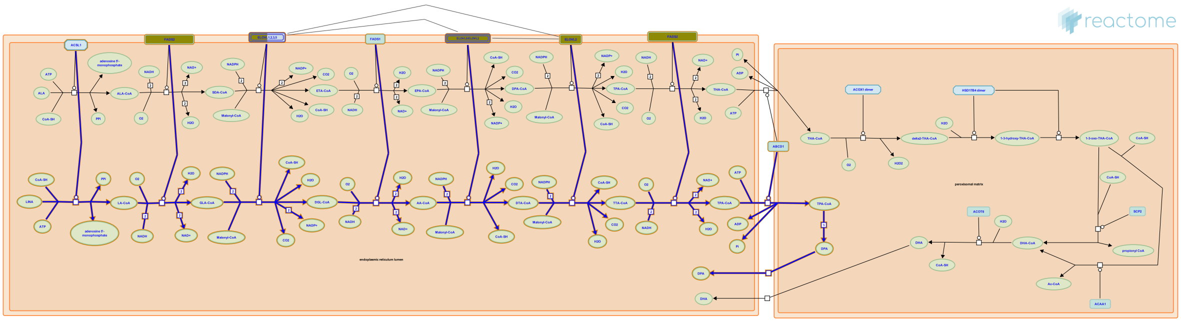

**Cellular compartments:** endoplasmic reticulum lumen, endoplasmic reticulum membrane, peroxisomal matrix, peroxisomal membrane.

Linoleic acid (LA, 18:2(n-6)) is an omega-6 fatty acid obtained through diet, mainly from vegetable oils. Omega-6 fatty acids helps stimulate skin and hair growth, maintain bone health, regulate metabolism, and maintain the reproductive system. All the desaturation and elongation steps occur in the endoplasmic reticulum (ER) except for the final step which requires translocation to peroxisomes for partial beta-oxidation. The linoleic acid pathway involves the following steps: 18:2(n-6)-->18:3(n-6)--> 20:3(n-6)-->20:4(n-6)-->22:4(n-6)-->24:4(n-6)-->24:5(n-6)-->22:5(n-6). Two desaturation enzymes are involved in this process: delta-6 desaturase which converts 18:2(n-6) to 18:3 (n-6) and 24:4(n-6) to 24:5(n-6) respectively, and delta-5 desaturase which converts 20:3(n-6) to 20:4(n-6). (Sprecher 2002).


##### Edit history

| Date | Action | Author |
|---|---|---|
| 2012-01-11 | Edited | Garapati P V |
| 2012-01-11 | Authored | Garapati P V |
| 2012-01-11 | Created | Garapati P V |
| 2021-05-22 | Modified | Shorser S |

##### Entities found in this pathway (2)

| Input | UniProt Id | Input | UniProt Id |
|---|---|---|---|
| ELOVL2 | Q9NXB9, Q9NYP7 | FADS2 | O95864 |

#### 16. Nuclear Envelope (NE) Reassembly (R-HSA-2995410)

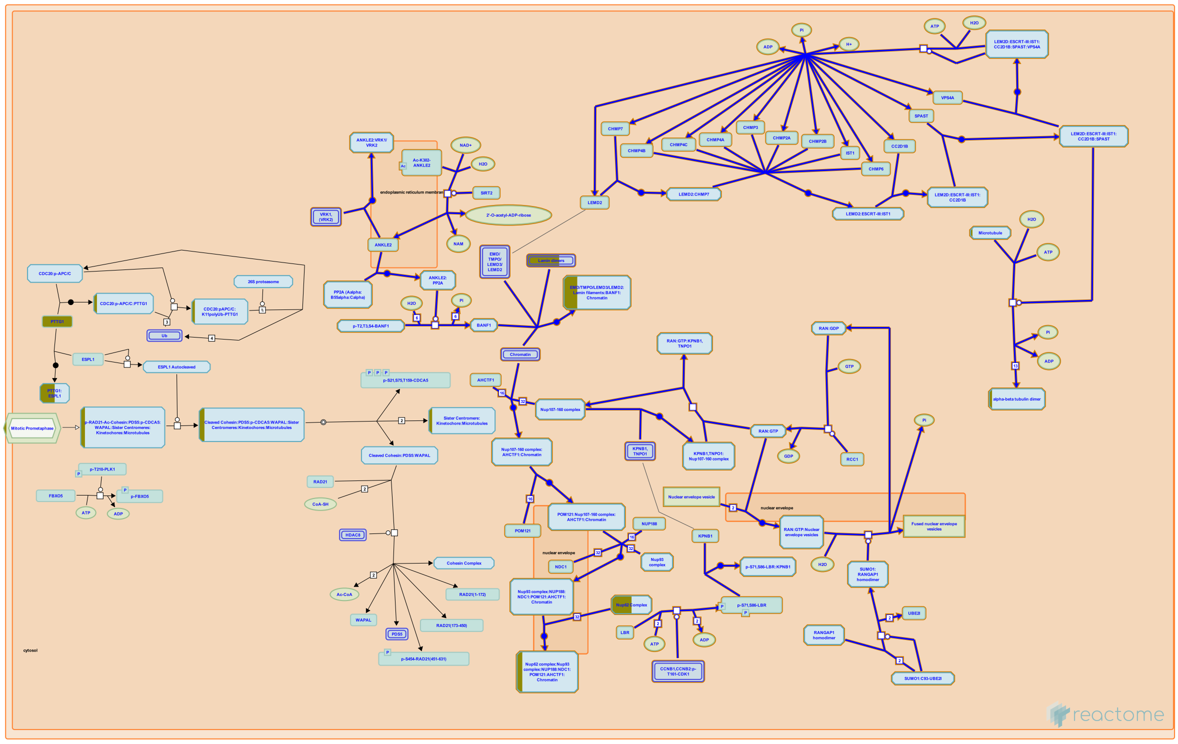

Reassembly of the nuclear envelope (NE) around separated sister chromatids begins in late anaphase and is completed in telophase (reviewed by Wandke and Kutay 2013). Characteristic proteins of the inner nuclear membrane and nuclear lamina accumulate at the reforming NE (reviewed by Wandke and Kutay 2013). Concurrently, nuclear pore complexes (NPCs) assemble and insert into the reforming NE, and the NE becomes sealed to reestablish the nucleocytoplasmic diffusion barrier (reviewed by Otsuka and Ellenberg 2018).


##### Edit history

| Date | Action | Author |
|---|---|---|
| 2013-01-23 | Edited | Gillespie ME |
| 2013-01-23 | Authored | Orlic-Milacic M |
| 2013-01-23 | Created | Orlic-Milacic M |
| 2019-11-07 | Authored | Gerace L |
| 2019-11-26 | Edited | Orlic-Milacic M |
| 2021-05-22 | Modified | Shorser S |

##### Entities found in this pathway (3)

| Input | UniProt Id | Input | UniProt Id | Input | UniProt Id |
|---|---|---|---|---|---|
| LMNA | P02545-1, P02545-2 | NUPL1 | Q9BVL2-1, Q9BVL2-2 | TUBA1A | Q71U36 |

#### 17. Non-integrin membrane-ECM interactions (R-HSA-3000171)

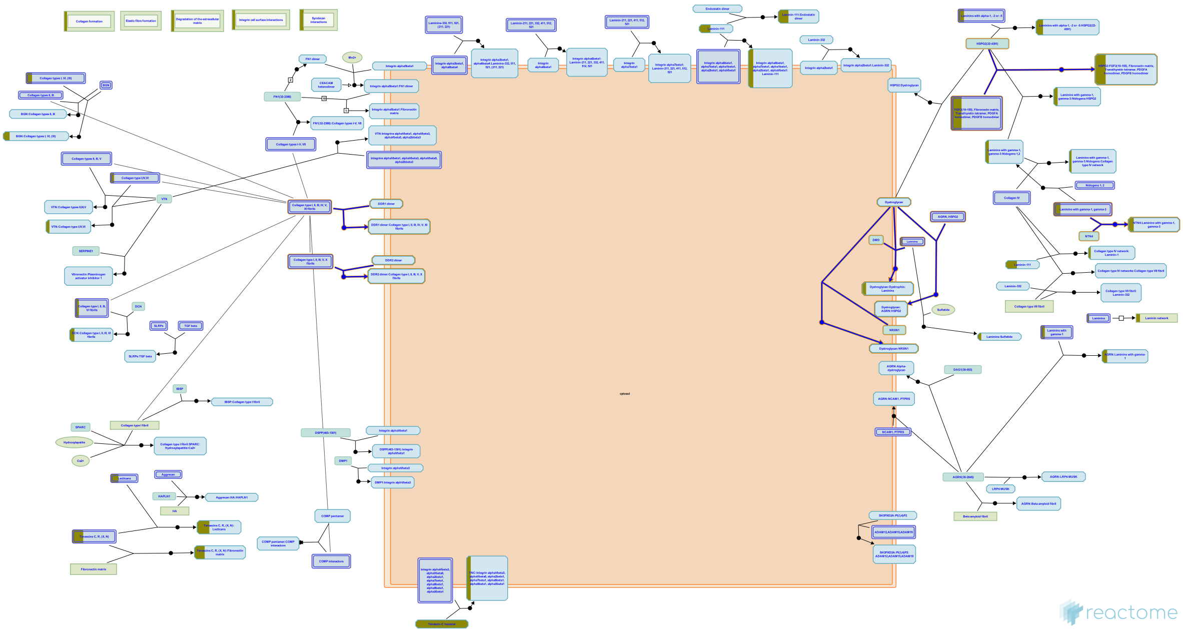

Several non-integrin membrane proteins interact with extracellular matrix proteins. Transmembrane proteoglycans may associate with integrins and growth factor receptors to influence their function, or they can signal independently, often influencing the actin cytoskeleton.


##### Edit history

| Date | Action | Author |
|---|---|---|
| 2012-07-31 | Authored | Jupe S |
| 2013-01-24 | Created | Jupe S |
| 2013-04-26 | Edited | Jupe S |
| 2013-05-22 | Reviewed | Ricard-Blum S |
| 2021-05-22 | Modified | Shorser S |

##### Entities found in this pathway (4)

| Input | UniProt Id |
|---|---|
| FGF2 | P09038 |
| SDC4 | P31431 |

| Input | UniProt Id |
|---|---|
| LAMA1 | P25391 |
| TNC | P24821 |

#### 18. FGFR1c ligand binding and activation (R-HSA-190373)

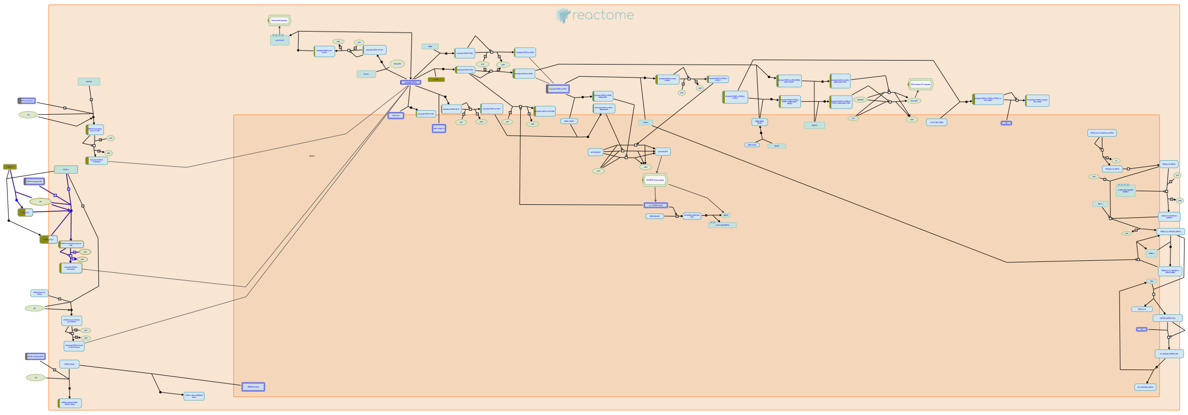

This pathway depicts the binding of an experimentally-verified range of ligands to FGFR1c. While binding affinities may vary considerably within this set, the ligands listed have been established to bring about receptor activation at their reported physiological concentrations.


##### Edit history

| Date | Action | Author |
|---|---|---|
| 2007-01-02 | Created | de Bono B |
| 2007-01-10 | Authored | de Bono B |
| 2007-02-07 | Reviewed | Mohammadi M |
| 2016-01-06 | Reviewed | Grose RP |
| 2021-05-22 | Modified | Shorser S |

##### Entities found in this pathway (2)

| Input | UniProt Id | Input | UniProt Id |
|---|---|---|---|
| FGF2 | P09038 | KAL1 | P23352 |

#### 19. Vitamin E (R-HSA-8877627)

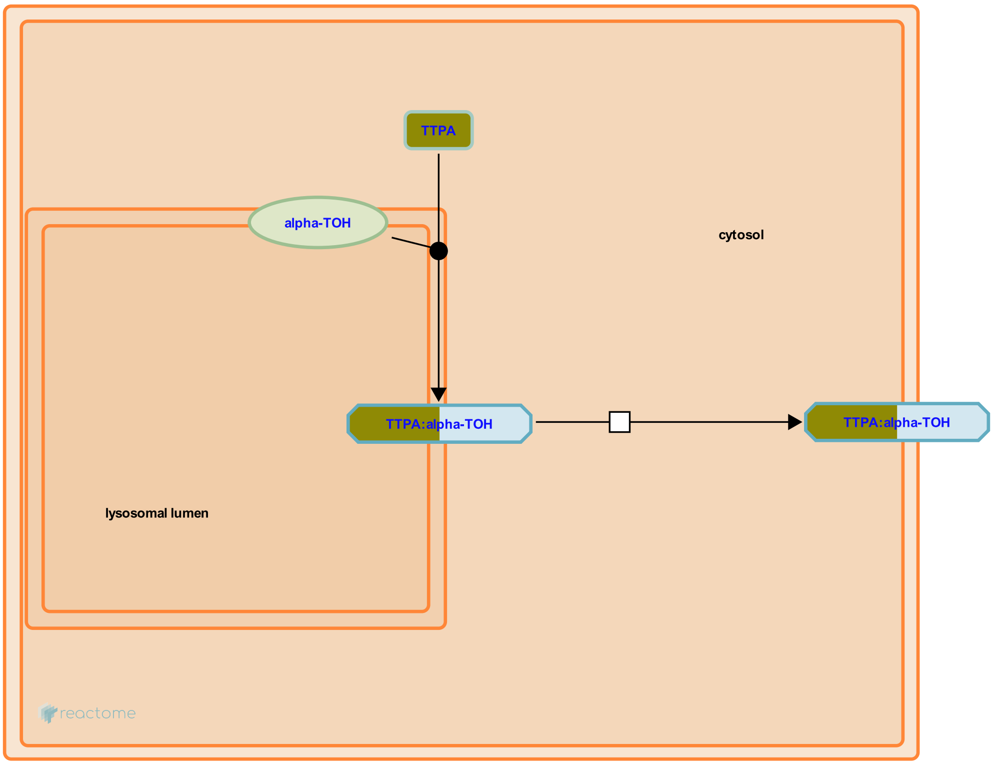


Vitamins A, D, E and K are lipophilic compounds, the so-called fat-soluble vitamins. Because of their lipophilicity, fat-soluble vitamins are solubilised and transported by intracellular carrier proteins to exert their actions. Alpha-tocopherol, the main form of vitamin E found in the body, is transported by alpha-tocopherol transfer protein (TTPA) in hepatic cells (Kono & Arai 2015, Schmolz et al. 2016).

##### Edit history

| Date | Action | Author |
|---|---|---|
| 2016-06-27 | Edited | Jassal B |
| 2016-06-27 | Authored | Jassal B |
| 2016-06-27 | Created | Jassal B |
| 2016-07-15 | Reviewed | D'Eustachio P |
| 2021-05-22 | Modified | Shorser S |

##### Entities found in this pathway (1)

| Input | UniProt Id |
|---|---|
| TTPA | P49638 |

#### 20. Nuclear Envelope Breakdown (R-HSA-2980766)

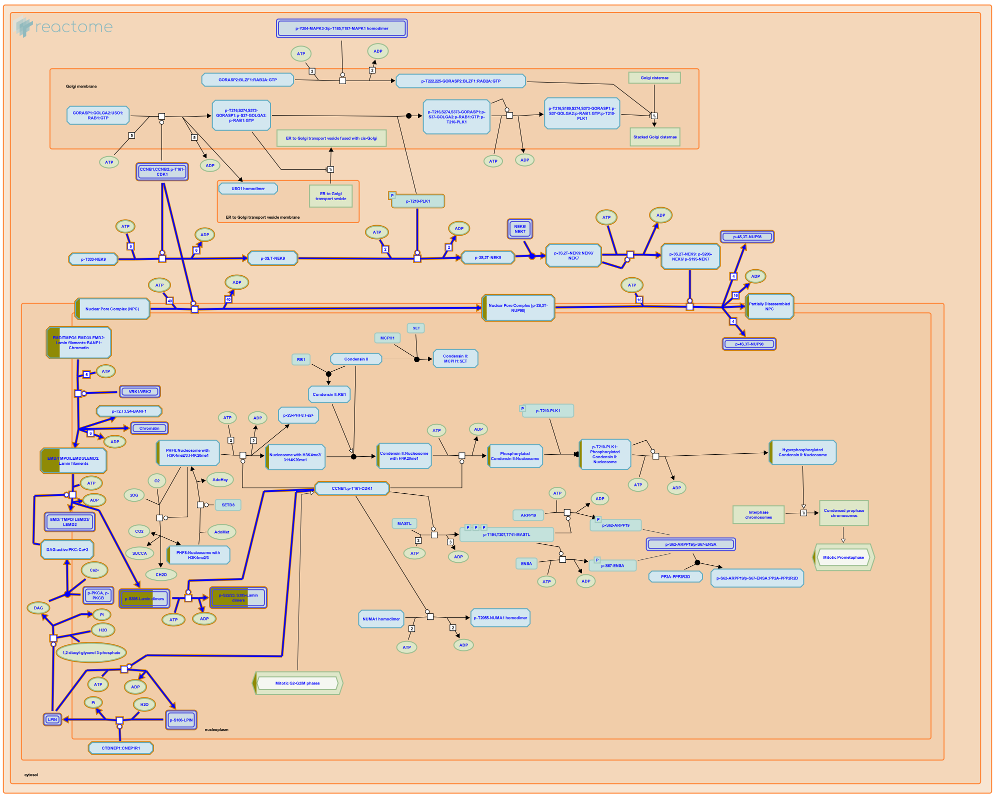

The nuclear envelope breakdown (NEBD) happens in late prophase of mitosis and involves disassembly of the nuclear pore complex, depolymerization of the nuclear lamina, and clearance of nuclear envelope from chromatin. NEBD allows mitotic spindle microtubules to access condensed chromosomes at kinetochores and enables nuclear division and segregation of genetic material to two daughter cells. For a recent review, please refer to Guttinger et al. 2009.

In mitotic prophase, chromatin detaches from the nuclear envelope, and this contributes to the nuclear envelope breakdown. VRK1 (and possibly VRK2) mediated phosphorylation of BANF1 (BAF), a protein that simultaneously interacts with DNA, LEM-domain inner nuclear membrane proteins, and lamins (Zheng et al. 2000, Shumaker et al. 2001, Haraguchi et al. 2001, Mansharamani and Wilson 2005, Brachner et al. 2005) is considered to be one of the key steps in the detachment of the nuclear envelope from chromatin (Bengtsson and Wilson 2006, Nichols et al. 2006, Gorjanacz et al. 2007).


##### Edit history

| Date | Action | Author |
|---|---|---|
| 2013-01-12 | Created | Orlic-Milacic M |
| 2013-01-23 | Edited | Gillespie ME |
| 2013-01-23 | Authored | Orlic-Milacic M |
| 2013-01-30 | Reviewed | Gorjánácz M, Mattaj IW |

| Date | Action | Author |
|---|---|---|
| 2021-05-22 | Modified | Shorser S |

#### Entities found in this pathway (2)

| Input | UniProt Id | Input | UniProt Id |
|---|---|---|---|
| LMNA | P02545-1, P02545-2 | NUPL1 | Q9BVL2-1, Q9BVL2-2 |

#### 21. Negative regulation of FGFR1 signaling (R-HSA-5654726)

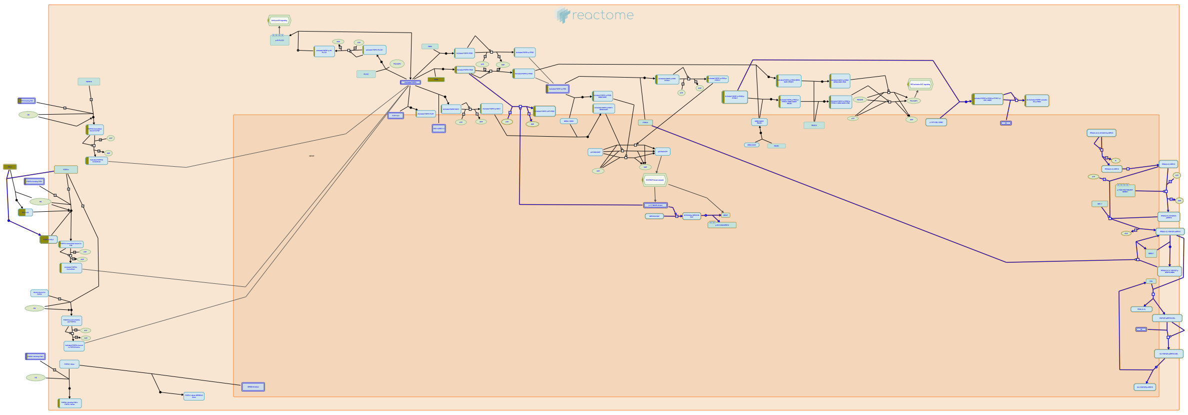

**Cellular compartments:** cytosol, extracellular region, plasma membrane.

Once activated, the FGFR signaling pathway is regulated by numerous negative feedback mechanisms. These include downregulation of receptors through CBL-mediated ubiquitination and endocytosis, ERK-mediated inhibition of FRS2-tyrosine phosphorylation and the attenuation of ERK signaling through the action of dual-specificity phosphatases, IL17RD/SEF, Sprouty and Spred proteins. A number of these inhibitors are themselves transcriptional targets of the activated FGFR pathway.


##### Edit history

| Date | Action | Author |
|---|---|---|
| 2011-08-15 | Authored | Rothfels K |
| 2011-08-26 | Reviewed | Gotoh N |
| 2014-12-04 | Created | Rothfels K |
| 2016-01-06 | Reviewed | Grose RP |
| 2021-05-22 | Modified | Shorser S |

##### Entities found in this pathway (3)

| Input | UniProt Id | Input | UniProt Id | Input | UniProt Id |
|---|---|---|---|---|---|
| FGF2 | P09038 | FRS2 | Q8WU20 | KAL1 | P23352 |

#### 22. Aquaporin-mediated transport (R-HSA-445717)

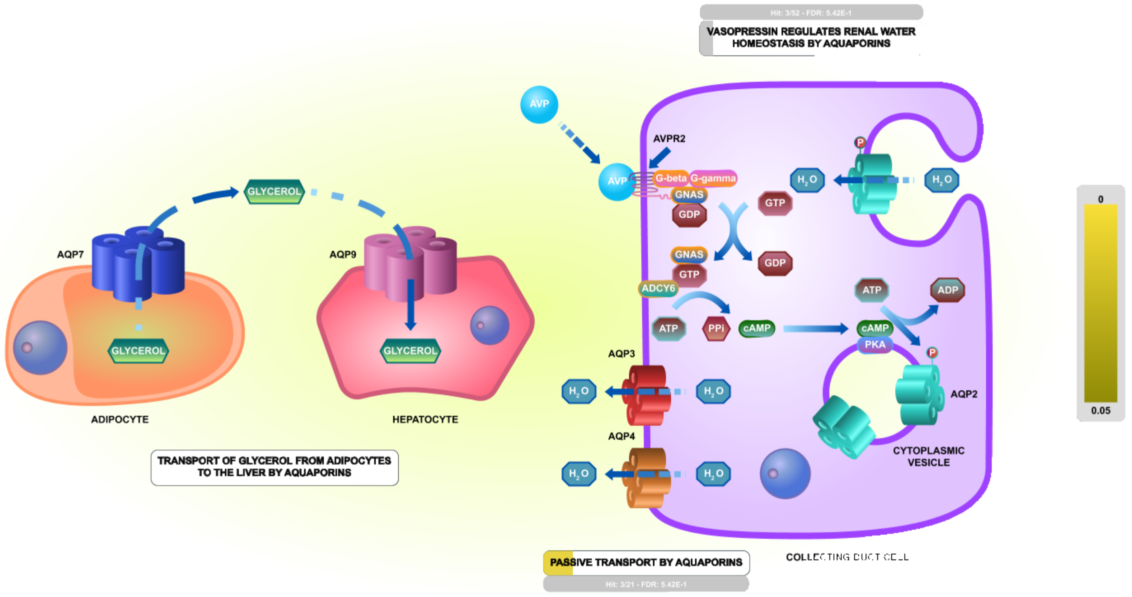


**Cellular compartments:** plasma membrane.

Aquaporins (AQP's) are six-pass transmembrane proteins that form channels in membranes. Each monomer contains a central channel formed in part by two asparagine-proline-alanine motifs (NPA boxes) that confer selectivity for water and/or solutes. The monomers assemble into tetramers. During passive transport by Aquaporins most aquaporins (i.e. AQP0/MIP, AQP1, AQP2, AQP3, AQP4, AQP5, AQP7, AQP8, AQP9, AQP10) transport water into and out of cells according to the osmotic gradient across the membrane. Four aquaporins (the aquaglyceroporins AQP3, AQP7, AQP9, AQP10) conduct glycerol, three aquaporins (AQP7, AQP9, AQP10) conduct urea, and one aquaporin (AQP6) conducts anions, especially nitrate. AQP8 also conducts ammonia in addition to water.


#### Edit history

| Date | Action | Author |
|---|---|---|
| 2009-11-01 | Edited | May B |
| 2009-11-01 | Authored | May B |
| 2009-11-05 | Created | May B |
| 2010-06-24 | Reviewed | Beitz E |
| 2010-07-15 | Reviewed | Calamita G |
| 2010-07-31 | Reviewed | Mathai JC, MacIver B |
| 2021-05-22 | Modified | Shorser S |

#### Entities found in this pathway (4)

| Input | UniProt Id |
|---|---|
| ADCY8 | P40145 |
| AQP4 | P55087 |

| Input | UniProt Id |
|---|---|
| AQP1 | P29972 |
| AQP6 | Q13520 |

#### 23. Amino acid transport across the plasma membrane (R-HSA-352230)

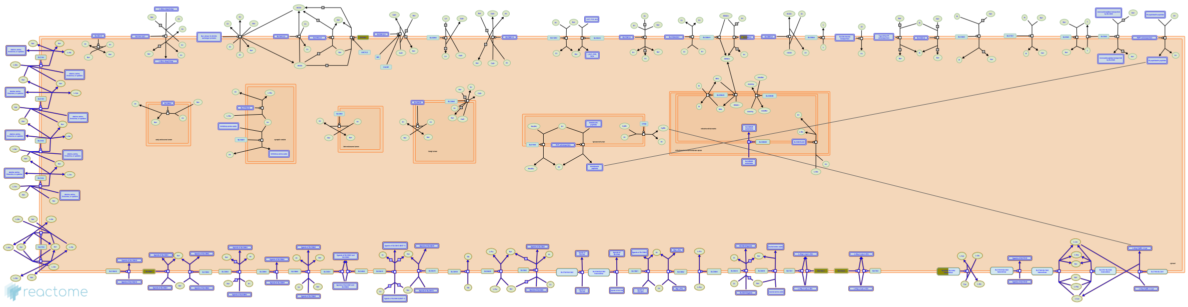

**Cellular compartments:** plasma membrane.

Amino acid transport across plasma membranes is critical to the uptake of these molecules from the gut, to their reabsortion in the kidney proximal tubulues, and to their distribution to cells in which they are required for the synthesis of proteins and of amino acid derived small molecules such as neurotransmitters. Physiological studies have defined 18 "systems" that mediate amino acid transport, each characterized by its amino acid substrates, as well as its pH sensitivity and its association (or not) with ion transport. More recently, molecular cloning studies have allowed the identification of the plasma membrane transport proteins that mediate these reactions. Amino acid uptake mediated by 17 of these transporters is annotated here (Broer 2008).


##### Edit history

| Date | Action | Author |
|---|---|---|
| 2008-06-02 | Created | D'Eustachio P |
| 2008-06-03 | Edited | D'Eustachio P |
| 2008-06-03 | Reviewed | Jassal B |
| 2008-06-03 | Authored | D'Eustachio P |
| 2021-05-22 | Modified | Shorser S |

##### Entities found in this pathway (3)

| Input | UniProt Id | Input | UniProt Id | Input | UniProt Id |
|---|---|---|---|---|---|
| SAT1 | Q9H2H9 | SLC7A11 | Q9UPY5 | SLC7A2 | P52569-1, P52569-2 |

#### 24. Glycogen metabolism (R-HSA-8982491)

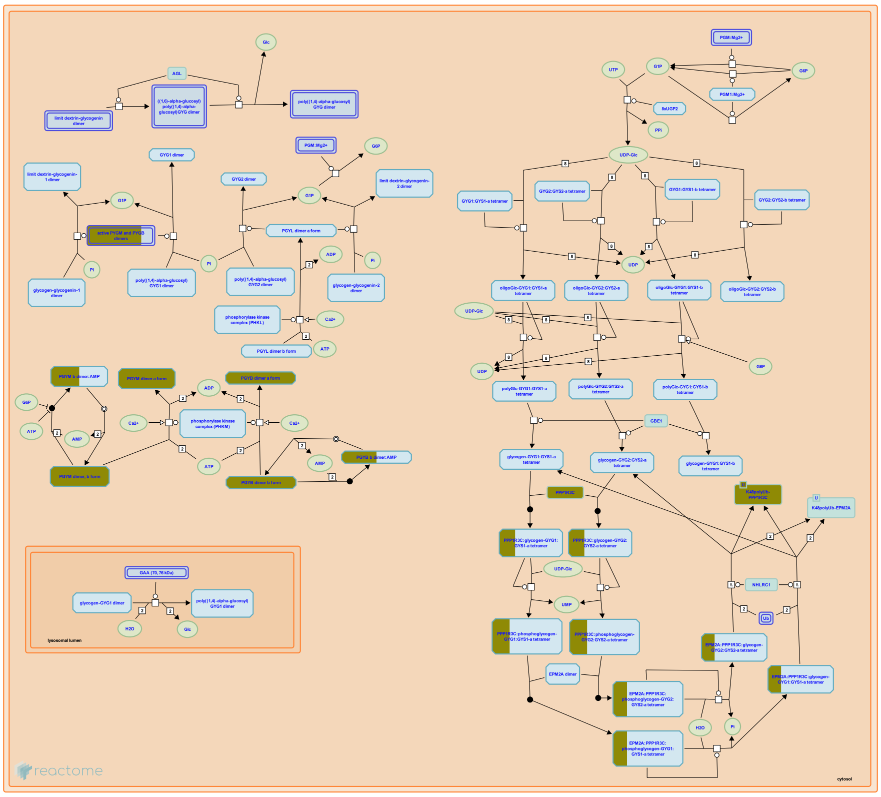

Glycogen, a highly branched glucose polymer, is formed and broken down in most human tissues, but is most abundant in liver and muscle, where it serves as a major stored fuel. Glycogen metabolism has been studied in most detail in liver and skeletal muscle. Glycogen metabolism in other tissues has not been studied as extensively, and is thought to resemble the muscle process.

Glycogen synthesis involves five reactions. The first two, conversion of glucose 6-phosphate to glucose 1-phosphate and synthesis of UDP-glucose from glucose 1-phosphate and UTP, are shared with several other pathways. The next three reactions, the auto-catalyzed synthesis of a glucose oligomer on glycogenin, the linear extension of the glucose oligomer catalyzed by glycogen synthase, and the formation of branches catalyzed by glycogen branching enzyme, are unique to glycogen synthesis. Repetition of the last two reactions generates large, extensively branched glycogen polymers. The catalysis of glycogenin glucosylation and oligoglucose chain extension by distinct isozymes in liver and nonhepatic tissues allows them to be regulated independently (Agius 2008; Bollen et al. 1998; Roach et al. 2012).

Cytosolic glycogen breakdown occurs via the same chemical steps in all tissues but is separately regulated via tissue specific isozymes and signaling pathways that enable distinct physiological fates for glycogen in liver and other tissues. Glycogen phosphorylase, which can be activated by phosphorylase kinase, catalyzes the removal of glucose residues as glucose 1-phosphate from the ends of glycogen branches. The final four residues of each branch are removed in two steps catalyzed by debranching enzyme, and further glycogen phosphorylase activity completes the process of glycogen breakdown. The first glucose residue in each branch is released as free glucose; all other residues are released as glucose 1-phosphate. The latter molecule can be converted to glucose 6-phosphate in a step shared with other pathways (Villar-Palasi & Larner 1970; Hers 1976).

Glycogen can also be taken up into lysosomes, where it is normally broken done by the action of a single enzyme, lysosomal alpha-glucosidase (GAA) (Brown et al. 1970).

#### Edit history

| Date | Action | Author |
|---|---|---|
| 2003-02-15 | Authored | |
| 2013-07-26 | Reviewed | Jassal B |
| 2017-03-18 | Edited | D'Eustachio P |
| 2017-03-18 | Created | D'Eustachio P |
| 2021-05-22 | Modified | Shorser S |

#### Entities found in this pathway (2)

| Input | UniProt Id | Input | UniProt Id |
|---|---|---|---|
| PPP1R3C | Q9UQK1 | PYGM | P11216, P11217 |

#### 25. Defective SLC4A4 causes renal tubular acidosis, proximal, with ocular abnormalities and mental retardation (pRTA-OA) (R-HSA-5619054)

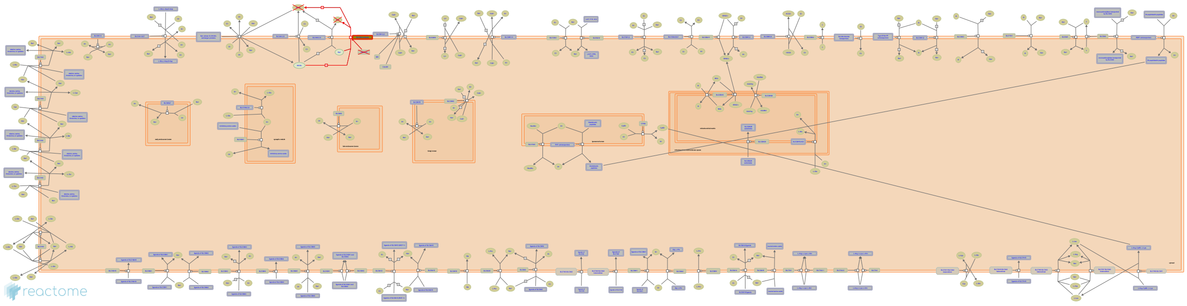

**Diseases:** renal tubular acidosis.

Members 4, 5, 7 and 9 of the SLC4A family couple the transport of bicarbonate (HCO3-) with sodium ions (Na+). SLC4A4 (aka NBCe1) is an electrogenic Na+/HCO3- cotransporter with a stoichiometry of 1:3. SLC4A4 is expressed in the kidney and pancreas, with lesser expression in many other tissues. Mutations in SLC4A4 can cause permanent isolated proximal renal tubular acidosis with ocular abnormalities and mental retardation (pRTA-OA), a rare autosomal recessive syndrome characterised by short stature, proximal renal tubular acidosis, mental retardation, bilateral glaucoma, cataracts and bandkeratopathy. pRTA results from the failure of the proximal tubular cells to reabsorb filtered HCO3- from urine, leading to urinary HCO3- wasting and subsequent acidemia. HCO3- also needs to move out of cells in the eye, thus failure to do so can affect ocular pressure homeostasis (Horita et al. 2005, Kurtz & Zhu 2013, Kurtz & Zhu 2013b, Seki et al. 2013).


##### Edit history

| Date | Action | Author |
|---|---|---|
| 2014-08-22 | Edited | Jassal B |
| 2014-08-22 | Authored | Jassal B |
| 2014-08-22 | Created | Jassal B |
| 2015-08-04 | Reviewed | Broer S |
| 2021-05-31 | Modified | Shorser S |

##### Entities found in this pathway (1)

| Input | UniProt Id |
| --- | --- |
| SLC4A4 | Q9Y6R1 |

### 6. Identifiers found

Below is a list of the input identifiers that have been found or mapped to an equivalent element in Reactome, classified by resource.

#### Entities (134)

| Input | UniProt Id | Input | UniProt Id | Input | UniProt Id |
|---|---|---|---|---|---|
| ABCB4 | P21439 | ABI1 | Q8IZP0 | ACSBG1 | Q96GR2 |
| ADCY8 | P40145 | AGT | P01019 | ALDH1A1 | P00352 |
| ALDH9A1 | P49189 | AMOT | Q4VCS5-1 | ANGPT1 | Q15389 |
| APOE | P02649 | AQP1 | P29972 | AQP4 | P55087 |
| AQP6 | Q13520 | ARC | Q7LC44 | ARHGAP24 | Q8N264 |
| ARHGEF6 | Q15052 | ARSF | P54793 | ATF3 | P18847 |
| BAG3 | O95817 | BBOX1 | O75936 | BCL2 | P10415 |
| CCL2 | P13500 | CCNH | P51946 | CCNL1 | P49736 |
| CD38 | P28907 | CDC37L1 | Q7L3B6 | CDC42EP4 | Q9H3Q1 |
| CDO1 | Q16878 | CLEC7A | Q9BXN2 | COL6A3 | P12111 |
| CSF2RA | P15509 | CTSH | P09668 | CX3CR1 | P49238 |
| CYP4F12 | Q9HCS2 | DAO | P14920 | DBI | P07108 |
| DDAH1 | O94760 | DIO2 | Q92813 | DUSP5 | Q16690 |
| EDNRB | P24530 | EID3 | Q8N140 | EIF4A3 | P38919 |
| ELOVL2 | Q9NXB9, Q9NYP7 | EZR | P15311 | F3 | P13726 |
| F5 | P12259 | FADS2 | O95864 | FGF2 | P09038 |
| FRS2 | Q8WU20 | GADD45B | P41440 | GFAP | P14136 |
| GLUD1 | P00367 | GPR161 | Q8N6U8 | H3F3B | P84243 |
| HK2 | P52789, P52790 | HSPB8 | Q9UJY1 | ID4 | P47928 |
| IDI1 | Q13907 | IL33 | O95760 | JAM2 | P57087 |
| JUN | P05412 | KAL1 | P23352 | KCNE1L | Q9UJ90 |
| KCNJ10 | P78508 | KCNN3 | Q9UGI6 | KLF4 | O43474 |
| LAMA1 | P25391 | LCP2 | Q13094 | LDLR | P01130 |
| LMNA | P02545-1, P02545-2 | LONRF1 | Q17RB8 | LRIG1 | Q96JA1 |
| MARCKS | P29966 | MASP1 | P48740 | MCL1 | Q07820 |
| MEI4 | P58012 | MGAT4C | Q9UBM8 | MLC1 | P60660 |
| MMP19 | Q99542 | MT1E | P04732 | MYO10 | Q9HD67 |
| NCAN | O14594 | NDEL1 | Q9GZM8 | NFIA | Q12857 |
| NPFFR1 | Q9GZQ6 | NUPL1 | Q9BVL2-1, Q9BVL2-2 | NWD1 | Q96DI7 |
| OLR1 | P78380 | P2RY1 | P47900 | PFKFB2 | O60825 |
| PIGA | P37287 | PLCD3 | Q8N3E9 | PLIN4 | O60664 |
| PMP2 | P15090 | PNO1 | Q9NRX1 | PPP1R3C | Q9UQK1 |
| PRCP | P42785 | PREX2 | Q70Z35 | PSAT1 | Q9Y617 |
| PTPMT1 | Q8WUK0 | PTTG1 | O95997 | PYGM | P11216, P11217 |
| RAB31 | Q13636 | RGMA | Q96B86 | RIN2 | Q8WYP3 |
| RND2 | P52198 | RNF182 | Q8N6D2 | RPE65 | Q16518 |
| SAT1 | Q9H2H9 | SDC4 | P31431 | SDS | P20132 |
| SERPINA3 | P01011, P29622 | SLC19A2 | O60779 | SLC4A4 | Q9Y6R1 |
| SLC7A11 | Q9UPY5 | SLC7A2 | P52569-1, P52569-2 | SLCO1C1 | Q9NYB5 |
| SPON1 | Q9HCB6 | SRSF7 | Q16629 | TG | P00488 |
| TNC | P24821 | TP53BP2 | Q13625 | TRIB1 | Q96RU8 |

| Input | UniProt Id | Input | UniProt Id | Input | UniProt Id |
|---|---|---|---|---|---|
| TRMT12 | Q53H54 | TTPA | P49638 | TUBA1A | Q71U36 |
| USH1C | Q9Y6N9 | ZFP36 | P26651 | ZNF10 | P21506 |
| ZNF184 | Q99676 | ZNRF3 | Q9ULT6 | | |

| Input | Ensembl Id | Input | Ensembl Id | Input | Ensembl Id |
|---|---|---|---|---|---|
| ABCB4 | ENSG00000005471 | AGT | ENSG00000135744 | APOE | ENSG00000130203 |
| ARC | ENSG00000198576 | ATF3 | ENSG00000162772 | BCL2 | ENSG00000171791 |
| CCL2 | ENSG00000108691 | F3 | ENSG00000117525 | FGF2 | ENSG00000138685 |
| GFAP | ENSG00000131095 | ID4 | ENSG00000172201 | IDI1 | ENSG00000067064 |
| KLF4 | ENSG00000136826 | LMNA | ENSG00000160789 | MCL1 | ENSG00000143384 |
| MT-TP | ENST00000387461 | RNU4ATAC | ENST00000580972 | TRIB1 | ENSG00000173334 |

| Input | miRBase Id |
|---|---|
| MIR24-2 | MI0000081 |

### 7. Identifiers not found

These 122 identifiers were not found neither mapped to any entity in Reactome.

| | | | | | | | |
|---|---|---|---|---|---|---|---|
| AC009499.1 | AC017002.2 | AC022498.1 | AC072062.1 | AL138815.1 | AL662800.1 | ARRDC4 | ATP13A4-AS1 |
| C1orf168 | C1orf51 | C2orf88 | CCDC141 | CCDC163P | CCDC80 | CCNB1IP1 | CDCA7L |
| CRISPLD1 | CSRNP1 | CSRP2 | CTC-244M17.1 | CTC-297N7.5 | CTD-2555I5.1 | CTD-3247F14.2 | DBX2 |
| DCLK2 | DCLRE1CP1 | DPY19L3 | EMP1 | ENKUR | ERRFI1 | EXPH5 | FAM167A |
| FAM181A | FAM189A2 | FAM198B | FAM69C | FAT1 | FIBIN | FJX1 | GAREML |
| GEM | GPR125 | HCG22 | HNRNPA1P61 | HNRNPLL | HSDL2 | IFRD1 | IGDCC4 |
| ITM2C | KLF6 | LARP1P1 | LINC00152 | LINC00511 | LINC00920 | LINC00963 | LINC01094 |
| LIX1 | MIR22HG | MIR548AV | MIR645 | MMD2 | MRPL35P1 | MYO16 | NDP |
| NEBL | NFKBIZ | NHSL1 | NKAIN3 | NKAIN4 | OGFRL1 | OSGIN2 | PAMR1 |
| PCDHGC3 | PDLIM3 | PEBP4 | PIH1D2 | PIRT | PRRX1 | RASL12 | RFX4 |
| RN7SL473P | RN7SL600P | RNF219-AS1 | RNU1-60P | RNU2-63P | RNVU1-15 | RP11-106M7.1 | RP11-1124B17.1 |
| RP11-141B14.1 | RP11-148L24.1 | RP11-179A16.2 | RP11-180C16.1 | RP11-258O13.1 | RP11-318M2.2 | RP11-38P22.2 | RP11-41O4.1 |
| RP11-572C15.6 | RP11-57K17.1 | RP11-659E9.4 | RP11-667K14.4 | RP11-686G23.2 | RP11-85M11.2 | RP4-555D20.2 | RP4-668G5.1 |
| RP5-855F14.1 | RP6-91H8.5 | RPS16P5 | RPS3AP35 | SCRG1 | SELK | SLC25A33 | SLC39A12 |
| SLC9A3R1 | SNORD3B-2 | SNORD3D | SRPX | SSPN | TMEM47 | TP53INP2 | U3 |
| XIRP1 | YBX2 | | | | | | |
