## Supplementary figures and images for "Compartment-specific total RNA profile of Hippocampal and Cortical cells from Mesial Temporal Lobe Epilepsy tissue"

### Cx-mTLE non-HS_Cyt_updownspecific_Homo sapiens.jpg

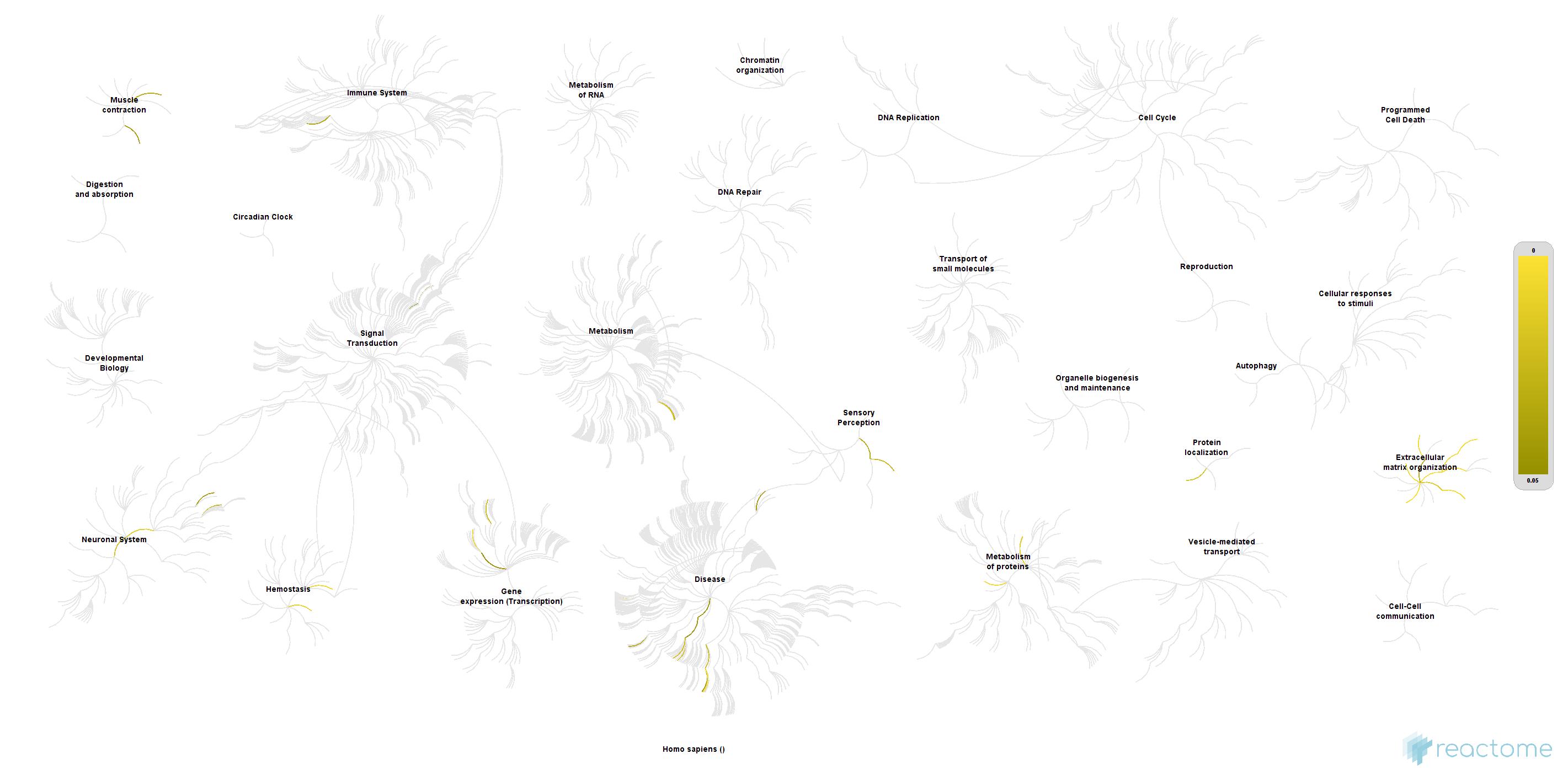

### Cx-mTLE non-HS_Nuc_updownspecific_Homo sapiens.jpg

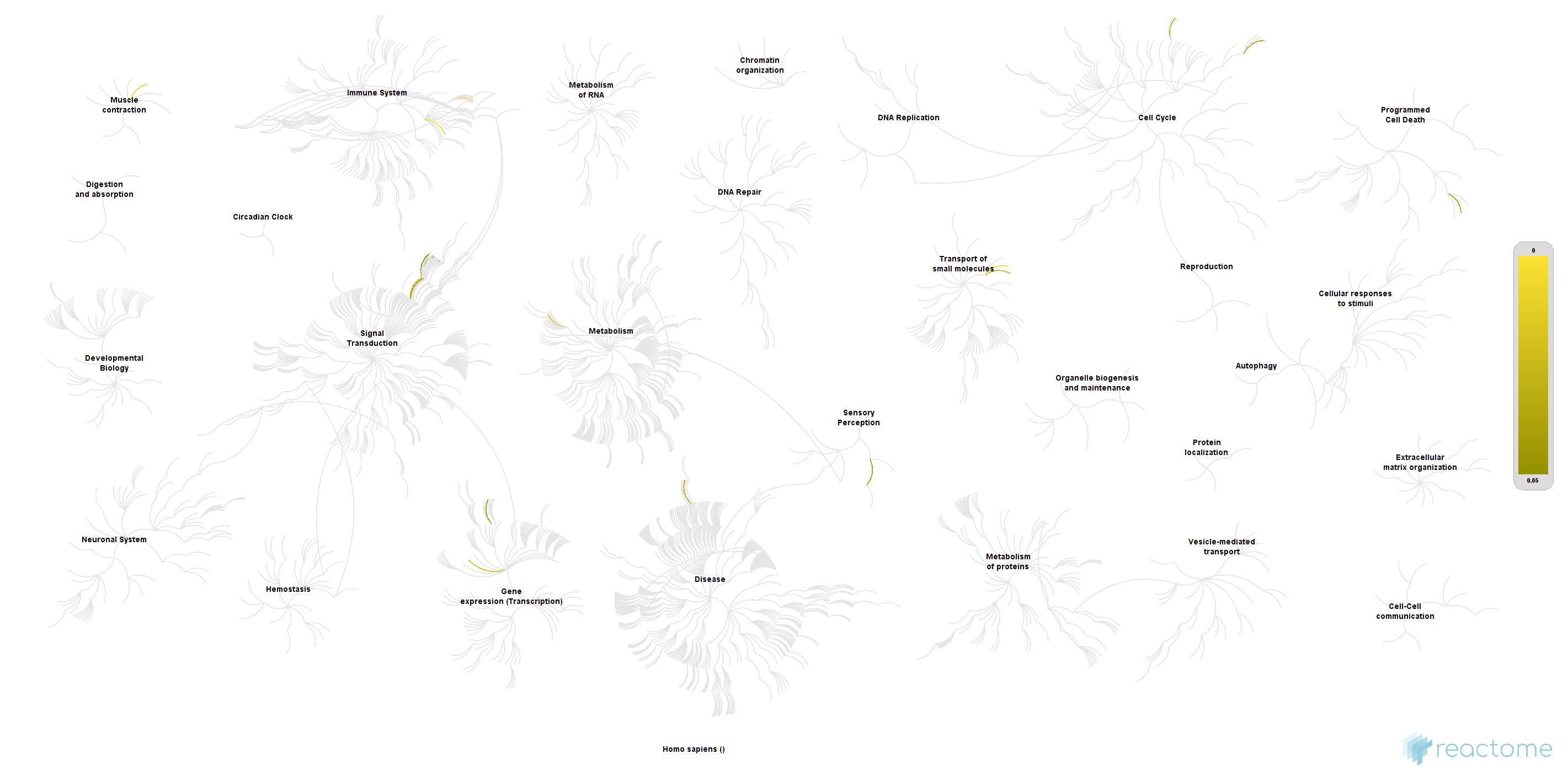

### Cx_mTLE+HS_Cyt_updownspecific_Homo sapiens.jpg

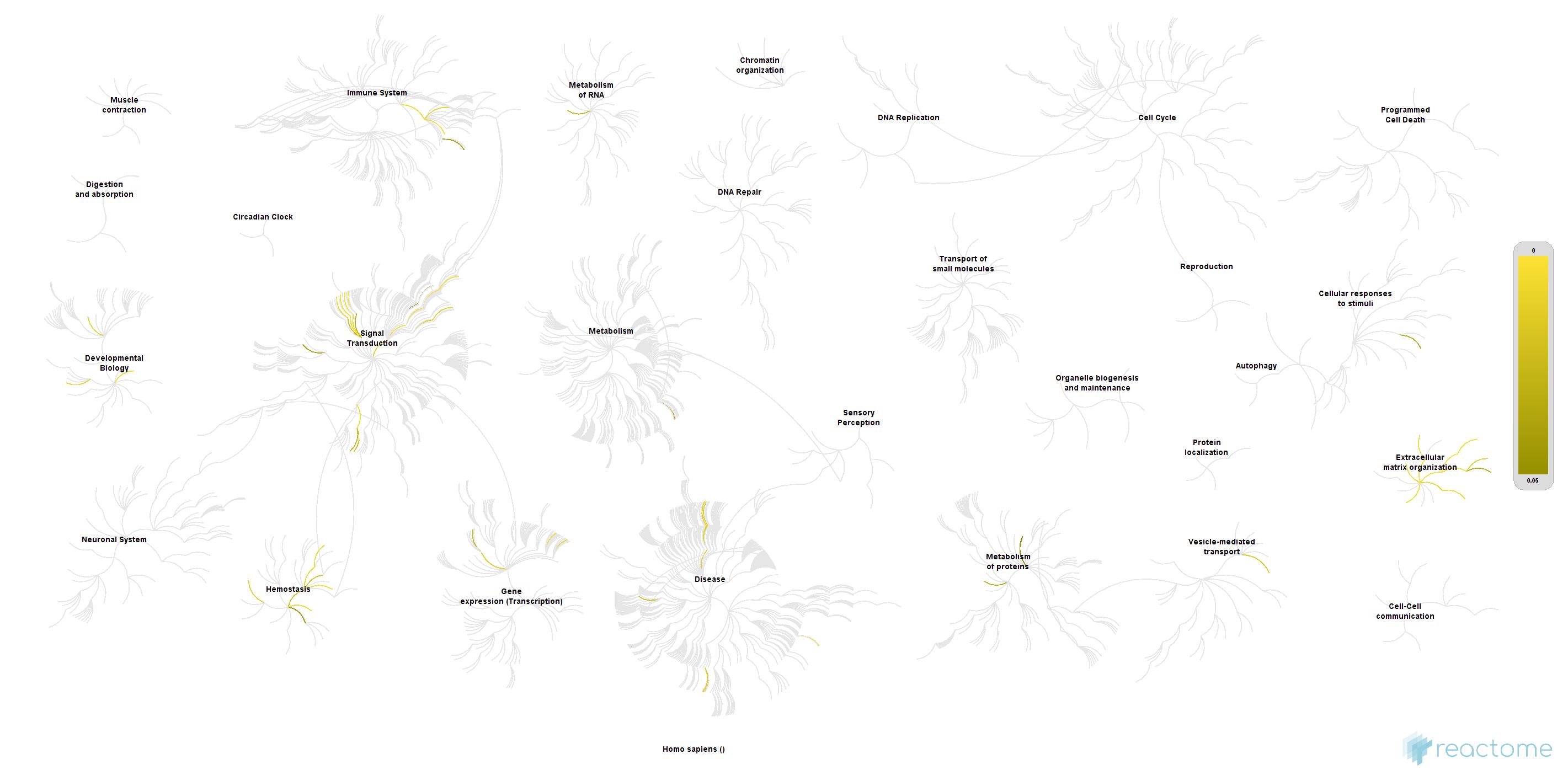

### Cx_mTLE+HS_Nuc_updownspecific_Homo sapiens.jpg

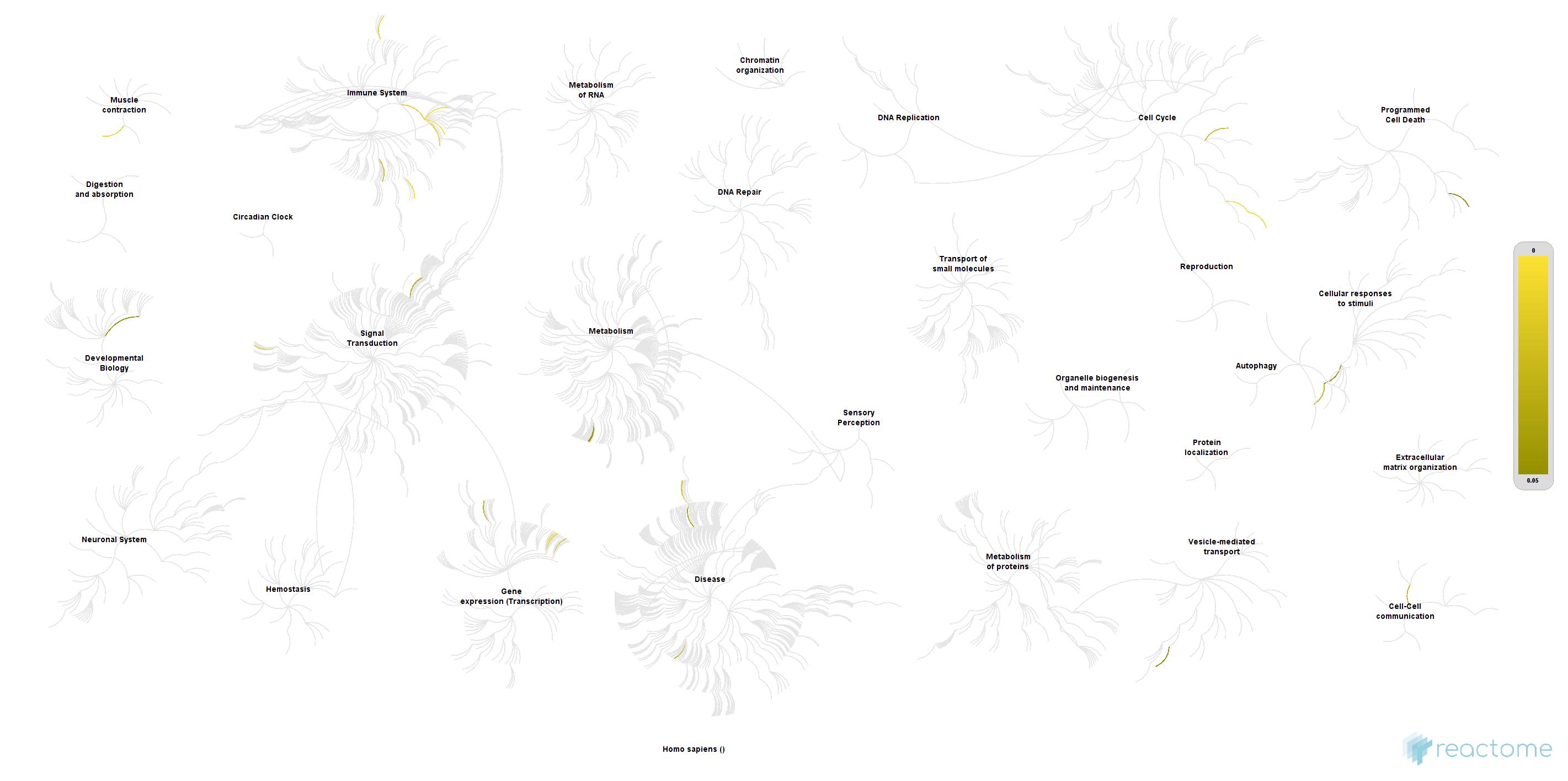

### HC_mTLE non-HS_Cytoplasm_DOWNspecific_Homo sapiens.jpg

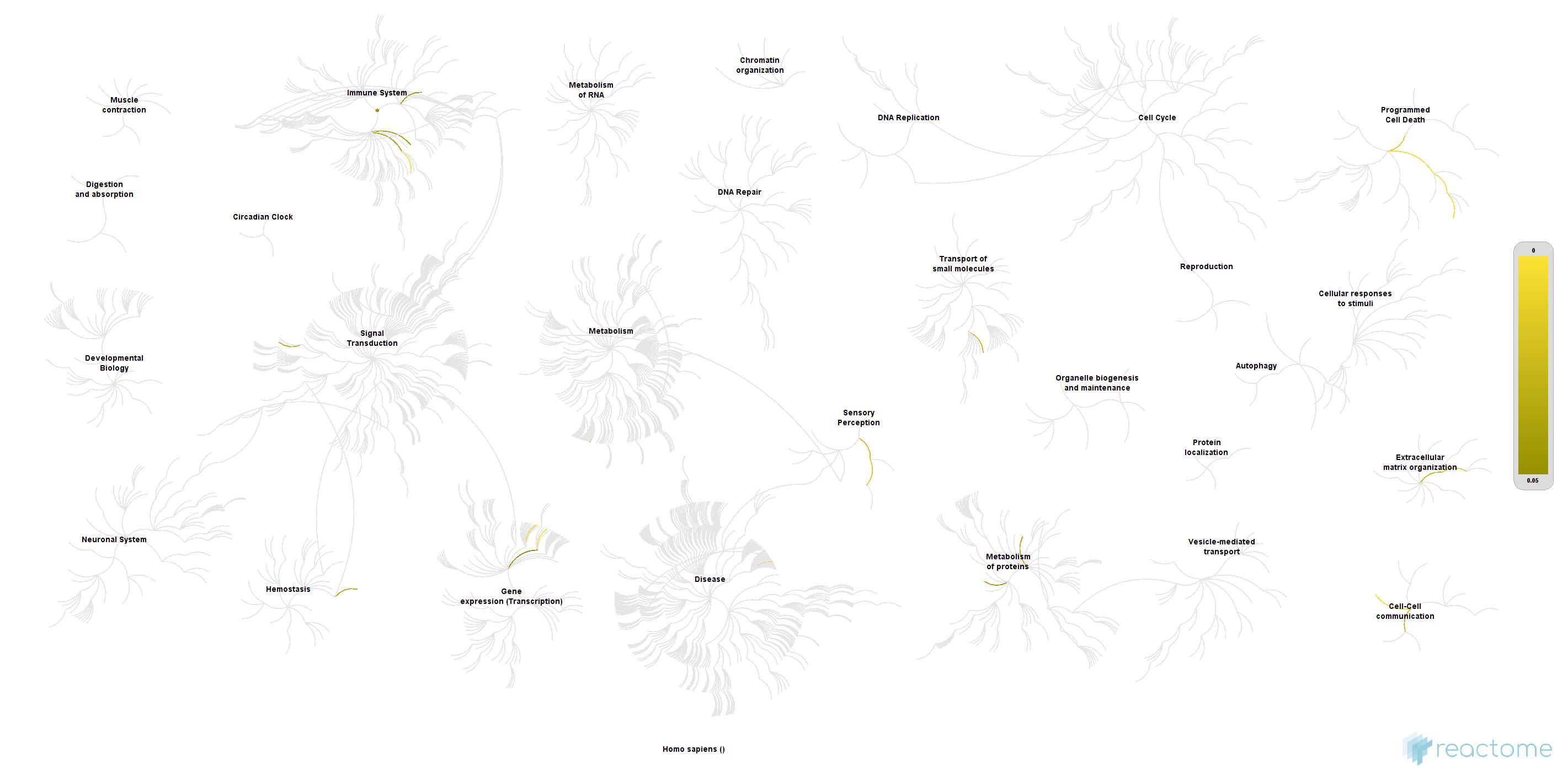

### HC_mTLE non-HS_Cytoplasm_UPspecific_Homo sapiens.jpg

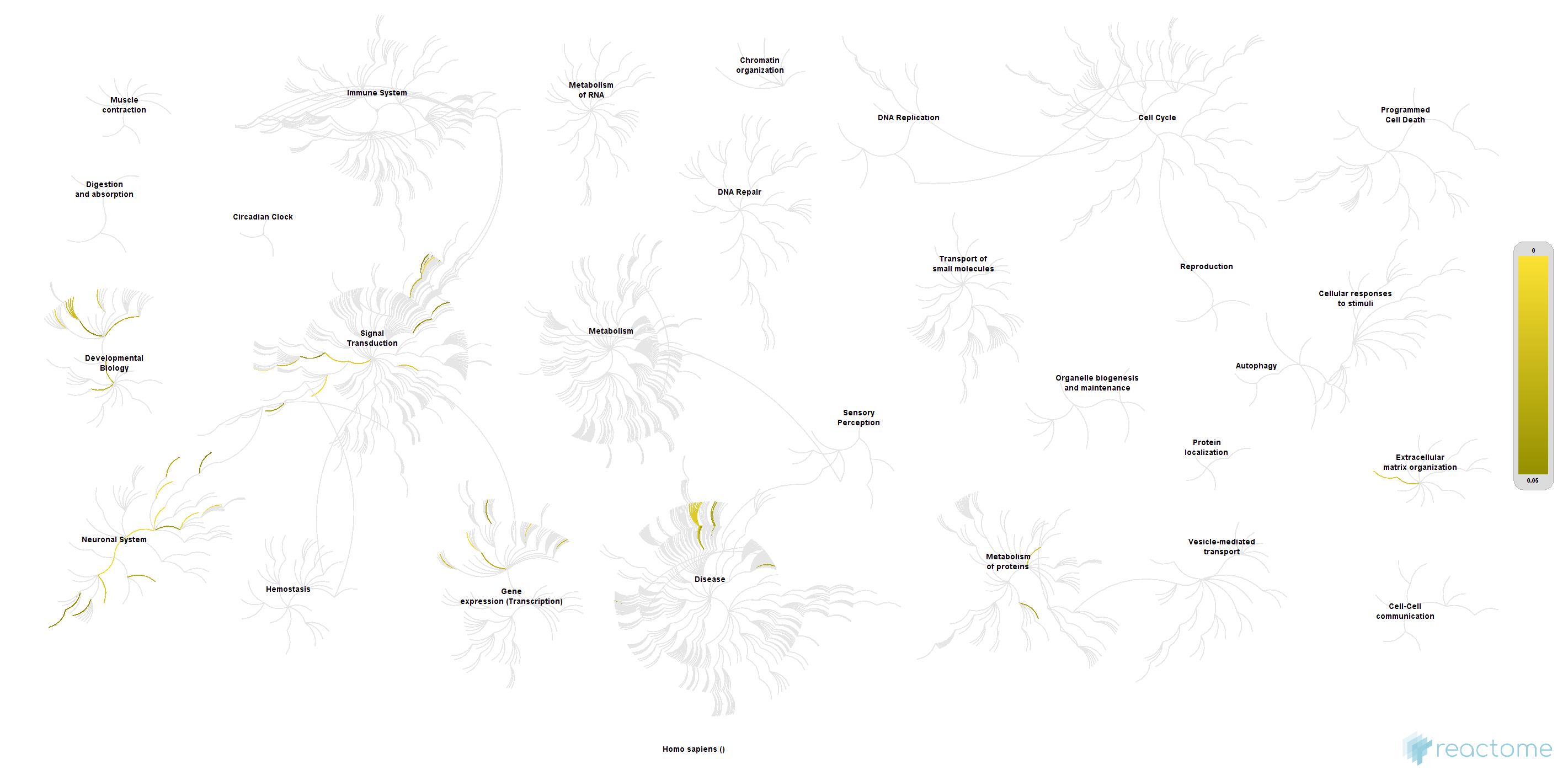

### HC_mTLE non-HS_Nucleus_DOWNspecific_Homo sapiens.jpg

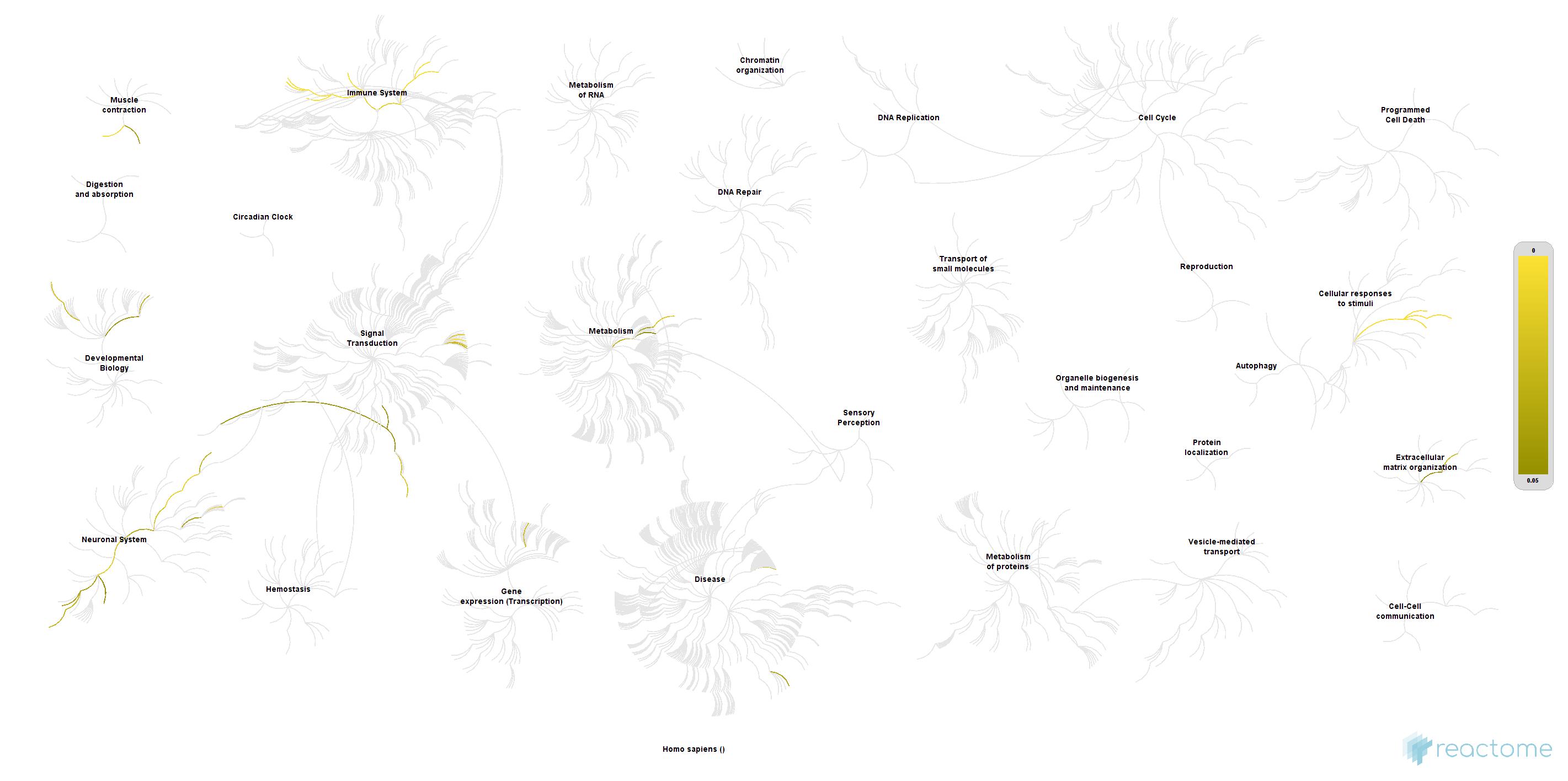

### HC_mTLE non-HS_Nucleus_UPspecific_Homo sapiens.jpg

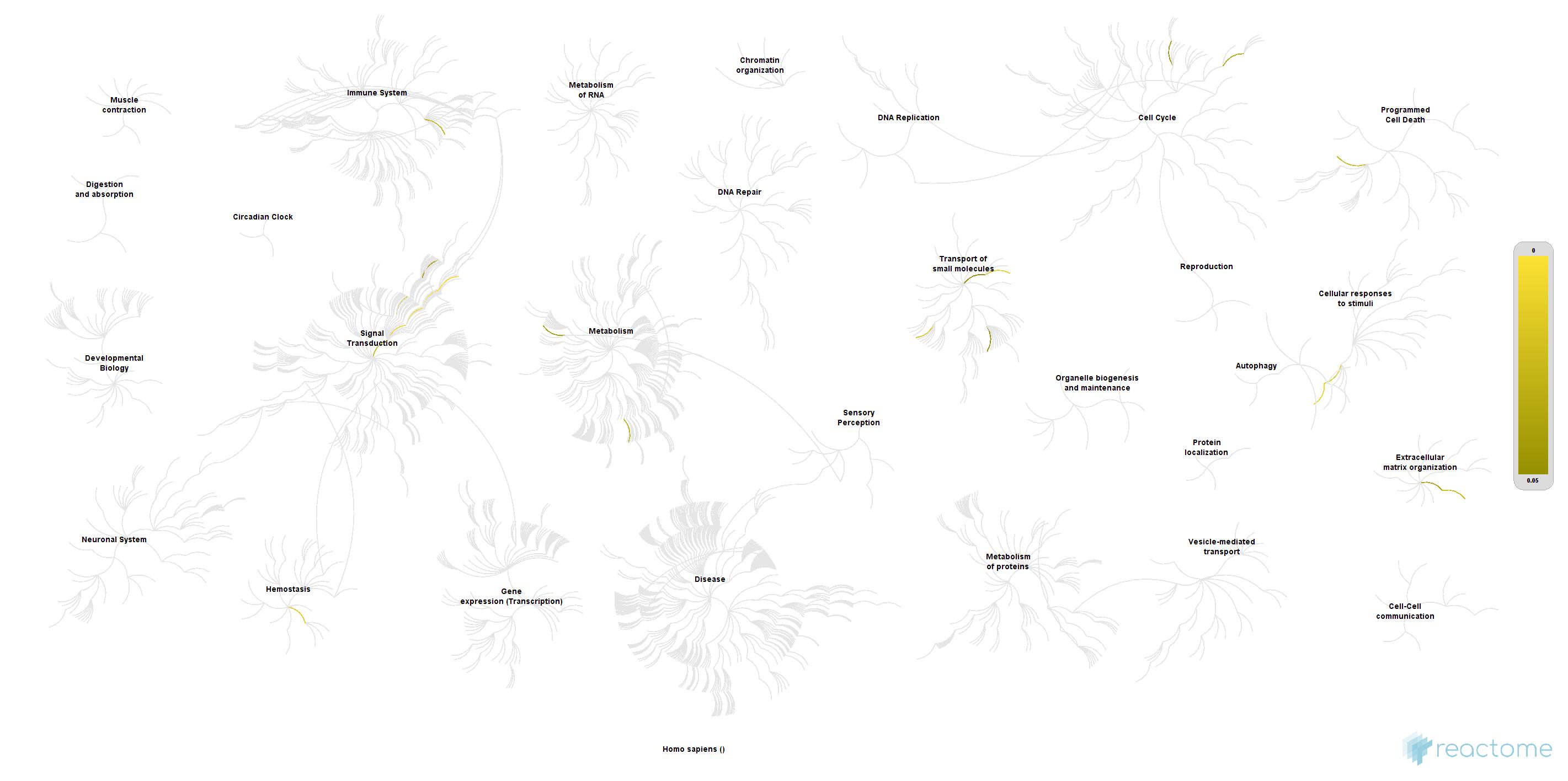
